# How age and underlying conditions affect incidence of hospitalization and death after documented COVID-19 diagnosis among individuals aged ≥20 years, a retrospective cohort study using PCORnet, April 2022 – March 2025

**DOI:** 10.64898/2026.09.18.26363440

**Authors:** Emilia H. Koumans, Gordana Derado, Melisa M. Shah, Nathan Graff, Christine Draper, Sharon Saydah, Melissa Briggs Hagen, Joshua L. Denson, Michael D. Kappelman, Deepika Thacker, Edward J. Schenck, Kshema Nagavedu, Tegan K. Boehmer, Diane Emerton, Thomas W. Carton, Pragna Patel, Jason P. Block, PCORnet^®^ Network Partners

## Abstract

**Background:** COVID-19 vaccinations, therapeutics, and immunity from infection have reduced COVID-19 severity. It is unknown to what extent previously identified risk factors – age and underlying medical conditions (UC)--remain associated with severe COVID-19 incidence.

**Methods:** Using electronic health record (EHR) data from PCORnet, we identified individuals aged ≥20 years with COVID-19 during April 2022-March 2025. We used Poisson regression with cluster-robust sandwich standard errors to estimate adjusted cumulative incidence (aCI) and aCI ratios (aCIR) for hospitalization within 16 days and mortality within 30 days of COVID-19 diagnosis to estimate the association of UCs and UC categories, comparing those with an UC to those without (adjusted for sociodemographic and health characteristics). We examined aCI and aCIR for cancer, cardiac, hepatic, immunosuppression, metabolic, neurologic, pulmonary, renal, psychiatric, and other conditions by age groups: 20-49, 50-64, 65-79, and 80+ years.

**Results:** Among 1,662,679 patients with COVID-19, the cumulative incidence of hospitalization (4.7%, 6.3%, 10.8%, 22.6%) and mortality (0.11%, 0.42%, 1.0%, 3.6%) increased across the 20-49, 50-64, 65-79, and 80+ year old age groups, respectively. Within each age group, aCI for hospitalization and mortality increased with number of UCs, with the highest aCI among those with 3 or more UCs. aCIR by age group demonstrated that UCs, especially 2 or more, were associated with incidence of both outcomes in all age groups.

**Conclusion:** Since 2022, older age has remained strongly associated with hospitalization and mortality after COVID-19 diagnosis. Adults with two or more UCs generally had higher incidence of both outcomes across age groups.

**summary:** Since 2022, older age has remained strongly associated with hospitalization and mortality with COVID-19. Having two or more of certain underlying conditions is associated with these outcomes among adults younger than 65 years.

## Introduction

During the early phase of the COVID-19 pandemic (2020–2021), older age and the presence of certain common underlying medical conditions (UC) emerged as risk factors for severe coronavirus disease 2019 (COVID-19), defined as hospitalization, ICU admission, intubation, or death.(1,2) Diabetes, obesity, chronic kidney disease, and immunosuppression were among the UC associated with severe COVID-19 outcomes.(3,4,5) These conditions are highly prevalent in the US population.(6) Age greater than 65 years was another important risk factor for severe COVID-19 and an indication for antiviral treatment, independent of UC.(7) A meta-analysis suggested a linearly increasing risk of mortality with increasing age after controlling for UC.(8) Other studies have shown that age was the main driver of risk for severe COVID-19 outcomes, and that there was an interaction between age and UC.(9,10)

Decreases in the incidence and severity of COVID-19 since 2021 have corresponded with several notable changes: availability of vaccines, increases in infection-induced immunity, the availability of antiviral medications, and variants associated with milder illness. The reported number of deaths from COVID-19 in the U.S. was estimated to be 463,267 in 2021 and 20,587 in 2025.(11) Hospitalization rates also declined, from 519 per 100,000 persons during the 2020-2021 to 22 per 100,000 during the 2025-2026 season.(12) Given the decrease in overall burden and severity of COVID-19, understanding the extent to which age and underlying conditions remain associated with incidence of severe disease since 2022 is important for informing strategies to prevent morbidity and mortality.

Defining risk factors for severe COVID-19 can direct patient and clinician education, inform vaccine policy, maximize prevention efforts, and encourage uptake of antiviral treatment among those with risk factors. However, understanding risk factors is complex, since UCs often co-occur, and UC prevalence increases with age. The extent to which age and UC remain similarly associated with severe COVID-19 infection since 2022 is unknown. Because there are interactions between age and UC(9), this study examined the role of underlying conditions in hospitalization and mortality after COVID-19, by age group, in 18 U.S. healthcare systems.

## Methods

### Study population

This study extracted electronic health record (EHR) data from ambulatory (outpatient, urgent care, telemedicine), emergency department, laboratory, hospital, and long-term care settings among 28,053,928 adults aged ≥20 years who were receiving care, defined as having had at least one encounter, at 18 U.S. health care systems participating in PCORnet during April 2022-March 2025.(13,14) PCORnet is a distributed research network that uses a Common Data Model in which sites collect, refresh, and curate routine clinical care data quarterly. The Common Data Model facilitates interoperability of data across sites, allowing for combination of data into a single analytic dataset. The requested PCORnet data was collected at each participating site and de-identified for three periods: April 2022-March 2023, April 2023-March 2024, and April 2024-March 2025. This activity was reviewed by the Centers for Disease Control and Prevention (CDC), deemed not research, and was conducted consistent with applicable federal law and CDC policy, US Department of Health and Human Services. (See e.g., 45 C.F.R. part 46.102(l)(2), 21 C.F.R. part 56; 42 U.S.C. §241(d); 5 U.S.C. §552a; 44 U.S.C. §3501 et seq).

Individuals aged ≥20 years were included if they had COVID-19, defined as meeting one of the following criteria: a laboratory-confirmed SARS-CoV-2 test result identified with Logical Observation Identifiers Names and Codes (LOINC)(Supplemental Material A), an International Classification of Diseases, Tenth Revision, Clinical Modification (ICD-10-CM)^§^ diagnostic code for COVID-19 (U07.1 or U07.2), or a prescription, administration, or procedure code for an outpatient COVID-19 treatment (Supplemental Material A). The COVID-19 diagnosis date(s) were determined based on date of diagnosis, prescription or treatment as described above. The index date was defined as the last or most recent COVID-19 diagnosis date during the three-year period (Figure 1) in cases where an individual had two or more index dates, to ensure identification of the most recent episode of illness and analyze the most recent incidence and risk factors.

**Figure 1.**
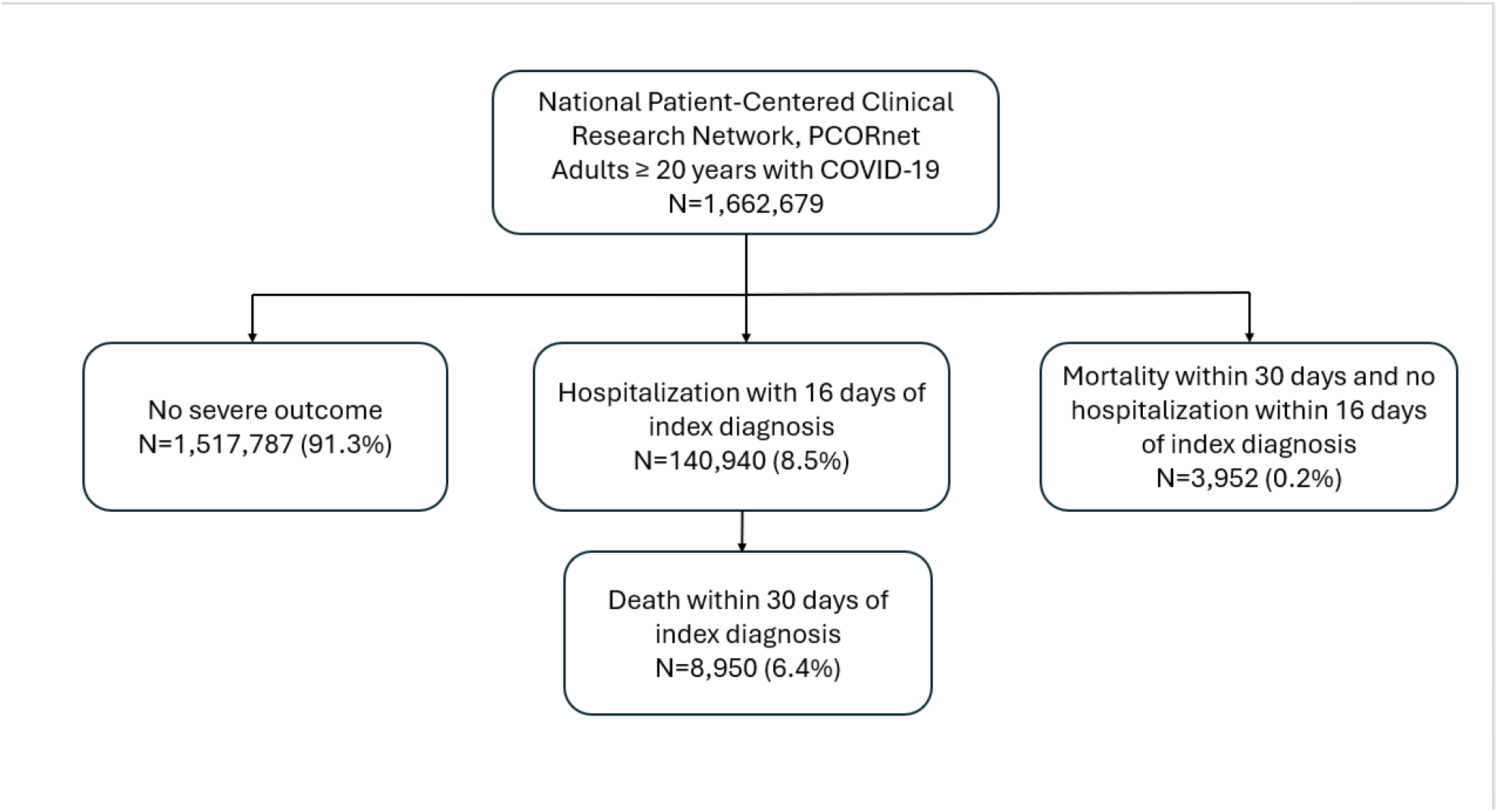
Flow diagram of individuals with COVID-19* meeting inclusion criteria and outcomes, PCORnet, April 2022-March 2025 *Adults aged ≥20 years with COVID-19 were defined as meeting one of the following criteria: 1) a laboratory-confirmed SARS-CoV-2 test result identified with Logical Observation Identifiers Names and Codes (LOINC), 2) an International Classification of Diseases, Tenth Revision, Clinical Modification (ICD-10-CM) diagnostic code for COVID-19 (U07.1 or U07.2), or 3) a prescription, administration, or procedure code of an outpatient COVID-19 treatment (nirmatrelvir-ritonavir, molnupiravir, monoclonal antibody, or remdesivir)

### Outcomes

Outcomes of COVID-19 severity included: any hospitalization within 16 days of the index date or mortality within 30 days of the index date. Hospitalizations were identified from inpatient records. Death was identified in the HER; although out-of-hospital deaths were reported, this was not systematic and varied by site. These time periods were chosen because COVID-19 can transition from mild to severe over several weeks, leading to hospitalization later in the course of illness; mortality may occur due to COVID-19 or a complication within a few weeks (15,16). One individual could contribute to one or both outcomes. Individuals were censored after 16 days for (last) hospitalization and 30 days for mortality.

### Underlying Conditions

The CDC list of underlying conditions that are risk factors for severe COVID-19 (22) included 29 conditions (23,24), which were captured by ICD-10-CM codes present in the EHR in the 3 years prior to the index date (Supplemental Material C). These 29 UCs were then categorized into 10 categories: (Table 2) cancer (all), cardiovascular (arrythmia, coronary artery disease, cardiac failure, cerebrovascular disease, peripheral vascular disease), severe liver disease (cirrhosis), immunosuppression (primary immunodeficiency, HIV, transplant, or immunosuppressing medication in the 365 days prior to index date [Supplementary Material D]), metabolic (severe obesity, BMI ≥30 kg/m^2^, type 1 diabetes, type 2 diabetes), neurologic (dementia, Parkinson’s disease), pulmonary (chronic pulmonary disease, chronic obstructive pulmonary disease [COPD], cystic fibrosis, pulmonary circulation disorder, ever or current smoking, asthma), renal (chronic renal failure, dialysis), psychiatric (schizophrenia, bipolar disorder, mood disorders), and other (pregnancy, tuberculosis [TB], Down Syndrome, hemiplegia).

**Table 1:**
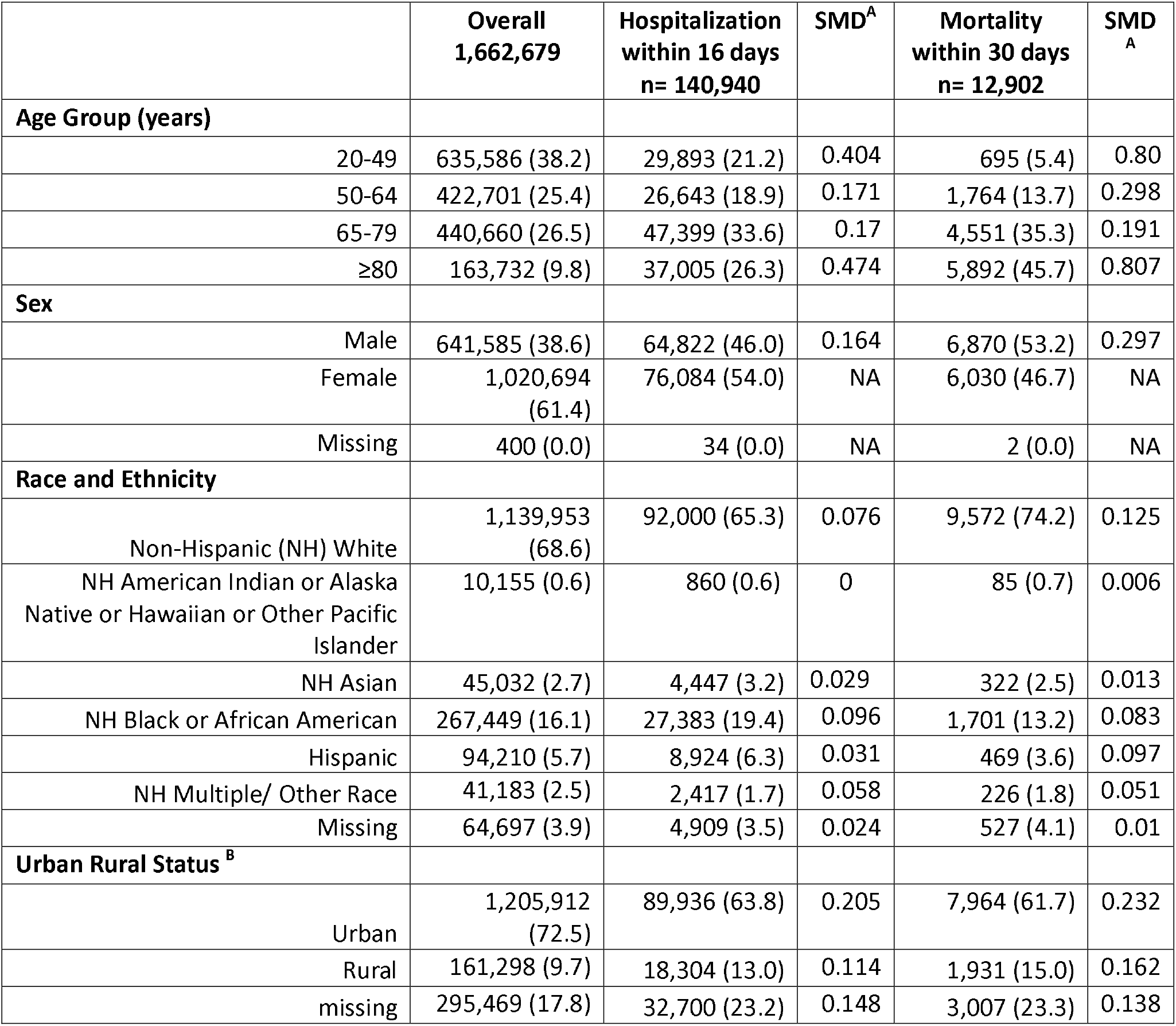
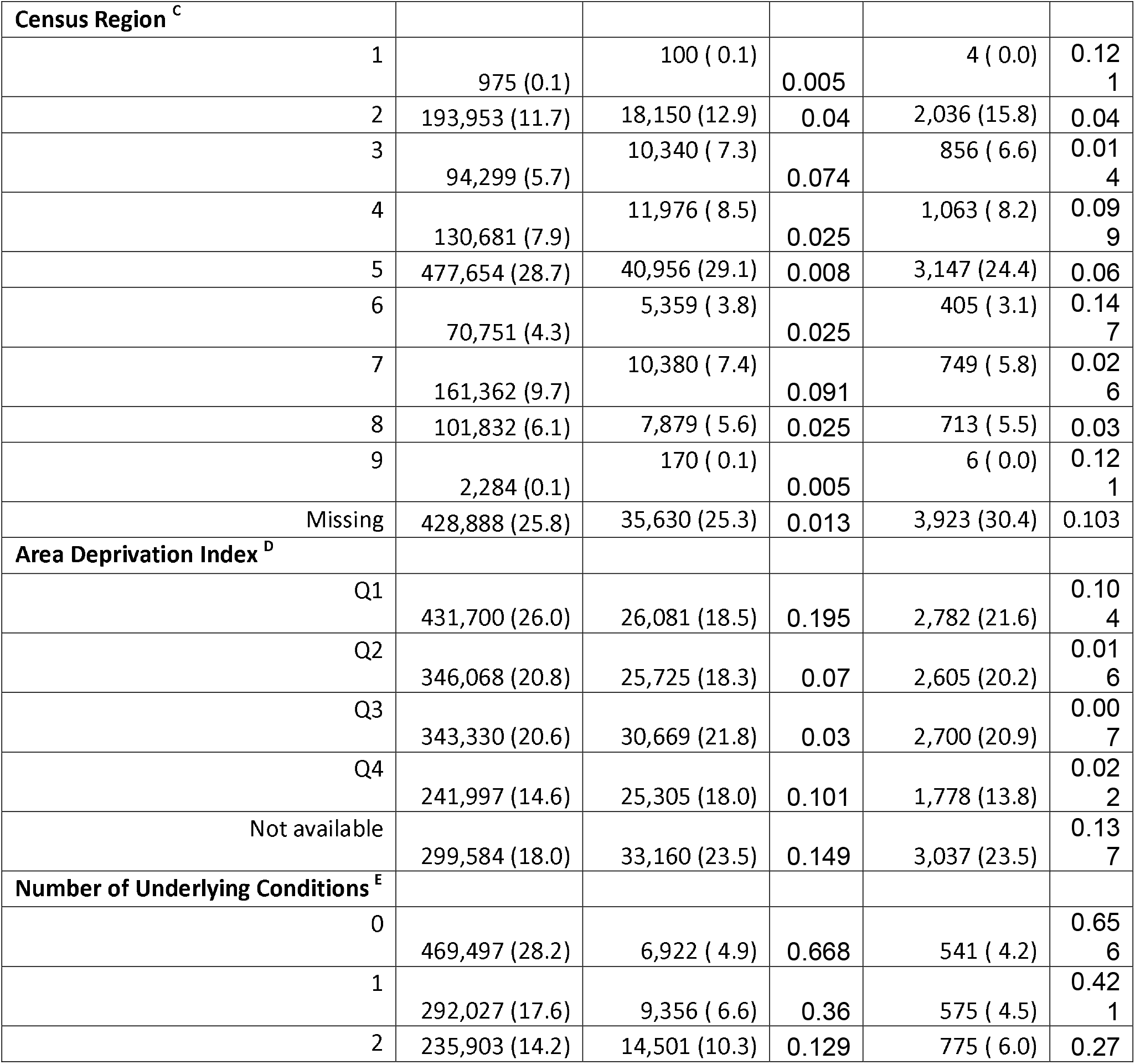
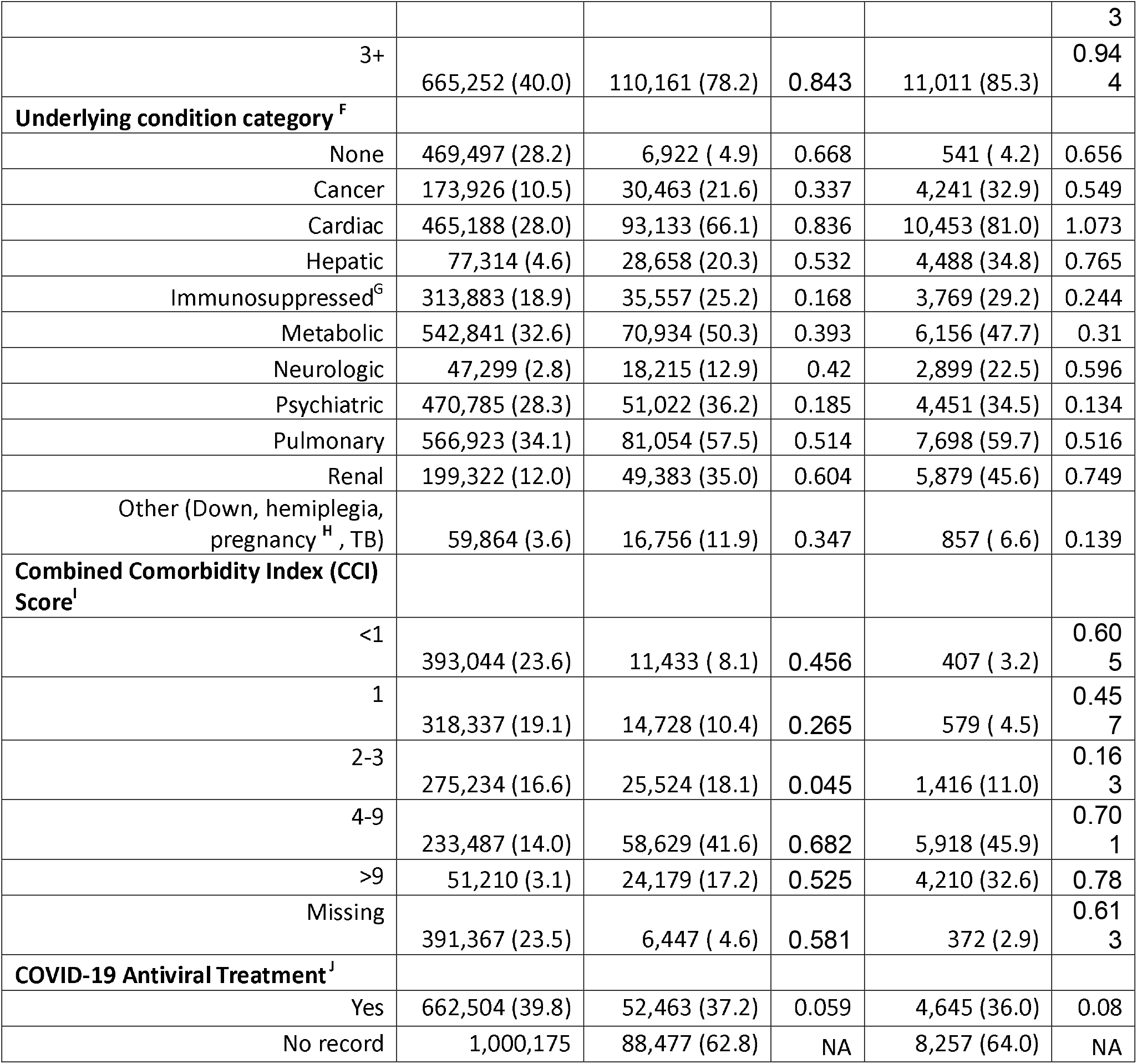
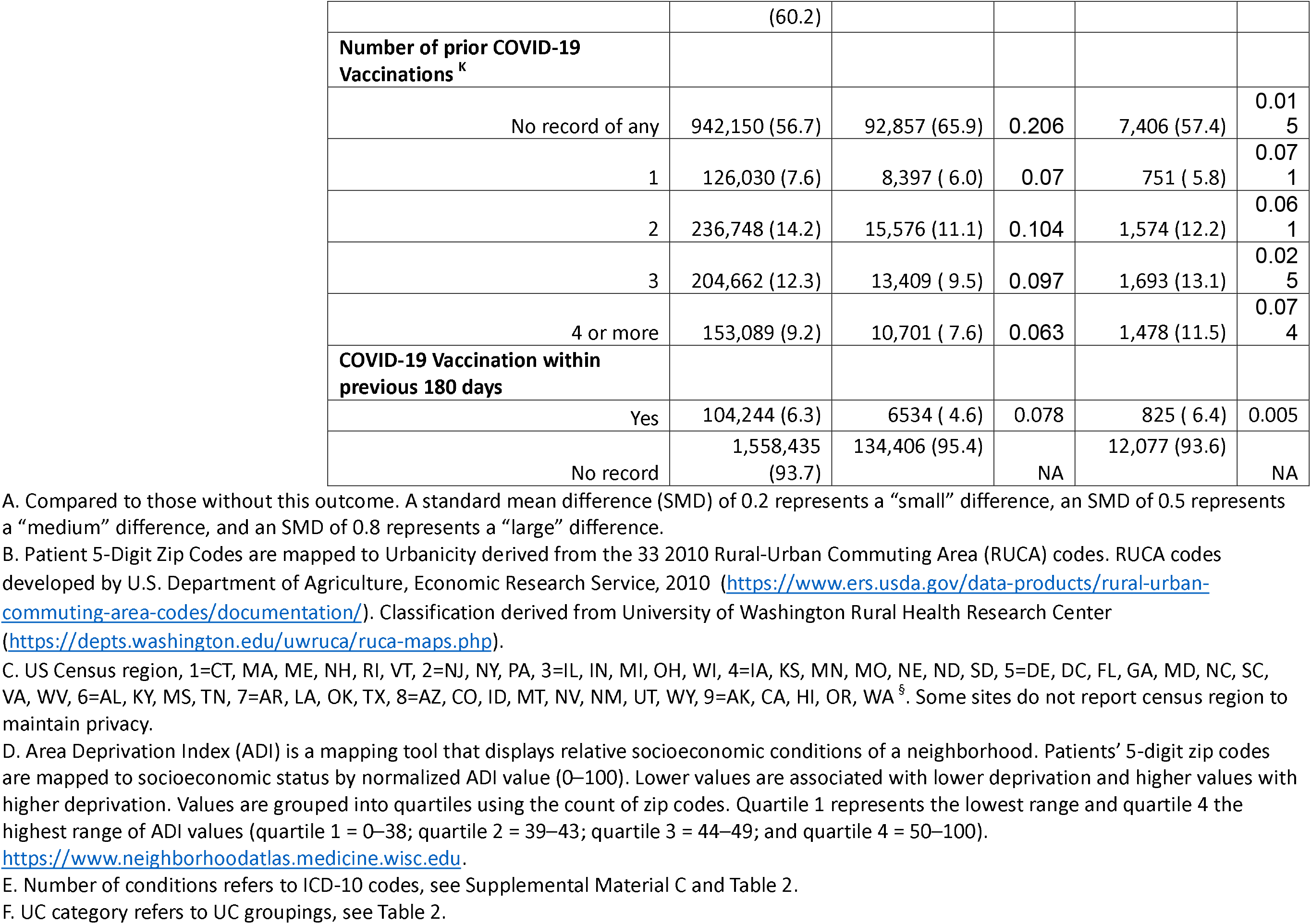
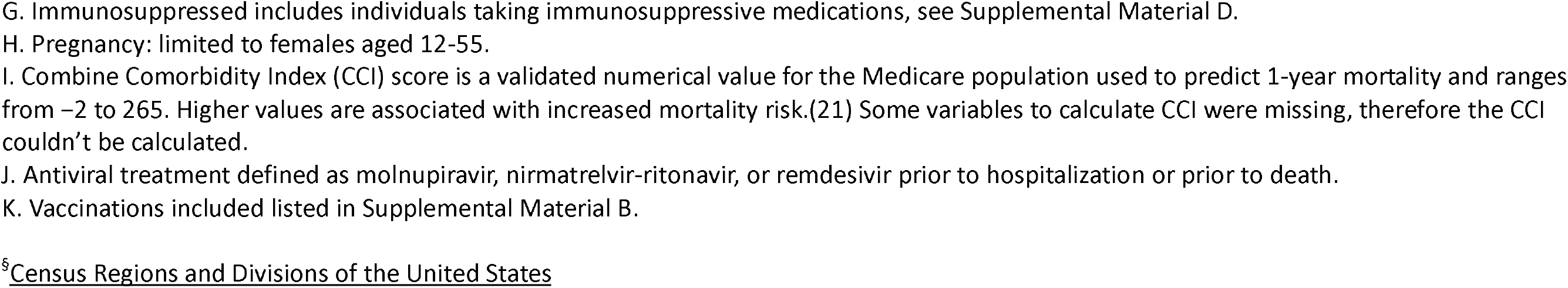
Demographic and health characteristics of adults aged ≥20 years with COVID-19, overall and by hospitalization within 16 days and mortality within 30 days of COVID-19 diagnosis, PCORnet, April 2022-March 2025, pre-mice imputation

**Table 2.** Underlying condition category, underlying condition, number and prevalence of each condition, and cumulative proportion with hospitalization within 16 days and mortality within 30 days among individuals aged ≥20 years with COVID-19, PCORnet, April 2022-March 2025

| Underlying Condition Category | Condition | Number with condition | Percent with this condition | Hospitalized within 16 days (%) | Mortality within 30 days (%) |
| --- | --- | --- | --- | --- | --- |
| Cancer | Cancer | 173,926 | 10.5 | 30,463 (17.5%) | 4,241 (2.4%) |
| Cardiovascular | Arrhythmia | 253,423 | 15.2 | 59,035 (23.3%) | 7,385 (2.9%) |
|  | Coronary Artery Disease | 205,678 | 12.4 | 47,170 (22.9%) | 5,535 (2.7%) |
|  | Cerebrovascular Disease | 141,386 | 8.5 | 36,243 (25.6%) | 4,365 (3.1%) |
|  | Congestive Heart Failure | 135,321 | 8.1 | 44,437 (32.8%) | 5,796 (4.3%) |
|  | Peripheral Vascular Disease | 139,117 | 8.4 | 34,408 (24.7%) | 4,280 (3.1%) |
| Hepatic | Severe Liver Disease (cirrhosis, liver failure) | 77,314 | 4.6 | 28,658 (37.1%) | 4,488 (5.8%) |
| Immunosuppression | Human Immunodeficiency Virus | 12,057 | 0.7 | 2,069 (17.2%) | 135 (1.1%) |
|  | Primary Immunodeficiency | 51,664 | 3.1 | 10,190 (19.7%) | 1,298 (2.5%) |
|  | Immunosuppressive Medication | 271,179 | 16.3 | 27,759 (10.2%) | 2,909 (1.1%) |
|  | Transplant | 26,521 | 1.6 | 6,419 (24.2%) | 579 (2.2%) |
| Metabolic | Obesity | 384,608 | 23.1 | 42,939 (11.2%) | 3,134 (0.8%) |
|  | Type 1 Diabetes | 18,280 | 1.1 | 3,590 (19.6%) | 245 (1.3%) |
|  | Type 2 Diabetes | 283,936 | 17.1 | 50,690 (17.9%) | 4,841 (1.7%) |
| Neurologic | Dementia | 39,082 | 2.4 | 16,437 (42.1%) | 2,710 (6.9%) |
|  | Parkinson's Disease | 11,472 | 0.7 | 3,204 (27.9%) | 405 (3.5%) |
| None | None | 469,497 | 28.3 | 6,925 (1.5%) | 541 (0.1%) |
| Other | Down Syndrome | 981 | 0.1 | 153 (15.6%) | 16 (1.6%) |
|  | Hemiplegia | 18,369 | 1.1 | 7,086 (38.6%) | 817 (4.4%) |
|  | Pregnancy | 39,578 | 2.4 | 9,343 (23.6%) | 5 (0.0%) |
|  | Tuberculosis | 1,059 | 0.1 | 219 (20.7%) | 22 (2.1%) |
| Pulmonary | Asthma | 196,062 | 11.8 | 18,475 (9.4%) | 1,168 (0.6%) |
|  | Chronic Pulmonary Disease | 338,166 | 20.3 | 47,907 (14.2%) | 4,805 (1.4%) |
|  | Chronic Obstructive Pulmonary | 124,854 | 7.5 | 33,064 (26.5%) | 3,873 (3.1%) |
|  | Disease |  |  |  |  |
|  | Cystic Fibrosis | 1,597 | 0.1 | 274 (17.2%) | 9 (0.6%) |
|  | Pulmonary Circulation Disorder | 32,471 | 2 | 9,503 (29.3%) | 1,267 (3.9%) |
|  | Ever Smoking | 352,705 | 21.2 | 61,768 (17.5%) | 5,744 (1.6%) |
| Renal | End Stage Renal Disease | 199,322 | 12 | 49,383 (24.8%) | 5,879 (2.9%) |
|  | Renal Dialysis | 25,012 | 1.5 | 10,095 (40.4%) | 1,236 (4.9%) |
| Psychiatric | Psychiatric | 470,785 | 28.3 | 51,022 (10.8%) | 4,451 (0.9%) |

### Sociodemographic and health characteristics

Sociodemographic and health characteristics were ascertained from the EHR. We included age group (20-49, 50-64, 65-79, and ≥80 years); sex (female, male); ethnicity and race (Non-Hispanic [NH] Asian, NH Black, NH Other, NH White, Hispanic); number of recorded COVID-19 vaccinations (Supplemental Material B) and recent COVID-19 vaccination, defined as a record of having received a COVID-19 vaccine within 180 days prior to index date; Area Deprivation Index (ADI) quartiles(17); U.S. Census region of residence(18); urban-rural residence(19,20); and prescription of COVID-19 antiviral treatment within 30 days of the index date (Supplemental Material A). We characterized individuals using the Combined Comorbidity Index (CCI),(21) a numerical value validated in the Medicare population used to predict 1-year mortality. CCI ranges from -2 to 265 with higher values associated with increased risk of mortality.

### Statistical analyses

We described the sociodemographic and health characteristics of individuals with COVID-19 overall and by each outcome (i.e., hospitalization and mortality). Crude prevalence estimates of sociodemographic and health characteristics, including UC, were calculated directly from observed data, and confidence intervals were derived using Wilson binomial methods. We calculated standard mean differences (SMD) for each sociodemographic characteristic or health condition comparing those who were hospitalized to those who were not hospitalized and those who died to those who did not die. We calculated Pearson’s correlation coefficients to assess the co-occurrence of two UC categories and plotted the results. Because there are interactions between age and UC(9) and UC categories were correlated (Figure 2), we calculated prevalences of each underlying condition, and cumulative incidence of hospitalization and mortality, by age group, using reported data.

**Figure 2.**
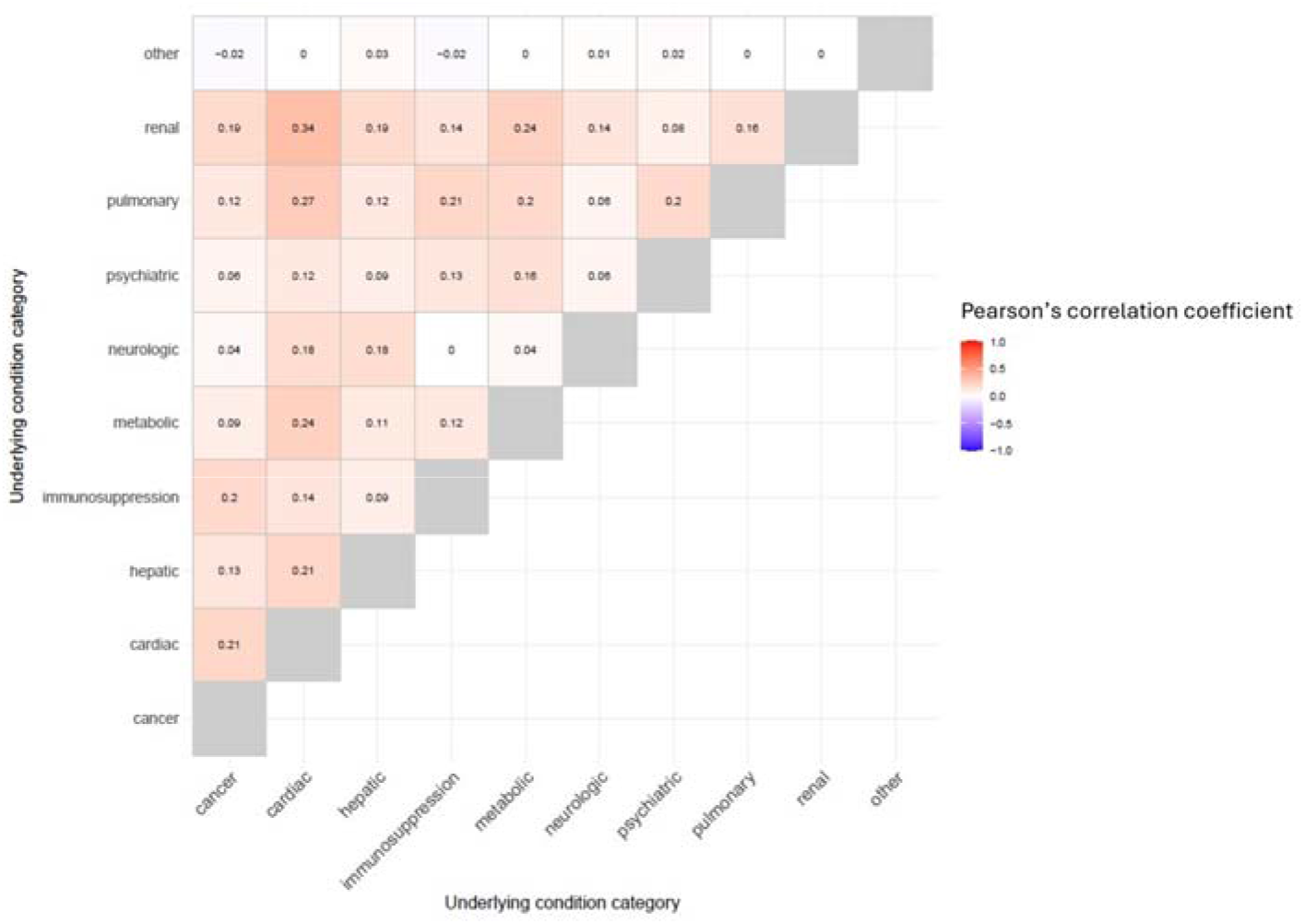
Co-occurrence of two underlying condition categories among individuals with COVID-19, graded color based on Pearson’s correlation coefficient, PCORnet, April 2022-March 2025

For modeling, missing covariate data were handled using multiple imputation by chained equations. Fifteen imputed datasets were generated using four iterations for each age group which converged. Race was imputed using multinomial logistic regression, binary variables including Hispanic ethnicity and urban-rural residence were imputed using logistic regression, and ordinal variables including ADI were imputed using proportional odds models. (26,27) Variables used as predictors in the imputation models included sex, total number of and recent vaccination, prescription for COVID-19 antiviral treatment, hospitalization, mortality, UCs, number of UCs, Census region, and site. The following variables were not themselves imputed: sex, age, vaccination and treatment indicators, outcome variables.(19,20,25,26,27) Estimates and variances were pooled across imputed datasets using Rubin’s rules.(28)

We determined the covariates for inclusion in models *a priori* based on pre-existing evidence for their association with COVID-19 disease severity, which included sex, ethnicity and race, ADI, urban-rural residence, recent vaccination, and prescription for COVID-19 treatment.(9,23,24) Separate age-group-specific Poisson regression models with a log link were fit for hospitalization and mortality outcomes, adjusting for sex, ethnicity and race, urban-rural residence, ADI, recent vaccination, and prescription for COVID-19 treatment including data that was imputed. Within each age group, adjusted cumulative incidence (aCI) of hospitalization and mortality were estimated using regression-based marginal standardization. Predicted outcome probabilities were averaged over the observed covariate distribution, yielding population-average (marginal) estimates for each underlying condition category.

We estimated adjusted cumulative incidence ratios (aCIR) of the outcomes adjusting for sex, ethnicity and race, rural-urban residence, ADI, recent vaccination and prescription for COVID-19 treatment, by age group. We used the adjusted models to estimate aCIR, by age group, for hospitalization and mortality. Variance estimation for all adjusted models for both outcomes accounted for clustering by site using cluster-robust sandwich estimators.

Because not all who were hospitalized died, and not all who died had been hospitalized, we conducted two sensitivity analyses by age. First, the aCIR of hospitalization by number of underlying conditions among those who were hospitalized and did not die. Second, the aCIR of mortality by number of underlying conditions among those who were not hospitalized and died. We used R software (version 4.4.2, R core team (2024)) for all analyses.

This study was reviewed by CDC, deemed not research, and conducted consistent with applicable federal law and CDC policy (45 CFR part 46.102(l)(2), 21 CFR part 56; 42 USC §241(d); 5 USC §552a; 44 USC §3501 et seq). This analysis followed the STrengthening the Reporting of OBservational Studies in Epidemiology (STROBE) reporting guidelines.(29) (Supplementary Material E)

## Results

### Description of study population

The study population included 1,662,679 unique individuals aged ≥20 years who had COVID-19 during April 2022 through March 2025 (Figure 1). A COVID-19 diagnostic code for COVID-19 was available for 1,245,560 (74.9%), a SARS-CoV-2 positive test for 619,380 (37.3%), and documented COVID-19 treatment for 573,195 (34.5%); 697,345 (41.9%) met two or more criteria. The 20-49 year age group comprised the largest proportion of individuals with COVID-19 (635,586, 38.2%); 1,116,575 (61.4%) patients were female (Table 1). The most common ethnicity and race group was Non-Hispanic White (68.6%), and 72.5% of individuals had an urban residence. Among included individuals, 6.3% of individuals had recent COVID-19 vaccination documented, and 39.8% had received a COVID-19 treatment within 30 days of the index date.

### Prevalence of underlying conditions and UC categories

Individuals with 3 or more underlying conditions comprised 40.0% of the cohort; 28.2% had no UC (Table 1). The prevalence of each UC category is provided in Table 1; 34.1%, 32.6%, and 28.0% of individuals had conditions in the pulmonary, metabolic, and cardiovascular categories, respectively. Of all individuals, 23.6% had a CCI score <1 and 18.9% had taken an immunosuppressive medication within the past year.

Multi-comorbidity was common. When examining the co-occurrence of UC categories, of the 619,837 individuals with a metabolic condition, 295,257 (47.6%) also had a pulmonary condition (Pearson’s correlation coefficient = 0.22) and 274,103 (44.2%) had a cardiovascular condition (coefficient=0.27); among the 600,727 individuals with a pulmonary condition, 275,905 (45.9%) also had a cardiovascular condition (coefficient=0.29) and 206,402 (34.4%) had a psychiatric condition (coefficient=0.23) (Figure 2).

While mental health disorders were the most prevalent UC in the 20-49, 50-64, and 65-79 year-old age groups, arrhythmias were the most prevalent in the ≥80 years age group. The prevalence of ever smoking, arrhythmias, coronary artery disease, chronic kidney disease, cancer, and COPD all increased with increasing age group (Figure 3).

**Figure 3.**
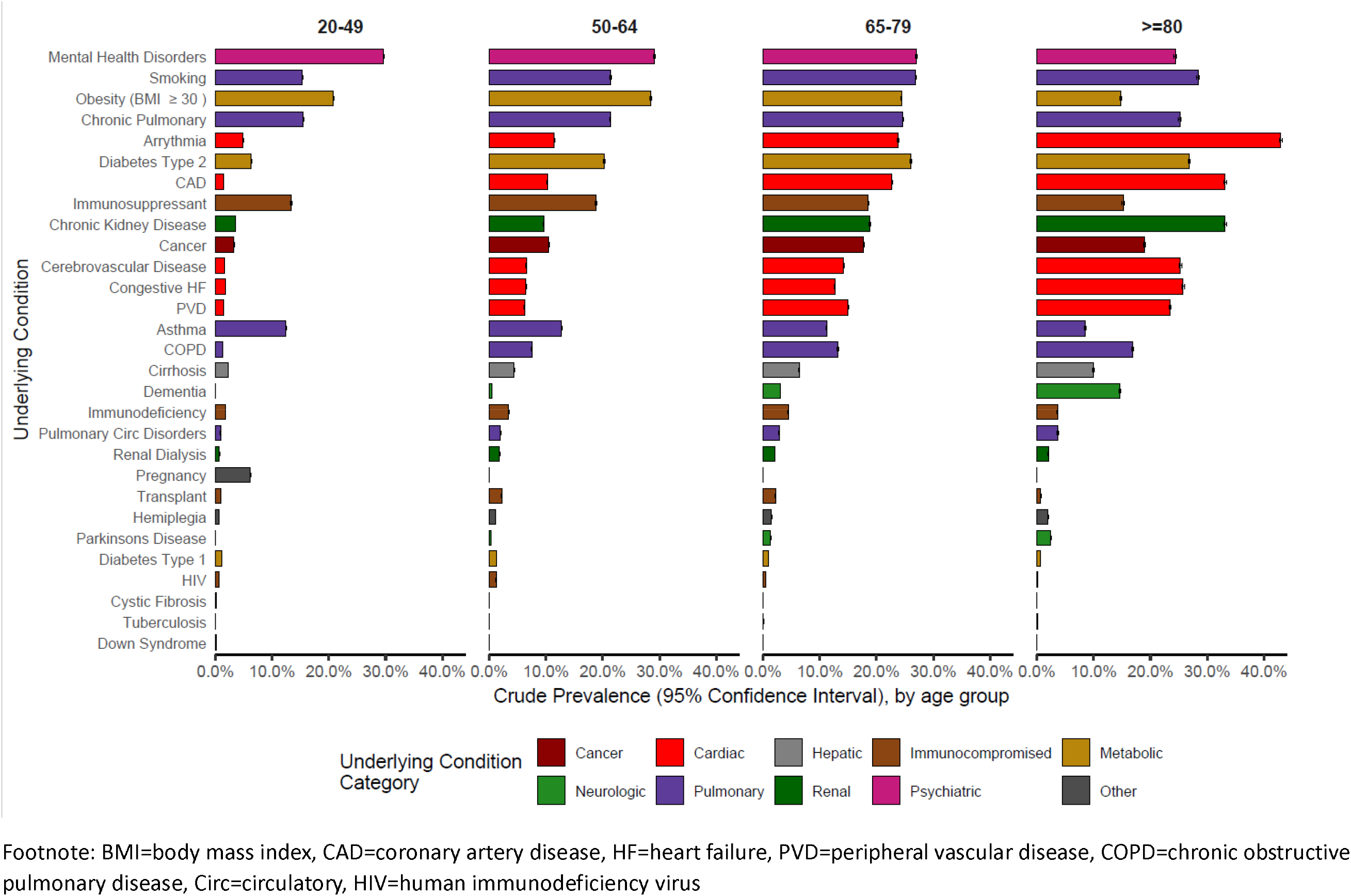
Crude prevalence and 95% Confidence Interval of 29 underlying conditions among individuals aged ≥20 years with COVID-19, by age group, PCORnet, April 2022-March 2025 Footnote: BMI=body mass index, CAD=coronary artery disease, HF=heart failure, PVD=peripheral vascular disease, COPD=chronic obstructive pulmonary disease, Circ=circulatory, HIV=human immunodeficiency virus

### Unadjusted cumulative incidence of hospitalization and mortality

Overall, 140,940 (8.5%) individuals were hospitalized within 16 days and 12,902 (0.8%) died within 30 days of the index date; a total of 144,892 (8.7%) were either hospitalized or died and 8,950 (69.4%) of those who died had a documented hospitalization (Figure 1). Individuals who were 65-79 years old were 33.6% of those hospitalized and 35.3% of those who died; individuals who were ≥80 years old comprised 26.3% of those hospitalized and 45.7% of those who died. Among those 20-49, 50-64, 65-79, and ≥80 years old, 4.7%, 6.3%, 10.8%, and 22.6%, were hospitalized - a more than 5-fold difference between the youngest and the oldest age groups - and 0.001%, 0.42%, 1.0%, and 3.6% died, respectively, a 36-fold difference.

### Underlying conditions, unadjusted, and adjusted (post-mice) cumulative incidence of hospitalization and mortality

Of the 469,497 individuals who had no recorded underlying condition, 6,922 (1.5%) were hospitalized and 541 (0.1%) died (Table 2). The highest unadjusted cumulative incidence of hospitalization across all age groups was among individuals with COVID-19 whose underlying conditions included renal dialysis (33.2%-48.4%), hemiplegia (29.8%-44.3%), dementia (29.2%-42.4%), and cirrhosis (27.8%-44.2%)(Figure 4). By UC category, the highest aCI of hospitalization across age groups was among those with conditions in the hepatic, neurologic, and renal UC categories (Figure 6). However, among all individuals who were hospitalized, the most common UCs were ever smoking (43.8%) and arrythmia (41.9%)(Table 2).

**Figure 4.**
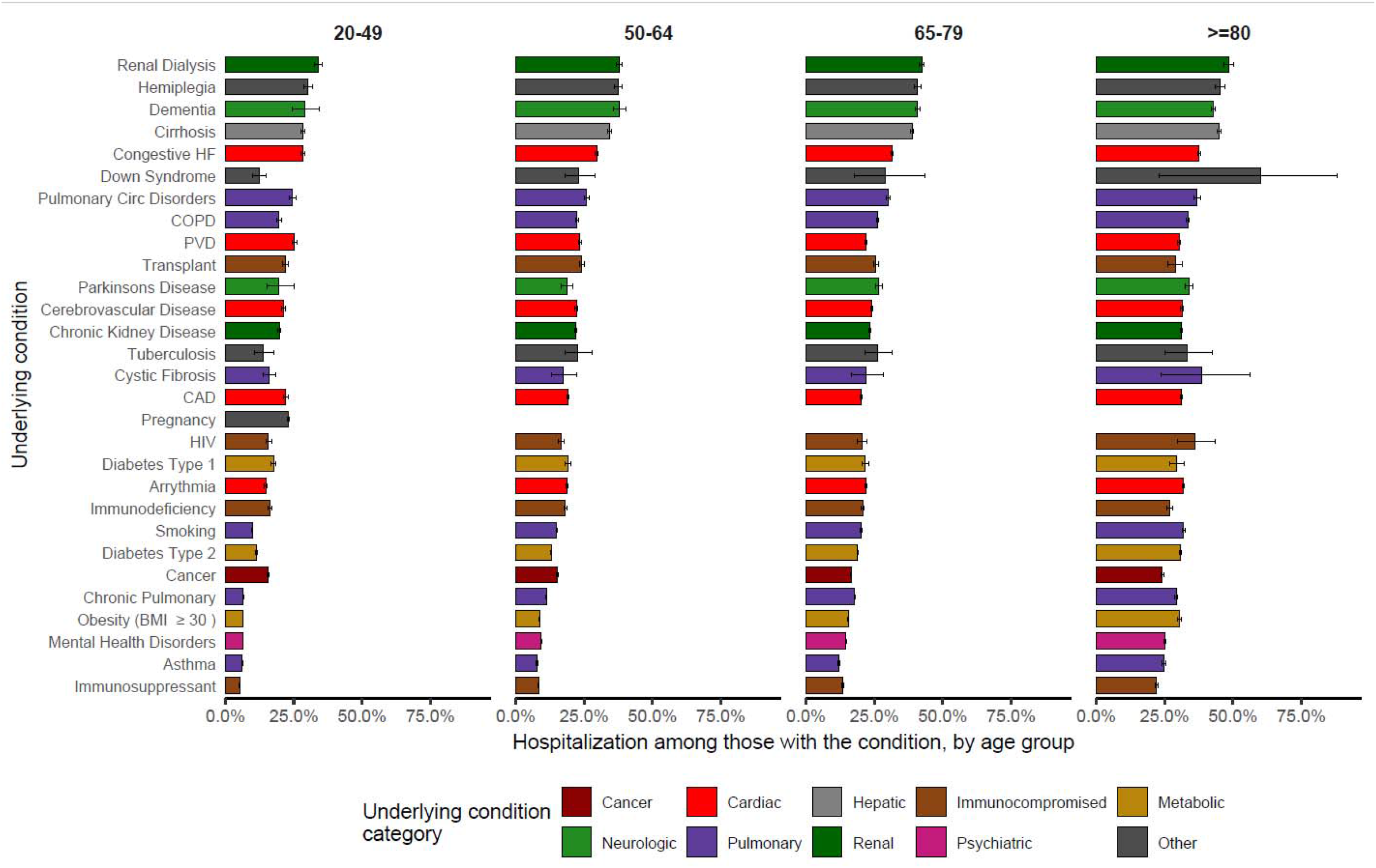
Crude incidence and 95% Confidence Interval of hospitalization within 16 days by age and underlying condition among individuals aged ≥20 years with COVID-19, PCORnet, April 2022-March 2025 Footnote: BMI=body mass index, CAD=coronary artery disease, HF=heart failure, PVD=peripheral vascular disease, COPD=chronic obstructive pulmonary disease, Circ=circulatory, HIV=human immunodeficiency virus

The highest unadjusted cumulative incidence of mortality across all age groups was among individuals with COVID-19 whose underlying conditions included cirrhosis (2.0%-9.4%), renal dialysis (1.2%-10.1%), hemiplegia (1.1%-8.8%), dementia (0.8%-8.0%), followed closely by pulmonary circulatory disorders and congestive heart failure (Figure 5). By UC category, the highest aCI of mortality across age groups was among those with conditions in the hepatic, neurologic, and renal UC categories (Figure 7). Among individuals who died, the most common UCs were arrythmia (57.2%) and chronic kidney disease (45.6%).

**Figure 5.**
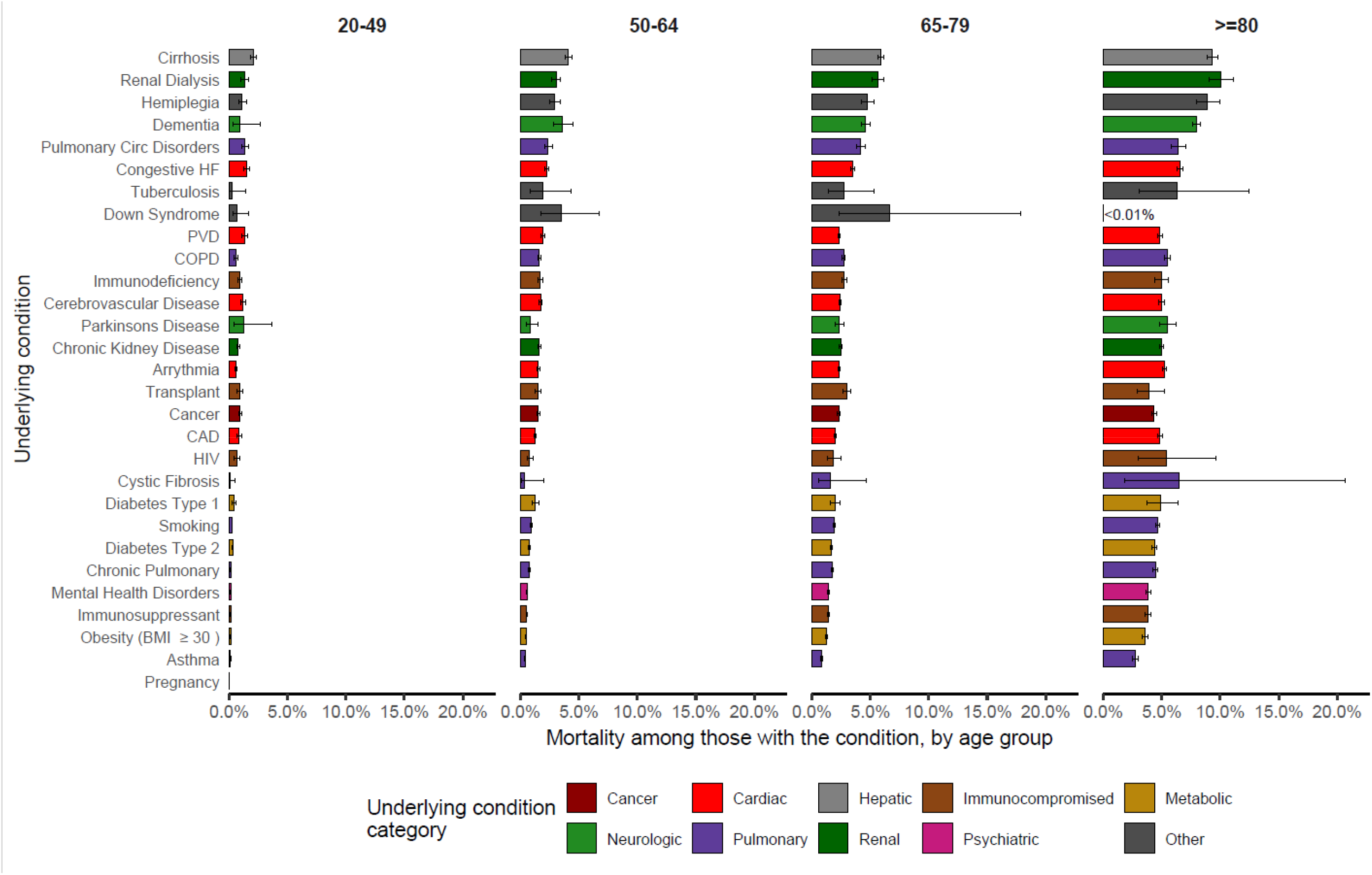
Crude incidence and 95% Confidence Interval of mortality within 30 days by age and underlying condition among individuals aged ≥20 years with COVID-19, PCORnet, April 2022-March 2025 Footnote: BMI=body mass index, CAD=coronary artery disease, HF=heart failure, PVD=peripheral vascular disease, COPD=chronic obstructive pulmonary disease, Circ=circulatory, HIV=human immunodeficiency virus Confidence Interval not shown for Down syndrome among persons aged ≥80 years because of sparse events; incidence was <0.01%

**Figure 6.**
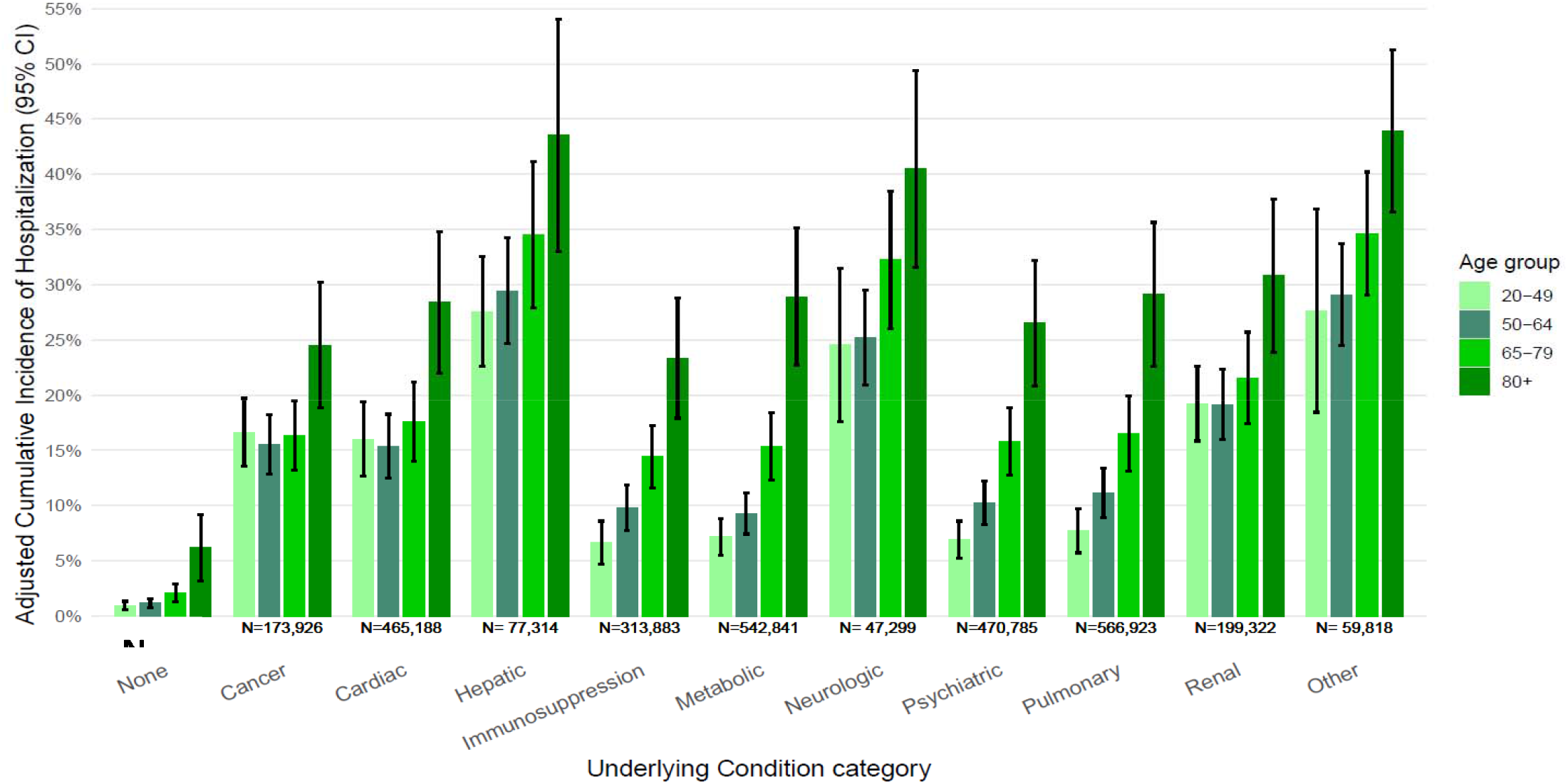
Adjusted cumulative incidence* and 95% Confidence Interval of hospitalization within 16 days of COVID-19 diagnosis by age and underlying condition category among individuals aged ≥20 years with COVID-19, PCORnet, April 2022-March 2025. * Separate age-group-specific Poisson regression models with a log link including imputed data were fit for hospitalization and mortality outcomes. All models were adjusted for sex, ethnicity and race, area deprivation index, rural urban residence, recent COVID-19 vaccination and COVID-19 antiviral treatment within 30 days of index COVID-19 diagnosis Caption: Individuals with multiple underlying conditions may be represented in more than one underlying condition category.

**Figure 7.**
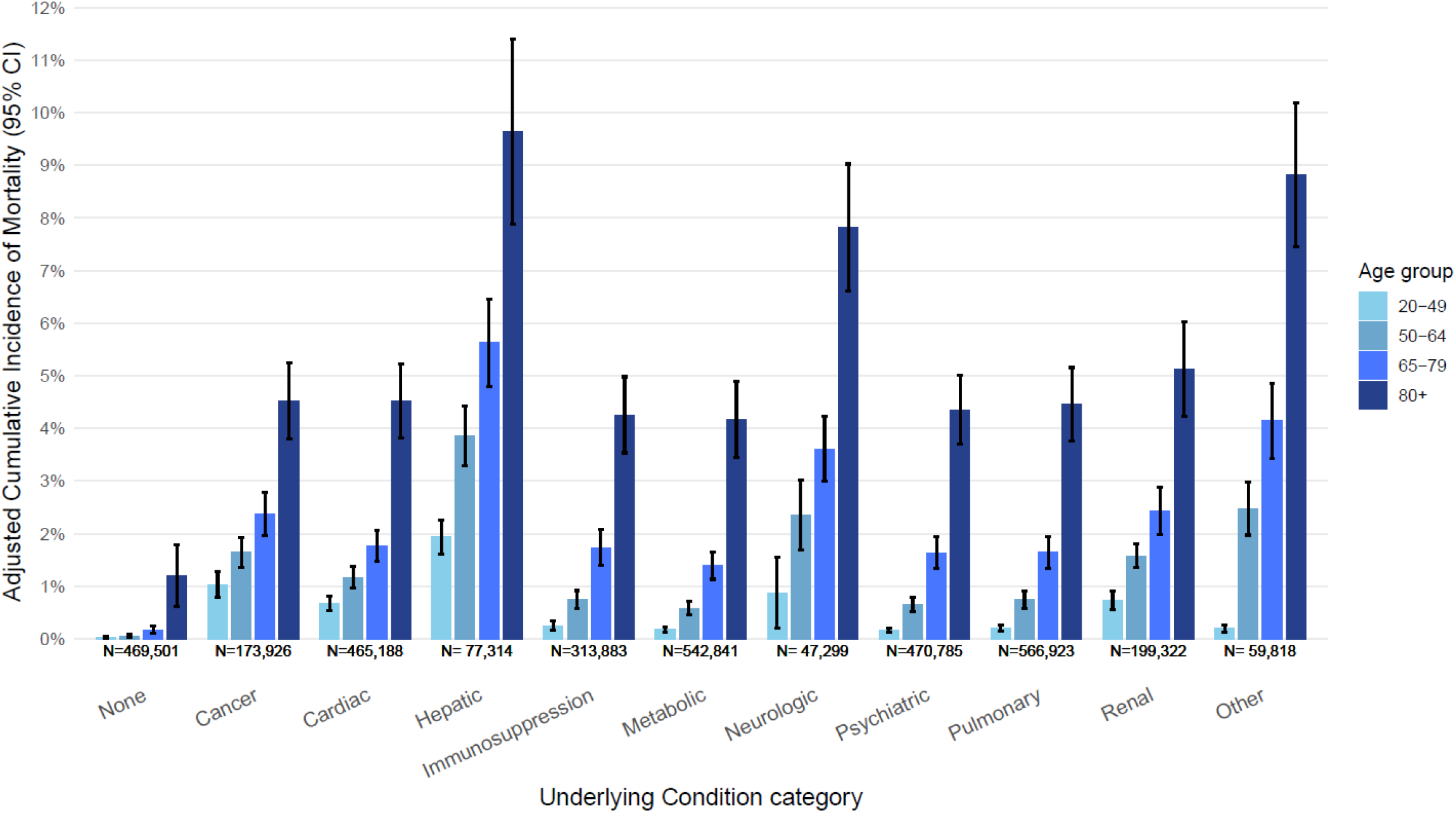
Adjusted cumulative incidence* and 95% Confidence Interval of mortality by age and underlying condition category among individuals aged ≥20 years with COVID-19, PCORnet, April 2022-March 2025. * Separate age-group-specific Poisson regression models with a log link including imputed data were fit for hospitalization and mortality outcomes. All models were adjusted for sex, ethnicity and race, area deprivation index, rural urban residence, recent COVID-19 vaccination and COVID-19 antiviral treatment within 30 days of index COVID-19 diagnosis. Caption: Individuals with multiple comorbidities may be represented in multiple categories; 29 underlying conditions were categorized into 10 categories within 3 years of COVID-19.

The aCI of hospitalization and mortality increased with increasing number of UC and with increasing CCI in each age group (Figures 8–11). The aCI of hospitalizations among those aged ≥80 years was 6.2%, 11.2%, 14.5%, and 29.5% for those with 0, 1, 2, or 3 or more UC, respectively (Figure 8). Among those with 1 or 2 UCs, the aCI of hospitalization was at least 50% higher than those without documented UC across all age groups except those aged 80 years or older. The aCI of mortality for the same age group was 1.2%, 1.6%, 2.0%, and 4.7% for those with 0, 1, 2, or 3 or more UC, respectively (Figure 9).

**Figure 8:**
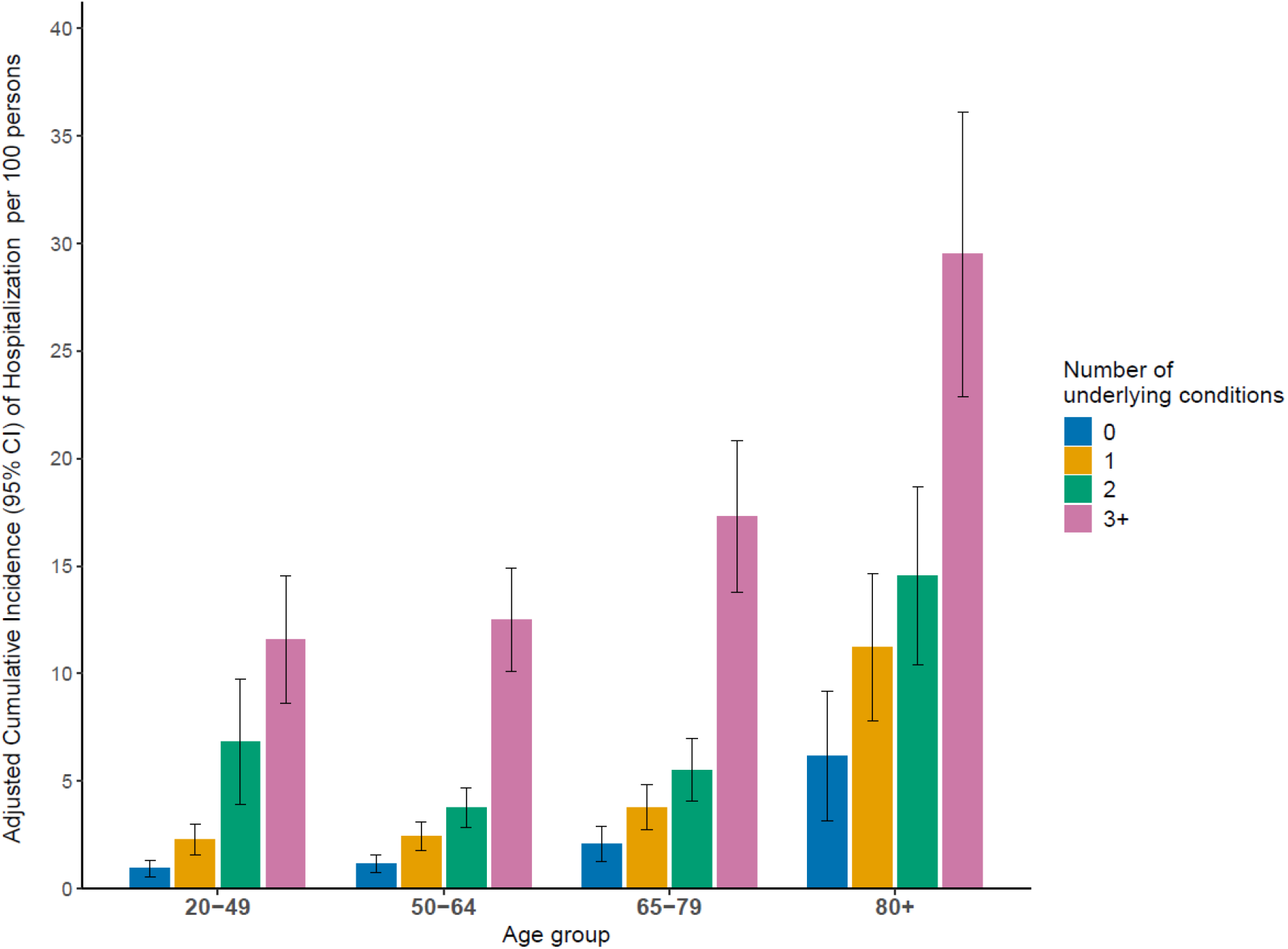
Adjusted cumulative incidence* and 95% Confidence Interval of hospitalization within 16 days among individuals aged ≥20 years with COVID-19 by age and number of underlying conditions (UC), PCORnet, April 2022-March 2025 * Separate age-group-specific Poisson regression models with a log link including imputed data were fit for hospitalization and mortality outcomes. All models were adjusted for sex, ethnicity and race, area deprivation index, rural urban residence, recent COVID-19 vaccination and COVID-19 antiviral treatment within 30 days of index COVID-19 diagnosis. Caption: The number of underlying conditions reflects how many of 29 underlying conditions were recorded within 3 years of COVID-19.

**Figure 9:**
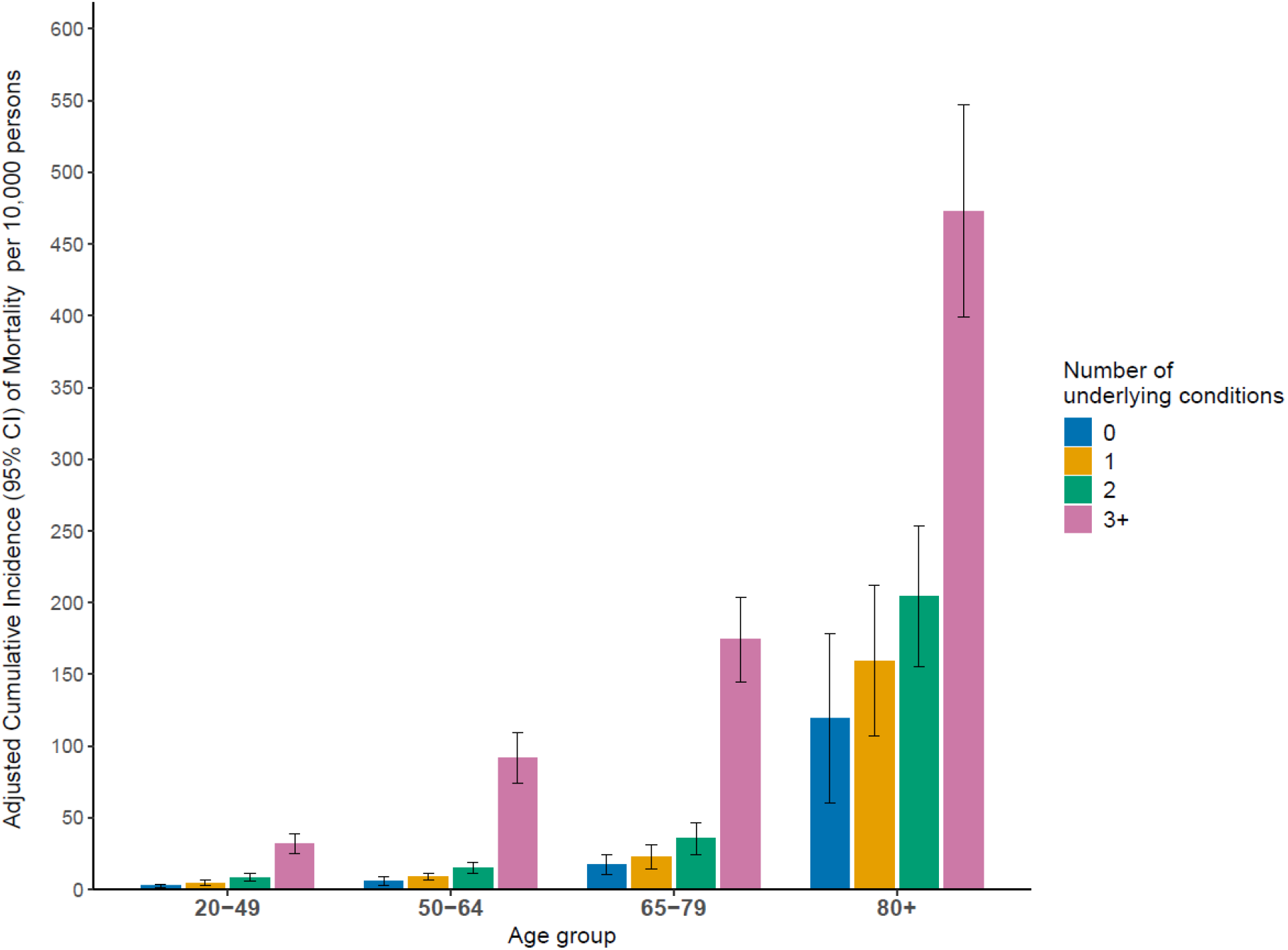
Adjusted cumulative incidence* and 95% Confidence Interval of mortality within 30 days among individuals aged ≥20 years with COVID-19 by age group and number of underlying conditions (UC), PCORnet, April 2022-March 2025 * Separate age-group-specific Poisson regression models with a log link including imputed data were fit for hospitalization and mortality outcomes. All models were adjusted for sex, ethnicity and race, area deprivation index, rural urban residence, recent COVID-19 vaccination and COVID-19 antiviral treatment within 30 days of index COVID-19 diagnosis. Caption: The number of underlying conditions reflects how many of 29 underlying conditions were recorded in the 3 years prior to COVID-19 index date.

**Figure 10:**
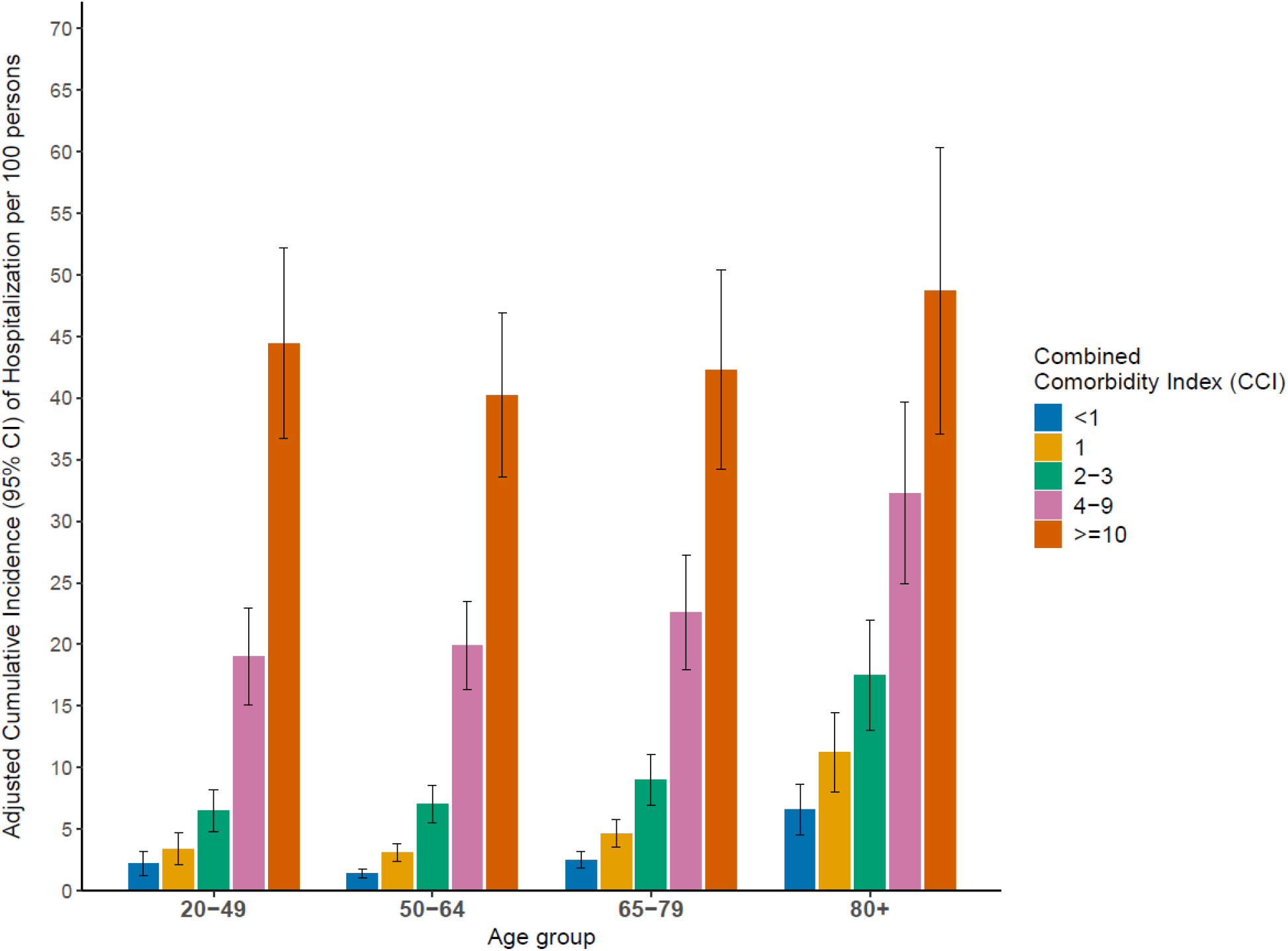
Adjusted cumulative incidence* and 95% Confidence Interval of hospitalization within 16 days among individuals aged ≥20 years with COVID-19 by age and Combined Comorbidity Index (CCI), PCORnet, April 2022-March 2025 * Separate age-group-specific Poisson regression models with a log link including imputed data were fit for hospitalization and mortality outcomes. All models were adjusted for sex, ethnicity and race, area deprivation index, rural urban residence, recent COVID-19 vaccination and COVID-19 antiviral treatment within 30 days of index COVID-19 diagnosis

**Figure 11:**
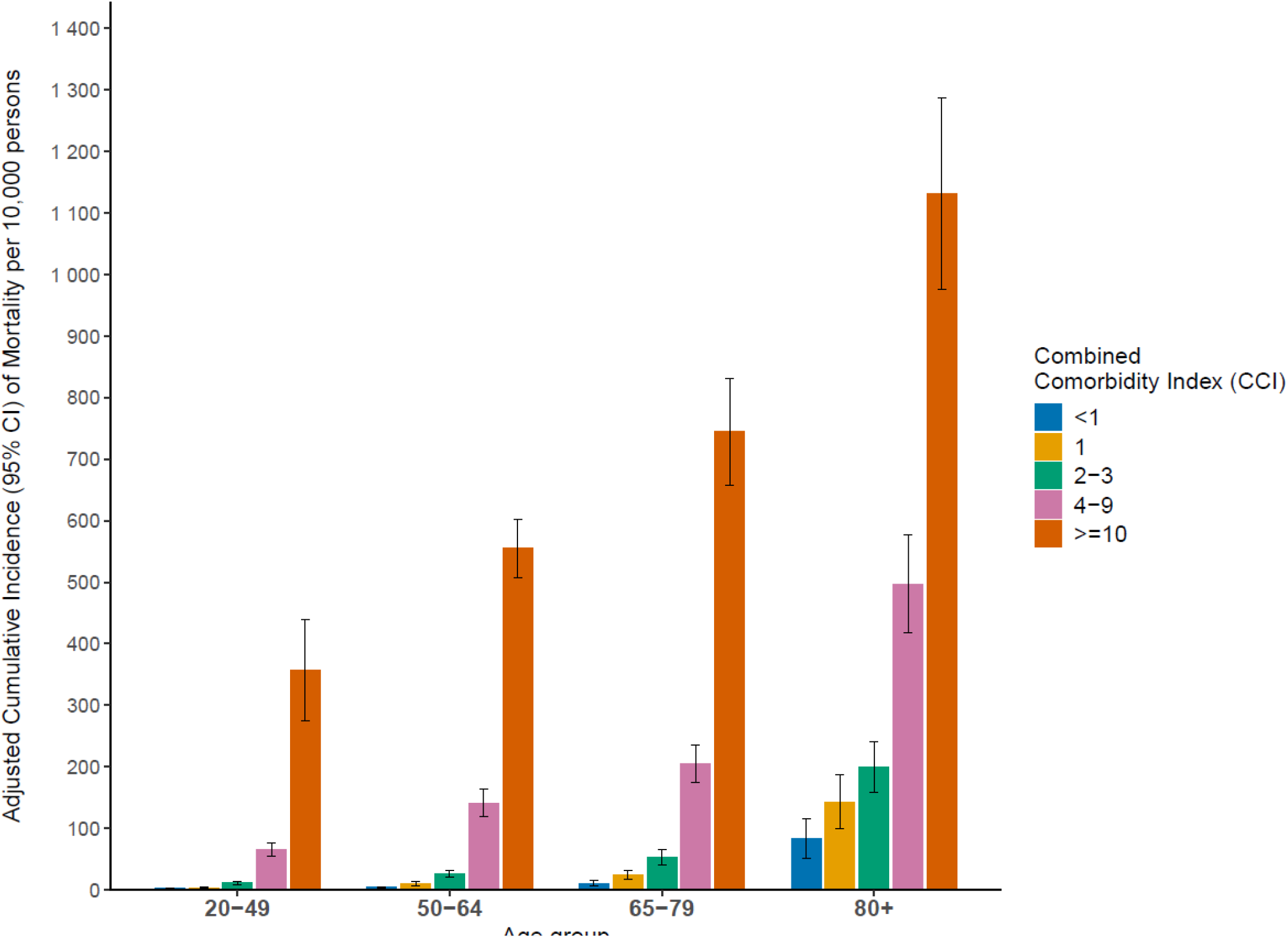
Adjusted cumulative incidence* and 95% Confidence Interval of mortality within 30 days among individuals aged ≥20 years with COVID-19 by age group and Combined Comorbidity Index (CCI), PCORnet, April 2022-March 2025. *Separate age-group-specific Poisson regression models with a log link including imputed data were fit for hospitalization and mortality outcomes. All models were adjusted for sex, ethnicity and race, area deprivation index, rural urban residence, recent COVID-19 vaccination and COVID-19 antiviral treatment within 30 days of index COVID-19 diagnosis.

### Adjusted cumulative incidence ratios

Adjusted cumulative incidence ratios (aCIR) indicated that within each age group, cumulative incidence of hospitalization and mortality increased with increasing number of underlying conditions (Figures 12–15, Table 3). However, as age group increased from the 20-49 year-olds to the ≥80 year-olds, the point estimate of the cumulative incidence of hospitalization generally declined and differences decreased with increasing number of UC or CCI increase. For example, among those with 1 (or 2 or 3 or more UCs), the aIR for hospitalization decreased among 20-49, 50-64, 65-79, and ≥80-year-olds. With one UC, the aIR was 2.5 (95% CI 1.5-4.1), 2.1 (1.4-3.3), 1.8 (1.1-2.9), 1.8 (1.0-3.2), and with 3 or more UCs: 12.36 (7.6-20.1), 10.8 (7.3-16.0), 8.3 (5.4-12.8), and 4.8 (2.8-8.2) among those 20-49, 50-64, 65-79, and ≥80 years old, respectively. Patterns were similar for mortality. (Table 3, Figure 12,13).

**Figure 12.**
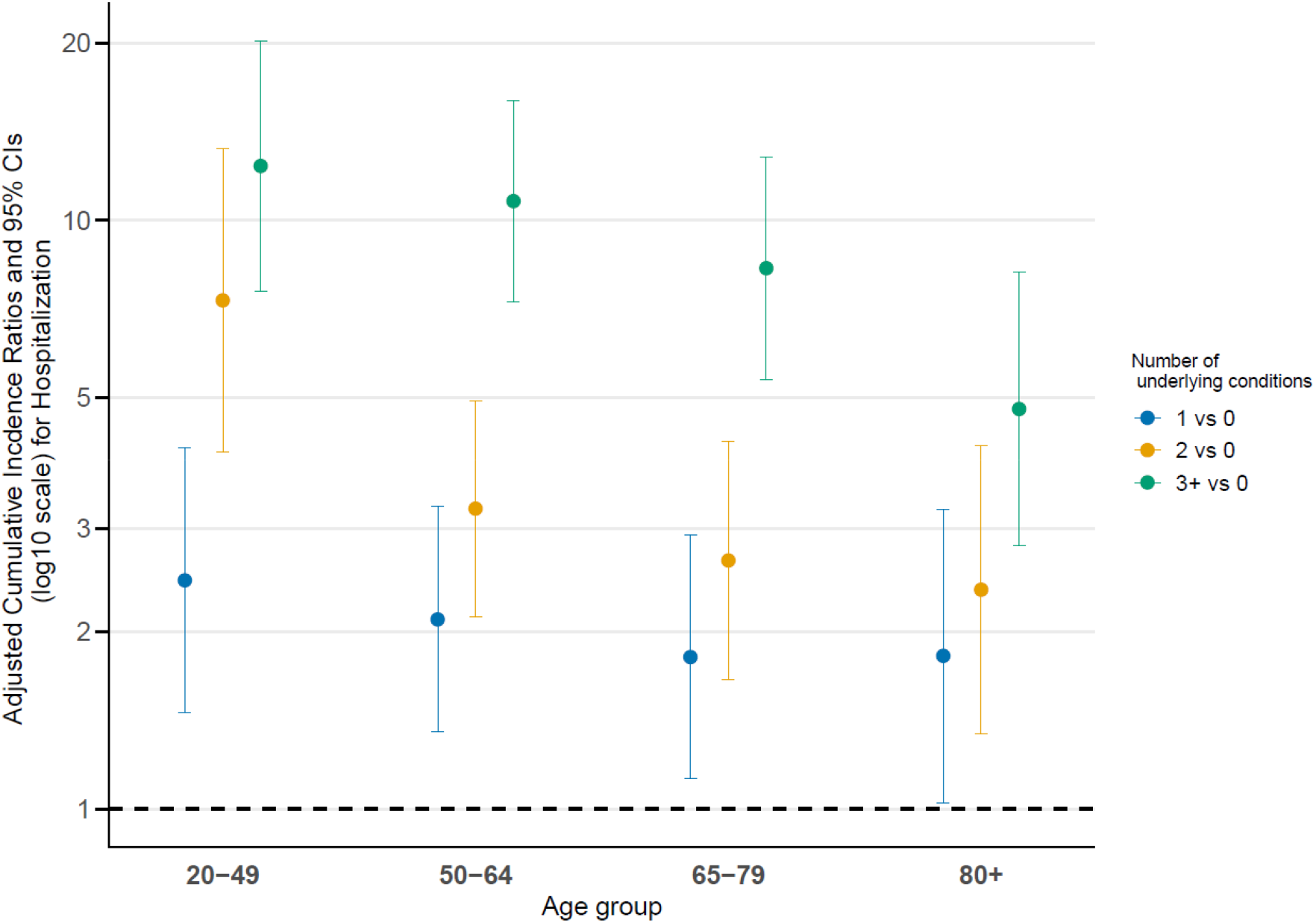
Adjusted cumulative incidence* ratios and 95% Confidence Interval of hospitalization within 16 days by age and number of underlying conditions among individuals aged ≥20 years with COVID-19, PCORnet, April 2022-March 2025. * Separate age-group-specific Poisson regression models with a log link including imputed data were fit for hospitalization and mortality outcomes. All models were adjusted for sex, ethnicity and race, area deprivation index, rural urban residence, recent COVID-19 vaccination and COVID-19 antiviral treatment within 30 days of index COVID-19 diagnosis.

**Figure 13.**
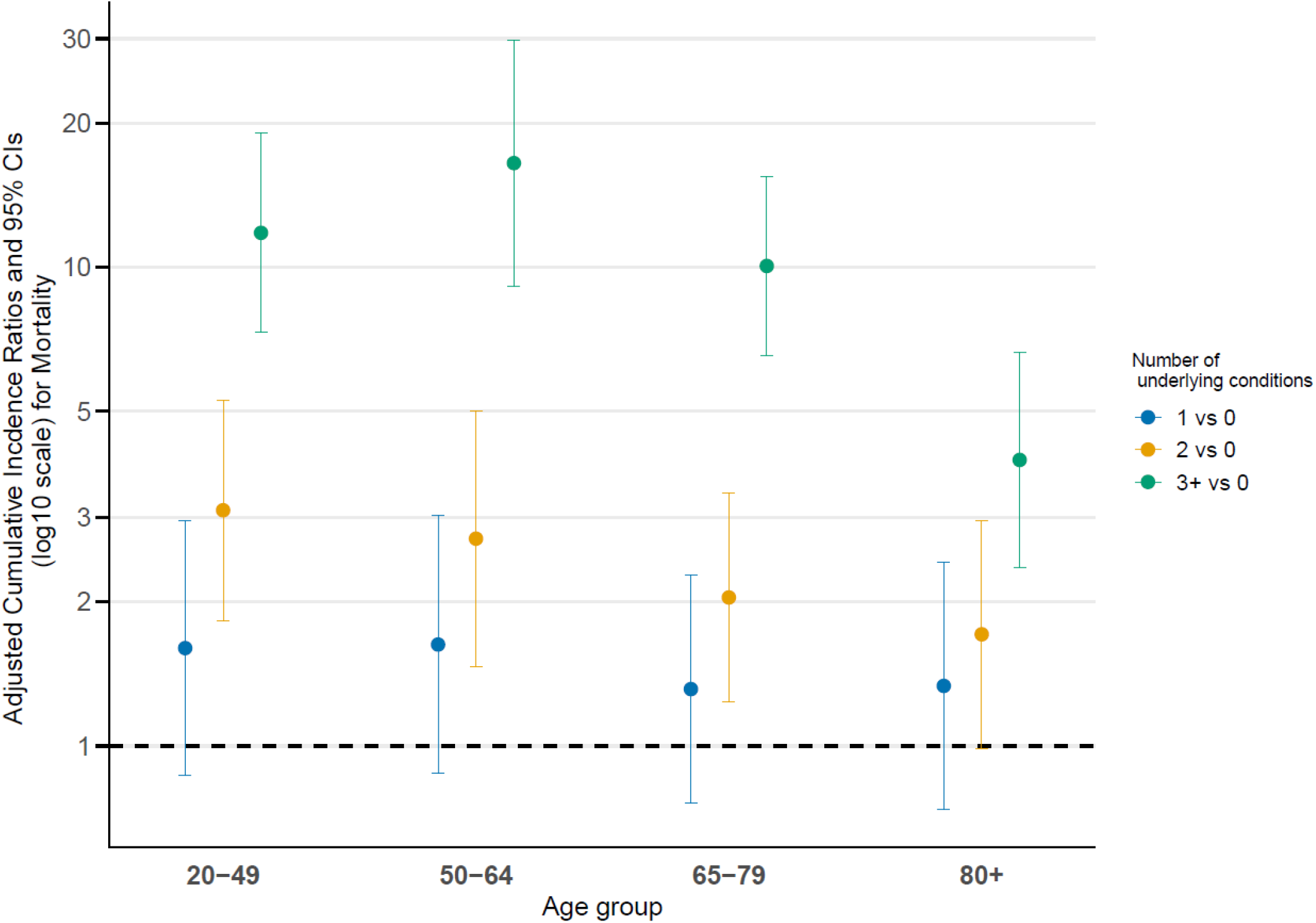
Adjusted cumulative incidence* ratios and 95% Confidence Interval of mortality within 30 days by age and number of underlying conditions among individuals aged ≥20 years with COVID-19, PCORnet April 2022-March 2025. * Separate age-group-specific Poisson regression models with a log link including imputed data were fit for hospitalization and mortality outcomes. All models were adjusted for sex, ethnicity and race, area deprivation index, rural urban residence, recent COVID-19 vaccination and COVID-19 antiviral treatment within 30 days of index COVID-19 diagnosis.

**Figure 14.**
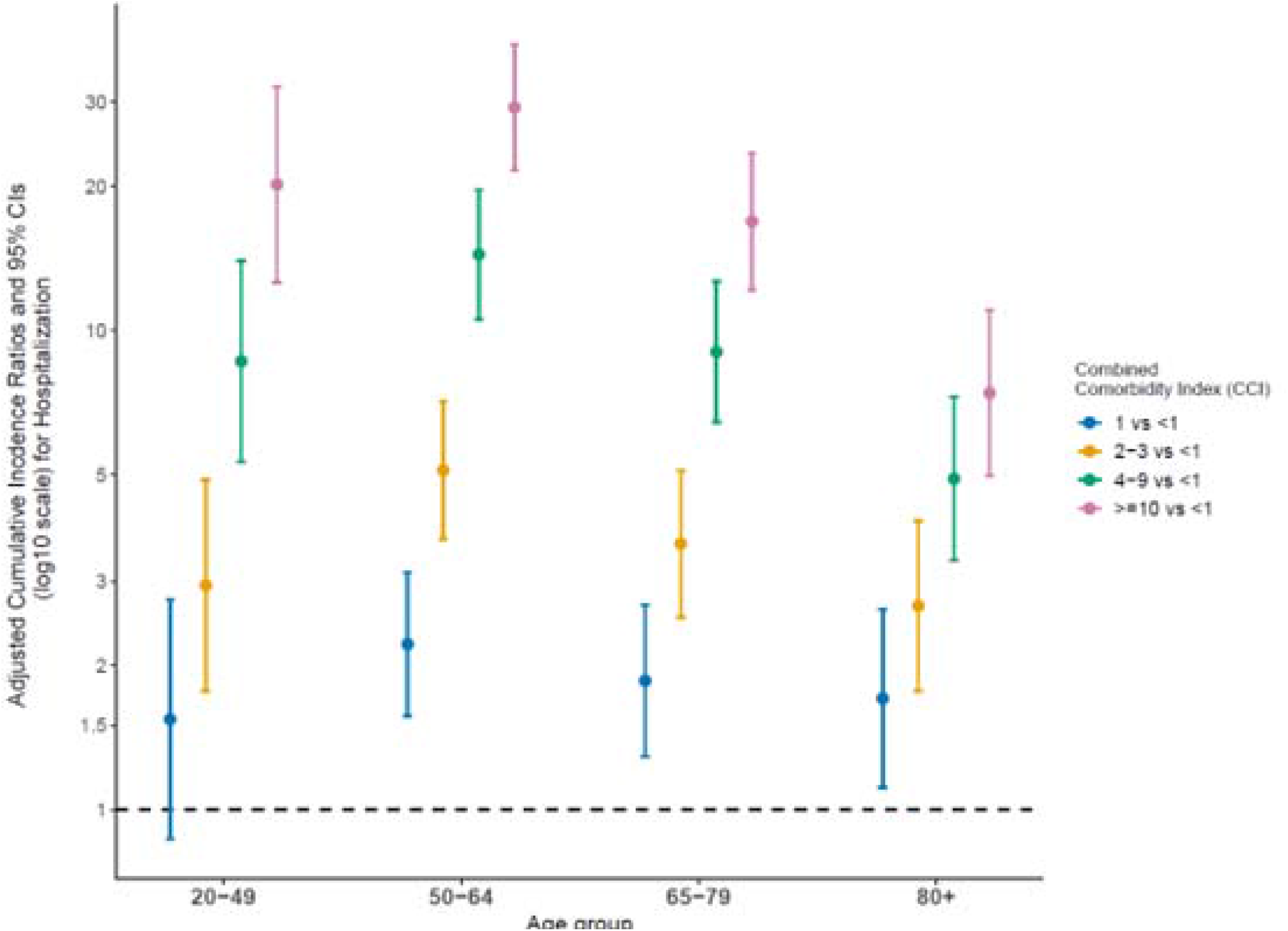
Adjusted cumulative incidence* ratios and 95% Confidence Interval of hospitalization within 16 days by age and by Combined Comorbidity Index (CCI) among individuals aged ≥20 years with COVID-19, PCORnet, April 2022-March 2025. * Separate age-group-specific Poisson regression models with a log link including imputed data were fit for hospitalization and mortality outcomes. All models were adjusted for sex, ethnicity and race, area deprivation index, rural urban residence, recent COVID-19 vaccination and COVID-19 antiviral treatment within 30 days of index COVID-19 diagnosis.

**Figure 15.**
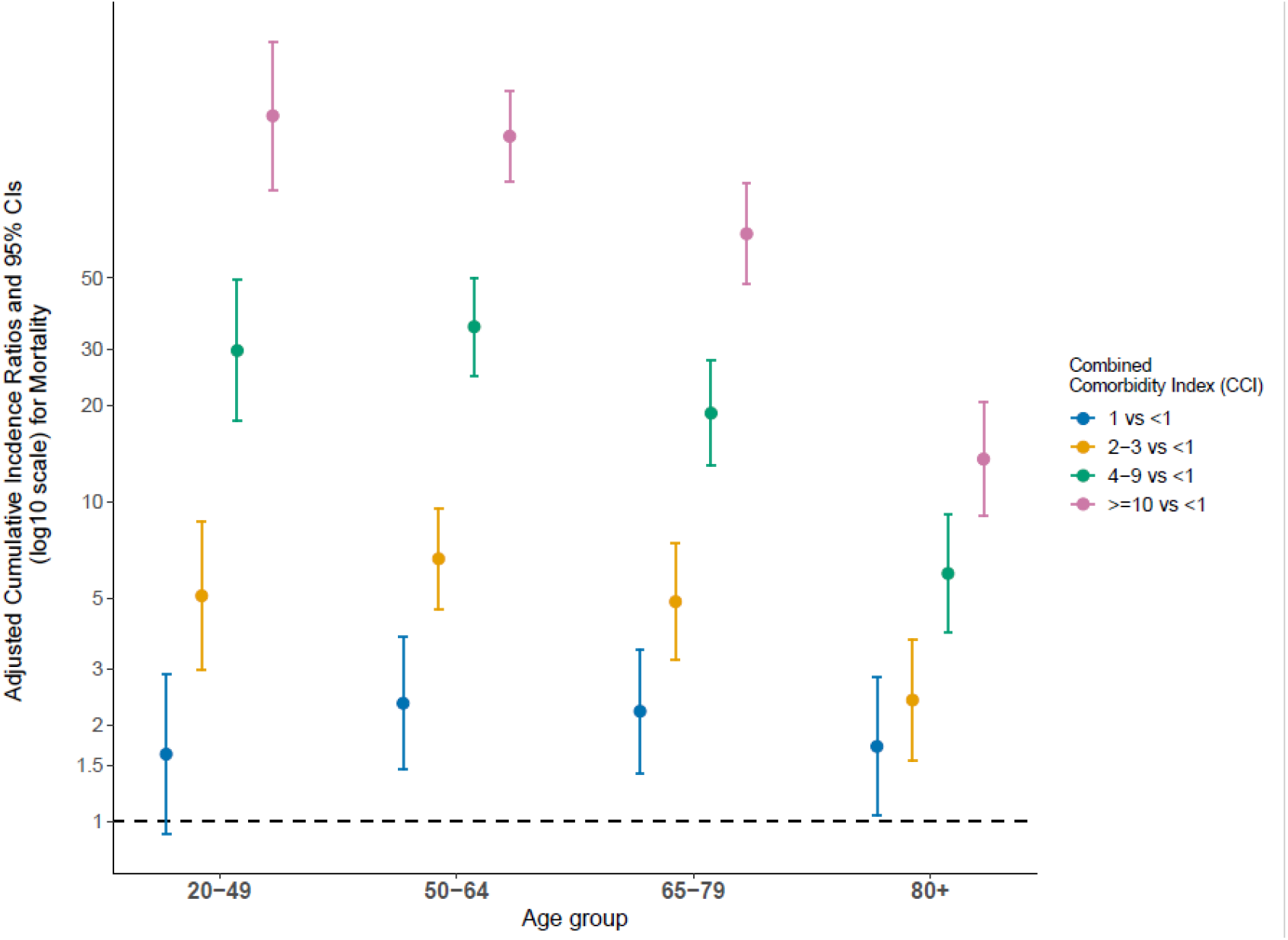
Adjusted cumulative incidence* ratios and 95% Confidence Interval of mortality within 30 days by age and Combined Comorbidity Index (CCI) among individuals aged ≥20 years with COVID-19, PCORnet, April 2022-March 2025. * Separate age-group-specific Poisson regression models with a log link including imputed data were fit for hospitalization and mortality outcomes. All models were adjusted for sex, ethnicity and race, area deprivation index, rural urban residence, recent COVID-19 vaccination and COVID-19 antiviral treatment within 30 days of index COVID-19 diagnosis.

**Table 3.**
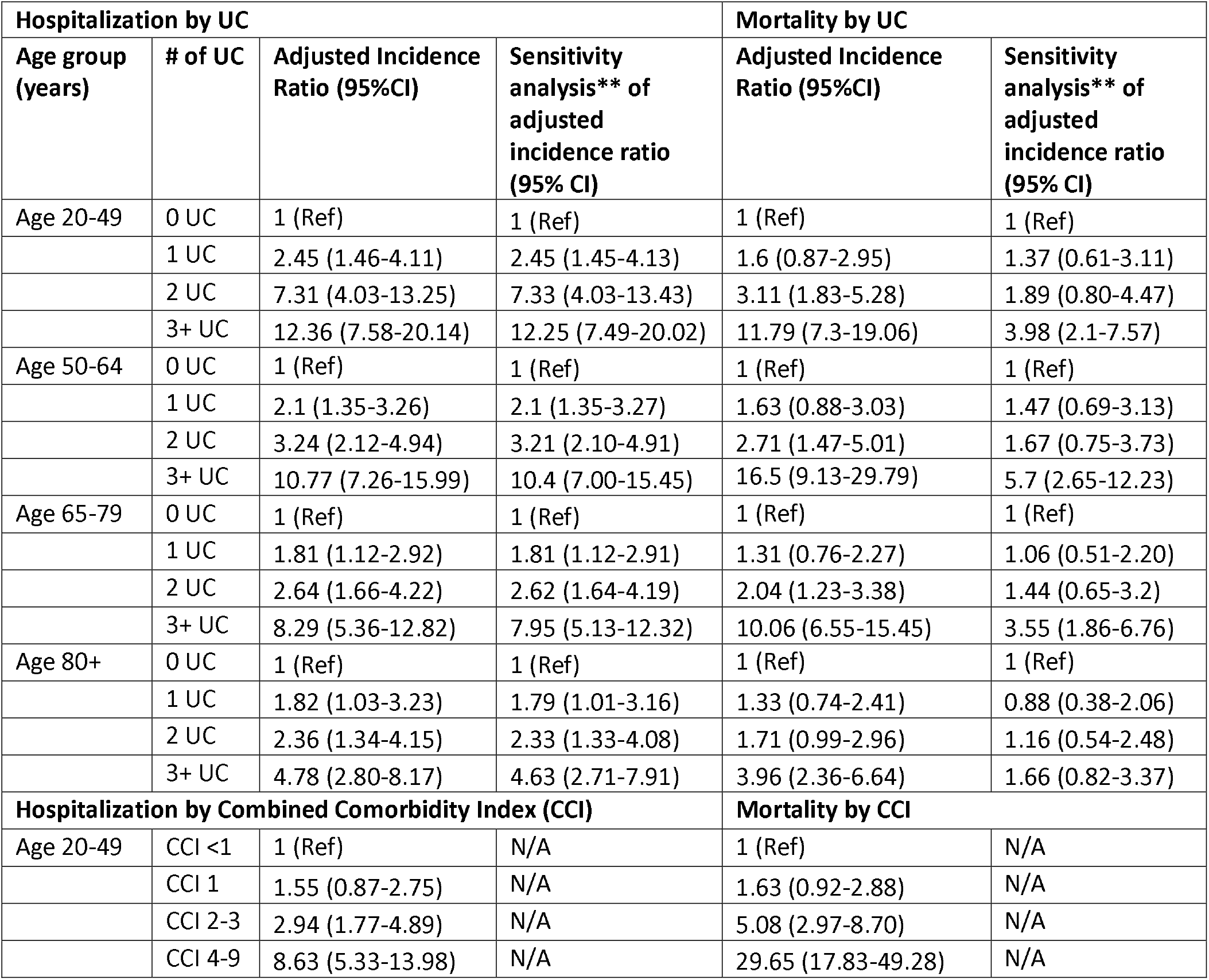
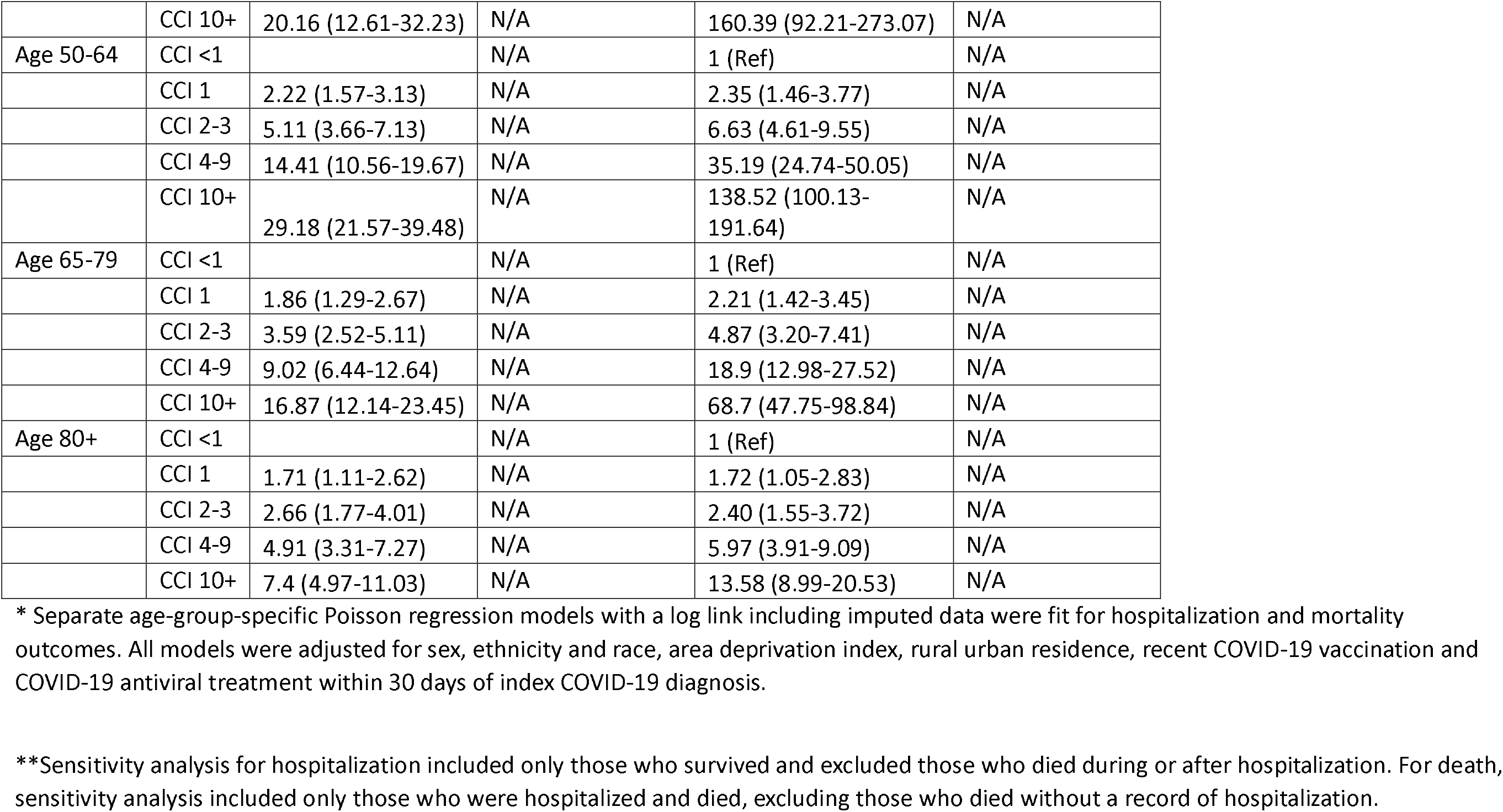
Adjusted cumulative incidence* ratios of hospitalization within 16 days and mortality within 30 days, 95% Confidence Interval, and sensitivity analysis** of hospitalization and mortality among individuals aged ≥20 years with COVID-19, by age group and number of underlying conditions (UC) and Combined Comorbidity Index (CCI), PCORnet, April 2022-March 2025.

Adjusted cumulative incidence ratios (aCIR) indicated that within each age group, cumulative incidence of hospitalization and mortality generally increased with increasing Combined Comorbidity Index. In the 20-49 and 50-64 year old age groups, as the CCI increased, the point estimate for the aCIRs for hospitalization and mortality increased; however, aCIR then declined in the 65-79 and ≥ 80 years age groups (Figures 14,15).

Sensitivity analyses showed that excluding people who had died during or after hospitalization from the hospitalization analyses did not appreciably change the aCIRs (Table 3). When excluding individuals with no hospitalization record from the analysis, aCIRs of mortality were lower and did not reach significance except for those with 3 or more UCs in each age group except those aged ≥80 year-olds (Table 3). A comparison of those who died during hospitalization to those who died without a record of hospitalization can be found in Supplemental Material F. The mortality group with no record of hospitalization had higher proportions of older, female, white individuals, higher proportions missing rural/urban, census region and ADI, higher proportions in Q1, lower number of UC, more with no recorded UC, lower proportion with any of the UC categories, and lower record of COVID-19 treatment. Vaccinations were similar.

## Discussion

Of the 1.6 million adult patients with COVID-19 diagnosis during 2022-2025 in this analysis, the cumulative incidence of hospitalization and mortality were 8.5% and 0.8%, respectively. There was a nearly 5-fold difference in aCI of hospitalization and over 30-fold difference in mortality from the youngest to the oldest age groups. While risk increased with increasing age, this analysis found that underlying conditions – and particularly having multiple conditions -- remain important risk factors in the incidence of severe disease for individuals aged younger than 65 years (21,30). Therefore, for individuals younger than 65 years of age, healthcare professionals may consider the contribution of co-occurring underlying conditions to the incidence of severe disease in their clinical decision-making. The higher incidence of hospitalization and mortality among the oldest individuals is similar to observations from the start of the COVID-19 pandemic.(21,30) Reports from 2020 showed a greater than 100-fold difference in incidence of hospitalization or mortality between individuals <50 years and those ≥50 years.(21,30)

Within each age group, aCI of hospitalization and mortality generally increased with increasing numbers of UCs. A similar pattern was evident when examining increases by CCI.In each age group, except for mortality among those ≥80-years, compared to those with no UC, those with 2 or more UCs had significantly increased incidence for hospitalization and mortality. The magnitude of the difference between 0 and 3 or more UC depended on the age group. For 20-49-year-olds, there was a 14-fold difference between none and 3 or more; there was a 5-fold difference for ≥80-year-olds.

We found that the aCI varied by UC category; for example, cardiovascular, hepatic, transplant, or neurologic UC categories had the highest aCIs for hospitalization and mortality. Individuals with hemiplegia, tuberculosis, COPD, a pulmonary circulatory disorder, or severe renal disease had higher hospitalization and mortality than other UC categories, andan adjusted incidence of hospitalization of 20% or higher.

The sensitivity analyses demonstrated that the hospitalization results were largely robust to excluding people who died during or after hospitalization. The sensitivity analysis for mortality, including only those who died without a record of hospitalization, showed that the aCIRs for the number of UCs were lower than those in the primary analysis. This may indicate that: a) some data were missing from those who died without a record of hospitalization, supported by Supplemental Material F, b) individuals with in-hospital death died despite having fewer UC, c) those excluded may have died from other causes.

The findings from this study are similar to other 2020-2021 reports on age- and UC-stratified risks for hospitalization and mortality among patients with COVID-19 (1,23,30–34). Some of these studies focused narrowly on specific conditions, such as cardiac disease(32), and others provided age-stratified risks for specific conditions (23,30–34). In 2020, Ge et al. examined the effect of underlying conditions and age on mortality in Ontario, Canada, and found that as age increased, the influence of higher numbers of UCs on mortality decreased(30) They also found that UCs had a larger effect on illness severity and mortality among individuals <50 years compared to those ≥50 years. Our analysis used similar UCs and UC categories as Ge et al. In our analysis, all of the UCs, except hypertension, that were found to be risk factors for hospitalization and mortality in Ge et al. in 2020 continued to have elevated cumulative incidence of hospitalization and mortality during 2022-2025, in particular the co-occurrence of UCs. In 2021, Molani et al. examined the risk of mortality among adults hospitalized with COVID-19 and found that for those aged 18-49 years, the UC risk factors for mortality included higher BMI, heart failure, and cardiomyopathy, while for those aged ≥50 years, the UC risk factors were higher BMI and dementia (33). Skarbinski et al., examined risk of severe COVID-19 during the late Delta (July 5—November 30, 2021) and early Omicron (December 18, 2021—January 7, 2022) periods, and reported that COPD, atherosclerotic heart disease, renal disease, cancer, rheumatologic disease, dementia, and obesity (BMI ≥30 kg/m^2^) remained significantly associated with hospitalization and mortality, with adjusted hazard ratios between 1.17 and 1.74, controlling for age(23). However, data were not stratified by age to examine specific risks by age group post-2021.

We found that the UCs that CDC currently lists as having strong evidence for severe COVID-19 disease early in the pandemic from systematic reviews or meta-analyses in 2020 and 2021 continue to play a role in COVID-19 severity.(22) Strengths of this analysis include: data from April 2022 until April 2025 when Omicron and Omicron -lineage sub-variants were circulating; the inclusion of established patients with a medical record with underlying conditions, vaccinations, and treatment; the large sample; and adjustment for several possible confounders. We were not able to assess the unique contribution of each UC or UC category to hospitalization and mortality, controlling for the other UCs, because of the common co-occurrence of conditions.

Several limitations apply to this analysis. During the study period, many patients with mild COVID-19 likely did not test for COVID-19, tested at home or outside the included settings and may not have presented for care at included settings, which may have biased the incidence upwards. (35, 36) Our study only captured those who received care for COVID-19 and had information about their infection documented in the EHR. As a result, our study likely overestimated the incidence of hospitalization and mortality due to COVID-19 and represents the incidence of hospitalization and mortality among those presenting for care. These analyses were based on available information contained in the EHRs of participating institutions. Individuals who also received care at other health care settings may be missing information. Therefore, some vaccination, treatment, UC, hospitalizations, and deaths may not have been recorded, particularly vaccinations, which are often administered in pharmacies.(38) The direction of the bias from missing information is unclear. We did not have information on the severity of UC, which may also differ by age and other UC, and may also affect incidence of hospitalization or mortality; the direction of the bias from missing UC severity is unclear. Missing health information could have led to misclassification of UC status. For example, if a UC had not been noted in the EHR in the 3 years prior to the COVID-19 diagnosis, an individual may have been mistakenly classified as having fewer UC. The effect of missing UC may have affected the estimates of aCIR downward (false lower risk); while the possible exclusions on the incidence of hospitalization and mortality are unclear, this limitation applies to other studies using EHR data. We did not adjust for age within each age stratum. We had no reason to conclude that misclassification differed by age; however, if it did, some of our inferences regarding the role of age and UC for severe COVID-19 disease outcomes could have been affected, the direction of the bias from missing UC severity is unclear.(39) Misclassification could also affect inferences where covariates such as COVID-19 vaccination and treatment were differentially captured by age or number of UC. Further, while we explored hospitalization and mortality following documented COVID-19, hospitalization or death may not have been caused by COVID-19, for example, a severe injury resulting in hospitalization or death. However, a 2023 analysis showed that among individuals ≥20 years who were hospitalized with a positive SARS-CoV-2 test, 78.1%-91.8% of those hospitalizations, varying by age, were determined to be COVID-19 related.(40) Hospitalizations and mortality caused by COVID-19 may also have happened after the 16-day or 30-day inclusion periods for each outcome, and out-of-hospital deaths within 30-days may not have been recorded; this may have biased the results for mortality downward. Generalizability is also limited because individuals in this study had access to care that facilitated COVID-19 testing and treatment and the measured incidences with UCs may not be applicable to those without similar healthcare access or care seeking behavior. Conditions that were managed may impart lower risk than unmanaged conditions.(37) However, the findings among those hospitalized are likely applicable since most people with severe COVID-19 during the analytic period were likely to be hospitalized. Lastly, trends in incidence of the outcomes during April 2022 through March 2025 could not be examined because encounter dates were removed.

In conclusion, among adults with COVID-19, in addition to age, cumulative incidence of hospitalization and mortality generally increased with increasing numbers of certain underlying conditions, namely cardiovascular, immunologic, hepatic, metabolic, oncologic, neurologic, pulmonary, renal, and a limited number of other conditions. Incidence increased substantially among those with 3 or more conditions, compared to having none. These results from EHRs from a large healthcare network highlight that older age and number of underlying conditions continue to be the major factors influencing incidence of severe COVID-19. This provides health care providers with valuable information for patients who may be at higher risk for hospitalization and death and who may benefit from assessment for recommended COVID-19 vaccinations and acute COVID-19 treatment.

## Data Availability

Data not publicly available as part of PCORnet privacy policy

## Acknowledgements

none

## Conflicts of interest

Michael Kappelman is a consultant for Eli Lilly, Takeda, and Roche Genentech and a shareholder in Johnson & Johnson. The remaining authors have no conflicts to declare.

Data not publicly available as part of PCORnet privacy policy.

## Authors’ contributions

EK: conceptualization, funding acquisition, investigation, methodology, project administration, supervision, validation, visualization, writing original, reviewing and editing

GD: formal analysis, investigation, software, validation, visualization, writing-review and editing

MS: conceptualization, data curation, investigation, visualization, writing original and review and editing

NG: data curation, formal analysis, investigation, software, validation, visualization, writing-review and editing

CD: data curation, project administration, writing-review and editing

SS: methodology, project administration, supervision, writing-review and editing

MBH: methodology, formal analysis, writing-review and editing

JD: resources, writing-review and editing

MK: Formal analysis, resources, writing-review and editing

DT: resources, writing-review and editing

ES: resources, writing-review and editing

KN: resources, writing-review and editing

TKB: funding acquisition, investigation, resources, writing-review and editing

DE: data curation, project administration, writing-review and editing

TC: resources, writing-review and editing PP: investigation, writing-review and editing

JB: conceptualization, investigation, methodology, project administration resources, writing-review and editing

## Footnotes

§ https://www.cdc.gov/nchs/icd/icd-10-cm/files.html

## Supplemental Materials

Supplemental Material A. LOINC codes and medications to treat COVID-19 used to identify COVID-19

Supplemental Material B. COVID-19 vaccination codes

Supplemental Material C. ICD-10 code list describing 29 underlying conditions

Supplemental Material D. Immunosuppressive medications

Supplemental Material E: STROBE checklist

Supplemental Material F: Comparison of individuals with COVID-19 who died within 30 days with or without record of hospitalization.

