## Supplemental Material A for "How age and underlying conditions affect incidence of hospitalization and death after documented COVID-19 diagnosis among individuals aged ≥20 years, a retrospective cohort study using PCORnet, April 2022 – March 2025"

| Code | Code type | Medication | Description |
|---|---|---|---|
| 2587906 | MA00 | Molupiravir | molnupiravir 200 MG Oral Capsule |
| 2587902 | MA00 | Molupiravir | molnupiravir 200 MG |
| 2587905 | MA00 | Molupiravir | molnupiravir Oral Capsule |
| 2587903 | MA00 | Molupiravir | molnupiravir Oral Product |
| 2587904 | MA00 | Molupiravir | molnupiravir Pill |
| 2587901 | MA00 | Molupiravir | molnupiravir |
| 6505507 | MA11 | Molupiravir | 200 mg molnupiravir |
| 6505506 | MA11 | Molupiravir | 200 mg molnupiravir |
| 2587906 | PR00 | Molupiravir | molnupiravir 200 MG Oral Capsule |
| 2587902 | PR00 | Molupiravir | molnupiravir 200 MG |
| 2587905 | PR00 | Molupiravir | molnupiravir Oral Capsule |
| 2587903 | PR00 | Molupiravir | molnupiravir Oral Product |
| 2587904 | PR00 | Molupiravir | molnupiravir Pill |
| 2587901 | PR00 | Molupiravir | molnupiravir |
| 2557245 | PR00 | Monocolonal antibodies | 10 ML casirivimab 60 MG/ML / imdevimab 60 MG/ML Injection |
| 2465246 | PR00 | Monocolonal antibodies | 11.1 ML casirivimab 120 MG/ML Injection |
| 2465248 | PR00 | Monocolonal antibodies | 2.5 ML casirivimab 120 MG/ML Injection |
| 2556915 | PR00 | Monocolonal antibodies | casirivimab / imdevimab Injectable Product |
| 2557241 | PR00 | Monocolonal antibodies | casirivimab / imdevimab Injection [Regen-Cov] |
| 2557237 | PR00 | Monocolonal antibodies | casirivimab / imdevimab Injection |
| 2557234 | PR00 | Monocolonal antibodies | casirivimab / imdevimab |
| 2465243 | PR00 | Monocolonal antibodies | casirivimab 120 MG/ML |
| 2557238 | PR00 | Monocolonal antibodies | casirivimab 60 MG/ML / imdevimab 60 MG/ML [Regen-Cov] |
| 2557235 | PR00 | Monocolonal antibodies | casirivimab 60 MG/ML |
| 2465244 | PR00 | Monocolonal antibodies | casirivimab Injectable Product |
| 2465245 | PR00 | Monocolonal antibodies | casirivimab Injection |
| 2465242 | PR00 | Monocolonal antibodies | casirivimab |
| 2557244 | PR00 | Monocolonal antibodies | REGEN-COV 600 MG / 600 MG per 10 ML Injection (EUA) |
| 2557236 | PR00 | Monocolonal antibodies | Regen-Cov Injectable Product |
| 2479150 | PR00 | Monocolonal antibodies | REGEN-COV, 2-cartons (casirivimab 1, imdevimab 1) (EUA) |
| 2479159 | PR00 | Monocolonal antibodies | REGEN-COV, 5-cartons (casirivimab 1, imdevimab 4) (EUA) |
| 2479156 | PR00 | Monocolonal antibodies | REGEN-COV, 5-cartons (casirivimab 4, imdevimab 1) (EUA) |
| 2479154 | PR00 | Monocolonal antibodies | REGEN-COV, 8-cartons (casirivimab 4, imdevimab 4) (EUA) |
| 2479149 | PR00 | Monocolonal antibodies | {1 (11.1 ML casirivimab 120 MG/ML Injection) / 1 (11.1 ML imdevimab 120 MG/ML Injection) } Pack |
| 2479158 | PR00 | Monocolonal antibodies | {1 (11.1 ML casirivimab 120 MG/ML Injection) / 4 (2.5 ML imdevimab 120 MG/ML Injection) } Pack |
| 2479155 | PR00 | Monocolonal antibodies | {1 (11.1 ML imdevimab 120 MG/ML Injection) / 4 (2.5 ML casirivimab 120 MG/ML Injection) } Pack |
| 2571851 | PR00 | Monocolonal antibodies | {1 (2.5 ML casirivimab 120 MG/ML Injection) / 1 (2.5 ML imdevimab 120 MG/ML Injection) } Pack |

| | | | |
|---|---|---|---|
| 2479153 | PR00 | Monocolonal antibodies | {4 (2.5 ML casirivimab 120 MG/ML Injection) / 4 (2.5 ML imdevimab 120 MG/ML Injection) } Pack |
| 2465253 | PR00 | Monocolonal antibodies | 11.1 ML imdevimab 120 MG/ML Injection |
| 2465255 | PR00 | Monocolonal antibodies | 2.5 ML imdevimab 120 MG/ML Injection |
| 2465250 | PR00 | Monocolonal antibodies | imdevimab 120 MG/ML |
| 2557239 | PR00 | Monocolonal antibodies | imdevimab 60 MG/ML |
| 2465251 | PR00 | Monocolonal antibodies | imdevimab Injectable Product |
| 2465252 | PR00 | Monocolonal antibodies | imdevimab Injection |
| 2465249 | PR00 | Monocolonal antibodies | imdevimab |
| 2463118 | PR00 | Monocolonal antibodies | 20 ML bamlanivimab 35 MG/ML Injection |
| 2463115 | PR00 | Monocolonal antibodies | bamlanivimab 35 MG/ML |
| 2463116 | PR00 | Monocolonal antibodies | bamlanivimab Injectable Product |
| 2463117 | PR00 | Monocolonal antibodies | bamlanivimab Injection |
| 2463114 | PR00 | Monocolonal antibodies | bamlanivimab |
| 2477902 | PR00 | Monocolonal antibodies | 20 ML etesevimab 35 MG/ML Injection |
| 2477899 | PR00 | Monocolonal antibodies | etesevimab 35 MG/ML |
| 2477900 | PR00 | Monocolonal antibodies | etesevimab Injectable Product |
| 2477901 | PR00 | Monocolonal antibodies | etesevimab Injection |
| 2477854 | PR00 | Monocolonal antibodies | etesevimab |
| 2550903 | PR00 | Monocolonal antibodies | 8 ML sotrovimab 62.5 MG/ML Injection |
| 2550900 | PR00 | Monocolonal antibodies | sotrovimab 62.5 MG/ML |
| 2550901 | PR00 | Monocolonal antibodies | sotrovimab Injectable Product |
| 2550902 | PR00 | Monocolonal antibodies | sotrovimab Injection |
| 2550731 | PR00 | Monocolonal antibodies | sotrovimab |
| 2592364 | PR00 | Monocolonal antibodies | 2 ML bebtelovimab 87.5 MG/ML Injection |
| 2592361 | PR00 | Monocolonal antibodies | bebtelovimab 87.5 MG/ML |
| 2592362 | PR00 | Monocolonal antibodies | bebtelovimab Injectable Product |
| 2592363 | PR00 | Monocolonal antibodies | bebtelovimab Injection |
| 2592360 | PR00 | Monocolonal antibodies | bebtelovimab |
| 2791001 | MA11 | Monocolonal antibodies | LY-COV555 (bamlanivimab) |
| 2463118 | MA00 | Monocolonal antibodies | 20 ML bamlanivimab 35 MG/ML Injection |
| 2463115 | MA00 | Monocolonal antibodies | bamlanivimab 35 MG/ML |
| 2463116 | MA00 | Monocolonal antibodies | bamlanivimab Injectable Product |
| 2463117 | MA00 | Monocolonal antibodies | bamlanivimab Injection |
| 2463114 | MA00 | Monocolonal antibodies | bamlanivimab |
| 2477902 | MA00 | Monocolonal antibodies | 20 ML etesevimab 35 MG/ML Injection |
| 2477899 | MA00 | Monocolonal antibodies | etesevimab 35 MG/ML |
| 2477900 | MA00 | Monocolonal antibodies | etesevimab Injectable Product |
| 2477901 | MA00 | Monocolonal antibodies | etesevimab Injection |
| 2477854 | MA00 | Monocolonal antibodies | etesevimab |
| 2795001 | MA11 | Monocolonal antibodies | Etesevimab 700 mg/20 mL (35 mg/mL) |
| 61755002401 | MA11 | Monocolonal antibodies | casirivimab, 1332 mg/11.1 mL (120 mg/mL) |
| 61755002601 | MA11 | Monocolonal antibodies | casirivimab, 300 mg/2.5 mL (120 mg/mL) |
| 61755002501 | MA11 | Monocolonal antibodies | imdevimab, 1332 mg/11.1 mL (120 mg/mL) |
| 61755002701 | MA11 | Monocolonal antibodies | imdevimab, 300 mg/2.5 mL (120 mg/mL) |

| | | | |
|---|---|---|---|
| 61755002400 | MA11 | Monocolonal antibodies | casirivimab |
| 61755002600 | MA11 | Monocolonal antibodies | casirivimab |
| 61755003502 | MA11 | Monocolonal antibodies | casirivimab/imdevimab |
| 61755003608 | MA11 | Monocolonal antibodies | casirivimab/imdevimab |
| 61755003705 | MA11 | Monocolonal antibodies | casirivimab/imdevimab |
| 61755003805 | MA11 | Monocolonal antibodies | casirivimab/imdevimab |
| 61755003900 | MA11 | Monocolonal antibodies | casirivimab/imdevimab |
| 61755003901 | MA11 | Monocolonal antibodies | casirivimab/imdevimab |
| 61755002500 | MA11 | Monocolonal antibodies | imdevimab |
| 61755002700 | MA11 | Monocolonal antibodies | imdevimab |
| 173090186 | MA11 | Monocolonal antibodies | sotrovimab |
| 2550903 | MA00 | Monocolonal antibodies | 8 ML sotrovimab 62.5 MG/ML Injection |
| 2550900 | MA00 | Monocolonal antibodies | sotrovimab 62.5 MG/ML |
| 2550901 | MA00 | Monocolonal antibodies | sotrovimab Injectable Product |
| 2550902 | MA00 | Monocolonal antibodies | sotrovimab Injection |
| 2550731 | MA00 | Monocolonal antibodies | sotrovimab |
| 2592364 | MA00 | Monocolonal antibodies | 2 ML bebtelovimab 87.5 MG/ML Injection |
| 2592361 | MA00 | Monocolonal antibodies | bebtelovimab 87.5 MG/ML |
| 2592362 | MA00 | Monocolonal antibodies | bebtelovimab Injectable Product |
| 2592363 | MA00 | Monocolonal antibodies | bebtelovimab Injection |
| 2592360 | MA00 | Monocolonal antibodies | bebtelovimab |
| 2557245 | MA00 | Monocolonal antibodies | 10 ML casirivimab 60 MG/ML / imdevimab 60 MG/ML Injection |
| 2465246 | MA00 | Monocolonal antibodies | 11.1 ML casirivimab 120 MG/ML Injection |
| 2465248 | MA00 | Monocolonal antibodies | 2.5 ML casirivimab 120 MG/ML Injection |
| 2556915 | MA00 | Monocolonal antibodies | casirivimab / imdevimab Injectable Product |
| 2557241 | MA00 | Monocolonal antibodies | casirivimab / imdevimab Injection [Regen-Cov] |
| 2557237 | MA00 | Monocolonal antibodies | casirivimab / imdevimab Injection |
| 2557234 | MA00 | Monocolonal antibodies | casirivimab / imdevimab |
| 2465243 | MA00 | Monocolonal antibodies | casirivimab 120 MG/ML |
| 2557238 | MA00 | Monocolonal antibodies | casirivimab 60 MG/ML / imdevimab 60 MG/ML [Regen-Cov] |
| 2557235 | MA00 | Monocolonal antibodies | casirivimab 60 MG/ML |
| 2465244 | MA00 | Monocolonal antibodies | casirivimab Injectable Product |
| 2465245 | MA00 | Monocolonal antibodies | casirivimab Injection |
| 2465242 | MA00 | Monocolonal antibodies | casirivimab |
| 2557244 | MA00 | Monocolonal antibodies | REGEN-COV 600 MG / 600 MG per 10 ML Injection (EUA) |
| 2557236 | MA00 | Monocolonal antibodies | Regen-Cov Injectable Product |
| 2479150 | MA00 | Monocolonal antibodies | REGEN-COV, 2-cartons (casirivimab 1, imdevimab 1) (EUA) |
| 2479159 | MA00 | Monocolonal antibodies | REGEN-COV, 5-cartons (casirivimab 1, imdevimab 4) (EUA) |
| 2479156 | MA00 | Monocolonal antibodies | REGEN-COV, 5-cartons (casirivimab 4, imdevimab 1) (EUA) |
| 2479154 | MA00 | Monocolonal antibodies | REGEN-COV, 8-cartons (casirivimab 4, imdevimab 4) (EUA) |
| 2479149 | MA00 | Monocolonal antibodies | {1 (11.1 ML casirivimab 120 MG/ML Injection) / 1 (11.1 ML imdevimab 120 MG/ML Injection) } Pack |
| 2479158 | MA00 | Monocolonal antibodies | {1 (11.1 ML casirivimab 120 MG/ML Injection) / 4 (2.5 ML imdevimab 120 MG/ML Injection) } Pack |

| | | | |
|---|---|---|---|
| 2479155 | MA00 | Monocolonal antibodies | {1 (11.1 ML imdevimab 120 MG/ML Injection) / 4 (2.5 ML casirivimab 120 MG/ML Injection) } Pack |
| 2571851 | MA00 | Monocolonal antibodies | {1 (2.5 ML casirivimab 120 MG/ML Injection) / 1 (2.5 ML imdevimab 120 MG/ML Injection) } Pack |
| 2479153 | MA00 | Monocolonal antibodies | {4 (2.5 ML casirivimab 120 MG/ML Injection) / 4 (2.5 ML imdevimab 120 MG/ML Injection) } Pack |
| 2465253 | MA00 | Monocolonal antibodies | 11.1 ML imdevimab 120 MG/ML Injection |
| 2465255 | MA00 | Monocolonal antibodies | 2.5 ML imdevimab 120 MG/ML Injection |
| 2465250 | MA00 | Monocolonal antibodies | imdevimab 120 MG/ML |
| 2557239 | MA00 | Monocolonal antibodies | imdevimab 60 MG/ML |
| 2465251 | MA00 | Monocolonal antibodies | imdevimab Injectable Product |
| 2465252 | MA00 | Monocolonal antibodies | imdevimab Injection |
| 2465249 | MA00 | Monocolonal antibodies | imdevimab |
| XW033H6 | PX10 | Monocolonal antibodies | Introduction of other new technology monoclonal antibody into peripheral vein, percutaneous approach, new technology group 6 |
| XW043H6 | PX10 | Monocolonal antibodies | Introduction of other new technology monoclonal antibody into central vein, percutaneous approach, new technology group 6 |
| Q0239 | PXCH | Monocolonal antibodies | Injection, bamlanivimab-xxxx, 700 mg |
| M0239 | PXCH | Monocolonal antibodies | Intravenous infusion, bamlanivimab, includes infusion and post administration monitoring |
| XW033F6 | PX10 | Monocolonal antibodies | Introduction of Bamlanivimab Monoclonal Antibody into Peripheral Vein, Percutaneous Approach, New Technology Group 6 |
| XW043F6 | PX10 | Monocolonal antibodies | Introduction of Bamlanivimab Monoclonal Antibody into Central Vein, Percutaneous Approach, New Technology Group 6 |
| Q0245 | PXCH | Monocolonal antibodies | Injection, bamlanivimab and etesevimab, 2100 mg |
| M0245 | PXCH | Monocolonal antibodies | intravenous infusion, bamlanivimab and etesevimab, includes infusion and post administration monitoring |
| Q0243 | PXCH | Monocolonal antibodies | Injection, casirivimab and imdevimab, 2400 mg |
| M0243 | PXCH | Monocolonal antibodies | intravenous infusion, casirivimab and imdevimab includes infusion and post administration monitoring |
| XW033G6 | PX10 | Monocolonal antibodies | Introduction of REGN-COV2 monoclonal antibody into peripheral vein, percutaneous approach, new technology group 6 |
| XW043G6 | PX10 | Monocolonal antibodies | Introduction of REGN-COV2 monoclonal antibody into central vein, percutaneous approach, new technology group 6 |

| | | | |
|---|---|---|---|
| M0248 | PXCH | Monocolonal antibodies | Intravenous infusion, sotrovimab, includes infusion and post administration monitoring in the home or residence; this includes a beneficiary's home that has been made provider-based to the hospital during the COVID-19 public health emergency |
| M0247 | PXCH | Monocolonal antibodies | Intravenous infusion, sotrovimab, includes infusion and post administration monitoring |
| Q0247 | PXCH | Monocolonal antibodies | Injection, sotrovimab, 500 mg |
| Q0222 | PXCH | Monocolonal antibodies | Injection, bebtelovimab, 175 mg |
| M0222 | PXCH | Monocolonal antibodies | Intravenous injection, bebtelovimab, includes injection and post administration monitoring |
| M0223 | PXCH | Monocolonal antibodies | Intravenous injection, bebtelovimab, includes injection and post administration monitoring in the home or residence; this includes a beneficiary’s home that has been made provider-based to the hospital during the covid-19 public health emergency |
| M0246 | PXCH | Monocolonal antibodies | Intravenous infusion, bamlanivimab and etesevimab, includes infusion and post administration monitoring in the home or residence; this includes a beneficiary’s home that has been made provider-based to the hospital during the COVID-19 public health emergency |
| M0241 | PXCH | Monocolonal antibodies | Intravenous infusion or subcutaneous injection, casirivimab and imdevimab, includes infusion or injection, and post administration monitoring in the home or residence. This includes a beneficiary's home that has been made provider-based to the hospital during the covid-19 public health emergency, subsequent repeat doses |
| M0244 | PXCH | Monocolonal antibodies | Intravenous infusion or subcutaneous injection, casirivimab and imdevimab, includes infusion or injection and post administration monitoring in the home or residence; this includes a beneficiary’s home that has been made provider-based to the hospital during the COVID-19 public health emergency |
| 2587897 | MA00 | PAXLOVID | nirmatrelvir 150 MG Oral Tablet |
| 2587893 | MA00 | PAXLOVID | nirmatrelvir 150 MG |
| 2587894 | MA00 | PAXLOVID | nirmatrelvir Oral Product |
| 2587896 | MA00 | PAXLOVID | nirmatrelvir Oral Tablet |
| 2587895 | MA00 | PAXLOVID | nirmatrelvir Pill |
| 2587892 | MA00 | PAXLOVID | nirmatrelvir |
| 2587899 | MA00 | PAXLOVID | Paxlovid 5-Day (EUA) |
| 2587898 | MA00 | PAXLOVID | {20 (nirmatrelvir 150 MG Oral Tablet) / 10 (ritonavir 100 MG Oral Tablet) } Pack |

| | | | |
|---|---|---|---|
| 69108530 | MA11 | PAXLOVID | nirmatrelvir/ritonavir 4 day blister card |
| 69108506 | MA11 | PAXLOVID | nirmatrelvir/ritonavir daily blister card |
| 2587897 | PR00 | PAXLOVID | nirmatrelvir 150 MG Oral Tablet |
| 2587893 | PR00 | PAXLOVID | nirmatrelvir 150 MG |
| 2587894 | PR00 | PAXLOVID | nirmatrelvir Oral Product |
| 2587896 | PR00 | PAXLOVID | nirmatrelvir Oral Tablet |
| 2587895 | PR00 | PAXLOVID | nirmatrelvir Pill |
| 2587892 | PR00 | PAXLOVID | nirmatrelvir |
| 2587899 | PR00 | PAXLOVID | Paxlovid 5-Day (EUA) |
| 2587898 | PR00 | PAXLOVID | {20 (nirmatrelvir 150 MG Oral Tablet) / 10 (ritonavir 100 MG Oral Tablet) } Pack |
| 69034506 | MA11 | PAXLOVID | nirmatrelvir/ritonavir |
| 69034530 | MA11 | PAXLOVID | nirmatrelvir/ritonavir |
| 69110104 | MA11 | PAXLOVID | nirmatrelvir/ritonavir |
| 69110120 | MA11 | PAXLOVID | nirmatrelvir/ritonavir |
| 61958999899 | MA11 | REMDESIVIR | REMDESIVIR INJ 100MG |
| 61958999999 | MA11 | REMDESIVIR | REMDESIVIR INJ 150MG |
| 61958290101 | MA11 | REMDESIVIR | SCD: remdesivir 100 MG Injection |
| 61958290201 | MA11 | REMDESIVIR | SCD: 20 ML remdesivir 5 MG/ML Injection |
| 61958290102 | MA11 | REMDESIVIR | SBD: remdesivir 100 MG Injection [Veklury] |
| 61958290202 | MA11 | REMDESIVIR | SBD: 20 ML remdesivir 5 MG/ML Injection [Veklury] |
| XW033E5 | PX10 | REMDESIVIR | Introduction of Remdesivir Anti-infective into Peripheral Vein, Percutaneous Approach, New Technology Group 5 |
| XW043E5 | PX10 | REMDESIVIR | Introduction of Remdesivir Anti-infective into Central Vein, Percutaneous Approach, New Technology Group 5 |
| 2367758 | PR00 | REMDESIVIR | 20 ML remdesivir 5 MG/ML Injection |
| 2284960 | PR00 | REMDESIVIR | remdesivir 100 MG Injection |
| 2395504 | PR00 | REMDESIVIR | 20 ML Veklury 5 MG/ML Injection |
| 2395499 | PR00 | REMDESIVIR | remdesivir 100 MG [Veklury] |
| 2284957 | PR00 | REMDESIVIR | remdesivir 100 MG |
| 2395503 | PR00 | REMDESIVIR | remdesivir 5 MG/ML Injection [Veklury] |
| 2367757 | PR00 | REMDESIVIR | remdesivir 5 MG/ML |
| 2284958 | PR00 | REMDESIVIR | remdesivir Injectable Product |
| 2395500 | PR00 | REMDESIVIR | remdesivir Injection [Veklury] |
| 2284959 | PR00 | REMDESIVIR | remdesivir Injection |
| 2284718 | PR00 | REMDESIVIR | remdesivir |
| 2395502 | PR00 | REMDESIVIR | Veklury 100 MG Injection |
| 2395501 | PR00 | REMDESIVIR | Veklury Injectable Product |
| 2395498 | PR00 | REMDESIVIR | Veklury |
| 2367758 | MA00 | REMDESIVIR | 20 ML remdesivir 5 MG/ML Injection |
| 2284960 | MA00 | REMDESIVIR | remdesivir 100 MG Injection |
| 2395504 | MA00 | REMDESIVIR | 20 ML Veklury 5 MG/ML Injection |
| 2395499 | MA00 | REMDESIVIR | remdesivir 100 MG [Veklury] |
| 2284957 | MA00 | REMDESIVIR | remdesivir 100 MG |
| 2395503 | MA00 | REMDESIVIR | remdesivir 5 MG/ML Injection [Veklury] |
| 2367757 | MA00 | REMDESIVIR | remdesivir 5 MG/ML |
| 2284958 | MA00 | REMDESIVIR | remdesivir Injectable Product |
| 2395500 | MA00 | REMDESIVIR | remdesivir Injection [Veklury] |
| 2284959 | MA00 | REMDESIVIR | remdesivir Injection |
| 2284718 | MA00 | REMDESIVIR | remdesivir |
| 2395502 | MA00 | REMDESIVIR | Veklury 100 MG Injection |
| 2395501 | MA00 | REMDESIVIR | Veklury Injectable Product |

| | | | |
|---|---|---|---|
| 2395498 | MA00 | REMDESIVIR | Veklury |

| Group |
| --- |
| Molupiravir |
| Molupiravir |
| Molupiravir |
| Molupiravir |
| Molupiravir |
| Molupiravir |
| Molupiravir |
| Molupiravir |
| Molupiravir |
| Molupiravir |
| Molupiravir |
| Molupiravir |
| Molupiravir |
| Molupiravir |
| casirivimab/imdevimab |
| casirivimab/imdevimab |
| casirivimab/imdevimab |
| casirivimab/imdevimab |
| casirivimab/imdevimab |
| casirivimab/imdevimab |
| casirivimab/imdevimab |
| casirivimab/imdevimab |
| casirivimab/imdevimab |
| casirivimab/imdevimab |
| casirivimab/imdevimab |
| casirivimab/imdevimab |
| casirivimab/imdevimab |
| casirivimab/imdevimab |
| casirivimab/imdevimab |
| casirivimab/imdevimab |
| casirivimab/imdevimab |
| casirivimab/imdevimab |
| casirivimab/imdevimab |
| casirivimab/imdevimab |
| casirivimab/imdevimab |
| casirivimab/imdevimab |
| casirivimab/imdevimab |

casirivimab/imdevimab

casirivimab/imdevimab

casirivimab/imdevimab

casirivimab/imdevimab
casirivimab/imdevimab
casirivimab/imdevimab
casirivimab/imdevimab
casirivimab/imdevimab
bamlanivimab

bamlanivimab
bamlanivimab
bamlanivimab
bamlanivimab
bamlanivimab/etesevimab
bamlanivimab/etesevimab
bamlanivimab/etesevimab
bamlanivimab/etesevimab
bamlanivimab/etesevimab
sotrovimab
sotrovimab
sotrovimab
sotrovimab
sotrovimab
Bebtelovimab

Bebtelovimab
Bebtelovimab
Bebtelovimab
Bebtelovimab
bamlanivimab
bamlanivimab

bamlanivimab
bamlanivimab
bamlanivimab
bamlanivimab
bamlanivimab/etesevimab
bamlanivimab/etesevimab
bamlanivimab/etesevimab
bamlanivimab/etesevimab
bamlanivimab/etesevimab
bamlanivimab/etesevimab
casirivimab/imdevimab

casirivimab/imdevimab

casirivimab/imdevimab

casirivimab/imdevimab

casirivimab/imdevimab
casirivimab/imdevimab
casirivimab/imdevimab
casirivimab/imdevimab
casirivimab/imdevimab
casirivimab/imdevimab
casirivimab/imdevimab
casirivimab/imdevimab
casirivimab/imdevimab
casirivimab/imdevimab
sotrovimab
sotrovimab
sotrovimab
sotrovimab
sotrovimab
sotrovimab
Bebtelovimab

Bebtelovimab
Bebtelovimab
Bebtelovimab
Bebtelovimab
casirivimab/imdevimab

casirivimab/imdevimab

casirivimab/imdevimab

casirivimab/imdevimab

casirivimab/imdevimab

casirivimab/imdevimab
casirivimab/imdevimab
casirivimab/imdevimab
casirivimab/imdevimab

casirivimab/imdevimab
casirivimab/imdevimab
casirivimab/imdevimab
casirivimab/imdevimab
casirivimab/imdevimab

casirivimab/imdevimab
casirivimab/imdevimab

casirivimab/imdevimab

casirivimab/imdevimab

casirivimab/imdevimab

casirivimab/imdevimab

casirivimab/imdevimab

casirivimab/imdevimab

casirivimab/imdevimab

casirivimab/imdevimab

casirivimab/imdevimab

casirivimab/imdevimab

casirivimab/imdevimab
casirivimab/imdevimab
casirivimab/imdevimab
casirivimab/imdevimab
casirivimab/imdevimab
Any_mAB

Any_mAB

bamlanivimab
bamlanivimab

bamlanivimab

bamlanivimab

bamlanivimab/etesevimab

bamlanivimab/etesevimab

casirivimab/imdevimab

casirivimab/imdevimab

casirivimab/imdevimab

casirivimab/imdevimab

sotrovimab

sotrovimab

sotrovimab
Bebtelovimab
Bebtelovimab

Bebtelovimab

bamlanivimab/etesevimab

casirivimab/imdevimab

casirivimab/imdevimab

PAXLOVID
PAXLOVID
PAXLOVID
PAXLOVID
PAXLOVID
PAXLOVID
PAXLOVID
PAXLOVID

PAXLOVID

PAXLOVID
PAXLOVID
PAXLOVID
PAXLOVID
PAXLOVID
PAXLOVID
PAXLOVID
PAXLOVID
PAXLOVID

PAXLOVID
PAXLOVID
PAXLOVID
PAXLOVID
REMDESIVIR
REMDESIVIR
REMDESIVIR
REMDESIVIR

REMDESIVIR

REMDESIVIR

REMDESIVIR

REMDESIVIR

REMDESIVIR
REMDESIVIR
REMDESIVIR
REMDESIVIR
REMDESIVIR
REMDESIVIR
REMDESIVIR
REMDESIVIR
REMDESIVIR
REMDESIVIR
REMDESIVIR
REMDESIVIR
REMDESIVIR
REMDESIVIR
REMDESIVIR
REMDESIVIR
REMDESIVIR
REMDESIVIR
REMDESIVIR
REMDESIVIR
REMDESIVIR
REMDESIVIR
REMDESIVIR
REMDESIVIR
REMDESIVIR
REMDESIVIR
REMDESIVIR

REMDESIVIR
