## Supplemental Material B for "How age and underlying conditions affect incidence of hospitalization and death after documented COVID-19 diagnosis among individuals aged ≥20 years, a retrospective cohort study using PCORnet, April 2022 – March 2025"

| Code | Codetype | Name of vaccination | Manufacturer |
|---|---|---|---|
| 80631010001 | VXND | COVID-19 vaccine, recombinant (Novavax)/adjuvant-Matrix/PF | Novavax |
| 80631010010 | VXND | COVID-19 vaccine, recombinant (Novavax)/adjuvant-Matrix/PF | Novavax |
| 91304 | VXCH | SARSCOV2 VAC 5MCG/0.5ML IM | Novavax |
| 0041A | VXCH | ADM SARSCOV2 5MCG/0.5ML 1ST | Novavax |
| 0042A | VXCH | ADM SARSCOV2 5MCG/0.5ML 2ND | Novavax |
| 211 | VXCX | SARS-COV-2 (COVID-19) vaccine, subunit, recombinant spike protein-nanoparticle+Matrix-M1 Adjuvant, preservative free, 0.5mL dose | Novavax |
| 212 | VXCX | [COVID-19]) vaccine, DNA, spike protein, adenovirus type 26 (Ad26) vector, preservative free, 5×1010 viral particles/0.5mL dosage | Janssen |
| 91303 | VXCH | Janssen COVID -19 | Janssen |
| 0031A | VXCH | Janssen Covid-19 Vaccine Administration - First Dose[3]*** | Janssen |
| 59676058005 | VXND | Janssen COVID -19 | Janssen |
| 2479835 | VXRX | Janssen COVID -19 | Janssen |
| 59676058015 | VXND | Janssen COVID -19 | Janssen |
| 0034A | VXCH | Janssen Covid-19 Vaccine Administration - Booster | Janssen |
| D1712 | VXCH | Janssen Covid-19 vaccine administration-booster dose | Janssen |
| D1707 | VXCH | Janssen Covid-19 vaccine administration | Janssen |
| 91301 | VXCH | Moderna COVID-19 Vaccine | Moderna |
| 0011A | VXCH | Moderna COVID-19 Vaccine- 1st Dose | Moderna |
| 0012A | VXCH | Moderna COVID-19 Vaccine- 2nd Dose | Moderna |
| 80777027310 | VXND | Moderna COVID-19 Vaccine | Moderna |
| 2470234 | VXRX | SARS-CoV-2 (COVID-19) vaccine, mRNA-1273 0.2 MG/ML Injectable Suspension | Moderna |
| 2470233 | VXRX | SARS-CoV-2 (COVID-19) vaccine, mRNA-1273 0.2 MG/ML | Moderna |
| 2470232 | VXRX | SARS-CoV-2 (COVID-19) vaccine, mRNA-1273 | Moderna |
| 207 | VXCX | Moderna Vaccine | Moderna |
| 0013A | VXCH | Moderna COVID-19 Vaccine- 3rd Dose | Moderna |
| 80777027315 | VXND | COVID-19 vaccine, mRNA, cx-024414, LNP-S (Moderna)/PF | Moderna |
| 80777027398 | VXND | COVID-19 vaccine, mRNA, cx-024414, LNP-S (Moderna)/PF | Moderna |
| 80777027399 | VXND | COVID-19 vaccine, mRNA, cx-024414, LNP-S (Moderna)/PF | Moderna |
| 91306 | VXCH | Moderna Covid-19 Vaccine (Low Dose) | Moderna |
| 0064A | VXCH | Moderna Covid-19 Vaccine (Low Dose) Administration - Booster | Moderna |
| 221 | VXCX | Moderna COVID-19 Vaccine Booster | Moderna |
| D1704 | VXCH | Moderna Covid-19 vaccine administration - second dose | Moderna |

| | | | |
|---|---|---|---|
| D1703 | VXCH | Moderna Covid-19 vaccine administration - first dose | Moderna |
| D1710 | VXCH | Moderna Covid-19 vaccine administration-third dose | Moderna |
| D1711 | VXCH | Moderna Covid-19 vaccine administration-booster dose | Moderna |
| 0094A | VXCH | Immunization administration by intramuscular injection of severe acute respiratory syndrome coronavirus 2 (SARS-CoV-2) (coronavirus disease [COVID-19]) vaccine, mRNA-LNP, spike protein, preservative free, 50 mcg/0.5 mL dosage, booster dose | Moderna |
| 91309 | VXCH | Severe acute respiratory syndrome coronavirus 2 (SARS-CoV-2) (coronavirus disease [COVID-19]) vaccine, mRNA-LNP, spike protein, preservative free, 50 mcg/0.5 mL dosage, for intramuscular use | Moderna |
| 91311 | VXCH | Severe acute respiratory syndrome coronavirus 2 (SARS-CoV-2) (coronavirus disease [COVID-19]) vaccine, mRNA-LNP, spike protein, preservative free, 25 mcg/0.25 mL dosage, for intramuscular use | Moderna |
| 0111A | VXCH | Moderna pediatric 1st dose booster | Moderna |
| 0112A | VXCH | Moderna pediatric 2nd dose booster | Moderna |
| 227 | VXCX | Pre-EUA Moderna Pediatric 6yr to<12 yr vaccine 2.5 mL vial, 50 mcg/0.5 mL dose | Moderna |
| 228 | VXCX | Pre-EUA Moderna Pediatric 6mo to <6yr 2.5 mL vial, 25 mcg/0.25 mL dose | Moderna |
| 0091A | VXCH | ADM SARSCOV2 50 MCG/.5 ML1ST | Moderna |
| 0092A | VXCH | ADM SARSCOV2 50 MCG/.5 ML2ND | Moderna |
| 0093A | VXCH | ADM SARSCOV2 50 MCG/.5 ML3RD | Moderna |
| 0113A | VXCH | ADM SARSCOV2 25MCG/0.25ML3RD | Moderna |
| 80777010011 | VXND | COVID-19 vaccine, mRNA, cx-024414, LNP-S (Moderna)/PF | Moderna |
| 80777010099 | VXND | COVID-19 vaccine, mRNA, cx-024414, LNP-S (Moderna)/PF | Moderna |
| 80777027505 | VXND | COVID-19 vaccine, mRNA, cx-024414, LNP-S (Moderna)/PF | Moderna |
| 80777027599 | VXND | COVID-19 vaccine, mRNA, cx-024414, LNP-S (Moderna)/PF | Moderna |
| 80777027705 | VXND | COVID-19 vaccine, mRNA, LNP-S, pediatric (Moderna)/PF | Moderna |
| 80777027799 | VXND | COVID-19 vaccine, mRNA, LNP-S, pediatric (Moderna)/PF | Moderna |
| 80777027905 | VXND | COVID-19 vaccine, mRNA, LNP-S, pediatric (Moderna)/PF | Moderna |
| 80777027999 | VXND | COVID-19 vaccine, mRNA, LNP-S, pediatric (Moderna)/PF | Moderna |
| 80777028005 | VXND | COVID-19 vaccine, bivalent, mRNA/preservative free | Moderna |
| 80777028099 | VXND | COVID-19 vaccine, bivalent, mRNA/preservative free | Moderna |
| 91300 | VXCH | Pfizer-BioNTech COVID-19 Vaccine | Pfizer |
| 0001A | VXCH | Pfizer-BioNTech COVID-19 Vaccine- 1st Dose | Pfizer |

| | | | |
|---|---|---|---|
| 0002A | VXCH | Pfizer-BioNTech COVID-19 Vaccine- 2nd Dose | Pfizer |
| 59267100001 | VXND | Pfizer-BioNTech COVID-19 Vaccine | Pfizer |
| 2468235 | VXRX | SARS-CoV-2 (COVID-19) vaccine, mRNA-BNT162b2 0.1 MG/ML Injectable Suspension | Pfizer |
| 2468232 | VXRX | SARS-CoV-2 (COVID-19) vaccine, mRNA-BNT162b2 0.1 MG/ML | Pfizer |
| 2468230 | VXRX | SARS-CoV-2 (COVID-19) vaccine, mRNA-BNT162b2 | Pfizer |
| 208 | VXCX | Pfizer_BionTech vaccine | Pfizer |
| 0003A | VXCH | Pfizer-BioNTech COVID-19 Vaccine- 3rd Dose | Pfizer |
| 0004A | VXCH | Pfizer-BioNTech COVID-19 Vaccine Booster | Pfizer |
| 59267100002 | VXND | COVID-19 vaccine, mRNA, BNT162b2, LNP-S (Pfizer)/PF | Pfizer |
| 59267100003 | VXND | COVID-19 vaccine, mRNA, BNT162b2, LNP-S (Pfizer)/PF | Pfizer |
| 91305 | VXCH | Pfizer SARSCOV2 VAC 30 MCG TRS-SUCR | Pfizer |
| 0051A | VXCH | Pfizer ADM SARSCV2 30MCG TRS-SUCR 1 | Pfizer |
| 0052A | VXCH | Pfizer ADM SARSCV2 30MCG TRS-SUCR 2 | Pfizer |
| 0053A | VXCH | Pfizer ADM SARSCV2 30MCG TRS-SUCR 3 | Pfizer |
| 0054A | VXCH | Pfizer ADM SARSCV2 30MCG TRS-SUCR B | Pfizer |
| 91307 | VXCH | Pfizer-BioNTech Covid-19 Pediatric Vaccine | Pfizer |
| 0071A | VXCH | Pfizer-BioNTech Covid-19 Pediatric Vaccine - Administration - First dose | Pfizer |
| 0072A | VXCH | Pfizer-BioNTech Covid-19 Pediatric Vaccine - Administration - Second dose | Pfizer |
| 217 | VXCX | Pfizer-BioNTech COVID-19 Vaccine for ages 12 yrs + | Pfizer |
| 218 | VXCX | Pfizer-BioNTech COVID-19 Vaccine (ORANGE CAP) for ages 5 yrs to < 12 yrs | Pfizer |
| 219 | VXCX | Pfizer-BioNTech COVID-19 Vaccine (MAROON CAP) for ages 6 mo to <5 yrs | Pfizer |
| D1714 | VXCH | Pfizer-BioNTech Covid-19 vaccine administration tris-sucrose pediatric-second dose | Pfizer |
| D1713 | VXCH | Pfizer-BioNTech Covid-19 vaccine administration tris-sucrose pediatric-first dose | Pfizer |
| D1702 | VXCH | Pfizer-BioNTech Covid-19 vaccine administration - second dose | Pfizer |
| D1701 | VXCH | Pfizer-BioNTech Covid-19 vaccine administration - first dose | Pfizer |
| D1708 | VXCH | Pfizer-BioNTech Covid-19 vaccine administration-third dose | Pfizer |
| D1709 | VXCH | Pfizer-BioNTech Covid-19 vaccine administration-booster dose | Pfizer |

| | | | |
|---|---|---|---|
| 91308 | VXCH | Severe acute respiratory syndrome coronavirus 2 (SARS-CoV-2) (coronavirus disease [COVID-19]) vaccine, mRNA-LNP, spike protein, preservative free, 3 mcg/0.2 mL dosage, diluent reconstituted, tris-sucrose formulation, for intramuscular use | Pfizer |
| 0082A | VXCH | Immunization administration by intramuscular injection of severe acute respiratory syndrome coronavirus 2 (SARS-CoV-2) (coronavirus disease [COVID-19]) vaccine, mRNA-LNP, spike protein, preservative free, 3 mcg/0.2 mL dosage, diluent reconstituted, tris-sucrose formulation; second dose | Pfizer |
| 0081A | VXCH | Immunization administration by intramuscular injection of severe acute respiratory syndrome coronavirus 2 (SARS-CoV-2) (coronavirus disease [COVID-19]) vaccine, mRNA-LNP, spike protein, preservative free, 3 mcg/0.2 mL dosage, diluent reconstituted, tris-sucrose formulation; first dose | Pfizer |
| 0073A | VXCH | Immunization administration by intramuscular injection of severe acute respiratory syndrome coronavirus 2 (SARS-CoV-2) (coronavirus disease [COVID-19]) vaccine, mRNA-LNP, spike protein, preservative free, 10 mcg/0.2 mL dosage, diluent reconstituted, tris-sucrose formulation; third dose | Pfizer |
| 0074A | VXCH | ADM SARSCV2 10MCG TRS-SUCR B | Pfizer |
| 0083A | VXCH | ADM SARSCOV2 3MCG TRS-SUCR 3 | Pfizer |
| 69100002 | VXND | COVID-19 vaccine, mRNA, BNT162b2, LNP-S (Pfizer)/PF | Pfizer |
| 69100003 | VXND | COVID-19 vaccine, mRNA, BNT162b2, LNP-S (Pfizer)/PF | Pfizer |
| 69202501 | VXND | COVID-19 vac mRNA,tris(Pfizer)/PF | Pfizer |
| 69202510 | VXND | COVID-19 vac mRNA,tris(Pfizer)/PF | Pfizer |
| 69202525 | VXND | COVID-19 vac mRNA,tris(Pfizer)/PF | Pfizer |
| 59267007801 | VXND | COVID-19 vac mRNA,tris(Pfizer)/PF | Pfizer |
| 59267007804 | VXND | COVID-19 vac mRNA,tris(Pfizer)/PF | Pfizer |
| 59267102501 | VXND | COVID-19 vac mRNA,tris(Pfizer)/PF | Pfizer |
| 59267102502 | VXND | COVID-19 vac mRNA,tris(Pfizer)/PF | Pfizer |
| 59267102503 | VXND | COVID-19 vac mRNA,tris(Pfizer)/PF | Pfizer |
| 59267102504 | VXND | COVID-19 vac mRNA,tris(Pfizer)/PF | Pfizer |
| 59267105501 | VXND | COVID-19 vac mRNA,tris(Pfizer)/PF | Pfizer |
| 59267105502 | VXND | COVID-19 vac mRNA,tris(Pfizer)/PF | Pfizer |
| 59267105504 | VXND | COVID-19 vac mRNA,tris(Pfizer)/PF | Pfizer |
| 91316 | VXCH | Moderna COVID-19 Vaccine, Bivalent Product (Aged 6 months through 5 years) (Dark Pink Cap and a label with a yellow box) | Moderna |
| 0164A | VXCH | Moderna COVID-19 Vaccine, Bivalent (Aged 6 months through 5 years) (Dark Pink Cap and label with a yellow box) Administration – Booster Dose | Moderna |

| | | | |
|---|---|---|---|
| 91313 | VXCH | Moderna COVID-19 Vaccine, Bivalent Product (Aged 12 years and older) (Dark Blue Cap with gray border)[6] | Moderna |
| 0134A | VXCH | Moderna COVID-19 Vaccine, Bivalent (Aged 12 years and older) (Dark Blue Cap with gray border) Administration – Booster Dose[6] | Moderna |
| 91314 | VXCH | Moderna COVID-19 Vaccine, Bivalent Product (Aged 6 years through 11 years) (Dark Blue Cap with gray border) | Moderna |
| 0144A | VXCH | Moderna COVID-19 Vaccine, Bivalent (Aged 6 years through 11 years) (Dark Blue Cap with gray border) Administration – Booster Dose | Moderna |
| 229 | VXCX | Moderna COVID-19 Vaccine Bivalent Booster | Moderna |
| 230 | VXCX | Moderna COVID-19 Vaccine Bivalent Booster for ages 6 months to < 6 yrs | Moderna |
| 80777028299 | VXND | Moderna COVID-19 Vaccine Bivalent Booster | Moderna |
| 80777028399 | VXND | Moderna COVID-19 Vaccine Bivalent Booster for ages 6 months to < 6 yrs | Moderna |
| 80777028205 | VXND | Moderna COVID-19 Vaccine Bivalent Booster | Moderna |
| 80777028302 | VXND | Moderna COVID-19 Vaccine Bivalent Booster for ages 6 months to < 6 yrs | Moderna |
| 91317 | VXCH | Pfizer-BioNTech COVID-19 Vaccine, Bivalent Product (Aged 6 months through 4 years) (Maroon Cap) | Pfizer |
| 0173A | VXCH | Pfizer-BioNTech Covid-19 Pediatric Vaccine (Aged 6 months through 4 years) (Maroon Cap) Administration - Third dose | Pfizer |
| 91315 | VXCH | Pfizer-BioNTech COVID-19 Vaccine, Bivalent Product (Aged 5 years through 11 years) (Orange Cap) | Pfizer |
| 0154A | VXCH | Pfizer-BioNTech COVID-19 Vaccine, Bivalent Product (Aged 5 years through 11 years) (Orange Cap) Administration – Booster Dose | Pfizer |
| 91312 | VXCH | Pfizer-BioNTech COVID-19 Vaccine, Bivalent Product (Aged 12 years and older) (Gray Cap) | Pfizer |
| 0124A | VXCH | Pfizer-BioNTech COVID-19 Vaccine, Bivalent (Gray Cap) Administration – Booster Dose | Pfizer |
| 300 | VXCX | Pfizer-BioNTech COVID-19 Vaccine Bivalent Booster | Pfizer |
| 301 | VXCX | Pfizer-BioNTech COVID-19 Vaccine Bivalent Booster | Pfizer |
| 302 | VXCX | Pfizer-BioNTech COVID-19 Vaccine Bivalent 6 months through 4 years (Tris-sucrose formulation) | Pfizer |
| 59267030402 | VXND | Pfizer-BioNTech COVID-19 Vaccine Bivalent Booster | Pfizer |
| 59267056502 | VXND | Pfizer-BioNTech COVID-19 Vaccine Bivalent Booster | Pfizer |
| 59267060902 | VXND | Pfizer-BioNTech COVID-19 Vaccine Bivalent 6 months through 4 years (Tris-sucrose formulation) | Pfizer |

| | | | |
|---|---|---|---|
| 59267030401 | VXND | Pfizer-BioNTech COVID-19 Vaccine Bivalent Booster | Pfizer |
| 59267056501 | VXND | Pfizer-BioNTech COVID-19 Vaccine Bivalent Booster | Pfizer |
| 59267060901 | VXND | Pfizer-BioNTech COVID-19 Vaccine Bivalent 6 months through 4 years (Tris-sucrose formulation) | Pfizer |
| 308 | VXCX | COVID-19, mRNA, LNP-S, PF, tris-sucrose, 3 mcg/0.3 mL | Pfizer |
| 309 | VXCX | COVID-19, mRNA, LNP-S, PF, tris-sucrose, 30 mcg/0.3 mL | Pfizer |
| 310 | VXCX | COVID-19, mRNA, LNP-S, PF, tris-sucrose, 10 mcg/0.3 mL | Pfizer |
| 91318 | VXCH | Severe acute respiratory syndrome coronavirus 2 (SARS-CoV-2) (coronavirus disease [COVID-19]) vaccine, mRNA-LNP, spike protein, 3 mcg/0.2 mL dosage, tris-sucrose formulation, for intramuscular use | Pfizer |
| 91320 | VXCH | Severe acute respiratory syndrome coronavirus 2 (SARS-CoV-2) (coronavirus disease [COVID-19]) vaccine, mRNA-LNP, spike protein, 30 mcg/0.3 mL dosage, tris-sucrose formulation, for intramuscular use | Pfizer |
| 91319 | VXCH | Severe acute respiratory syndrome coronavirus 2 (SARS-CoV-2) (coronavirus disease [COVID-19]) vaccine, mRNA-LNP, spike protein, 10 mcg/0.2 mL dosage, tris-sucrose formulation, for intramuscular use | Pfizer |
| 59267433102 | VXND | COVID-19, mRNA, LNP-S, PF, tris-sucrose, 10 mcg/0.3 mL | Pfizer |
| 69236210 | VXND | COVID-19, mRNA, LNP-S, PF, tris-sucrose, 30 mcg/0.3 mL | Pfizer |
| 59267431502 | VXND | COVID-19, mRNA, LNP-S, PF, tris-sucrose, 3 mcg/0.3 mL | Pfizer |
| 69239210 | VXND | COVID-19, mRNA, LNP-S, PF, tris-sucrose, 30 mcg/0.3 mL | Pfizer |
| 312 | VXCX | COVID-19, mRNA, LNP-S, PF, 50 mcg/0.5 mL | Moderna |
| 311 | VXCX | COVID-19, mRNA, LNP-S, PF, 25 mcg/0.25 mL | Moderna |
| 91322 | VXCH | Severe acute respiratory syndrome coronavirus 2 (SARS-CoV-2) (coronavirus disease [COVID-19]) vaccine, mRNA-LNP, 50 mcg/0.5 mL dosage, for intramuscular use | Moderna |
| 91321 | VXCH | Severe acute respiratory syndrome coronavirus 2 (SARS-CoV-2) (coronavirus disease [COVID-19]) vaccine, mRNA-LNP, 25 mcg/0.25 mL dosage, for intramuscular use | Moderna |
| 80777010295 | VXND | COVID-19, mRNA, LNP-S, PF, 50 mcg/0.5 mL | Moderna |
| 80777010293 | VXND | COVID-19, mRNA, LNP-S, PF, 50 mcg/0.5 mL | Moderna |
| 80777010296 | VXND | COVID-19, mRNA, LNP-S, PF, 50 mcg/0.5 mL | Moderna |

| | | | |
|---|---|---|---|
| 80777028792 | VXND | COVID-19, mRNA, LNP-S, PF, 25 mcg/0.25 mL | Moderna |
| 313 | VXCX | COVID-19, subunit, rS-nanoparticle, adjuvanted, PF, 5 mcg/0.5 mL | Novavax |
| 80631010502 | VXND | NOVAVAX COVID-19 Vaccine, Adjuvanted | Novavax |
| 2623378 | VXRX | Moderna COVID-19 Vaccine, Bivalent for children 6 months through 5 years of age | Moderna |
| 2610328 | VXRX | Moderna Covid-19 Vaccine, Bivalent | Moderna |
| 2598700 | VXRX | Second Booster Dose of the Moderna COVID-19 Vaccine | Moderna |
| 2601552 | VXRX | Spikevax | Moderna |
| 2623382 | VXRX | Pfizer-BioNTech COVID-19 Vaccine, Bivalent for children 6 months through 4 years of age | Pfizer |
| 2610319 | VXRX | Pfizer-BioNTech Covid-19 Vaccine, Bivalent | Pfizer |
| 2610347 | VXRX | Pfizer-BioNTech Covid-19 Vaccine, Bivalent | Pfizer |
| 2593847 | VXRX | Comirnaty | Pfizer |
| 2583743 | VXRX | Pfizer-BioNTech COVID-19 Vaccine for children 5 through 11 years of age | Pfizer |
| 2606078 | VXRX | Novavax COVID-19 Vaccine, Adjuvanted | Novavax |
| 91304 | PXCH | SARSCOV2 VAC 5MCG/0.5ML IM | Novavax |
| 0041A | PXCH | ADM SARSCOV2 5MCG/0.5ML 1ST | Novavax |
| 0042A | PXCH | ADM SARSCOV2 5MCG/0.5ML 2ND | Novavax |
| 91303 | PXCH | Janssen COVID -19 | Janssen |
| 0031A | PXCH | Janssen Covid-19 Vaccine Administration - First Dose[3]*** | Janssen |
| 0034A | PXCH | Janssen Covid-19 Vaccine Administration - Booster | Janssen |
| D1712 | PXCH | Janssen Covid-19 vaccine administration-booster dose | Janssen |
| D1707 | PXCH | Janssen Covid-19 vaccine administration | Janssen |
| 91301 | PXCH | Moderna COVID-19 Vaccine | Moderna |
| 0011A | PXCH | Moderna COVID-19 Vaccine- 1st Dose | Moderna |
| 0012A | PXCH | Moderna COVID-19 Vaccine- 2nd Dose | Moderna |
| 0013A | PXCH | Moderna COVID-19 Vaccine- 3rd Dose | Moderna |
| 91306 | PXCH | Moderna Covid-19 Vaccine (Low Dose) | Moderna |
| 0064A | PXCH | Moderna Covid-19 Vaccine (Low Dose) Administration - Booster | Moderna |
| D1704 | PXCH | Moderna Covid-19 vaccine administration - second dose | Moderna |
| D1703 | PXCH | Moderna Covid-19 vaccine administration - first dose | Moderna |
| D1710 | PXCH | Moderna Covid-19 vaccine administration-third dose | Moderna |
| D1711 | PXCH | Moderna Covid-19 vaccine administration-booster dose | Moderna |

| | | | |
|---|---|---|---|
| 0094A | PXCH | Immunization administration by intramuscular injection of severe acute respiratory syndrome coronavirus 2 (SARS-CoV-2) (coronavirus disease [COVID-19]) vaccine, mRNA-LNP, spike protein, preservative free, 50 mcg/0.5 mL dosage, booster dose | Moderna |
| 91309 | PXCH | Severe acute respiratory syndrome coronavirus 2 (SARS-CoV-2) (coronavirus disease [COVID-19]) vaccine, mRNA-LNP, spike protein, preservative free, 50 mcg/0.5 mL dosage, for intramuscular use | Moderna |
| 91311 | PXCH | Severe acute respiratory syndrome coronavirus 2 (SARS-CoV-2) (coronavirus disease [COVID-19]) vaccine, mRNA-LNP, spike protein, preservative free, 25 mcg/0.25 mL dosage, for intramuscular use | Moderna |
| 0111A | PXCH | Moderna pediatric 1st dose booster | Moderna |
| 0112A | PXCH | Moderna pediatric 2nd dose booster | Moderna |
| 0091A | PXCH | ADM SARSCOV2 50 MCG/.5 ML1ST | Moderna |
| 0092A | PXCH | ADM SARSCOV2 50 MCG/.5 ML2ND | Moderna |
| 0093A | PXCH | ADM SARSCOV2 50 MCG/.5 ML3RD | Moderna |
| 0113A | PXCH | ADM SARSCOV2 25MCG/0.25ML3RD | Moderna |
| 91300 | PXCH | Pfizer-BioNTech COVID-19 Vaccine | Pfizer |
| 0001A | PXCH | Pfizer-BioNTech COVID-19 Vaccine- 1st Dose | Pfizer |
| 0002A | PXCH | Pfizer-BioNTech COVID-19 Vaccine- 2nd Dose | Pfizer |
| 0003A | PXCH | Pfizer-BioNTech COVID-19 Vaccine- 3rd Dose | Pfizer |
| 0004A | PXCH | Pfizer-BioNTech COVID-19 Vaccine Booster | Pfizer |
| 91305 | PXCH | Pfizer SARSCOV2 VAC 30 MCG TRS-SUCR | Pfizer |
| 0051A | PXCH | Pfizer ADM SARSCV2 30MCG TRS-SUCR 1 | Pfizer |
| 0052A | PXCH | Pfizer ADM SARSCV2 30MCG TRS-SUCR 2 | Pfizer |
| 0053A | PXCH | Pfizer ADM SARSCV2 30MCG TRS-SUCR 3 | Pfizer |
| 0054A | PXCH | Pfizer ADM SARSCV2 30MCG TRS-SUCR B | Pfizer |
| 91307 | PXCH | Pfizer-BioNTech Covid-19 Pediatric Vaccine | Pfizer |
| 0071A | PXCH | Pfizer-BioNTech Covid-19 Pediatric Vaccine - Administration - First dose | Pfizer |
| 0072A | PXCH | Pfizer-BioNTech Covid-19 Pediatric Vaccine - Administration - Second dose | Pfizer |
| D1714 | PXCH | Pfizer-BioNTech Covid-19 vaccine administration tris-sucrose pediatric-second dose | Pfizer |
| D1713 | PXCH | Pfizer-BioNTech Covid-19 vaccine administration tris-sucrose pediatric-first dose | Pfizer |
| D1702 | PXCH | Pfizer-BioNTech Covid-19 vaccine administration - second dose | Pfizer |

| | | | |
|---|---|---|---|
| D1701 | PXCH | Pfizer-BioNTech Covid-19 vaccine administration - first dose | Pfizer |
| D1708 | PXCH | Pfizer-BioNTech Covid-19 vaccine administration-third dose | Pfizer |
| D1709 | PXCH | Pfizer-BioNTech Covid-19 vaccine administration-booster dose | Pfizer |
| 91308 | PXCH | Severe acute respiratory syndrome coronavirus 2 (SARS-CoV-2) (coronavirus disease [COVID-19]) vaccine, mRNA-LNP, spike protein, preservative free, 3 mcg/0.2 mL dosage, diluent reconstituted, tris-sucrose formulation, for intramuscular use | Pfizer |
| 0082A | PXCH | Immunization administration by intramuscular injection of severe acute respiratory syndrome coronavirus 2 (SARS-CoV-2) (coronavirus disease [COVID-19]) vaccine, mRNA-LNP, spike protein, preservative free, 3 mcg/0.2 mL dosage, diluent reconstituted, tris-sucrose formulation; second dose | Pfizer |
| 0081A | PXCH | Immunization administration by intramuscular injection of severe acute respiratory syndrome coronavirus 2 (SARS-CoV-2) (coronavirus disease [COVID-19]) vaccine, mRNA-LNP, spike protein, preservative free, 3 mcg/0.2 mL dosage, diluent reconstituted, tris-sucrose formulation; first dose | Pfizer |
| 0073A | PXCH | Immunization administration by intramuscular injection of severe acute respiratory syndrome coronavirus 2 (SARS-CoV-2) (coronavirus disease [COVID-19]) vaccine, mRNA-LNP, spike protein, preservative free, 10 mcg/0.2 mL dosage, diluent reconstituted, tris-sucrose formulation; third dose | Pfizer |
| 0074A | PXCH | ADM SARSCV2 10MCG TRS-SUCR B | Pfizer |
| 0083A | PXCH | ADM SARSCOV2 3MCG TRS-SUCR 3 | Pfizer |
| 91316 | PXCH | Moderna COVID-19 Vaccine, Bivalent Product (Aged 6 months through 5 years) (Dark Pink Cap and a label with a yellow box) | Moderna |
| 0164A | PXCH | Moderna COVID-19 Vaccine, Bivalent (Aged 6 months through 5 years) (Dark Pink Cap and label with a yellow box) Administration – Booster Dose | Moderna |
| 91313 | PXCH | Moderna COVID-19 Vaccine, Bivalent Product (Aged 12 years and older) (Dark Blue Cap with gray border)[6] | Moderna |
| 0134A | PXCH | Moderna COVID-19 Vaccine, Bivalent (Aged 12 years and older) (Dark Blue Cap with gray border) Administration – Booster Dose[6] | Moderna |
| 91314 | PXCH | Moderna COVID-19 Vaccine, Bivalent Product (Aged 6 years through 11 years) (Dark Blue Cap with gray border) | Moderna |

| | | | |
|---|---|---|---|
| 0144A | PXCH | Moderna COVID-19 Vaccine, Bivalent (Aged 6 years through 11 years) (Dark Blue Cap with gray border) Administration – Booster Dose | Moderna |
| 91317 | PXCH | Pfizer-BioNTech COVID-19 Vaccine, Bivalent Product (Aged 6 months through 4 years) (Maroon Cap) | Pfizer |
| 0173A | PXCH | Pfizer-BioNTech Covid-19 Pediatric Vaccine (Aged 6 months through 4 years) (Maroon Cap) Administration - Third dose | Pfizer |
| 91315 | PXCH | Pfizer-BioNTech COVID-19 Vaccine, Bivalent Product (Aged 5 years through 11 years) (Orange Cap) | Pfizer |
| 0154A | PXCH | Pfizer-BioNTech COVID-19 Vaccine, Bivalent Product (Aged 5 years through 11 years) (Orange Cap) Administration – Booster Dose | Pfizer |
| 91312 | PXCH | Pfizer-BioNTech COVID-19 Vaccine, Bivalent Product (Aged 12 years and older) (Gray Cap) | Pfizer |
| 0124A | PXCH | Pfizer-BioNTech COVID-19 Vaccine, Bivalent (Gray Cap) Administration – Booster Dose | Pfizer |
| 91318 | PXCH | Severe acute respiratory syndrome coronavirus 2 (SARS-CoV-2) (coronavirus disease [COVID-19]) vaccine, mRNA-LNP, spike protein, 3 mcg/0.2 mL dosage, tris-sucrose formulation, for intramuscular use | Pfizer |
| 91320 | PXCH | Severe acute respiratory syndrome coronavirus 2 (SARS-CoV-2) (coronavirus disease [COVID-19]) vaccine, mRNA-LNP, spike protein, 30 mcg/0.3 mL dosage, tris-sucrose formulation, for intramuscular use | Pfizer |
| 91319 | PXCH | Severe acute respiratory syndrome coronavirus 2 (SARS-CoV-2) (coronavirus disease [COVID-19]) vaccine, mRNA-LNP, spike protein, 10 mcg/0.2 mL dosage, tris-sucrose formulation, for intramuscular use | Pfizer |
| 91322 | PXCH | Severe acute respiratory syndrome coronavirus 2 (SARS-CoV-2) (coronavirus disease [COVID-19]) vaccine, mRNA-LNP, 50 mcg/0.5 mL dosage, for intramuscular use | Moderna |
| 91321 | PXCH | Severe acute respiratory syndrome coronavirus 2 (SARS-CoV-2) (coronavirus disease [COVID-19]) vaccine, mRNA-LNP, 25 mcg/0.25 mL dosage, for intramuscular use | Moderna |
