## Supplemental Material C for "How age and underlying conditions affect incidence of hospitalization and death after documented COVID-19 diagnosis among individuals aged ≥20 years, a retrospective cohort study using PCORnet, April 2022 – March 2025"

| Code | Code type | Description | Dx_Category | Code2 |
|---|---|---|---|---|
| 42610 | DX10 | ARRHYT_ELIX | Arrythmia | 42610 |
| 42611 | DX10 | ARRHYT_ELIX | Arrythmia | 42611 |
| 42613 | DX10 | ARRHYT_ELIX | Arrythmia | 42613 |
| 4262* | DX10 | ARRHYT_ELIX | Arrythmia | 4262* |
| 4263* | DX10 | ARRHYT_ELIX | Arrythmia | 4263* |
| 4264* | DX10 | ARRHYT_ELIX | Arrythmia | 4264* |
| 42650 | DX10 | ARRHYT_ELIX | Arrythmia | 42650 |
| 42651 | DX10 | ARRHYT_ELIX | Arrythmia | 42651 |
| 42652 | DX10 | ARRHYT_ELIX | Arrythmia | 42652 |
| 42653 | DX10 | ARRHYT_ELIX | Arrythmia | 42653 |
| 4266* | DX10 | ARRHYT_ELIX | Arrythmia | 4266* |
| 4267* | DX10 | ARRHYT_ELIX | Arrythmia | 4267* |
| 4268* | DX10 | ARRHYT_ELIX | Arrythmia | 4268* |
| 4270* | DX10 | ARRHYT_ELIX | Arrythmia | 4270* |
| 4272* | DX10 | ARRHYT_ELIX | Arrythmia | 4272* |
| 42731 | DX10 | ARRHYT_ELIX | Arrythmia | 42731 |
| 42760 | DX10 | ARRHYT_ELIX | Arrythmia | 42760 |
| 4279* | DX10 | ARRHYT_ELIX | Arrythmia | 4279* |
| 7850* | DX10 | ARRHYT_ELIX | Arrythmia | 7850* |
| V450* | DX10 | ARRHYT_ELIX | Arrythmia | V450* |
| V533* | DX10 | ARRHYT_ELIX | Arrythmia | V533* |
| I440 | DX10 | ARRHYT_ELIX | Arrythmia | I440 |
| I441 | DX10 | ARRHYT_ELIX | Arrythmia | I441 |
| I443 | DX10 | ARRHYT_ELIX | Arrythmia | I443 |
| I4430 | DX10 | ARRHYT_ELIX | Arrythmia | I4430 |
| I4439 | DX10 | ARRHYT_ELIX | Arrythmia | I4439 |
| I444 | DX10 | ARRHYT_ELIX | Arrythmia | I444 |
| I445 | DX10 | ARRHYT_ELIX | Arrythmia | I445 |
| I446 | DX10 | ARRHYT_ELIX | Arrythmia | I446 |
| I4460 | DX10 | ARRHYT_ELIX | Arrythmia | I4460 |
| I4469 | DX10 | ARRHYT_ELIX | Arrythmia | I4469 |
| I447 | DX10 | ARRHYT_ELIX | Arrythmia | I447 |
| I450 | DX10 | ARRHYT_ELIX | Arrythmia | I450 |
| I451 | DX10 | ARRHYT_ELIX | Arrythmia | I451 |
| I4510 | DX10 | ARRHYT_ELIX | Arrythmia | I4510 |
| I4519 | DX10 | ARRHYT_ELIX | Arrythmia | I4519 |
| I452 | DX10 | ARRHYT_ELIX | Arrythmia | I452 |
| I454 | DX10 | ARRHYT_ELIX | Arrythmia | I454 |
| I455 | DX10 | ARRHYT_ELIX | Arrythmia | I455 |
| I456 | DX10 | ARRHYT_ELIX | Arrythmia | I456 |
| I458 | DX10 | ARRHYT_ELIX | Arrythmia | I458 |
| I4581 | DX10 | ARRHYT_ELIX | Arrythmia | I4581 |
| I4589 | DX10 | ARRHYT_ELIX | Arrythmia | I4589 |
| I459 | DX10 | ARRHYT_ELIX | Arrythmia | I459 |
| I47 | DX10 | ARRHYT_ELIX | Arrythmia | I47 |
| I470 | DX10 | ARRHYT_ELIX | Arrythmia | I470 |
| I471 | DX10 | ARRHYT_ELIX | Arrythmia | I471 |
| I472 | DX10 | ARRHYT_ELIX | Arrythmia | I472 |
| I479 | DX10 | ARRHYT_ELIX | Arrythmia | I479 |
| I48 | DX10 | ARRHYT_ELIX | Arrythmia | I48 |
| I480 | DX10 | ARRHYT_ELIX | Arrythmia | I480 |
| I481 | DX10 | ARRHYT_ELIX | Arrythmia | I481 |
| I482 | DX10 | ARRHYT_ELIX | Arrythmia | I482 |
| I483 | DX10 | ARRHYT_ELIX | Arrythmia | I483 |
| I484 | DX10 | ARRHYT_ELIX | Arrythmia | I484 |
| I489 | DX10 | ARRHYT_ELIX | Arrythmia | I489 |
| I4891 | DX10 | ARRHYT_ELIX | Arrythmia | I4891 |
| I4892 | DX10 | ARRHYT_ELIX | Arrythmia | I4892 |

| | | | | |
|---|---|---|---|---|
| I49 | DX10 | ARRHYT_ELIX | Arrythmia | I49 |
| I490 | DX10 | ARRHYT_ELIX | Arrythmia | I490 |
| I4901 | DX10 | ARRHYT_ELIX | Arrythmia | I4901 |
| I4902 | DX10 | ARRHYT_ELIX | Arrythmia | I4902 |
| I491 | DX10 | ARRHYT_ELIX | Arrythmia | I491 |
| I492 | DX10 | ARRHYT_ELIX | Arrythmia | I492 |
| I493 | DX10 | ARRHYT_ELIX | Arrythmia | I493 |
| I494 | DX10 | ARRHYT_ELIX | Arrythmia | I494 |
| I4940 | DX10 | ARRHYT_ELIX | Arrythmia | I4940 |
| I4949 | DX10 | ARRHYT_ELIX | Arrythmia | I4949 |
| I495 | DX10 | ARRHYT_ELIX | Arrythmia | I495 |
| I498 | DX10 | ARRHYT_ELIX | Arrythmia | I498 |
| I499 | DX10 | ARRHYT_ELIX | Arrythmia | I499 |
| R008 | DX10 | ARRHYT_ELIX | Arrythmia | R008 |
| T821 | DX10 | ARRHYT_ELIX | Arrythmia | T821 |
| T8211 | DX10 | ARRHYT_ELIX | Arrythmia | T8211 |
| T82110 | DX10 | ARRHYT_ELIX | Arrythmia | T82110 |
| T82110A | DX10 | ARRHYT_ELIX | Arrythmia | T82110A |
| T82110D | DX10 | ARRHYT_ELIX | Arrythmia | T82110D |
| T82110S | DX10 | ARRHYT_ELIX | Arrythmia | T82110S |
| T82111 | DX10 | ARRHYT_ELIX | Arrythmia | T82111 |
| T82111A | DX10 | ARRHYT_ELIX | Arrythmia | T82111A |
| T82111D | DX10 | ARRHYT_ELIX | Arrythmia | T82111D |
| T82111S | DX10 | ARRHYT_ELIX | Arrythmia | T82111S |
| T82118 | DX10 | ARRHYT_ELIX | Arrythmia | T82118 |
| T82118A | DX10 | ARRHYT_ELIX | Arrythmia | T82118A |
| T82118D | DX10 | ARRHYT_ELIX | Arrythmia | T82118D |
| T82118S | DX10 | ARRHYT_ELIX | Arrythmia | T82118S |
| T82119 | DX10 | ARRHYT_ELIX | Arrythmia | T82119 |
| T82119A | DX10 | ARRHYT_ELIX | Arrythmia | T82119A |
| T82119D | DX10 | ARRHYT_ELIX | Arrythmia | T82119D |
| T82119S | DX10 | ARRHYT_ELIX | Arrythmia | T82119S |
| T8212 | DX10 | ARRHYT_ELIX | Arrythmia | T8212 |
| T82120 | DX10 | ARRHYT_ELIX | Arrythmia | T82120 |
| T82120A | DX10 | ARRHYT_ELIX | Arrythmia | T82120A |
| T82120D | DX10 | ARRHYT_ELIX | Arrythmia | T82120D |
| T82120S | DX10 | ARRHYT_ELIX | Arrythmia | T82120S |
| T82121 | DX10 | ARRHYT_ELIX | Arrythmia | T82121 |
| T82121A | DX10 | ARRHYT_ELIX | Arrythmia | T82121A |
| T82121D | DX10 | ARRHYT_ELIX | Arrythmia | T82121D |
| T82121S | DX10 | ARRHYT_ELIX | Arrythmia | T82121S |
| T82128 | DX10 | ARRHYT_ELIX | Arrythmia | T82128 |
| T82128A | DX10 | ARRHYT_ELIX | Arrythmia | T82128A |
| T82128D | DX10 | ARRHYT_ELIX | Arrythmia | T82128D |
| T82128S | DX10 | ARRHYT_ELIX | Arrythmia | T82128S |
| T82129 | DX10 | ARRHYT_ELIX | Arrythmia | T82129 |
| T82129A | DX10 | ARRHYT_ELIX | Arrythmia | T82129A |
| T82129D | DX10 | ARRHYT_ELIX | Arrythmia | T82129D |
| T82129S | DX10 | ARRHYT_ELIX | Arrythmia | T82129S |
| T8219 | DX10 | ARRHYT_ELIX | Arrythmia | T8219 |
| T82190 | DX10 | ARRHYT_ELIX | Arrythmia | T82190 |
| T82190A | DX10 | ARRHYT_ELIX | Arrythmia | T82190A |
| T82190D | DX10 | ARRHYT_ELIX | Arrythmia | T82190D |
| T82190S | DX10 | ARRHYT_ELIX | Arrythmia | T82190S |
| T82191 | DX10 | ARRHYT_ELIX | Arrythmia | T82191 |
| T82191A | DX10 | ARRHYT_ELIX | Arrythmia | T82191A |
| T82191D | DX10 | ARRHYT_ELIX | Arrythmia | T82191D |
| T82191S | DX10 | ARRHYT_ELIX | Arrythmia | T82191S |
| T82198 | DX10 | ARRHYT_ELIX | Arrythmia | T82198 |

| | | | | |
|---|---|---|---|---|
| T82198A | DX10 | ARRHYT_ELIX | Arrythmia | T82198A |
| T82198D | DX10 | ARRHYT_ELIX | Arrythmia | T82198D |
| T82198S | DX10 | ARRHYT_ELIX | Arrythmia | T82198S |
| T82199 | DX10 | ARRHYT_ELIX | Arrythmia | T82199 |
| T82199A | DX10 | ARRHYT_ELIX | Arrythmia | T82199A |
| T82199D | DX10 | ARRHYT_ELIX | Arrythmia | T82199D |
| T82199S | DX10 | ARRHYT_ELIX | Arrythmia | T82199S |
| Z450 | DX10 | ARRHYT_ELIX | Arrythmia | Z450 |
| Z4501 | DX10 | ARRHYT_ELIX | Arrythmia | Z4501 |
| Z45010 | DX10 | ARRHYT_ELIX | Arrythmia | Z45010 |
| Z45018 | DX10 | ARRHYT_ELIX | Arrythmia | Z45018 |
| Z4502 | DX10 | ARRHYT_ELIX | Arrythmia | Z4502 |
| Z4509 | DX10 | ARRHYT_ELIX | Arrythmia | Z4509 |
| Z950 | DX10 | ARRHYT_ELIX | Arrythmia | Z950 |
| Z95810 | DX10 | ARRHYT_ELIX | Arrythmia | Z95810 |
| Z95818 | DX10 | ARRHYT_ELIX | Arrythmia | Z95818 |
| Z959 | DX10 | ARRHYT_ELIX | Arrythmia | Z959 |
| J45 | DX10 | Asthma | Asthma | J45 |
| J45.2 | DX10 | Mild intermittent asthma | Asthma | J452 |
| J45.20 | DX10 | Mild intermittent asthma, uncomplicated | Asthma | J4520 |
| J45.21 | DX10 | Mild intermittent asthma with (acute) exacerbation | Asthma | J4521 |
| J45.22 | DX10 | Mild intermittent asthma with status asthmaticus | Asthma | J4522 |
| J45.3 | DX10 | Mild persistent asthma | Asthma | J453 |
| J45.30 | DX10 | Mild persistent asthma, uncomplicated | Asthma | J4530 |
| J45.31 | DX10 | Mild persistent asthma with (acute) exacerbation | Asthma | J4531 |
| J45.32 | DX10 | Mild persistent asthma with status asthmaticus | Asthma | J4532 |
| J45.4 | DX10 | Moderate persistent asthma | Asthma | J454 |
| J45.40 | DX10 | Moderate persistent asthma, uncomplicated | Asthma | J4540 |
| J45.41 | DX10 | Moderate persistent asthma with (acute) exacerbation | Asthma | J4541 |
| J45.42 | DX10 | Moderate persistent asthma with status asthmaticus | Asthma | J4542 |
| J45.5 | DX10 | Severe persistent asthma | Asthma | J455 |
| J45.50 | DX10 | Severe persistent asthma, uncomplicated | Asthma | J4550 |
| J45.51 | DX10 | Severe persistent asthma with (acute) exacerbation | Asthma | J4551 |
| J45.52 | DX10 | Severe persistent asthma with status asthmaticus | Asthma | J4552 |
| J45.9 | DX10 | Other and unspecified asthma | Asthma | J459 |
| J45.90 | DX10 | Unspecified asthma | Asthma | J4590 |
| J45.901 | DX10 | Unspecified asthma with (acute) exacerbation | Asthma | J45901 |
| J45.902 | DX10 | Unspecified asthma with status asthmaticus | Asthma | J45902 |
| J45.909 | DX10 | Unspecified asthma, uncomplicated | Asthma | J45909 |
| J45.99 | DX10 | Other asthma | Asthma | J4599 |
| J45.990 | DX10 | Exercise induced bronchospasm | Asthma | J45990 |
| J45.991 | DX10 | Cough variant asthma | Asthma | J45991 |
| J45.998 | DX10 | Other asthma | Asthma | J45998 |
| J82.83 | DX10 | Eosinophilic asthma | Asthma | J8283 |
| C00 | DX10 | Malignant neoplasm of lip | Cancer | C00 |
| C00.0 | DX10 | Malignant neoplasm of external upper lip | Cancer | C000 |
| C00.1 | DX10 | Malignant neoplasm of external lower lip | Cancer | C001 |
| C00.2 | DX10 | Malignant neoplasm of external lip, unspecified | Cancer | C002 |
| C00.3 | DX10 | Malignant neoplasm of upper lip, inner aspect | Cancer | C003 |
| C00.4 | DX10 | Malignant neoplasm of lower lip, inner aspect | Cancer | C004 |
| C00.5 | DX10 | Malignant neoplasm of lip, unspecified, inner aspect | Cancer | C005 |
| C00.6 | DX10 | Malignant neoplasm of commissure of lip, unspecified | Cancer | C006 |
| C00.8 | DX10 | Malignant neoplasm of overlapping sites of lip | Cancer | C008 |
| C00.9 | DX10 | Malignant neoplasm of lip, unspecified | Cancer | C009 |
| C01 | DX10 | Malignant neoplasm of base of tongue | Cancer | C01 |
| C02 | DX10 | Malignant neoplasm of other and unspecified parts of tongue | Cancer | C02 |
| C02.0 | DX10 | Malignant neoplasm of dorsal surface of tongue | Cancer | C020 |
| C02.1 | DX10 | Malignant neoplasm of border of tongue | Cancer | C021 |

| | | | | |
|---|---|---|---|---|
| C02.2 | DX10 | Malignant neoplasm of ventral surface of tongue | Cancer | C022 |
| C02.3 | DX10 | Malig neoplasm of anterior two-thirds of tongue, part unsp | Cancer | C023 |
| C02.4 | DX10 | Malignant neoplasm of lingual tonsil | Cancer | C024 |
| C02.8 | DX10 | Malignant neoplasm of overlapping sites of tongue | Cancer | C028 |
| C02.9 | DX10 | Malignant neoplasm of tongue, unspecified | Cancer | C029 |
| C03 | DX10 | Malignant neoplasm of gum | Cancer | C03 |
| C03.0 | DX10 | Malignant neoplasm of upper gum | Cancer | C030 |
| C03.1 | DX10 | Malignant neoplasm of lower gum | Cancer | C031 |
| C03.9 | DX10 | Malignant neoplasm of gum, unspecified | Cancer | C039 |
| C04 | DX10 | Malignant neoplasm of floor of mouth | Cancer | C04 |
| C04.0 | DX10 | Malignant neoplasm of anterior floor of mouth | Cancer | C040 |
| C04.1 | DX10 | Malignant neoplasm of lateral floor of mouth | Cancer | C041 |
| C04.8 | DX10 | Malignant neoplasm of overlapping sites of floor of mouth | Cancer | C048 |
| C04.9 | DX10 | Malignant neoplasm of floor of mouth, unspecified | Cancer | C049 |
| C05 | DX10 | Malignant neoplasm of palate | Cancer | C05 |
| C05.0 | DX10 | Malignant neoplasm of hard palate | Cancer | C050 |
| C05.1 | DX10 | Malignant neoplasm of soft palate | Cancer | C051 |
| C05.2 | DX10 | Malignant neoplasm of uvula | Cancer | C052 |
| C05.8 | DX10 | Malignant neoplasm of overlapping sites of palate | Cancer | C058 |
| C05.9 | DX10 | Malignant neoplasm of palate, unspecified | Cancer | C059 |
| C06 | DX10 | Malignant neoplasm of other and unspecified parts of mouth | Cancer | C06 |
| C06.0 | DX10 | Malignant neoplasm of cheek mucosa | Cancer | C060 |
| C06.1 | DX10 | Malignant neoplasm of vestibule of mouth | Cancer | C061 |
| C06.2 | DX10 | Malignant neoplasm of retromolar area | Cancer | C062 |
| C06.8 | DX10 | Malignant neoplasm of ovrlp sites of and unsp parts of mouth | Cancer | C068 |
| C06.80 | DX10 | Malignant neoplasm of ovrlp sites of unsp parts of mouth | Cancer | C0680 |
| C06.89 | DX10 | Malignant neoplasm of overlapping sites of oth prt mouth | Cancer | C0689 |
| C06.9 | DX10 | Malignant neoplasm of mouth, unspecified | Cancer | C069 |
| C07 | DX10 | Malignant neoplasm of parotid gland | Cancer | C07 |
| C08 | DX10 | Malignant neoplasm of other and unsp major salivary glands | Cancer | C08 |
| C08.0 | DX10 | Malignant neoplasm of submandibular gland | Cancer | C080 |
| C08.1 | DX10 | Malignant neoplasm of sublingual gland | Cancer | C081 |
| C08.9 | DX10 | Malignant neoplasm of major salivary gland, unspecified | Cancer | C089 |
| C09 | DX10 | Malignant neoplasm of tonsil | Cancer | C09 |
| C09.0 | DX10 | Malignant neoplasm of tonsillar fossa | Cancer | C090 |
| C09.1 | DX10 | Malig neoplasm of tonsillar pillar (anterior) (posterior) | Cancer | C091 |
| C09.8 | DX10 | Malignant neoplasm of overlapping sites of tonsil | Cancer | C098 |
| C09.9 | DX10 | Malignant neoplasm of tonsil, unspecified | Cancer | C099 |
| C10 | DX10 | Malignant neoplasm of oropharynx | Cancer | C10 |
| C10.0 | DX10 | Malignant neoplasm of vallecula | Cancer | C100 |
| C10.1 | DX10 | Malignant neoplasm of anterior surface of epiglottis | Cancer | C101 |
| C10.2 | DX10 | Malignant neoplasm of lateral wall of oropharynx | Cancer | C102 |
| C10.3 | DX10 | Malignant neoplasm of posterior wall of oropharynx | Cancer | C103 |
| C10.4 | DX10 | Malignant neoplasm of branchial cleft | Cancer | C104 |
| C10.8 | DX10 | Malignant neoplasm of overlapping sites of oropharynx | Cancer | C108 |
| C10.9 | DX10 | Malignant neoplasm of oropharynx, unspecified | Cancer | C109 |
| C11 | DX10 | Malignant neoplasm of nasopharynx | Cancer | C11 |
| C11.0 | DX10 | Malignant neoplasm of superior wall of nasopharynx | Cancer | C110 |
| C11.1 | DX10 | Malignant neoplasm of posterior wall of nasopharynx | Cancer | C111 |
| C11.2 | DX10 | Malignant neoplasm of lateral wall of nasopharynx | Cancer | C112 |
| C11.3 | DX10 | Malignant neoplasm of anterior wall of nasopharynx | Cancer | C113 |

| | | | | |
|---|---|---|---|---|
| C11.8 | DX10 | Malignant neoplasm of overlapping sites of nasopharynx | Cancer | C118 |
| C11.9 | DX10 | Malignant neoplasm of nasopharynx, unspecified | Cancer | C119 |
| C12 | DX10 | Malignant neoplasm of pyriform sinus | Cancer | C12 |
| C13 | DX10 | Malignant neoplasm of hypopharynx | Cancer | C13 |
| C13.0 | DX10 | Malignant neoplasm of postcricoid region | Cancer | C130 |
| C13.1 | DX10 | Malig neoplasm of aryepiglottic fold, hypopharyngeal aspect | Cancer | C131 |
| C13.2 | DX10 | Malignant neoplasm of posterior wall of hypopharynx | Cancer | C132 |
| C14 | DX10 | Malig neoplasm of sites in the lip, oral cavity and pharynx | Cancer | C14 |
| C14.0 | DX10 | Malignant neoplasm of pharynx, unspecified | Cancer | C140 |
| C14.2 | DX10 | Malignant neoplasm of Waldeyer's ring | Cancer | C142 |
| C14.8 | DX10 | Malig neoplm of ovrlp sites of lip, oral cavity and pharynx | Cancer | C148 |
| C15 | DX10 | Malignant neoplasm of esophagus | Cancer | C15 |
| C15.3 | DX10 | Malignant neoplasm of upper third of esophagus | Cancer | C153 |
| C15.4 | DX10 | Malignant neoplasm of middle third of esophagus | Cancer | C154 |
| C15.5 | DX10 | Malignant neoplasm of lower third of esophagus | Cancer | C155 |
| C15.8 | DX10 | Malignant neoplasm of overlapping sites of esophagus | Cancer | C158 |
| C15.9 | DX10 | Malignant neoplasm of esophagus, unspecified | Cancer | C159 |
| C16 | DX10 | Malignant neoplasm of stomach | Cancer | C16 |
| C16.0 | DX10 | Malignant neoplasm of cardia | Cancer | C160 |
| C16.1 | DX10 | Malignant neoplasm of fundus of stomach | Cancer | C161 |
| C16.2 | DX10 | Malignant neoplasm of body of stomach | Cancer | C162 |
| C16.3 | DX10 | Malignant neoplasm of pyloric antrum | Cancer | C163 |
| C16.4 | DX10 | Malignant neoplasm of pylorus | Cancer | C164 |
| C16.5 | DX10 | Malignant neoplasm of lesser curvature of stomach, unsp | Cancer | C165 |
| C16.6 | DX10 | Malignant neoplasm of greater curvature of stomach, unsp | Cancer | C166 |
| C16.8 | DX10 | Malignant neoplasm of overlapping sites of stomach | Cancer | C168 |
| C16.9 | DX10 | Malignant neoplasm of stomach, unspecified | Cancer | C169 |
| C17 | DX10 | Malignant neoplasm of small intestine | Cancer | C17 |
| C17.0 | DX10 | Malignant neoplasm of duodenum | Cancer | C170 |
| C17.1 | DX10 | Malignant neoplasm of jejunum | Cancer | C171 |
| C17.2 | DX10 | Malignant neoplasm of ileum | Cancer | C172 |
| C17.3 | DX10 | Meckel's diverticulum, malignant | Cancer | C173 |
| C17.8 | DX10 | Malignant neoplasm of overlapping sites of small intestine | Cancer | C178 |
| C17.9 | DX10 | Malignant neoplasm of small intestine, unspecified | Cancer | C179 |
| C18 | DX10 | Malignant neoplasm of colon | Cancer | C18 |
| C18.0 | DX10 | Malignant neoplasm of cecum | Cancer | C180 |
| C18.1 | DX10 | Malignant neoplasm of appendix | Cancer | C181 |
| C18.2 | DX10 | Malignant neoplasm of ascending colon | Cancer | C182 |
| C18.3 | DX10 | Malignant neoplasm of hepatic flexure | Cancer | C183 |
| C18.4 | DX10 | Malignant neoplasm of transverse colon | Cancer | C184 |
| C18.5 | DX10 | Malignant neoplasm of splenic flexure | Cancer | C185 |
| C18.6 | DX10 | Malignant neoplasm of descending colon | Cancer | C186 |
| C18.7 | DX10 | Malignant neoplasm of sigmoid colon | Cancer | C187 |
| C18.8 | DX10 | Malignant neoplasm of overlapping sites of colon | Cancer | C188 |
| C18.9 | DX10 | Malignant neoplasm of colon, unspecified | Cancer | C189 |
| C19 | DX10 | Malignant neoplasm of rectosigmoid junction | Cancer | C19 |
| C20 | DX10 | Malignant neoplasm of rectum | Cancer | C20 |
| C21 | DX10 | Malignant neoplasm of anus and anal canal | Cancer | C21 |
| C21.0 | DX10 | Malignant neoplasm of anus, unspecified | Cancer | C210 |
| C21.1 | DX10 | Malignant neoplasm of anal canal | Cancer | C211 |
| C21.2 | DX10 | Malignant neoplasm of cloacogenic zone | Cancer | C212 |

| | | | | |
|---|---|---|---|---|
| C21.8 | DX10 | Malig neoplasm of ovrlp sites of rectum, anus and anal canal | Cancer | C218 |
| C22 | DX10 | Malignant neoplasm of liver and intrahepatic bile ducts | Cancer | C22 |
| C22.0 | DX10 | Liver cell carcinoma | Cancer | C220 |
| C22.1 | DX10 | Intrahepatic bile duct carcinoma | Cancer | C221 |
| C22.2 | DX10 | Hepatoblastoma | Cancer | C222 |
| C22.3 | DX10 | Angiosarcoma of liver | Cancer | C223 |
| C22.4 | DX10 | Other sarcomas of liver | Cancer | C224 |
| C22.7 | DX10 | Other specified carcinomas of liver | Cancer | C227 |
| C22.8 | DX10 | Malignant neoplasm of liver, primary, unspecified as to type | Cancer | C228 |
| C22.9 | DX10 | Malig neoplasm of liver, not specified as primary or sec | Cancer | C229 |
| C23 | DX10 | Malignant neoplasm of gallbladder | Cancer | C23 |
| C24 | DX10 | Malignant neoplasm of other and unsp parts of biliary tract | Cancer | C24 |
| C24.0 | DX10 | Malignant neoplasm of extrahepatic bile duct | Cancer | C240 |
| C24.1 | DX10 | Malignant neoplasm of ampulla of Vater | Cancer | C241 |
| C24.8 | DX10 | Malignant neoplasm of overlapping sites of biliary tract | Cancer | C248 |
| C24.9 | DX10 | Malignant neoplasm of biliary tract, unspecified | Cancer | C249 |
| C25 | DX10 | Malignant neoplasm of pancreas | Cancer | C25 |
| C25.0 | DX10 | Malignant neoplasm of head of pancreas | Cancer | C250 |
| C25.1 | DX10 | Malignant neoplasm of body of pancreas | Cancer | C251 |
| C25.2 | DX10 | Malignant neoplasm of tail of pancreas | Cancer | C252 |
| C25.3 | DX10 | Malignant neoplasm of pancreatic duct | Cancer | C253 |
| C25.4 | DX10 | Malignant neoplasm of endocrine pancreas | Cancer | C254 |
| C25.7 | DX10 | Malignant neoplasm of other parts of pancreas | Cancer | C257 |
| C25.8 | DX10 | Malignant neoplasm of overlapping sites of pancreas | Cancer | C258 |
| C25.9 | DX10 | Malignant neoplasm of pancreas, unspecified | Cancer | C259 |
| C26 | DX10 | Malignant neoplasm of other and ill-defined digestive organs | Cancer | C26 |
| C26.0 | DX10 | Malignant neoplasm of intestinal tract, part unspecified | Cancer | C260 |
| C26.1 | DX10 | Malignant neoplasm of spleen | Cancer | C261 |
| C26.9 | DX10 | Malignant neoplasm of ill-defined sites within the dgstv sys | Cancer | C269 |
| C30 | DX10 | Malignant neoplasm of nasal cavity and middle ear | Cancer | C30 |
| C30.0 | DX10 | Malignant neoplasm of nasal cavity | Cancer | C300 |
| C30.1 | DX10 | Malignant neoplasm of middle ear | Cancer | C301 |
| C31 | DX10 | Malignant neoplasm of accessory sinuses | Cancer | C31 |
| C31.0 | DX10 | Malignant neoplasm of maxillary sinus | Cancer | C310 |
| C31.1 | DX10 | Malignant neoplasm of ethmoidal sinus | Cancer | C311 |
| C31.2 | DX10 | Malignant neoplasm of frontal sinus | Cancer | C312 |
| C31.3 | DX10 | Malignant neoplasm of sphenoid sinus | Cancer | C313 |
| C31.8 | DX10 | Malignant neoplasm of overlapping sites of accessory sinuses | Cancer | C318 |
| C31.9 | DX10 | Malignant neoplasm of accessory sinus, unspecified | Cancer | C319 |
| C32 | DX10 | Malignant neoplasm of larynx | Cancer | C32 |
| C32.0 | DX10 | Malignant neoplasm of glottis | Cancer | C320 |
| C32.1 | DX10 | Malignant neoplasm of supraglottis | Cancer | C321 |
| C32.2 | DX10 | Malignant neoplasm of subglottis | Cancer | C322 |
| C32.3 | DX10 | Malignant neoplasm of laryngeal cartilage | Cancer | C323 |
| C32.8 | DX10 | Malignant neoplasm of overlapping sites of larynx | Cancer | C328 |
| C32.9 | DX10 | Malignant neoplasm of larynx, unspecified | Cancer | C329 |
| C33 | DX10 | Malignant neoplasm of trachea | Cancer | C33 |
| C34 | DX10 | Malignant neoplasm of bronchus and lung | Cancer | C34 |
| C34.0 | DX10 | Malignant neoplasm of main bronchus | Cancer | C340 |
| C34.00 | DX10 | Malignant neoplasm of unspecified main bronchus | Cancer | C3400 |
| C34.01 | DX10 | Malignant neoplasm of right main bronchus | Cancer | C3401 |
| C34.02 | DX10 | Malignant neoplasm of left main bronchus | Cancer | C3402 |
| C34.1 | DX10 | Malignant neoplasm of upper lobe, bronchus or lung | Cancer | C341 |

| | | | | |
|---|---|---|---|---|
| C34.10 | DX10 | Malignant neoplasm of upper lobe, unsp bronchus or lung | Cancer | C3410 |
| C34.11 | DX10 | Malignant neoplasm of upper lobe, right bronchus or lung | Cancer | C3411 |
| C34.12 | DX10 | Malignant neoplasm of upper lobe, left bronchus or lung | Cancer | C3412 |
| C34.2 | DX10 | Malignant neoplasm of middle lobe, bronchus or lung | Cancer | C342 |
| C34.3 | DX10 | Malignant neoplasm of lower lobe, bronchus or lung | Cancer | C343 |
| C34.30 | DX10 | Malignant neoplasm of lower lobe, unsp bronchus or lung | Cancer | C3430 |
| C34.31 | DX10 | Malignant neoplasm of lower lobe, right bronchus or lung | Cancer | C3431 |
| C34.32 | DX10 | Malignant neoplasm of lower lobe, left bronchus or lung | Cancer | C3432 |
| C34.8 | DX10 | Malignant neoplasm of overlapping sites of bronchus and lung | Cancer | C348 |
| C34.80 | DX10 | Malignant neoplasm of ovrlp sites of unsp bronchus and lung | Cancer | C3480 |
| C34.81 | DX10 | Malignant neoplasm of ovrlp sites of right bronchus and lung | Cancer | C3481 |
| C34.82 | DX10 | Malignant neoplasm of ovrlp sites of left bronchus and lung | Cancer | C3482 |
| C34.9 | DX10 | Malignant neoplasm of unspecified part of bronchus or lung | Cancer | C349 |
| C34.90 | DX10 | Malignant neoplasm of unsp part of unsp bronchus or lung | Cancer | C3490 |
| C34.91 | DX10 | Malignant neoplasm of unsp part of right bronchus or lung | Cancer | C3491 |
| C34.92 | DX10 | Malignant neoplasm of unsp part of left bronchus or lung | Cancer | C3492 |
| C37 | DX10 | Malignant neoplasm of thymus | Cancer | C37 |
| C38 | DX10 | Malignant neoplasm of heart, mediastinum and pleura | Cancer | C38 |
| C38.0 | DX10 | Malignant neoplasm of heart | Cancer | C380 |
| C38.1 | DX10 | Malignant neoplasm of anterior mediastinum | Cancer | C381 |
| C38.2 | DX10 | Malignant neoplasm of posterior mediastinum | Cancer | C382 |
| C38.3 | DX10 | Malignant neoplasm of mediastinum, part unspecified | Cancer | C383 |
| C38.4 | DX10 | Malignant neoplasm of pleura | Cancer | C384 |
| C38.8 | DX10 | Malig neoplm of ovrlp sites of heart, mediastinum and pleura | Cancer | C388 |
| C39 | DX10 | Malig neoplm of sites in the resp sys and intrathorac organs | Cancer | C39 |
| C39.0 | DX10 | Malignant neoplasm of upper respiratory tract, part unsp | Cancer | C390 |
| C39.9 | DX10 | Malignant neoplasm of lower respiratory tract, part unsp | Cancer | C399 |
| C40 | DX10 | Malignant neoplasm of bone and articular cartilage of limbs | Cancer | C40 |
| C40.0 | DX10 | Malignant neoplasm of scapula and long bones of upper limb | Cancer | C400 |
| C40.00 | DX10 | Malig neoplasm of scapula and long bones of unsp upper limb | Cancer | C4000 |
| C40.01 | DX10 | Malig neoplasm of scapula and long bones of right upper limb | Cancer | C4001 |
| C40.02 | DX10 | Malig neoplasm of scapula and long bones of left upper limb | Cancer | C4002 |
| C40.1 | DX10 | Malignant neoplasm of short bones of upper limb | Cancer | C401 |
| C40.10 | DX10 | Malignant neoplasm of short bones of unspecified upper limb | Cancer | C4010 |
| C40.11 | DX10 | Malignant neoplasm of short bones of right upper limb | Cancer | C4011 |
| C40.12 | DX10 | Malignant neoplasm of short bones of left upper limb | Cancer | C4012 |

| | | | | |
|---|---|---|---|---|
| C40.2 | DX10 | Malignant neoplasm of long bones of lower limb | Cancer | C402 |
| C40.20 | DX10 | Malignant neoplasm of long bones of unspecified lower limb | Cancer | C4020 |
| C40.21 | DX10 | Malignant neoplasm of long bones of right lower limb | Cancer | C4021 |
| C40.22 | DX10 | Malignant neoplasm of long bones of left lower limb | Cancer | C4022 |
| C40.3 | DX10 | Malignant neoplasm of short bones of lower limb | Cancer | C403 |
| C40.30 | DX10 | Malignant neoplasm of short bones of unspecified lower limb | Cancer | C4030 |
| C40.31 | DX10 | Malignant neoplasm of short bones of right lower limb | Cancer | C4031 |
| C40.32 | DX10 | Malignant neoplasm of short bones of left lower limb | Cancer | C4032 |
| C40.8 | DX10 | Malig neoplasm of ovrlp sites of bone/artic cartl of limb | Cancer | C408 |
| C40.80 | DX10 | Malig neoplm of ovrlp sites of bone/artic cartl of unsp limb | Cancer | C4080 |
| C40.81 | DX10 | Malig neoplm of ovrlp sites of bone/artic cartl of r limb | Cancer | C4081 |
| C40.82 | DX10 | Malig neoplm of ovrlp sites of bone/artic cartl of left limb | Cancer | C4082 |
| C40.9 | DX10 | Malignant neoplasm of unsp bones and artic cartilage of limb | Cancer | C409 |
| C40.90 | DX10 | Malig neoplasm of unsp bones and artic cartlg of unsp limb | Cancer | C4090 |
| C40.91 | DX10 | Malig neoplasm of unsp bones and artic cartlg of right limb | Cancer | C4091 |
| C40.92 | DX10 | Malig neoplasm of unsp bones and artic cartlg of left limb | Cancer | C4092 |
| C41 | DX10 | Malignant neoplasm of bone/artic cartl of and unsp sites | Cancer | C41 |
| C41.0 | DX10 | Malignant neoplasm of bones of skull and face | Cancer | C410 |
| C41.1 | DX10 | Malignant neoplasm of mandible | Cancer | C411 |
| C41.2 | DX10 | Malignant neoplasm of vertebral column | Cancer | C412 |
| C41.3 | DX10 | Malignant neoplasm of ribs, sternum and clavicle | Cancer | C413 |
| C41.4 | DX10 | Malignant neoplasm of pelvic bones, sacrum and coccyx | Cancer | C414 |
| C41.9 | DX10 | Malignant neoplasm of bone and articular cartilage, unsp | Cancer | C419 |
| C43 | DX10 | Malignant melanoma of skin | Cancer | C43 |
| C43.0 | DX10 | Malignant melanoma of lip | Cancer | C430 |
| C43.1 | DX10 | Malignant melanoma of eyelid, including canthus | Cancer | C431 |
| C43.10 | DX10 | Malignant melanoma of unspecified eyelid, including canthus | Cancer | C4310 |
| C43.11 | DX10 | Malignant melanoma of right eyelid, including canthus | Cancer | C4311 |
| C43.111 | DX10 | | Cancer | C43111 |
| C43.112 | DX10 | | Cancer | C43112 |
| C43.12 | DX10 | Malignant melanoma of left eyelid, including canthus | Cancer | C4312 |
| C43.121 | DX10 | | Cancer | C43121 |
| C43.122 | DX10 | | Cancer | C43122 |
| C43.2 | DX10 | Malignant melanoma of ear and external auricular canal | Cancer | C432 |
| C43.20 | DX10 | Malignant melanoma of unsp ear and external auricular canal | Cancer | C4320 |
| C43.21 | DX10 | Malignant melanoma of right ear and external auricular canal | Cancer | C4321 |
| C43.22 | DX10 | Malignant melanoma of left ear and external auricular canal | Cancer | C4322 |
| C43.3 | DX10 | Malignant melanoma of other and unspecified parts of face | Cancer | C433 |
| C43.30 | DX10 | Malignant melanoma of unspecified part of face | Cancer | C4330 |
| C43.31 | DX10 | Malignant melanoma of nose | Cancer | C4331 |
| C43.39 | DX10 | Malignant melanoma of other parts of face | Cancer | C4339 |
| C43.4 | DX10 | Malignant melanoma of scalp and neck | Cancer | C434 |
| C43.5 | DX10 | Malignant melanoma of trunk | Cancer | C435 |

| | | | | |
|---|---|---|---|---|
| C43.51 | DX10 | Malignant melanoma of anal skin | Cancer | C4351 |
| C43.52 | DX10 | Malignant melanoma of skin of breast | Cancer | C4352 |
| C43.59 | DX10 | Malignant melanoma of other part of trunk | Cancer | C4359 |
| C43.6 | DX10 | Malignant melanoma of upper limb, including shoulder | Cancer | C436 |
| C43.60 | DX10 | Malignant melanoma of unsp upper limb, including shoulder | Cancer | C4360 |
| C43.61 | DX10 | Malignant melanoma of right upper limb, including shoulder | Cancer | C4361 |
| C43.62 | DX10 | Malignant melanoma of left upper limb, including shoulder | Cancer | C4362 |
| C43.7 | DX10 | Malignant melanoma of lower limb, including hip | Cancer | C437 |
| C43.70 | DX10 | Malignant melanoma of unspecified lower limb, including hip | Cancer | C4370 |
| C43.71 | DX10 | Malignant melanoma of right lower limb, including hip | Cancer | C4371 |
| C43.72 | DX10 | Malignant melanoma of left lower limb, including hip | Cancer | C4372 |
| C43.8 | DX10 | Malignant melanoma of overlapping sites of skin | Cancer | C438 |
| C43.9 | DX10 | Malignant melanoma of skin, unspecified | Cancer | C439 |
| C45 | DX10 | Mesothelioma | Cancer | C45 |
| C45.0 | DX10 | Mesothelioma of pleura | Cancer | C450 |
| C45.1 | DX10 | Mesothelioma of peritoneum | Cancer | C451 |
| C45.2 | DX10 | Mesothelioma of pericardium | Cancer | C452 |
| C45.7 | DX10 | Mesothelioma of other sites | Cancer | C457 |
| C45.9 | DX10 | Mesothelioma, unspecified | Cancer | C459 |
| C46 | DX10 | Kaposi's sarcoma | Cancer | C46 |
| C46.0 | DX10 | Kaposi's sarcoma of skin | Cancer | C460 |
| C46.1 | DX10 | Kaposi's sarcoma of soft tissue | Cancer | C461 |
| C46.2 | DX10 | Kaposi's sarcoma of palate | Cancer | C462 |
| C46.3 | DX10 | Kaposi's sarcoma of lymph nodes | Cancer | C463 |
| C46.4 | DX10 | Kaposi's sarcoma of gastrointestinal sites | Cancer | C464 |
| C46.5 | DX10 | Kaposi's sarcoma of lung | Cancer | C465 |
| C46.50 | DX10 | Kaposi's sarcoma of unspecified lung | Cancer | C4650 |
| C46.51 | DX10 | Kaposi's sarcoma of right lung | Cancer | C4651 |
| C46.52 | DX10 | Kaposi's sarcoma of left lung | Cancer | C4652 |
| C46.7 | DX10 | Kaposi's sarcoma of other sites | Cancer | C467 |
| C46.9 | DX10 | Kaposi's sarcoma, unspecified | Cancer | C469 |
| C47 | DX10 | Malignant neoplasm of prph nerves and autonomic nervous sys | Cancer | C47 |
| C47.0 | DX10 | Malignant neoplasm of prph nerves of head, face and neck | Cancer | C470 |
| C47.1 | DX10 | Malig neoplasm of prph nerves of upper limb, inc shoulder | Cancer | C471 |
| C47.10 | DX10 | Malig neoplm of prph nerves of unsp upper limb, inc shoulder | Cancer | C4710 |
| C47.11 | DX10 | Malig neoplm of prph nerves of right upper limb, inc shldr | Cancer | C4711 |
| C47.12 | DX10 | Malig neoplm of prph nerves of left upper limb, inc shoulder | Cancer | C4712 |
| C47.2 | DX10 | Malignant neoplasm of prph nerves of lower limb, inc hip | Cancer | C472 |
| C47.20 | DX10 | Malig neoplasm of prph nerves of unsp lower limb, inc hip | Cancer | C4720 |
| C47.21 | DX10 | Malig neoplasm of prph nerves of right lower limb, inc hip | Cancer | C4721 |
| C47.22 | DX10 | Malig neoplasm of prph nerves of left lower limb, inc hip | Cancer | C4722 |
| C47.3 | DX10 | Malignant neoplasm of peripheral nerves of thorax | Cancer | C473 |
| C47.4 | DX10 | Malignant neoplasm of peripheral nerves of abdomen | Cancer | C474 |
| C47.5 | DX10 | Malignant neoplasm of peripheral nerves of pelvis | Cancer | C475 |
| C47.6 | DX10 | Malignant neoplasm of peripheral nerves of trunk, unsp | Cancer | C476 |

| | | | | |
|---|---|---|---|---|
| C47.8 | DX10 | Malig neoplm of ovrlp sites of prph nrv and autonm nrv sys | Cancer | C478 |
| C47.9 | DX10 | Malig neoplasm of prph nerves and autonm nervous sys, unsp | Cancer | C479 |
| C48 | DX10 | Malignant neoplasm of retroperitoneum and peritoneum | Cancer | C48 |
| C48.0 | DX10 | Malignant neoplasm of retroperitoneum | Cancer | C480 |
| C48.1 | DX10 | Malignant neoplasm of specified parts of peritoneum | Cancer | C481 |
| C48.2 | DX10 | Malignant neoplasm of peritoneum, unspecified | Cancer | C482 |
| C48.8 | DX10 | Malig neoplasm of ovrlp sites of retroperiton and peritoneum | Cancer | C488 |
| C49 | DX10 | Malignant neoplasm of other connective and soft tissue | Cancer | C49 |
| C49.0 | DX10 | Malig neoplm of conn and soft tissue of head, face and neck | Cancer | C490 |
| C49.1 | DX10 | Malig neoplm of conn and soft tiss of upper limb, inc shldr | Cancer | C491 |
| C49.10 | DX10 | Malig neoplm of conn & soft tiss of unsp upr lmb, inc shldr | Cancer | C4910 |
| C49.11 | DX10 | Malig neoplm of conn and soft tiss of r upr limb, inc shldr | Cancer | C4911 |
| C49.12 | DX10 | Malig neoplm of conn and soft tiss of l upr limb, inc shldr | Cancer | C4912 |
| C49.2 | DX10 | Malig neoplm of conn and soft tissue of lower limb, inc hip | Cancer | C492 |
| C49.20 | DX10 | Malig neoplm of conn and soft tiss of unsp low limb, inc hip | Cancer | C4920 |
| C49.21 | DX10 | Malig neoplm of conn and soft tiss of r low limb, inc hip | Cancer | C4921 |
| C49.22 | DX10 | Malig neoplm of conn and soft tiss of left low limb, inc hip | Cancer | C4922 |
| C49.3 | DX10 | Malignant neoplasm of connective and soft tissue of thorax | Cancer | C493 |
| C49.4 | DX10 | Malignant neoplasm of connective and soft tissue of abdomen | Cancer | C494 |
| C49.5 | DX10 | Malignant neoplasm of connective and soft tissue of pelvis | Cancer | C495 |
| C49.6 | DX10 | Malignant neoplasm of conn and soft tissue of trunk, unsp | Cancer | C496 |
| C49.8 | DX10 | Malignant neoplasm of ovrlp sites of conn and soft tissue | Cancer | C498 |
| C49.9 | DX10 | Malignant neoplasm of connective and soft tissue, unsp | Cancer | C499 |
| C49.A | DX10 | Gastrointestinal stromal tumor | Cancer | C49A |
| C49.A0 | DX10 | Gastrointestinal stromal tumor, unspecified site | Cancer | C49A0 |
| C49.A1 | DX10 | Gastrointestinal stromal tumor of esophagus | Cancer | C49A1 |
| C49.A2 | DX10 | Gastrointestinal stromal tumor of stomach | Cancer | C49A2 |
| C49.A3 | DX10 | Gastrointestinal stromal tumor of small intestine | Cancer | C49A3 |
| C49.A4 | DX10 | Gastrointestinal stromal tumor of large intestine | Cancer | C49A4 |
| C49.A5 | DX10 | Gastrointestinal stromal tumor of rectum | Cancer | C49A5 |
| C49.A9 | DX10 | Gastrointestinal stromal tumor of other sites | Cancer | C49A9 |
| C50 | DX10 | Malignant neoplasm of breast | Cancer | C50 |
| C50.0 | DX10 | Malignant neoplasm of nipple and areola | Cancer | C500 |
| C50.01 | DX10 | Malignant neoplasm of nipple and areola, female | Cancer | C5001 |
| C50.011 | DX10 | Malignant neoplasm of nipple and areola, right female breast | Cancer | C50011 |
| C50.012 | DX10 | Malignant neoplasm of nipple and areola, left female breast | Cancer | C50012 |
| C50.019 | DX10 | Malignant neoplasm of nipple and areola, unsp female breast | Cancer | C50019 |
| C50.02 | DX10 | Malignant neoplasm of nipple and areola, male | Cancer | C5002 |

| | | | | |
|---|---|---|---|---|
| C50.021 | DX10 | Malignant neoplasm of nipple and areola, right male breast | Cancer | C50021 |
| C50.022 | DX10 | Malignant neoplasm of nipple and areola, left male breast | Cancer | C50022 |
| C50.029 | DX10 | Malignant neoplasm of nipple and areola, unsp male breast | Cancer | C50029 |
| C50.1 | DX10 | Malignant neoplasm of central portion of breast | Cancer | C501 |
| C50.11 | DX10 | Malignant neoplasm of central portion of breast, female | Cancer | C5011 |
| C50.111 | DX10 | Malignant neoplasm of central portion of right female breast | Cancer | C50111 |
| C50.112 | DX10 | Malignant neoplasm of central portion of left female breast | Cancer | C50112 |
| C50.119 | DX10 | Malignant neoplasm of central portion of unsp female breast | Cancer | C50119 |
| C50.12 | DX10 | Malignant neoplasm of central portion of breast, male | Cancer | C5012 |
| C50.121 | DX10 | Malignant neoplasm of central portion of right male breast | Cancer | C50121 |
| C50.122 | DX10 | Malignant neoplasm of central portion of left male breast | Cancer | C50122 |
| C50.129 | DX10 | Malignant neoplasm of central portion of unsp male breast | Cancer | C50129 |
| C50.2 | DX10 | Malignant neoplasm of upper-inner quadrant of breast | Cancer | C502 |
| C50.21 | DX10 | Malignant neoplasm of upper-inner quadrant of breast, female | Cancer | C5021 |
| C50.211 | DX10 | Malig neoplm of upper-inner quadrant of right female breast | Cancer | C50211 |
| C50.212 | DX10 | Malig neoplasm of upper-inner quadrant of left female breast | Cancer | C50212 |
| C50.219 | DX10 | Malig neoplasm of upper-inner quadrant of unsp female breast | Cancer | C50219 |
| C50.22 | DX10 | Malignant neoplasm of upper-inner quadrant of breast, male | Cancer | C5022 |
| C50.221 | DX10 | Malig neoplasm of upper-inner quadrant of right male breast | Cancer | C50221 |
| C50.222 | DX10 | Malig neoplasm of upper-inner quadrant of left male breast | Cancer | C50222 |
| C50.229 | DX10 | Malig neoplasm of upper-inner quadrant of unsp male breast | Cancer | C50229 |
| C50.3 | DX10 | Malignant neoplasm of lower-inner quadrant of breast | Cancer | C503 |
| C50.31 | DX10 | Malignant neoplasm of lower-inner quadrant of breast, female | Cancer | C5031 |
| C50.311 | DX10 | Malig neoplm of lower-inner quadrant of right female breast | Cancer | C50311 |
| C50.312 | DX10 | Malig neoplasm of lower-inner quadrant of left female breast | Cancer | C50312 |
| C50.319 | DX10 | Malig neoplasm of lower-inner quadrant of unsp female breast | Cancer | C50319 |
| C50.32 | DX10 | Malignant neoplasm of lower-inner quadrant of breast, male | Cancer | C5032 |
| C50.321 | DX10 | Malig neoplasm of lower-inner quadrant of right male breast | Cancer | C50321 |
| C50.322 | DX10 | Malig neoplasm of lower-inner quadrant of left male breast | Cancer | C50322 |
| C50.329 | DX10 | Malig neoplasm of lower-inner quadrant of unsp male breast | Cancer | C50329 |
| C50.4 | DX10 | Malignant neoplasm of upper-outer quadrant of breast | Cancer | C504 |
| C50.41 | DX10 | Malignant neoplasm of upper-outer quadrant of breast, female | Cancer | C5041 |

| | | | | |
|---|---|---|---|---|
| C50.411 | DX10 | Malig neoplm of upper-outer quadrant of right female breast | Cancer | C50411 |
| C50.412 | DX10 | Malig neoplasm of upper-outer quadrant of left female breast | Cancer | C50412 |
| C50.419 | DX10 | Malig neoplasm of upper-outer quadrant of unsp female breast | Cancer | C50419 |
| C50.42 | DX10 | Malignant neoplasm of upper-outer quadrant of breast, male | Cancer | C5042 |
| C50.421 | DX10 | Malig neoplasm of upper-outer quadrant of right male breast | Cancer | C50421 |
| C50.422 | DX10 | Malig neoplasm of upper-outer quadrant of left male breast | Cancer | C50422 |
| C50.429 | DX10 | Malig neoplasm of upper-outer quadrant of unsp male breast | Cancer | C50429 |
| C50.5 | DX10 | Malignant neoplasm of lower-outer quadrant of breast | Cancer | C505 |
| C50.51 | DX10 | Malignant neoplasm of lower-outer quadrant of breast, female | Cancer | C5051 |
| C50.511 | DX10 | Malig neoplm of lower-outer quadrant of right female breast | Cancer | C50511 |
| C50.512 | DX10 | Malig neoplasm of lower-outer quadrant of left female breast | Cancer | C50512 |
| C50.519 | DX10 | Malig neoplasm of lower-outer quadrant of unsp female breast | Cancer | C50519 |
| C50.52 | DX10 | Malignant neoplasm of lower-outer quadrant of breast, male | Cancer | C5052 |
| C50.521 | DX10 | Malig neoplasm of lower-outer quadrant of right male breast | Cancer | C50521 |
| C50.522 | DX10 | Malig neoplasm of lower-outer quadrant of left male breast | Cancer | C50522 |
| C50.529 | DX10 | Malig neoplasm of lower-outer quadrant of unsp male breast | Cancer | C50529 |
| C50.6 | DX10 | Malignant neoplasm of axillary tail of breast | Cancer | C506 |
| C50.61 | DX10 | Malignant neoplasm of axillary tail of breast, female | Cancer | C5061 |
| C50.611 | DX10 | Malignant neoplasm of axillary tail of right female breast | Cancer | C50611 |
| C50.612 | DX10 | Malignant neoplasm of axillary tail of left female breast | Cancer | C50612 |
| C50.619 | DX10 | Malignant neoplasm of axillary tail of unsp female breast | Cancer | C50619 |
| C50.62 | DX10 | Malignant neoplasm of axillary tail of breast, male | Cancer | C5062 |
| C50.621 | DX10 | Malignant neoplasm of axillary tail of right male breast | Cancer | C50621 |
| C50.622 | DX10 | Malignant neoplasm of axillary tail of left male breast | Cancer | C50622 |
| C50.629 | DX10 | Malignant neoplasm of axillary tail of unsp male breast | Cancer | C50629 |
| C50.8 | DX10 | Malignant neoplasm of overlapping sites of breast | Cancer | C508 |
| C50.81 | DX10 | Malignant neoplasm of overlapping sites of breast, female | Cancer | C5081 |
| C50.811 | DX10 | Malignant neoplasm of ovrlp sites of right female breast | Cancer | C50811 |
| C50.812 | DX10 | Malignant neoplasm of ovrlp sites of left female breast | Cancer | C50812 |
| C50.819 | DX10 | Malignant neoplasm of ovrlp sites of unsp female breast | Cancer | C50819 |
| C50.82 | DX10 | Malignant neoplasm of overlapping sites of breast, male | Cancer | C5082 |
| C50.821 | DX10 | Malignant neoplasm of overlapping sites of right male breast | Cancer | C50821 |
| C50.822 | DX10 | Malignant neoplasm of overlapping sites of left male breast | Cancer | C50822 |
| C50.829 | DX10 | Malignant neoplasm of overlapping sites of unsp male breast | Cancer | C50829 |
| C50.9 | DX10 | Malignant neoplasm of breast of unspecified site | Cancer | C509 |
| C50.91 | DX10 | Malignant neoplasm of breast of unspecified site, female | Cancer | C5091 |
| C50.911 | DX10 | Malignant neoplasm of unsp site of right female breast | Cancer | C50911 |

| | | | | |
|---|---|---|---|---|
| C50.912 | DX10 | Malignant neoplasm of unspecified site of left female breast | Cancer | C50912 |
| C50.919 | DX10 | Malignant neoplasm of unsp site of unspecified female breast | Cancer | C50919 |
| C50.92 | DX10 | Malignant neoplasm of breast of unspecified site, male | Cancer | C5092 |
| C50.921 | DX10 | Malignant neoplasm of unspecified site of right male breast | Cancer | C50921 |
| C50.922 | DX10 | Malignant neoplasm of unspecified site of left male breast | Cancer | C50922 |
| C50.929 | DX10 | Malignant neoplasm of unsp site of unspecified male breast | Cancer | C50929 |
| C51 | DX10 | Malignant neoplasm of vulva | Cancer | C51 |
| C51.0 | DX10 | Malignant neoplasm of labium majus | Cancer | C510 |
| C51.1 | DX10 | Malignant neoplasm of labium minus | Cancer | C511 |
| C51.2 | DX10 | Malignant neoplasm of clitoris | Cancer | C512 |
| C51.8 | DX10 | Malignant neoplasm of overlapping sites of vulva | Cancer | C518 |
| C51.9 | DX10 | Malignant neoplasm of vulva, unspecified | Cancer | C519 |
| C52 | DX10 | Malignant neoplasm of vagina | Cancer | C52 |
| C53 | DX10 | Malignant neoplasm of cervix uteri | Cancer | C53 |
| C53.0 | DX10 | Malignant neoplasm of endocervix | Cancer | C530 |
| C53.1 | DX10 | Malignant neoplasm of exocervix | Cancer | C531 |
| C53.8 | DX10 | Malignant neoplasm of overlapping sites of cervix uteri | Cancer | C538 |
| C53.9 | DX10 | Malignant neoplasm of cervix uteri, unspecified | Cancer | C539 |
| C54 | DX10 | Malignant neoplasm of corpus uteri | Cancer | C54 |
| C54.0 | DX10 | Malignant neoplasm of isthmus uteri | Cancer | C540 |
| C54.1 | DX10 | Malignant neoplasm of endometrium | Cancer | C541 |
| C54.2 | DX10 | Malignant neoplasm of myometrium | Cancer | C542 |
| C54.3 | DX10 | Malignant neoplasm of fundus uteri | Cancer | C543 |
| C54.8 | DX10 | Malignant neoplasm of overlapping sites of corpus uteri | Cancer | C548 |
| C54.9 | DX10 | Malignant neoplasm of corpus uteri, unspecified | Cancer | C549 |
| C55 | DX10 | Malignant neoplasm of uterus, part unspecified | Cancer | C55 |
| C56 | DX10 | Malignant neoplasm of ovary | Cancer | C56 |
| C56.1 | DX10 | Malignant neoplasm of right ovary | Cancer | C561 |
| C56.2 | DX10 | Malignant neoplasm of left ovary | Cancer | C562 |
| C56.9 | DX10 | Malignant neoplasm of unspecified ovary | Cancer | C569 |
| C57 | DX10 | Malignant neoplasm of other and unsp female genital organs | Cancer | C57 |
| C57.0 | DX10 | Malignant neoplasm of fallopian tube | Cancer | C570 |
| C57.00 | DX10 | Malignant neoplasm of unspecified fallopian tube | Cancer | C5700 |
| C57.01 | DX10 | Malignant neoplasm of right fallopian tube | Cancer | C5701 |
| C57.02 | DX10 | Malignant neoplasm of left fallopian tube | Cancer | C5702 |
| C57.1 | DX10 | Malignant neoplasm of broad ligament | Cancer | C571 |
| C57.10 | DX10 | Malignant neoplasm of unspecified broad ligament | Cancer | C5710 |
| C57.11 | DX10 | Malignant neoplasm of right broad ligament | Cancer | C5711 |
| C57.12 | DX10 | Malignant neoplasm of left broad ligament | Cancer | C5712 |
| C57.2 | DX10 | Malignant neoplasm of round ligament | Cancer | C572 |
| C57.20 | DX10 | Malignant neoplasm of unspecified round ligament | Cancer | C5720 |
| C57.21 | DX10 | Malignant neoplasm of right round ligament | Cancer | C5721 |
| C57.22 | DX10 | Malignant neoplasm of left round ligament | Cancer | C5722 |
| C57.3 | DX10 | Malignant neoplasm of parametrium | Cancer | C573 |
| C57.4 | DX10 | Malignant neoplasm of uterine adnexa, unspecified | Cancer | C574 |
| C57.7 | DX10 | Malignant neoplasm of other specified female genital organs | Cancer | C577 |
| C57.8 | DX10 | Malignant neoplasm of ovrlp sites of female genital organs | Cancer | C578 |
| C57.9 | DX10 | Malignant neoplasm of female genital organ, unspecified | Cancer | C579 |
| C58 | DX10 | Malignant neoplasm of placenta | Cancer | C58 |
| C60 | DX10 | Malignant neoplasm of penis | Cancer | C60 |

| | | | | |
|---|---|---|---|---|
| C60.0 | DX10 | Malignant neoplasm of prepuce | Cancer | C600 |
| C60.1 | DX10 | Malignant neoplasm of glans penis | Cancer | C601 |
| C60.2 | DX10 | Malignant neoplasm of body of penis | Cancer | C602 |
| C60.8 | DX10 | Malignant neoplasm of overlapping sites of penis | Cancer | C608 |
| C60.9 | DX10 | Malignant neoplasm of penis, unspecified | Cancer | C609 |
| C61 | DX10 | Malignant neoplasm of prostate | Cancer | C61 |
| C62 | DX10 | Malignant neoplasm of testis | Cancer | C62 |
| C62.0 | DX10 | Malignant neoplasm of undescended testis | Cancer | C620 |
| C62.00 | DX10 | Malignant neoplasm of unspecified undescended testis | Cancer | C6200 |
| C62.01 | DX10 | Malignant neoplasm of undescended right testis | Cancer | C6201 |
| C62.02 | DX10 | Malignant neoplasm of undescended left testis | Cancer | C6202 |
| C62.1 | DX10 | Malignant neoplasm of descended testis | Cancer | C621 |
| C62.10 | DX10 | Malignant neoplasm of unspecified descended testis | Cancer | C6210 |
| C62.11 | DX10 | Malignant neoplasm of descended right testis | Cancer | C6211 |
| C62.12 | DX10 | Malignant neoplasm of descended left testis | Cancer | C6212 |
| C62.9 | DX10 | Malignant neoplasm of testis, unsp descended or undescended | Cancer | C629 |
| C62.90 | DX10 | Malig neoplasm of unsp testis, unsp descended or undescended | Cancer | C6290 |
| C62.91 | DX10 | Malig neoplm of right testis, unsp descended or undescended | Cancer | C6291 |
| C62.92 | DX10 | Malig neoplasm of left testis, unsp descended or undescended | Cancer | C6292 |
| C63 | DX10 | Malignant neoplasm of other and unsp male genital organs | Cancer | C63 |
| C63.0 | DX10 | Malignant neoplasm of epididymis | Cancer | C630 |
| C63.00 | DX10 | Malignant neoplasm of unspecified epididymis | Cancer | C6300 |
| C63.01 | DX10 | Malignant neoplasm of right epididymis | Cancer | C6301 |
| C63.02 | DX10 | Malignant neoplasm of left epididymis | Cancer | C6302 |
| C63.1 | DX10 | Malignant neoplasm of spermatic cord | Cancer | C631 |
| C63.10 | DX10 | Malignant neoplasm of unspecified spermatic cord | Cancer | C6310 |
| C63.11 | DX10 | Malignant neoplasm of right spermatic cord | Cancer | C6311 |
| C63.12 | DX10 | Malignant neoplasm of left spermatic cord | Cancer | C6312 |
| C63.2 | DX10 | Malignant neoplasm of scrotum | Cancer | C632 |
| C63.7 | DX10 | Malignant neoplasm of other specified male genital organs | Cancer | C637 |
| C63.8 | DX10 | Malignant neoplasm of ovrlp sites of male genital organs | Cancer | C638 |
| C63.9 | DX10 | Malignant neoplasm of male genital organ, unspecified | Cancer | C639 |
| C64 | DX10 | Malignant neoplasm of kidney, except renal pelvis | Cancer | C64 |
| C64.1 | DX10 | Malignant neoplasm of right kidney, except renal pelvis | Cancer | C641 |
| C64.2 | DX10 | Malignant neoplasm of left kidney, except renal pelvis | Cancer | C642 |
| C64.9 | DX10 | Malignant neoplasm of unsp kidney, except renal pelvis | Cancer | C649 |
| C65 | DX10 | Malignant neoplasm of renal pelvis | Cancer | C65 |
| C65.1 | DX10 | Malignant neoplasm of right renal pelvis | Cancer | C651 |
| C65.2 | DX10 | Malignant neoplasm of left renal pelvis | Cancer | C652 |
| C65.9 | DX10 | Malignant neoplasm of unspecified renal pelvis | Cancer | C659 |
| C66 | DX10 | Malignant neoplasm of ureter | Cancer | C66 |
| C66.1 | DX10 | Malignant neoplasm of right ureter | Cancer | C661 |
| C66.2 | DX10 | Malignant neoplasm of left ureter | Cancer | C662 |
| C66.9 | DX10 | Malignant neoplasm of unspecified ureter | Cancer | C669 |
| C67 | DX10 | Malignant neoplasm of bladder | Cancer | C67 |
| C67.0 | DX10 | Malignant neoplasm of trigone of bladder | Cancer | C670 |
| C67.1 | DX10 | Malignant neoplasm of dome of bladder | Cancer | C671 |
| C67.2 | DX10 | Malignant neoplasm of lateral wall of bladder | Cancer | C672 |
| C67.3 | DX10 | Malignant neoplasm of anterior wall of bladder | Cancer | C673 |
| C67.4 | DX10 | Malignant neoplasm of posterior wall of bladder | Cancer | C674 |
| C67.5 | DX10 | Malignant neoplasm of bladder neck | Cancer | C675 |
| C67.6 | DX10 | Malignant neoplasm of ureteric orifice | Cancer | C676 |

| | | | | |
|---|---|---|---|---|
| C67.7 | DX10 | Malignant neoplasm of urachus | Cancer | C677 |
| C67.8 | DX10 | Malignant neoplasm of overlapping sites of bladder | Cancer | C678 |
| C67.9 | DX10 | Malignant neoplasm of bladder, unspecified | Cancer | C679 |
| C68 | DX10 | Malignant neoplasm of other and unspecified urinary organs | Cancer | C68 |
| C68.0 | DX10 | Malignant neoplasm of urethra | Cancer | C680 |
| C68.1 | DX10 | Malignant neoplasm of paraurethral glands | Cancer | C681 |
| C68.2 | DX10 | | Cancer | C682 |
| C68.8 | DX10 | Malignant neoplasm of overlapping sites of urinary organs | Cancer | C688 |
| C68.9 | DX10 | Malignant neoplasm of urinary organ, unspecified | Cancer | C689 |
| C69 | DX10 | Malignant neoplasm of eye and adnexa | Cancer | C69 |
| C69.0 | DX10 | Malignant neoplasm of conjunctiva | Cancer | C690 |
| C69.00 | DX10 | Malignant neoplasm of unspecified conjunctiva | Cancer | C6900 |
| C69.01 | DX10 | Malignant neoplasm of right conjunctiva | Cancer | C6901 |
| C69.02 | DX10 | Malignant neoplasm of left conjunctiva | Cancer | C6902 |
| C69.1 | DX10 | Malignant neoplasm of cornea | Cancer | C691 |
| C69.10 | DX10 | Malignant neoplasm of unspecified cornea | Cancer | C6910 |
| C69.11 | DX10 | Malignant neoplasm of right cornea | Cancer | C6911 |
| C69.12 | DX10 | Malignant neoplasm of left cornea | Cancer | C6912 |
| C69.2 | DX10 | Malignant neoplasm of retina | Cancer | C692 |
| C69.20 | DX10 | Malignant neoplasm of unspecified retina | Cancer | C6920 |
| C69.21 | DX10 | Malignant neoplasm of right retina | Cancer | C6921 |
| C69.22 | DX10 | Malignant neoplasm of left retina | Cancer | C6922 |
| C69.3 | DX10 | Malignant neoplasm of choroid | Cancer | C693 |
| C69.30 | DX10 | Malignant neoplasm of unspecified choroid | Cancer | C6930 |
| C69.31 | DX10 | Malignant neoplasm of right choroid | Cancer | C6931 |
| C69.32 | DX10 | Malignant neoplasm of left choroid | Cancer | C6932 |
| C69.4 | DX10 | Malignant neoplasm of ciliary body | Cancer | C694 |
| C69.40 | DX10 | Malignant neoplasm of unspecified ciliary body | Cancer | C6940 |
| C69.41 | DX10 | Malignant neoplasm of right ciliary body | Cancer | C6941 |
| C69.42 | DX10 | Malignant neoplasm of left ciliary body | Cancer | C6942 |
| C69.5 | DX10 | Malignant neoplasm of lacrimal gland and duct | Cancer | C695 |
| C69.50 | DX10 | Malignant neoplasm of unspecified lacrimal gland and duct | Cancer | C6950 |
| C69.51 | DX10 | Malignant neoplasm of right lacrimal gland and duct | Cancer | C6951 |
| C69.52 | DX10 | Malignant neoplasm of left lacrimal gland and duct | Cancer | C6952 |
| C69.6 | DX10 | Malignant neoplasm of orbit | Cancer | C696 |
| C69.60 | DX10 | Malignant neoplasm of unspecified orbit | Cancer | C6960 |
| C69.61 | DX10 | Malignant neoplasm of right orbit | Cancer | C6961 |
| C69.62 | DX10 | Malignant neoplasm of left orbit | Cancer | C6962 |
| C69.8 | DX10 | Malignant neoplasm of overlapping sites of eye and adnexa | Cancer | C698 |
| C69.80 | DX10 | Malignant neoplasm of ovrlp sites of unsp eye and adnexa | Cancer | C6980 |
| C69.81 | DX10 | Malignant neoplasm of ovrlp sites of right eye and adnexa | Cancer | C6981 |
| C69.82 | DX10 | Malignant neoplasm of ovrlp sites of left eye and adnexa | Cancer | C6982 |
| C69.9 | DX10 | Malignant neoplasm of unspecified site of eye | Cancer | C699 |
| C69.90 | DX10 | Malignant neoplasm of unspecified site of unspecified eye | Cancer | C6990 |
| C69.91 | DX10 | Malignant neoplasm of unspecified site of right eye | Cancer | C6991 |
| C69.92 | DX10 | Malignant neoplasm of unspecified site of left eye | Cancer | C6992 |
| C70 | DX10 | Malignant neoplasm of meninges | Cancer | C70 |
| C70.0 | DX10 | Malignant neoplasm of cerebral meninges | Cancer | C700 |
| C70.1 | DX10 | Malignant neoplasm of spinal meninges | Cancer | C701 |
| C70.9 | DX10 | Malignant neoplasm of meninges, unspecified | Cancer | C709 |
| C71 | DX10 | Malignant neoplasm of brain | Cancer | C71 |

| | | | | |
|---|---|---|---|---|
| C71.0 | DX10 | Malignant neoplasm of cerebrum, except lobes and ventricles | Cancer | C710 |
| C71.1 | DX10 | Malignant neoplasm of frontal lobe | Cancer | C711 |
| C71.2 | DX10 | Malignant neoplasm of temporal lobe | Cancer | C712 |
| C71.3 | DX10 | Malignant neoplasm of parietal lobe | Cancer | C713 |
| C71.4 | DX10 | Malignant neoplasm of occipital lobe | Cancer | C714 |
| C71.5 | DX10 | Malignant neoplasm of cerebral ventricle | Cancer | C715 |
| C71.6 | DX10 | Malignant neoplasm of cerebellum | Cancer | C716 |
| C71.7 | DX10 | Malignant neoplasm of brain stem | Cancer | C717 |
| C71.8 | DX10 | Malignant neoplasm of overlapping sites of brain | Cancer | C718 |
| C71.9 | DX10 | Malignant neoplasm of brain, unspecified | Cancer | C719 |
| C72 | DX10 | Malig neoplm of spinal cord, cranial nerves and oth prt cnsl | Cancer | C72 |
| C72.0 | DX10 | Malignant neoplasm of spinal cord | Cancer | C720 |
| C72.1 | DX10 | Malignant neoplasm of cauda equina | Cancer | C721 |
| C72.2 | DX10 | Malignant neoplasm of olfactory nerve | Cancer | C722 |
| C72.20 | DX10 | Malignant neoplasm of unspecified olfactory nerve | Cancer | C7220 |
| C72.21 | DX10 | Malignant neoplasm of right olfactory nerve | Cancer | C7221 |
| C72.22 | DX10 | Malignant neoplasm of left olfactory nerve | Cancer | C7222 |
| C72.3 | DX10 | Malignant neoplasm of optic nerve | Cancer | C723 |
| C72.30 | DX10 | Malignant neoplasm of unspecified optic nerve | Cancer | C7230 |
| C72.31 | DX10 | Malignant neoplasm of right optic nerve | Cancer | C7231 |
| C72.32 | DX10 | Malignant neoplasm of left optic nerve | Cancer | C7232 |
| C72.4 | DX10 | Malignant neoplasm of acoustic nerve | Cancer | C724 |
| C72.40 | DX10 | Malignant neoplasm of unspecified acoustic nerve | Cancer | C7240 |
| C72.41 | DX10 | Malignant neoplasm of right acoustic nerve | Cancer | C7241 |
| C72.42 | DX10 | Malignant neoplasm of left acoustic nerve | Cancer | C7242 |
| C72.5 | DX10 | Malignant neoplasm of other and unspecified cranial nerves | Cancer | C725 |
| C72.50 | DX10 | Malignant neoplasm of unspecified cranial nerve | Cancer | C7250 |
| C72.59 | DX10 | Malignant neoplasm of other cranial nerves | Cancer | C7259 |
| C72.9 | DX10 | Malignant neoplasm of central nervous system, unspecified | Cancer | C729 |
| C73 | DX10 | Malignant neoplasm of thyroid gland | Cancer | C73 |
| C74 | DX10 | Malignant neoplasm of adrenal gland | Cancer | C74 |
| C74.0 | DX10 | Malignant neoplasm of cortex of adrenal gland | Cancer | C740 |
| C74.00 | DX10 | Malignant neoplasm of cortex of unspecified adrenal gland | Cancer | C7400 |
| C74.01 | DX10 | Malignant neoplasm of cortex of right adrenal gland | Cancer | C7401 |
| C74.02 | DX10 | Malignant neoplasm of cortex of left adrenal gland | Cancer | C7402 |
| C74.1 | DX10 | Malignant neoplasm of medulla of adrenal gland | Cancer | C741 |
| C74.10 | DX10 | Malignant neoplasm of medulla of unspecified adrenal gland | Cancer | C7410 |
| C74.11 | DX10 | Malignant neoplasm of medulla of right adrenal gland | Cancer | C7411 |
| C74.12 | DX10 | Malignant neoplasm of medulla of left adrenal gland | Cancer | C7412 |
| C74.9 | DX10 | Malignant neoplasm of unspecified part of adrenal gland | Cancer | C749 |
| C74.90 | DX10 | Malignant neoplasm of unsp part of unspecified adrenal gland | Cancer | C7490 |
| C74.91 | DX10 | Malignant neoplasm of unsp part of right adrenal gland | Cancer | C7491 |
| C74.92 | DX10 | Malignant neoplasm of unspecified part of left adrenal gland | Cancer | C7492 |
| C75 | DX10 | Malignant neoplasm of endo glands and related structures | Cancer | C75 |
| C75.0 | DX10 | Malignant neoplasm of parathyroid gland | Cancer | C750 |
| C75.1 | DX10 | Malignant neoplasm of pituitary gland | Cancer | C751 |
| C75.2 | DX10 | Malignant neoplasm of craniopharyngeal duct | Cancer | C752 |
| C75.3 | DX10 | Malignant neoplasm of pineal gland | Cancer | C753 |
| C75.4 | DX10 | Malignant neoplasm of carotid body | Cancer | C754 |

| | | | | |
|---|---|---|---|---|
| C75.5 | DX10 | Malignant neoplasm of aortic body and other paraganglia | Cancer | C755 |
| C75.8 | DX10 | Malignant neoplasm with pluriglandular involvement, unsp | Cancer | C758 |
| C75.9 | DX10 | Malignant neoplasm of endocrine gland, unspecified | Cancer | C759 |
| C76 | DX10 | Malignant neoplasm of other and ill-defined sites | Cancer | C76 |
| C76.0 | DX10 | Malignant neoplasm of head, face and neck | Cancer | C760 |
| C76.1 | DX10 | Malignant neoplasm of thorax | Cancer | C761 |
| C76.2 | DX10 | Malignant neoplasm of abdomen | Cancer | C762 |
| C76.3 | DX10 | Malignant neoplasm of pelvis | Cancer | C763 |
| C76.4 | DX10 | Malignant neoplasm of upper limb | Cancer | C764 |
| C76.40 | DX10 | Malignant neoplasm of unspecified upper limb | Cancer | C7640 |
| C76.41 | DX10 | Malignant neoplasm of right upper limb | Cancer | C7641 |
| C76.42 | DX10 | Malignant neoplasm of left upper limb | Cancer | C7642 |
| C76.5 | DX10 | Malignant neoplasm of lower limb | Cancer | C765 |
| C76.50 | DX10 | Malignant neoplasm of unspecified lower limb | Cancer | C7650 |
| C76.51 | DX10 | Malignant neoplasm of right lower limb | Cancer | C7651 |
| C76.52 | DX10 | Malignant neoplasm of left lower limb | Cancer | C7652 |
| C76.8 | DX10 | Malignant neoplasm of other specified ill-defined sites | Cancer | C768 |
| C80 | DX10 | Malignant neoplasm without specification of site | Cancer | C80 |
| C80.0 | DX10 | Disseminated malignant neoplasm, unspecified | Cancer | C800 |
| C80.1 | DX10 | Malignant (primary) neoplasm, unspecified | Cancer | C801 |
| C80.2 | DX10 | Malignant neoplasm associated with transplanted organ | Cancer | C802 |
| C7A | DX10 | Malignant neuroendocrine tumors | Cancer | C7A |
| C7A.0 | DX10 | Malignant carcinoid tumors | Cancer | C7A0 |
| C7A.00 | DX10 | Malignant carcinoid tumor of unspecified site | Cancer | C7A00 |
| C7A.01 | DX10 | Malignant carcinoid tumors of the small intestine | Cancer | C7A01 |
| C7A.010 | DX10 | Malignant carcinoid tumor of the duodenum | Cancer | C7A010 |
| C7A.011 | DX10 | Malignant carcinoid tumor of the jejunum | Cancer | C7A011 |
| C7A.012 | DX10 | Malignant carcinoid tumor of the ileum | Cancer | C7A012 |
| C7A.019 | DX10 | Malignant carcinoid tumor of the sm int, unsp portion | Cancer | C7A019 |
| C7A.02 | DX10 | Malig carcinoid tumors of the appendix, lg int, and rectum | Cancer | C7A02 |
| C7A.020 | DX10 | Malignant carcinoid tumor of the appendix | Cancer | C7A020 |
| C7A.021 | DX10 | Malignant carcinoid tumor of the cecum | Cancer | C7A021 |
| C7A.022 | DX10 | Malignant carcinoid tumor of the ascending colon | Cancer | C7A022 |
| C7A.023 | DX10 | Malignant carcinoid tumor of the transverse colon | Cancer | C7A023 |
| C7A.024 | DX10 | Malignant carcinoid tumor of the descending colon | Cancer | C7A024 |
| C7A.025 | DX10 | Malignant carcinoid tumor of the sigmoid colon | Cancer | C7A025 |
| C7A.026 | DX10 | Malignant carcinoid tumor of the rectum | Cancer | C7A026 |
| C7A.029 | DX10 | Malignant carcinoid tumor of the lg int, unsp portion | Cancer | C7A029 |
| C7A.09 | DX10 | Malignant carcinoid tumors of other sites | Cancer | C7A09 |
| C7A.090 | DX10 | Malignant carcinoid tumor of the bronchus and lung | Cancer | C7A090 |
| C7A.091 | DX10 | Malignant carcinoid tumor of the thymus | Cancer | C7A091 |
| C7A.092 | DX10 | Malignant carcinoid tumor of the stomach | Cancer | C7A092 |
| C7A.093 | DX10 | Malignant carcinoid tumor of the kidney | Cancer | C7A093 |
| C7A.094 | DX10 | Malignant carcinoid tumor of the foregut, unspecified | Cancer | C7A094 |
| C7A.095 | DX10 | Malignant carcinoid tumor of the midgut, unspecified | Cancer | C7A095 |
| C7A.096 | DX10 | Malignant carcinoid tumor of the hindgut, unspecified | Cancer | C7A096 |
| C7A.098 | DX10 | Malignant carcinoid tumors of other sites | Cancer | C7A098 |
| C7A.1 | DX10 | Malignant poorly differentiated neuroendocrine tumors | Cancer | C7A1 |
| C7A.8 | DX10 | Other malignant neuroendocrine tumors | Cancer | C7A8 |
| C7B.8 | DX10 | Other secondary neuroendocrine tumors | Cancer | C7B8 |
| C81 | DX10 | Hodgkin lymphoma | Cancer | C81 |
| C81.0 | DX10 | Nodular lymphocyte predominant Hodgkin lymphoma | Cancer | C810 |
| C81.00 | DX10 | Nodular lymphocyte predominant Hodgkin lymphoma, unsp site | Cancer | C8100 |
| C81.01 | DX10 | Nodlr lymphocy predom Hdgkn lymph, nodes of head, face, & nk | Cancer | C8101 |

| | | | | |
|---|---|---|---|---|
| C81.02 | DX10 | Nodular lymphocy predom Hodgkin lymphoma, intrathorac nodes | Cancer | C8102 |
| C81.03 | DX10 | Nodular lymphocyte predom Hodgkin lymphoma, intra-abd nodes | Cancer | C8103 |
| C81.04 | DX10 | Nodlr lymphocy predom Hdgkn lymph, nodes of axla and upr lmb | Cancer | C8104 |
| C81.05 | DX10 | Nodlr lymphocy predom Hdgkn lymph,nodes of ing rgn & low lmb | Cancer | C8105 |
| C81.06 | DX10 | Nodular lymphocyte predom Hodgkin lymphoma, intrapelv nodes | Cancer | C8106 |
| C81.07 | DX10 | Nodular lymphocyte predominant Hodgkin lymphoma, spleen | Cancer | C8107 |
| C81.08 | DX10 | Nodular lymphocyte predom Hodgkin lymphoma, nodes mult site | Cancer | C8108 |
| C81.09 | DX10 | Nodlr lymphocy predom Hdgkn lymph, extrnod & solid org site | Cancer | C8109 |
| C81.1 | DX10 | Nodular sclerosis Hodgkin lymphoma | Cancer | C811 |
| C81.10 | DX10 | Nodular sclerosis Hodgkin lymphoma, unspecified site | Cancer | C8110 |
| C81.11 | DX10 | Nodular scler Hodgkin lymph, nodes of head, face, and neck | Cancer | C8111 |
| C81.12 | DX10 | Nodular sclerosis Hodgkin lymphoma, intrathorac lymph nodes | Cancer | C8112 |
| C81.13 | DX10 | Nodular sclerosis Hodgkin lymphoma, intra-abd lymph nodes | Cancer | C8113 |
| C81.14 | DX10 | Nodular scler Hodgkin lymph, nodes of axilla and upper limb | Cancer | C8114 |
| C81.15 | DX10 | Nodlr scler Hdgkn lymph, nodes of ing region and lower limb | Cancer | C8115 |
| C81.16 | DX10 | Nodular sclerosis Hodgkin lymphoma, intrapelvic lymph nodes | Cancer | C8116 |
| C81.17 | DX10 | Nodular sclerosis Hodgkin lymphoma, spleen | Cancer | C8117 |
| C81.18 | DX10 | Nodular sclerosis Hodgkin lymphoma, lymph nodes mult site | Cancer | C8118 |
| C81.19 | DX10 | Nodular scler Hodgkin lymph, extrnod and solid organ sites | Cancer | C8119 |
| C81.2 | DX10 | Mixed cellularity Hodgkin lymphoma | Cancer | C812 |
| C81.20 | DX10 | Mixed cellularity Hodgkin lymphoma, unspecified site | Cancer | C8120 |
| C81.21 | DX10 | Mixed cellular Hodgkin lymph, nodes of head, face, and neck | Cancer | C8121 |
| C81.22 | DX10 | Mixed cellularity Hodgkin lymphoma, intrathorac lymph nodes | Cancer | C8122 |
| C81.23 | DX10 | Mixed cellularity Hodgkin lymphoma, intra-abd lymph nodes | Cancer | C8123 |
| C81.24 | DX10 | Mixed cellular Hodgkin lymph, nodes of axilla and upper limb | Cancer | C8124 |
| C81.25 | DX10 | Mixed cellular Hdgkn lymph, nodes of ing rgn and lower limb | Cancer | C8125 |
| C81.26 | DX10 | Mixed cellularity Hodgkin lymphoma, intrapelvic lymph nodes | Cancer | C8126 |
| C81.27 | DX10 | Mixed cellularity Hodgkin lymphoma, spleen | Cancer | C8127 |
| C81.28 | DX10 | Mixed cellularity Hodgkin lymphoma, lymph nodes mult site | Cancer | C8128 |
| C81.29 | DX10 | Mixed cellular Hodgkin lymph, extrnod and solid organ sites | Cancer | C8129 |
| C81.3 | DX10 | Lymphocyte depleted Hodgkin lymphoma | Cancer | C813 |
| C81.30 | DX10 | Lymphocyte depleted Hodgkin lymphoma, unspecified site | Cancer | C8130 |
| C81.31 | DX10 | Lymphocy deplet Hodgkin lymph, nodes of head, face, and neck | Cancer | C8131 |

| | | | | |
|---|---|---|---|---|
| C81.32 | DX10 | Lymphocyte depleted Hodgkin lymphoma, intrathorac nodes | Cancer | C8132 |
| C81.33 | DX10 | Lymphocyte depleted Hodgkin lymphoma, intra-abd lymph nodes | Cancer | C8133 |
| C81.34 | DX10 | Lymphocy deplet Hdgkn lymph, nodes of axilla and upper limb | Cancer | C8134 |
| C81.35 | DX10 | Lymphocy deplet Hdgkn lymph, nodes of ing rgn and lower limb | Cancer | C8135 |
| C81.36 | DX10 | Lymphocyte depleted Hodgkin lymphoma, intrapelv lymph nodes | Cancer | C8136 |
| C81.37 | DX10 | Lymphocyte depleted Hodgkin lymphoma, spleen | Cancer | C8137 |
| C81.38 | DX10 | Lymphocyte depleted Hodgkin lymphoma, lymph nodes mult site | Cancer | C8138 |
| C81.39 | DX10 | Lymphocy deplet Hodgkin lymph, extrnod and solid organ sites | Cancer | C8139 |
| C81.40 | DX10 | Lymphocyte-rich Hodgkin lymphoma, unspecified site | Cancer | C8140 |
| C81.41 | DX10 | Lymp-rich Hodgkin lymphoma, nodes of head, face, and neck | Cancer | C8141 |
| C81.42 | DX10 | Lymphocyte-rich Hodgkin lymphoma, intrathoracic lymph nodes | Cancer | C8142 |
| C81.43 | DX10 | Lymphocyte-rich Hodgkin lymphoma, intra-abd lymph nodes | Cancer | C8143 |
| C81.44 | DX10 | Lymp-rich Hodgkin lymphoma, nodes of axilla and upper limb | Cancer | C8144 |
| C81.45 | DX10 | Lymp-rich Hodgkin lymph, nodes of ing region and lower limb | Cancer | C8145 |
| C81.46 | DX10 | Lymphocyte-rich Hodgkin lymphoma, intrapelvic lymph nodes | Cancer | C8146 |
| C81.47 | DX10 | Lymphocyte-rich Hodgkin lymphoma, spleen | Cancer | C8147 |
| C81.48 | DX10 | Lymphocyte-rich Hodgkin lymphoma, lymph nodes mult site | Cancer | C8148 |
| C81.49 | DX10 | Lymp-rich Hodgkin lymphoma, extranodal and solid organ sites | Cancer | C8149 |
| C81.7 | DX10 | Other Hodgkin lymphoma | Cancer | C817 |
| C81.70 | DX10 | Other Hodgkin lymphoma, unspecified site | Cancer | C8170 |
| C81.71 | DX10 | Other Hodgkin lymphoma, lymph nodes of head, face, and neck | Cancer | C8171 |
| C81.72 | DX10 | Other Hodgkin lymphoma, intrathoracic lymph nodes | Cancer | C8172 |
| C81.73 | DX10 | Other Hodgkin lymphoma, intra-abdominal lymph nodes | Cancer | C8173 |
| C81.74 | DX10 | Other Hodgkin lymphoma, lymph nodes of axilla and upper limb | Cancer | C8174 |
| C8.75 | DX10 | | Cancer | C875 |
| C81.76 | DX10 | Other Hodgkin lymphoma, intrapelvic lymph nodes | Cancer | C8176 |
| C81.77 | DX10 | Other Hodgkin lymphoma, spleen | Cancer | C8177 |
| C81.78 | DX10 | Other Hodgkin lymphoma, lymph nodes of multiple sites | Cancer | C8178 |
| C81.79 | DX10 | Other Hodgkin lymphoma, extranodal and solid organ sites | Cancer | C8179 |
| C81.9 | DX10 | Hodgkin lymphoma, unspecified | Cancer | C819 |
| C81.90 | DX10 | Hodgkin lymphoma, unspecified, unspecified site | Cancer | C8190 |
| C81.91 | DX10 | Hodgkin lymphoma, unsp, lymph nodes of head, face, and neck | Cancer | C8191 |
| C81.92 | DX10 | Hodgkin lymphoma, unspecified, intrathoracic lymph nodes | Cancer | C8192 |
| C81.93 | DX10 | Hodgkin lymphoma, unspecified, intra-abdominal lymph nodes | Cancer | C8193 |
| C81.94 | DX10 | Hodgkin lymphoma, unsp, lymph nodes of axilla and upper limb | Cancer | C8194 |

| | | | | |
|---|---|---|---|---|
| C81.95 | DX10 | Hodgkin lymphoma, unsp, nodes of ing region and lower limb | Cancer | C8195 |
| C81.96 | DX10 | Hodgkin lymphoma, unspecified, intrapelvic lymph nodes | Cancer | C8196 |
| C81.97 | DX10 | Hodgkin lymphoma, unspecified, spleen | Cancer | C8197 |
| C81.98 | DX10 | Hodgkin lymphoma, unspecified, lymph nodes of multiple sites | Cancer | C8198 |
| C81.99 | DX10 | Hodgkin lymphoma, unsp, extranodal and solid organ sites | Cancer | C8199 |
| C82 | DX10 | Follicular lymphoma | Cancer | C82 |
| C82.0 | DX10 | Follicular lymphoma grade I | Cancer | C820 |
| C82.00 | DX10 | Follicular lymphoma grade I, unspecified site | Cancer | C8200 |
| C82.01 | DX10 | Follicular lymphoma grade I, nodes of head, face, and neck | Cancer | C8201 |
| C82.02 | DX10 | Follicular lymphoma grade I, intrathoracic lymph nodes | Cancer | C8202 |
| C82.03 | DX10 | Follicular lymphoma grade I, intra-abdominal lymph nodes | Cancer | C8203 |
| C82.04 | DX10 | Follicular lymphoma grade I, nodes of axilla and upper limb | Cancer | C8204 |
| C82.05 | DX10 | Foliclar lymph grade I, nodes of ing region and lower limb | Cancer | C8205 |
| C82.06 | DX10 | Follicular lymphoma grade I, intrapelvic lymph nodes | Cancer | C8206 |
| C82.07 | DX10 | Follicular lymphoma grade I, spleen | Cancer | C8207 |
| C82.08 | DX10 | Follicular lymphoma grade I, lymph nodes of multiple sites | Cancer | C8208 |
| C82.09 | DX10 | Follicular lymphoma grade I, extrnod and solid organ sites | Cancer | C8209 |
| C82.1 | DX10 | Follicular lymphoma grade II | Cancer | C821 |
| C82.10 | DX10 | Follicular lymphoma grade II, unspecified site | Cancer | C8210 |
| C82.11 | DX10 | Follicular lymphoma grade II, nodes of head, face, and neck | Cancer | C8211 |
| C82.12 | DX10 | Follicular lymphoma grade II, intrathoracic lymph nodes | Cancer | C8212 |
| C82.13 | DX10 | Follicular lymphoma grade II, intra-abdominal lymph nodes | Cancer | C8213 |
| C82.14 | DX10 | Follicular lymphoma grade II, nodes of axilla and upper limb | Cancer | C8214 |
| C82.15 | DX10 | Foliclar lymph grade II, nodes of ing region and lower limb | Cancer | C8215 |
| C82.16 | DX10 | Follicular lymphoma grade II, intrapelvic lymph nodes | Cancer | C8216 |
| C82.17 | DX10 | Follicular lymphoma grade II, spleen | Cancer | C8217 |
| C82.18 | DX10 | Follicular lymphoma grade II, lymph nodes of multiple sites | Cancer | C8218 |
| C82.19 | DX10 | Follicular lymphoma grade II, extrnod and solid organ sites | Cancer | C8219 |
| C82.2 | DX10 | Follicular lymphoma grade III, unspecified | Cancer | C822 |
| C82.20 | DX10 | Follicular lymphoma grade III, unspecified, unspecified site | Cancer | C8220 |
| C82.21 | DX10 | Foliclar lymph grade III, unsp, nodes of head, face, and nk | Cancer | C8221 |
| C82.22 | DX10 | Follicular lymphoma grade III, unsp, intrathorac lymph nodes | Cancer | C8222 |
| C82.23 | DX10 | Follicular lymphoma grade III, unsp, intra-abd lymph nodes | Cancer | C8223 |
| C82.24 | DX10 | Foliclar lymph grade III, unsp, nodes of axla and upper limb | Cancer | C8224 |
| C82.25 | DX10 | Foliclar lymph grade III, unsp, nodes of ing rgn and low lmb | Cancer | C8225 |

| | | | | |
|---|---|---|---|---|
| C82.26 | DX10 | Follicular lymphoma grade III, unsp, intrapelvic lymph nodes | Cancer | C8226 |
| C82.27 | DX10 | Follicular lymphoma grade III, unspecified, spleen | Cancer | C8227 |
| C82.28 | DX10 | Follicular lymphoma grade III, unsp, lymph nodes mult site | Cancer | C8228 |
| C82.29 | DX10 | Foliclar lymph grade III, unsp, extrnod and solid org sites | Cancer | C8229 |
| C82.3 | DX10 | Follicular lymphoma grade IIIa | Cancer | C823 |
| C82.30 | DX10 | Follicular lymphoma grade IIIa, unspecified site | Cancer | C8230 |
| C82.31 | DX10 | Foliclar lymphoma grade IIIa, nodes of head, face, and neck | Cancer | C8231 |
| C82.32 | DX10 | Follicular lymphoma grade IIIa, intrathoracic lymph nodes | Cancer | C8232 |
| C82.33 | DX10 | Follicular lymphoma grade IIIa, intra-abdominal lymph nodes | Cancer | C8233 |
| C82.34 | DX10 | Foliclar lymphoma grade IIIa, nodes of axilla and upper limb | Cancer | C8234 |
| C82.35 | DX10 | Foliclar lymph grade IIIa, nodes of ing rgn and lower limb | Cancer | C8235 |
| C82.36 | DX10 | Follicular lymphoma grade IIIa, intrapelvic lymph nodes | Cancer | C8236 |
| C82.37 | DX10 | Follicular lymphoma grade IIIa, spleen | Cancer | C8237 |
| C82.38 | DX10 | Follicular lymphoma grade IIIa, lymph nodes mult site | Cancer | C8238 |
| C82.39 | DX10 | Foliclar lymphoma grade IIIa, extrnod and solid organ sites | Cancer | C8239 |
| C82.4 | DX10 | Follicular lymphoma grade IIIb | Cancer | C824 |
| C82.40 | DX10 | Follicular lymphoma grade IIIb, unspecified site | Cancer | C8240 |
| C82.41 | DX10 | Foliclar lymphoma grade IIIb, nodes of head, face, and neck | Cancer | C8241 |
| C82.42 | DX10 | Follicular lymphoma grade IIIb, intrathoracic lymph nodes | Cancer | C8242 |
| C82.43 | DX10 | Follicular lymphoma grade IIIb, intra-abdominal lymph nodes | Cancer | C8243 |
| C82.44 | DX10 | Foliclar lymphoma grade IIIb, nodes of axilla and upper limb | Cancer | C8244 |
| C82.45 | DX10 | Foliclar lymph grade IIIb, nodes of ing rgn and lower limb | Cancer | C8245 |
| C82.46 | DX10 | Follicular lymphoma grade IIIb, intrapelvic lymph nodes | Cancer | C8246 |
| C82.47 | DX10 | Follicular lymphoma grade IIIb, spleen | Cancer | C8247 |
| C82.48 | DX10 | Follicular lymphoma grade IIIb, lymph nodes mult site | Cancer | C8248 |
| C82.49 | DX10 | Foliclar lymphoma grade IIIb, extrnod and solid organ sites | Cancer | C8249 |
| C82.5 | DX10 | Diffuse follicle center lymphoma | Cancer | C825 |
| C82.50 | DX10 | Diffuse follicle center lymphoma, unspecified site | Cancer | C8250 |
| C82.51 | DX10 | Diffuse folicl center lymph, nodes of head, face, and neck | Cancer | C8251 |
| C82.52 | DX10 | Diffuse follicle center lymphoma, intrathoracic lymph nodes | Cancer | C8252 |
| C82.53 | DX10 | Diffuse follicle center lymphoma, intra-abd lymph nodes | Cancer | C8253 |
| C82.54 | DX10 | Diffuse follicle center lymphoma, lymph nodes axilla upper limb | Cancer | C8254 |
| C82.55 | DX10 | Diffus folicl cntr lymph, nodes of ing region and lower limb | Cancer | C8255 |
| C82.56 | DX10 | Diffuse follicle center lymphoma, intrapelvic lymph nodes | Cancer | C8256 |
| C82.57 | DX10 | Diffuse follicle center lymphoma, spleen | Cancer | C8257 |

| | | | | |
|---|---|---|---|---|
| C82.58 | DX10 | Diffuse follicle center lymphoma, lymph nodes mult site | Cancer | C8258 |
| C82.59 | DX10 | Diffuse folicl center lymph, extrnod and solid organ sites | Cancer | C8259 |
| C82.6 | DX10 | Cutaneous follicle center lymphoma | Cancer | C826 |
| C82.60 | DX10 | Cutaneous follicle center lymphoma, unspecified site | Cancer | C8260 |
| C82.61 | DX10 | Cutan folicl center lymphoma, nodes of head, face, and neck | Cancer | C8261 |
| C82.62 | DX10 | Cutaneous follicle center lymphoma, intrathorac lymph nodes | Cancer | C8262 |
| C82.63 | DX10 | Cutaneous follicle center lymphoma, intra-abd lymph nodes | Cancer | C8263 |
| C82.64 | DX10 | Cutan folicl center lymphoma, nodes of axilla and upper limb | Cancer | C8264 |
| C82.65 | DX10 | Cutan folicl cntr lymph, nodes of ing region and lower limb | Cancer | C8265 |
| C82.66 | DX10 | Cutaneous follicle center lymphoma, intrapelvic lymph nodes | Cancer | C8266 |
| C82.67 | DX10 | Cutaneous follicle center lymphoma, spleen | Cancer | C8267 |
| C82.68 | DX10 | Cutaneous follicle center lymphoma, lymph nodes mult site | Cancer | C8268 |
| C82.69 | DX10 | Cutan folicl center lymphoma, extrnod and solid organ sites | Cancer | C8269 |
| C82.8 | DX10 | Other types of follicular lymphoma | Cancer | C828 |
| C82.80 | DX10 | Other types of follicular lymphoma, unspecified site | Cancer | C8280 |
| C82.81 | DX10 | Oth types of foliclar lymph, nodes of head, face, and neck | Cancer | C8281 |
| C82.82 | DX10 | Oth types of follicular lymphoma, intrathoracic lymph nodes | Cancer | C8282 |
| C82.83 | DX10 | Oth types of follicular lymphoma, intra-abd lymph nodes | Cancer | C8283 |
| C82.84 | DX10 | Oth types of foliclar lymph, nodes of axilla and upper limb | Cancer | C8284 |
| C82.85 | DX10 | Oth types of foliclar lymph, nodes of ing rgn and lower limb | Cancer | C8285 |
| C82.86 | DX10 | Other types of follicular lymphoma, intrapelvic lymph nodes | Cancer | C8286 |
| C82.87 | DX10 | Other types of follicular lymphoma, spleen | Cancer | C8287 |
| C82.88 | DX10 | Oth types of follicular lymphoma, lymph nodes mult site | Cancer | C8288 |
| C82.89 | DX10 | Oth types of foliclar lymph, extrnod and solid organ sites | Cancer | C8289 |
| C82.9 | DX10 | Follicular lymphoma, unspecified | Cancer | C829 |
| C82.90 | DX10 | Follicular lymphoma, unspecified, unspecified site | Cancer | C8290 |
| C82.91 | DX10 | Follicular lymphoma, unsp, nodes of head, face, and neck | Cancer | C8291 |
| C82.92 | DX10 | Follicular lymphoma, unspecified, intrathoracic lymph nodes | Cancer | C8292 |
| C82.93 | DX10 | Follicular lymphoma, unsp, intra-abdominal lymph nodes | Cancer | C8293 |
| C82.94 | DX10 | Follicular lymphoma, unsp, nodes of axilla and upper limb | Cancer | C8294 |
| C82.95 | DX10 | Foliclar lymphoma, unsp, nodes of ing region and lower limb | Cancer | C8295 |
| C82.96 | DX10 | Follicular lymphoma, unspecified, intrapelvic lymph nodes | Cancer | C8296 |
| C82.97 | DX10 | Follicular lymphoma, unspecified, spleen | Cancer | C8297 |
| C82.98 | DX10 | Follicular lymphoma, unsp, lymph nodes of multiple sites | Cancer | C8298 |

| | | | | |
|---|---|---|---|---|
| C82.99 | DX10 | Follicular lymphoma, unsp, extranodal and solid organ sites | Cancer | C8299 |
| C83 | DX10 | Non-follicular lymphoma | Cancer | C83 |
| C83.0 | DX10 | Small cell B-cell lymphoma | Cancer | C830 |
| C83.00 | DX10 | Small cell B-cell lymphoma, unspecified site | Cancer | C8300 |
| C83.01 | DX10 | Small cell B-cell lymphoma, nodes of head, face, and neck | Cancer | C8301 |
| C83.02 | DX10 | Small cell B-cell lymphoma, intrathoracic lymph nodes | Cancer | C8302 |
| C83.03 | DX10 | Small cell B-cell lymphoma, intra-abdominal lymph nodes | Cancer | C8303 |
| C83.04 | DX10 | Small cell B-cell lymphoma, nodes of axilla and upper limb | Cancer | C8304 |
| C83.05 | DX10 | Small cell B-cell lymph, nodes of ing region and lower limb | Cancer | C8305 |
| C83.06 | DX10 | Small cell B-cell lymphoma, intrapelvic lymph nodes | Cancer | C8306 |
| C83.07 | DX10 | Small cell B-cell lymphoma, spleen | Cancer | C8307 |
| C83.08 | DX10 | Small cell B-cell lymphoma, lymph nodes of multiple sites | Cancer | C8308 |
| C83.09 | DX10 | Small cell B-cell lymphoma, extranodal and solid organ sites | Cancer | C8309 |
| C83.1 | DX10 | Mantle cell lymphoma | Cancer | C831 |
| C83.10 | DX10 | Mantle cell lymphoma, unspecified site | Cancer | C8310 |
| C83.11 | DX10 | Mantle cell lymphoma, lymph nodes of head, face, and neck | Cancer | C8311 |
| C83.12 | DX10 | Mantle cell lymphoma, intrathoracic lymph nodes | Cancer | C8312 |
| C83.13 | DX10 | Mantle cell lymphoma, intra-abdominal lymph nodes | Cancer | C8313 |
| C83.14 | DX10 | Mantle cell lymphoma, lymph nodes of axilla and upper limb | Cancer | C8314 |
| C83.15 | DX10 | Mantle cell lymphoma, nodes of ing region and lower limb | Cancer | C8315 |
| C83.16 | DX10 | Mantle cell lymphoma, intrapelvic lymph nodes | Cancer | C8316 |
| C83.17 | DX10 | Mantle cell lymphoma, spleen | Cancer | C8317 |
| C83.18 | DX10 | Mantle cell lymphoma, lymph nodes of multiple sites | Cancer | C8318 |
| C83.19 | DX10 | Mantle cell lymphoma, extranodal and solid organ sites | Cancer | C8319 |
| C83.3 | DX10 | Diffuse large B-cell lymphoma | Cancer | C833 |
| C83.30 | DX10 | Diffuse large B-cell lymphoma, unspecified site | Cancer | C8330 |
| C83.31 | DX10 | Diffuse large B-cell lymphoma, nodes of head, face, and neck | Cancer | C8331 |
| C83.32 | DX10 | Diffuse large B-cell lymphoma, intrathoracic lymph nodes | Cancer | C8332 |
| C83.33 | DX10 | Diffuse large B-cell lymphoma, intra-abdominal lymph nodes | Cancer | C8333 |
| C83.34 | DX10 | Diffuse large B-cell lymph, nodes of axilla and upper limb | Cancer | C8334 |
| C83.35 | DX10 | Diffus large B-cell lymph, nodes of ing rgn and lower limb | Cancer | C8335 |
| C83.36 | DX10 | Diffuse large B-cell lymphoma, intrapelvic lymph nodes | Cancer | C8336 |
| C83.37 | DX10 | Diffuse large B-cell lymphoma, spleen | Cancer | C8337 |
| C83.38 | DX10 | Diffuse large B-cell lymphoma, lymph nodes of multiple sites | Cancer | C8338 |
| C83.39 | DX10 | Diffuse large B-cell lymphoma, extrnod and solid organ sites | Cancer | C8339 |
| C83.5 | DX10 | Lymphoblastic (diffuse) lymphoma | Cancer | C835 |
| C83.50 | DX10 | Lymphoblastic (diffuse) lymphoma, unspecified site | Cancer | C8350 |
| C83.51 | DX10 | Lymphoblastic lymphoma, nodes of head, face, and neck | Cancer | C8351 |
| C83.52 | DX10 | Lymphoblastic (diffuse) lymphoma, intrathoracic lymph nodes | Cancer | C8352 |

| | | | | |
|---|---|---|---|---|
| C83.53 | DX10 | Lymphoblastic (diffuse) lymphoma, intra-abd lymph nodes | Cancer | C8353 |
| C83.54 | DX10 | Lymphoblastic lymphoma, nodes of axilla and upper limb | Cancer | C8354 |
| C83.55 | DX10 | Lymphoblastic lymphoma, nodes of ing region and lower limb | Cancer | C8355 |
| C83.56 | DX10 | Lymphoblastic (diffuse) lymphoma, intrapelvic lymph nodes | Cancer | C8356 |
| C83.57 | DX10 | Lymphoblastic (diffuse) lymphoma, spleen | Cancer | C8357 |
| C83.58 | DX10 | Lymphoblastic (diffuse) lymphoma, lymph nodes mult site | Cancer | C8358 |
| C83.59 | DX10 | Lymphoblastic lymphoma, extrnod and solid organ sites | Cancer | C8359 |
| C83.7 | DX10 | Burkitt lymphoma | Cancer | C837 |
| C83.70 | DX10 | Burkitt lymphoma, unspecified site | Cancer | C8370 |
| C83.71 | DX10 | Burkitt lymphoma, lymph nodes of head, face, and neck | Cancer | C8371 |
| C83.72 | DX10 | Burkitt lymphoma, intrathoracic lymph nodes | Cancer | C8372 |
| C83.73 | DX10 | Burkitt lymphoma, intra-abdominal lymph nodes | Cancer | C8373 |
| C83.74 | DX10 | Burkitt lymphoma, lymph nodes of axilla and upper limb | Cancer | C8374 |
| C83.75 | DX10 | Burkitt lymphoma, nodes of inguinal region and lower limb | Cancer | C8375 |
| C83.76 | DX10 | Burkitt lymphoma, intrapelvic lymph nodes | Cancer | C8376 |
| C83.77 | DX10 | Burkitt lymphoma, spleen | Cancer | C8377 |
| C83.78 | DX10 | Burkitt lymphoma, lymph nodes of multiple sites | Cancer | C8378 |
| C83.79 | DX10 | Burkitt lymphoma, extranodal and solid organ sites | Cancer | C8379 |
| C83.8 | DX10 | Other non-follicular lymphoma | Cancer | C838 |
| C83.80 | DX10 | Other non-follicular lymphoma, unspecified site | Cancer | C8380 |
| C83.81 | DX10 | Oth non-follic lymphoma, lymph nodes of head, face, and neck | Cancer | C8381 |
| C38.82 | DX10 | | Cancer | C3882 |
| C83.83 | DX10 | Other non-follicular lymphoma, intra-abdominal lymph nodes | Cancer | C8383 |
| C83.84 | DX10 | Oth non-follic lymphoma, nodes of axilla and upper limb | Cancer | C8384 |
| C83.85 | DX10 | Oth non-follic lymphoma, nodes of ing region and lower limb | Cancer | C8385 |
| C83.86 | DX10 | Other non-follicular lymphoma, intrapelvic lymph nodes | Cancer | C8386 |
| C83.87 | DX10 | Other non-follicular lymphoma, spleen | Cancer | C8387 |
| C83.88 | DX10 | Other non-follicular lymphoma, lymph nodes of multiple sites | Cancer | C8388 |
| C83.89 | DX10 | Oth non-follic lymphoma, extranodal and solid organ sites | Cancer | C8389 |
| C83.9 | DX10 | Non-follicular (diffuse) lymphoma, unspecified | Cancer | C839 |
| C83.90 | DX10 | Non-follicular (diffuse) lymphoma, unsp, unspecified site | Cancer | C8390 |
| C83.91 | DX10 | Non-follic lymphoma, unsp, nodes of head, face, and neck | Cancer | C8391 |
| C83.92 | DX10 | Non-follic (diffuse) lymphoma, unsp, intrathorac lymph nodes | Cancer | C8392 |
| C83.93 | DX10 | Non-follic (diffuse) lymphoma, unsp, intra-abd lymph nodes | Cancer | C8393 |
| C83.94 | DX10 | Non-follic lymphoma, unsp, nodes of axilla and upper limb | Cancer | C8394 |
| C83.95 | DX10 | Non-follic lymph, unsp, nodes of ing region and lower limb | Cancer | C8395 |

| | | | | |
|---|---|---|---|---|
| C83.96 | DX10 | Non-follic (diffuse) lymphoma, unsp, intrapelvic lymph nodes | Cancer | C8396 |
| C83.97 | DX10 | Non-follicular (diffuse) lymphoma, unspecified, spleen | Cancer | C8397 |
| C83.98 | DX10 | Non-follic (diffuse) lymphoma, unsp, lymph nodes mult site | Cancer | C8398 |
| C83.99 | DX10 | Non-follic lymphoma, unsp, extrnod and solid organ sites | Cancer | C8399 |
| C84 | DX10 | Mature T/NK-cell lymphomas | Cancer | C84 |
| C84.0 | DX10 | Mycosis fungoides | Cancer | C840 |
| C84.00 | DX10 | Mycosis fungoides, unspecified site | Cancer | C8400 |
| C84.01 | DX10 | Mycosis fungoides, lymph nodes of head, face, and neck | Cancer | C8401 |
| C84.02 | DX10 | Mycosis fungoides, intrathoracic lymph nodes | Cancer | C8402 |
| C84.03 | DX10 | Mycosis fungoides, intra-abdominal lymph nodes | Cancer | C8403 |
| C84.04 | DX10 | Mycosis fungoides, lymph nodes of axilla and upper limb | Cancer | C8404 |
| C84.05 | DX10 | Mycosis fungoides, nodes of inguinal region and lower limb | Cancer | C8405 |
| C84.06 | DX10 | Mycosis fungoides, intrapelvic lymph nodes | Cancer | C8406 |
| C84.07 | DX10 | Mycosis fungoides, spleen | Cancer | C8407 |
| C84.08 | DX10 | Mycosis fungoides, lymph nodes of multiple sites | Cancer | C8408 |
| C84.09 | DX10 | Mycosis fungoides, extranodal and solid organ sites | Cancer | C8409 |
| C84.1 | DX10 | Sezary disease | Cancer | C841 |
| C84.10 | DX10 | Sezary disease, unspecified site | Cancer | C8410 |
| C84.11 | DX10 | Sezary disease, lymph nodes of head, face, and neck | Cancer | C8411 |
| C84.12 | DX10 | Sezary disease, intrathoracic lymph nodes | Cancer | C8412 |
| C84.13 | DX10 | Sezary disease, intra-abdominal lymph nodes | Cancer | C8413 |
| C84.14 | DX10 | Sezary disease, lymph nodes of axilla and upper limb | Cancer | C8414 |
| C84.15 | DX10 | Sezary disease, nodes of inguinal region and lower limb | Cancer | C8415 |
| C84.16 | DX10 | Sezary disease, intrapelvic lymph nodes | Cancer | C8416 |
| C84.17 | DX10 | Sezary disease, spleen | Cancer | C8417 |
| C84.18 | DX10 | Sezary disease, lymph nodes of multiple sites | Cancer | C8418 |
| C84.19 | DX10 | Sezary disease, extranodal and solid organ sites | Cancer | C8419 |
| C84.4 | DX10 | Peripheral T-cell lymphoma, not classified | Cancer | C844 |
| C84.40 | DX10 | Peripheral T-cell lymphoma, not classified, unspecified site | Cancer | C8440 |
| C84.41 | DX10 | Prph T-cell lymph, not class, nodes of head, face, and neck | Cancer | C8441 |
| C84.42 | DX10 | Peripheral T-cell lymphoma, not class, intrathorac nodes | Cancer | C8442 |
| C84.43 | DX10 | Peripheral T-cell lymphoma, not classified, intra-abd nodes | Cancer | C8443 |
| C84.44 | DX10 | Prph T-cell lymph, not class, nodes of axilla and upper limb | Cancer | C8444 |
| C84.45 | DX10 | Prph T-cell lymph, not class, nodes of ing rgn and low limb | Cancer | C8445 |
| C84.46 | DX10 | Peripheral T-cell lymphoma, not classified, intrapelv nodes | Cancer | C8446 |
| C84.47 | DX10 | Peripheral T-cell lymphoma, not classified, spleen | Cancer | C8447 |
| C84.48 | DX10 | Peripheral T-cell lymphoma, not classified, nodes mult site | Cancer | C8448 |
| C84.49 | DX10 | Prph T-cell lymph, not class, extrnod and solid organ sites | Cancer | C8449 |
| C84.6 | DX10 | Anaplastic large cell lymphoma, ALK-positive | Cancer | C846 |
| C84.60 | DX10 | Anaplastic large cell lymphoma, ALK-positive, unsp site | Cancer | C8460 |
| C84.61 | DX10 | Anaplstc lg cell lymph, ALK-pos, nodes of head, face, and nk | Cancer | C8461 |
| C84.62 | DX10 | Anaplastic large cell lymphoma, ALK-pos, intrathorac nodes | Cancer | C8462 |

| | | | | |
|---|---|---|---|---|
| C84.63 | DX10 | Anaplastic large cell lymphoma, ALK-pos, intra-abd nodes | Cancer | C8463 |
| C84.64 | DX10 | Anaplstc lg cell lymph, ALK-pos, nodes of axla and upr limb | Cancer | C8464 |
| C84.65 | DX10 | Anaplstc lg cell lymph, ALK-pos, nodes of ing rgn & low lmb | Cancer | C8465 |
| C84.66 | DX10 | Anaplastic large cell lymphoma, ALK-pos, intrapelv nodes | Cancer | C8466 |
| C84.67 | DX10 | Anaplastic large cell lymphoma, ALK-positive, spleen | Cancer | C8467 |
| C84.68 | DX10 | Anaplastic large cell lymphoma, ALK-pos, nodes mult site | Cancer | C8468 |
| C84.69 | DX10 | Anaplstc lg cell lymph, ALK-pos, extrnod and solid org sites | Cancer | C8469 |
| C84.7 | DX10 | Anaplastic large cell lymphoma, ALK-negative | Cancer | C847 |
| C84.70 | DX10 | Anaplastic large cell lymphoma, ALK-negative, unsp site | Cancer | C8470 |
| C84.71 | DX10 | Anaplstc lg cell lymph, ALK-neg, nodes of head, face, and nk | Cancer | C8471 |
| C84.72 | DX10 | Anaplastic large cell lymphoma, ALK-neg, intrathorac nodes | Cancer | C8472 |
| C84.73 | DX10 | Anaplastic large cell lymphoma, ALK-neg, intra-abd nodes | Cancer | C8473 |
| C84.74 | DX10 | Anaplstc lg cell lymph, ALK-neg, nodes of axla and upr limb | Cancer | C8474 |
| C84.75 | DX10 | Anaplstc lg cell lymph, ALK-neg, nodes of ing rgn & low lmb | Cancer | C8475 |
| C84.76 | DX10 | Anaplastic large cell lymphoma, ALK-neg, intrapelv nodes | Cancer | C8476 |
| C84.77 | DX10 | Anaplastic large cell lymphoma, ALK-negative, spleen | Cancer | C8477 |
| C84.78 | DX10 | Anaplastic large cell lymphoma, ALK-neg, nodes mult site | Cancer | C8478 |
| C84.79 | DX10 | Anaplstc lg cell lymph, ALK-neg, extrnod and solid org sites | Cancer | C8479 |
| C84.A | DX10 | Cutaneous T-cell lymphoma, unspecified | Cancer | C84A |
| C84.A0 | DX10 | Cutaneous T-cell lymphoma, unspecified, unspecified site | Cancer | C84A0 |
| C84.A1 | DX10 | Cutan T-cell lymphoma, unsp nodes of head, face, and neck | Cancer | C84A1 |
| C84.A2 | DX10 | Cutaneous T-cell lymphoma, unsp, intrathoracic lymph nodes | Cancer | C84A2 |
| C84.A3 | DX10 | Cutaneous T-cell lymphoma, unsp, intra-abdominal lymph nodes | Cancer | C84A3 |
| C84.A4 | DX10 | Cutan T-cell lymphoma, unsp, nodes of axilla and upper limb | Cancer | C84A4 |
| C84.A5 | DX10 | Cutan T-cell lymph, unsp, nodes of ing region and lower limb | Cancer | C84A5 |
| C84.A6 | DX10 | Cutaneous T-cell lymphoma, unsp, intrapelvic lymph nodes | Cancer | C84A6 |
| C84.A7 | DX10 | Cutaneous T-cell lymphoma, unspecified, spleen | Cancer | C84A7 |
| C84.A8 | DX10 | Cutaneous T-cell lymphoma, unsp, lymph nodes mult site | Cancer | C84A8 |
| C84.A9 | DX10 | Cutan T-cell lymphoma, unsp, extrnod and solid organ sites | Cancer | C84A9 |
| C84.Z | DX10 | Other mature T/NK-cell lymphomas | Cancer | C84Z |
| C84.Z0 | DX10 | Other mature T/NK-cell lymphomas, unspecified site | Cancer | C84Z0 |
| C84.Z1 | DX10 | Oth mature T/NK-cell lymph, nodes of head, face, and neck | Cancer | C84Z1 |
| C84.Z2 | DX10 | Other mature T/NK-cell lymphomas, intrathoracic lymph nodes | Cancer | C84Z2 |

| | | | | |
|---|---|---|---|---|
| C84.Z3 | DX10 | Oth mature T/NK-cell lymphomas, intra-abdominal lymph nodes | Cancer | C84Z3 |
| C84.Z4 | DX10 | Oth mature T/NK-cell lymph, nodes of axilla and upper limb | Cancer | C84Z4 |
| C84.Z5 | DX10 | Oth mature T/NK-cell lymph, nodes of ing rgn and lower limb | Cancer | C84Z5 |
| C84.Z6 | DX10 | Other mature T/NK-cell lymphomas, intrapelvic lymph nodes | Cancer | C84Z6 |
| C84.Z7 | DX10 | Other mature T/NK-cell lymphomas, spleen | Cancer | C84Z7 |
| C84.Z8 | DX10 | Oth mature T/NK-cell lymphomas, lymph nodes mult site | Cancer | C84Z8 |
| C84.Z9 | DX10 | Oth mature T/NK-cell lymph, extrnod and solid organ sites | Cancer | C84Z9 |
| C84.9 | DX10 | Mature T/NK-cell lymphomas, unspecified | Cancer | C849 |
| C84.90 | DX10 | Mature T/NK-cell lymphomas, unspecified, unspecified site | Cancer | C8490 |
| C84.91 | DX10 | Mature T/NK-cell lymph, unsp, nodes of head, face, and neck | Cancer | C8491 |
| C84.92 | DX10 | Mature T/NK-cell lymphomas, unsp, intrathoracic lymph nodes | Cancer | C8492 |
| C84.93 | DX10 | Mature T/NK-cell lymphomas, unsp, intra-abd lymph nodes | Cancer | C8493 |
| C84.94 | DX10 | Mature T/NK-cell lymph, unsp, nodes of axilla and upper limb | Cancer | C8494 |
| C84.95 | DX10 | Mature T/NK-cell lymph, unsp, nodes of ing rgn and low limb | Cancer | C8495 |
| C84.96 | DX10 | Mature T/NK-cell lymphomas, unsp, intrapelvic lymph nodes | Cancer | C8496 |
| C84.97 | DX10 | Mature T/NK-cell lymphomas, unspecified, spleen | Cancer | C8497 |
| C84.98 | DX10 | Mature T/NK-cell lymphomas, unsp, lymph nodes mult site | Cancer | C8498 |
| C84.99 | DX10 | Mature T/NK-cell lymph, unsp, extrnod and solid organ sites | Cancer | C8499 |
| C85 | DX10 | Oth and unspecified types of non-Hodgkin lymphoma | Cancer | C85 |
| C85.1 | DX10 | Unspecified B-cell lymphoma | Cancer | C851 |
| C85.10 | DX10 | Unspecified B-cell lymphoma, unspecified site | Cancer | C8510 |
| C85.11 | DX10 | Unsp B-cell lymphoma, lymph nodes of head, face, and neck | Cancer | C8511 |
| C85.12 | DX10 | Unspecified B-cell lymphoma, intrathoracic lymph nodes | Cancer | C8512 |
| C85.13 | DX10 | Unspecified B-cell lymphoma, intra-abdominal lymph nodes | Cancer | C8513 |
| C85.14 | DX10 | Unsp B-cell lymphoma, lymph nodes of axilla and upper limb | Cancer | C8514 |
| C85.15 | DX10 | Unsp B-cell lymphoma, nodes of ing region and lower limb | Cancer | C8515 |
| C85.16 | DX10 | Unspecified B-cell lymphoma, intrapelvic lymph nodes | Cancer | C8516 |
| C85.17 | DX10 | Unspecified B-cell lymphoma, spleen | Cancer | C8517 |
| C85.18 | DX10 | Unspecified B-cell lymphoma, lymph nodes of multiple sites | Cancer | C8518 |
| C85.19 | DX10 | Unsp B-cell lymphoma, extranodal and solid organ sites | Cancer | C8519 |
| C85.2 | DX10 | Mediastinal (thymic) large B-cell lymphoma | Cancer | C852 |
| C85.20 | DX10 | Mediastinal (thymic) large B-cell lymphoma, unspecified site | Cancer | C8520 |
| C85.21 | DX10 | Mediastnl large B-cell lymph, nodes of head, face, and neck | Cancer | C8521 |
| C85.22 | DX10 | Mediastnl (thymic) large B-cell lymphoma, intrathorac nodes | Cancer | C8522 |

| | | | | |
|---|---|---|---|---|
| C85.23 | DX10 | Mediastinal (thymic) large B-cell lymphoma, intra-abd nodes | Cancer | C8523 |
| C85.24 | DX10 | Mediastnl large B-cell lymph, nodes of axilla and upper limb | Cancer | C8524 |
| C85.25 | DX10 | Mediastnl lg B-cell lymph, nodes of ing rgn and lower limb | Cancer | C8525 |
| C85.26 | DX10 | Mediastinal (thymic) large B-cell lymphoma, intrapelv nodes | Cancer | C8526 |
| C85.27 | DX10 | Mediastinal (thymic) large B-cell lymphoma, spleen | Cancer | C8527 |
| C85.28 | DX10 | Mediastinal (thymic) large B-cell lymphoma, nodes mult site | Cancer | C8528 |
| C85.29 | DX10 | Mediastnl large B-cell lymph, extrnod and solid organ sites | Cancer | C8529 |
| C85.8 | DX10 | Other specified types of non-Hodgkin lymphoma | Cancer | C858 |
| C85.80 | DX10 | Oth types of non-Hodgkin lymphoma, unspecified site | Cancer | C8580 |
| C85.81 | DX10 | Oth types of non-hodg lymph, nodes of head, face, and neck | Cancer | C8581 |
| C85.82 | DX10 | Oth types of non-Hodgkin lymphoma, intrathoracic lymph nodes | Cancer | C8582 |
| C85.83 | DX10 | Oth types of non-Hodgkin lymphoma, intra-abd lymph nodes | Cancer | C8583 |
| C85.84 | DX10 | Oth types of non-hodg lymph, nodes of axilla and upper limb | Cancer | C8584 |
| C85.85 | DX10 | Oth types of non-hodg lymph, nodes of ing rgn and lower limb | Cancer | C8585 |
| C85.86 | DX10 | Oth types of non-Hodgkin lymphoma, intrapelvic lymph nodes | Cancer | C8586 |
| C85.87 | DX10 | Other specified types of non-Hodgkin lymphoma, spleen | Cancer | C8587 |
| C85.88 | DX10 | Oth types of non-Hodgkin lymphoma, lymph nodes mult site | Cancer | C8588 |
| C85.89 | DX10 | Oth types of non-hodg lymph, extrnod and solid organ sites | Cancer | C8589 |
| C85.9 | DX10 | Non-Hodgkin lymphoma, unspecified | Cancer | C859 |
| C85.90 | DX10 | Non-Hodgkin lymphoma, unspecified, unspecified site | Cancer | C8590 |
| C85.91 | DX10 | Non-Hodgkin lymphoma, unsp, nodes of head, face, and neck | Cancer | C8591 |
| C85.92 | DX10 | Non-Hodgkin lymphoma, unspecified, intrathoracic lymph nodes | Cancer | C8592 |
| C85.93 | DX10 | Non-Hodgkin lymphoma, unsp, intra-abdominal lymph nodes | Cancer | C8593 |
| C85.94 | DX10 | Non-Hodgkin lymphoma, unsp, nodes of axilla and upper limb | Cancer | C8594 |
| C85.95 | DX10 | Non-hodg lymphoma, unsp, nodes of ing region and lower limb | Cancer | C8595 |
| C85.96 | DX10 | Non-Hodgkin lymphoma, unspecified, intrapelvic lymph nodes | Cancer | C8596 |
| C85.97 | DX10 | Non-Hodgkin lymphoma, unspecified, spleen | Cancer | C8597 |
| C85.98 | DX10 | Non-Hodgkin lymphoma, unsp, lymph nodes of multiple sites | Cancer | C8598 |
| C85.99 | DX10 | Non-Hodgkin lymphoma, unsp, extranodal and solid organ sites | Cancer | C8599 |
| C86 | DX10 | Other specified types of T/NK-cell lymphoma | Cancer | C86 |
| C86.0 | DX10 | Extranodal NK/T-cell lymphoma, nasal type | Cancer | C860 |
| C86.1 | DX10 | Hepatosplenic T-cell lymphoma | Cancer | C861 |
| C86.2 | DX10 | Enteropathy-type (intestinal) T-cell lymphoma | Cancer | C862 |
| C86.3 | DX10 | Subcutaneous panniculitis-like T-cell lymphoma | Cancer | C863 |
| C86.4 | DX10 | Blastic NK-cell lymphoma | Cancer | C864 |
| C86.5 | DX10 | Angioimmunoblastic T-cell lymphoma | Cancer | C865 |

| | | | | |
|---|---|---|---|---|
| C86.6 | DX10 | Primary cutaneous CD30-positive T-cell proliferations | Cancer | C866 |
| C88 | DX10 | Malig immunoproliferative dis and certain oth B-cell lymph | Cancer | C88 |
| C88.0 | DX10 | Waldenstrom macroglobulinemia | Cancer | C880 |
| C88.2 | DX10 | Heavy chain disease | Cancer | C882 |
| C88.3 | DX10 | Immunoproliferative small intestinal disease | Cancer | C883 |
| C88.4 | DX10 | Extrnod mrgnl zn B-cell lymph of mucosa-assoc lymphoid tiss | Cancer | C884 |
| C88.8 | DX10 | Other malignant immunoproliferative diseases | Cancer | C888 |
| C88.9 | DX10 | Malignant immunoproliferative disease, unspecified | Cancer | C889 |
| C90 | DX10 | Multiple myeloma and malignant plasma cell neoplasms | Cancer | C90 |
| C90.0 | DX10 | Multiple myeloma | Cancer | C900 |
| C90.00 | DX10 | Multiple myeloma not having achieved remission | Cancer | C9000 |
| C90.01 | DX10 | Multiple myeloma in remission | Cancer | C9001 |
| C90.02 | DX10 | Multiple myeloma in relapse | Cancer | C9002 |
| C90.1 | DX10 | Plasma cell leukemia | Cancer | C901 |
| C90.10 | DX10 | Plasma cell leukemia not having achieved remission | Cancer | C9010 |
| C90.11 | DX10 | Plasma cell leukemia in remission | Cancer | C9011 |
| C90.12 | DX10 | Plasma cell leukemia in relapse | Cancer | C9012 |
| C90.2 | DX10 | Extramedullary plasmacytoma | Cancer | C902 |
| C90.20 | DX10 | Extramedullary plasmacytoma not having achieved remission | Cancer | C9020 |
| C90.21 | DX10 | Extramedullary plasmacytoma in remission | Cancer | C9021 |
| C90.22 | DX10 | Extramedullary plasmacytoma in relapse | Cancer | C9022 |
| C90.3 | DX10 | Solitary plasmacytoma | Cancer | C903 |
| C90.30 | DX10 | Solitary plasmacytoma not having achieved remission | Cancer | C9030 |
| C90.31 | DX10 | Solitary plasmacytoma in remission | Cancer | C9031 |
| C90.32 | DX10 | Solitary plasmacytoma in relapse | Cancer | C9032 |
| C91 | DX10 | Lymphoid leukemia | Cancer | C91 |
| C91.0 | DX10 | Acute lymphoblastic leukemia [ALL] | Cancer | C910 |
| C91.00 | DX10 | Acute lymphoblastic leukemia not having achieved remission | Cancer | C9100 |
| C91.01 | DX10 | Acute lymphoblastic leukemia, in remission | Cancer | C9101 |
| C91.02 | DX10 | Acute lymphoblastic leukemia, in relapse | Cancer | C9102 |
| C91.1 | DX10 | Chronic lymphocytic leukemia of B-cell type | Cancer | C911 |
| C91.10 | DX10 | Chronic lymphocytic leuk of B-cell type not achieve remis | Cancer | C9110 |
| C91.11 | DX10 | Chronic lymphocytic leukemia of B-cell type in remission | Cancer | C9111 |
| C91.12 | DX10 | Chronic lymphocytic leukemia of B-cell type in relapse | Cancer | C9112 |
| C91.3 | DX10 | Prolymphocytic leukemia of B-cell type | Cancer | C913 |
| C91.30 | DX10 | Prolymphocytic leukemia of B-cell type not achieve remission | Cancer | C9130 |
| C91.31 | DX10 | Prolymphocytic leukemia of B-cell type, in remission | Cancer | C9131 |
| C91.32 | DX10 | Prolymphocytic leukemia of B-cell type, in relapse | Cancer | C9132 |
| C91.4 | DX10 | Hairy cell leukemia | Cancer | C914 |
| C91.40 | DX10 | Hairy cell leukemia not having achieved remission | Cancer | C9140 |
| C91.41 | DX10 | Hairy cell leukemia, in remission | Cancer | C9141 |
| C91.42 | DX10 | Hairy cell leukemia, in relapse | Cancer | C9142 |
| C91.5 | DX10 | Adult T-cell lymphoma/leukemia (HTLV-1-associated) | Cancer | C915 |
| C91.50 | DX10 | Adult T-cell lymph/leuk (HTLV-1-assoc) not achieve remission | Cancer | C9150 |
| C91.51 | DX10 | Adult T-cell lymphoma/leukemia (HTLV-1-assoc), in remission | Cancer | C9151 |
| C91.52 | DX10 | Adult T-cell lymphoma/leukemia (HTLV-1-assoc), in relapse | Cancer | C9152 |
| C91.6 | DX10 | Prolymphocytic leukemia of T-cell type | Cancer | C916 |

| | | | | |
|---|---|---|---|---|
| C91.60 | DX10 | Prolymphocytic leukemia of T-cell type not achieve remission | Cancer | C9160 |
| C91.61 | DX10 | Prolymphocytic leukemia of T-cell type, in remission | Cancer | C9161 |
| C91.62 | DX10 | Prolymphocytic leukemia of T-cell type, in relapse | Cancer | C9162 |
| C91.A | DX10 | Mature B-cell leukemia Burkitt-type | Cancer | C91A |
| C91.A0 | DX10 | Mature B-cell leukemia Burkitt-type not achieve remission | Cancer | C91A0 |
| C91.A1 | DX10 | Mature B-cell leukemia Burkitt-type, in remission | Cancer | C91A1 |
| C91.A2 | DX10 | Mature B-cell leukemia Burkitt-type, in relapse | Cancer | C91A2 |
| C91.Z | DX10 | Other lymphoid leukemia | Cancer | C91Z |
| C91.Z0 | DX10 | Other lymphoid leukemia not having achieved remission | Cancer | C91Z0 |
| C91.Z1 | DX10 | Other lymphoid leukemia, in remission | Cancer | C91Z1 |
| C91.Z2 | DX10 | Other lymphoid leukemia, in relapse | Cancer | C91Z2 |
| C91.9 | DX10 | Lymphoid leukemia, unspecified | Cancer | C919 |
| C91.90 | DX10 | Lymphoid leukemia, unspecified not having achieved remission | Cancer | C9190 |
| C91.91 | DX10 | Lymphoid leukemia, unspecified, in remission | Cancer | C9191 |
| C91.92 | DX10 | Lymphoid leukemia, unspecified, in relapse | Cancer | C9192 |
| C92 | DX10 | Myeloid leukemia | Cancer | C92 |
| C92.0 | DX10 | Acute myeloblastic leukemia | Cancer | C920 |
| C92.00 | DX10 | Acute myeloblastic leukemia, not having achieved remission | Cancer | C9200 |
| C92.01 | DX10 | Acute myeloblastic leukemia, in remission | Cancer | C9201 |
| C92.02 | DX10 | Acute myeloblastic leukemia, in relapse | Cancer | C9202 |
| C92.1 | DX10 | Chronic myeloid leukemia, BCR/ABL-positive | Cancer | C921 |
| C92.10 | DX10 | Chronic myeloid leuk, BCR/ABL-positive, not achieve remis | Cancer | C9210 |
| C92.11 | DX10 | Chronic myeloid leukemia, BCR/ABL-positive, in remission | Cancer | C9211 |
| C92.12 | DX10 | Chronic myeloid leukemia, BCR/ABL-positive, in relapse | Cancer | C9212 |
| C92.2 | DX10 | Atypical chronic myeloid leukemia, BCR/ABL-negative | Cancer | C922 |
| C92.20 | DX10 | Atyp chronic myeloid leuk, BCR/ABL-neg, not achieve remis | Cancer | C9220 |
| C92.21 | DX10 | Atypical chronic myeloid leukemia, BCR/ABL-neg, in remission | Cancer | C9221 |
| C92.22 | DX10 | Atypical chronic myeloid leukemia, BCR/ABL-neg, in relapse | Cancer | C9222 |
| C92.3 | DX10 | Myeloid sarcoma | Cancer | C923 |
| C92.30 | DX10 | Myeloid sarcoma, not having achieved remission | Cancer | C9230 |
| C92.31 | DX10 | Myeloid sarcoma, in remission | Cancer | C9231 |
| C92.32 | DX10 | Myeloid sarcoma, in relapse | Cancer | C9232 |
| C92.4 | DX10 | Acute promyelocytic leukemia | Cancer | C924 |
| C92.40 | DX10 | Acute promyelocytic leukemia, not having achieved remission | Cancer | C9240 |
| C92.41 | DX10 | Acute promyelocytic leukemia, in remission | Cancer | C9241 |
| C92.42 | DX10 | Acute promyelocytic leukemia, in relapse | Cancer | C9242 |
| C92.5 | DX10 | Acute myelomonocytic leukemia | Cancer | C925 |
| C92.50 | DX10 | Acute myelomonocytic leukemia, not having achieved remission | Cancer | C9250 |
| C92.51 | DX10 | Acute myelomonocytic leukemia, in remission | Cancer | C9251 |
| C92.52 | DX10 | Acute myelomonocytic leukemia, in relapse | Cancer | C9252 |
| C92.6 | DX10 | Acute myeloid leukemia with 11q23-abnormality | Cancer | C926 |
| C92.60 | DX10 | Acute myeloid leukemia w 11q23-abnormality not achieve remis | Cancer | C9260 |
| C92.61 | DX10 | Acute myeloid leukemia with 11q23-abnormality in remission | Cancer | C9261 |
| C92.62 | DX10 | Acute myeloid leukemia with 11q23-abnormality in relapse | Cancer | C9262 |

| | | | | |
|---|---|---|---|---|
| C92.A | DX10 | Acute myeloid leukemia with multilineage dysplasia | Cancer | C92A |
| C92.A0 | DX10 | Acute myeloid leuk w multilin dysplasia, not achieve remis | Cancer | C92A0 |
| C92.A1 | DX10 | Acute myeloid leukemia w multilin dysplasia, in remission | Cancer | C92A1 |
| C92.A2 | DX10 | Acute myeloid leukemia w multilineage dysplasia, in relapse | Cancer | C92A2 |
| C92.Z | DX10 | Other myeloid leukemia | Cancer | C92Z |
| C92.Z0 | DX10 | Other myeloid leukemia not having achieved remission | Cancer | C92Z0 |
| C92.Z1 | DX10 | Other myeloid leukemia, in remission | Cancer | C92Z1 |
| C92.Z2 | DX10 | Other myeloid leukemia, in relapse | Cancer | C92Z2 |
| C92.9 | DX10 | Myeloid leukemia, unspecified | Cancer | C929 |
| C92.90 | DX10 | Myeloid leukemia, unspecified, not having achieved remission | Cancer | C9290 |
| C92.91 | DX10 | Myeloid leukemia, unspecified in remission | Cancer | C9291 |
| C92.92 | DX10 | Myeloid leukemia, unspecified in relapse | Cancer | C9292 |
| C93 | DX10 | Monocytic leukemia | Cancer | C93 |
| C93.0 | DX10 | Acute monoblastic/monocytic leukemia | Cancer | C930 |
| C93.00 | DX10 | Acute monoblastic/monocytic leukemia, not achieve remission | Cancer | C9300 |
| C93.01 | DX10 | Acute monoblastic/monocytic leukemia, in remission | Cancer | C9301 |
| C93.02 | DX10 | Acute monoblastic/monocytic leukemia, in relapse | Cancer | C9302 |
| C93.1 | DX10 | Chronic myelomonocytic leukemia | Cancer | C931 |
| C93.10 | DX10 | Chronic myelomonocytic leukemia not achieve remission | Cancer | C9310 |
| C93.11 | DX10 | Chronic myelomonocytic leukemia, in remission | Cancer | C9311 |
| C93.12 | DX10 | Chronic myelomonocytic leukemia, in relapse | Cancer | C9312 |
| C93.3 | DX10 | Juvenile myelomonocytic leukemia | Cancer | C933 |
| C93.30 | DX10 | Juvenile myelomonocytic leukemia, not achieve remission | Cancer | C9330 |
| C93.31 | DX10 | Juvenile myelomonocytic leukemia, in remission | Cancer | C9331 |
| C93.32 | DX10 | Juvenile myelomonocytic leukemia, in relapse | Cancer | C9332 |
| C93.Z | DX10 | Other monocytic leukemia | Cancer | C93Z |
| C93.Z0 | DX10 | Other monocytic leukemia, not having achieved remission | Cancer | C93Z0 |
| C93.Z1 | DX10 | Other monocytic leukemia, in remission | Cancer | C93Z1 |
| C93.Z2 | DX10 | Other monocytic leukemia, in relapse | Cancer | C93Z2 |
| C93.9 | DX10 | Monocytic leukemia, unspecified | Cancer | C939 |
| C93.90 | DX10 | Monocytic leukemia, unsp, not having achieved remission | Cancer | C9390 |
| C93.91 | DX10 | Monocytic leukemia, unspecified in remission | Cancer | C9391 |
| C93.92 | DX10 | Monocytic leukemia, unspecified in relapse | Cancer | C9392 |
| C94 | DX10 | Other leukemias of specified cell type | Cancer | C94 |
| C94.0 | DX10 | Acute erythroid leukemia | Cancer | C940 |
| C94.00 | DX10 | Acute erythroid leukemia, not having achieved remission | Cancer | C9400 |
| C94.01 | DX10 | Acute erythroid leukemia, in remission | Cancer | C9401 |
| C94.02 | DX10 | Acute erythroid leukemia, in relapse | Cancer | C9402 |
| C94.2 | DX10 | Acute megakaryoblastic leukemia | Cancer | C942 |
| C94.20 | DX10 | Acute megakaryoblastic leukemia not achieve remission | Cancer | C9420 |
| C94.21 | DX10 | Acute megakaryoblastic leukemia, in remission | Cancer | C9421 |
| C94.22 | DX10 | Acute megakaryoblastic leukemia, in relapse | Cancer | C9422 |
| C94.3 | DX10 | Mast cell leukemia | Cancer | C943 |
| C94.30 | DX10 | Mast cell leukemia not having achieved remission | Cancer | C9430 |
| C94.31 | DX10 | Mast cell leukemia, in remission | Cancer | C9431 |
| C94.32 | DX10 | Mast cell leukemia, in relapse | Cancer | C9432 |
| C94.4 | DX10 | Acute panmyelosis with myelofibrosis | Cancer | C944 |
| C94.40 | DX10 | Acute panmyelosis w myelofibrosis not achieve remission | Cancer | C9440 |

| | | | | |
|---|---|---|---|---|
| C94.41 | DX10 | Acute panmyelosis with myelofibrosis, in remission | Cancer | C9441 |
| C94.42 | DX10 | Acute panmyelosis with myelofibrosis, in relapse | Cancer | C9442 |
| C94.6 | DX10 | Myelodysplastic disease, not classified | Cancer | C946 |
| C94.8 | DX10 | Other specified leukemias | Cancer | C948 |
| C94.80 | DX10 | Other specified leukemias not having achieved remission | Cancer | C9480 |
| C94.81 | DX10 | Other specified leukemias, in remission | Cancer | C9481 |
| C94.82 | DX10 | Other specified leukemias, in relapse | Cancer | C9482 |
| C95 | DX10 | Leukemia of unspecified cell type | Cancer | C95 |
| C95.0 | DX10 | Acute leukemia of unspecified cell type | Cancer | C950 |
| C95.00 | DX10 | Acute leukemia of unsp cell type not achieve remission | Cancer | C9500 |
| C95.01 | DX10 | Acute leukemia of unspecified cell type, in remission | Cancer | C9501 |
| C95.02 | DX10 | Acute leukemia of unspecified cell type, in relapse | Cancer | C9502 |
| C95.1 | DX10 | Chronic leukemia of unspecified cell type | Cancer | C951 |
| C95.10 | DX10 | Chronic leukemia of unsp cell type not achieve remission | Cancer | C9510 |
| C95.11 | DX10 | Chronic leukemia of unspecified cell type, in remission | Cancer | C9511 |
| C95.12 | DX10 | Chronic leukemia of unspecified cell type, in relapse | Cancer | C9512 |
| C95.9 | DX10 | Leukemia, unspecified | Cancer | C959 |
| C95.90 | DX10 | Leukemia, unspecified not having achieved remission | Cancer | C9590 |
| C95.91 | DX10 | Leukemia, unspecified, in remission | Cancer | C9591 |
| C95.92 | DX10 | Leukemia, unspecified, in relapse | Cancer | C9592 |
| C96 | DX10 | Oth & unsp malig neoplm of lymphoid, hematpoetc and rel tiss | Cancer | C96 |
| C96.0 | DX10 | Multifocal and multisystemic Langerhans-cell histiocytosis | Cancer | C960 |
| C96.2 | DX10 | Malignant mast cell tumor | Cancer | C962 |
| C96.20 | DX10 | | Cancer | C9620 |
| C96.21 | DX10 | | Cancer | C9621 |
| C96.22 | DX10 | | Cancer | C9622 |
| C96.29 | DX10 | | Cancer | C9629 |
| C96.4 | DX10 | Sarcoma of dendritic cells (accessory cells) | Cancer | C964 |
| C96.5 | DX10 | Multifocal and unisystemic Langerhans-cell histiocytosis | Cancer | C965 |
| C96.6 | DX10 | Unifocal Langerhans-cell histiocytosis | Cancer | C966 |
| C96.A | DX10 | Histiocytic sarcoma | Cancer | C96A |
| C96.Z | DX10 | Oth malig neoplm of lymphoid, hematpoetc and related tissue | Cancer | C96Z |
| C96.9 | DX10 | Malig neoplm of lymphoid, hematpoetc and rel tissue, unsp | Cancer | C969 |
| D09.0 | DX10 | Carcinoma in situ of bladder | Cancer | D090 |
| D45 | DX10 | Polycythemia vera | Cancer | D45 |
| C4A | DX10 | Merkel Cell Carcinoma | Cancer | C4A |
| C4A.0 | DX10 | Merkel cell carcinoma of lip | Cancer | C4A0 |
| C4A.1 | DX10 | Merkel cell carcinoma of eyelid, including canthus | Cancer | C4A1 |
| C4A.2 | DX10 | Merkel cell carcinoma of ear and external auricular canal | Cancer | C4A2 |
| C4A.3 | DX10 | Merkel cell carcinoma of unspecified part sof face | Cancer | C4A3 |
| C4A.4 | DX10 | Merkel cell carcinoma of scalp and neck | Cancer | C4A4 |
| C4A.5 | DX10 | Merkel cell carcinoma of trunk | Cancer | C4A5 |
| C4A.6 | DX10 | Merkel cell carcinoma of upper limb, including shoulder | Cancer | C4A6 |
| C4A.7 | DX10 | Merkel cell carcinoma of skin of lower limb, including hip | Cancer | C4A7 |
| C4A.8 | DX10 | Merkel cell carcinoma of overlapping sites | Cancer | C4A8 |
| C4A.9 | DX10 | Merkel cell carcinoma, unspecified | Cancer | C4A9 |
| C94.1 | DX10 | Chronic erythremia | Cancer | C941 |
| I60 | DX10 | Nontraumatic subarachnoid hemorrhage | Cerebrovascular_Disease | I60 |
| I60.0 | DX10 | Ntrm subarach hemorrhage from carotid siphon and bifurcation | Cerebrovascular_Disease | I600 |

| | | | | |
|---|---|---|---|---|
| I60.00 | DX10 | Ntrm subarach hemorrhage from unsp carotid siphon and bifurc | Cerebrovascular_Disease | I6000 |
| I60.01 | DX10 | Ntrm subarach hemor from right carotid siphon and bifurc | Cerebrovascular_Disease | I6001 |
| I60.02 | DX10 | Ntrm subarach hemorrhage from left carotid siphon and bifurc | Cerebrovascular_Disease | I6002 |
| I60.1 | DX10 | Ntrm subarachnoid hemorrhage from middle cerebral artery | Cerebrovascular_Disease | I601 |
| I60.10 | DX10 | Ntrm subarach hemorrhage from unsp middle cerebral artery | Cerebrovascular_Disease | I6010 |
| I60.11 | DX10 | Ntrm subarach hemorrhage from right middle cerebral artery | Cerebrovascular_Disease | I6011 |
| I60.12 | DX10 | Ntrm subarach hemorrhage from left middle cerebral artery | Cerebrovascular_Disease | I6012 |
| I60.2 | DX10 | Ntrm subarach hemorrhage from anterior communicating artery | Cerebrovascular_Disease | I602 |
| I60.20 | DX10 | Ntrm subarach hemor from unsp anterior communicating artery | Cerebrovascular_Disease | I6020 |
| I60.21 | DX10 | Ntrm subarach hemor from right anterior communicating artery | Cerebrovascular_Disease | I6021 |
| I60.22 | DX10 | Ntrm subarach hemor from left anterior communicating artery | Cerebrovascular_Disease | I6022 |
| I60.3 | DX10 | Ntrm subarach hemorrhage from posterior communicating artery | Cerebrovascular_Disease | I603 |
| I60.30 | DX10 | Ntrm subarach hemor from unsp posterior communicating artery | Cerebrovascular_Disease | I6030 |
| I60.31 | DX10 | Ntrm subarach hemor from right post communicating artery | Cerebrovascular_Disease | I6031 |
| I60.32 | DX10 | Ntrm subarach hemor from left posterior communicating artery | Cerebrovascular_Disease | I6032 |
| I60.4 | DX10 | Nontraumatic subarachnoid hemorrhage from basilar artery | Cerebrovascular_Disease | I604 |
| I60.5 | DX10 | Nontraumatic subarachnoid hemorrhage from vertebral artery | Cerebrovascular_Disease | I605 |
| I60.50 | DX10 | Nontraumatic subarachnoid hemorrhage from unsp verteb art | Cerebrovascular_Disease | I6050 |
| I60.51 | DX10 | Nontraumatic subarachnoid hemorrhage from r verteb art | Cerebrovascular_Disease | I6051 |
| I60.52 | DX10 | Nontraumatic subarachnoid hemorrhage from l verteb art | Cerebrovascular_Disease | I6052 |
| I60.6 | DX10 | Nontraumatic subarachnoid hemorrhage from oth intracran art | Cerebrovascular_Disease | I606 |
| I60.7 | DX10 | Nontraumatic subarachnoid hemorrhage from unsp intracran art | Cerebrovascular_Disease | I607 |
| I60.8 | DX10 | Other nontraumatic subarachnoid hemorrhage | Cerebrovascular_Disease | I608 |
| I60.9 | DX10 | Nontraumatic subarachnoid hemorrhage, unspecified | Cerebrovascular_Disease | I609 |
| I61 | DX10 | Nontraumatic intracerebral hemorrhage | Cerebrovascular_Disease | I61 |
| I61.0 | DX10 | Nontraumatic intcrbl hemorrhage in hemisphere, subcortical | Cerebrovascular_Disease | I610 |
| I61.1 | DX10 | Nontraumatic intcrbl hemorrhage in hemisphere, cortical | Cerebrovascular_Disease | I611 |
| I61.2 | DX10 | Nontraumatic intracerebral hemorrhage in hemisphere, unsp | Cerebrovascular_Disease | I612 |
| I61.3 | DX10 | Nontraumatic intracerebral hemorrhage in brain stem | Cerebrovascular_Disease | I613 |

| | | | | |
|---|---|---|---|---|
| I61.4 | DX10 | Nontraumatic intracerebral hemorrhage in cerebellum | Cerebrovascular_Disease | I614 |
| I61.5 | DX10 | Nontraumatic intracerebral hemorrhage, intraventricular | Cerebrovascular_Disease | I615 |
| I61.6 | DX10 | Nontraumatic intracerebral hemorrhage, multiple localized | Cerebrovascular_Disease | I616 |
| I61.8 | DX10 | Other nontraumatic intracerebral hemorrhage | Cerebrovascular_Disease | I618 |
| I61.9 | DX10 | Nontraumatic intracerebral hemorrhage, unspecified | Cerebrovascular_Disease | I619 |
| I62 | DX10 | Other and unspecified nontraumatic intracranial hemorrhage | Cerebrovascular_Disease | I62 |
| I62.0 | DX10 | Nontraumatic subdural hemorrhage | Cerebrovascular_Disease | I620 |
| I62.00 | DX10 | Nontraumatic subdural hemorrhage, unspecified | Cerebrovascular_Disease | I6200 |
| I62.01 | DX10 | Nontraumatic acute subdural hemorrhage | Cerebrovascular_Disease | I6201 |
| I62.02 | DX10 | Nontraumatic subacute subdural hemorrhage | Cerebrovascular_Disease | I6202 |
| I62.03 | DX10 | Nontraumatic chronic subdural hemorrhage | Cerebrovascular_Disease | I6203 |
| I62.1 | DX10 | Nontraumatic extradural hemorrhage | Cerebrovascular_Disease | I621 |
| I62.9 | DX10 | Nontraumatic intracranial hemorrhage, unspecified | Cerebrovascular_Disease | I629 |
| I63 | DX10 | Cerebral infarction | Cerebrovascular_Disease | I63 |
| I63.0 | DX10 | Cerebral infarction due to thrombosis of precerebral arteries | Cerebrovascular_Disease | I630 |
| I63.00 | DX10 | Cerebral infarction due to thombos unsp precerebral artery | Cerebrovascular_Disease | I6300 |
| I63.01 | DX10 | Cerebral infarction due to thrombosis of vertebral artery | Cerebrovascular_Disease | I6301 |
| I63.011 | DX10 | Cerebral infarction due to thrombosis of r verteb art | Cerebrovascular_Disease | I63011 |
| I63.012 | DX10 | Cerebral infarction due to thrombosis of l verteb art | Cerebrovascular_Disease | I63012 |
| I63.013 | DX10 | Cerebral infrc due to thrombosis of bilateral verteb art | Cerebrovascular_Disease | I63013 |
| I63.019 | DX10 | Cerebral infarction due to thombos unsp vertebral artery | Cerebrovascular_Disease | I63019 |
| I63.02 | DX10 | Cerebral infarction due to thrombosis of basilar artery | Cerebrovascular_Disease | I6302 |
| I63.03 | DX10 | Cerebral infarction due to thrombosis of carotid artery | Cerebrovascular_Disease | I6303 |
| I63.031 | DX10 | Cerebral infrc due to thrombosis of right carotid artery | Cerebrovascular_Disease | I63031 |
| I63.032 | DX10 | Cerebral infarction due to thrombosis of left carotid artery | Cerebrovascular_Disease | I63032 |
| I63.033 | DX10 | Cerebral infrc due to thombos of bilateral carotid arteries | Cerebrovascular_Disease | I63033 |
| I63.039 | DX10 | Cerebral infarction due to thrombosis of unsp carotid artery | Cerebrovascular_Disease | I63039 |
| I63.09 | DX10 | Cerebral infarction due to thrombosis of precerebral artery | Cerebrovascular_Disease | I6309 |
| I63.1 | DX10 | Cerebral infarction due to embolism of precerebral arteries | Cerebrovascular_Disease | I631 |

| | | | | |
|---|---|---|---|---|
| I63.10 | DX10 | Cerebral infarction due to embolism of precerebral arteries | Cerebrovascular_Disease | I6310 |
| I63.11 | DX10 | Cerebral infarction due to embolism of vertebral artery | Cerebrovascular_Disease | I6311 |
| I63.111 | DX10 | Cerebral infarction due to embolism of r verteb art | Cerebrovascular_Disease | I63111 |
| I63.112 | DX10 | Cerebral infarction due to embolism of left vertebral artery | Cerebrovascular_Disease | I63112 |
| I63.113 | DX10 | Cerebral infarction due to embolism of bilateral verteb art | Cerebrovascular_Disease | I63113 |
| I63.119 | DX10 | Cerebral infarction due to embolism of unsp vertebral artery | Cerebrovascular_Disease | I63119 |
| I63.12 | DX10 | Cerebral infarction due to embolism of basilar artery | Cerebrovascular_Disease | I6312 |
| I63.13 | DX10 | Cerebral infarction due to embolism of carotid artery | Cerebrovascular_Disease | I6313 |
| I63.131 | DX10 | Cerebral infarction due to embolism of right carotid artery | Cerebrovascular_Disease | I63131 |
| I63.132 | DX10 | Cerebral infarction due to embolism of left carotid artery | Cerebrovascular_Disease | I63132 |
| I63.133 | DX10 | Cerebral infrc due to embolism of bilateral carotid arteries | Cerebrovascular_Disease | I63133 |
| I63.139 | DX10 | Cerebral infarction due to embolism of unsp carotid artery | Cerebrovascular_Disease | I63139 |
| I63.19 | DX10 | Cerebral infarction due to embolism of precerebral artery | Cerebrovascular_Disease | I6319 |
| I63.2 | DX10 | Cerebral infrc due to unsp occls or stenosis of precerb art | Cerebrovascular_Disease | I632 |
| I63.20 | DX10 | Cerebral infrc due to unsp occls or stenosis of precerb art | Cerebrovascular_Disease | I6320 |
| I63.21 | DX10 | Cerebral infrc due to unsp occls or stenosis of verteb art | Cerebrovascular_Disease | I6321 |
| I63.211 | DX10 | Cereb infrc due to unsp occls or stenos of right verteb art | Cerebrovascular_Disease | I63211 |
| I63.212 | DX10 | Cereb infrc due to unsp occls or stenosis of left verteb art | Cerebrovascular_Disease | I63212 |
| I63.213 | DX10 | Cereb infrc due to unsp occls or stenosis of bi verteb art | Cerebrovascular_Disease | I63213 |
| I63.219 | DX10 | Cereb infrc due to unsp occls or stenosis of unsp verteb art | Cerebrovascular_Disease | I63219 |
| I63.22 | DX10 | Cerebral infrc due to unsp occls or stenosis of basilar art | Cerebrovascular_Disease | I6322 |
| I63.23 | DX10 | Cerebral infrc due to unsp occls or stenosis of carotid art | Cerebrovascular_Disease | I6323 |
| I63.231 | DX10 | Cereb infrc due to unsp occls or stenos of right carotid art | Cerebrovascular_Disease | I63231 |
| I63.232 | DX10 | Cereb infrc due to unsp occls or stenos of left carotid art | Cerebrovascular_Disease | I63232 |
| I63.233 | DX10 | Cereb infrc due to unsp occls or stenosis of bi carotid art | Cerebrovascular_Disease | I63233 |
| I63.239 | DX10 | Cereb infrc due to unsp occls or stenos of unsp carotid art | Cerebrovascular_Disease | I63239 |
| I63.29 | DX10 | Cerebral infrc due to unsp occls or stenosis of precerb art | Cerebrovascular_Disease | I6329 |
| I63.3 | DX10 | Cerebral infarction due to thrombosis of cerebral arteries | Cerebrovascular_Disease | I633 |
| I63.30 | DX10 | Cerebral infarction due to thrombosis of cerebral arteries | Cerebrovascular_Disease | I6330 |

| | | | | |
|---|---|---|---|---|
| I63.31 | DX10 | Cerebral infrc due to thrombosis of middle cerebral artery | Cerebrovascular_Disease | I6331 |
| I63.311 | DX10 | Cereb infrc due to thombos of right middle cerebral artery | Cerebrovascular_Disease | I63311 |
| I63.312 | DX10 | Cerebral infrc due to thombos of left middle cerebral artery | Cerebrovascular_Disease | I63312 |
| I63.313 | DX10 | Cerebral infrc due to thombos of bi middle cerebral arteries | Cerebrovascular_Disease | I63313 |
| I63.319 | DX10 | Cerebral infrc due to thombos unsp middle cerebral artery | Cerebrovascular_Disease | I63319 |
| I63.32 | DX10 | Cerebral infrc due to thrombosis of anterior cerebral artery | Cerebrovascular_Disease | I6332 |
| I63.321 | DX10 | Cerebral infrc due to thombos of right ant cerebral artery | Cerebrovascular_Disease | I63321 |
| I63.322 | DX10 | Cerebral infrc due to thombos of left ant cerebral artery | Cerebrovascular_Disease | I63322 |
| I63.323 | DX10 | Cerebral infrc due to thombos of bilateral anterior arteries | Cerebrovascular_Disease | I63323 |
| I63.329 | DX10 | Cerebral infrc due to thombos unsp anterior cerebral artery | Cerebrovascular_Disease | I63329 |
| I63.33 | DX10 | Cerebral infrc due to thombos of posterior cerebral artery | Cerebrovascular_Disease | I6333 |
| I63.331 | DX10 | Cerebral infrc due to thombos of right post cerebral artery | Cerebrovascular_Disease | I63331 |
| I63.332 | DX10 | Cerebral infrc due to thombos of left post cerebral artery | Cerebrovascular_Disease | I63332 |
| I63.333 | DX10 | Cerebral infrc to thrombosis of bilateral posterior arteries | Cerebrovascular_Disease | I63333 |
| I63.339 | DX10 | Cerebral infrc due to thombos unsp posterior cerebral artery | Cerebrovascular_Disease | I63339 |
| I63.34 | DX10 | Cerebral infarction due to thrombosis of cerebellar artery | Cerebrovascular_Disease | I6334 |
| I63.341 | DX10 | Cerebral infrc due to thrombosis of right cereblr artery | Cerebrovascular_Disease | I63341 |
| I63.342 | DX10 | Cerebral infarction due to thrombosis of left cereblr artery | Cerebrovascular_Disease | I63342 |
| I63.343 | DX10 | Cerebral infrc to thrombosis of bilateral cereblr arteries | Cerebrovascular_Disease | I63343 |
| I63.349 | DX10 | Cerebral infarction due to thombos unsp cerebellar artery | Cerebrovascular_Disease | I63349 |
| I63.39 | DX10 | Cerebral infarction due to thrombosis of oth cerebral artery | Cerebrovascular_Disease | I6339 |
| I63.4 | DX10 | Cerebral infarction due to embolism of cerebral arteries | Cerebrovascular_Disease | I634 |
| I63.40 | DX10 | Cerebral infarction due to embolism of cerebral arteries | Cerebrovascular_Disease | I6340 |
| I63.41 | DX10 | Cerebral infrc due to embolism of middle cerebral artery | Cerebrovascular_Disease | I6341 |
| I63.411 | DX10 | Cereb infrc due to embolism of right middle cerebral artery | Cerebrovascular_Disease | I63411 |
| I63.412 | DX10 | Cereb infrc due to embolism of left middle cerebral artery | Cerebrovascular_Disease | I63412 |
| I63.413 | DX10 | Cerebral infrc due to embolism of bi middle cerebral art | Cerebrovascular_Disease | I63413 |
| I63.419 | DX10 | Cereb infrc due to embolism of unsp middle cerebral artery | Cerebrovascular_Disease | I63419 |
| I63.42 | DX10 | Cerebral infrc due to embolism of anterior cerebral artery | Cerebrovascular_Disease | I6342 |

| | | | | |
|---|---|---|---|---|
| I63.421 | DX10 | Cerebral infrc due to embolism of right ant cerebral artery | Cerebrovascular_Disease | I63421 |
| I63.422 | DX10 | Cerebral infrc due to embolism of left ant cerebral artery | Cerebrovascular_Disease | I63422 |
| I63.423 | DX10 | Cerebral infrc due to embolism of bi ant cerebral arteries | Cerebrovascular_Disease | I63423 |
| I63.429 | DX10 | Cerebral infrc due to embolism of unsp ant cerebral artery | Cerebrovascular_Disease | I63429 |
| I63.43 | DX10 | Cerebral infrc due to embolism of posterior cerebral artery | Cerebrovascular_Disease | I6343 |
| I63.431 | DX10 | Cerebral infrc due to embolism of right post cerebral artery | Cerebrovascular_Disease | I63431 |
| I63.432 | DX10 | Cerebral infrc due to embolism of left post cerebral artery | Cerebrovascular_Disease | I63432 |
| I63.433 | DX10 | Cerebral infrc due to embolism of bi post cerebral arteries | Cerebrovascular_Disease | I63433 |
| I63.439 | DX10 | Cerebral infrc due to embolism of unsp post cerebral artery | Cerebrovascular_Disease | I63439 |
| I63.44 | DX10 | Cerebral infarction due to embolism of cerebellar artery | Cerebrovascular_Disease | I6344 |
| I63.441 | DX10 | Cerebral infarction due to embolism of right cereblr artery | Cerebrovascular_Disease | I63441 |
| I63.442 | DX10 | Cerebral infarction due to embolism of left cereblr artery | Cerebrovascular_Disease | I63442 |
| I63.443 | DX10 | Cerebral infrc due to embolism of bilateral cereblr arteries | Cerebrovascular_Disease | I63443 |
| I63.449 | DX10 | Cerebral infarction due to embolism of unsp cereblr artery | Cerebrovascular_Disease | I63449 |
| I63.49 | DX10 | Cerebral infarction due to embolism of other cerebral artery | Cerebrovascular_Disease | I6349 |
| I63.5 | DX10 | Cerebral infrc due to unsp occls or stenosis of cerebral art | Cerebrovascular_Disease | I635 |
| I63.50 | DX10 | Cerebral infrc due to unsp occls or stenosis of cerebral art | Cerebrovascular_Disease | I6350 |
| I63.51 | DX10 | Cereb infrc due to unsp occls or stenos of middle cereb art | Cerebrovascular_Disease | I6351 |
| I63.511 | DX10 | Cereb infrc d/t unsp occls or stenos of right mid cereb art | Cerebrovascular_Disease | I63511 |
| I63.512 | DX10 | Cereb infrc d/t unsp occls or stenos of left mid cereb art | Cerebrovascular_Disease | I63512 |
| I63.513 | DX10 | Cereb infrc due to unsp occls or stenosis of bi middle art | Cerebrovascular_Disease | I63513 |
| I63.519 | DX10 | Cereb infrc d/t unsp occls or stenos of unsp mid cereb art | Cerebrovascular_Disease | I63519 |
| I63.52 | DX10 | Cereb infrc due to unsp occls or stenos of ant cereb artery | Cerebrovascular_Disease | I6352 |
| I63.521 | DX10 | Cereb infrc d/t unsp occls or stenos of right ant cereb art | Cerebrovascular_Disease | I63521 |
| I63.522 | DX10 | Cereb infrc d/t unsp occls or stenos of left ant cereb art | Cerebrovascular_Disease | I63522 |
| I63.523 | DX10 | Cerebral infrc due to unsp occls or stenosis of bi ant art | Cerebrovascular_Disease | I63523 |
| I63.529 | DX10 | Cereb infrc d/t unsp occls or stenos of unsp ant cereb art | Cerebrovascular_Disease | I63529 |
| I63.53 | DX10 | Cereb infrc due to unsp occls or stenos of post cereb artery | Cerebrovascular_Disease | I6353 |
| I63.531 | DX10 | Cereb infrc d/t unsp occls or stenos of right post cereb art | Cerebrovascular_Disease | I63531 |

| | | | | |
|---|---|---|---|---|
| I63.532 | DX10 | Cereb infrc d/t unsp occls or stenos of left post cereb art | Cerebrovascular_Disease | I63532 |
| I63.533 | DX10 | Cerebral infrc due to unsp occls or stenosis of bi post art | Cerebrovascular_Disease | I63533 |
| I63.539 | DX10 | Cereb infrc d/t unsp occls or stenos of unsp post cereb art | Cerebrovascular_Disease | I63539 |
| I63.54 | DX10 | Cereb infrc due to unsp occls or stenosis of cereblr artery | Cerebrovascular_Disease | I6354 |
| I63.541 | DX10 | Cereb infrc due to unsp occls or stenos of right cereblr art | Cerebrovascular_Disease | I63541 |
| I63.542 | DX10 | Cereb infrc due to unsp occls or stenos of left cereblr art | Cerebrovascular_Disease | I63542 |
| I63.543 | DX10 | Cereb infrc due to unsp occls or stenosis of bi cereblr art | Cerebrovascular_Disease | I63543 |
| I63.549 | DX10 | Cereb infrc due to unsp occls or stenos of unsp cereblr art | Cerebrovascular_Disease | I63549 |
| I63.59 | DX10 | Cereb infrc due to unsp occls or stenosis of cerebral artery | Cerebrovascular_Disease | I6359 |
| I65 | DX10 | Occls and stenosis of precerb art, not rslt in cereb infrc | Cerebrovascular_Disease | I65 |
| I65.0 | DX10 | Occls and stenosis of precerb art, not rslt in cereb infrc | Cerebrovascular_Disease | I650 |
| I65.01 | DX10 | Occlusion and stenosis of right vertebral artery | Cerebrovascular_Disease | I6501 |
| I65.02 | DX10 | Occlusion and stenosis of left vertebral artery | Cerebrovascular_Disease | I6502 |
| I65.03 | DX10 | Occlusion and stenosis of bilateral vertebral arteries | Cerebrovascular_Disease | I6503 |
| I65.09 | DX10 | Occlusion and stenosis of unspecified vertebral artery | Cerebrovascular_Disease | I6509 |
| I65.1 | DX10 | Occlusion and stenosis of basilar artery | Cerebrovascular_Disease | I651 |
| I65.2 | DX10 | Occlusion and stenosis of carotid artery | Cerebrovascular_Disease | I652 |
| I65.21 | DX10 | Occlusion and stenosis of right carotid artery | Cerebrovascular_Disease | I6521 |
| I65.22 | DX10 | Occlusion and stenosis of left carotid artery | Cerebrovascular_Disease | I6522 |
| I65.23 | DX10 | Occlusion and stenosis of bilateral carotid arteries | Cerebrovascular_Disease | I6523 |
| I65.29 | DX10 | Occlusion and stenosis of unspecified carotid artery | Cerebrovascular_Disease | I6529 |
| I65.8 | DX10 | Occlusion and stenosis of other precerebral arteries | Cerebrovascular_Disease | I658 |
| I65.9 | DX10 | Occlusion and stenosis of unspecified precerebral artery | Cerebrovascular_Disease | I659 |
| I66 | DX10 | Occls and stenosis of cereb art, not rslt in cerebral infrc | Cerebrovascular_Disease | I66 |
| I66.0 | DX10 | Occls and stenosis of middle cereb art | Cerebrovascular_Disease | I660 |
| I66.01 | DX10 | Occlusion and stenosis of right middle cerebral artery | Cerebrovascular_Disease | I6601 |
| I66.02 | DX10 | Occlusion and stenosis of left middle cerebral artery | Cerebrovascular_Disease | I6602 |
| I66.03 | DX10 | Occlusion and stenosis of bilateral middle cerebral arteries | Cerebrovascular_Disease | I6603 |
| I66.09 | DX10 | Occlusion and stenosis of unspecified middle cerebral artery | Cerebrovascular_Disease | I6609 |

| | | | | |
|---|---|---|---|---|
| I66.1 | DX10 | Occlusion and stenosis of anterior cerebral artery | Cerebrovascular_Disease | I661 |
| I66.11 | DX10 | Occlusion and stenosis of right anterior cerebral artery | Cerebrovascular_Disease | I6611 |
| I66.12 | DX10 | Occlusion and stenosis of left anterior cerebral artery | Cerebrovascular_Disease | I6612 |
| I66.13 | DX10 | Occlusion and stenosis of bi anterior cerebral arteries | Cerebrovascular_Disease | I6613 |
| I66.19 | DX10 | Occlusion and stenosis of unsp anterior cerebral artery | Cerebrovascular_Disease | I6619 |
| I66.2 | DX10 | Occlusion and stenosis of posterior cerebral artery | Cerebrovascular_Disease | I662 |
| I66.21 | DX10 | Occlusion and stenosis of right posterior cerebral artery | Cerebrovascular_Disease | I6621 |
| I66.22 | DX10 | Occlusion and stenosis of left posterior cerebral artery | Cerebrovascular_Disease | I6622 |
| I66.23 | DX10 | Occlusion and stenosis of bi posterior cerebral arteries | Cerebrovascular_Disease | I6623 |
| I66.29 | DX10 | Occlusion and stenosis of unsp posterior cerebral artery | Cerebrovascular_Disease | I6629 |
| I66.3 | DX10 | Occlusion and stenosis of cerebellar arteries | Cerebrovascular_Disease | I663 |
| I66.8 | DX10 | Occlusion and stenosis of other cerebral arteries | Cerebrovascular_Disease | I668 |
| I66.9 | DX10 | Occlusion and stenosis of unspecified cerebral artery | Cerebrovascular_Disease | I669 |
| I67.2 | DX10 | Cerebral atherosclerosis | Cerebrovascular_Disease | I672 |
| I67.81 | DX10 | Acute cerebrovascular insufficiency | Cerebrovascular_Disease | I6781 |
| I67.82 | DX10 | Cerebral ischemia | Cerebrovascular_Disease | I6782 |
| I67.83 | DX10 | Posterior reversible encephalopathy syndrome | Cerebrovascular_Disease | I6783 |
| I67.84 | DX10 | Cerebral vasospasm and vasoconstriction | Cerebrovascular_Disease | I6784 |
| I67.841 | DX10 | Reversible cerebrovascular vasoconstriction syndrome | Cerebrovascular_Disease | I67841 |
| I67.848 | DX10 | Other cerebrovascular vasospasm and vasoconstriction | Cerebrovascular_Disease | I67848 |
| I67.85 | DX10 | Hereditary cerebrovascular diseases | Cerebrovascular_Disease | I6785 |
| I67.850 | DX10 | Cerebral autosomal dominant arteriopathy with subcortical infarcts and leukoencephalopathy | Cerebrovascular_Disease | I67850 |
| I69 | DX10 | Sequelae of cerebrovascular disease | Cerebrovascular_Disease | I69 |
| I69.0 | DX10 | Sequelae of nontraumatic subarachnoid hemorrhage | Cerebrovascular_Disease | I690 |
| I69.00 | DX10 | Sequelae of nontraumatic subarachnoid hemorrhage, unspecified | Cerebrovascular_Disease | I6900 |
| I69.01 | DX10 | Cognitive deficits following ntrm subarachnoid hemorrhage | Cerebrovascular_Disease | I6901 |
| I69.011 | DX10 | Memory deficit following ntrm subarachnoid hemorrhage | Cerebrovascular_Disease | I69011 |
| I69.012 | DX10 | Vis def/sptl nglct following ntrm subarachnoid hemorrhage | Cerebrovascular_Disease | I69012 |
| I69.013 | DX10 | Psychomotor deficit following ntrm subarachnoid hemorrhage | Cerebrovascular_Disease | I69013 |

| | | | | |
|---|---|---|---|---|
| I69.014 | DX10 | Fntl lb and exec fcn def following ntrm subarach hemorrhage | Cerebrovascular_Disease | I69014 |
| I69.015 | DX10 | Cognitive social or emo def following ntrm subarach hemor | Cerebrovascular_Disease | I69015 |
| I69.018 | DX10 | Other symp and signs w cogn fnctns fol ntrm subarach hemor | Cerebrovascular_Disease | I69018 |
| I69.019 | DX10 | Unsp symp and signs w cogn fnctns fol ntrm subarach hemor | Cerebrovascular_Disease | I69019 |
| I69.02 | DX10 | Speech/lang deficits following ntrm subarachnoid hemorrhage | Cerebrovascular_Disease | I6902 |
| I69.021 | DX10 | Dysphasia following nontraumatic subarachnoid hemorrhage | Cerebrovascular_Disease | I69021 |
| I69.022 | DX10 | Dysarthria following nontraumatic subarachnoid hemorrhage | Cerebrovascular_Disease | I69022 |
| I69.023 | DX10 | Fluency disorder following ntrm subarachnoid hemorrhage | Cerebrovascular_Disease | I69023 |
| I69.028 | DX10 | Oth speech/lang deficits following ntrm subarach hemorrhage | Cerebrovascular_Disease | I69028 |
| I69.03 | DX10 | Monoplg upr lmb following ntrm subarachnoid hemorrhage | Cerebrovascular_Disease | I6903 |
| I69.031 | DX10 | Monoplg upr lmb fol ntrm subarach hemor aff right dom side | Cerebrovascular_Disease | I69031 |
| I69.032 | DX10 | Monoplg upr lmb fol ntrm subarach hemor aff left dom side | Cerebrovascular_Disease | I69032 |
| I69.033 | DX10 | Monoplg upr lmb fol ntrm subarach hemor aff r nondom side | Cerebrovascular_Disease | I69033 |
| I69.034 | DX10 | Monoplg upr lmb fol ntrm subarach hemor aff left nondom side | Cerebrovascular_Disease | I69034 |
| I69.039 | DX10 | Monoplg upr lmb following ntrm subarach hemor aff unsp side | Cerebrovascular_Disease | I69039 |
| I69.04 | DX10 | Monoplg low lmb following ntrm subarachnoid hemorrhage | Cerebrovascular_Disease | I6904 |
| I69.041 | DX10 | Monoplg low lmb fol ntrm subarach hemor aff right dom side | Cerebrovascular_Disease | I69041 |
| I69.042 | DX10 | Monoplg low lmb fol ntrm subarach hemor aff left dom side | Cerebrovascular_Disease | I69042 |
| I69.043 | DX10 | Monoplg low lmb fol ntrm subarach hemor aff r nondom side | Cerebrovascular_Disease | I69043 |
| I69.044 | DX10 | Monoplg low lmb fol ntrm subarach hemor aff left nondom side | Cerebrovascular_Disease | I69044 |
| I69.049 | DX10 | Monoplg low lmb following ntrm subarach hemor aff unsp side | Cerebrovascular_Disease | I69049 |
| I69.05 | DX10 | Hemiplga following nontraumatic subarachnoid hemorrhage | Cerebrovascular_Disease | I6905 |
| I69.051 | DX10 | Hemiplga fol ntrm subarach hemor aff right dominant side | Cerebrovascular_Disease | I69051 |
| I69.052 | DX10 | Hemiplga fol ntrm subarach hemor aff left dominant side | Cerebrovascular_Disease | I69052 |
| I69.053 | DX10 | Hemiplga following ntrm subarach hemor aff right nondom side | Cerebrovascular_Disease | I69053 |
| I69.054 | DX10 | Hemiplga following ntrm subarach hemor aff left nondom side | Cerebrovascular_Disease | I69054 |
| I69.059 | DX10 | Hemiplga following ntrm subarach hemor affecting unsp side | Cerebrovascular_Disease | I69059 |
| I69.06 | DX10 | Oth paralytic syndrome following ntrm subarach hemorrhage | Cerebrovascular_Disease | I6906 |
| I69.061 | DX10 | Oth parlyt synd fol ntrm subarach hemor aff right dom side | Cerebrovascular_Disease | I69061 |

| | | | | |
|---|---|---|---|---|
| I69.062 | DX10 | Oth parlyt synd fol ntrm subarach hemor aff left dom side | Cerebrovascular_Disease | I69062 |
| I69.063 | DX10 | Oth parlyt synd fol ntrm subarach hemor aff r nondom side | Cerebrovascular_Disease | I69063 |
| I69.064 | DX10 | Oth parlyt synd fol ntrm subarach hemor aff left nondom side | Cerebrovascular_Disease | I69064 |
| I69.065 | DX10 | Oth paralytic syndrome following ntrm subarach hemor, bi | Cerebrovascular_Disease | I69065 |
| I69.069 | DX10 | Oth paralytic syndrome fol ntrm subarach hemor aff unsp side | Cerebrovascular_Disease | I69069 |
| I69.09 | DX10 | Other sequelae of nontraumatic subarachnoid hemorrhage | Cerebrovascular_Disease | I6909 |
| I69.1 | DX10 | Sequelae of nontraumatic intracerebral hemorrhage | Cerebrovascular_Disease | I691 |
| I69.10 | DX10 | Sequelae of nontraumatic intracerebral hemorrhage, unspecified | Cerebrovascular_Disease | I6910 |
| I69.11 | DX10 | Cognitive deficits following nontraumatic intcrbl hemorrhage | Cerebrovascular_Disease | I6911 |
| I69.11 | DX10 | Cognitive deficits following nontraumatic intcrbl hemorrhage | Cerebrovascular_Disease | I6911 |
| I69.111 | DX10 | Memory deficit following nontraumatic intcrbl hemorrhage | Cerebrovascular_Disease | I69111 |
| I69.112 | DX10 | Vis def/sptl nglct following nontraumatic intcrbl hemorrhage | Cerebrovascular_Disease | I69112 |
| I69.113 | DX10 | Psychomotor deficit following ntrm intcrbl hemorrhage | Cerebrovascular_Disease | I69113 |
| I69.114 | DX10 | Fntl lb and exec fcn def following ntrm intcrbl hemorrhage | Cerebrovascular_Disease | I69114 |
| I69.115 | DX10 | Cognitive social or emo def following ntrm intcrbl hemor | Cerebrovascular_Disease | I69115 |
| I69.118 | DX10 | Other symp and signs w cogn fnctns fol ntrm intcrbl hemor | Cerebrovascular_Disease | I69118 |
| I69.119 | DX10 | Unsp symptoms and signs w cogn fnctns fol ntrm intcrbl hemor | Cerebrovascular_Disease | I69119 |
| I69.12 | DX10 | Speech/lang deficits following ntrm intcrbl hemorrhage | Cerebrovascular_Disease | I6912 |
| I69.12 | DX10 | Speech/lang deficits following ntrm intcrbl hemorrhage | Cerebrovascular_Disease | I6912 |
| I69.121 | DX10 | Dysphasia following nontraumatic intracerebral hemorrhage | Cerebrovascular_Disease | I69121 |
| I69.122 | DX10 | Dysarthria following nontraumatic intracerebral hemorrhage | Cerebrovascular_Disease | I69122 |
| I69.123 | DX10 | Fluency disorder following nontraumatic intcrbl hemorrhage | Cerebrovascular_Disease | I69123 |
| I69.128 | DX10 | Oth speech/lang deficits following ntrm intcrbl hemorrhage | Cerebrovascular_Disease | I69128 |
| I69.13 | DX10 | Monoplg upr lmb following nontraumatic intcrbl hemorrhage | Cerebrovascular_Disease | I6913 |
| I69.131 | DX10 | Monoplg upr lmb fol ntrm intcrbl hemor aff right dom side | Cerebrovascular_Disease | I69131 |
| I69.132 | DX10 | Monoplg upr lmb fol ntrm intcrbl hemor aff left dom side | Cerebrovascular_Disease | I69132 |
| I69.133 | DX10 | Monoplg upr lmb fol ntrm intcrbl hemor aff right nondom side | Cerebrovascular_Disease | I69133 |
| I69.134 | DX10 | Monoplg upr lmb fol ntrm intcrbl hemor aff left nondom side | Cerebrovascular_Disease | I69134 |
| I69.139 | DX10 | Monoplg upr lmb following ntrm intcrbl hemor aff unsp side | Cerebrovascular_Disease | I69139 |

| | | | | |
|---|---|---|---|---|
| I69.14 | DX10 | Monoplg low lmb following nontraumatic intcrbl hemorrhage | Cerebrovascular_Disease | I6914 |
| I69.141 | DX10 | Monoplg low lmb fol ntrm intcrbl hemor aff right dom side | Cerebrovascular_Disease | I69141 |
| I69.142 | DX10 | Monoplg low lmb fol ntrm intcrbl hemor aff left dom side | Cerebrovascular_Disease | I69142 |
| I69.143 | DX10 | Monoplg low lmb fol ntrm intcrbl hemor aff right nondom side | Cerebrovascular_Disease | I69143 |
| I69.144 | DX10 | Monoplg low lmb fol ntrm intcrbl hemor aff left nondom side | Cerebrovascular_Disease | I69144 |
| I69.149 | DX10 | Monoplg low lmb following ntrm intcrbl hemor aff unsp side | Cerebrovascular_Disease | I69149 |
| I69.15 | DX10 | Hemiplga following nontraumatic intracerebral hemorrhage | Cerebrovascular_Disease | I6915 |
| I69.151 | DX10 | Hemiplga fol ntrm intcrbl hemor aff right dominant side | Cerebrovascular_Disease | I69151 |
| I69.152 | DX10 | Hemiplga following ntrm intcrbl hemor aff left dominant side | Cerebrovascular_Disease | I69152 |
| I69.153 | DX10 | Hemiplga following ntrm intcrbl hemor aff right nondom side | Cerebrovascular_Disease | I69153 |
| I69.154 | DX10 | Hemiplga following ntrm intcrbl hemor aff left nondom side | Cerebrovascular_Disease | I69154 |
| I69.159 | DX10 | Hemiplga following ntrm intcrbl hemor affecting unsp side | Cerebrovascular_Disease | I69159 |
| I69.16 | DX10 | Oth paralytic syndrome following ntrm intcrbl hemorrhage | Cerebrovascular_Disease | I6916 |
| I69.161 | DX10 | Oth parlyt synd fol ntrm intcrbl hemor aff right dom side | Cerebrovascular_Disease | I69161 |
| I69.162 | DX10 | Oth parlyt syndrome fol ntrm intcrbl hemor aff left dom side | Cerebrovascular_Disease | I69162 |
| I69.163 | DX10 | Oth parlyt synd fol ntrm intcrbl hemor aff right nondom side | Cerebrovascular_Disease | I69163 |
| I69.164 | DX10 | Oth parlyt synd fol ntrm intcrbl hemor aff left nondom side | Cerebrovascular_Disease | I69164 |
| I69.165 | DX10 | Oth paralytic syndrome following ntrm intcrbl hemor, bi | Cerebrovascular_Disease | I69165 |
| I69.169 | DX10 | Oth paralytic syndrome following ntrm intcrbl hemor, unsp side | Cerebrovascular_Disease | I69169 |
| I69.19 | DX10 | Other sequelae of nontraumatic intracerebral hemorrhage | Cerebrovascular_Disease | I6919 |
| I69.19 | DX10 | Other sequelae of nontraumatic intracerebral hemorrhage | Cerebrovascular_Disease | I6919 |
| I69.2 | DX10 | Sequelae of other nontraumatic intracranial hemorrhage | Cerebrovascular_Disease | I692 |
| I69.20 | DX10 | Sequelae of other nontraumatic intracranial hemorrhage, unspecified | Cerebrovascular_Disease | I6920 |
| I69.21 | DX10 | Cognitive deficits following oth ntrm intcrn hemorrhage | Cerebrovascular_Disease | I6921 |
| I69.21 | DX10 | Cognitive deficits following oth ntrm intcrn hemorrhage | Cerebrovascular_Disease | I6921 |
| I69.211 | DX10 | Memory deficit following other ntrm intcrn hemorrhage | Cerebrovascular_Disease | I69211 |
| I69.212 | DX10 | Vis def/sptl nglct following other ntrm intcrn hemorrhage | Cerebrovascular_Disease | I69212 |
| I69.213 | DX10 | Psychomotor deficit following other ntrm intcrn hemorrhage | Cerebrovascular_Disease | I69213 |
| I69.214 | DX10 | Fntl lb and exec fcn def following other ntrm intcrn hemor | Cerebrovascular_Disease | I69214 |

| | | | | |
|---|---|---|---|---|
| I69.215 | DX10 | Cognitive social or emo def fol other ntrm intcrn hemor | Cerebrovascular_Disease | I69215 |
| I69.218 | DX10 | Oth symp and signs w cogn fnctns fol other ntrm intcrn hemor | Cerebrovascular_Disease | I69218 |
| I69.219 | DX10 | Unsp symp and signs w cogn fnctns fol oth ntrm intcrn hemor | Cerebrovascular_Disease | I69219 |
| I69.22 | DX10 | Speech/lang deficits following oth ntrm intcrn hemorrhage | Cerebrovascular_Disease | I6922 |
| I69.22 | DX10 | Speech/lang deficits following oth ntrm intcrn hemorrhage | Cerebrovascular_Disease | I6922 |
| I69.221 | DX10 | Dysphasia following oth nontraumatic intracranial hemorrhage | Cerebrovascular_Disease | I69221 |
| I69.222 | DX10 | Dysarthria following oth nontraumatic intcrn hemorrhage | Cerebrovascular_Disease | I69222 |
| I69.223 | DX10 | Fluency disorder following oth ntrm intcrn hemorrhage | Cerebrovascular_Disease | I69223 |
| I69.228 | DX10 | Oth speech/lang deficits following oth ntrm intcrn hemor | Cerebrovascular_Disease | I69228 |
| I69.23 | DX10 | Monoplg upr lmb following oth nontraumatic intcrn hemorrhage | Cerebrovascular_Disease | I6923 |
| I69.231 | DX10 | Monoplg upr lmb fol oth ntrm intcrn hemor aff right dom side | Cerebrovascular_Disease | I69231 |
| I69.232 | DX10 | Monoplg upr lmb fol oth ntrm intcrn hemor aff left dom side | Cerebrovascular_Disease | I69232 |
| I69.233 | DX10 | Monoplg upr lmb fol oth ntrm intcrn hemor aff r nondom side | Cerebrovascular_Disease | I69233 |
| I69.234 | DX10 | Monoplg upr lmb fol oth ntrm intcrn hemor aff l nondom side | Cerebrovascular_Disease | I69234 |
| I69.239 | DX10 | Monoplg upr lmb fol oth ntrm intcrn hemor aff unsp side | Cerebrovascular_Disease | I69239 |
| I69.24 | DX10 | Monoplg low lmb following oth nontraumatic intcrn hemorrhage | Cerebrovascular_Disease | I6924 |
| I69.241 | DX10 | Monoplg low lmb fol oth ntrm intcrn hemor aff right dom side | Cerebrovascular_Disease | I69241 |
| I69.242 | DX10 | Monoplg low lmb fol oth ntrm intcrn hemor aff left dom side | Cerebrovascular_Disease | I69242 |
| I69.243 | DX10 | Monoplg low lmb fol oth ntrm intcrn hemor aff r nondom side | Cerebrovascular_Disease | I69243 |
| I69.244 | DX10 | Monoplg low lmb fol oth ntrm intcrn hemor aff l nondom side | Cerebrovascular_Disease | I69244 |
| I69.249 | DX10 | Monoplg low lmb fol oth ntrm intcrn hemor aff unsp side | Cerebrovascular_Disease | I69249 |
| I69.25 | DX10 | Hemiplga following oth nontraumatic intracranial hemorrhage | Cerebrovascular_Disease | I6925 |
| I69.251 | DX10 | Hemiplga fol oth ntrm intcrn hemor aff right dominant side | Cerebrovascular_Disease | I69251 |
| I69.252 | DX10 | Hemiplga fol oth ntrm intcrn hemor aff left dominant side | Cerebrovascular_Disease | I69252 |
| I69.253 | DX10 | Hemiplga fol oth ntrm intcrn hemor aff right nondom side | Cerebrovascular_Disease | I69253 |
| I69.254 | DX10 | Hemiplga fol oth ntrm intcrn hemor aff left nondom side | Cerebrovascular_Disease | I69254 |
| I69.259 | DX10 | Hemiplga following oth ntrm intcrn hemor affecting unsp side | Cerebrovascular_Disease | I69259 |
| I69.26 | DX10 | Oth paralytic syndrome following oth ntrm intcrn hemorrhage | Cerebrovascular_Disease | I6926 |
| I69.261 | DX10 | Oth parlyt synd fol oth ntrm intcrn hemor aff right dom side | Cerebrovascular_Disease | I69261 |

| | | | | |
|---|---|---|---|---|
| I69.262 | DX10 | Oth parlyt synd fol oth ntrm intcrn hemor aff left dom side | Cerebrovascular_Disease | I69262 |
| I69.263 | DX10 | Oth parlyt synd fol oth ntrm intcrn hemor aff r nondom side | Cerebrovascular_Disease | I69263 |
| I69.264 | DX10 | Oth parlyt synd fol oth ntrm intcrn hemor aff l nondom side | Cerebrovascular_Disease | I69264 |
| I69.265 | DX10 | Oth paralytic syndrome following oth ntrm intcrn hemor, bi | Cerebrovascular_Disease | I69265 |
| I69.269 | DX10 | Oth parlyt syndrome fol oth ntrm intcrn hemor aff unsp side | Cerebrovascular_Disease | I69269 |
| I69.29 | DX10 | Other sequelae of other nontraumatic intracranial hemorrhage | Cerebrovascular_Disease | I6929 |
| I69.29 | DX10 | Other sequelae of other nontraumatic intracranial hemorrhage | Cerebrovascular_Disease | I6929 |
| I69.3 | DX10 | Sequelae of cerebral infarction | Cerebrovascular_Disease | I693 |
| I69.30 | DX10 | Sequelae of cerebral infarction, unspecified | Cerebrovascular_Disease | I6930 |
| I69.31 | DX10 | Cognitive deficits following cerebral infarction | Cerebrovascular_Disease | I6931 |
| I69.310 | DX10 | Cognitive deficits following cerebral infarction | Cerebrovascular_Disease | I69310 |
| I69.311 | DX10 | Memory deficit following cerebral infarction | Cerebrovascular_Disease | I69311 |
| I69.312 | DX10 | Vis def/sptl nglct following cerebral infarction | Cerebrovascular_Disease | I69312 |
| I69.313 | DX10 | Psychomotor deficit following cerebral infarction | Cerebrovascular_Disease | I69313 |
| I69.314 | DX10 | Frontal lobe and exec fcn def following cerebral infarction | Cerebrovascular_Disease | I69314 |
| I69.315 | DX10 | Cognitive social or emo def following cerebral infarction | Cerebrovascular_Disease | I69315 |
| I69.318 | DX10 | Other symptoms and signs w cogn fnctns fol cerebral infrc | Cerebrovascular_Disease | I69318 |
| I69.319 | DX10 | Unsp symptoms and signs w cogn fnctns fol cerebral infrc | Cerebrovascular_Disease | I69319 |
| I69.32 | DX10 | Speech and language deficits following cerebral infarction | Cerebrovascular_Disease | I6932 |
| I69.32 | DX10 | Speech and language deficits following cerebral infarction | Cerebrovascular_Disease | I6932 |
| I69.321 | DX10 | Dysphasia following cerebral infarction | Cerebrovascular_Disease | I69321 |
| I69.322 | DX10 | Dysarthria following cerebral infarction | Cerebrovascular_Disease | I69322 |
| I69.323 | DX10 | Fluency disorder following cerebral infarction | Cerebrovascular_Disease | I69323 |
| I69.328 | DX10 | Oth speech/lang deficits following cerebral infarction | Cerebrovascular_Disease | I69328 |
| I69.33 | DX10 | Monoplegia of upper limb following cerebral infarction | Cerebrovascular_Disease | I6933 |
| I69.331 | DX10 | Monoplg upr lmb fol cerebral infrc aff right dominant side | Cerebrovascular_Disease | I69331 |
| I69.332 | DX10 | Monoplg upr lmb fol cerebral infrc aff left dominant side | Cerebrovascular_Disease | I69332 |
| I69.333 | DX10 | Monoplg upr lmb fol cerebral infrc aff right nondom side | Cerebrovascular_Disease | I69333 |
| I69.334 | DX10 | Monoplg upr lmb fol cerebral infrc aff left nondom side | Cerebrovascular_Disease | I69334 |

| | | | | |
|---|---|---|---|---|
| I69.339 | DX10 | Monoplg upr lmb following cerebral infrc affecting unsp side | Cerebrovascular_Disease | I69339 |
| I69.34 | DX10 | Monoplegia of lower limb following cerebral infarction | Cerebrovascular_Disease | I6934 |
| I69.341 | DX10 | Monoplg low lmb fol cerebral infrc aff right dominant side | Cerebrovascular_Disease | I69341 |
| I69.342 | DX10 | Monoplg low lmb fol cerebral infrc aff left dominant side | Cerebrovascular_Disease | I69342 |
| I69.343 | DX10 | Monoplg low lmb fol cerebral infrc aff right nondom side | Cerebrovascular_Disease | I69343 |
| I69.344 | DX10 | Monoplg low lmb fol cerebral infrc aff left nondom side | Cerebrovascular_Disease | I69344 |
| I69.349 | DX10 | Monoplg low lmb following cerebral infrc affecting unsp side | Cerebrovascular_Disease | I69349 |
| I69.35 | DX10 | Hemiplegia and hemiparesis following cerebral infarction | Cerebrovascular_Disease | I6935 |
| I69.351 | DX10 | Hemiplga following cerebral infrc aff right dominant side | Cerebrovascular_Disease | I69351 |
| I69.352 | DX10 | Hemiplga following cerebral infrc aff left dominant side | Cerebrovascular_Disease | I69352 |
| I69.353 | DX10 | Hemiplga following cerebral infrc aff right nondom side | Cerebrovascular_Disease | I69353 |
| I69.354 | DX10 | Hemiplga following cerebral infrc affecting left nondom side | Cerebrovascular_Disease | I69354 |
| I69.359 | DX10 | Hemiplga following cerebral infarction affecting unsp side | Cerebrovascular_Disease | I69359 |
| I69.36 | DX10 | Other paralytic syndrome following cerebral infarction | Cerebrovascular_Disease | I6936 |
| I69.361 | DX10 | Oth parlyt syndrome fol cereb infrc aff right dominant side | Cerebrovascular_Disease | I69361 |
| I69.362 | DX10 | Oth parlyt syndrome fol cereb infrc aff left dominant side | Cerebrovascular_Disease | I69362 |
| I69.363 | DX10 | Oth parlyt syndrome fol cerebral infrc aff right nondom side | Cerebrovascular_Disease | I69363 |
| I69.364 | DX10 | Oth parlyt syndrome fol cerebral infrc aff left nondom side | Cerebrovascular_Disease | I69364 |
| I69.365 | DX10 | Oth paralytic syndrome following cerebral infrc, bilateral | Cerebrovascular_Disease | I69365 |
| I69.369 | DX10 | Oth paralytic syndrome fol cerebral infrc aff unsp side | Cerebrovascular_Disease | I69369 |
| I69.39 | DX10 | Other sequelae of cerebral infarction | Cerebrovascular_Disease | I6939 |
| I69.390 | DX10 | Other sequelae of cerebral infarction | Cerebrovascular_Disease | I69390 |
| I69.8 | DX10 | Sequelae of other cerebrovascular diseases | Cerebrovascular_Disease | I698 |
| I69.80 | DX10 | Sequelae of other cerebrovascular diseases, unspecified | Cerebrovascular_Disease | I6980 |
| I69.81 | DX10 | Cognitive deficits following other cerebrovascular disease | Cerebrovascular_Disease | I6981 |
| I69.810 | DX10 | Cognitive deficits following other cerebrovascular disease | Cerebrovascular_Disease | I69810 |
| I69.811 | DX10 | Memory deficit following other cerebrovascular disease | Cerebrovascular_Disease | I69811 |
| I69.812 | DX10 | Vis def/sptl nglct following other cerebrovascular disease | Cerebrovascular_Disease | I69812 |
| I69.813 | DX10 | Psychomotor deficit following other cerebrovascular disease | Cerebrovascular_Disease | I69813 |

| | | | | |
|---|---|---|---|---|
| I69.814 | DX10 | Fntl lb and exec fcn def following other cerebvasc disease | Cerebrovascular_Disease | I69814 |
| I69.815 | DX10 | Cognitive social or emo def fol other cerebvasc disease | Cerebrovascular_Disease | I69815 |
| I69.818 | DX10 | Other symp and signs w cogn fnctns fol other cerebvasc dis | Cerebrovascular_Disease | I69818 |
| I69.819 | DX10 | Unsp symp and signs w cogn fnctns fol other cerebvasc dis | Cerebrovascular_Disease | I69819 |
| I69.82 | DX10 | Speech/lang deficits following oth cerebrovascular disease | Cerebrovascular_Disease | I6982 |
| I69.820 | DX10 | Speech/lang deficits following oth cerebrovascular disease | Cerebrovascular_Disease | I69820 |
| I69.821 | DX10 | Dysphasia following other cerebrovascular disease | Cerebrovascular_Disease | I69821 |
| I69.822 | DX10 | Dysarthria following other cerebrovascular disease | Cerebrovascular_Disease | I69822 |
| I69.823 | DX10 | Fluency disorder following other cerebrovascular disease | Cerebrovascular_Disease | I69823 |
| I69.828 | DX10 | Oth speech/lang deficits following oth cerebvasc disease | Cerebrovascular_Disease | I69828 |
| I69.83 | DX10 | Monoplg upr lmb following oth cerebrovascular disease | Cerebrovascular_Disease | I6983 |
| I69.831 | DX10 | Monoplg upr lmb fol oth cerebvasc disease aff right dom side | Cerebrovascular_Disease | I69831 |
| I69.832 | DX10 | Monoplg upr lmb fol oth cerebvasc disease aff left dom side | Cerebrovascular_Disease | I69832 |
| I69.833 | DX10 | Monoplg upr lmb fol oth cerebvasc dis aff right nondom side | Cerebrovascular_Disease | I69833 |
| I69.834 | DX10 | Monoplg upr lmb fol oth cerebvasc dis aff left nondom side | Cerebrovascular_Disease | I69834 |
| I69.839 | DX10 | Monoplg upr lmb fol oth cerebvasc disease aff unsp side | Cerebrovascular_Disease | I69839 |
| I69.84 | DX10 | Monoplg low lmb following oth cerebrovascular disease | Cerebrovascular_Disease | I6984 |
| I69.841 | DX10 | Monoplg low lmb fol oth cerebvasc disease aff right dom side | Cerebrovascular_Disease | I69841 |
| I69.842 | DX10 | Monoplg low lmb fol oth cerebvasc disease aff left dom side | Cerebrovascular_Disease | I69842 |
| I69.843 | DX10 | Monoplg low lmb fol oth cerebvasc dis aff right nondom side | Cerebrovascular_Disease | I69843 |
| I69.844 | DX10 | Monoplg low lmb fol oth cerebvasc dis aff left nondom side | Cerebrovascular_Disease | I69844 |
| I69.849 | DX10 | Monoplg low lmb fol oth cerebvasc disease aff unsp side | Cerebrovascular_Disease | I69849 |
| I69.85 | DX10 | Hemiplga following oth cerebrovascular disease | Cerebrovascular_Disease | I6985 |
| I69.851 | DX10 | Hemiplga fol oth cerebvasc disease aff right dominant side | Cerebrovascular_Disease | I69851 |
| I69.852 | DX10 | Hemiplga fol oth cerebvasc disease aff left dominant side | Cerebrovascular_Disease | I69852 |
| I69.853 | DX10 | Hemiplga fol oth cerebvasc disease aff right nondom side | Cerebrovascular_Disease | I69853 |
| I69.854 | DX10 | Hemiplga fol oth cerebvasc disease aff left nondom side | Cerebrovascular_Disease | I69854 |
| I69.859 | DX10 | Hemiplga following oth cerebvasc disease affecting unsp side | Cerebrovascular_Disease | I69859 |
| I69.86 | DX10 | Oth paralytic syndrome following oth cerebrovascular disease | Cerebrovascular_Disease | I6986 |

| | | | | |
|---|---|---|---|---|
| I69.861 | DX10 | Oth parlyt synd fol oth cerebvasc disease aff right dom side | Cerebrovascular_Disease | I69861 |
| I69.862 | DX10 | Oth parlyt synd fol oth cerebvasc disease aff left dom side | Cerebrovascular_Disease | I69862 |
| I69.863 | DX10 | Oth parlyt synd fol oth cerebvasc dis aff right nondom side | Cerebrovascular_Disease | I69863 |
| I69.864 | DX10 | Oth parlyt synd fol oth cerebvasc dis aff left nondom side | Cerebrovascular_Disease | I69864 |
| I69.865 | DX10 | Oth paralytic syndrome following oth cerebvasc disease, bi | Cerebrovascular_Disease | I69865 |
| I69.869 | DX10 | Oth parlyt syndrome fol oth cerebvasc disease aff unsp side | Cerebrovascular_Disease | I69869 |
| I69.89 | DX10 | Other sequelae of other cerebrovascular disease | Cerebrovascular_Disease | I6989 |
| I69.890 | DX10 | Other sequelae of other cerebrovascular disease | Cerebrovascular_Disease | I69890 |
| I69.9 | DX10 | Sequelae of unspecified cerebrovascular diseases | Cerebrovascular_Disease | I699 |
| I69.90 | DX10 | Sequelae of unspecified cerebrovascular diseases, unspecified | Cerebrovascular_Disease | I6990 |
| I69.91 | DX10 | Cognitive deficits following unsp cerebrovascular disease | Cerebrovascular_Disease | I6991 |
| I69.91 | DX10 | Cognitive deficits following unsp cerebrovascular disease | Cerebrovascular_Disease | I6991 |
| I69.911 | DX10 | Memory deficit following unspecified cerebrovascular disease | Cerebrovascular_Disease | I69911 |
| I69.912 | DX10 | Vis def/sptl nglct following unspecified cerebvasc disease | Cerebrovascular_Disease | I69912 |
| I69.913 | DX10 | Psychomotor deficit following unspecified cerebvasc disease | Cerebrovascular_Disease | I69913 |
| I69.914 | DX10 | Fntl lb and exec fcn def following unsp cerebvasc disease | Cerebrovascular_Disease | I69914 |
| I69.915 | DX10 | Cognitive social or emo def following unsp cerebvasc disease | Cerebrovascular_Disease | I69915 |
| I69.918 | DX10 | Other symp and signs w cogn fnctns fol unsp cerebvasc dis | Cerebrovascular_Disease | I69918 |
| I69.919 | DX10 | Unsp symp and signs w cogn fnctns fol unsp cerebvasc disease | Cerebrovascular_Disease | I69919 |
| I69.92 | DX10 | Speech/lang deficits following unsp cerebrovascular disease | Cerebrovascular_Disease | I6992 |
| I69.920 | DX10 | Speech/lang deficits following unsp cerebrovascular disease | Cerebrovascular_Disease | I69920 |
| I69.921 | DX10 | Dysphasia following unspecified cerebrovascular disease | Cerebrovascular_Disease | I69921 |
| I69.922 | DX10 | Dysarthria following unspecified cerebrovascular disease | Cerebrovascular_Disease | I69922 |
| I69.923 | DX10 | Fluency disorder following unsp cerebrovascular disease | Cerebrovascular_Disease | I69923 |
| I69.928 | DX10 | Oth speech/lang deficits following unsp cerebvasc disease | Cerebrovascular_Disease | I69928 |
| I69.93 | DX10 | Monoplg upr lmb following unsp cerebrovascular disease | Cerebrovascular_Disease | I6993 |
| I69.931 | DX10 | Monoplg upr lmb fol unsp cerebvasc dis aff right dom side | Cerebrovascular_Disease | I69931 |
| I69.932 | DX10 | Monoplg upr lmb fol unsp cerebvasc disease aff left dom side | Cerebrovascular_Disease | I69932 |
| I69.933 | DX10 | Monoplg upr lmb fol unsp cerebvasc dis aff right nondom side | Cerebrovascular_Disease | I69933 |

| | | | | |
|---|---|---|---|---|
| I69.934 | DX10 | Monoplg upr lmb fol unsp cerebvasc dis aff left nondom side | Cerebrovascular_Disease | I69934 |
| I69.939 | DX10 | Monoplg upr lmb fol unsp cerebvasc disease aff unsp side | Cerebrovascular_Disease | I69939 |
| I69.94 | DX10 | Monoplg low lmb following unsp cerebrovascular disease | Cerebrovascular_Disease | I6994 |
| I69.941 | DX10 | Monoplg low lmb fol unsp cerebvasc dis aff right dom side | Cerebrovascular_Disease | I69941 |
| I69.942 | DX10 | Monoplg low lmb fol unsp cerebvasc disease aff left dom side | Cerebrovascular_Disease | I69942 |
| I69.943 | DX10 | Monoplg low lmb fol unsp cerebvasc dis aff right nondom side | Cerebrovascular_Disease | I69943 |
| I69.944 | DX10 | Monoplg low lmb fol unsp cerebvasc dis aff left nondom side | Cerebrovascular_Disease | I69944 |
| I69.949 | DX10 | Monoplg low lmb fol unsp cerebvasc disease aff unsp side | Cerebrovascular_Disease | I69949 |
| I69.95 | DX10 | Hemiplga following unsp cerebrovascular disease | Cerebrovascular_Disease | I6995 |
| I69.951 | DX10 | Hemiplga fol unsp cerebvasc disease aff right dominant side | Cerebrovascular_Disease | I69951 |
| I69.952 | DX10 | Hemiplga fol unsp cerebvasc disease aff left dominant side | Cerebrovascular_Disease | I69952 |
| I69.953 | DX10 | Hemiplga fol unsp cerebvasc disease aff right nondom side | Cerebrovascular_Disease | I69953 |
| I69.954 | DX10 | Hemiplga fol unsp cerebvasc disease aff left nondom side | Cerebrovascular_Disease | I69954 |
| I69.959 | DX10 | Hemiplga following unsp cerebvasc disease aff unsp side | Cerebrovascular_Disease | I69959 |
| I69.96 | DX10 | Oth paralytic syndrome following unsp cerebvasc disease | Cerebrovascular_Disease | I6996 |
| I69.961 | DX10 | Oth parlyt synd fol unsp cerebvasc dis aff right dom side | Cerebrovascular_Disease | I69961 |
| I69.962 | DX10 | Oth parlyt synd fol unsp cerebvasc disease aff left dom side | Cerebrovascular_Disease | I69962 |
| I69.963 | DX10 | Oth parlyt synd fol unsp cerebvasc dis aff right nondom side | Cerebrovascular_Disease | I69963 |
| I69.964 | DX10 | Oth parlyt synd fol unsp cerebvasc dis aff left nondom side | Cerebrovascular_Disease | I69964 |
| I69.965 | DX10 | Oth paralytic syndrome following unsp cerebvasc disease, bi | Cerebrovascular_Disease | I69965 |
| I69.969 | DX10 | Oth parlyt syndrome fol unsp cerebvasc disease aff unsp side | Cerebrovascular_Disease | I69969 |
| I69.99 | DX10 | Other sequelae of unspecified cerebrovascular disease | Cerebrovascular_Disease | I6999 |
| I69.990 | DX10 | Other sequelae of unspecified cerebrovascular disease | Cerebrovascular_Disease | I69990 |
| G45.0 | DX10 | Vertebro-basilar artery syndrome | Cerebrovascular_Disease | G450 |
| G45.1 | DX10 | Carotid artery syndrome (hemispheric) | Cerebrovascular_Disease | G451 |
| G45.2 | DX10 | Multiple and bilateral precerebral artery syndromes | Cerebrovascular_Disease | G452 |
| G45.8 | DX10 | Oth transient cerebral ischemic attacks and related synd | Cerebrovascular_Disease | G458 |
| G45.9 | DX10 | Transient cerebral ischemic attack, unspecified | Cerebrovascular_Disease | G459 |
| G46.0 | DX10 | Middle cerebral artery syndrome | Cerebrovascular_Disease | G460 |

| | | | | |
|---|---|---|---|---|
| G46.1 | DX10 | Anterior cerebral artery syndrome | Cerebrovascular_Disease | G461 |
| G46.2 | DX10 | Posterior cerebral artery syndrome | Cerebrovascular_Disease | G462 |
| G46.3 | DX10 | Brain stem stroke syndrome | Cerebrovascular_Disease | G463 |
| G46.4 | DX10 | Cerebellar stroke syndrome | Cerebrovascular_Disease | G464 |
| G46.5 | DX10 | Pure motor lacunar syndrome | Cerebrovascular_Disease | G465 |
| G46.6 | DX10 | Pure sensory lacunar syndrome | Cerebrovascular_Disease | G466 |
| G46.7 | DX10 | Other lacunar syndromes | Cerebrovascular_Disease | G467 |
| G46.8 | DX10 | Oth vascular syndromes of brain in cerebrovascular diseases | Cerebrovascular_Disease | G468 |
| G97.31 | DX10 | Intraop hemor/hemtom of a nervous sys org comp nrv sys proc | Cerebrovascular_Disease | G9731 |
| G97.32 | DX10 | Intraop hemor/hemtom of a nervous sys org comp oth procedure | Cerebrovascular_Disease | G9732 |
| I97.810 | DX10 | Intraoperative cerebvasc infarction during cardiac surgery | Cerebrovascular_Disease | I97810 |
| I97.811 | DX10 | Intraoperative cerebrovascular infarction during oth surgery | Cerebrovascular_Disease | I97811 |
| I97.820 | DX10 | Postproc cerebvasc infarction following cardiac surgery | Cerebrovascular_Disease | I97820 |
| I97.820 | DX10 | Postproc cerebvasc infarction following cardiac surgery | Cerebrovascular_Disease | I97820 |
| I97.821 | DX10 | Postprocedural cerebvasc infarction following other surgery | Cerebrovascular_Disease | I97821 |
| Z86.73 | DX10 | Personal history of TIA and cerebral infarction without residual deficits | Cerebrovascular_Disease | Z8673 |
| D59.3 | DX10 | Hemolytic-uremic syndrome | Chronic_Kidney_Disease | D593 |
| E08.2 | DX10 | Diabetes due to underlying condition w kidney complications | Chronic_Kidney_Disease | E082 |
| E08.21 | DX10 | Diabetes due to underlying condition w diabetic nephropathy | Chronic_Kidney_Disease | E0821 |
| E08.22 | DX10 | Diabetes due to undrl cond w diabetic chronic kidney disease | Chronic_Kidney_Disease | E0822 |
| E08.29 | DX10 | Diabetes due to undrl condition w oth diabetic kidney comp | Chronic_Kidney_Disease | E0829 |
| E09.2 | DX10 | Drug/chem diabetes mellitus w kidney complications | Chronic_Kidney_Disease | E092 |
| E09.21 | DX10 | Drug/chem diabetes mellitus w diabetic nephropathy | Chronic_Kidney_Disease | E0921 |
| E09.22 | DX10 | Drug/chem diabetes w diabetic chronic kidney disease | Chronic_Kidney_Disease | E0922 |
| E09.29 | DX10 | Drug/chem diabetes w oth diabetic kidney complication | Chronic_Kidney_Disease | E0929 |
| E10.2 | DX10 | Type 1 diabetes mellitus with kidney complications | Chronic_Kidney_Disease | E102 |
| E10.21 | DX10 | Type 1 diabetes mellitus with diabetic nephropathy | Chronic_Kidney_Disease | E1021 |
| E10.22 | DX10 | Type 1 diabetes mellitus w diabetic chronic kidney disease | Chronic_Kidney_Disease | E1022 |
| E10.29 | DX10 | Type 1 diabetes mellitus w oth diabetic kidney complication | Chronic_Kidney_Disease | E1029 |

| | | | | |
|---|---|---|---|---|
| E11.2 | DX10 | Type 2 diabetes mellitus with kidney complications | Chronic_Kidney_Disease | E112 |
| E11.21 | DX10 | Type 2 diabetes mellitus with diabetic nephropathy | Chronic_Kidney_Disease | E1121 |
| E11.22 | DX10 | Type 2 diabetes mellitus w diabetic chronic kidney disease | Chronic_Kidney_Disease | E1122 |
| E11.29 | DX10 | Type 2 diabetes mellitus w oth diabetic kidney complication | Chronic_Kidney_Disease | E1129 |
| E13.2 | DX10 | Other specified diabetes mellitus with kidney complications | Chronic_Kidney_Disease | E132 |
| E13.21 | DX10 | Other specified diabetes mellitus with diabetic nephropathy | Chronic_Kidney_Disease | E1321 |
| E13.22 | DX10 | Oth diabetes mellitus with diabetic chronic kidney disease | Chronic_Kidney_Disease | E1322 |
| E13.29 | DX10 | Oth diabetes mellitus with oth diabetic kidney complication | Chronic_Kidney_Disease | E1329 |
| I12 | DX10 | Hypertensive chronic kidney disease | Chronic_Kidney_Disease | I12 |
| I12.0 | DX10 | Hyp chr kidney disease w stage 5 chr kidney disease or ESRD | Chronic_Kidney_Disease | I120 |
| I12.9 | DX10 | Hypertensive chronic kidney disease w stg 1-4/unsp chr kdny | Chronic_Kidney_Disease | I129 |
| I13.0 | DX10 | Hyp hrt & chr kdny dis w hrt fail and stg 1-4/unsp chr kdny | Chronic_Kidney_Disease | I130 |
| I13.1 | DX10 | Hypertensive heart and chronic kidney disease w/o hrt fail | Chronic_Kidney_Disease | I131 |
| I13.10 | DX10 | Hyp hrt & chr kdny dis w/o hrt fail, w stg 1-4/unsp chr kdny | Chronic_Kidney_Disease | I1310 |
| I13.11 | DX10 | Hyp hrt and chr kdny dis w/o hrt fail, w stg 5 chr kdny/ESRD | Chronic_Kidney_Disease | I1311 |
| I13.2 | DX10 | Hyp hrt & chr kdny dis w hrt fail and w stg 5 chr kdny/ESRD | Chronic_Kidney_Disease | I132 |
| K76.7 | DX10 | Hepatorenal syndrome | Chronic_Kidney_Disease | K767 |
| M10.3 | DX10 | Gout due to renal impairment | Chronic_Kidney_Disease | M103 |
| M10.30 | DX10 | Gout due to renal impairment, unspecified site | Chronic_Kidney_Disease | M1030 |
| M10.31 | DX10 | Gout due to renal impairment, shoulder | Chronic_Kidney_Disease | M1031 |
| M10.311 | DX10 | Gout due to renal impairment, right shoulder | Chronic_Kidney_Disease | M10311 |
| M10.312 | DX10 | Gout due to renal impairment, left shoulder | Chronic_Kidney_Disease | M10312 |
| M10.319 | DX10 | Gout due to renal impairment, unspecified shoulder | Chronic_Kidney_Disease | M10319 |
| M10.32 | DX10 | Gout due to renal impairment, elbow | Chronic_Kidney_Disease | M1032 |
| M10.321 | DX10 | Gout due to renal impairment, right elbow | Chronic_Kidney_Disease | M10321 |
| M10.322 | DX10 | Gout due to renal impairment, left elbow | Chronic_Kidney_Disease | M10322 |
| M10.329 | DX10 | Gout due to renal impairment, unspecified elbow | Chronic_Kidney_Disease | M10329 |
| M10.33 | DX10 | Gout due to renal impairment, wrist | Chronic_Kidney_Disease | M1033 |
| M10.331 | DX10 | Gout due to renal impairment, right wrist | Chronic_Kidney_Disease | M10331 |

| | | | | |
|---|---|---|---|---|
| M10.332 | DX10 | Gout due to renal impairment, left wrist | Chronic_Kidney_Disease | M10332 |
| M10.339 | DX10 | Gout due to renal impairment, unspecified wrist | Chronic_Kidney_Disease | M10339 |
| M10.34 | DX10 | Gout due to renal impairment, hand | Chronic_Kidney_Disease | M1034 |
| M10.341 | DX10 | Gout due to renal impairment, right hand | Chronic_Kidney_Disease | M10341 |
| M10.342 | DX10 | Gout due to renal impairment, left hand | Chronic_Kidney_Disease | M10342 |
| M10.349 | DX10 | Gout due to renal impairment, unspecified hand | Chronic_Kidney_Disease | M10349 |
| M10.35 | DX10 | Gout due to renal impairment, hip | Chronic_Kidney_Disease | M1035 |
| M10.351 | DX10 | Gout due to renal impairment, right hip | Chronic_Kidney_Disease | M10351 |
| M10.352 | DX10 | Gout due to renal impairment, left hip | Chronic_Kidney_Disease | M10352 |
| M10.359 | DX10 | Gout due to renal impairment, unspecified hip | Chronic_Kidney_Disease | M10359 |
| M10.36 | DX10 | Gout due to renal impairment, knee | Chronic_Kidney_Disease | M1036 |
| M10.361 | DX10 | Gout due to renal impairment, right knee | Chronic_Kidney_Disease | M10361 |
| M10.362 | DX10 | Gout due to renal impairment, left knee | Chronic_Kidney_Disease | M10362 |
| M10.369 | DX10 | Gout due to renal impairment, unspecified knee | Chronic_Kidney_Disease | M10369 |
| M10.37 | DX10 | Gout due to renal impairment, ankle and foot | Chronic_Kidney_Disease | M1037 |
| M10.371 | DX10 | Gout due to renal impairment, right ankle and foot | Chronic_Kidney_Disease | M10371 |
| M10.372 | DX10 | Gout due to renal impairment, left ankle and foot | Chronic_Kidney_Disease | M10372 |
| M10.379 | DX10 | Gout due to renal impairment, unspecified ankle and foot | Chronic_Kidney_Disease | M10379 |
| M10.38 | DX10 | Gout due to renal impairment, vertebrae | Chronic_Kidney_Disease | M1038 |
| M10.39 | DX10 | Gout due to renal impairment, multiple sites | Chronic_Kidney_Disease | M1039 |
| M32.14 | DX10 | Glomerular disease in systemic lupus erythematosus | Chronic_Kidney_Disease | M3214 |
| M32.15 | DX10 | Tubulo-interstitial neuropath in sys lupus erythematosus | Chronic_Kidney_Disease | M3215 |
| N02 | DX10 | Recurrent and persistent hematuria | Chronic_Kidney_Disease | N02 |
| N02.0 | DX10 | Recurrent and persistent hematuria w minor glomerular abnlt | Chronic_Kidney_Disease | N020 |
| N02.1 | DX10 | Recur and perst hematur w focal and seg glomerular lesions | Chronic_Kidney_Disease | N021 |
| N02.2 | DX10 | Recurrent and perst hematur w diffuse membranous glomrlneph | Chronic_Kidney_Disease | N022 |
| N02.3 | DX10 | Recur and perst hematur w diffus mesangial prolif glomrlneph | Chronic_Kidney_Disease | N023 |
| N02.4 | DX10 | Recur & perst hematur w diffus endocaplry prolif glomrlneph | Chronic_Kidney_Disease | N024 |
| N02.5 | DX10 | Recurrent and perst hematur w diffuse mesangiocap glomrlneph | Chronic_Kidney_Disease | N025 |

| | | | | |
|---|---|---|---|---|
| N02.6 | DX10 | Recurrent and persistent hematuria w dense deposit disease | Chronic_Kidney_Disease | N026 |
| N02.7 | DX10 | Recurrent and perst hematur w diffuse crescentic glomrlneph | Chronic_Kidney_Disease | N027 |
| N02.8 | DX10 | Recurrent and persistent hematuria w oth morphologic changes | Chronic_Kidney_Disease | N028 |
| N02.9 | DX10 | Recurrent and perst hematuria w unsp morphologic changes | Chronic_Kidney_Disease | N029 |
| N02.A | DX10 | | Chronic_Kidney_Disease | N02A |
| N03 | DX10 | Chronic nephritic syndrome | Chronic_Kidney_Disease | N03 |
| N03.0 | DX10 | Chronic nephritic syndrome with minor glomerular abnormality | Chronic_Kidney_Disease | N030 |
| N03.1 | DX10 | Chronic neph syndrome w focal and seg glomerular lesions | Chronic_Kidney_Disease | N031 |
| N03.2 | DX10 | Chronic nephritic syndrome w diffuse membranous glomrlneph | Chronic_Kidney_Disease | N032 |
| N03.3 | DX10 | Chronic neph syndrome w diffuse mesangial prolif glomrlneph | Chronic_Kidney_Disease | N033 |
| N03.4 | DX10 | Chronic neph syndrome w diffuse endocaplry prolif glomrlneph | Chronic_Kidney_Disease | N034 |
| N03.5 | DX10 | Chronic nephritic syndrome w diffuse mesangiocap glomrlneph | Chronic_Kidney_Disease | N035 |
| N03.6 | DX10 | Chronic nephritic syndrome with dense deposit disease | Chronic_Kidney_Disease | N036 |
| N03.7 | DX10 | Chronic nephritic syndrome w diffuse crescentic glomrlneph | Chronic_Kidney_Disease | N037 |
| N03.8 | DX10 | Chronic nephritic syndrome with other morphologic changes | Chronic_Kidney_Disease | N038 |
| N03.9 | DX10 | Chronic nephritic syndrome with unsp morphologic changes | Chronic_Kidney_Disease | N039 |
| N03.A | DX10 | | Chronic_Kidney_Disease | N03A |
| N04 | DX10 | Nephrotic syndrome | Chronic_Kidney_Disease | N04 |
| N04.0 | DX10 | Nephrotic syndrome with minor glomerular abnormality | Chronic_Kidney_Disease | N040 |
| N04.1 | DX10 | Nephrotic syndrome w focal and segmental glomerular lesions | Chronic_Kidney_Disease | N041 |
| N04.2 | DX10 | Nephrotic syndrome w diffuse membranous glomerulonephritis | Chronic_Kidney_Disease | N042 |
| N04.3 | DX10 | Nephrotic syndrome w diffuse mesangial prolif glomrlneph | Chronic_Kidney_Disease | N043 |
| N04.4 | DX10 | Nephrotic syndrome w diffuse endocaplry prolif glomrlneph | Chronic_Kidney_Disease | N044 |
| N04.5 | DX10 | Nephrotic syndrome w diffuse mesangiocapillary glomrlneph | Chronic_Kidney_Disease | N045 |
| N04.6 | DX10 | Nephrotic syndrome with dense deposit disease | Chronic_Kidney_Disease | N046 |
| N04.7 | DX10 | Nephrotic syndrome w diffuse crescentic glomerulonephritis | Chronic_Kidney_Disease | N047 |
| N04.8 | DX10 | Nephrotic syndrome with other morphologic changes | Chronic_Kidney_Disease | N048 |
| N04.9 | DX10 | Nephrotic syndrome with unspecified morphologic changes | Chronic_Kidney_Disease | N049 |
| N04.A | DX10 | | Chronic_Kidney_Disease | N04A |

| | | | | |
|---|---|---|---|---|
| N05 | DX10 | Unspecified nephritic syndrome | Chronic_Kidney_Disease | N05 |
| N05.0 | DX10 | Unsp nephritic syndrome with minor glomerular abnormality | Chronic_Kidney_Disease | N050 |
| N05.1 | DX10 | Unsp neph syndrome w focal and segmental glomerular lesions | Chronic_Kidney_Disease | N051 |
| N05.2 | DX10 | Unsp nephritic syndrome w diffuse membranous glomrlneph | Chronic_Kidney_Disease | N052 |
| N05.3 | DX10 | Unsp neph syndrome w diffuse mesangial prolif glomrlneph | Chronic_Kidney_Disease | N053 |
| N05.4 | DX10 | Unsp neph syndrome w diffuse endocaplry prolif glomrlneph | Chronic_Kidney_Disease | N054 |
| N05.5 | DX10 | Unsp nephritic syndrome w diffuse mesangiocap glomrlneph | Chronic_Kidney_Disease | N055 |
| N05.6 | DX10 | Unspecified nephritic syndrome with dense deposit disease | Chronic_Kidney_Disease | N056 |
| N05.7 | DX10 | Unsp nephritic syndrome w diffuse crescentic glomrlneph | Chronic_Kidney_Disease | N057 |
| N05.8 | DX10 | Unsp nephritic syndrome with other morphologic changes | Chronic_Kidney_Disease | N058 |
| N05.9 | DX10 | Unsp nephritic syndrome with unspecified morphologic changes | Chronic_Kidney_Disease | N059 |
| N05.A | DX10 | | Chronic_Kidney_Disease | N05A |
| N06 | DX10 | Isolated proteinuria with specified morphological lesion | Chronic_Kidney_Disease | N06 |
| N06.0 | DX10 | Isolated proteinuria with minor glomerular abnormality | Chronic_Kidney_Disease | N060 |
| N06.1 | DX10 | Isolated protein w focal and segmental glomerular lesions | Chronic_Kidney_Disease | N061 |
| N06.2 | DX10 | Isolated proteinuria w diffuse membranous glomerulonephritis | Chronic_Kidney_Disease | N062 |
| N06.3 | DX10 | Isolated proteinuria w diffuse mesangial prolif glomrlneph | Chronic_Kidney_Disease | N063 |
| N06.4 | DX10 | Isolated proteinuria w diffuse endocaplry prolif glomrlneph | Chronic_Kidney_Disease | N064 |
| N06.5 | DX10 | Isolated proteinuria w diffuse mesangiocapillary glomrlneph | Chronic_Kidney_Disease | N065 |
| N06.6 | DX10 | Isolated proteinuria with dense deposit disease | Chronic_Kidney_Disease | N066 |
| N06.7 | DX10 | Isolated proteinuria w diffuse crescentic glomerulonephritis | Chronic_Kidney_Disease | N067 |
| N06.8 | DX10 | Isolated proteinuria with other morphologic lesion | Chronic_Kidney_Disease | N068 |
| N06.9 | DX10 | Isolated proteinuria with unspecified morphologic lesion | Chronic_Kidney_Disease | N069 |
| N06.A | DX10 | | Chronic_Kidney_Disease | N06A |
| N07 | DX10 | Hereditary nephropathy, not elsewhere classified | Chronic_Kidney_Disease | N07 |
| N07.0 | DX10 | Hereditary nephropathy, NEC w minor glomerular abnormality | Chronic_Kidney_Disease | N070 |
| N07.1 | DX10 | Heredit neuropath, NEC w focal and seg glomerular lesions | Chronic_Kidney_Disease | N071 |
| N07.2 | DX10 | Hereditary nephropathy, NEC w diffuse membranous glomrlneph | Chronic_Kidney_Disease | N072 |
| N07.3 | DX10 | Heredit neuropath, NEC w diffuse mesangial prolif glomrlneph | Chronic_Kidney_Disease | N073 |

| | | | | |
|---|---|---|---|---|
| N07.4 | DX10 | Heredit neuropath, NEC w diffus endocaplry prolif glomrlneph | Chronic_Kidney_Disease | N074 |
| N07.5 | DX10 | Hereditary nephropathy, NEC w diffuse mesangiocap glomrlneph | Chronic_Kidney_Disease | N075 |
| N07.6 | DX10 | Hereditary nephropathy, NEC w dense deposit disease | Chronic_Kidney_Disease | N076 |
| N07.7 | DX10 | Hereditary nephropathy, NEC w diffuse crescentic glomrlneph | Chronic_Kidney_Disease | N077 |
| N07.8 | DX10 | Hereditary nephropathy, NEC w oth morphologic lesions | Chronic_Kidney_Disease | N078 |
| N07.9 | DX10 | Hereditary nephropathy, NEC w unsp morphologic lesions | Chronic_Kidney_Disease | N079 |
| N07.A | DX10 | | Chronic_Kidney_Disease | N07A |
| N08 | DX10 | Glomerular disorders in diseases classified elsewhere | Chronic_Kidney_Disease | N08 |
| N11 | DX10 | Chronic tubulo-interstitial nephritis | Chronic_Kidney_Disease | N11 |
| N11.0 | DX10 | Nonobstructive reflux-associated chronic pyelonephritis | Chronic_Kidney_Disease | N110 |
| N11.1 | DX10 | Chronic obstructive pyelonephritis | Chronic_Kidney_Disease | N111 |
| N11.8 | DX10 | Other chronic tubulo-interstitial nephritis | Chronic_Kidney_Disease | N118 |
| N11.9 | DX10 | Chronic tubulo-interstitial nephritis, unspecified | Chronic_Kidney_Disease | N119 |
| N12 | DX10 | Tubulo-interstitial nephritis, not spcf as acute or chronic | Chronic_Kidney_Disease | N12 |
| N14 | DX10 | Drug- & heavy-metal-induced tubulo-interstitial & tublr cond | Chronic_Kidney_Disease | N14 |
| N14.0 | DX10 | Analgesic nephropathy | Chronic_Kidney_Disease | N140 |
| N14.1 | DX10 | Nephropathy induced by oth drug/meds/biol subst | Chronic_Kidney_Disease | N141 |
| N14.2 | DX10 | Neuropath induced by unsp drug, medicament or biolg sub | Chronic_Kidney_Disease | N142 |
| N14.3 | DX10 | Nephropathy induced by heavy metals | Chronic_Kidney_Disease | N143 |
| N14.4 | DX10 | Toxic nephropathy, not elsewhere classified | Chronic_Kidney_Disease | N144 |
| N15 | DX10 | Other renal tubulo-interstitial diseases | Chronic_Kidney_Disease | N15 |
| N15.0 | DX10 | Balkan nephropathy | Chronic_Kidney_Disease | N150 |
| N15.8 | DX10 | Other specified renal tubulo-interstitial diseases | Chronic_Kidney_Disease | N158 |
| N15.9 | DX10 | Renal tubulo-interstitial disease, unspecified | Chronic_Kidney_Disease | N159 |
| N16 | DX10 | Renal tubulo-interstitial disord in diseases classd elswhr | Chronic_Kidney_Disease | N16 |
| N18 | DX10 | Chronic kidney disease (CKD) | Chronic_Kidney_Disease | N18 |
| N18.1 | DX10 | Chronic kidney disease, stage 1 | Chronic_Kidney_Disease | N181 |
| N18.2 | DX10 | Chronic kidney disease, stage 2 (mild) | Chronic_Kidney_Disease | N182 |
| N18.3 | DX10 | Chronic kidney disease, stage 3 (moderate) | Chronic_Kidney_Disease | N183 |

| | | | | |
|---|---|---|---|---|
| N18.30 | DX10 | Chronic kidney disease, stage 3 unspecified | Chronic_Kidney_Disease | N1830 |
| N18.31 | DX10 | Chronic kidney disease, stage 3a | Chronic_Kidney_Disease | N1831 |
| N18.32 | DX10 | Chronic kidney disease, stage 3b | Chronic_Kidney_Disease | N1832 |
| N18.4 | DX10 | Chronic kidney disease, stage 4 (severe) | Chronic_Kidney_Disease | N184 |
| N18.5 | DX10 | Chronic kidney disease, stage 5 | Chronic_Kidney_Disease | N185 |
| N18.6 | DX10 | End stage renal disease | Chronic_Kidney_Disease | N186 |
| N18.9 | DX10 | Chronic kidney disease, unspecified | Chronic_Kidney_Disease | N189 |
| N19 | DX10 | Unspecified kidney failure | Chronic_Kidney_Disease | N19 |
| N25 | DX10 | Disorders resulting from impaired renal tubular function | Chronic_Kidney_Disease | N25 |
| N26.1 | DX10 | Atrophy of kidney (terminal) | Chronic_Kidney_Disease | N261 |
| N26.9 | DX10 | Renal sclerosis, unspecified | Chronic_Kidney_Disease | N269 |
| O26.83 | DX10 | Pregnancy related renal disease | Chronic_Kidney_Disease | O2683 |
| Q61.1 | DX10 | Polycystic kidney, infantile type | Chronic_Kidney_Disease | Q611 |
| Q61.11 | DX10 | Cystic dilatation of collecting ducts | Chronic_Kidney_Disease | Q6111 |
| Q61.19 | DX10 | Other polycystic kidney, infantile type | Chronic_Kidney_Disease | Q6119 |
| Q61.2 | DX10 | Polycystic kidney, adult type | Chronic_Kidney_Disease | Q612 |
| Q61.3 | DX10 | Polycystic kidney, unspecified | Chronic_Kidney_Disease | Q613 |
| Q61.4 | DX10 | Renal dysplasia | Chronic_Kidney_Disease | Q614 |
| I70.1 | DX10 | Atherosclerosis of renal artery | Chronic_Kidney_Disease | I701 |
| I72.2 | DX10 | Aneurysm of renal artery | Chronic_Kidney_Disease | I722 |
| M35.04 | DX10 | Sicca syndrome with tubulo-interstitial nephropathy | Chronic_Kidney_Disease | M3504 |
| N13.1 | DX10 | Hydronephrosis w ureteral stricture, NEC | Chronic_Kidney_Disease | N131 |
| N13.2 | DX10 | Hydronephrosis with renal and ureteral calculous obstruction | Chronic_Kidney_Disease | N132 |
| N13.30 | DX10 | Unspecified hydronephrosis | Chronic_Kidney_Disease | N1330 |
| N13.39 | DX10 | Other hydronephrosis | Chronic_Kidney_Disease | N1339 |
| N25.0 | DX10 | Renal osteodystrophy | Chronic_Kidney_Disease | N250 |
| N25.1 | DX10 | Nephrogenic diabetes insipidus | Chronic_Kidney_Disease | N251 |
| N25.81 | DX10 | Secondary hyperparathyroidism of renal origin | Chronic_Kidney_Disease | N2581 |
| N25.89 | DX10 | Oth disorders resulting from impaired renal tubular function | Chronic_Kidney_Disease | N2589 |

| | | | | |
|---|---|---|---|---|
| N25.9 | DX10 | Disorder rslt from impaired renal tubular function, unsp | Chronic_Kidney_Disease | N259 |
| Q62.0 | DX10 | Congenital hydronephrosis | Chronic_Kidney_Disease | Q620 |
| Q62.10 | DX10 | Congenital occlusion of ureter, unspecified | Chronic_Kidney_Disease | Q6210 |
| Q62.11 | DX10 | Congenital occlusion of ureteropelvic junction | Chronic_Kidney_Disease | Q6211 |
| Q62.12 | DX10 | Congenital occlusion of ureterovesical orifice | Chronic_Kidney_Disease | Q6212 |
| Q62.2 | DX10 | Congenital megaureter | Chronic_Kidney_Disease | Q622 |
| Q62.31 | DX10 | Congenital ureterocele, orthotopic | Chronic_Kidney_Disease | Q6231 |
| Q62.32 | DX10 | Cecoureterocele | Chronic_Kidney_Disease | Q6232 |
| Q62.39 | DX10 | Other obstructive defects of renal pelvis and ureter | Chronic_Kidney_Disease | Q6239 |
| R94.4 | DX10 | Abnormal results of kidney function studies | Chronic_Kidney_Disease | R944 |
| P96.0 | DX10 | Congenital renal failure | Chronic_Kidney_Disease | P960 |
| Z99.2 | DX10 | Dependence on renal dialysis | Chronic_Kidney_Disease | Z992 |
| 4150* | DX10 | PULMON_ROM | Chronic_Pulmonary_Disorder | 4150* |
| 491* | DX10 | PULMON_ROM | Chronic_Pulmonary_Disorder | 491* |
| 492* | DX10 | PULMON_ROM | Chronic_Pulmonary_Disorder | 492* |
| 493* | DX10 | PULMON_ROM | Chronic_Pulmonary_Disorder | 493* |
| 494* | DX10 | PULMON_ROM | Chronic_Pulmonary_Disorder | 494* |
| 496* | DX10 | PULMON_ROM | Chronic_Pulmonary_Disorder | 496* |
| I260 | DX10 | PULMON_ROM | Chronic_Pulmonary_Disorder | I260 |
| I2601 | DX10 | PULMON_ROM | Chronic_Pulmonary_Disorder | I2601 |
| I2602 | DX10 | PULMON_ROM | Chronic_Pulmonary_Disorder | I2602 |
| I2609 | DX10 | PULMON_ROM | Chronic_Pulmonary_Disorder | I2609 |
| I272 | DX10 | PULMON_ROM | Chronic_Pulmonary_Disorder | I272 |
| I278 | DX10 | PULMON_ROM | Chronic_Pulmonary_Disorder | I278 |
| I2781 | DX10 | PULMON_ROM | Chronic_Pulmonary_Disorder | I2781 |
| I2782 | DX10 | PULMON_ROM | Chronic_Pulmonary_Disorder | I2782 |
| I2789 | DX10 | PULMON_ROM | Chronic_Pulmonary_Disorder | I2789 |
| I279 | DX10 | PULMON_ROM | Chronic_Pulmonary_Disorder | I279 |
| J40 | DX10 | PULMON_ROM | Chronic_Pulmonary_Disorder | J40 |

| | | | | |
|---|---|---|---|---|
| J41 | DX10 | PULMON_ROM | Chronic_Pulmonary_Disorder | J41 |
| J410 | DX10 | PULMON_ROM | Chronic_Pulmonary_Disorder | J410 |
| J411 | DX10 | PULMON_ROM | Chronic_Pulmonary_Disorder | J411 |
| J418 | DX10 | PULMON_ROM | Chronic_Pulmonary_Disorder | J418 |
| J42 | DX10 | PULMON_ROM | Chronic_Pulmonary_Disorder | J42 |
| J43 | DX10 | PULMON_ROM | Chronic_Pulmonary_Disorder | J43 |
| J430 | DX10 | PULMON_ROM | Chronic_Pulmonary_Disorder | J430 |
| J431 | DX10 | PULMON_ROM | Chronic_Pulmonary_Disorder | J431 |
| J432 | DX10 | PULMON_ROM | Chronic_Pulmonary_Disorder | J432 |
| J438 | DX10 | PULMON_ROM | Chronic_Pulmonary_Disorder | J438 |
| J439 | DX10 | PULMON_ROM | Chronic_Pulmonary_Disorder | J439 |
| J44 | DX10 | PULMON_ROM | Chronic_Pulmonary_Disorder | J44 |
| J440 | DX10 | PULMON_ROM | Chronic_Pulmonary_Disorder | J440 |
| J441 | DX10 | PULMON_ROM | Chronic_Pulmonary_Disorder | J441 |
| J449 | DX10 | PULMON_ROM | Chronic_Pulmonary_Disorder | J449 |
| J45 | DX10 | PULMON_ROM | Chronic_Pulmonary_Disorder | J45 |
| J45.2 | DX10 | PULMON_ROM | Chronic_Pulmonary_Disorder | J452 |
| J45.20 | DX10 | PULMON_ROM | Chronic_Pulmonary_Disorder | J4520 |
| J45.21 | DX10 | PULMON_ROM | Chronic_Pulmonary_Disorder | J4521 |
| J45.22 | DX10 | PULMON_ROM | Chronic_Pulmonary_Disorder | J4522 |
| J45.3 | DX10 | PULMON_ROM | Chronic_Pulmonary_Disorder | J453 |
| J45.30 | DX10 | PULMON_ROM | Chronic_Pulmonary_Disorder | J4530 |
| J45.31 | DX10 | PULMON_ROM | Chronic_Pulmonary_Disorder | J4531 |
| J45.32 | DX10 | PULMON_ROM | Chronic_Pulmonary_Disorder | J4532 |
| J45.4 | DX10 | PULMON_ROM | Chronic_Pulmonary_Disorder | J454 |
| J45.40 | DX10 | PULMON_ROM | Chronic_Pulmonary_Disorder | J4540 |
| J45.41 | DX10 | PULMON_ROM | Chronic_Pulmonary_Disorder | J4541 |
| J45.42 | DX10 | PULMON_ROM | Chronic_Pulmonary_Disorder | J4542 |
| J45.5 | DX10 | PULMON_ROM | Chronic_Pulmonary_Disorder | J455 |

| | | | | |
|---|---|---|---|---|
| J45.50 | DX10 | PULMON_ROM | Chronic_Pulmonary_Disorder | J4550 |
| J45.51 | DX10 | PULMON_ROM | Chronic_Pulmonary_Disorder | J4551 |
| J45.52 | DX10 | PULMON_ROM | Chronic_Pulmonary_Disorder | J4552 |
| J45.9 | DX10 | PULMON_ROM | Chronic_Pulmonary_Disorder | J459 |
| J45.90 | DX10 | PULMON_ROM | Chronic_Pulmonary_Disorder | J4590 |
| J45.901 | DX10 | PULMON_ROM | Chronic_Pulmonary_Disorder | J45901 |
| J45.902 | DX10 | PULMON_ROM | Chronic_Pulmonary_Disorder | J45902 |
| J45.909 | DX10 | PULMON_ROM | Chronic_Pulmonary_Disorder | J45909 |
| J45.99 | DX10 | PULMON_ROM | Chronic_Pulmonary_Disorder | J4599 |
| J45.990 | DX10 | PULMON_ROM | Chronic_Pulmonary_Disorder | J45990 |
| J45.991 | DX10 | PULMON_ROM | Chronic_Pulmonary_Disorder | J45991 |
| J45.998 | DX10 | PULMON_ROM | Chronic_Pulmonary_Disorder | J45998 |
| J47 | DX10 | PULMON_ROM | Chronic_Pulmonary_Disorder | J47 |
| J470 | DX10 | PULMON_ROM | Chronic_Pulmonary_Disorder | J470 |
| J471 | DX10 | PULMON_ROM | Chronic_Pulmonary_Disorder | J471 |
| J479 | DX10 | PULMON_ROM | Chronic_Pulmonary_Disorder | J479 |
| J60 | DX10 | PULMON_ROM | Chronic_Pulmonary_Disorder | J60 |
| J61 | DX10 | PULMON_ROM | Chronic_Pulmonary_Disorder | J61 |
| J62 | DX10 | PULMON_ROM | Chronic_Pulmonary_Disorder | J62 |
| J620 | DX10 | PULMON_ROM | Chronic_Pulmonary_Disorder | J620 |
| J628 | DX10 | PULMON_ROM | Chronic_Pulmonary_Disorder | J628 |
| J63 | DX10 | PULMON_ROM | Chronic_Pulmonary_Disorder | J63 |
| J630 | DX10 | PULMON_ROM | Chronic_Pulmonary_Disorder | J630 |
| J631 | DX10 | PULMON_ROM | Chronic_Pulmonary_Disorder | J631 |
| J632 | DX10 | PULMON_ROM | Chronic_Pulmonary_Disorder | J632 |
| J633 | DX10 | PULMON_ROM | Chronic_Pulmonary_Disorder | J633 |
| J634 | DX10 | PULMON_ROM | Chronic_Pulmonary_Disorder | J634 |
| J635 | DX10 | PULMON_ROM | Chronic_Pulmonary_Disorder | J635 |
| J636 | DX10 | PULMON_ROM | Chronic_Pulmonary_Disorder | J636 |

| | | | | |
|---|---|---|---|---|
| J64 | DX10 | PULMON_ROM | Chronic_Pulmonary_Disorder | J64 |
| J65 | DX10 | PULMON_ROM | Chronic_Pulmonary_Disorder | J65 |
| J66 | DX10 | PULMON_ROM | Chronic_Pulmonary_Disorder | J66 |
| J660 | DX10 | PULMON_ROM | Chronic_Pulmonary_Disorder | J660 |
| J661 | DX10 | PULMON_ROM | Chronic_Pulmonary_Disorder | J661 |
| J662 | DX10 | PULMON_ROM | Chronic_Pulmonary_Disorder | J662 |
| J668 | DX10 | PULMON_ROM | Chronic_Pulmonary_Disorder | J668 |
| J67 | DX10 | PULMON_ROM | Chronic_Pulmonary_Disorder | J67 |
| J670 | DX10 | PULMON_ROM | Chronic_Pulmonary_Disorder | J670 |
| J671 | DX10 | PULMON_ROM | Chronic_Pulmonary_Disorder | J671 |
| J672 | DX10 | PULMON_ROM | Chronic_Pulmonary_Disorder | J672 |
| J673 | DX10 | PULMON_ROM | Chronic_Pulmonary_Disorder | J673 |
| J674 | DX10 | PULMON_ROM | Chronic_Pulmonary_Disorder | J674 |
| J675 | DX10 | PULMON_ROM | Chronic_Pulmonary_Disorder | J675 |
| J676 | DX10 | PULMON_ROM | Chronic_Pulmonary_Disorder | J676 |
| J677 | DX10 | PULMON_ROM | Chronic_Pulmonary_Disorder | J677 |
| J678 | DX10 | PULMON_ROM | Chronic_Pulmonary_Disorder | J678 |
| J679 | DX10 | PULMON_ROM | Chronic_Pulmonary_Disorder | J679 |
| J684 | DX10 | PULMON_ROM | Chronic_Pulmonary_Disorder | J684 |
| J701 | DX10 | PULMON_ROM | Chronic_Pulmonary_Disorder | J701 |
| J703 | DX10 | PULMON_ROM | Chronic_Pulmonary_Disorder | J703 |
| 571.2 | DX10 | Alcoholic cirrhosis of liver | Cirrhosis | 5712 |
| 571.5 | DX10 | Cirrhosis of liver without mention of alcohol | Cirrhosis | 5715 |
| 571.6 | DX10 | Biliary cirrhosis | Cirrhosis | 5716 |
| 777.8 | DX10 | Other specified perinatal disorder of digestive system | Cirrhosis | 7778 |
| K70.30 | DX10 | Alcoholic cirrhosis of liver without ascites | Cirrhosis | K7030 |
| K74.60 | DX10 | Unspecified cirrhosis of liver | Cirrhosis | K7460 |
| K74.69 | DX10 | Other cirrhosis of liver | Cirrhosis | K7469 |
| K74.3 | DX10 | Primary biliary cirrhosis | Cirrhosis | K743 |
| K74.4 | DX10 | Secondary biliary cirrhosis | Cirrhosis | K744 |
| K74.5 | DX10 | Biliary cirrhosis, unspecified | Cirrhosis | K745 |
| P78.81 | DX10 | Congenital cirrhosis (of liver) | Cirrhosis | P7881 |
| K70.3 | DX10 | Alcoholic cirrhosis of liver | Cirrhosis | K703 |
| K70.31 | DX10 | Alcoholic cirrhosis of liver with ascites | Cirrhosis | K7031 |
| K71.7 | DX10 | Toxic liver disease with fibrosis and cirrhosis of liver | Cirrhosis | K717 |
| K74 | DX10 | Fibrosis and cirrhosis of liver | Cirrhosis | K74 |
| K74.6 | DX10 | Other and unspecified cirrhosis of liver | Cirrhosis | K746 |

| | | | | |
|---|---|---|---|---|
| 398.91 | DX10 | RHEUMATIC HEART FAILURE | Congestive_Heart_Failure | 39891 |
| 402.01 | DX10 | MALIG HYPERTENSIVE HEART DISEASE W/HEART FAILURE | Congestive_Heart_Failure | 40201 |
| 402.11 | DX10 | BEN HYPERTENSIVE HEART DISEASE W/HEART FAILURE | Congestive_Heart_Failure | 40211 |
| 402.91 | DX10 | HYPERTENSIVE HEART DISEASE UNSPEC W/HEART FAIL | Congestive_Heart_Failure | 40291 |
| 404.01 | DX10 | HTN HRT & CKD MAL W/HF & W/CKD STAGE 1-IV/UNS | Congestive_Heart_Failure | 40401 |
| 404.03 | DX10 | HTN HRT & CKD MALIG W/HF & W/CKD STAGE V/ESRD | Congestive_Heart_Failure | 40403 |
| 404.11 | DX10 | HTN HRT & CKD BEN W/HF & W/CKD STAGE I-IV/UNS | Congestive_Heart_Failure | 40411 |
| 404.13 | DX10 | HTN HEART & CKD BEN W/HF & CKD STAGE V/ESRD | Congestive_Heart_Failure | 40413 |
| 404.91 | DX10 | HTN HRT & CKD UNS W/HF & W/CKD STAGE I - IV/UNS | Congestive_Heart_Failure | 40491 |
| 404.93 | DX10 | HTN HEART & CKD UNSPEC W/HF & CKD STAGE V/ESRD | Congestive_Heart_Failure | 40493 |
| 428 | DX10 | HEART FAILURE | Congestive_Heart_Failure | 428 |
| 428.0 | DX10 | CONGESTIVE HEART FAILURE UNSPECIFIED | Congestive_Heart_Failure | 4280 |
| 428.1 | DX10 | LEFT HEART FAILURE | Congestive_Heart_Failure | 4281 |
| 428.2 | DX10 | SYSTOLIC HEART FAILURE | Congestive_Heart_Failure | 4282 |
| 428.20 | DX10 | UNSPECIFIED SYSTOLIC HEART FAILURE | Congestive_Heart_Failure | 42820 |
| 428.21 | DX10 | ACUTE SYSTOLIC HEART FAILURE | Congestive_Heart_Failure | 42821 |
| 428.22 | DX10 | CHRONIC SYSTOLIC HEART FAILURE | Congestive_Heart_Failure | 42822 |
| 428.23 | DX10 | ACUTE ON CHRONIC SYSTOLIC HEART FAILURE | Congestive_Heart_Failure | 42823 |
| 428.3 | DX10 | DIASTOLIC HEART FAILURE | Congestive_Heart_Failure | 4283 |
| 428.30 | DX10 | UNSPECIFIED DIASTOLIC HEART FAILURE | Congestive_Heart_Failure | 42830 |
| 428.31 | DX10 | ACUTE DIASTOLIC HEART FAILURE | Congestive_Heart_Failure | 42831 |
| 428.32 | DX10 | CHRONIC DIASTOLIC HEART FAILURE | Congestive_Heart_Failure | 42832 |
| 428.33 | DX10 | ACUTE ON CHRONIC DIASTOLIC HEART FAILURE | Congestive_Heart_Failure | 42833 |
| 428.4 | DX10 | COMBINED SYSTOLIC AND DIASTOLIC HEART FAILURE | Congestive_Heart_Failure | 4284 |
| 428.40 | DX10 | UNSPEC COMBINED SYSTOLIC&DIASTOLIC HEART FAILURE | Congestive_Heart_Failure | 42840 |
| 428.41 | DX10 | ACUTE COMBINED SYSTOLIC&DIASTOLIC HEART FAILURE | Congestive_Heart_Failure | 42841 |
| 428.42 | DX10 | CHRONIC COMB SYSTOLIC&DIASTOLIC HEART FAILURE | Congestive_Heart_Failure | 42842 |
| 428.43 | DX10 | ACUTE CHRONIC COMB SYSTOLIC&DIASTOLIC HEART FAIL | Congestive_Heart_Failure | 42843 |
| 428.9 | DX10 | UNSPECIFIED HEART FAILURE | Congestive_Heart_Failure | 4289 |

| | | | | |
|---|---|---|---|---|
| I09.81 | DX10 | RHEUMATIC HEART FAILURE | Congestive_Heart_Failure | I0981 |
| I11.0 | DX10 | HYPERTENSIVE HEART DISEASE WITH HEART FAILURE | Congestive_Heart_Failure | I110 |
| I13.0 | DX10 | HTN HEART & CKD W/HF & CKD STAGE 1-4 OR UNS CKD | Congestive_Heart_Failure | I130 |
| I13.2 | DX10 | HTN HEART & CKD W/HF W/STAGE 5 CKD OR ESRD | Congestive_Heart_Failure | I132 |
| I50 | DX10 | HEART FAILURE | Congestive_Heart_Failure | I50 |
| I50.1 | DX10 | LEFT VENTRICULAR FAILURE UNSPECIFIED | Congestive_Heart_Failure | I501 |
| I50.2 | DX10 | SYSTOLIC CONGESTIVE HEART FAILURE | Congestive_Heart_Failure | I502 |
| I50.20 | DX10 | UNSPECIFIED SYSTOLIC CONGESTIVE HEART FAILURE | Congestive_Heart_Failure | I5020 |
| I50.21 | DX10 | ACUTE SYSTOLIC CONGESTIVE HEART FAILURE | Congestive_Heart_Failure | I5021 |
| I50.22 | DX10 | CHRONIC SYSTOLIC CONGESTIVE HEART FAILURE | Congestive_Heart_Failure | I5022 |
| I50.23 | DX10 | ACUTE CHRON SYSTOLIC HEART FAILURE | Congestive_Heart_Failure | I5023 |
| I50.3 | DX10 | DIASTOLIC CONGESTIVE HEART FAILURE | Congestive_Heart_Failure | I503 |
| I50.30 | DX10 | UNSPECIFIED DIASTOLIC CONGESTIVE HEART FAILURE | Congestive_Heart_Failure | I5030 |
| I50.31 | DX10 | ACUTE DIASTOLIC CONGESTIVE HEART FAILURE | Congestive_Heart_Failure | I5031 |
| I50.32 | DX10 | CHRONIC DIASTOLIC CONGESTIVE HEART FAILURE | Congestive_Heart_Failure | I5032 |
| I50.33 | DX10 | ACUTE ON CHRON DIASTOLIC CONGESTIV HEART FAILURE | Congestive_Heart_Failure | I5033 |
| I50.4 | DX10 | COMBINED SYSTOLIC & DIASTOLIC HEART FAILURE | Congestive_Heart_Failure | I504 |
| I50.40 | DX10 | UNSPECIFIED COMBINED SYSTOLIC & DIASTOLIC CHF | Congestive_Heart_Failure | I5040 |
| I50.41 | DX10 | ACUTE COMBINED SYSTOLIC AND DIASTOLIC CHF | Congestive_Heart_Failure | I5041 |
| I50.42 | DX10 | CHRONIC COMBINED SYSTOLIC AND DIASTOLIC CHF | Congestive_Heart_Failure | I5042 |
| I50.43 | DX10 | ACUTE ON CHRONIC COMB SYSTOLIC & DIASTOLIC CHF | Congestive_Heart_Failure | I5043 |
| I50.8 | DX10 | OTHER HEART FAILURE | Congestive_Heart_Failure | I508 |
| I50.81 | DX10 | RIGHT HEART FAILURE | Congestive_Heart_Failure | I5081 |
| I50.810 | DX10 | RIGHT HEART FAILURE UNSPECIFIED | Congestive_Heart_Failure | I50810 |
| I50.811 | DX10 | ACUTE RIGHT HEART FAILURE | Congestive_Heart_Failure | I50811 |
| I50.812 | DX10 | CHRONIC RIGHT HEART FAILURE | Congestive_Heart_Failure | I50812 |
| I50.813 | DX10 | ACUTE ON CHRONIC RIGHT HEART FAILURE | Congestive_Heart_Failure | I50813 |
| I50.814 | DX10 | RIGHT HEART FAILURE DUE TO LEFT HEART FAILURE | Congestive_Heart_Failure | I50814 |
| I50.82 | DX10 | BIVENTRICULAR HEART FAILURE | Congestive_Heart_Failure | I5082 |

| | | | | |
|---|---|---|---|---|
| I50.83 | DX10 | HIGH OUTPUT HEART FAILURE | Congestive_Heart_Failure | I5083 |
| I50.84 | DX10 | END STAGE HEART FAILURE | Congestive_Heart_Failure | I5084 |
| I50.89 | DX10 | OTHER HEART FAILURE | Congestive_Heart_Failure | I5089 |
| I50.9 | DX10 | HEART FAILURE UNSPECIFIED | Congestive_Heart_Failure | I509 |
| I97.11 | DX10 | POSTPROCEDURAL CARDIAC INSUFFICIENCY | Congestive_Heart_Failure | I9711 |
| I97.110 | DX10 | POSTPROC CARD INSUFFICIENCY FOLLOW CARD SURGERY | Congestive_Heart_Failure | I97110 |
| I97.111 | DX10 | POSTPROC CARD INSUFFICIENCY FOLLOW OTH SURGERY | Congestive_Heart_Failure | I97111 |
| I97.13 | DX10 | POSTPROCEDURAL HEART FAILURE | Congestive_Heart_Failure | I9713 |
| I97.130 | DX10 | POSTPROC HEART FAILURE FOLLOWING CARDIAC SURGERY | Congestive_Heart_Failure | I97130 |
| I97.131 | DX10 | POSTPROC HEART FAILURE FOLLOWING OTH SURGERY | Congestive_Heart_Failure | I97131 |
| 425.5 | DX10 | Alcoholic cardiomyopathy | Congestive_Heart_Failure | 4255 |
| 402.1 | DX10 | Benign hypertensive heart disease | Congestive_Heart_Failure | 4021 |
| 425.8 | DX10 | Cardiomyopathy in other diseases classified elsewhere | Congestive_Heart_Failure | 4258 |
| 425.7 | DX10 | Nutritional and metabolic cardiomyopathy | Congestive_Heart_Failure | 4257 |
| 425.4 | DX10 | Other primary cardiomyopathies | Congestive_Heart_Failure | 4254 |
| 425.9 | DX10 | Unspecified secondary cardiomyopathy | Congestive_Heart_Failure | 4259 |
| I11 | DX10 | Hypertensive heart disease | Congestive_Heart_Failure | I11 |
| I25.5 | DX10 | Ischemic cardiomyopathy | Congestive_Heart_Failure | I255 |
| I42.0 | DX10 | Dilated cardiomyopathy | Congestive_Heart_Failure | I420 |
| I42.5 | DX10 | Other restrictive cardiomyopathy | Congestive_Heart_Failure | I425 |
| I42.6 | DX10 | Alcoholic cardiomyopathy | Congestive_Heart_Failure | I426 |
| I42.7 | DX10 | Cardiomyopathy due to drug and external agent | Congestive_Heart_Failure | I427 |
| I42.8 | DX10 | Other cardiomyopathies | Congestive_Heart_Failure | I428 |
| I42.9 | DX10 | Cardiomyopathy, unspecified | Congestive_Heart_Failure | I429 |
| I43 | DX10 | Cardiomyopathy in diseases classified elsewhere | Congestive_Heart_Failure | I43 |
| 414.0 | DX10 | Coronary atherosclerosis | Coronary_Artery_Disease | 4140 |
| 414.00 | DX10 | Coronary atherosclerosis of unspecified type of vessel, native or graft | Coronary_Artery_Disease | 41400 |
| 414.01 | DX10 | Coronary atherosclerosis of native coronary artery | Coronary_Artery_Disease | 41401 |
| 414.02 | DX10 | Coronary atherosclerosis of autologous vein bypass graft | Coronary_Artery_Disease | 41402 |

| | | | | |
|---|---|---|---|---|
| 414.03 | DX10 | Coronary atherosclerosis of nonautologous biological bypass graft | Coronary_Artery_Disease | 41403 |
| 414.04 | DX10 | Coronary atherosclerosis of artery bypass graft | Coronary_Artery_Disease | 41404 |
| 414.05 | DX10 | Coronary atherosclerosis of unspecified bypass graft | Coronary_Artery_Disease | 41405 |
| 414.06 | DX10 | Coronary atherosclerosis of native coronary artery of transplanted heart | Coronary_Artery_Disease | 41406 |
| 414.07 | DX10 | Coronary atherosclerosis of bypass graft (artery) (vein) of transplanted heart | Coronary_Artery_Disease | 41407 |
| 414.11 | DX10 | Aneurysm of coronary vessels | Coronary_Artery_Disease | 41411 |
| 414.12 | DX10 | Dissection of coronary artery | Coronary_Artery_Disease | 41412 |
| 414.2 | DX10 | Chronic total occlusion of coronary artery | Coronary_Artery_Disease | 4142 |
| 414.3 | DX10 | Coronary atherosclerosis due to lipid rich plaque | Coronary_Artery_Disease | 4143 |
| 414.8 | DX10 | Other specified forms of chronic ischemic heart disease | Coronary_Artery_Disease | 4148 |
| 414.9 | DX10 | Chronic ischemic heart disease, unspecified | Coronary_Artery_Disease | 4149 |
| 411.81 | DX10 | Acute coronary occlusion without myocardial infarction | Coronary_Artery_Disease | 41181 |
| 411.89 | DX10 | Other acute and subacute forms of ischemic heart disease, other | Coronary_Artery_Disease | 41189 |
| 746.85 | DX10 | Coronary artery anomaly | Coronary_Artery_Disease | 74685 |
| 996 | DX10 | Mechanical complication of unspecified cardiac device, implant, and graft | Coronary_Artery_Disease | 996 |
| 996.03 | DX10 | Mechanical complication due to coronary bypass graft | Coronary_Artery_Disease | 99603 |
| V45.81 | DX10 | Aortocoronary bypass status | Coronary_Artery_Disease | V4581 |
| V45.82 | DX10 | Percutaneous transluminal coronary angioplasty status | Coronary_Artery_Disease | V4582 |
| I24.0 | DX10 | Acute coronary thrombosis not resulting in myocardial infarction | Coronary_Artery_Disease | I240 |
| I24.8 | DX10 | Other forms of acute ischemic heart disease | Coronary_Artery_Disease | I248 |
| I24.9 | DX10 | Acute ischemic heart disease, unspecified | Coronary_Artery_Disease | I249 |
| I25.10 | DX10 | Atherosclerotic heart disease of native coronary artery without angina pectoris | Coronary_Artery_Disease | I2510 |
| I25.110 | DX10 | Atherosclerotic heart disease of native coronary artery with unstable angina pectoris | Coronary_Artery_Disease | I25110 |
| I25.111 | DX10 | Atherosclerotic heart disease of native coronary artery with angina pectoris with documented spasm | Coronary_Artery_Disease | I25111 |
| I25.118 | DX10 | Atherosclerotic heart disease of native coronary artery with other forms of angina pectoris | Coronary_Artery_Disease | I25118 |
| I25.119 | DX10 | Atherosclerotic heart disease of native coronary artery with unspecified angina pectoris | Coronary_Artery_Disease | I25119 |
| I25.41 | DX10 | Coronary artery aneurysm | Coronary_Artery_Disease | I2541 |
| I25.42 | DX10 | Coronary artery dissection | Coronary_Artery_Disease | I2542 |
| I25.5 | DX10 | Ischemic cardiomyopathy | Coronary_Artery_Disease | I255 |

| | | | | |
|---|---|---|---|---|
| I25.6 | DX10 | Silent myocardial ischemia | Coronary_Artery_Disease | I256 |
| I25.700 | DX10 | Atherosclerosis of coronary artery bypass graft(s), unspecified, with unstable angina pectoris | Coronary_Artery_Disease | I25700 |
| I25.701 | DX10 | Atherosclerosis of coronary artery bypass graft(s), unspecified, with angina pectoris with documented spasm | Coronary_Artery_Disease | I25701 |
| I25.708 | DX10 | Atherosclerosis of coronary artery bypass graft(s), unspecified, with other forms of angina pectoris | Coronary_Artery_Disease | I25708 |
| I25.709 | DX10 | Atherosclerosis of coronary artery bypass graft(s), unspecified, with unspecified angina pectoris | Coronary_Artery_Disease | I25709 |
| I25.710 | DX10 | Atherosclerosis of autologous vein coronary artery bypass graft(s) with unstable angina pectoris | Coronary_Artery_Disease | I25710 |
| I25.711 | DX10 | Atherosclerosis of autologous vein coronary artery bypass graft(s) with angina pectoris with documented spasm | Coronary_Artery_Disease | I25711 |
| I25.718 | DX10 | Atherosclerosis of autologous vein coronary artery bypass graft(s) with other forms of angina pectoris | Coronary_Artery_Disease | I25718 |
| I25.719 | DX10 | Atherosclerosis of autologous vein coronary artery bypass graft(s) with unspecified angina pectoris | Coronary_Artery_Disease | I25719 |
| I25.720 | DX10 | Atherosclerosis of autologous artery coronary artery bypass graft(s) with unstable angina pectoris | Coronary_Artery_Disease | I25720 |
| I25.721 | DX10 | Atherosclerosis of autologous artery coronary artery bypass graft(s) with angina pectoris with documented spasm | Coronary_Artery_Disease | I25721 |
| I25.728 | DX10 | Atherosclerosis of autologous artery coronary artery bypass graft(s) with other forms of angina pectoris | Coronary_Artery_Disease | I25728 |
| I25.729 | DX10 | Atherosclerosis of autologous artery coronary artery bypass graft(s) with unspecified angina pectoris | Coronary_Artery_Disease | I25729 |
| I25.730 | DX10 | Atherosclerosis of nonautologous biological coronary artery bypass graft(s) with unstable angina pectoris | Coronary_Artery_Disease | I25730 |
| I25.731 | DX10 | Atherosclerosis of nonautologous biological coronary artery bypass graft(s) with angina pectoris with documented spasm | Coronary_Artery_Disease | I25731 |
| I25.738 | DX10 | Atherosclerosis of nonautologous biological coronary artery bypass graft(s) with other forms of angina pectoris | Coronary_Artery_Disease | I25738 |
| I25.739 | DX10 | Atherosclerosis of nonautologous biological coronary artery bypass graft(s) with unspecified angina pectoris | Coronary_Artery_Disease | I25739 |
| I25.750 | DX10 | Atherosclerosis of native coronary artery of transplanted heart with unstable angina | Coronary_Artery_Disease | I25750 |
| I25.751 | DX10 | Atherosclerosis of native coronary artery of transplanted heart with angina pectoris with documented spasm | Coronary_Artery_Disease | I25751 |
| I25.758 | DX10 | Atherosclerosis of native coronary artery of transplanted heart with other forms of angina pectoris | Coronary_Artery_Disease | I25758 |
| I25.759 | DX10 | Atherosclerosis of native coronary artery of transplanted heart with unspecified angina pectoris | Coronary_Artery_Disease | I25759 |
| I25.760 | DX10 | Atherosclerosis of bypass graft of coronary artery of transplanted heart with unstable angina | Coronary_Artery_Disease | I25760 |
| I25.761 | DX10 | Atherosclerosis of bypass graft of coronary artery of transplanted heart with angina pectoris with documented spasm | Coronary_Artery_Disease | I25761 |
| I25.768 | DX10 | Atherosclerosis of bypass graft of coronary artery of transplanted heart with other forms of angina pectoris | Coronary_Artery_Disease | I25768 |
| I25.769 | DX10 | Atherosclerosis of bypass graft of coronary artery of transplanted heart with unspecified angina pectoris | Coronary_Artery_Disease | I25769 |
| I25.790 | DX10 | Atherosclerosis of other coronary artery bypass graft(s) with unstable angina pectoris | Coronary_Artery_Disease | I25790 |
| I25.791 | DX10 | Atherosclerosis of other coronary artery bypass graft(s) with angina pectoris with documented spasm | Coronary_Artery_Disease | I25791 |

| | | | | |
|---|---|---|---|---|
| I25.798 | DX10 | Atherosclerosis of other coronary artery bypass graft(s) with other forms of angina pectoris | Coronary_Artery_Disease | I25798 |
| I25.799 | DX10 | Atherosclerosis of other coronary artery bypass graft(s) with unspecified angina pectoris | Coronary_Artery_Disease | I25799 |
| I25.810 | DX10 | Atherosclerosis of coronary artery bypass graft(s) without angina pectoris | Coronary_Artery_Disease | I25810 |
| I25.811 | DX10 | Atherosclerosis of native coronary artery of transplanted heart without angina pectoris | Coronary_Artery_Disease | I25811 |
| I25.812 | DX10 | Atherosclerosis of bypass graft of coronary artery of transplanted heart without angina pectoris | Coronary_Artery_Disease | I25812 |
| I25.82 | DX10 | Chronic total occlusion of coronary artery | Coronary_Artery_Disease | I2582 |
| I25.83 | DX10 | Coronary atherosclerosis due to lipid rich plaque | Coronary_Artery_Disease | I2583 |
| I25.84 | DX10 | Coronary atherosclerosis due to calcified coronary lesion | Coronary_Artery_Disease | I2584 |
| I25.89 | DX10 | Other forms of chronic ischemic heart disease | Coronary_Artery_Disease | I2589 |
| I25.9 | DX10 | Chronic ischemic heart disease, unspecified | Coronary_Artery_Disease | I259 |
| Q24.5 | DX10 | Malformation of coronary vessels | Coronary_Artery_Disease | Q245 |
| T82.211A | DX10 | Breakdown (mechanical) of coronary artery bypass graft, initial encounter | Coronary_Artery_Disease | T82211A |
| T82.211D | DX10 | Breakdown (mechanical) of coronary artery bypass graft, subsequent encounter | Coronary_Artery_Disease | T82211D |
| T82.211S | DX10 | Breakdown (mechanical) of coronary artery bypass graft, sequela | Coronary_Artery_Disease | T82211S |
| T82.212A | DX10 | Displacement of coronary artery bypass graft, initial encounter | Coronary_Artery_Disease | T82212A |
| T82.212D | DX10 | Displacement of coronary artery bypass graft, subsequent encounter | Coronary_Artery_Disease | T82212D |
| T82.212S | DX10 | Displacement of coronary artery bypass graft, sequela | Coronary_Artery_Disease | T82212S |
| T82.213A | DX10 | Leakage of coronary artery bypass graft, initial encounter | Coronary_Artery_Disease | T82213A |
| T82.213D | DX10 | Leakage of coronary artery bypass graft, subsequent encounter | Coronary_Artery_Disease | T82213D |
| T82.213S | DX10 | Leakage of coronary artery bypass graft, sequela | Coronary_Artery_Disease | T82213S |
| T82.218A | DX10 | Other mechanical complication of coronary artery bypass graft, initial encounter | Coronary_Artery_Disease | T82218A |
| T82.218D | DX10 | Other mechanical complication of coronary artery bypass graft, subsequent encounter | Coronary_Artery_Disease | T82218D |
| T82.218S | DX10 | Other mechanical complication of coronary artery bypass graft, sequela | Coronary_Artery_Disease | T82218S |
| Z95.1 | DX10 | Presence of aortocoronary bypass graft | Coronary_Artery_Disease | Z951 |
| Z95.5 | DX10 | Presence of coronary angioplasty implant and graft | Coronary_Artery_Disease | Z955 |
| Z98.61 | DX10 | Coronary angioplasty status | Coronary_Artery_Disease | Z9861 |
| 491.20 | DX10 | Obstructive chronic bronchitis, without exacerbation | CPOD | 49120 |
| 491.21 | DX10 | Obstructive chronic bronchitis, with (acute) exacerbation | CPOD | 49121 |
| 491.22 | DX10 | Obstructive chronic bronchitis with acute bronchitis | CPOD | 49122 |
| 491.8 | DX10 | Other chronic bronchitis | CPOD | 4918 |
| 491.9 | DX10 | Unspecified chronic bronchitis | CPOD | 4919 |
| 492.8 | DX10 | Other emphysema | CPOD | 4928 |
| 493.2 | DX10 | Chronic obstructive asthma | CPOD | 4932 |

| | | | | |
|---|---|---|---|---|
| 493.20 | DX10 | Chronic obstructive asthma, unspecified | CPOD | 49320 |
| 493.21 | DX10 | Chronic obstructive asthma with status asthmaticus | CPOD | 49321 |
| 493.22 | DX10 | Chronic obstructive asthma, with (acute) exacerbation | CPOD | 49322 |
| 496 | DX10 | Chronic airway obstruction, not elsewhere classified | CPOD | 496 |
| J44 | DX10 | Other chronic obstructive pulmonary disease | CPOD | J44 |
| J44.0 | DX10 | Chronic obstructive pulmonary disease with (acute) lower respiratory infection | CPOD | J440 |
| J44.1 | DX10 | Chronic obstructive pulmonary disease with (acute) exacerbation | CPOD | J441 |
| J44.9 | DX10 | Chronic obstructive pulmonary disease, unspecified | CPOD | J449 |
| E84 | DX10 | Cystic fibrosis | Cystic_Fibrosis | E84 |
| E84.0 | DX10 | Cystic fibrosis with pulmonary manifestations | Cystic_Fibrosis | E840 |
| E84.1 | DX10 | Cystic fibrosis with intestinal manifestations | Cystic_Fibrosis | E841 |
| E84.11 | DX10 | Meconium ileus in cystic fibrosis | Cystic_Fibrosis | E8411 |
| E84.19 | DX10 | Cystic fibrosis with other intestinal manifestations | Cystic_Fibrosis | E8419 |
| E84.8 | DX10 | Cystic fibrosis with other manifestations | Cystic_Fibrosis | E848 |
| E84.9 | DX10 | Cystic fibrosis, unspecified | Cystic_Fibrosis | E849 |
| F01 | DX10 | Vascular dementia | Dementia | F01 |
| F01.5 | DX10 | Vascular dementia | Dementia | F015 |
| F01.50 | DX10 | Vascular dementia without behavioral disturbance | Dementia | F0150 |
| F01.51 | DX10 | Vascular dementia with behavioral disturbance | Dementia | F0151 |
| F02 | DX10 | Dementia in other diseases classified elsewhere | Dementia | F02 |
| F02.8 | DX10 | Dementia in other diseases classified elsewhere | Dementia | F028 |
| F02.80 | DX10 | Dementia in oth diseases classd elswhr w/o behavrl disturb | Dementia | F0280 |
| F02.81 | DX10 | Dementia in oth diseases classd elswhr w behavioral disturb | Dementia | F0281 |
| F03 | DX10 | Unspecified dementia | Dementia | F03 |
| F03.9 | DX10 | Unspecified dementia | Dementia | F039 |
| F03.90 | DX10 | Unspecified dementia without behavioral disturbance | Dementia | F0390 |
| F03.91 | DX10 | Unspecified dementia with behavioral disturbance | Dementia | F0391 |
| G30 | DX10 | Alzheimer's disease | Dementia | G30 |
| G30.0 | DX10 | Alzheimer's disease with early onset | Dementia | G300 |
| G30.1 | DX10 | Alzheimer's disease with late onset | Dementia | G301 |
| G30.8 | DX10 | Other Alzheimer's disease | Dementia | G308 |
| G30.9 | DX10 | Alzheimer's disease, unspecified | Dementia | G309 |
| G31.0 | DX10 | Frontotemporal dementia | Dementia | G310 |
| G31.01 | DX10 | Pick's disease | Dementia | G3101 |
| G31.09 | DX10 | Other frontotemporal dementia | Dementia | G3109 |
| G31.83 | DX10 | Dementia with Lewy bodies | Dementia | G3183 |
| E10.10 | DX10 | Type 1 diabetes mellitus with ketoacidosis without coma | Diabetes_Type1 | E1010 |
| E10.11 | DX10 | Type 1 diabetes mellitus with ketoacidosis with coma | Diabetes_Type1 | E1011 |
| E10.21 | DX10 | Type 1 diabetes mellitus with diabetic nephropathy | Diabetes_Type1 | E1021 |
| E10.22 | DX10 | Type 1 diabetes mellitus with diabetic chronic kidney disease | Diabetes_Type1 | E1022 |
| E10.29 | DX10 | Type 1 diabetes mellitus with other diabetic kidney complication | Diabetes_Type1 | E1029 |
| E10.311 | DX10 | Type 1 diabetes mellitus with unspecified diabetic retinopathy with macular edema | Diabetes_Type1 | E10311 |
| E10.319 | DX10 | Type 1 diabetes mellitus with unspecified diabetic retinopathy without macular edema | Diabetes_Type1 | E10319 |
| E10.3211 | DX10 | Type 1 diabetes mellitus with mild nonproliferative diabetic retinopathy with macular edema | Diabetes_Type1 | E103211 |
| E10.3212 | DX10 | Type 1 diabetes mellitus with mild nonproliferative diabetic retinopathy with macular edema | Diabetes_Type1 | E103212 |
| E10.3213 | DX10 | Type 1 diabetes mellitus with mild nonproliferative diabetic retinopathy with macular edema | Diabetes_Type1 | E103213 |

| | | | | |
|---|---|---|---|---|
| E10.3219 | DX10 | Type 1 diabetes mellitus with mild nonproliferative diabetic retinopathy with macular edema | Diabetes_Type1 | E103219 |
| E10.3291 | DX10 | Type 1 diabetes mellitus with mild nonproliferative diabetic retinopathy without macular edema | Diabetes_Type1 | E103291 |
| E10.3292 | DX10 | Type 1 diabetes mellitus with mild nonproliferative diabetic retinopathy without macular edema | Diabetes_Type1 | E103292 |
| E10.3293 | DX10 | Type 1 diabetes mellitus with mild nonproliferative diabetic retinopathy without macular edema | Diabetes_Type1 | E103293 |
| E10.3299 | DX10 | Type 1 diabetes mellitus with mild nonproliferative diabetic retinopathy without macular edema | Diabetes_Type1 | E103299 |
| E10.3311 | DX10 | Type 1 diabetes mellitus with moderate nonproliferative diabetic retinopathy with macular edema | Diabetes_Type1 | E103311 |
| E10.3312 | DX10 | Type 1 diabetes mellitus with moderate nonproliferative diabetic retinopathy with macular edema | Diabetes_Type1 | E103312 |
| E10.3313 | DX10 | Type 1 diabetes mellitus with moderate nonproliferative diabetic retinopathy with macular edema | Diabetes_Type1 | E103313 |
| E10.3319 | DX10 | Type 1 diabetes mellitus with moderate nonproliferative diabetic retinopathy with macular edema | Diabetes_Type1 | E103319 |
| E10.3391 | DX10 | Type 1 diabetes mellitus with moderate nonproliferative diabetic retinopathy without macular edema | Diabetes_Type1 | E103391 |
| E10.3392 | DX10 | Type 1 diabetes mellitus with moderate nonproliferative diabetic retinopathy without macular edema | Diabetes_Type1 | E103392 |
| E10.3393 | DX10 | Type 1 diabetes mellitus with moderate nonproliferative diabetic retinopathy without macular edema | Diabetes_Type1 | E103393 |
| E10.3399 | DX10 | Type 1 diabetes mellitus with moderate nonproliferative diabetic retinopathy without macular edema | Diabetes_Type1 | E103399 |
| E10.3411 | DX10 | Type 1 diabetes mellitus with severe nonproliferative diabetic retinopathy with macular edema | Diabetes_Type1 | E103411 |
| E10.3412 | DX10 | Type 1 diabetes mellitus with severe nonproliferative diabetic retinopathy with macular edema | Diabetes_Type1 | E103412 |
| E10.3413 | DX10 | Type 1 diabetes mellitus with severe nonproliferative diabetic retinopathy with macular edema | Diabetes_Type1 | E103413 |
| E10.3419 | DX10 | Type 1 diabetes mellitus with severe nonproliferative diabetic retinopathy with macular edema | Diabetes_Type1 | E103419 |
| E10.3491 | DX10 | Type 1 diabetes mellitus with severe nonproliferative diabetic retinopathy without macular edema | Diabetes_Type1 | E103491 |
| E10.3492 | DX10 | Type 1 diabetes mellitus with severe nonproliferative diabetic retinopathy without macular edema | Diabetes_Type1 | E103492 |
| E10.3493 | DX10 | Type 1 diabetes mellitus with severe nonproliferative diabetic retinopathy without macular edema | Diabetes_Type1 | E103493 |
| E10.3499 | DX10 | Type 1 diabetes mellitus with severe nonproliferative diabetic retinopathy without macular edema | Diabetes_Type1 | E103499 |
| E10.3511 | DX10 | Type 1 diabetes mellitus with proliferative diabetic retinopathy with macular edema | Diabetes_Type1 | E103511 |
| E10.3512 | DX10 | Type 1 diabetes mellitus with proliferative diabetic retinopathy with macular edema | Diabetes_Type1 | E103512 |
| E10.3513 | DX10 | Type 1 diabetes mellitus with proliferative diabetic retinopathy with macular edema | Diabetes_Type1 | E103513 |
| E10.3519 | DX10 | Type 1 diabetes mellitus with proliferative diabetic retinopathy with macular edema | Diabetes_Type1 | E103519 |
| E10.3521 | DX10 | Type 1 diabetes mellitus with proliferative diabetic retinopathy with traction retinal detachment involving the macula | Diabetes_Type1 | E103521 |
| E10.3522 | DX10 | Type 1 diabetes mellitus with proliferative diabetic retinopathy with traction retinal detachment involving the macula | Diabetes_Type1 | E103522 |
| E10.3523 | DX10 | Type 1 diabetes mellitus with proliferative diabetic retinopathy with traction retinal detachment involving the macula | Diabetes_Type1 | E103523 |

| | | | | |
|---|---|---|---|---|
| E10.3529 | DX10 | Type 1 diabetes mellitus with proliferative diabetic retinopathy with traction retinal detachment involving the macula | Diabetes_Type1 | E103529 |
| E10.3531 | DX10 | Type 1 diabetes mellitus with proliferative diabetic retinopathy with traction retinal detachment not involving the macula | Diabetes_Type1 | E103531 |
| E10.3532 | DX10 | Type 1 diabetes mellitus with proliferative diabetic retinopathy with traction retinal detachment not involving the macula | Diabetes_Type1 | E103532 |
| E10.3533 | DX10 | Type 1 diabetes mellitus with proliferative diabetic retinopathy with traction retinal detachment not involving the macula | Diabetes_Type1 | E103533 |
| E10.3539 | DX10 | Type 1 diabetes mellitus with proliferative diabetic retinopathy with traction retinal detachment not involving the macula | Diabetes_Type1 | E103539 |
| E10.3541 | DX10 | Type 1 diabetes mellitus with proliferative diabetic retinopathy with combined traction retinal detachment and rhegmatogenous retinal detachment | Diabetes_Type1 | E103541 |
| E10.3542 | DX10 | Type 1 diabetes mellitus with proliferative diabetic retinopathy with combined traction retinal detachment and rhegmatogenous retinal detachment | Diabetes_Type1 | E103542 |
| E10.3543 | DX10 | Type 1 diabetes mellitus with proliferative diabetic retinopathy with combined traction retinal detachment and rhegmatogenous retinal detachment | Diabetes_Type1 | E103543 |
| E10.3549 | DX10 | Type 1 diabetes mellitus with proliferative diabetic retinopathy with combined traction retinal detachment and rhegmatogenous retinal detachment | Diabetes_Type1 | E103549 |
| E10.3551 | DX10 | Type 1 diabetes mellitus with stable proliferative diabetic retinopathy | Diabetes_Type1 | E103551 |
| E10.3552 | DX10 | Type 1 diabetes mellitus with stable proliferative diabetic retinopathy | Diabetes_Type1 | E103552 |
| E10.3553 | DX10 | Type 1 diabetes mellitus with stable proliferative diabetic retinopathy | Diabetes_Type1 | E103553 |
| E10.3559 | DX10 | Type 1 diabetes mellitus with stable proliferative diabetic retinopathy | Diabetes_Type1 | E103559 |
| E10.3591 | DX10 | Type 1 diabetes mellitus with proliferative diabetic retinopathy without macular edema | Diabetes_Type1 | E103591 |
| E10.3592 | DX10 | Type 1 diabetes mellitus with proliferative diabetic retinopathy without macular edema | Diabetes_Type1 | E103592 |
| E10.3593 | DX10 | Type 1 diabetes mellitus with proliferative diabetic retinopathy without macular edema | Diabetes_Type1 | E103593 |
| E10.3599 | DX10 | Type 1 diabetes mellitus with proliferative diabetic retinopathy without macular edema | Diabetes_Type1 | E103599 |
| E10.36 | DX10 | Type 1 diabetes mellitus with diabetic cataract | Diabetes_Type1 | E1036 |
| E10.37X1 | DX10 | Type 1 diabetes mellitus with diabetic macular edema, resolved following treatment | Diabetes_Type1 | E1037X1 |
| E10.37X2 | DX10 | Type 1 diabetes mellitus with diabetic macular edema, resolved following treatment | Diabetes_Type1 | E1037X2 |
| E10.37X3 | DX10 | Type 1 diabetes mellitus with diabetic macular edema, resolved following treatment | Diabetes_Type1 | E1037X3 |
| E10.37X9 | DX10 | Type 1 diabetes mellitus with diabetic macular edema, resolved following treatment | Diabetes_Type1 | E1037X9 |
| E10.39 | DX10 | Type 1 diabetes mellitus with other diabetic ophthalmic complication | Diabetes_Type1 | E1039 |
| E10.40 | DX10 | Type 1 diabetes mellitus with diabetic neuropathy, unspecified | Diabetes_Type1 | E1040 |
| E10.41 | DX10 | Type 1 diabetes mellitus with diabetic mononeuropathy | Diabetes_Type1 | E1041 |
| E10.42 | DX10 | Type 1 diabetes mellitus with diabetic polyneuropathy | Diabetes_Type1 | E1042 |

| | | | | |
|---|---|---|---|---|
| E10.43 | DX10 | Type 1 diabetes mellitus with diabetic autonomic (poly)neuropathy | Diabetes_Type1 | E1043 |
| E10.44 | DX10 | Type 1 diabetes mellitus with diabetic amyotrophy | Diabetes_Type1 | E1044 |
| E10.49 | DX10 | Type 1 diabetes mellitus with other diabetic neurological complication | Diabetes_Type1 | E1049 |
| E10.51 | DX10 | Type 1 diabetes mellitus with diabetic peripheral angiopathy without gangrene | Diabetes_Type1 | E1051 |
| E10.52 | DX10 | Type 1 diabetes mellitus with diabetic peripheral angiopathy with gangrene | Diabetes_Type1 | E1052 |
| E10.59 | DX10 | Type 1 diabetes mellitus with other circulatory complications | Diabetes_Type1 | E1059 |
| E10.610 | DX10 | Type 1 diabetes mellitus with diabetic neuropathic arthropathy | Diabetes_Type1 | E10610 |
| E10.618 | DX10 | Type 1 diabetes mellitus with other diabetic arthropathy | Diabetes_Type1 | E10618 |
| E10.620 | DX10 | Type 1 diabetes mellitus with diabetic dermatitis | Diabetes_Type1 | E10620 |
| E10.621 | DX10 | Type 1 diabetes mellitus with foot ulcer | Diabetes_Type1 | E10621 |
| E10.622 | DX10 | Type 1 diabetes mellitus with other skin ulcer | Diabetes_Type1 | E10622 |
| E10.628 | DX10 | Type 1 diabetes mellitus with other skin complications | Diabetes_Type1 | E10628 |
| E10.630 | DX10 | Type 1 diabetes mellitus with periodontal disease | Diabetes_Type1 | E10630 |
| E10.638 | DX10 | Type 1 diabetes mellitus with other oral complications | Diabetes_Type1 | E10638 |
| E10.641 | DX10 | Type 1 diabetes mellitus with hypoglycemia with coma | Diabetes_Type1 | E10641 |
| E10.649 | DX10 | Type 1 diabetes mellitus with hypoglycemia without coma | Diabetes_Type1 | E10649 |
| E10.65 | DX10 | Type 1 diabetes mellitus with hyperglycemia | Diabetes_Type1 | E1065 |
| E10.69 | DX10 | Type 1 diabetes mellitus with other specified complication | Diabetes_Type1 | E1069 |
| E10.8 | DX10 | Type 1 diabetes mellitus with unspecified complications | Diabetes_Type1 | E108 |
| E10.9 | DX10 | Type 1 diabetes mellitus without complications | Diabetes_Type1 | E109 |
| 250.00 | DX10 | DIAB W/O COMP TYPE II/UNS NOT STATED UNCNTRL | Diabetes_Type2 | 25000 |
| 250.02 | DX10 | DIAB W/O MENTION COMP TYPE II/UNS TYPE UNCNTRL | Diabetes_Type2 | 2502 |
| 250.10 | DX10 | DIAB W/KETOACIDOS TYPE II/UNS NOT STATED UNCNTRL | Diabetes_Type2 | 25010 |
| 250.12 | DX10 | DIABETES W/KETOACIDOSIS TYPE II/UNS TYPE UNCNTRL | Diabetes_Type2 | 25012 |
| 250.20 | DX10 | DIAB W/HYPEROSMOLARITY TYPE II/UNS NOT UNCNTRL | Diabetes_Type2 | 25020 |
| 250.22 | DX10 | DIAB W/HYPEROSMOLARITY TYPE II/UNS TYPE UNCNTRL | Diabetes_Type2 | 25022 |
| 250.30 | DX10 | DIAB W/OTH COMA TYPE II/UNS NOT STATED UNCNTRL | Diabetes_Type2 | 25030 |
| 250.32 | DX10 | DIABETES W/OTH COMA TYPE II/UNS UNCONTROLLED | Diabetes_Type2 | 25032 |
| 250.40 | DX10 | DIAB W/RENAL MANIFESTS TYPE II/UNS NOT UNCNTRL | Diabetes_Type2 | 25040 |
| 250.42 | DX10 | DIAB W/RENAL MANIFESTS TYPE II/UNS TYPE UNCNTRL | Diabetes_Type2 | 25042 |
| 250.50 | DX10 | DIAB W/OPHTH MANIFESTS TYPE II/UNS NOT UNCNTRL | Diabetes_Type2 | 25050 |
| 250.52 | DX10 | DIAB W/OPHTH MANIFESTS TYPE II/UNS TYPE UNCNTRL | Diabetes_Type2 | 25052 |
| 250.60 | DX10 | DIAB W/NEURO MANIFESTS TYPE II/UNS NOT UNCNTRL | Diabetes_Type2 | 25060 |
| 250.62 | DX10 | DIAB W/NEURO MANIFESTS TYPE II/UNS TYPE UNCNTRL | Diabetes_Type2 | 25062 |
| 250.70 | DX10 | DIAB W/PERIPH CIRC D/O TYPE II/UNS NOT UNCNTRL | Diabetes_Type2 | 25070 |
| 250.72 | DX10 | DIAB W/PERIPH CIRC D/O TYPE II/UNS TYPE UNCNTRL | Diabetes_Type2 | 25072 |
| 250.80 | DX10 | DIAB W/OTH MANIFESTS TYPE II/UNS NOT UNCNTRL | Diabetes_Type2 | 25080 |
| 250.82 | DX10 | DIAB W/OTH MANIFESTS TYPE II/UNS TYPE UNCNTRL | Diabetes_Type2 | 25082 |
| 250.90 | DX10 | DIAB W/UNS COMP TYPE II/UNS NOT STATED UNCNTRL | Diabetes_Type2 | 25090 |
| 250.92 | DX10 | DIAB W/UNSPEC COMP TYPE II/UNSPEC TYPE UNCNTRL | Diabetes_Type2 | 25092 |
| E11 | DX10 | TYPE 2 DIABETES MELLITUS | Diabetes_Type2 | E11 |

| | | | | |
|---|---|---|---|---|
| E11.0 | DX10 | TYPE 2 DIABETES MELLITUS WITH HYPEROSMOLARITY | Diabetes_Type2 | E110 |
| E11.00 | DX10 | TYPE 2 DM W/HYPEROSMOLARITY W/O NKHHC | Diabetes_Type2 | E1100 |
| E11.01 | DX10 | TYPE 2 DIAB MELLITUS W/HYPEROSMOLARITY W/COMA | Diabetes_Type2 | E111 |
| E11.1 | DX10 | TYPE 2 DIABETES MELLITUS WITH KETOACIDOSIS | Diabetes_Type2 | E111 |
| E11.10 | DX10 | TYPE 2 DIABETES MELLITUS W/KETOACIDOSIS W/O COMA | Diabetes_Type2 | E1110 |
| E11.11 | DX10 | TYPE 2 DIABETES MELLITUS WITH KETOACIDOSIS COMA | Diabetes_Type2 | E1111 |
| E11.2 | DX10 | TYPE 2 DIABETES MELLITUS W/KIDNEY COMPLICATIONS | Diabetes_Type2 | E112 |
| E11.21 | DX10 | TYPE 2 DIABETES MELLITUS W/DIABETIC NEPHROPATHY | Diabetes_Type2 | E1121 |
| E11.22 | DX10 | TYPE 2 DIABETES MELLITUS W/DIAB CHRON KIDNEY DZ | Diabetes_Type2 | E1122 |
| E11.29 | DX10 | TYPE 2 DIABETES MELLITUS W/OTH DIAB KIDNEY COMP | Diabetes_Type2 | E1129 |
| E11.3 | DX10 | TYPE 2 DIABETES MELLITUS W/OPHTHALMIC COMP | Diabetes_Type2 | E113 |
| E11.31 | DX10 | TYPE 2 DIABETES MELLITUS W/UNS DIAB RETINOPATHY | Diabetes_Type2 | E1131 |
| E11.311 | DX10 | TYPE 2 DM W/UNS DIAB RETINPATHY W/MACULAR EDEMA | Diabetes_Type2 | E11311 |
| E11.319 | DX10 | TYPE 2 DM W/UNS DIAB RETINPATH W/O MACULAR EDEMA | Diabetes_Type2 | E11319 |
| E11.32 | DX10 | TYPE 2 DM W/MILD NONPROLIF DIABETIC RETINPATHY | Diabetes_Type2 | E1132 |
| E11.321 | DX10 | TYPE 2 DM W/MILD NONPROLIF DIAB RETINOPATHY W/ME | Diabetes_Type2 | E11321 |
| E11.3211 | DX10 | TYPE 2 DIABETES MELLITUS MILD NPD MACULAR ED OD | Diabetes_Type2 | E113211 |
| E11.3212 | DX10 | TYPE 2 DIABETES MELLITUS MILD NPD MACULAR ED OS | Diabetes_Type2 | E113212 |
| E11.3213 | DX10 | TYPE 2 DIABETES MELLITUS MILD NPD MACULAR ED BIL | Diabetes_Type2 | E113213 |
| E11.3219 | DX10 | TYPE 2 DIAB MELLITUS MILD NPD MACULAR ED UNS EYE | Diabetes_Type2 | E113219 |
| E11.329 | DX10 | TYPE 2 DM W/MILD NONPROLIF DIAB RETINPATH W/O ME | Diabetes_Type2 | E11329 |
| E11.3291 | DX10 | TYPE 2 DIABETES MELLITUS MILD NPDR W/O MAC ED OD | Diabetes_Type2 | E113291 |
| E11.3292 | DX10 | TYPE 2 DIABETES MELLITUS MILD NPDR W/O MAC ED OS | Diabetes_Type2 | E113292 |
| E11.3293 | DX10 | TYPE 2 DIABETES MELLITUS MILD NPDR W/O MAC ED BL | Diabetes_Type2 | E113293 |
| E11.3299 | DX10 | TYPE 2 DIABETES MELLITUS MLD NPDR W/O ME UNS EYE | Diabetes_Type2 | E113299 |
| E11.33 | DX10 | TYPE 2 DM W/MOD NONPROLIF DIABETIC RETINOPATHY | Diabetes_Type2 | E1133 |
| E11.331 | DX10 | TYPE 2 DM W/MOD NONPROLIF DIAB RETINOPATHY W/ME | Diabetes_Type2 | E11331 |
| E11.3311 | DX10 | TYPE 2 DIABETES MELLITUS MOD NPDR MACULAR ED OD | Diabetes_Type2 | E113311 |
| E11.3312 | DX10 | TYPE 2 DIABETES MELLITUS MOD NPDR MACULAR ED OS | Diabetes_Type2 | E113312 |
| E11.3313 | DX10 | TYPE 2 DIABETES MELLITUS MOD NPDR MACULAR ED BIL | Diabetes_Type2 | E113313 |
| E11.3319 | DX10 | TYPE 2 DIABETES MELLITUS MOD NPDR MAC ED UNS EYE | Diabetes_Type2 | E113319 |
| E11.339 | DX10 | TYPE 2 DM W/MOD NONPROLIF DM RETINOPATHY W/O ME | Diabetes_Type2 | E11339 |

| | | | | |
|---|---|---|---|---|
| E11.3391 | DX10 | TYPE 2 DIABETES MELLITUS MOD NPDR W/O MAC ED OD | Diabetes_Type2 | E113391 |
| E11.3392 | DX10 | TYPE 2 DIABETES MELLITUS MOD NPDR W/O MAC ED OS | Diabetes_Type2 | E113392 |
| E11.3393 | DX10 | TYPE 2 DIABETES MELLITUS MOD NPDR W/O MAC ED BIL | Diabetes_Type2 | E113393 |
| E11.3399 | DX10 | TYPE 2 DIABETES MELLITUS MOD NPDR W/O ME UNS EYE | Diabetes_Type2 | E113399 |
| E11.34 | DX10 | TYPE 2 DM W/SEVERE NONPROLIF DIAB RETINOPATHY | Diabetes_Type2 | E1134 |
| E11.341 | DX10 | TYPE 2 DM W/SEV NONPROLIF DIAB RETINOPATHY W/ME | Diabetes_Type2 | E11341 |
| E11.3411 | DX10 | TYPE 2 DIABETES MELLITUS SVR NPDR MACULAR ED OD | Diabetes_Type2 | E113411 |
| E11.3412 | DX10 | TYPE 2 DIABETES MELLITUS SVR NPDR MACULAR ED OS | Diabetes_Type2 | E113412 |
| E11.3413 | DX10 | TYPE 2 DIABETES MELLITUS SVR NPDR MACULAR ED BIL | Diabetes_Type2 | E113413 |
| E11.3419 | DX10 | TYPE 2 DIABETES MELLITUS SVR NPDR MAC ED UNS EYE | Diabetes_Type2 | E113419 |
| E11.349 | DX10 | TYPE 2 DM W/SEV NONPROLIF DIAB RETINOPATH W/O ME | Diabetes_Type2 | E11349 |
| E11.3491 | DX10 | TYPE 2 DIABETES MELLITUS SVR NPDR W/O MAC ED OD | Diabetes_Type2 | E113491 |
| E11.3492 | DX10 | TYPE 2 DIABETES MELLITUS SVR NPDR W/O MAC ED OS | Diabetes_Type2 | E113492 |
| E11.3493 | DX10 | TYPE 2 DIABETES MELLITUS SVR NPDR W/O MAC ED BIL | Diabetes_Type2 | E113493 |
| E11.3499 | DX10 | TYPE 2 DIABETES MELLITUS SVR NPDR W/O ME UNS EYE | Diabetes_Type2 | E113499 |
| E11.35 | DX10 | TYPE 2 DM W/PROLIFERATIVE DIABETIC RETINOPATHY | Diabetes_Type2 | E1135 |
| E11.351 | DX10 | TYPE 2 DM W/PROLIFERATIVE DIAB RETINOPATHY W/ME | Diabetes_Type2 | E11351 |
| E11.3511 | DX10 | TYPE 2 DIABETES MELLITUS PDR MACULAR EDEMA OD | Diabetes_Type2 | E113511 |
| E11.3512 | DX10 | TYPE 2 DIABETES MELLITUS PDR MACULAR EDEMA OS | Diabetes_Type2 | E113512 |
| E11.3513 | DX10 | TYPE 2 DIABETES MELLITUS PDR MACULAR EDEMA BILAT | Diabetes_Type2 | E113513 |
| E11.3519 | DX10 | TYPE 2 DIABETES MELLITUS PDR MACULAR ED UNS EYE | Diabetes_Type2 | E113519 |
| E11.359 | DX10 | TYPE 2 DM PROLIF DM RETINOPATHY NO MACULAR EDEMA | Diabetes_Type2 | E11359 |
| E11.3591 | DX10 | TYPE 2 DIABETES MELLITUS PDR W/O MACULAR ED OD | Diabetes_Type2 | E113591 |
| E11.3592 | DX10 | TYPE 2 DIABETES MELLITUS PDR W/O MACULAR ED OS | Diabetes_Type2 | E113592 |
| E11.3593 | DX10 | TYPE 2 DIABETES MELLITUS PDR W/O MACULAR ED BIL | Diabetes_Type2 | E113593 |
| E11.3599 | DX10 | TYPE 2 DIABETES MELLITUS PDR W/O MAC ED UNS EYE | Diabetes_Type2 | E113599 |
| E11.36 | DX10 | TYPE 2 DIABETES MELLITUS WITH DIABETIC CATARACT | Diabetes_Type2 | E1136 |
| E11.37 | DX10 | TYPE 2 DIAB MELLITUS DIAB MAC EDEMA RSLVD FLW TX | Diabetes_Type2 | E1137 |
| E11.37X1 | DX10 | TYPE 2 DIABETES MELLITUS DIAB ME RSLVD FLW TX OD | Diabetes_Type2 | E1137X1 |
| E11.37X2 | DX10 | TYPE 2 DIABETES MELLITUS DIAB ME RSLVD FLW TX OS | Diabetes_Type2 | E1137X2 |
| E11.37X3 | DX10 | TYPE 2 DIABETES MELLITUS DIAB ME RSLVD FLW TX BL | Diabetes_Type2 | E1137X3 |
| E11.37X9 | DX10 | TYPE 2 DIAB MELLITUS DB ME RSLVD FLW TX UNS EYE | Diabetes_Type2 | E1137X9 |
| E11.39 | DX10 | TYPE 2 DIABETES MELLITUS OTH DIAB OPHTHALM COMP | Diabetes_Type2 | E1139 |
| E11.4 | DX10 | TYPE 2 DIABETES MELLITUS W/NEUROLOGICAL COMP | Diabetes_Type2 | E114 |

| | | | | |
|---|---|---|---|---|
| E11.40 | DX10 | TYPE 2 DM WITH DIABETIC NEUROPATHY UNSPECIFIED | Diabetes_Type2 | E1140 |
| E11.41 | DX10 | TYPE 2 DIABETES MELLITUS W/DIAB MONONEUROPATHY | Diabetes_Type2 | E1141 |
| E11.42 | DX10 | TYPE 2 DIABETES MELLITUS W/DIAB POLYNEUROPATHY | Diabetes_Type2 | E1142 |
| E11.43 | DX10 | TYPE 2 DM W/DIABETIC AUTONOMIC POLYNEUROPATHY | Diabetes_Type2 | E1143 |
| E11.44 | DX10 | TYPE 2 DIABETES MELLITUS W/DIABETIC AMYOTROPHY | Diabetes_Type2 | E1144 |
| E11.49 | DX10 | TYPE 2 DIABETES MELLITUS W/OTH DIAB NEURO COMP | Diabetes_Type2 | E1149 |
| E11.5 | DX10 | TYPE 2 DIABETES MELLITUS W/CIRCULATORY COMP | Diabetes_Type2 | E115 |
| E11.51 | DX10 | TYPE 2 DM W/DIAB PERIPH ANGIOPATHY W/O GANGRENE | Diabetes_Type2 | E1151 |
| E11.52 | DX10 | TYPE 2 DM W/DIAB PERIPH ANGIOPATHY W/GANGRENE | Diabetes_Type2 | E1152 |
| E11.59 | DX10 | TYPE 2 DIABETES MELLITUS W/OTH CIRCULATORY COMP | Diabetes_Type2 | E1159 |
| E11.6 | DX10 | TYPE 2 DIABETES MELLITUS W/OTH SPEC COMPLICATION | Diabetes_Type2 | E116 |
| E11.61 | DX10 | TYPE 2 DIABETES MELLITUS W/DIABETIC ARTHROPATHY | Diabetes_Type2 | E1161 |
| E11.610 | DX10 | TYPE 2 DM W/DIABETIC NEUROPATHIC ARTHROPATHY | Diabetes_Type2 | E11610 |
| E11.618 | DX10 | TYPE 2 DIABETES MELLITUS W/OTH DIAB ARTHROPATHY | Diabetes_Type2 | E11618 |
| E11.62 | DX10 | TYPE 2 DIABETES MELLITUS WITH SKIN COMPLICATIONS | Diabetes_Type2 | E1162 |
| E11.620 | DX10 | TYPE 2 DIABETES MELLITUS W/DIABETIC DERMATITIS | Diabetes_Type2 | E11620 |
| E11.621 | DX10 | TYPE 2 DIABETES MELLITUS WITH FOOT ULCER | Diabetes_Type2 | E11621 |
| E11.622 | DX10 | TYPE 2 DIABETES MELLITUS WITH OTHER SKIN ULCER | Diabetes_Type2 | E11622 |
| E11.628 | DX10 | TYPE 2 DIABETES MELLITUS W/OTH SKIN COMP | Diabetes_Type2 | E11628 |
| E11.63 | DX10 | TYPE 2 DIABETES MELLITUS WITH ORAL COMPLICATIONS | Diabetes_Type2 | E1163 |
| E11.630 | DX10 | TYPE 2 DIABETES MELLITUS W/PERIODONTAL DISEASE | Diabetes_Type2 | E11630 |
| E11.638 | DX10 | TYPE 2 DIABETES MELLITUS W/OTH ORAL COMP | Diabetes_Type2 | E11638 |
| E11.64 | DX10 | TYPE 2 DIABETES MELLITUS WITH HYPOGLYCEMIA | Diabetes_Type2 | E1164 |
| E11.641 | DX10 | TYPE 2 DIABETES MELLITUS W/HYPOGLYCEMIA W/COMA | Diabetes_Type2 | E11641 |
| E11.649 | DX10 | TYPE 2 DIABETES MELLITUS W/HYPOGLYCEMIA W/O COMA | Diabetes_Type2 | E11649 |
| E11.65 | DX10 | TYPE 2 DIABETES MELLITUS WITH HYPERGLYCEMIA | Diabetes_Type2 | E1165 |
| E11.69 | DX10 | TYPE 2 DIABETES MELLITUS W/OTH SPEC COMPLICATION | Diabetes_Type2 | E1169 |
| E11.8 | DX10 | TYPE 2 DIABETES MELLITUS W/UNS COMPLICATIONS | Diabetes_Type2 | E118 |
| E11.9 | DX10 | TYPE 2 DIABETES MELLITUS WITHOUT COMPLICATIONS | Diabetes_Type2 | E119 |
| Q90* | DX10 | down syndrom | Down_Syndrom | Q90* |
| 585.6 | DX10 | End stage renal disease | EndStageRenalDisease_Dialysis | 5856 |
| 38.95 | DX10 | Venous catheritazation for dialysis | EndStageRenalDisease_Dialysis | 3895 |
| 403.01 | DX10 | Hypertensive chronic kidney disease, malignant, with chronic kidney disease stage V or end stage renal disease | EndStageRenalDisease_Dialysis | 40301 |
| 403.11 | DX10 | Hypertensive chronic kidney disease, benign, with chronic kidney disease stage V or end stage renal disease | EndStageRenalDisease_Dialysis | 40311 |

| | | | | |
|---|---|---|---|---|
| 403.91 | DX10 | Hypertensive chronic kidney disease, unspecified, with chronic kidney disease stage V or end stage renal disease | EndStageRenalDisease_Dialysis | 40391 |
| 404.02 | DX10 | Hypertensive heart and chronic kidney disease, malignant, without heart failure and with chronic kidney disease stage V or end stage renal disease | EndStageRenalDisease_Dialysis | 40402 |
| 404.03 | DX10 | Hypertensive heart and chronic kidney disease, malignant, with heart failure and with chronic kidney disease stage V or end stage renal disease | EndStageRenalDisease_Dialysis | 40403 |
| 404.12 | DX10 | Hypertensive heart and chronic kidney disease, benign, without heart failure and with chronic kidney disease stage V or end stage renal disease | EndStageRenalDisease_Dialysis | 40412 |
| 404.13 | DX10 | Hypertensive heart and chronic kidney disease, benign, with heart failure and chronic kidney disease stage V or end stage renal disease | EndStageRenalDisease_Dialysis | 40413 |
| 404.92 | DX10 | Hypertensive heart and chronic kidney disease, unspecified, without heart failure and with chronic kidney disease stage V or end stage renal disease | EndStageRenalDisease_Dialysis | 40492 |
| 404.93 | DX10 | Hypertensive heart and chronic kidney disease, unspecified, with heart failure and chronic kidney disease stage V or end stage renal disease | EndStageRenalDisease_Dialysis | 40493 |
| 585.5 | DX10 | Chronic kidney disease, Stage V | EndStageRenalDisease_Dialysis | 5855 |
| N18.6 | DX10 | End stage renal disease | EndStageRenalDisease_Dialysis | N186 |
| I12.0 | DX10 | Hypertensive chronic kidney disease with stage 5 chronic kidney disease or end stage renal disease | EndStageRenalDisease_Dialysis | I120 |
| I13.11 | DX10 | Hypertensive heart and chronic kidney disease without heart failure with stage 5 chronic kidney disease, or end stage renal disease | EndStageRenalDisease_Dialysis | I1311 |
| I13.2 | DX10 | Hypertensive heart and chronic kidney disease with heart failure and with stage 5 chronic kidney disease, or end stage renal disease | EndStageRenalDisease_Dialysis | I132 |
| N18.5 | DX10 | Chronic kidney disease, Stage V | EndStageRenalDisease_Dialysis | N185 |
| 342* | DX10 | HEMIPL_ROM | Hemiplegia | 342* |
| 344* | DX10 | HEMIPL_ROM | Hemiplegia | 344* |
| G041 | DX10 | HEMIPL_ROM | Hemiplegia | G041 |
| G114 | DX10 | HEMIPL_ROM | Hemiplegia | G114 |
| G801 | DX10 | HEMIPL_ROM | Hemiplegia | G801 |
| G802 | DX10 | HEMIPL_ROM | Hemiplegia | G802 |
| G81 | DX10 | HEMIPL_ROM | Hemiplegia | G81 |
| G810 | DX10 | HEMIPL_ROM | Hemiplegia | G810 |
| G8100 | DX10 | HEMIPL_ROM | Hemiplegia | G8100 |
| G8101 | DX10 | HEMIPL_ROM | Hemiplegia | G8101 |
| G8102 | DX10 | HEMIPL_ROM | Hemiplegia | G8102 |
| G8103 | DX10 | HEMIPL_ROM | Hemiplegia | G8103 |
| G8104 | DX10 | HEMIPL_ROM | Hemiplegia | G8104 |
| G811 | DX10 | HEMIPL_ROM | Hemiplegia | G811 |
| G8110 | DX10 | HEMIPL_ROM | Hemiplegia | G8110 |
| G8111 | DX10 | HEMIPL_ROM | Hemiplegia | G8111 |
| G8112 | DX10 | HEMIPL_ROM | Hemiplegia | G8112 |
| G8113 | DX10 | HEMIPL_ROM | Hemiplegia | G8113 |
| G8114 | DX10 | HEMIPL_ROM | Hemiplegia | G8114 |
| G819 | DX10 | HEMIPL_ROM | Hemiplegia | G819 |
| G8190 | DX10 | HEMIPL_ROM | Hemiplegia | G8190 |
| G8191 | DX10 | HEMIPL_ROM | Hemiplegia | G8191 |
| G8192 | DX10 | HEMIPL_ROM | Hemiplegia | G8192 |
| G8193 | DX10 | HEMIPL_ROM | Hemiplegia | G8193 |

| | | | | |
|---|---|---|---|---|
| G8194 | DX10 | HEMIPL_ROM | Hemiplegia | G8194 |
| G82 | DX10 | HEMIPL_ROM | Hemiplegia | G82 |
| G822 | DX10 | HEMIPL_ROM | Hemiplegia | G822 |
| G8220 | DX10 | HEMIPL_ROM | Hemiplegia | G8220 |
| G8221 | DX10 | HEMIPL_ROM | Hemiplegia | G8221 |
| G8222 | DX10 | HEMIPL_ROM | Hemiplegia | G8222 |
| G825 | DX10 | HEMIPL_ROM | Hemiplegia | G825 |
| G8250 | DX10 | HEMIPL_ROM | Hemiplegia | G8250 |
| G8251 | DX10 | HEMIPL_ROM | Hemiplegia | G8251 |
| G8252 | DX10 | HEMIPL_ROM | Hemiplegia | G8252 |
| G8253 | DX10 | HEMIPL_ROM | Hemiplegia | G8253 |
| G8254 | DX10 | HEMIPL_ROM | Hemiplegia | G8254 |
| G83 | DX10 | HEMIPL_ROM | Hemiplegia | G83 |
| G830 | DX10 | HEMIPL_ROM | Hemiplegia | G830 |
| G831 | DX10 | HEMIPL_ROM | Hemiplegia | G831 |
| G8310 | DX10 | HEMIPL_ROM | Hemiplegia | G8310 |
| G8311 | DX10 | HEMIPL_ROM | Hemiplegia | G8311 |
| G8312 | DX10 | HEMIPL_ROM | Hemiplegia | G8312 |
| G8313 | DX10 | HEMIPL_ROM | Hemiplegia | G8313 |
| G8314 | DX10 | HEMIPL_ROM | Hemiplegia | G8314 |
| G832 | DX10 | HEMIPL_ROM | Hemiplegia | G832 |
| G8320 | DX10 | HEMIPL_ROM | Hemiplegia | G8320 |
| G8321 | DX10 | HEMIPL_ROM | Hemiplegia | G8321 |
| G8322 | DX10 | HEMIPL_ROM | Hemiplegia | G8322 |
| G8323 | DX10 | HEMIPL_ROM | Hemiplegia | G8323 |
| G8324 | DX10 | HEMIPL_ROM | Hemiplegia | G8324 |
| G833 | DX10 | HEMIPL_ROM | Hemiplegia | G833 |
| G8330 | DX10 | HEMIPL_ROM | Hemiplegia | G8330 |
| G8331 | DX10 | HEMIPL_ROM | Hemiplegia | G8331 |
| G8332 | DX10 | HEMIPL_ROM | Hemiplegia | G8332 |
| G8333 | DX10 | HEMIPL_ROM | Hemiplegia | G8333 |
| G8334 | DX10 | HEMIPL_ROM | Hemiplegia | G8334 |
| G834 | DX10 | HEMIPL_ROM | Hemiplegia | G834 |
| G835 | DX10 | HEMIPL_ROM | Hemiplegia | G835 |
| G838 | DX10 | HEMIPL_ROM | Hemiplegia | G838 |
| G8381 | DX10 | HEMIPL_ROM | Hemiplegia | G8381 |
| G8382 | DX10 | HEMIPL_ROM | Hemiplegia | G8382 |
| G8383 | DX10 | HEMIPL_ROM | Hemiplegia | G8383 |
| G8384 | DX10 | HEMIPL_ROM | Hemiplegia | G8384 |
| G8389 | DX10 | HEMIPL_ROM | Hemiplegia | G8389 |
| G839 | DX10 | HEMIPL_ROM | Hemiplegia | G839 |
| B20 | DX10 | Human immunodeficiency virus [HIV] disease | HIV | B20 |
| B97.35 | DX10 | Human immunodeficiency virus type 2 as cause of diseases classified elsewhere | HIV | B9735 |
| Z21 | DX10 | Asymptomatic HIV infection status | HIV | Z21 |
| O98.7 | DX10 | HIV in pregnancy | HIV | O987 |
| O98.71 | DX10 | HIV disease complicating pregnancy | HIV | O9871 |
| O98.72 | DX10 | HIV disease complicating childbirth | HIV | O9872 |
| O98.73 | DX10 | HIV disease complicating the puerperium | HIV | O9873 |
| D80 | DX10 | Immunodeficiency with predominantly antibody defects | Immunodeficiency | D80 |
| D80.0 | DX10 | Hereditary hypogammaglobulinemia | Immunodeficiency | D800 |
| D80.1 | DX10 | Nonfamilial hypogammaglobulinemia | Immunodeficiency | D801 |
| D80.2 | DX10 | Selective deficiency of immunoglobulin A [IgA] | Immunodeficiency | D802 |
| D80.3 | DX10 | Selective deficiency of immunoglobulin G [IgG] subclasses | Immunodeficiency | D803 |

| | | | | |
|---|---|---|---|---|
| D80.4 | DX10 | Selective deficiency of immunoglobulin M [IgM] | Immunodeficiency | D804 |
| D80.5 | DX10 | Immunodeficiency with increased immunoglobulin M [IgM] | Immunodeficiency | D805 |
| D80.6 | DX10 | Antibody defic w near-norm immunoglob or w hyperimmunoglob | Immunodeficiency | D806 |
| D80.7 | DX10 | Transient hypogammaglobulinemia of infancy | Immunodeficiency | D807 |
| D80.8 | DX10 | Other immunodeficiencies with predominantly antibody defects | Immunodeficiency | D808 |
| D80.9 | DX10 | Immunodeficiency with predominantly antibody defects, unsp | Immunodeficiency | D809 |
| D81 | DX10 | Combined immunodeficiencies | Immunodeficiency | D81 |
| D81.0 | DX10 | Severe combined immunodeficiency with reticular dysgenesis | Immunodeficiency | D810 |
| D81.1 | DX10 | Severe combined immunodeficiency w low T- and B-cell numbers | Immunodeficiency | D811 |
| D81.2 | DX10 | Severe combined immunodef w low or normal B-cell numbers | Immunodeficiency | D812 |
| D81.3 | DX10 | Adenosine deaminase [ADA] deficiency | Immunodeficiency | D813 |
| D81.30 | DX10 | Adenosine deaminase [ADA] deficiency, unspecified | Immunodeficiency | D8130 |
| D81.31 | DX10 | Severe combined immunodeficiency due to ADA deficiency | Immunodeficiency | D8131 |
| D81.32 | DX10 | Adenosine deaminase 2 deficiency | Immunodeficiency | D8132 |
| D81.39 | DX10 | Other adenosine deaminase deficiency | Immunodeficiency | D8139 |
| D81.4 | DX10 | Nezelof's syndrome | Immunodeficiency | D814 |
| D81.5 | DX10 | Purine nucleoside phosphorylase [PNP] deficiency | Immunodeficiency | D815 |
| D81.6 | DX10 | Major histocompatibility complex class I deficiency | Immunodeficiency | D816 |
| D81.7 | DX10 | Major histocompatibility complex class II deficiency | Immunodeficiency | D817 |
| D81.8 | DX10 | Other combined immunodeficiencies | Immunodeficiency | D818 |
| D81.81 | DX10 | Biotin-dependent carboxylase deficiency | Immunodeficiency | D8181 |
| D81.810 | DX10 | Biotinidase deficiency | Immunodeficiency | D81810 |
| D81.818 | DX10 | Other biotin-dependent carboxylase deficiency | Immunodeficiency | D81818 |
| D81.819 | DX10 | Biotin-dependent carboxylase deficiency, unspecified | Immunodeficiency | D81819 |
| D81.89 | DX10 | Other combined immunodeficiencies | Immunodeficiency | D8189 |
| D81.9 | DX10 | Combined immunodeficiency, unspecified | Immunodeficiency | D819 |
| D82 | DX10 | Immunodeficiency associated with other major defects | Immunodeficiency | D82 |
| D82.0 | DX10 | Wiskott-Aldrich syndrome | Immunodeficiency | D820 |
| D82.1 | DX10 | Di George's syndrome | Immunodeficiency | D821 |

| | | | | |
|---|---|---|---|---|
| D82.2 | DX10 | Immunodeficiency with short-limbed stature | Immunodeficiency | D822 |
| D82.3 | DX10 | Immunodef fol heredit defctv response to Epstein-Barr virus | Immunodeficiency | D823 |
| D82.4 | DX10 | Hyperimmunoglobulin E [IgE] syndrome | Immunodeficiency | D824 |
| D82.8 | DX10 | Immunodeficiency associated with oth major defects | Immunodeficiency | D828 |
| D82.9 | DX10 | Immunodeficiency associated with major defect, unspecified | Immunodeficiency | D829 |
| D83 | DX10 | Common variable immunodeficiency | Immunodeficiency | D83 |
| D83.0 | DX10 | Com variab immunodef w predom abnlt of B-cell nums & functn | Immunodeficiency | D830 |
| D83.1 | DX10 | Com variab immunodef w predom immunoreg T-cell disorders | Immunodeficiency | D831 |
| D83.2 | DX10 | Common variable immunodef w autoantibodies to B- or T-cells | Immunodeficiency | D832 |
| D83.8 | DX10 | Other common variable immunodeficiencies | Immunodeficiency | D838 |
| D83.9 | DX10 | Common variable immunodeficiency, unspecified | Immunodeficiency | D839 |
| D84.0 | DX10 | Lymphocyte function antigen-1 [LFA-1] defect | Immunodeficiency | D840 |
| D84.1 | DX10 | Defects in the complement system | Immunodeficiency | D841 |
| D89.0 | DX10 | Polyclonal hypergammaglobulinemia | Immunodeficiency | D890 |
| D89.1 | DX10 | Cryoglobulinemia | Immunodeficiency | D891 |
| D89.2 | DX10 | Hypergammaglobulinemia, unspecified | Immunodeficiency | D892 |
| D89.3 | DX10 | Immune reconstitution syndrome | Immunodeficiency | D893 |
| D89.82 | DX10 | Autoimmune lymphoproliferative syndrome [ALPS] | Immunodeficiency | D8982 |
| D89.83 | DX10 | Cytokine release syndrome | Immunodeficiency | D8983 |
| D89.831 | DX10 | Cytokine release syndrome, grade 1 | Immunodeficiency | D89831 |
| D89.832 | DX10 | Cytokine release syndrome, grade 2 | Immunodeficiency | D89832 |
| D89.833 | DX10 | Cytokine release syndrome, grade 3 | Immunodeficiency | D89833 |
| D89.834 | DX10 | Cytokine release syndrome, grade 4 | Immunodeficiency | D89834 |
| D89.835 | DX10 | Cytokine release syndrome, grade 5 | Immunodeficiency | D89835 |
| D89.839 | DX10 | Cytokine release syndrome, grade unspecified | Immunodeficiency | D89839 |
| D89.89 | DX10 | Oth disrd involving the immune mechanism, NEC | Immunodeficiency | D8989 |
| D89.9 | DX10 | Disorder involving the immune mechanism, unspecified | Immunodeficiency | D899 |
| D70.0 | DX10 | Congenital agranulocytosis | Immunodeficiency | D700 |
| D70.1 | DX10 | Agranulocytosis secondary to cancer chemotherapy | Immunodeficiency | D701 |

| | | | | |
|---|---|---|---|---|
| D70.2 | DX10 | Other drug-induced agranulocytosis | Immunodeficiency | D702 |
| D70.3 | DX10 | Neutropenia due to infection | Immunodeficiency | D703 |
| D70.4 | DX10 | Cyclic neutropenia | Immunodeficiency | D704 |
| D70.8 | DX10 | Other neutropenia | Immunodeficiency | D708 |
| D70.9 | DX10 | Neutropenia, unspecified | Immunodeficiency | D709 |
| D71 | DX10 | Functional disorders of polymorphonuclear neutrophils | Immunodeficiency | D71 |
| D61.81 | DX10 | Pancytopenia | Immunodeficiency | D6181 |
| D70 | DX10 | Neutropenia | Immunodeficiency | D70 |
| D72.0 | DX10 | Genetic anomalies of leukocytes | Immunodeficiency | D720 |
| D72.81 | DX10 | Lymphocytopenia | Immunodeficiency | D7281 |
| D75.81 | DX10 | Myelofibrosis | Immunodeficiency | D7581 |
| D75.89 | DX10 | Other specified diseases of blood and blood-forming organs | Immunodeficiency | D7589 |
| D75.9 | DX10 | Blood diseases NOS | Immunodeficiency | D759 |
| D89 | DX10 | Polyclonal hypergammaglobulinemia | Immunodeficiency | D89 |
| Z85 | DX10 | Personal history of malignant neoplasm of unspecified digestive organ | Immunodeficiency | Z85 |
| F32.0 | DX10 | Major depressive disorder, single episode, mild | Mental_Health_Disorder | F320 |
| F32.A | DX10 | 0 | Mental_Health_Disorder | F32A |
| F32.1 | DX10 | Major depressive disorder, single episode, moderate | Mental_Health_Disorder | F321 |
| F32.2 | DX10 | Major depressv disord, single epsd, sev w/o psych features | Mental_Health_Disorder | F322 |
| F32.3 | DX10 | Major depressv disord, single epsd, severe w psych features | Mental_Health_Disorder | F323 |
| F32.4 | DX10 | Major depressv disorder, single episode, in partial remis | Mental_Health_Disorder | F324 |
| F32.81 | DX10 | Premenstrual dysphoric disorder | Mental_Health_Disorder | F3281 |
| F32.89 | DX10 | Other specified depressive episodes | Mental_Health_Disorder | F3289 |
| F32.9 | DX10 | Major depressive disorder, single episode, unspecified | Mental_Health_Disorder | F329 |
| F33.0 | DX10 | Major depressive disorder, recurrent, mild | Mental_Health_Disorder | F330 |
| F33.1 | DX10 | Major depressive disorder, recurrent, moderate | Mental_Health_Disorder | F331 |
| F33.2 | DX10 | Major depressv disorder, recurrent severe w/o psych features | Mental_Health_Disorder | F332 |
| F33.3 | DX10 | Major depressv disorder, recurrent, severe w psych symptoms | Mental_Health_Disorder | F333 |
| F33.40 | DX10 | Major depressive disorder, recurrent, in remission, unsp | Mental_Health_Disorder | F3340 |

| | | | | |
|---|---|---|---|---|
| F33.41 | DX10 | Major depressive disorder, recurrent, in partial remission | Mental_Health_Disorder | F3341 |
| F33.8 | DX10 | Other recurrent depressive disorders | Mental_Health_Disorder | F338 |
| F33.9 | DX10 | Major depressive disorder, recurrent, unspecified | Mental_Health_Disorder | F339 |
| F34.0 | DX10 | Cyclothymic disorder | Mental_Health_Disorder | F340 |
| F34.1 | DX10 | Dysthymic disorder | Mental_Health_Disorder | F341 |
| F34.9 | DX10 | Persistent mood [affective] disorder, unspecified | Mental_Health_Disorder | F349 |
| F39 | DX10 | Unspecified mood [affective] disorder | Mental_Health_Disorder | F39 |
| F43.21 | DX10 | Adjustment disorder with depressed mood | Mental_Health_Disorder | F4321 |
| F43.23 | DX10 | Adjustment disorder with mixed anxiety and depressed mood | Mental_Health_Disorder | F4323 |
| F30.10 | DX10 | Manic episode without psychotic symptoms, unspecified | Mental_Health_Disorder | F3010 |
| F30.11 | DX10 | Manic episode without psychotic symptoms, mild | Mental_Health_Disorder | F3011 |
| F30.12 | DX10 | Manic episode without psychotic symptoms, moderate | Mental_Health_Disorder | F3012 |
| F30.13 | DX10 | Manic episode, severe, without psychotic symptoms | Mental_Health_Disorder | F3013 |
| F30.2 | DX10 | Manic episode, severe with psychotic symptoms | Mental_Health_Disorder | F302 |
| F30.3 | DX10 | Manic episode in partial remission | Mental_Health_Disorder | F303 |
| F30.8 | DX10 | Other manic episodes | Mental_Health_Disorder | F308 |
| F30.9 | DX10 | Manic episode, unspecified | Mental_Health_Disorder | F309 |
| F31.0 | DX10 | Bipolar disorder, current episode hypomanic | Mental_Health_Disorder | F310 |
| F31.10 | DX10 | Bipolar disord, crnt episode manic w/o psych features, unsp | Mental_Health_Disorder | F3110 |
| F31.11 | DX10 | Bipolar disord, crnt episode manic w/o psych features, mild | Mental_Health_Disorder | F3111 |
| F31.12 | DX10 | Bipolar disord, crnt episode manic w/o psych features, mod | Mental_Health_Disorder | F3112 |
| F31.13 | DX10 | Bipolar disord, crnt epsd manic w/o psych features, severe | Mental_Health_Disorder | F3113 |
| F31.2 | DX10 | Bipolar disord, crnt episode manic severe w psych features | Mental_Health_Disorder | F312 |
| F31.3 | DX10 | 0 | Mental_Health_Disorder | F313 |
| F31.30 | DX10 | Bipolar disord, crnt epsd depress, mild or mod severt, unsp | Mental_Health_Disorder | F3130 |
| F31.31 | DX10 | Bipolar disorder, current episode depressed, mild | Mental_Health_Disorder | F3131 |
| F31.32 | DX10 | Bipolar disorder, current episode depressed, moderate | Mental_Health_Disorder | F3132 |
| F31.4 | DX10 | Bipolar disord, crnt epsd depress, sev, w/o psych features | Mental_Health_Disorder | F314 |
| F31.5 | DX10 | Bipolar disord, crnt epsd depress, severe, w psych features | Mental_Health_Disorder | F315 |

| | | | | |
|---|---|---|---|---|
| F31.6 | DX10 | 0 | Mental_Health_Disorder | F316 |
| F31.60 | DX10 | Bipolar disorder, current episode mixed, unspecified | Mental_Health_Disorder | F3160 |
| F31.61 | DX10 | Bipolar disorder, current episode mixed, mild | Mental_Health_Disorder | F3161 |
| F31.62 | DX10 | Bipolar disorder, current episode mixed, moderate | Mental_Health_Disorder | F3162 |
| F31.63 | DX10 | Bipolar disord, crnt epsd mixed, severe, w/o psych features | Mental_Health_Disorder | F3163 |
| F31.64 | DX10 | Bipolar disord, crnt episode mixed, severe, w psych features | Mental_Health_Disorder | F3164 |
| F31.70 | DX10 | Bipolar disord, currently in remis, most recent episode unsp | Mental_Health_Disorder | F3170 |
| F31.71 | DX10 | Bipolar disord, in partial remis, most recent epsd hypomanic | Mental_Health_Disorder | F3171 |
| F31.73 | DX10 | Bipolar disord, in partial remis, most recent episode manic | Mental_Health_Disorder | F3173 |
| F31.75 | DX10 | Bipolar disord, in partial remis, most recent epsd depress | Mental_Health_Disorder | F3175 |
| F31.77 | DX10 | Bipolar disord, in partial remis, most recent episode mixed | Mental_Health_Disorder | F3177 |
| F31.81 | DX10 | Bipolar II disorder | Mental_Health_Disorder | F3181 |
| F31.89 | DX10 | Other bipolar disorder | Mental_Health_Disorder | F3189 |
| F31.9 | DX10 | Bipolar disorder, unspecified | Mental_Health_Disorder | F319 |
| F20.0 | DX10 | Paranoid schizophrenia | Mental_Health_Disorder | F200 |
| F20.1 | DX10 | Disorganized schizophrenia | Mental_Health_Disorder | F201 |
| F20.2 | DX10 | Catatonic schizophrenia | Mental_Health_Disorder | F202 |
| F20.3 | DX10 | Undifferentiated schizophrenia | Mental_Health_Disorder | F203 |
| F20.5 | DX10 | Residual schizophrenia | Mental_Health_Disorder | F205 |
| F20.81 | DX10 | Schizophreniform disorder | Mental_Health_Disorder | F2081 |
| F20.89 | DX10 | Other schizophrenia | Mental_Health_Disorder | F2089 |
| F20.9 | DX10 | Schizophrenia, unspecified | Mental_Health_Disorder | F209 |
| F25.0 | DX10 | Schizoaffective disorder, bipolar type | Mental_Health_Disorder | F250 |
| F25.1 | DX10 | Schizoaffective disorder, depressive type | Mental_Health_Disorder | F251 |
| F25.8 | DX10 | Other schizoaffective disorders | Mental_Health_Disorder | F258 |
| F25.9 | DX10 | Schizoaffective disorder, unspecified | Mental_Health_Disorder | F259 |
| F21 | DX10 | Schizotypal disorder | Mental_Health_Disorder | F21 |
| O90.6 | DX10 | Mental health disorder | Mental_Health_Disorder | O906 |
| E66.0 | DX10 | Obesity due to excess calories | Obesity_BMI_ge30 | E660 |

| | | | | |
|---|---|---|---|---|
| E66.01 | DX10 | Morbid (severe) obesity due to excess calories | Obesity_BMI_ge30 | E6601 |
| E66.09 | DX10 | Other obesity due to excess calories | Obesity_BMI_ge30 | E6609 |
| E66.1 | DX10 | Drug-induced obesity | Obesity_BMI_ge30 | E661 |
| E66.2 | DX10 | Morbid (severe) obesity with alveolar hypoventilation | Obesity_BMI_ge30 | E662 |
| E66.8 | DX10 | Other obesity | Obesity_BMI_ge30 | E668 |
| E66.9 | DX10 | Obesity, unspecified | Obesity_BMI_ge30 | E669 |
| Z68.30 | DX10 | Body mass index (BMI) 30.0-30.9, adult | Obesity_BMI_ge30 | Z6830 |
| Z68.31 | DX10 | Body mass index (BMI) 31.0-31.9, adult | Obesity_BMI_ge30 | Z6831 |
| Z68.32 | DX10 | Body mass index (BMI) 32.0-32.9, adult | Obesity_BMI_ge30 | Z6832 |
| Z68.33 | DX10 | Body mass index (BMI) 33.0-33.9, adult | Obesity_BMI_ge30 | Z6833 |
| Z68.34 | DX10 | Body mass index (BMI) 34.0-34.9, adult | Obesity_BMI_ge30 | Z6834 |
| Z68.35 | DX10 | Body mass index (BMI) 35.0-35.9, adult | Obesity_BMI_ge30 | Z6835 |
| Z68.36 | DX10 | Body mass index (BMI) 36.0-36.9, adult | Obesity_BMI_ge30 | Z6836 |
| Z68.37 | DX10 | Body mass index (BMI) 37.0-37.9, adult | Obesity_BMI_ge30 | Z6837 |
| Z68.38 | DX10 | Body mass index (BMI) 38.0-38.9, adult | Obesity_BMI_ge30 | Z6838 |
| Z68.39 | DX10 | Body mass index (BMI) 39.0-39.9, adult | Obesity_BMI_ge30 | Z6839 |
| Z68.4 | DX10 | Body mass index (BMI) 40 or greater, adult | Obesity_BMI_ge30 | Z684 |
| Z68.41 | DX10 | Body mass index (BMI) 40.0-44.9, adult | Obesity_BMI_ge30 | Z6841 |
| Z68.43 | DX10 | Body mass index (BMI) 50-59.9 , adult | Obesity_BMI_ge30 | Z6843 |
| Z68.44 | DX10 | Body mass index (BMI) 60.0-69.9, adult | Obesity_BMI_ge30 | Z6844 |
| Z68.45 | DX10 | Body mass index (BMI) 70 or greater, adult | Obesity_BMI_ge30 | Z6845 |
| Z68.54 | DX10 | BMI pediatric, greater than or equal to 95% for age | Obesity_BMI_ge30 | Z6854 |
| 332.0 | DX10 | Paralysis agitans | Parkinsons_Disease | 3320 |
| 332.1 | DX10 | Secondary Parkinsonism | Parkinsons_Disease | 3321 |
| 333.92 | DX10 | Neuroleptic malignant syndrome | Parkinsons_Disease | 33392 |
| G20 | DX10 | Parkinson's disease | Parkinsons_Disease | G20 |
| G21.4 | DX10 | Vascular parkinsonism | Parkinsons_Disease | G214 |
| G21.11 | DX10 | Neuroleptic induced parkinsonism | Parkinsons_Disease | G2111 |
| G21.19 | DX10 | Other drug induced secondary parkinsonism | Parkinsons_Disease | G2119 |

| | | | | |
|---|---|---|---|---|
| G21.2 | DX10 | Secondary parkinsonism due to other external agents | Parkinsons_Disease | G212 |
| G21.3 | DX10 | Postencephalitic parkinsonism | Parkinsons_Disease | G213 |
| G21.8 | DX10 | Other secondary parkinsonism | Parkinsons_Disease | G218 |
| G21.9 | DX10 | Secondary parkinsonism, unspecified | Parkinsons_Disease | G219 |
| G21.0 | DX10 | Malignant neuroleptic syndrome | Parkinsons_Disease | G210 |
| G21 | DX10 | Secondary parkinsonism | Parkinsons_Disease | G21 |
| G21.1 | DX10 | Other drug-induced secondary parkinsonism | Parkinsons_Disease | G211 |
| 440* | DX10 | PVD_ELIX | Peripheral_Vascular_Disorder | 440* |
| 4412* | DX10 | PVD_ELIX | Peripheral_Vascular_Disorder | 4412* |
| 4414* | DX10 | PVD_ELIX | Peripheral_Vascular_Disorder | 4414* |
| 4417* | DX10 | PVD_ELIX | Peripheral_Vascular_Disorder | 4417* |
| 4419* | DX10 | PVD_ELIX | Peripheral_Vascular_Disorder | 4419* |
| 4431* | DX10 | PVD_ELIX | Peripheral_Vascular_Disorder | 4431* |
| 4432* | DX10 | PVD_ELIX | Peripheral_Vascular_Disorder | 4432* |
| 4438* | DX10 | PVD_ELIX | Peripheral_Vascular_Disorder | 4438* |
| 4439* | DX10 | PVD_ELIX | Peripheral_Vascular_Disorder | 4439* |
| 4471* | DX10 | PVD_ELIX | Peripheral_Vascular_Disorder | 4471* |
| 5571* | DX10 | PVD_ELIX | Peripheral_Vascular_Disorder | 5571* |
| 5579* | DX10 | PVD_ELIX | Peripheral_Vascular_Disorder | 5579* |
| V434* | DX10 | PVD_ELIX | Peripheral_Vascular_Disorder | V434* |
| E0851 | DX10 | PVD_ELIX | Peripheral_Vascular_Disorder | E0851 |
| E0852 | DX10 | PVD_ELIX | Peripheral_Vascular_Disorder | E0852 |
| E0951 | DX10 | PVD_ELIX | Peripheral_Vascular_Disorder | E0951 |
| E0952 | DX10 | PVD_ELIX | Peripheral_Vascular_Disorder | E0952 |
| E1051 | DX10 | PVD_ELIX | Peripheral_Vascular_Disorder | E1051 |
| E1052 | DX10 | PVD_ELIX | Peripheral_Vascular_Disorder | E1052 |
| E1151 | DX10 | PVD_ELIX | Peripheral_Vascular_Disorder | E1151 |
| E1351 | DX10 | PVD_ELIX | Peripheral_Vascular_Disorder | E1351 |
| E1352 | DX10 | PVD_ELIX | Peripheral_Vascular_Disorder | E1352 |

| | | | | |
|---|---|---|---|---|
| I670 | DX10 | PVD_ELIX | Peripheral_Vascular_Disorder | I670 |
| I70 | DX10 | PVD_ELIX | Peripheral_Vascular_Disorder | I70 |
| I700 | DX10 | PVD_ELIX | Peripheral_Vascular_Disorder | I700 |
| I70.1 | DX10 | PVD_ELIX | Peripheral_Vascular_Disorder | I701 |
| I702 | DX10 | PVD_ELIX | Peripheral_Vascular_Disorder | I702 |
| I7020 | DX10 | PVD_ELIX | Peripheral_Vascular_Disorder | I7020 |
| I70201 | DX10 | PVD_ELIX | Peripheral_Vascular_Disorder | I70201 |
| I70202 | DX10 | PVD_ELIX | Peripheral_Vascular_Disorder | I70202 |
| I70203 | DX10 | PVD_ELIX | Peripheral_Vascular_Disorder | I70203 |
| I70208 | DX10 | PVD_ELIX | Peripheral_Vascular_Disorder | I70208 |
| I70209 | DX10 | PVD_ELIX | Peripheral_Vascular_Disorder | I70209 |
| I7021 | DX10 | PVD_ELIX | Peripheral_Vascular_Disorder | I7021 |
| I70211 | DX10 | PVD_ELIX | Peripheral_Vascular_Disorder | I70211 |
| I70212 | DX10 | PVD_ELIX | Peripheral_Vascular_Disorder | I70212 |
| I70213 | DX10 | PVD_ELIX | Peripheral_Vascular_Disorder | I70213 |
| I70218 | DX10 | PVD_ELIX | Peripheral_Vascular_Disorder | I70218 |
| I70219 | DX10 | PVD_ELIX | Peripheral_Vascular_Disorder | I70219 |
| I7022 | DX10 | PVD_ELIX | Peripheral_Vascular_Disorder | I7022 |
| I70221 | DX10 | PVD_ELIX | Peripheral_Vascular_Disorder | I70221 |
| I70222 | DX10 | PVD_ELIX | Peripheral_Vascular_Disorder | I70222 |
| I70223 | DX10 | PVD_ELIX | Peripheral_Vascular_Disorder | I70223 |
| I70228 | DX10 | PVD_ELIX | Peripheral_Vascular_Disorder | I70228 |
| I70229 | DX10 | PVD_ELIX | Peripheral_Vascular_Disorder | I70229 |
| I7023 | DX10 | PVD_ELIX | Peripheral_Vascular_Disorder | I7023 |
| I70231 | DX10 | PVD_ELIX | Peripheral_Vascular_Disorder | I70231 |
| I70232 | DX10 | PVD_ELIX | Peripheral_Vascular_Disorder | I70232 |
| I70233 | DX10 | PVD_ELIX | Peripheral_Vascular_Disorder | I70233 |
| I70234 | DX10 | PVD_ELIX | Peripheral_Vascular_Disorder | I70234 |
| I70235 | DX10 | PVD_ELIX | Peripheral_Vascular_Disorder | I70235 |

| | | | | |
|---|---|---|---|---|
| I70238 | DX10 | PVD_ELIX | Peripheral_Vascular_ Disorder | I70238 |
| I70239 | DX10 | PVD_ELIX | Peripheral_Vascular_ Disorder | I70239 |
| I7024 | DX10 | PVD_ELIX | Peripheral_Vascular_ Disorder | I7024 |
| I70241 | DX10 | PVD_ELIX | Peripheral_Vascular_ Disorder | I70241 |
| I70242 | DX10 | PVD_ELIX | Peripheral_Vascular_ Disorder | I70242 |
| I70243 | DX10 | PVD_ELIX | Peripheral_Vascular_ Disorder | I70243 |
| I70244 | DX10 | PVD_ELIX | Peripheral_Vascular_ Disorder | I70244 |
| I70245 | DX10 | PVD_ELIX | Peripheral_Vascular_ Disorder | I70245 |
| I70248 | DX10 | PVD_ELIX | Peripheral_Vascular_ Disorder | I70248 |
| I70249 | DX10 | PVD_ELIX | Peripheral_Vascular_ Disorder | I70249 |
| I7025 | DX10 | PVD_ELIX | Peripheral_Vascular_ Disorder | I7025 |
| I7026 | DX10 | PVD_ELIX | Peripheral_Vascular_ Disorder | I7026 |
| I70261 | DX10 | PVD_ELIX | Peripheral_Vascular_ Disorder | I70261 |
| I70262 | DX10 | PVD_ELIX | Peripheral_Vascular_ Disorder | I70262 |
| I70263 | DX10 | PVD_ELIX | Peripheral_Vascular_ Disorder | I70263 |
| I70268 | DX10 | PVD_ELIX | Peripheral_Vascular_ Disorder | I70268 |
| I70269 | DX10 | PVD_ELIX | Peripheral_Vascular_ Disorder | I70269 |
| I7029 | DX10 | PVD_ELIX | Peripheral_Vascular_ Disorder | I7029 |
| I70291 | DX10 | PVD_ELIX | Peripheral_Vascular_ Disorder | I70291 |
| I70292 | DX10 | PVD_ELIX | Peripheral_Vascular_ Disorder | I70292 |
| I70293 | DX10 | PVD_ELIX | Peripheral_Vascular_ Disorder | I70293 |
| I70298 | DX10 | PVD_ELIX | Peripheral_Vascular_ Disorder | I70298 |
| I70299 | DX10 | PVD_ELIX | Peripheral_Vascular_ Disorder | I70299 |
| I703 | DX10 | PVD_ELIX | Peripheral_Vascular_ Disorder | I703 |
| I7030 | DX10 | PVD_ELIX | Peripheral_Vascular_ Disorder | I7030 |
| I70301 | DX10 | PVD_ELIX | Peripheral_Vascular_ Disorder | I70301 |
| I70302 | DX10 | PVD_ELIX | Peripheral_Vascular_ Disorder | I70302 |
| I70303 | DX10 | PVD_ELIX | Peripheral_Vascular_ Disorder | I70303 |
| I70308 | DX10 | PVD_ELIX | Peripheral_Vascular_ Disorder | I70308 |

| | | | |
|---|---|---|---|
| I70309 | DX10 | PVD_ELIX | Peripheral_Vascular_Disorder I70309 |
| I7031 | DX10 | PVD_ELIX | Peripheral_Vascular_Disorder I7031 |
| I70311 | DX10 | PVD_ELIX | Peripheral_Vascular_Disorder I70311 |
| I70312 | DX10 | PVD_ELIX | Peripheral_Vascular_Disorder I70312 |
| I70313 | DX10 | PVD_ELIX | Peripheral_Vascular_Disorder I70313 |
| I70318 | DX10 | PVD_ELIX | Peripheral_Vascular_Disorder I70318 |
| I70319 | DX10 | PVD_ELIX | Peripheral_Vascular_Disorder I70319 |
| I7032 | DX10 | PVD_ELIX | Peripheral_Vascular_Disorder I7032 |
| I70321 | DX10 | PVD_ELIX | Peripheral_Vascular_Disorder I70321 |
| I70322 | DX10 | PVD_ELIX | Peripheral_Vascular_Disorder I70322 |
| I70323 | DX10 | PVD_ELIX | Peripheral_Vascular_Disorder I70323 |
| I70328 | DX10 | PVD_ELIX | Peripheral_Vascular_Disorder I70328 |
| I70329 | DX10 | PVD_ELIX | Peripheral_Vascular_Disorder I70329 |
| I7033 | DX10 | PVD_ELIX | Peripheral_Vascular_Disorder I7033 |
| I70331 | DX10 | PVD_ELIX | Peripheral_Vascular_Disorder I70331 |
| I70332 | DX10 | PVD_ELIX | Peripheral_Vascular_Disorder I70332 |
| I70333 | DX10 | PVD_ELIX | Peripheral_Vascular_Disorder I70333 |
| I70334 | DX10 | PVD_ELIX | Peripheral_Vascular_Disorder I70334 |
| I70335 | DX10 | PVD_ELIX | Peripheral_Vascular_Disorder I70335 |
| I70338 | DX10 | PVD_ELIX | Peripheral_Vascular_Disorder I70338 |
| I70339 | DX10 | PVD_ELIX | Peripheral_Vascular_Disorder I70339 |
| I7034 | DX10 | PVD_ELIX | Peripheral_Vascular_Disorder I7034 |
| I70341 | DX10 | PVD_ELIX | Peripheral_Vascular_Disorder I70341 |
| I70342 | DX10 | PVD_ELIX | Peripheral_Vascular_Disorder I70342 |
| I70343 | DX10 | PVD_ELIX | Peripheral_Vascular_Disorder I70343 |
| I70344 | DX10 | PVD_ELIX | Peripheral_Vascular_Disorder I70344 |
| I70345 | DX10 | PVD_ELIX | Peripheral_Vascular_Disorder I70345 |
| I70348 | DX10 | PVD_ELIX | Peripheral_Vascular_Disorder I70348 |
| I70349 | DX10 | PVD_ELIX | Peripheral_Vascular_Disorder I70349 |

| | | | | |
|---|---|---|---|---|
| I7035 | DX10 | PVD_ELIX | Peripheral_Vascular_Disorder | I7035 |
| I7036 | DX10 | PVD_ELIX | Peripheral_Vascular_Disorder | I7036 |
| I70361 | DX10 | PVD_ELIX | Peripheral_Vascular_Disorder | I70361 |
| I70362 | DX10 | PVD_ELIX | Peripheral_Vascular_Disorder | I70362 |
| I70363 | DX10 | PVD_ELIX | Peripheral_Vascular_Disorder | I70363 |
| I70368 | DX10 | PVD_ELIX | Peripheral_Vascular_Disorder | I70368 |
| I70369 | DX10 | PVD_ELIX | Peripheral_Vascular_Disorder | I70369 |
| I7039 | DX10 | PVD_ELIX | Peripheral_Vascular_Disorder | I7039 |
| I70391 | DX10 | PVD_ELIX | Peripheral_Vascular_Disorder | I70391 |
| I70392 | DX10 | PVD_ELIX | Peripheral_Vascular_Disorder | I70392 |
| I70393 | DX10 | PVD_ELIX | Peripheral_Vascular_Disorder | I70393 |
| I70398 | DX10 | PVD_ELIX | Peripheral_Vascular_Disorder | I70398 |
| I70399 | DX10 | PVD_ELIX | Peripheral_Vascular_Disorder | I70399 |
| I704 | DX10 | PVD_ELIX | Peripheral_Vascular_Disorder | I704 |
| I7040 | DX10 | PVD_ELIX | Peripheral_Vascular_Disorder | I7040 |
| I70401 | DX10 | PVD_ELIX | Peripheral_Vascular_Disorder | I70401 |
| I70402 | DX10 | PVD_ELIX | Peripheral_Vascular_Disorder | I70402 |
| I70403 | DX10 | PVD_ELIX | Peripheral_Vascular_Disorder | I70403 |
| I70408 | DX10 | PVD_ELIX | Peripheral_Vascular_Disorder | I70408 |
| I70409 | DX10 | PVD_ELIX | Peripheral_Vascular_Disorder | I70409 |
| I7041 | DX10 | PVD_ELIX | Peripheral_Vascular_Disorder | I7041 |
| I70411 | DX10 | PVD_ELIX | Peripheral_Vascular_Disorder | I70411 |
| I70412 | DX10 | PVD_ELIX | Peripheral_Vascular_Disorder | I70412 |
| I70413 | DX10 | PVD_ELIX | Peripheral_Vascular_Disorder | I70413 |
| I70418 | DX10 | PVD_ELIX | Peripheral_Vascular_Disorder | I70418 |
| I70419 | DX10 | PVD_ELIX | Peripheral_Vascular_Disorder | I70419 |
| I7042 | DX10 | PVD_ELIX | Peripheral_Vascular_Disorder | I7042 |
| I70421 | DX10 | PVD_ELIX | Peripheral_Vascular_Disorder | I70421 |
| I70422 | DX10 | PVD_ELIX | Peripheral_Vascular_Disorder | I70422 |

| | | | | |
|---|---|---|---|---|
| I70423 | DX10 | PVD_ELIX | Peripheral_Vascular_Disorder | I70423 |
| I70428 | DX10 | PVD_ELIX | Peripheral_Vascular_Disorder | I70428 |
| I70429 | DX10 | PVD_ELIX | Peripheral_Vascular_Disorder | I70429 |
| I7043 | DX10 | PVD_ELIX | Peripheral_Vascular_Disorder | I7043 |
| I70431 | DX10 | PVD_ELIX | Peripheral_Vascular_Disorder | I70431 |
| I70432 | DX10 | PVD_ELIX | Peripheral_Vascular_Disorder | I70432 |
| I70433 | DX10 | PVD_ELIX | Peripheral_Vascular_Disorder | I70433 |
| I70434 | DX10 | PVD_ELIX | Peripheral_Vascular_Disorder | I70434 |
| I70435 | DX10 | PVD_ELIX | Peripheral_Vascular_Disorder | I70435 |
| I70438 | DX10 | PVD_ELIX | Peripheral_Vascular_Disorder | I70438 |
| I70439 | DX10 | PVD_ELIX | Peripheral_Vascular_Disorder | I70439 |
| I7044 | DX10 | PVD_ELIX | Peripheral_Vascular_Disorder | I7044 |
| I70441 | DX10 | PVD_ELIX | Peripheral_Vascular_Disorder | I70441 |
| I70442 | DX10 | PVD_ELIX | Peripheral_Vascular_Disorder | I70442 |
| I70443 | DX10 | PVD_ELIX | Peripheral_Vascular_Disorder | I70443 |
| I70444 | DX10 | PVD_ELIX | Peripheral_Vascular_Disorder | I70444 |
| I70445 | DX10 | PVD_ELIX | Peripheral_Vascular_Disorder | I70445 |
| I70448 | DX10 | PVD_ELIX | Peripheral_Vascular_Disorder | I70448 |
| I70449 | DX10 | PVD_ELIX | Peripheral_Vascular_Disorder | I70449 |
| I7045 | DX10 | PVD_ELIX | Peripheral_Vascular_Disorder | I7045 |
| I7046 | DX10 | PVD_ELIX | Peripheral_Vascular_Disorder | I7046 |
| I70461 | DX10 | PVD_ELIX | Peripheral_Vascular_Disorder | I70461 |
| I70462 | DX10 | PVD_ELIX | Peripheral_Vascular_Disorder | I70462 |
| I70463 | DX10 | PVD_ELIX | Peripheral_Vascular_Disorder | I70463 |
| I70468 | DX10 | PVD_ELIX | Peripheral_Vascular_Disorder | I70468 |
| I70469 | DX10 | PVD_ELIX | Peripheral_Vascular_Disorder | I70469 |
| I7049 | DX10 | PVD_ELIX | Peripheral_Vascular_Disorder | I7049 |
| I70491 | DX10 | PVD_ELIX | Peripheral_Vascular_Disorder | I70491 |
| I70492 | DX10 | PVD_ELIX | Peripheral_Vascular_Disorder | I70492 |

| | | | | |
|---|---|---|---|---|
| I70493 | DX10 | PVD_ELIX | Peripheral_Vascular_Disorder | I70493 |
| I70498 | DX10 | PVD_ELIX | Peripheral_Vascular_Disorder | I70498 |
| I70499 | DX10 | PVD_ELIX | Peripheral_Vascular_Disorder | I70499 |
| I705 | DX10 | PVD_ELIX | Peripheral_Vascular_Disorder | I705 |
| I7050 | DX10 | PVD_ELIX | Peripheral_Vascular_Disorder | I7050 |
| I70501 | DX10 | PVD_ELIX | Peripheral_Vascular_Disorder | I70501 |
| I70502 | DX10 | PVD_ELIX | Peripheral_Vascular_Disorder | I70502 |
| I70503 | DX10 | PVD_ELIX | Peripheral_Vascular_Disorder | I70503 |
| I70508 | DX10 | PVD_ELIX | Peripheral_Vascular_Disorder | I70508 |
| I70509 | DX10 | PVD_ELIX | Peripheral_Vascular_Disorder | I70509 |
| I7051 | DX10 | PVD_ELIX | Peripheral_Vascular_Disorder | I7051 |
| I70511 | DX10 | PVD_ELIX | Peripheral_Vascular_Disorder | I70511 |
| I70512 | DX10 | PVD_ELIX | Peripheral_Vascular_Disorder | I70512 |
| I70513 | DX10 | PVD_ELIX | Peripheral_Vascular_Disorder | I70513 |
| I70518 | DX10 | PVD_ELIX | Peripheral_Vascular_Disorder | I70518 |
| I70519 | DX10 | PVD_ELIX | Peripheral_Vascular_Disorder | I70519 |
| I7052 | DX10 | PVD_ELIX | Peripheral_Vascular_Disorder | I7052 |
| I70521 | DX10 | PVD_ELIX | Peripheral_Vascular_Disorder | I70521 |
| I70522 | DX10 | PVD_ELIX | Peripheral_Vascular_Disorder | I70522 |
| I70523 | DX10 | PVD_ELIX | Peripheral_Vascular_Disorder | I70523 |
| I70528 | DX10 | PVD_ELIX | Peripheral_Vascular_Disorder | I70528 |
| I70529 | DX10 | PVD_ELIX | Peripheral_Vascular_Disorder | I70529 |
| I7053 | DX10 | PVD_ELIX | Peripheral_Vascular_Disorder | I7053 |
| I70531 | DX10 | PVD_ELIX | Peripheral_Vascular_Disorder | I70531 |
| I70532 | DX10 | PVD_ELIX | Peripheral_Vascular_Disorder | I70532 |
| I70533 | DX10 | PVD_ELIX | Peripheral_Vascular_Disorder | I70533 |
| I70534 | DX10 | PVD_ELIX | Peripheral_Vascular_Disorder | I70534 |
| I70535 | DX10 | PVD_ELIX | Peripheral_Vascular_Disorder | I70535 |
| I70538 | DX10 | PVD_ELIX | Peripheral_Vascular_Disorder | I70538 |

| | | | | |
|---|---|---|---|---|
| I70539 | DX10 | PVD_ELIX | Peripheral_Vascular_Disorder | I70539 |
| I7054 | DX10 | PVD_ELIX | Peripheral_Vascular_Disorder | I7054 |
| I70541 | DX10 | PVD_ELIX | Peripheral_Vascular_Disorder | I70541 |
| I70542 | DX10 | PVD_ELIX | Peripheral_Vascular_Disorder | I70542 |
| I70543 | DX10 | PVD_ELIX | Peripheral_Vascular_Disorder | I70543 |
| I70544 | DX10 | PVD_ELIX | Peripheral_Vascular_Disorder | I70544 |
| I70545 | DX10 | PVD_ELIX | Peripheral_Vascular_Disorder | I70545 |
| I70548 | DX10 | PVD_ELIX | Peripheral_Vascular_Disorder | I70548 |
| I70549 | DX10 | PVD_ELIX | Peripheral_Vascular_Disorder | I70549 |
| I7055 | DX10 | PVD_ELIX | Peripheral_Vascular_Disorder | I7055 |
| I7056 | DX10 | PVD_ELIX | Peripheral_Vascular_Disorder | I7056 |
| I70561 | DX10 | PVD_ELIX | Peripheral_Vascular_Disorder | I70561 |
| I70562 | DX10 | PVD_ELIX | Peripheral_Vascular_Disorder | I70562 |
| I70563 | DX10 | PVD_ELIX | Peripheral_Vascular_Disorder | I70563 |
| I70568 | DX10 | PVD_ELIX | Peripheral_Vascular_Disorder | I70568 |
| I70569 | DX10 | PVD_ELIX | Peripheral_Vascular_Disorder | I70569 |
| I7059 | DX10 | PVD_ELIX | Peripheral_Vascular_Disorder | I7059 |
| I70591 | DX10 | PVD_ELIX | Peripheral_Vascular_Disorder | I70591 |
| I70592 | DX10 | PVD_ELIX | Peripheral_Vascular_Disorder | I70592 |
| I70593 | DX10 | PVD_ELIX | Peripheral_Vascular_Disorder | I70593 |
| I70598 | DX10 | PVD_ELIX | Peripheral_Vascular_Disorder | I70598 |
| I70599 | DX10 | PVD_ELIX | Peripheral_Vascular_Disorder | I70599 |
| I706 | DX10 | PVD_ELIX | Peripheral_Vascular_Disorder | I706 |
| I7060 | DX10 | PVD_ELIX | Peripheral_Vascular_Disorder | I7060 |
| I70601 | DX10 | PVD_ELIX | Peripheral_Vascular_Disorder | I70601 |
| I70602 | DX10 | PVD_ELIX | Peripheral_Vascular_Disorder | I70602 |
| I70603 | DX10 | PVD_ELIX | Peripheral_Vascular_Disorder | I70603 |
| I70608 | DX10 | PVD_ELIX | Peripheral_Vascular_Disorder | I70608 |
| I70609 | DX10 | PVD_ELIX | Peripheral_Vascular_Disorder | I70609 |

| | | | | |
|---|---|---|---|---|
| I7061 | DX10 | PVD_ELIX | Peripheral_Vascular_Disorder | I7061 |
| I70611 | DX10 | PVD_ELIX | Peripheral_Vascular_Disorder | I70611 |
| I70612 | DX10 | PVD_ELIX | Peripheral_Vascular_Disorder | I70612 |
| I70613 | DX10 | PVD_ELIX | Peripheral_Vascular_Disorder | I70613 |
| I70618 | DX10 | PVD_ELIX | Peripheral_Vascular_Disorder | I70618 |
| I70619 | DX10 | PVD_ELIX | Peripheral_Vascular_Disorder | I70619 |
| I7062 | DX10 | PVD_ELIX | Peripheral_Vascular_Disorder | I7062 |
| I70621 | DX10 | PVD_ELIX | Peripheral_Vascular_Disorder | I70621 |
| I70622 | DX10 | PVD_ELIX | Peripheral_Vascular_Disorder | I70622 |
| I70623 | DX10 | PVD_ELIX | Peripheral_Vascular_Disorder | I70623 |
| I70628 | DX10 | PVD_ELIX | Peripheral_Vascular_Disorder | I70628 |
| I70629 | DX10 | PVD_ELIX | Peripheral_Vascular_Disorder | I70629 |
| I7063 | DX10 | PVD_ELIX | Peripheral_Vascular_Disorder | I7063 |
| I70631 | DX10 | PVD_ELIX | Peripheral_Vascular_Disorder | I70631 |
| I70632 | DX10 | PVD_ELIX | Peripheral_Vascular_Disorder | I70632 |
| I70633 | DX10 | PVD_ELIX | Peripheral_Vascular_Disorder | I70633 |
| I70634 | DX10 | PVD_ELIX | Peripheral_Vascular_Disorder | I70634 |
| I70635 | DX10 | PVD_ELIX | Peripheral_Vascular_Disorder | I70635 |
| I70638 | DX10 | PVD_ELIX | Peripheral_Vascular_Disorder | I70638 |
| I70639 | DX10 | PVD_ELIX | Peripheral_Vascular_Disorder | I70639 |
| I7064 | DX10 | PVD_ELIX | Peripheral_Vascular_Disorder | I7064 |
| I70641 | DX10 | PVD_ELIX | Peripheral_Vascular_Disorder | I70641 |
| I70642 | DX10 | PVD_ELIX | Peripheral_Vascular_Disorder | I70642 |
| I70643 | DX10 | PVD_ELIX | Peripheral_Vascular_Disorder | I70643 |
| I70644 | DX10 | PVD_ELIX | Peripheral_Vascular_Disorder | I70644 |
| I70645 | DX10 | PVD_ELIX | Peripheral_Vascular_Disorder | I70645 |
| I70648 | DX10 | PVD_ELIX | Peripheral_Vascular_Disorder | I70648 |
| I70649 | DX10 | PVD_ELIX | Peripheral_Vascular_Disorder | I70649 |
| I7065 | DX10 | PVD_ELIX | Peripheral_Vascular_Disorder | I7065 |

| | | | | |
|---|---|---|---|---|
| I7066 | DX10 | PVD_ELIX | Peripheral_Vascular_Disorder | I7066 |
| I70661 | DX10 | PVD_ELIX | Peripheral_Vascular_Disorder | I70661 |
| I70662 | DX10 | PVD_ELIX | Peripheral_Vascular_Disorder | I70662 |
| I70663 | DX10 | PVD_ELIX | Peripheral_Vascular_Disorder | I70663 |
| I70668 | DX10 | PVD_ELIX | Peripheral_Vascular_Disorder | I70668 |
| I70669 | DX10 | PVD_ELIX | Peripheral_Vascular_Disorder | I70669 |
| I7069 | DX10 | PVD_ELIX | Peripheral_Vascular_Disorder | I7069 |
| I70691 | DX10 | PVD_ELIX | Peripheral_Vascular_Disorder | I70691 |
| I70692 | DX10 | PVD_ELIX | Peripheral_Vascular_Disorder | I70692 |
| I70693 | DX10 | PVD_ELIX | Peripheral_Vascular_Disorder | I70693 |
| I70698 | DX10 | PVD_ELIX | Peripheral_Vascular_Disorder | I70698 |
| I70699 | DX10 | PVD_ELIX | Peripheral_Vascular_Disorder | I70699 |
| I707 | DX10 | PVD_ELIX | Peripheral_Vascular_Disorder | I707 |
| I7070 | DX10 | PVD_ELIX | Peripheral_Vascular_Disorder | I7070 |
| I70701 | DX10 | PVD_ELIX | Peripheral_Vascular_Disorder | I70701 |
| I70702 | DX10 | PVD_ELIX | Peripheral_Vascular_Disorder | I70702 |
| I70703 | DX10 | PVD_ELIX | Peripheral_Vascular_Disorder | I70703 |
| I70708 | DX10 | PVD_ELIX | Peripheral_Vascular_Disorder | I70708 |
| I70709 | DX10 | PVD_ELIX | Peripheral_Vascular_Disorder | I70709 |
| I7071 | DX10 | PVD_ELIX | Peripheral_Vascular_Disorder | I7071 |
| I70711 | DX10 | PVD_ELIX | Peripheral_Vascular_Disorder | I70711 |
| I70712 | DX10 | PVD_ELIX | Peripheral_Vascular_Disorder | I70712 |
| I70713 | DX10 | PVD_ELIX | Peripheral_Vascular_Disorder | I70713 |
| I70718 | DX10 | PVD_ELIX | Peripheral_Vascular_Disorder | I70718 |
| I70719 | DX10 | PVD_ELIX | Peripheral_Vascular_Disorder | I70719 |
| I7072 | DX10 | PVD_ELIX | Peripheral_Vascular_Disorder | I7072 |
| I70721 | DX10 | PVD_ELIX | Peripheral_Vascular_Disorder | I70721 |
| I70722 | DX10 | PVD_ELIX | Peripheral_Vascular_Disorder | I70722 |
| I70723 | DX10 | PVD_ELIX | Peripheral_Vascular_Disorder | I70723 |

| | | | | |
|---|---|---|---|---|
| I70728 | DX10 | PVD_ELIX | Peripheral_Vascular_ Disorder | I70728 |
| I70729 | DX10 | PVD_ELIX | Peripheral_Vascular_ Disorder | I70729 |
| I7073 | DX10 | PVD_ELIX | Peripheral_Vascular_ Disorder | I7073 |
| I70731 | DX10 | PVD_ELIX | Peripheral_Vascular_ Disorder | I70731 |
| I70732 | DX10 | PVD_ELIX | Peripheral_Vascular_ Disorder | I70732 |
| I70733 | DX10 | PVD_ELIX | Peripheral_Vascular_ Disorder | I70733 |
| I70734 | DX10 | PVD_ELIX | Peripheral_Vascular_ Disorder | I70734 |
| I70735 | DX10 | PVD_ELIX | Peripheral_Vascular_ Disorder | I70735 |
| I70738 | DX10 | PVD_ELIX | Peripheral_Vascular_ Disorder | I70738 |
| I70739 | DX10 | PVD_ELIX | Peripheral_Vascular_ Disorder | I70739 |
| I7074 | DX10 | PVD_ELIX | Peripheral_Vascular_ Disorder | I7074 |
| I70741 | DX10 | PVD_ELIX | Peripheral_Vascular_ Disorder | I70741 |
| I70742 | DX10 | PVD_ELIX | Peripheral_Vascular_ Disorder | I70742 |
| I70743 | DX10 | PVD_ELIX | Peripheral_Vascular_ Disorder | I70743 |
| I70744 | DX10 | PVD_ELIX | Peripheral_Vascular_ Disorder | I70744 |
| I70745 | DX10 | PVD_ELIX | Peripheral_Vascular_ Disorder | I70745 |
| I70748 | DX10 | PVD_ELIX | Peripheral_Vascular_ Disorder | I70748 |
| I70749 | DX10 | PVD_ELIX | Peripheral_Vascular_ Disorder | I70749 |
| I7075 | DX10 | PVD_ELIX | Peripheral_Vascular_ Disorder | I7075 |
| I7076 | DX10 | PVD_ELIX | Peripheral_Vascular_ Disorder | I7076 |
| I70761 | DX10 | PVD_ELIX | Peripheral_Vascular_ Disorder | I70761 |
| I70762 | DX10 | PVD_ELIX | Peripheral_Vascular_ Disorder | I70762 |
| I70763 | DX10 | PVD_ELIX | Peripheral_Vascular_ Disorder | I70763 |
| I70768 | DX10 | PVD_ELIX | Peripheral_Vascular_ Disorder | I70768 |
| I70769 | DX10 | PVD_ELIX | Peripheral_Vascular_ Disorder | I70769 |
| I7079 | DX10 | PVD_ELIX | Peripheral_Vascular_ Disorder | I7079 |
| I70791 | DX10 | PVD_ELIX | Peripheral_Vascular_ Disorder | I70791 |
| I70792 | DX10 | PVD_ELIX | Peripheral_Vascular_ Disorder | I70792 |
| I70793 | DX10 | PVD_ELIX | Peripheral_Vascular_ Disorder | I70793 |

| | | | | |
|---|---|---|---|---|
| I70798 | DX10 | PVD_ELIX | Peripheral_Vascular_Disorder | I70798 |
| I70799 | DX10 | PVD_ELIX | Peripheral_Vascular_Disorder | I70799 |
| I708 | DX10 | PVD_ELIX | Peripheral_Vascular_Disorder | I708 |
| I709 | DX10 | PVD_ELIX | Peripheral_Vascular_Disorder | I709 |
| I7090 | DX10 | PVD_ELIX | Peripheral_Vascular_Disorder | I7090 |
| I7091 | DX10 | PVD_ELIX | Peripheral_Vascular_Disorder | I7091 |
| I7092 | DX10 | PVD_ELIX | Peripheral_Vascular_Disorder | I7092 |
| I71 | DX10 | PVD_ELIX | Peripheral_Vascular_Disorder | I71 |
| I710 | DX10 | PVD_ELIX | Peripheral_Vascular_Disorder | I710 |
| I7100 | DX10 | PVD_ELIX | Peripheral_Vascular_Disorder | I7100 |
| I7101 | DX10 | PVD_ELIX | Peripheral_Vascular_Disorder | I7101 |
| I7102 | DX10 | PVD_ELIX | Peripheral_Vascular_Disorder | I7102 |
| I7103 | DX10 | PVD_ELIX | Peripheral_Vascular_Disorder | I7103 |
| I711 | DX10 | PVD_ELIX | Peripheral_Vascular_Disorder | I711 |
| I712 | DX10 | PVD_ELIX | Peripheral_Vascular_Disorder | I712 |
| I713 | DX10 | PVD_ELIX | Peripheral_Vascular_Disorder | I713 |
| I714 | DX10 | PVD_ELIX | Peripheral_Vascular_Disorder | I714 |
| I715 | DX10 | PVD_ELIX | Peripheral_Vascular_Disorder | I715 |
| I716 | DX10 | PVD_ELIX | Peripheral_Vascular_Disorder | I716 |
| I718 | DX10 | PVD_ELIX | Peripheral_Vascular_Disorder | I718 |
| I719 | DX10 | PVD_ELIX | Peripheral_Vascular_Disorder | I719 |
| I731 | DX10 | PVD_ELIX | Peripheral_Vascular_Disorder | I731 |
| I738 | DX10 | PVD_ELIX | Peripheral_Vascular_Disorder | I738 |
| I7381 | DX10 | PVD_ELIX | Peripheral_Vascular_Disorder | I7381 |
| I7389 | DX10 | PVD_ELIX | Peripheral_Vascular_Disorder | I7389 |
| I739 | DX10 | PVD_ELIX | Peripheral_Vascular_Disorder | I739 |
| I771 | DX10 | PVD_ELIX | Peripheral_Vascular_Disorder | I771 |
| I7771 | DX10 | PVD_ELIX | Peripheral_Vascular_Disorder | I7771 |
| I7772 | DX10 | PVD_ELIX | Peripheral_Vascular_Disorder | I7772 |

| | | | | |
|---|---|---|---|---|
| I7773 | DX10 | PVD_ELIX | Peripheral_Vascular_ Disorder | I7773 |
| I7774 | DX10 | PVD_ELIX | Peripheral_Vascular_ Disorder | I7774 |
| I7779 | DX10 | PVD_ELIX | Peripheral_Vascular_ Disorder | I7779 |
| I79 | DX10 | PVD_ELIX | Peripheral_Vascular_ Disorder | I79 |
| I790 | DX10 | PVD_ELIX | Peripheral_Vascular_ Disorder | I790 |
| I791 | DX10 | PVD_ELIX | Peripheral_Vascular_ Disorder | I791 |
| I798 | DX10 | PVD_ELIX | Peripheral_Vascular_ Disorder | I798 |
| K551 | DX10 | PVD_ELIX | Peripheral_Vascular_ Disorder | K551 |
| K558 | DX10 | PVD_ELIX | Peripheral_Vascular_ Disorder | K558 |
| K559 | DX10 | PVD_ELIX | Peripheral_Vascular_ Disorder | K559 |
| Z9582 | DX10 | PVD_ELIX | Peripheral_Vascular_ Disorder | Z9582 |
| Z95820 | DX10 | PVD_ELIX | Peripheral_Vascular_ Disorder | Z95820 |
| Z95828 | DX10 | PVD_ELIX | Peripheral_Vascular_ Disorder | Z95828 |
| Z959 | DX10 | PVD_ELIX | Peripheral_Vascular_ Disorder | Z959 |
| O00.1 | DX10 | Tubal pregnancy | Pregnancy | O001 |
| O00.10 | DX10 | TUBAL PREGNANCY WITHOUT INTRAUTERINE PREGNANCY | Pregnancy | O0010 |
| O00.101 | DX10 | Right tubal pregnancy without intrauterine pregnancy | Pregnancy | O00101 |
| O00.102 | DX10 | Left tubal pregnancy without intrauterine pregnancy | Pregnancy | O00102 |
| O00.109 | DX10 | Unspecified tubal pregnancy without intrauterine pregnancy | Pregnancy | O00109 |
| O36.4XX0 | DX10 | Maternal care for intrauterine death, not applicable or unsp | Pregnancy | O364XX0 |
| O36.4XX1 | DX10 | Maternal care for intrauterine death, fetus 1 | Pregnancy | O364XX1 |
| O36.4XX2 | DX10 | Maternal care for intrauterine death, fetus 2 | Pregnancy | O364XX2 |
| O36.4XX3 | DX10 | Maternal care for intrauterine death, fetus 3 | Pregnancy | O364XX3 |
| O36.4XX4 | DX10 | Maternal care for intrauterine death, fetus 4 | Pregnancy | O364XX4 |
| O36.4XX5 | DX10 | Maternal care for intrauterine death, fetus 5 | Pregnancy | O364XX5 |
| O36.4XX9 | DX10 | Maternal care for intrauterine death, other fetus | Pregnancy | O364XX9 |
| O75.82 | DX10 | Onset labor 37-39 weeks, w del by (planned) cesarean section | Pregnancy | O7582 |
| O80 | DX10 | Encounter for full-term uncomplicated delivery | Pregnancy | O80 |
| Z37.0 | DX10 | Single live birth | Pregnancy | Z370 |
| Z37.1 | DX10 | Single stillbirth | Pregnancy | Z371 |
| Z37.2 | DX10 | Twins, both liveborn | Pregnancy | Z372 |
| Z37.3 | DX10 | Twins, one liveborn and one stillborn | Pregnancy | Z373 |
| Z37.4 | DX10 | Twins, both stillborn | Pregnancy | Z374 |
| Z37.50 | DX10 | Multiple births, unspecified, all liveborn | Pregnancy | Z3750 |
| Z37.51 | DX10 | Triplets, all liveborn | Pregnancy | Z3751 |
| Z37.52 | DX10 | Quadruplets, all liveborn | Pregnancy | Z3752 |
| Z37.53 | DX10 | Quintuplets, all liveborn | Pregnancy | Z3753 |
| Z37.54 | DX10 | Sextuplets, all liveborn | Pregnancy | Z3754 |
| Z37.59 | DX10 | Other multiple births, all liveborn | Pregnancy | Z3759 |
| Z37.60 | DX10 | Multiple births, unspecified, some liveborn | Pregnancy | Z3760 |
| Z37.61 | DX10 | Triplets, some liveborn | Pregnancy | Z3761 |

| | | | | |
|---|---|---|---|---|
| Z37.62 | DX10 | Quadruplets, some liveborn | Pregnancy | Z3762 |
| Z37.63 | DX10 | Quintuplets, some liveborn | Pregnancy | Z3763 |
| Z37.64 | DX10 | Sextuplets, some liveborn | Pregnancy | Z3764 |
| Z37.69 | DX10 | Other multiple births, some liveborn | Pregnancy | Z3769 |
| Z37.7 | DX10 | Other multiple births, all stillborn | Pregnancy | Z377 |
| Z37.9 | DX10 | Outcome of delivery, unspecified | Pregnancy | Z379 |
| A34 | DX10 | Obstetrical tetanus | Pregnancy | A34 |
| O00.0 | DX10 | Abdominal pregnancy | Pregnancy | O000 |
| O00.00 | DX10 | Abdominal pregnancy without intrauterine pregnancy | Pregnancy | O0000 |
| O00.01 | DX10 | ABDOMINAL PREGNANCY WITH INTRAUTERINE PREGNANCY | Pregnancy | O0001 |
| O00.11 | DX10 | Tubal pregnancy with intrauterine pregnancy | Pregnancy | O0011 |
| O00.111 | DX10 | Right tubal pregnancy with intrauterine pregnancy | Pregnancy | O00111 |
| O00.112 | DX10 | Left tubal pregnancy with intrauterine pregnancy | Pregnancy | O00112 |
| O00.119 | DX10 | Unsp tubal pregnancy with intrauterine pregnancy | Pregnancy | O00119 |
| O00.20 | DX10 | Ovarian pregnancy without intrauterine pregnancy | Pregnancy | O0020 |
| O00.201 | DX10 | Right Ovarian pregnancy without intrauterine pregnancy | Pregnancy | O00201 |
| O00.202 | DX10 | Left Ovarian pregnancy without intrauterine pregnancy | Pregnancy | O00202 |
| O00.209 | DX10 | Unsp Ovarian pregnancy without intrauterine pregnancy | Pregnancy | O00209 |
| O00.21 | DX10 | Ovarian pregnancy with intrauterine pregnancy | Pregnancy | O0021 |
| O00.211 | DX10 | Ovarian pregnancy with intrauterine pregnancy | Pregnancy | O00211 |
| O00.212 | DX10 | Ovarian pregnancy with intrauterine pregnancy | Pregnancy | O00212 |
| O00.219 | DX10 | Ovarian pregnancy with intrauterine pregnancy | Pregnancy | O00219 |
| O00.8 | DX10 | Other ectopic pregnancy | Pregnancy | O008 |
| O00.80 | DX10 | OTHER ECTOPIC PREGNANCY W/O INTRAUTERINE PREG | Pregnancy | O0080 |
| O00.81 | DX10 | OTHER ECTOPIC PREGNANCY W/INTRAUTERINE PREGNANCY | Pregnancy | O0081 |
| O00.9 | DX10 | Ectopic pregnancy, unspecified | Pregnancy | O009 |
| O00.90 | DX10 | UNSPEC ECTOPIC PREGNANCY W/O INTRAUTERINE PREG | Pregnancy | O0090 |
| O00.91 | DX10 | Unspecified ectopic pregnancy with intrauterine pregnancy | Pregnancy | O0091 |
| O01.0 | DX10 | Classical hydatidiform mole | Pregnancy | O010 |
| O01.1 | DX10 | Incomplete and partial hydatidiform mole | Pregnancy | O011 |
| O01.9 | DX10 | Hydatidiform mole, unspecified | Pregnancy | O019 |
| O02.0 | DX10 | Blighted ovum and nonhydatidiform mole | Pregnancy | O020 |
| O02.1 | DX10 | Missed abortion | Pregnancy | O021 |
| O03.0 | DX10 | Genitl trct and pelvic infection fol incmpl spon abortion | Pregnancy | O030 |
| O03.1 | DX10 | Delayed or excessive hemor following incmpl spon abortion | Pregnancy | O031 |
| O03.2 | DX10 | Embolism following incomplete spontaneous abortion | Pregnancy | O032 |
| O03.30 | DX10 | Unsp complication following incomplete spontaneous abortion | Pregnancy | O0330 |
| O03.31 | DX10 | Shock following incomplete spontaneous abortion | Pregnancy | O0331 |
| O03.32 | DX10 | Renal failure following incomplete spontaneous abortion | Pregnancy | O0332 |
| O03.33 | DX10 | Metabolic disorder following incomplete spontaneous abortion | Pregnancy | O0333 |
| O03.34 | DX10 | Damage to pelvic organs following incomplete spon abortion | Pregnancy | O0334 |
| O03.35 | DX10 | Oth venous comp following incomplete spontaneous abortion | Pregnancy | O0335 |
| O03.36 | DX10 | Cardiac arrest following incomplete spontaneous abortion | Pregnancy | O0336 |
| O03.37 | DX10 | Sepsis following incomplete spontaneous abortion | Pregnancy | O0337 |
| O03.38 | DX10 | Urinary tract infection following incomplete spon abortion | Pregnancy | O0338 |

| | | | | |
|---|---|---|---|---|
| O03.39 | DX10 | Incomplete spontaneous abortion with other complications | Pregnancy | O0339 |
| O03.4 | DX10 | Incomplete spontaneous abortion without complication | Pregnancy | O034 |
| O03.5 | DX10 | Genitl trct and pelvic infct fol complete or unsp spon abort | Pregnancy | O035 |
| O03.6 | DX10 | Delayed or excess hemor fol complete or unsp spon abortion | Pregnancy | O036 |
| O03.7 | DX10 | Embolism following complete or unsp spontaneous abortion | Pregnancy | O037 |
| O03.80 | DX10 | Unsp comp following complete or unsp spontaneous abortion | Pregnancy | O0380 |
| O03.81 | DX10 | Shock following complete or unspecified spontaneous abortion | Pregnancy | O0381 |
| O03.82 | DX10 | Renal failure following complete or unsp spon abortion | Pregnancy | O0382 |
| O03.83 | DX10 | Metabolic disorder following complete or unsp spon abortion | Pregnancy | O0383 |
| O03.84 | DX10 | Damage to pelvic organs fol complete or unsp spon abortion | Pregnancy | O0384 |
| O03.85 | DX10 | Oth venous comp following complete or unsp spon abortion | Pregnancy | O0385 |
| O03.86 | DX10 | Cardiac arrest following complete or unsp spon abortion | Pregnancy | O0386 |
| O03.87 | DX10 | Sepsis following complete or unsp spontaneous abortion | Pregnancy | O0387 |
| O03.88 | DX10 | Urinary tract infection fol complete or unsp spon abortion | Pregnancy | O0388 |
| O03.89 | DX10 | Complete or unsp spontaneous abortion with oth complications | Pregnancy | O0389 |
| O03.9 | DX10 | Complete or unsp spontaneous abortion without complication | Pregnancy | O039 |
| O04.5 | DX10 | Genitl trct and pelvic infct fol (induced) term of pregnancy | Pregnancy | O045 |
| O04.6 | DX10 | Delayed or excess hemor fol (induced) term of pregnancy | Pregnancy | O046 |
| O04.7 | DX10 | Embolism following (induced) termination of pregnancy | Pregnancy | O047 |
| O04.80 | DX10 | (Induced) termination of pregnancy with unsp complications | Pregnancy | O0480 |
| O04.81 | DX10 | Shock following (induced) termination of pregnancy | Pregnancy | O0481 |
| O04.82 | DX10 | Renal failure following (induced) termination of pregnancy | Pregnancy | O0482 |
| O04.83 | DX10 | Metabolic disorder following (induced) term of pregnancy | Pregnancy | O0483 |
| O04.84 | DX10 | Damage to pelvic organs fol (induced) term of pregnancy | Pregnancy | O0484 |
| O04.85 | DX10 | Oth venous comp following (induced) termination of pregnancy | Pregnancy | O0485 |
| O04.86 | DX10 | Cardiac arrest following (induced) termination of pregnancy | Pregnancy | O0486 |
| O04.87 | DX10 | Sepsis following (induced) termination of pregnancy | Pregnancy | O0487 |
| O04.88 | DX10 | Urinary tract infection fol (induced) term of pregnancy | Pregnancy | O0488 |
| O04.89 | DX10 | (Induced) termination of pregnancy with other complications | Pregnancy | O0489 |
| O07.0 | DX10 | Genitl trct and pelvic infct fol failed attempt term of preg | Pregnancy | O070 |
| O07.1 | DX10 | Delayed or excess hemor fol failed attempt term of pregnancy | Pregnancy | O071 |
| O07.2 | DX10 | Embolism following failed attempted termination of pregnancy | Pregnancy | O072 |

| | | | | |
|---|---|---|---|---|
| O07.30 | DX10 | Failed attempted termination of pregnancy w unsp comp | Pregnancy | O0730 |
| O07.31 | DX10 | Shock following failed attempted termination of pregnancy | Pregnancy | O0731 |
| O07.32 | DX10 | Renal failure following failed attempted term of pregnancy | Pregnancy | O0732 |
| O07.33 | DX10 | Metabolic disorder fol failed attempt term of pregnancy | Pregnancy | O0733 |
| O07.34 | DX10 | Damage to pelvic organs fol failed attempt term of pregnancy | Pregnancy | O0734 |
| O07.35 | DX10 | Oth venous comp following failed attempted term of pregnancy | Pregnancy | O0735 |
| O07.36 | DX10 | Cardiac arrest following failed attempted term of pregnancy | Pregnancy | O0736 |
| O07.37 | DX10 | Sepsis following failed attempted termination of pregnancy | Pregnancy | O0737 |
| O07.38 | DX10 | Urinary tract infection fol failed attempt term of pregnancy | Pregnancy | O0738 |
| O07.39 | DX10 | Failed attempted termination of pregnancy w oth comp | Pregnancy | O0739 |
| O07.4 | DX10 | Failed attempted termination of pregnancy w/o complication | Pregnancy | O074 |
| O08.0 | DX10 | Genitl trct and pelvic infct fol ectopic and molar pregnancy | Pregnancy | O080 |
| O08.1 | DX10 | Delayed or excess hemor fol ectopic and molar pregnancy | Pregnancy | O081 |
| O08.2 | DX10 | Embolism following ectopic and molar pregnancy | Pregnancy | O082 |
| O08.3 | DX10 | Shock following ectopic and molar pregnancy | Pregnancy | O083 |
| O08.4 | DX10 | Renal failure following ectopic and molar pregnancy | Pregnancy | O084 |
| O08.5 | DX10 | Metabolic disorders following an ectopic and molar pregnancy | Pregnancy | O085 |
| O08.6 | DX10 | Damage to pelvic organs and tiss fol an ect and molar preg | Pregnancy | O086 |
| O08.7 | DX10 | Oth venous comp following an ectopic and molar pregnancy | Pregnancy | O087 |
| O08.81 | DX10 | Cardiac arrest following an ectopic and molar pregnancy | Pregnancy | O0881 |
| O08.82 | DX10 | Sepsis following ectopic and molar pregnancy | Pregnancy | O0882 |
| O08.83 | DX10 | Urinary tract infection fol an ectopic and molar pregnancy | Pregnancy | O0883 |
| O08.89 | DX10 | Other complications following an ectopic and molar pregnancy | Pregnancy | O0889 |
| O08.9 | DX10 | Unsp complication following an ectopic and molar pregnancy | Pregnancy | O089 |
| O09.00 | DX10 | Suprvsn of preg w history of infertility, unsp trimester | Pregnancy | O0900 |
| O09.01 | DX10 | Suprvsn of preg w history of infertility, first trimester | Pregnancy | O0901 |
| O09.02 | DX10 | Suprvsn of preg w history of infertility, second trimester | Pregnancy | O0902 |
| O09.03 | DX10 | Suprvsn of preg w history of infertility, third trimester | Pregnancy | O0903 |
| O09.10 | DX10 | Suprvsn of preg w history of ectopic or molar preg, unsp tri | Pregnancy | O0910 |
| O09.11 | DX10 | Suprvsn of preg w history of ect or molar preg, first tri | Pregnancy | O0911 |
| O09.12 | DX10 | Suprvsn of preg w history of ect or molar preg, second tri | Pregnancy | O0912 |
| O09.13 | DX10 | Suprvsn of preg w history of ect or molar preg, third tri | Pregnancy | O0913 |
| O09.211 | DX10 | Suprvsn of preg w history of pre-term labor, first trimester | Pregnancy | O09211 |
| O09.212 | DX10 | Suprvsn of preg w history of pre-term labor, second tri | Pregnancy | O09212 |
| O09.213 | DX10 | Suprvsn of preg w history of pre-term labor, third trimester | Pregnancy | O09213 |
| O09.219 | DX10 | Suprvsn of preg w history of pre-term labor, unsp trimester | Pregnancy | O09219 |
| O09.291 | DX10 | Suprvsn of preg w poor reprodctv or obstet hx, first tri | Pregnancy | O09291 |

| | | | | |
|---|---|---|---|---|
| O09.292 | DX10 | Suprvsn of preg w poor reprodctv or obstet hx, second tri | Pregnancy | O09292 |
| O09.293 | DX10 | Suprvsn of preg w poor reprodctv or obstet hx, third tri | Pregnancy | O09293 |
| O09.299 | DX10 | Suprvsn of preg w poor reprodctv or obstet history, unsp tri | Pregnancy | O09299 |
| O09.30 | DX10 | Suprvsn of preg w insufficient antenat care, unsp trimester | Pregnancy | O0930 |
| O09.31 | DX10 | Suprvsn of preg w insufficient antenat care, first trimester | Pregnancy | O0931 |
| O09.32 | DX10 | Suprvsn of preg w insufficient antenat care, second tri | Pregnancy | O0932 |
| O09.33 | DX10 | Suprvsn of preg w insufficient antenat care, third trimester | Pregnancy | O0933 |
| O09.40 | DX10 | Supervision of pregnancy w grand multiparity, unsp trimester | Pregnancy | O0940 |
| O09.41 | DX10 | Suprvsn of pregnancy w grand multiparity, first trimester | Pregnancy | O0941 |
| O09.42 | DX10 | Suprvsn of pregnancy w grand multiparity, second trimester | Pregnancy | O0942 |
| O09.43 | DX10 | Suprvsn of pregnancy w grand multiparity, third trimester | Pregnancy | O0943 |
| O09.511 | DX10 | Supervision of elderly primigravida, first trimester | Pregnancy | O09511 |
| O09.512 | DX10 | Supervision of elderly primigravida, second trimester | Pregnancy | O09512 |
| O09.513 | DX10 | Supervision of elderly primigravida, third trimester | Pregnancy | O09513 |
| O09.519 | DX10 | Supervision of elderly primigravida, unspecified trimester | Pregnancy | O09519 |
| O09.521 | DX10 | Supervision of elderly multigravida, first trimester | Pregnancy | O09521 |
| O09.522 | DX10 | Supervision of elderly multigravida, second trimester | Pregnancy | O09522 |
| O09.523 | DX10 | Supervision of elderly multigravida, third trimester | Pregnancy | O09523 |
| O09.529 | DX10 | Supervision of elderly multigravida, unspecified trimester | Pregnancy | O09529 |
| O09.611 | DX10 | Supervision of young primigravida, first trimester | Pregnancy | O09611 |
| O09.612 | DX10 | Supervision of young primigravida, second trimester | Pregnancy | O09612 |
| O09.613 | DX10 | Supervision of young primigravida, third trimester | Pregnancy | O09613 |
| O09.619 | DX10 | Supervision of young primigravida, unspecified trimester | Pregnancy | O09619 |
| O09.621 | DX10 | Supervision of young multigravida, first trimester | Pregnancy | O09621 |
| O09.622 | DX10 | Supervision of young multigravida, second trimester | Pregnancy | O09622 |
| O09.623 | DX10 | Supervision of young multigravida, third trimester | Pregnancy | O09623 |
| O09.629 | DX10 | Supervision of young multigravida, unspecified trimester | Pregnancy | O09629 |
| O09.70 | DX10 | Suprvsn of high risk preg due to social problems, unsp tri | Pregnancy | O0970 |
| O09.71 | DX10 | Suprvsn of high risk preg due to social problems, first tri | Pregnancy | O0971 |
| O09.72 | DX10 | Suprvsn of high risk preg due to social problems, second tri | Pregnancy | O0972 |
| O09.73 | DX10 | Suprvsn of high risk preg due to social problems, third tri | Pregnancy | O0973 |
| O09.811 | DX10 | Suprvsn of preg rslt from assisted reprodctv tech, first tri | Pregnancy | O09811 |
| O09.812 | DX10 | Suprvsn of preg rslt from assist reprodctv tech, second tri | Pregnancy | O09812 |
| O09.813 | DX10 | Suprvsn of preg rslt from assisted reprodctv tech, third tri | Pregnancy | O09813 |
| O09.819 | DX10 | Suprvsn of preg rslt from assisted reprodctv tech, unsp tri | Pregnancy | O09819 |
| O09.821 | DX10 | Suprvsn of preg w hx of in utero proc dur prev preg, 1st tri | Pregnancy | O09821 |
| O09.822 | DX10 | Suprvsn of preg w hx of in utero proc dur prev preg, 2nd tri | Pregnancy | O09822 |
| O09.823 | DX10 | Suprvsn of preg w hx of in utero proc dur prev preg, 3rd tri | Pregnancy | O09823 |
| O09.829 | DX10 | Suprvsn of preg w hx of in utero proc dur prev preg,unsp tri | Pregnancy | O09829 |

| | | | | |
|---|---|---|---|---|
| O09.891 | DX10 | Supervision of other high risk pregnancies, first trimester | Pregnancy | O09891 |
| O09.892 | DX10 | Supervision of other high risk pregnancies, second trimester | Pregnancy | O09892 |
| O09.893 | DX10 | Supervision of other high risk pregnancies, third trimester | Pregnancy | O09893 |
| O09.899 | DX10 | Supervision of other high risk pregnancies, unsp trimester | Pregnancy | O09899 |
| O09.90 | DX10 | Supervision of high risk pregnancy, unsp, unsp trimester | Pregnancy | O0990 |
| O09.91 | DX10 | Supervision of high risk pregnancy, unsp, first trimester | Pregnancy | O0991 |
| O09.92 | DX10 | Supervision of high risk pregnancy, unsp, second trimester | Pregnancy | O0992 |
| O09.93 | DX10 | Supervision of high risk pregnancy, unsp, third trimester | Pregnancy | O0993 |
| O09.A0 | DX10 | Supervision of pregnancy with history of molar pregnancy, unspecified trimester | Pregnancy | O09A0 |
| O09.A1 | DX10 | Supervision of pregnancy with history of molar pregnancy, first trimester | Pregnancy | O09A1 |
| O09.A2 | DX10 | Supervision of pregnancy with history of molar pregnancy, second trimester | Pregnancy | O09A2 |
| O09.A3 | DX10 | Supervision of pregnancy with history of molar pregnancy, third trimester | Pregnancy | O09A3 |
| O10.011 | DX10 | Pre-existing essential htn comp pregnancy, first trimester | Pregnancy | O10011 |
| O10.012 | DX10 | Pre-existing essential htn comp pregnancy, second trimester | Pregnancy | O10012 |
| O10.013 | DX10 | Pre-existing essential htn comp pregnancy, third trimester | Pregnancy | O10013 |
| O10.019 | DX10 | Pre-existing essential htn comp pregnancy, unsp trimester | Pregnancy | O10019 |
| O10.02 | DX10 | Pre-existing essential hypertension complicating childbirth | Pregnancy | O1002 |
| O10.03 | DX10 | Pre-existing essential hypertension comp the puerperium | Pregnancy | O1003 |
| O10.111 | DX10 | Pre-exist hyp heart disease comp pregnancy, first trimester | Pregnancy | O10111 |
| O10.112 | DX10 | Pre-exist hyp heart disease comp pregnancy, second trimester | Pregnancy | O10112 |
| O10.113 | DX10 | Pre-exist hyp heart disease comp pregnancy, third trimester | Pregnancy | O10113 |
| O10.119 | DX10 | Pre-exist hyp heart disease comp pregnancy, unsp trimester | Pregnancy | O10119 |
| O10.12 | DX10 | Pre-existing hypertensive heart disease comp childbirth | Pregnancy | O1012 |
| O10.13 | DX10 | Pre-existing hypertensive heart disease comp the puerperium | Pregnancy | O1013 |
| O10.211 | DX10 | Pre-exist hyp chronic kidney disease comp preg, first tri | Pregnancy | O10211 |
| O10.212 | DX10 | Pre-exist hyp chronic kidney disease comp preg, second tri | Pregnancy | O10212 |
| O10.213 | DX10 | Pre-exist hyp chronic kidney disease comp preg, third tri | Pregnancy | O10213 |
| O10.219 | DX10 | Pre-exist hyp chronic kidney disease comp preg, unsp tri | Pregnancy | O10219 |
| O10.22 | DX10 | Pre-existing hyp chronic kidney disease comp childbirth | Pregnancy | O1022 |
| O10.23 | DX10 | Pre-existing hyp chronic kidney disease comp the puerperium | Pregnancy | O1023 |
| O10.311 | DX10 | Pre-exist hyp heart and chr kidney dis comp preg, first tri | Pregnancy | O10311 |
| O10.312 | DX10 | Pre-exist hyp heart and chr kidney dis comp preg, second tri | Pregnancy | O10312 |
| O10.313 | DX10 | Pre-exist hyp heart and chr kidney dis comp preg, third tri | Pregnancy | O10313 |
| O10.319 | DX10 | Pre-exist hyp heart and chr kidney dis comp preg, unsp tri | Pregnancy | O10319 |
| O10.32 | DX10 | Pre-exist hyp heart and chronic kidney disease comp chldbrth | Pregnancy | O1032 |

| | | | | |
|---|---|---|---|---|
| O10.33 | DX10 | Pre-exist hyp heart and chr kidney disease comp the puerp | Pregnancy | O1033 |
| O10.411 | DX10 | Pre-existing secondary htn comp pregnancy, first trimester | Pregnancy | O10411 |
| O10.412 | DX10 | Pre-existing secondary htn comp pregnancy, second trimester | Pregnancy | O10412 |
| O10.413 | DX10 | Pre-existing secondary htn comp pregnancy, third trimester | Pregnancy | O10413 |
| O10.419 | DX10 | Pre-existing secondary htn comp pregnancy, unsp trimester | Pregnancy | O10419 |
| O10.42 | DX10 | Pre-existing secondary hypertension complicating childbirth | Pregnancy | O1042 |
| O10.43 | DX10 | Pre-existing secondary hypertension comp the puerperium | Pregnancy | O1043 |
| O10.911 | DX10 | Unsp pre-existing htn comp pregnancy, first trimester | Pregnancy | O10911 |
| O10.912 | DX10 | Unsp pre-existing htn comp pregnancy, second trimester | Pregnancy | O10912 |
| O10.913 | DX10 | Unsp pre-existing htn comp pregnancy, third trimester | Pregnancy | O10913 |
| O10.919 | DX10 | Unsp pre-existing htn comp pregnancy, unsp trimester | Pregnancy | O10919 |
| O10.92 | DX10 | Unsp pre-existing hypertension complicating childbirth | Pregnancy | O1092 |
| O10.93 | DX10 | Unsp pre-existing hypertension complicating the puerperium | Pregnancy | O1093 |
| O11.1 | DX10 | Pre-existing hypertension w pre-eclampsia, first trimester | Pregnancy | O111 |
| O11.2 | DX10 | Pre-existing hypertension w pre-eclampsia, second trimester | Pregnancy | O112 |
| O11.3 | DX10 | Pre-existing hypertension w pre-eclampsia, third trimester | Pregnancy | O113 |
| O11.4 | DX10 | Pre-existing hypertension with pre-eclampsia, complicating childbirth | Pregnancy | O114 |
| O11.5 | DX10 | Pre-existing hypertension with pre-eclampsia, complicating the puerperium | Pregnancy | O115 |
| O11.9 | DX10 | Pre-existing hypertension with pre-eclampsia, unsp trimester | Pregnancy | O119 |
| O12.00 | DX10 | Gestational edema, unspecified trimester | Pregnancy | O1200 |
| O12.01 | DX10 | Gestational edema, first trimester | Pregnancy | O1201 |
| O12.02 | DX10 | Gestational edema, second trimester | Pregnancy | O1202 |
| O12.03 | DX10 | Gestational edema, third trimester | Pregnancy | O1203 |
| O12.04 | DX10 | Gestational edema, complicating childbirth | Pregnancy | O1204 |
| O12.05 | DX10 | Gestational edema, complicating the puerperium | Pregnancy | O1205 |
| O12.10 | DX10 | Gestational proteinuria, unspecified trimester | Pregnancy | O1210 |
| O12.11 | DX10 | Gestational proteinuria, first trimester | Pregnancy | O1211 |
| O12.12 | DX10 | Gestational proteinuria, second trimester | Pregnancy | O1212 |
| O12.13 | DX10 | Gestational proteinuria, third trimester | Pregnancy | O1213 |
| O12.14 | DX10 | Gestational proteinuria, complicating childbirth | Pregnancy | O1214 |
| O12.15 | DX10 | Gestational proteinuria, complicating the puerperium | Pregnancy | O1215 |
| O12.20 | DX10 | Gestational edema with proteinuria, unspecified trimester | Pregnancy | O1220 |
| O12.21 | DX10 | Gestational edema with proteinuria, first trimester | Pregnancy | O1221 |
| O12.22 | DX10 | Gestational edema with proteinuria, second trimester | Pregnancy | O1222 |
| O12.23 | DX10 | Gestational edema with proteinuria, third trimester | Pregnancy | O1223 |
| O12.24 | DX10 | Gestational edema with proteinuria, complicating childbirth | Pregnancy | O1224 |
| O12.25 | DX10 | Gestational edema with proteinuria, complicating the puerperium | Pregnancy | O1225 |
| O13.1 | DX10 | Gestational htn w/o significant proteinuria, first trimester | Pregnancy | O131 |
| O13.2 | DX10 | Gestatnl htn w/o significant proteinuria, second trimester | Pregnancy | O132 |
| O13.3 | DX10 | Gestational htn w/o significant proteinuria, third trimester | Pregnancy | O133 |

| | | | | |
|---|---|---|---|---|
| O13.4 | DX10 | GESTATIONAL HTN W/O SIGNIF PROTEINURIA COMP CB | Pregnancy | O134 |
| O13.5 | DX10 | Gestational [pregnancy-induced] hypertension without significant proteinuria, complicating the puerperium | Pregnancy | O135 |
| O13.9 | DX10 | Gestational htn w/o significant proteinuria, unsp trimester | Pregnancy | O139 |
| O14.00 | DX10 | Mild to moderate pre-eclampsia, unspecified trimester | Pregnancy | O1400 |
| O14.02 | DX10 | Mild to moderate pre-eclampsia, second trimester | Pregnancy | O1402 |
| O14.03 | DX10 | Mild to moderate pre-eclampsia, third trimester | Pregnancy | O1403 |
| O14.04 | DX10 | Mild to moderate pre-eclampsia, complicating childbirth | Pregnancy | O1404 |
| O14.05 | DX10 | Mild to moderate pre-eclampsia, complicating the puerperium | Pregnancy | O1405 |
| O14.10 | DX10 | Severe pre-eclampsia, unspecified trimester | Pregnancy | O1410 |
| O14.12 | DX10 | Severe pre-eclampsia, second trimester | Pregnancy | O1412 |
| O14.13 | DX10 | Severe pre-eclampsia, third trimester | Pregnancy | O1413 |
| O14.14 | DX10 | Severe pre-eclampsia complicating childbirth | Pregnancy | O1414 |
| O14.20 | DX10 | HELLP syndrome (HELLP), unspecified trimester | Pregnancy | O1420 |
| O14.22 | DX10 | HELLP syndrome (HELLP), second trimester | Pregnancy | O1422 |
| O14.23 | DX10 | HELLP syndrome (HELLP), third trimester | Pregnancy | O1423 |
| O14.24 | DX10 | HELLP syndrome, complicating childbirth | Pregnancy | O1424 |
| O14.90 | DX10 | Unspecified pre-eclampsia, unspecified trimester | Pregnancy | O1490 |
| O14.92 | DX10 | Unspecified pre-eclampsia, second trimester | Pregnancy | O1492 |
| O14.93 | DX10 | Unspecified pre-eclampsia, third trimester | Pregnancy | O1493 |
| O14.94 | DX10 | UNSPECIFIED PRE-ECLAMPSIA COMP CHILDBIRTH | Pregnancy | O1494 |
| O14.95 | DX10 | UNSPECIFIED PRE-ECLAMPSIA COMP THE PUERPERIUM | Pregnancy | O1495 |
| O1415 | DX10 | Severe pre-eclampsia, complicating the puerperium | Pregnancy | O1415 |
| O1425 | DX10 | HELLP syndrome, complicating the puerperium | Pregnancy | O1425 |
| O15.00 | DX10 | Eclampsia in pregnancy, unspecified trimester | Pregnancy | O1500 |
| O15.02 | DX10 | Eclampsia in pregnancy, second trimester | Pregnancy | O1502 |
| O15.03 | DX10 | Eclampsia in pregnancy, third trimester | Pregnancy | O1503 |
| O15.1 | DX10 | Eclampsia in labor | Pregnancy | O151 |
| O15.2 | DX10 | ECLAMPSIA IN THE PUERPERIUM | Pregnancy | O152 |
| O15.9 | DX10 | Eclampsia, unspecified as to time period | Pregnancy | O159 |
| O16.1 | DX10 | Unspecified maternal hypertension, first trimester | Pregnancy | O161 |
| O16.2 | DX10 | Unspecified maternal hypertension, second trimester | Pregnancy | O162 |
| O16.3 | DX10 | Unspecified maternal hypertension, third trimester | Pregnancy | O163 |
| O16.4 | DX10 | Unspecified maternal hypertension, complicating childbirth | Pregnancy | O164 |
| O16.5 | DX10 | UNSPECIFIED MATERNAL HTN COMPLICATING PUERPERIUM | Pregnancy | O165 |
| O16.9 | DX10 | Unspecified maternal hypertension, unspecified trimester | Pregnancy | O169 |
| O20.0 | DX10 | Threatened abortion | Pregnancy | O200 |
| O20.8 | DX10 | Other hemorrhage in early pregnancy | Pregnancy | O208 |
| O20.9 | DX10 | Hemorrhage in early pregnancy, unspecified | Pregnancy | O209 |
| O21.0 | DX10 | Mild hyperemesis gravidarum | Pregnancy | O210 |
| O21.1 | DX10 | Hyperemesis gravidarum with metabolic disturbance | Pregnancy | O211 |
| O21.2 | DX10 | Late vomiting of pregnancy | Pregnancy | O212 |
| O21.8 | DX10 | Other vomiting complicating pregnancy | Pregnancy | O218 |
| O21.9 | DX10 | Vomiting of pregnancy, unspecified | Pregnancy | O219 |
| O22.00 | DX10 | Varicose veins of low extrm in pregnancy, unsp trimester | Pregnancy | O2200 |
| O22.01 | DX10 | Varicose veins of low extrm in pregnancy, first trimester | Pregnancy | O2201 |
| O22.02 | DX10 | Varicose veins of low extrm in pregnancy, second trimester | Pregnancy | O2202 |
| O22.03 | DX10 | Varicose veins of low extrm in pregnancy, third trimester | Pregnancy | O2203 |
| O22.10 | DX10 | Genital varices in pregnancy, unspecified trimester | Pregnancy | O2210 |
| O22.11 | DX10 | Genital varices in pregnancy, first trimester | Pregnancy | O2211 |
| O22.12 | DX10 | Genital varices in pregnancy, second trimester | Pregnancy | O2212 |

| | | | | |
|---|---|---|---|---|
| O22.13 | DX10 | Genital varices in pregnancy, third trimester | Pregnancy | O2213 |
| O22.20 | DX10 | Superficial thrombophlebitis in pregnancy, unsp trimester | Pregnancy | O2220 |
| O22.21 | DX10 | Superficial thrombophlebitis in pregnancy, first trimester | Pregnancy | O2221 |
| O22.22 | DX10 | Superficial thrombophlebitis in pregnancy, second trimester | Pregnancy | O2222 |
| O22.23 | DX10 | Superficial thrombophlebitis in pregnancy, third trimester | Pregnancy | O2223 |
| O22.30 | DX10 | Deep phlebothrombosis in pregnancy, unspecified trimester | Pregnancy | O2230 |
| O22.31 | DX10 | Deep phlebothrombosis in pregnancy, first trimester | Pregnancy | O2231 |
| O22.32 | DX10 | Deep phlebothrombosis in pregnancy, second trimester | Pregnancy | O2232 |
| O22.33 | DX10 | Deep phlebothrombosis in pregnancy, third trimester | Pregnancy | O2233 |
| O22.40 | DX10 | Hemorrhoids in pregnancy, unspecified trimester | Pregnancy | O2240 |
| O22.41 | DX10 | Hemorrhoids in pregnancy, first trimester | Pregnancy | O2241 |
| O22.42 | DX10 | Hemorrhoids in pregnancy, second trimester | Pregnancy | O2242 |
| O22.43 | DX10 | Hemorrhoids in pregnancy, third trimester | Pregnancy | O2243 |
| O22.50 | DX10 | Cerebral venous thrombosis in pregnancy, unsp trimester | Pregnancy | O2250 |
| O22.51 | DX10 | Cerebral venous thrombosis in pregnancy, first trimester | Pregnancy | O2251 |
| O22.52 | DX10 | Cerebral venous thrombosis in pregnancy, second trimester | Pregnancy | O2252 |
| O22.53 | DX10 | Cerebral venous thrombosis in pregnancy, third trimester | Pregnancy | O2253 |
| O22.8X1 | DX10 | Other venous complications in pregnancy, first trimester | Pregnancy | O228X1 |
| O22.8X2 | DX10 | Other venous complications in pregnancy, second trimester | Pregnancy | O228X2 |
| O22.8X3 | DX10 | Other venous complications in pregnancy, third trimester | Pregnancy | O228X3 |
| O22.8X9 | DX10 | Other venous complications in pregnancy, unsp trimester | Pregnancy | O228X9 |
| O22.90 | DX10 | Venous complication in pregnancy, unsp, unsp trimester | Pregnancy | O2290 |
| O22.91 | DX10 | Venous complication in pregnancy, unsp, first trimester | Pregnancy | O2291 |
| O22.92 | DX10 | Venous complication in pregnancy, unsp, second trimester | Pregnancy | O2292 |
| O22.93 | DX10 | Venous complication in pregnancy, unsp, third trimester | Pregnancy | O2293 |
| O23.00 | DX10 | Infections of kidney in pregnancy, unspecified trimester | Pregnancy | O2300 |
| O23.01 | DX10 | Infections of kidney in pregnancy, first trimester | Pregnancy | O2301 |
| O23.02 | DX10 | Infections of kidney in pregnancy, second trimester | Pregnancy | O2302 |
| O23.03 | DX10 | Infections of kidney in pregnancy, third trimester | Pregnancy | O2303 |
| O23.10 | DX10 | Infections of bladder in pregnancy, unspecified trimester | Pregnancy | O2310 |
| O23.11 | DX10 | Infections of bladder in pregnancy, first trimester | Pregnancy | O2311 |
| O23.12 | DX10 | Infections of bladder in pregnancy, second trimester | Pregnancy | O2312 |
| O23.13 | DX10 | Infections of bladder in pregnancy, third trimester | Pregnancy | O2313 |
| O23.20 | DX10 | Infections of urethra in pregnancy, unspecified trimester | Pregnancy | O2320 |
| O23.21 | DX10 | Infections of urethra in pregnancy, first trimester | Pregnancy | O2321 |
| O23.22 | DX10 | Infections of urethra in pregnancy, second trimester | Pregnancy | O2322 |
| O23.23 | DX10 | Infections of urethra in pregnancy, third trimester | Pregnancy | O2323 |
| O23.30 | DX10 | Infections of prt urinary tract in pregnancy, unsp trimester | Pregnancy | O2330 |
| O23.31 | DX10 | Infect of prt urinary tract in pregnancy, first trimester | Pregnancy | O2331 |
| O23.32 | DX10 | Infect of prt urinary tract in pregnancy, second trimester | Pregnancy | O2332 |
| O23.33 | DX10 | Infect of prt urinary tract in pregnancy, third trimester | Pregnancy | O2333 |
| O23.40 | DX10 | Unsp infection of urinary tract in pregnancy, unsp trimester | Pregnancy | O2340 |
| O23.41 | DX10 | Unsp infct of urinary tract in pregnancy, first trimester | Pregnancy | O2341 |

| | | | | |
|---|---|---|---|---|
| O23.42 | DX10 | Unsp infct of urinary tract in pregnancy, second trimester | Pregnancy | O2342 |
| O23.43 | DX10 | Unsp infct of urinary tract in pregnancy, third trimester | Pregnancy | O2343 |
| O23.511 | DX10 | Infections of cervix in pregnancy, first trimester | Pregnancy | O23511 |
| O23.512 | DX10 | Infections of cervix in pregnancy, second trimester | Pregnancy | O23512 |
| O23.513 | DX10 | Infections of cervix in pregnancy, third trimester | Pregnancy | O23513 |
| O23.519 | DX10 | Infections of cervix in pregnancy, unspecified trimester | Pregnancy | O23519 |
| O23.521 | DX10 | Salpingo-oophoritis in pregnancy, first trimester | Pregnancy | O23521 |
| O23.522 | DX10 | Salpingo-oophoritis in pregnancy, second trimester | Pregnancy | O23522 |
| O23.523 | DX10 | Salpingo-oophoritis in pregnancy, third trimester | Pregnancy | O23523 |
| O23.529 | DX10 | Salpingo-oophoritis in pregnancy, unspecified trimester | Pregnancy | O23529 |
| O23.591 | DX10 | Infection oth prt genitl trct in pregnancy, first trimester | Pregnancy | O23591 |
| O23.592 | DX10 | Infection oth prt genitl trct in pregnancy, second trimester | Pregnancy | O23592 |
| O23.593 | DX10 | Infection oth prt genitl trct in pregnancy, third trimester | Pregnancy | O23593 |
| O23.599 | DX10 | Infection oth prt genital tract in pregnancy, unsp trimester | Pregnancy | O23599 |
| O23.90 | DX10 | Unsp GU tract infection in pregnancy, unsp trimester | Pregnancy | O2390 |
| O23.91 | DX10 | Unsp GU tract infection in pregnancy, first trimester | Pregnancy | O2391 |
| O23.92 | DX10 | Unsp GU tract infection in pregnancy, second trimester | Pregnancy | O2392 |
| O23.93 | DX10 | Unsp GU tract infection in pregnancy, third trimester | Pregnancy | O2393 |
| O24.011 | DX10 | Pre-existing diabetes, type 1, in pregnancy, first trimester | Pregnancy | O24011 |
| O24.012 | DX10 | Pre-exist diabetes, type 1, in pregnancy, second trimester | Pregnancy | O24012 |
| O24.013 | DX10 | Pre-existing diabetes, type 1, in pregnancy, third trimester | Pregnancy | O24013 |
| O24.019 | DX10 | Pre-existing diabetes, type 1, in pregnancy, unsp trimester | Pregnancy | O24019 |
| O24.02 | DX10 | Pre-existing diabetes mellitus, type 1, in childbirth | Pregnancy | O2402 |
| O24.03 | DX10 | Pre-existing diabetes mellitus, type 1, in the puerperium | Pregnancy | O2403 |
| O24.111 | DX10 | Pre-existing diabetes, type 2, in pregnancy, first trimester | Pregnancy | O24111 |
| O24.112 | DX10 | Pre-exist diabetes, type 2, in pregnancy, second trimester | Pregnancy | O24112 |
| O24.113 | DX10 | Pre-existing diabetes, type 2, in pregnancy, third trimester | Pregnancy | O24113 |
| O24.119 | DX10 | Pre-existing diabetes, type 2, in pregnancy, unsp trimester | Pregnancy | O24119 |
| O24.12 | DX10 | Pre-existing diabetes mellitus, type 2, in childbirth | Pregnancy | O2412 |
| O24.13 | DX10 | Pre-existing diabetes mellitus, type 2, in the puerperium | Pregnancy | O2413 |
| O24.311 | DX10 | Unsp pre-existing diabetes in pregnancy, first trimester | Pregnancy | O24311 |
| O24.312 | DX10 | Unsp pre-existing diabetes in pregnancy, second trimester | Pregnancy | O24312 |
| O24.313 | DX10 | Unsp pre-existing diabetes in pregnancy, third trimester | Pregnancy | O24313 |
| O24.319 | DX10 | Unsp pre-existing diabetes in pregnancy, unsp trimester | Pregnancy | O24319 |
| O24.32 | DX10 | Unspecified pre-existing diabetes mellitus in childbirth | Pregnancy | O2432 |
| O24.33 | DX10 | Unspecified pre-existing diabetes mellitus in the puerperium | Pregnancy | O2433 |
| O24.410 | DX10 | Gestational diabetes mellitus in pregnancy, diet controlled | Pregnancy | O24410 |
| O24.414 | DX10 | Gestational diabetes in pregnancy, insulin controlled | Pregnancy | O24414 |
| O24.415 | DX10 | GESTATIONAL DM PREGNANCY CONTROLLED ORAL HG RX | Pregnancy | O24415 |
| O24.419 | DX10 | Gestational diabetes mellitus in pregnancy, unsp control | Pregnancy | O24419 |
| O24.420 | DX10 | Gestational diabetes mellitus in childbirth, diet controlled | Pregnancy | O24420 |
| O24.424 | DX10 | Gestational diabetes in childbirth, insulin controlled | Pregnancy | O24424 |
| O24.425 | DX10 | Gestational diabetes mellitus in childbirth, controlled by oral hypoglycemic drugs | Pregnancy | O24425 |

| | | | | |
|---|---|---|---|---|
| O24.429 | DX10 | Gestational diabetes mellitus in childbirth, unsp control | Pregnancy | O24429 |
| O24.430 | DX10 | Gestational diabetes in the puerperium, diet controlled | Pregnancy | O24430 |
| O24.434 | DX10 | Gestational diabetes in the puerperium, insulin controlled | Pregnancy | O24434 |
| O24.435 | DX10 | Gestational diabetes mellitus in puerperium, controlled by oral hypoglycemic drugs | Pregnancy | O24435 |
| O24.439 | DX10 | Gestational diabetes in the puerperium, unsp control | Pregnancy | O24439 |
| O24.811 | DX10 | Oth pre-existing diabetes in pregnancy, first trimester | Pregnancy | O24811 |
| O24.812 | DX10 | Oth pre-existing diabetes in pregnancy, second trimester | Pregnancy | O24812 |
| O24.813 | DX10 | Oth pre-existing diabetes in pregnancy, third trimester | Pregnancy | O24813 |
| O24.819 | DX10 | Oth pre-existing diabetes in pregnancy, unsp trimester | Pregnancy | O24819 |
| O24.82 | DX10 | Other pre-existing diabetes mellitus in childbirth | Pregnancy | O2482 |
| O24.83 | DX10 | Other pre-existing diabetes mellitus in the puerperium | Pregnancy | O2483 |
| O24.911 | DX10 | Unspecified diabetes mellitus in pregnancy, first trimester | Pregnancy | O24911 |
| O24.912 | DX10 | Unspecified diabetes mellitus in pregnancy, second trimester | Pregnancy | O24912 |
| O24.913 | DX10 | Unspecified diabetes mellitus in pregnancy, third trimester | Pregnancy | O24913 |
| O24.919 | DX10 | Unsp diabetes mellitus in pregnancy, unspecified trimester | Pregnancy | O24919 |
| O24.92 | DX10 | Unspecified diabetes mellitus in childbirth | Pregnancy | O2492 |
| O24.93 | DX10 | UNSPECIFIED DIABETES MELLITUS IN THE PUERPERIUM | Pregnancy | O2493 |
| O25.10 | DX10 | Malnutrition in pregnancy, unspecified trimester | Pregnancy | O2510 |
| O25.11 | DX10 | Malnutrition in pregnancy, first trimester | Pregnancy | O2511 |
| O25.12 | DX10 | Malnutrition in pregnancy, second trimester | Pregnancy | O2512 |
| O25.13 | DX10 | Malnutrition in pregnancy, third trimester | Pregnancy | O2513 |
| O25.2 | DX10 | Malnutrition in childbirth | Pregnancy | O252 |
| O25.3 | DX10 | Malnutrition in the puerperium | Pregnancy | O253 |
| O26.00 | DX10 | Excessive weight gain in pregnancy, unspecified trimester | Pregnancy | O2600 |
| O26.01 | DX10 | Excessive weight gain in pregnancy, first trimester | Pregnancy | O2601 |
| O26.02 | DX10 | Excessive weight gain in pregnancy, second trimester | Pregnancy | O2602 |
| O26.03 | DX10 | Excessive weight gain in pregnancy, third trimester | Pregnancy | O2603 |
| O26.10 | DX10 | Low weight gain in pregnancy, unspecified trimester | Pregnancy | O2610 |
| O26.11 | DX10 | Low weight gain in pregnancy, first trimester | Pregnancy | O2611 |
| O26.12 | DX10 | Low weight gain in pregnancy, second trimester | Pregnancy | O2612 |
| O26.13 | DX10 | Low weight gain in pregnancy, third trimester | Pregnancy | O2613 |
| O26.20 | DX10 | Preg care for patient w recurrent preg loss, unsp trimester | Pregnancy | O2620 |
| O26.21 | DX10 | Preg care for patient w recurrent preg loss, first trimester | Pregnancy | O2621 |
| O26.22 | DX10 | Preg care for patient w recur preg loss, second trimester | Pregnancy | O2622 |
| O26.23 | DX10 | Preg care for patient w recurrent preg loss, third trimester | Pregnancy | O2623 |
| O26.30 | DX10 | Retained uterin contracep dev in pregnancy, unsp trimester | Pregnancy | O2630 |
| O26.31 | DX10 | Retained uterin contracep dev in pregnancy, first trimester | Pregnancy | O2631 |
| O26.32 | DX10 | Retained uterin contracep dev in pregnancy, second trimester | Pregnancy | O2632 |
| O26.33 | DX10 | Retained uterin contracep dev in pregnancy, third trimester | Pregnancy | O2633 |
| O26.40 | DX10 | Herpes gestationis, unspecified trimester | Pregnancy | O2640 |
| O26.41 | DX10 | Herpes gestationis, first trimester | Pregnancy | O2641 |
| O26.42 | DX10 | Herpes gestationis, second trimester | Pregnancy | O2642 |
| O26.43 | DX10 | Herpes gestationis, third trimester | Pregnancy | O2643 |
| O26.50 | DX10 | Maternal hypotension syndrome, unspecified trimester | Pregnancy | O2650 |
| O26.51 | DX10 | Maternal hypotension syndrome, first trimester | Pregnancy | O2651 |
| O26.52 | DX10 | Maternal hypotension syndrome, second trimester | Pregnancy | O2652 |

| | | | | |
|---|---|---|---|---|
| O26.53 | DX10 | Maternal hypotension syndrome, third trimester | Pregnancy | O2653 |
| O26.611 | DX10 | Liver and biliary tract disord in pregnancy, first trimester | Pregnancy | O26611 |
| O26.612 | DX10 | Liver and biliary tract disord in preg, second trimester | Pregnancy | O26612 |
| O26.613 | DX10 | Liver and biliary tract disord in pregnancy, third trimester | Pregnancy | O26613 |
| O26.619 | DX10 | Liver and biliary tract disord in pregnancy, unsp trimester | Pregnancy | O26619 |
| O26.62 | DX10 | Liver and biliary tract disorders in childbirth | Pregnancy | O2662 |
| O26.63 | DX10 | Liver and biliary tract disorders in the puerperium | Pregnancy | O2663 |
| O26.711 | DX10 | Sublux of symphysis (pubis) in pregnancy, first trimester | Pregnancy | O26711 |
| O26.712 | DX10 | Sublux of symphysis (pubis) in pregnancy, second trimester | Pregnancy | O26712 |
| O26.713 | DX10 | Sublux of symphysis (pubis) in pregnancy, third trimester | Pregnancy | O26713 |
| O26.719 | DX10 | Sublux of symphysis (pubis) in pregnancy, unsp trimester | Pregnancy | O26719 |
| O26.72 | DX10 | Subluxation of symphysis (pubis) in childbirth | Pregnancy | O2672 |
| O26.73 | DX10 | Subluxation of symphysis (pubis) in the puerperium | Pregnancy | O2673 |
| O26.811 | DX10 | Pregnancy related exhaustion and fatigue, first trimester | Pregnancy | O26811 |
| O26.812 | DX10 | Pregnancy related exhaustion and fatigue, second trimester | Pregnancy | O26812 |
| O26.813 | DX10 | Pregnancy related exhaustion and fatigue, third trimester | Pregnancy | O26813 |
| O26.819 | DX10 | Pregnancy related exhaustion and fatigue, unsp trimester | Pregnancy | O26819 |
| O26.821 | DX10 | Pregnancy related peripheral neuritis, first trimester | Pregnancy | O26821 |
| O26.822 | DX10 | Pregnancy related peripheral neuritis, second trimester | Pregnancy | O26822 |
| O26.823 | DX10 | Pregnancy related peripheral neuritis, third trimester | Pregnancy | O26823 |
| O26.829 | DX10 | Pregnancy related peripheral neuritis, unspecified trimester | Pregnancy | O26829 |
| O26.831 | DX10 | Pregnancy related renal disease, first trimester | Pregnancy | O26831 |
| O26.832 | DX10 | Pregnancy related renal disease, second trimester | Pregnancy | O26832 |
| O26.833 | DX10 | Pregnancy related renal disease, third trimester | Pregnancy | O26833 |
| O26.839 | DX10 | Pregnancy related renal disease, unspecified trimester | Pregnancy | O26839 |
| O26.841 | DX10 | Uterine size-date discrepancy, first trimester | Pregnancy | O26841 |
| O26.842 | DX10 | Uterine size-date discrepancy, second trimester | Pregnancy | O26842 |
| O26.843 | DX10 | Uterine size-date discrepancy, third trimester | Pregnancy | O26843 |
| O26.849 | DX10 | Uterine size-date discrepancy, unspecified trimester | Pregnancy | O26849 |
| O26.851 | DX10 | Spotting complicating pregnancy, first trimester | Pregnancy | O26851 |
| O26.852 | DX10 | Spotting complicating pregnancy, second trimester | Pregnancy | O26852 |
| O26.853 | DX10 | Spotting complicating pregnancy, third trimester | Pregnancy | O26853 |
| O26.859 | DX10 | Spotting complicating pregnancy, unspecified trimester | Pregnancy | O26859 |
| O26.86 | DX10 | Pruritic urticarial papules and plaques of pregnancy (PUPPP) | Pregnancy | O2686 |
| O26.872 | DX10 | Cervical shortening, second trimester | Pregnancy | O26872 |
| O26.873 | DX10 | Cervical shortening, third trimester | Pregnancy | O26873 |
| O26.879 | DX10 | Cervical shortening, unspecified trimester | Pregnancy | O26879 |
| O26.891 | DX10 | Oth pregnancy related conditions, first trimester | Pregnancy | O26891 |
| O26.892 | DX10 | Oth pregnancy related conditions, second trimester | Pregnancy | O26892 |
| O26.893 | DX10 | Oth pregnancy related conditions, third trimester | Pregnancy | O26893 |
| O26.899 | DX10 | Oth pregnancy related conditions, unspecified trimester | Pregnancy | O26899 |
| O26.90 | DX10 | Pregnancy related conditions, unsp, unspecified trimester | Pregnancy | O2690 |
| O26.91 | DX10 | Pregnancy related conditions, unspecified, first trimester | Pregnancy | O2691 |
| O26.92 | DX10 | Pregnancy related conditions, unspecified, second trimester | Pregnancy | O2692 |
| O26.93 | DX10 | Pregnancy related conditions, unspecified, third trimester | Pregnancy | O2693 |
| O28.0 | DX10 | Abnormal hematolog finding on antenatal screening of mother | Pregnancy | O280 |
| O28.1 | DX10 | Abnormal biochemical finding on antenat screening of mother | Pregnancy | O281 |

| | | | | |
|---|---|---|---|---|
| O28.2 | DX10 | Abnormal cytolog finding on antenatal screening of mother | Pregnancy | O282 |
| O28.3 | DX10 | Abnormal ultrasonic finding on antenatal screening of mother | Pregnancy | O283 |
| O28.4 | DX10 | Abnormal radiolog finding on antenatal screening of mother | Pregnancy | O284 |
| O28.5 | DX10 | Abn chromsoml and genetic find on antenat screen of mother | Pregnancy | O285 |
| O28.8 | DX10 | Other abnormal findings on antenatal screening of mother | Pregnancy | O288 |
| O28.9 | DX10 | Unsp abnormal findings on antenatal screening of mother | Pregnancy | O289 |
| O29.011 | DX10 | Aspirat pneumonitis due to anesth during preg, first tri | Pregnancy | O29011 |
| O29.012 | DX10 | Aspirat pneumonitis due to anesth during preg, second tri | Pregnancy | O29012 |
| O29.013 | DX10 | Aspirat pneumonitis due to anesth during preg, third tri | Pregnancy | O29013 |
| O29.019 | DX10 | Aspirat pneumonitis due to anesth during preg, unsp tri | Pregnancy | O29019 |
| O29.021 | DX10 | Pressr collapse of lung due to anesth during preg, first tri | Pregnancy | O29021 |
| O29.022 | DX10 | Pressr collapse of lung d/t anesth during preg, second tri | Pregnancy | O29022 |
| O29.023 | DX10 | Pressr collapse of lung due to anesth during preg, third tri | Pregnancy | O29023 |
| O29.029 | DX10 | Pressr collapse of lung due to anesth during preg, unsp tri | Pregnancy | O29029 |
| O29.091 | DX10 | Oth pulmonary comp of anesth during preg, first trimester | Pregnancy | O29091 |
| O29.092 | DX10 | Oth pulmonary comp of anesth during preg, second trimester | Pregnancy | O29092 |
| O29.093 | DX10 | Oth pulmonary comp of anesth during preg, third trimester | Pregnancy | O29093 |
| O29.099 | DX10 | Oth pulmonary comp of anesth during preg, unsp trimester | Pregnancy | O29099 |
| O29.111 | DX10 | Cardiac arrest due to anesth during preg, first trimester | Pregnancy | O29111 |
| O29.112 | DX10 | Cardiac arrest due to anesth during preg, second trimester | Pregnancy | O29112 |
| O29.113 | DX10 | Cardiac arrest due to anesth during preg, third trimester | Pregnancy | O29113 |
| O29.119 | DX10 | Cardiac arrest due to anesth during preg, unsp trimester | Pregnancy | O29119 |
| O29.121 | DX10 | Cardiac failure due to anesth during preg, first trimester | Pregnancy | O29121 |
| O29.122 | DX10 | Cardiac failure due to anesth during preg, second trimester | Pregnancy | O29122 |
| O29.123 | DX10 | Cardiac failure due to anesth during preg, third trimester | Pregnancy | O29123 |
| O29.129 | DX10 | Cardiac failure due to anesth during preg, unsp trimester | Pregnancy | O29129 |
| O29.191 | DX10 | Oth cardiac comp of anesth during pregnancy, first trimester | Pregnancy | O29191 |
| O29.192 | DX10 | Oth cardiac comp of anesth during preg, second trimester | Pregnancy | O29192 |
| O29.193 | DX10 | Oth cardiac comp of anesth during pregnancy, third trimester | Pregnancy | O29193 |
| O29.199 | DX10 | Oth cardiac comp of anesth during pregnancy, unsp trimester | Pregnancy | O29199 |
| O29.211 | DX10 | Cerebral anoxia due to anesth during preg, first trimester | Pregnancy | O29211 |
| O29.212 | DX10 | Cerebral anoxia due to anesth during preg, second trimester | Pregnancy | O29212 |
| O29.213 | DX10 | Cerebral anoxia due to anesth during preg, third trimester | Pregnancy | O29213 |
| O29.219 | DX10 | Cerebral anoxia due to anesth during preg, unsp trimester | Pregnancy | O29219 |
| O29.291 | DX10 | Oth cnsl comp of anesth during pregnancy, first trimester | Pregnancy | O29291 |

| | | | | |
|---|---|---|---|---|
| O29.292 | DX10 | Oth cnsl comp of anesth during pregnancy, second trimester | Pregnancy | O29292 |
| O29.293 | DX10 | Oth cnsl comp of anesth during pregnancy, third trimester | Pregnancy | O29293 |
| O29.299 | DX10 | Oth cnsl comp of anesthesia during pregnancy, unsp trimester | Pregnancy | O29299 |
| O29.3X1 | DX10 | Toxic reaction to local anesth during preg, first trimester | Pregnancy | O293X1 |
| O29.3X2 | DX10 | Toxic reaction to local anesth during preg, second trimester | Pregnancy | O293X2 |
| O29.3X3 | DX10 | Toxic reaction to local anesth during preg, third trimester | Pregnancy | O293X3 |
| O29.3X9 | DX10 | Toxic reaction to local anesth during preg, unsp trimester | Pregnancy | O293X9 |
| O29.40 | DX10 | Spinal and epidur anesth induce hdache during preg, unsp tri | Pregnancy | O2940 |
| O29.41 | DX10 | Spinal and epidur anesth induce hdache dur preg, first tri | Pregnancy | O2941 |
| O29.42 | DX10 | Spinal and epidur anesth induce hdache dur preg, second tri | Pregnancy | O2942 |
| O29.43 | DX10 | Spinal and epidur anesth induce hdache dur preg, third tri | Pregnancy | O2943 |
| O29.5X1 | DX10 | Oth comp of spinal and epidur anesth during preg, first tri | Pregnancy | O295X1 |
| O29.5X2 | DX10 | Oth comp of spinal and epidur anesth during preg, second tri | Pregnancy | O295X2 |
| O29.5X3 | DX10 | Oth comp of spinal and epidur anesth during preg, third tri | Pregnancy | O295X3 |
| O29.5X9 | DX10 | Oth comp of spinal and epidural anesth during preg, unsp tri | Pregnancy | O295X9 |
| O29.60 | DX10 | Failed or difficult intubation for anesth dur preg, unsp tri | Pregnancy | O2960 |
| O29.61 | DX10 | Fail or difficult intubation for anesth dur preg, first tri | Pregnancy | O2961 |
| O29.62 | DX10 | Fail or difficult intubation for anesth dur preg, second tri | Pregnancy | O2962 |
| O29.63 | DX10 | Fail or difficult intubation for anesth dur preg, third tri | Pregnancy | O2963 |
| O29.8X1 | DX10 | Oth comp of anesthesia during pregnancy, first trimester | Pregnancy | O298X1 |
| O29.8X2 | DX10 | Oth comp of anesthesia during pregnancy, second trimester | Pregnancy | O298X2 |
| O29.8X3 | DX10 | Oth comp of anesthesia during pregnancy, third trimester | Pregnancy | O298X3 |
| O29.8X9 | DX10 | Oth comp of anesthesia during pregnancy, unsp trimester | Pregnancy | O298X9 |
| O29.90 | DX10 | Unsp comp of anesthesia during pregnancy, unsp trimester | Pregnancy | O2990 |
| O29.91 | DX10 | Unsp comp of anesthesia during pregnancy, first trimester | Pregnancy | O2991 |
| O29.92 | DX10 | Unsp comp of anesthesia during pregnancy, second trimester | Pregnancy | O2992 |
| O29.93 | DX10 | Unsp comp of anesthesia during pregnancy, third trimester | Pregnancy | O2993 |
| O30.001 | DX10 | Twin preg, unsp num plcnta & amnio sacs, first trimester | Pregnancy | O30001 |
| O30.002 | DX10 | Twin preg, unsp num plcnta & amnio sacs, second trimester | Pregnancy | O30002 |
| O30.003 | DX10 | Twin preg, unsp num plcnta & amnio sacs, third trimester | Pregnancy | O30003 |
| O30.009 | DX10 | Twin pregnancy, unsp num plcnta & amnio sacs, unsp trimester | Pregnancy | O30009 |
| O30.011 | DX10 | Twin pregnancy, monochorionic/monoamniotic, first trimester | Pregnancy | O30011 |
| O30.012 | DX10 | Twin pregnancy, monochorionic/monoamniotic, second trimester | Pregnancy | O30012 |

| | | | | |
|---|---|---|---|---|
| O30.013 | DX10 | Twin pregnancy, monochorionic/monoamniotic, third trimester | Pregnancy | O30013 |
| O30.019 | DX10 | Twin pregnancy, monochorionic/monoamniotic, unsp trimester | Pregnancy | O30019 |
| O30.021 | DX10 | Conjoined twin pregnancy, first trimester | Pregnancy | O30021 |
| O30.022 | DX10 | Conjoined twin pregnancy, second trimester | Pregnancy | O30022 |
| O30.023 | DX10 | Conjoined twin pregnancy, third trimester | Pregnancy | O30023 |
| O30.029 | DX10 | Conjoined twin pregnancy, unspecified trimester | Pregnancy | O30029 |
| O30.031 | DX10 | Twin pregnancy, monochorionic/diamniotic, first trimester | Pregnancy | O30031 |
| O30.032 | DX10 | Twin pregnancy, monochorionic/diamniotic, second trimester | Pregnancy | O30032 |
| O30.033 | DX10 | Twin pregnancy, monochorionic/diamniotic, third trimester | Pregnancy | O30033 |
| O30.039 | DX10 | Twin pregnancy, monochorionic/diamniotic, unsp trimester | Pregnancy | O30039 |
| O30.041 | DX10 | Twin pregnancy, dichorionic/diamniotic, first trimester | Pregnancy | O30041 |
| O30.042 | DX10 | Twin pregnancy, dichorionic/diamniotic, second trimester | Pregnancy | O30042 |
| O30.043 | DX10 | Twin pregnancy, dichorionic/diamniotic, third trimester | Pregnancy | O30043 |
| O30.049 | DX10 | Twin pregnancy, dichorionic/diamniotic, unsp trimester | Pregnancy | O30049 |
| O30.091 | DX10 | Twin preg, unable to dtrm num plcnta & amnio sacs, first tri | Pregnancy | O30091 |
| O30.092 | DX10 | Twin preg, unable to dtrm num plcnta & amnio sacs, 2nd tri | Pregnancy | O30092 |
| O30.093 | DX10 | Twin preg, unable to dtrm num plcnta & amnio sacs, third tri | Pregnancy | O30093 |
| O30.099 | DX10 | Twin preg, unable to dtrm num plcnta & amnio sacs, unsp tri | Pregnancy | O30099 |
| O30.101 | DX10 | Triplet preg, unsp num plcnta & amnio sacs, first trimester | Pregnancy | O30101 |
| O30.102 | DX10 | Triplet preg, unsp num plcnta & amnio sacs, second trimester | Pregnancy | O30102 |
| O30.103 | DX10 | Triplet preg, unsp num plcnta & amnio sacs, third trimester | Pregnancy | O30103 |
| O30.109 | DX10 | Triplet preg, unsp num plcnta & amnio sacs, unsp trimester | Pregnancy | O30109 |
| O30.111 | DX10 | Triplet preg w two or more monochorionic fetuses, first tri | Pregnancy | O30111 |
| O30.112 | DX10 | Triplet preg w two or more monochorionic fetuses, second tri | Pregnancy | O30112 |
| O30.113 | DX10 | Triplet preg w two or more monochorionic fetuses, third tri | Pregnancy | O30113 |
| O30.119 | DX10 | Triplet preg w two or more monochorionic fetuses, unsp tri | Pregnancy | O30119 |
| O30.121 | DX10 | Triplet preg w two or more monoamnio fetuses, first tri | Pregnancy | O30121 |
| O30.122 | DX10 | Triplet preg w two or more monoamnio fetuses, second tri | Pregnancy | O30122 |
| O30.123 | DX10 | Triplet preg w two or more monoamnio fetuses, third tri | Pregnancy | O30123 |
| O30.129 | DX10 | Triplet preg w two or more monoamnio fetuses, unsp trimester | Pregnancy | O30129 |
| O30.191 | DX10 | Trp preg, unable to dtrm num plcnta & amnio sacs, first tri | Pregnancy | O30191 |
| O30.192 | DX10 | Trp preg, unable to dtrm num plcnta & amnio sacs, second tri | Pregnancy | O30192 |
| O30.193 | DX10 | Trp preg, unable to dtrm num plcnta & amnio sacs, third tri | Pregnancy | O30193 |
| O30.199 | DX10 | Trp preg, unable to dtrm num plcnta & amnio sacs, unsp tri | Pregnancy | O30199 |

| | | | | |
|---|---|---|---|---|
| O30.201 | DX10 | Quad preg, unsp num plcnta & amnio sacs, first trimester | Pregnancy | O30201 |
| O30.202 | DX10 | Quad preg, unsp num plcnta & amnio sacs, second trimester | Pregnancy | O30202 |
| O30.203 | DX10 | Quad preg, unsp num plcnta & amnio sacs, third trimester | Pregnancy | O30203 |
| O30.209 | DX10 | Quad pregnancy, unsp num plcnta & amnio sacs, unsp trimester | Pregnancy | O30209 |
| O30.211 | DX10 | Quad preg w two or more monochorionic fetuses, first tri | Pregnancy | O30211 |
| O30.212 | DX10 | Quad preg w two or more monochorionic fetuses, second tri | Pregnancy | O30212 |
| O30.213 | DX10 | Quad preg w two or more monochorionic fetuses, third tri | Pregnancy | O30213 |
| O30.219 | DX10 | Quad preg w two or more monochorionic fetuses, unsp tri | Pregnancy | O30219 |
| O30.221 | DX10 | Quad preg w two or more monoamnio fetuses, first trimester | Pregnancy | O30221 |
| O30.222 | DX10 | Quad preg w two or more monoamnio fetuses, second trimester | Pregnancy | O30222 |
| O30.223 | DX10 | Quad preg w two or more monoamnio fetuses, third trimester | Pregnancy | O30223 |
| O30.229 | DX10 | Quad preg w two or more monoamnio fetuses, unsp trimester | Pregnancy | O30229 |
| O30.291 | DX10 | Quad preg, unable to dtrm num plcnta & amnio sacs, first tri | Pregnancy | O30291 |
| O30.292 | DX10 | Quad preg, unable to dtrm num plcnta & amnio sacs, 2nd tri | Pregnancy | O30292 |
| O30.293 | DX10 | Quad preg, unable to dtrm num plcnta & amnio sacs, third tri | Pregnancy | O30293 |
| O30.299 | DX10 | Quad preg, unable to dtrm num plcnta & amnio sacs, unsp tri | Pregnancy | O30299 |
| O30.801 | DX10 | Oth multiple gest, unsp num plcnta & amnio sacs, first tri | Pregnancy | O30801 |
| O30.802 | DX10 | Oth multiple gest, unsp num plcnta & amnio sacs, second tri | Pregnancy | O30802 |
| O30.803 | DX10 | Oth multiple gest, unsp num plcnta & amnio sacs, third tri | Pregnancy | O30803 |
| O30.809 | DX10 | Oth multiple gest, unsp num plcnta & amnio sacs, unsp tri | Pregnancy | O30809 |
| O30.811 | DX10 | Oth mult gest w two or more monochorionic fetuses, first tri | Pregnancy | O30811 |
| O30.812 | DX10 | Oth mult gest w two or more monochorionic fetuses, 2nd tri | Pregnancy | O30812 |
| O30.813 | DX10 | Oth mult gest w two or more monochorionic fetuses, third tri | Pregnancy | O30813 |
| O30.819 | DX10 | Oth mult gest w two or more monochorionic fetuses, unsp tri | Pregnancy | O30819 |
| O30.821 | DX10 | Oth multiple gest w two or more monoamnio fetuses, first tri | Pregnancy | O30821 |
| O30.822 | DX10 | Oth mult gest w two or more monoamnio fetuses, second tri | Pregnancy | O30822 |
| O30.823 | DX10 | Oth multiple gest w two or more monoamnio fetuses, third tri | Pregnancy | O30823 |
| O30.829 | DX10 | Oth multiple gest w two or more monoamnio fetuses, unsp tri | Pregnancy | O30829 |
| O30.891 | DX10 | Oth mult gest, unab to dtrm num plcnta & amnio sacs, 1st tri | Pregnancy | O30891 |
| O30.892 | DX10 | Oth mult gest, unab to dtrm num plcnta & amnio sacs, 2nd tri | Pregnancy | O30892 |

| | | | | |
|---|---|---|---|---|
| O30.893 | DX10 | Oth mult gest, unab to dtrm num plcnta & amnio sacs, 3rd tri | Pregnancy | O30893 |
| O30.899 | DX10 | Oth mult gest,unab to dtrm num plcnta & amnio sacs, unsp tri | Pregnancy | O30899 |
| O30.90 | DX10 | Multiple gestation, unspecified, unspecified trimester | Pregnancy | O3090 |
| O30.91 | DX10 | Multiple gestation, unspecified, first trimester | Pregnancy | O3091 |
| O30.92 | DX10 | Multiple gestation, unspecified, second trimester | Pregnancy | O3092 |
| O30.93 | DX10 | Multiple gestation, unspecified, third trimester | Pregnancy | O3093 |
| O31.00X0 | DX10 | Papyraceous fetus, unsp trimester, not applicable or unsp | Pregnancy | O3100X0 |
| O31.00X1 | DX10 | Papyraceous fetus, unspecified trimester, fetus 1 | Pregnancy | O3100X1 |
| O31.00X2 | DX10 | Papyraceous fetus, unspecified trimester, fetus 2 | Pregnancy | O3100X2 |
| O31.00X3 | DX10 | Papyraceous fetus, unspecified trimester, fetus 3 | Pregnancy | O3100X3 |
| O31.00X4 | DX10 | Papyraceous fetus, unspecified trimester, fetus 4 | Pregnancy | O3100X4 |
| O31.00X5 | DX10 | Papyraceous fetus, unspecified trimester, fetus 5 | Pregnancy | O3100X5 |
| O31.00X9 | DX10 | Papyraceous fetus, unspecified trimester, other fetus | Pregnancy | O3100X9 |
| O31.01X0 | DX10 | Papyraceous fetus, first trimester, not applicable or unsp | Pregnancy | O3101X0 |
| O31.01X1 | DX10 | Papyraceous fetus, first trimester, fetus 1 | Pregnancy | O3101X1 |
| O31.01X2 | DX10 | Papyraceous fetus, first trimester, fetus 2 | Pregnancy | O3101X2 |
| O31.01X3 | DX10 | Papyraceous fetus, first trimester, fetus 3 | Pregnancy | O3101X3 |
| O31.01X4 | DX10 | Papyraceous fetus, first trimester, fetus 4 | Pregnancy | O3101X4 |
| O31.01X5 | DX10 | Papyraceous fetus, first trimester, fetus 5 | Pregnancy | O3101X5 |
| O31.01X9 | DX10 | Papyraceous fetus, first trimester, other fetus | Pregnancy | O3101X9 |
| O31.02X0 | DX10 | Papyraceous fetus, second trimester, not applicable or unsp | Pregnancy | O3102X0 |
| O31.02X1 | DX10 | Papyraceous fetus, second trimester, fetus 1 | Pregnancy | O3102X1 |
| O31.02X2 | DX10 | Papyraceous fetus, second trimester, fetus 2 | Pregnancy | O3102X2 |
| O31.02X3 | DX10 | Papyraceous fetus, second trimester, fetus 3 | Pregnancy | O3102X3 |
| O31.02X4 | DX10 | Papyraceous fetus, second trimester, fetus 4 | Pregnancy | O3102X4 |
| O31.02X5 | DX10 | Papyraceous fetus, second trimester, fetus 5 | Pregnancy | O3102X5 |
| O31.02X9 | DX10 | Papyraceous fetus, second trimester, other fetus | Pregnancy | O3102X9 |
| O31.03X0 | DX10 | Papyraceous fetus, third trimester, not applicable or unsp | Pregnancy | O3103X0 |
| O31.03X1 | DX10 | Papyraceous fetus, third trimester, fetus 1 | Pregnancy | O3103X1 |
| O31.03X2 | DX10 | Papyraceous fetus, third trimester, fetus 2 | Pregnancy | O3103X2 |
| O31.03X3 | DX10 | Papyraceous fetus, third trimester, fetus 3 | Pregnancy | O3103X3 |
| O31.03X4 | DX10 | Papyraceous fetus, third trimester, fetus 4 | Pregnancy | O3103X4 |
| O31.03X5 | DX10 | Papyraceous fetus, third trimester, fetus 5 | Pregnancy | O3103X5 |
| O31.03X9 | DX10 | Papyraceous fetus, third trimester, other fetus | Pregnancy | O3103X9 |
| O31.10X0 | DX10 | Cont preg aft spon abort of one fts or more, unsp tri, unsp | Pregnancy | O3110X0 |
| O31.10X1 | DX10 | Cont preg aft spon abort of one fts or more, unsp tri, fts1 | Pregnancy | O3110X1 |
| O31.10X2 | DX10 | Cont preg aft spon abort of one fts or more, unsp tri, fts2 | Pregnancy | O3110X2 |
| O31.10X3 | DX10 | Cont preg aft spon abort of one fts or more, unsp tri, fts3 | Pregnancy | O3110X3 |
| O31.10X4 | DX10 | Cont preg aft spon abort of one fts or more, unsp tri, fts4 | Pregnancy | O3110X4 |
| O31.10X5 | DX10 | Cont preg aft spon abort of one fts or more, unsp tri, fts5 | Pregnancy | O3110X5 |
| O31.10X9 | DX10 | Cont preg aft spon abort of one fetus or more, unsp tri, oth | Pregnancy | O3110X9 |
| O31.11X0 | DX10 | Cont preg aft spon abort of one fts or more, first tri, unsp | Pregnancy | O3111X0 |
| O31.11X1 | DX10 | Cont preg aft spon abort of one fts or more, first tri, fts1 | Pregnancy | O3111X1 |
| O31.11X2 | DX10 | Cont preg aft spon abort of one fts or more, first tri, fts2 | Pregnancy | O3111X2 |
| O31.11X3 | DX10 | Cont preg aft spon abort of one fts or more, first tri, fts3 | Pregnancy | O3111X3 |
| O31.11X4 | DX10 | Cont preg aft spon abort of one fts or more, first tri, fts4 | Pregnancy | O3111X4 |

| | | | | |
|---|---|---|---|---|
| O31.11X5 | DX10 | Cont preg aft spon abort of one fts or more, first tri, fts5 | Pregnancy | O3111X5 |
| O31.11X9 | DX10 | Cont preg aft spon abort of one fts or more, first tri, oth | Pregnancy | O3111X9 |
| O31.12X0 | DX10 | Cont preg aft spon abort of one fetus or more, 2nd tri, unsp | Pregnancy | O3112X0 |
| O31.12X1 | DX10 | Cont preg aft spon abort of one fetus or more, 2nd tri, fts1 | Pregnancy | O3112X1 |
| O31.12X2 | DX10 | Cont preg aft spon abort of one fetus or more, 2nd tri, fts2 | Pregnancy | O3112X2 |
| O31.12X3 | DX10 | Cont preg aft spon abort of one fetus or more, 2nd tri, fts3 | Pregnancy | O3112X3 |
| O31.12X4 | DX10 | Cont preg aft spon abort of one fetus or more, 2nd tri, fts4 | Pregnancy | O3112X4 |
| O31.12X5 | DX10 | Cont preg aft spon abort of one fetus or more, 2nd tri, fts5 | Pregnancy | O3112X5 |
| O31.12X9 | DX10 | Cont preg aft spon abort of one fetus or more, 2nd tri, oth | Pregnancy | O3112X9 |
| O31.13X0 | DX10 | Cont preg aft spon abort of one fts or more, third tri, unsp | Pregnancy | O3113X0 |
| O31.13X1 | DX10 | Cont preg aft spon abort of one fts or more, third tri, fts1 | Pregnancy | O3113X1 |
| O31.13X2 | DX10 | Cont preg aft spon abort of one fts or more, third tri, fts2 | Pregnancy | O3113X2 |
| O31.13X3 | DX10 | Cont preg aft spon abort of one fts or more, third tri, fts3 | Pregnancy | O3113X3 |
| O31.13X4 | DX10 | Cont preg aft spon abort of one fts or more, third tri, fts4 | Pregnancy | O3113X4 |
| O31.13X5 | DX10 | Cont preg aft spon abort of one fts or more, third tri, fts5 | Pregnancy | O3113X5 |
| O31.13X9 | DX10 | Cont preg aft spon abort of one fts or more, third tri, oth | Pregnancy | O3113X9 |
| O31.20X0 | DX10 | Cont preg aft uterin dth of one fts or more, unsp tri, unsp | Pregnancy | O3120X0 |
| O31.20X1 | DX10 | Cont preg aft uterin dth of one fts or more, unsp tri, fts1 | Pregnancy | O3120X1 |
| O31.20X2 | DX10 | Cont preg aft uterin dth of one fts or more, unsp tri, fts2 | Pregnancy | O3120X2 |
| O31.20X3 | DX10 | Cont preg aft uterin dth of one fts or more, unsp tri, fts3 | Pregnancy | O3120X3 |
| O31.20X4 | DX10 | Cont preg aft uterin dth of one fts or more, unsp tri, fts4 | Pregnancy | O3120X4 |
| O31.20X5 | DX10 | Cont preg aft uterin dth of one fts or more, unsp tri, fts5 | Pregnancy | O3120X5 |
| O31.20X9 | DX10 | Cont preg aft uterin dth of one fetus or more, unsp tri, oth | Pregnancy | O3120X9 |
| O31.21X0 | DX10 | Cont preg aft uterin dth of one fts or more, first tri, unsp | Pregnancy | O3121X0 |
| O31.21X1 | DX10 | Cont preg aft uterin dth of one fts or more, first tri, fts1 | Pregnancy | O3121X1 |
| O31.21X2 | DX10 | Cont preg aft uterin dth of one fts or more, first tri, fts2 | Pregnancy | O3121X2 |
| O31.21X3 | DX10 | Cont preg aft uterin dth of one fts or more, first tri, fts3 | Pregnancy | O3121X3 |
| O31.21X4 | DX10 | Cont preg aft uterin dth of one fts or more, first tri, fts4 | Pregnancy | O3121X4 |
| O31.21X5 | DX10 | Cont preg aft uterin dth of one fts or more, first tri, fts5 | Pregnancy | O3121X5 |
| O31.21X9 | DX10 | Cont preg aft uterin dth of one fts or more, first tri, oth | Pregnancy | O3121X9 |
| O31.22X0 | DX10 | Cont preg aft uterin dth of one fetus or more, 2nd tri, unsp | Pregnancy | O3122X0 |
| O31.22X1 | DX10 | Cont preg aft uterin dth of one fetus or more, 2nd tri, fts1 | Pregnancy | O3122X1 |
| O31.22X2 | DX10 | Cont preg aft uterin dth of one fetus or more, 2nd tri, fts2 | Pregnancy | O3122X2 |
| O31.22X3 | DX10 | Cont preg aft uterin dth of one fetus or more, 2nd tri, fts3 | Pregnancy | O3122X3 |
| O31.22X4 | DX10 | Cont preg aft uterin dth of one fetus or more, 2nd tri, fts4 | Pregnancy | O3122X4 |
| O31.22X5 | DX10 | Cont preg aft uterin dth of one fetus or more, 2nd tri, fts5 | Pregnancy | O3122X5 |
| O31.22X9 | DX10 | Cont preg aft uterin dth of one fetus or more, 2nd tri, oth | Pregnancy | O3122X9 |
| O31.23X0 | DX10 | Cont preg aft uterin dth of one fts or more, third tri, unsp | Pregnancy | O3123X0 |
| O31.23X1 | DX10 | Cont preg aft uterin dth of one fts or more, third tri, fts1 | Pregnancy | O3123X1 |
| O31.23X2 | DX10 | Cont preg aft uterin dth of one fts or more, third tri, fts2 | Pregnancy | O3123X2 |
| O31.23X3 | DX10 | Cont preg aft uterin dth of one fts or more, third tri, fts3 | Pregnancy | O3123X3 |
| O31.23X4 | DX10 | Cont preg aft uterin dth of one fts or more, third tri, fts4 | Pregnancy | O3123X4 |
| O31.23X5 | DX10 | Cont preg aft uterin dth of one fts or more, third tri, fts5 | Pregnancy | O3123X5 |

| | | | | |
|---|---|---|---|---|
| O31.23X9 | DX10 | Cont preg aft uterin dth of one fts or more, third tri, oth | Pregnancy | O3123X9 |
| O31.30X0 | DX10 | Cont preg aft elctv fetl rdct of 1 fts or more,unsp tri,unsp | Pregnancy | O3130X0 |
| O31.30X1 | DX10 | Cont preg aft elctv fetl rdct of 1 fts or more,unsp tri,fts1 | Pregnancy | O3130X1 |
| O31.30X2 | DX10 | Cont preg aft elctv fetl rdct of 1 fts or more,unsp tri,fts2 | Pregnancy | O3130X2 |
| O31.30X3 | DX10 | Cont preg aft elctv fetl rdct of 1 fts or more,unsp tri,fts3 | Pregnancy | O3130X3 |
| O31.30X4 | DX10 | Cont preg aft elctv fetl rdct of 1 fts or more,unsp tri,fts4 | Pregnancy | O3130X4 |
| O31.30X5 | DX10 | Cont preg aft elctv fetl rdct of 1 fts or more,unsp tri,fts5 | Pregnancy | O3130X5 |
| O31.30X9 | DX10 | Cont preg aft elctv fetl rdct of 1 fts or more,unsp tri, oth | Pregnancy | O3130X9 |
| O31.31X0 | DX10 | Cont preg aft elctv fetl rdct of 1 fts or more,1st tri, unsp | Pregnancy | O3131X0 |
| O31.31X1 | DX10 | Cont preg aft elctv fetl rdct of 1 fts or more,1st tri, fts1 | Pregnancy | O3131X1 |
| O31.31X2 | DX10 | Cont preg aft elctv fetl rdct of 1 fts or more,1st tri, fts2 | Pregnancy | O3131X2 |
| O31.31X3 | DX10 | Cont preg aft elctv fetl rdct of 1 fts or more,1st tri, fts3 | Pregnancy | O3131X3 |
| O31.31X4 | DX10 | Cont preg aft elctv fetl rdct of 1 fts or more,1st tri, fts4 | Pregnancy | O3131X4 |
| O31.31X5 | DX10 | Cont preg aft elctv fetl rdct of 1 fts or more,1st tri, fts5 | Pregnancy | O3131X5 |
| O31.31X9 | DX10 | Cont preg aft elctv fetl rdct of 1 fts or more, 1st tri, oth | Pregnancy | O3131X9 |
| O31.32X0 | DX10 | Cont preg aft elctv fetl rdct of 1 fts or more,2nd tri, unsp | Pregnancy | O3132X0 |
| O31.32X1 | DX10 | Cont preg aft elctv fetl rdct of 1 fts or more,2nd tri, fts1 | Pregnancy | O3132X1 |
| O31.32X2 | DX10 | Cont preg aft elctv fetl rdct of 1 fts or more,2nd tri, fts2 | Pregnancy | O3132X2 |
| O31.32X3 | DX10 | Cont preg aft elctv fetl rdct of 1 fts or more,2nd tri, fts3 | Pregnancy | O3132X3 |
| O31.32X4 | DX10 | Cont preg aft elctv fetl rdct of 1 fts or more,2nd tri, fts4 | Pregnancy | O3132X4 |
| O31.32X5 | DX10 | Cont preg aft elctv fetl rdct of 1 fts or more,2nd tri, fts5 | Pregnancy | O3132X5 |
| O31.32X9 | DX10 | Cont preg aft elctv fetl rdct of 1 fts or more, 2nd tri, oth | Pregnancy | O3132X9 |
| O31.33X0 | DX10 | Cont preg aft elctv fetl rdct of 1 fts or more,3rd tri, unsp | Pregnancy | O3133X0 |
| O31.33X1 | DX10 | Cont preg aft elctv fetl rdct of 1 fts or more,3rd tri, fts1 | Pregnancy | O3133X1 |
| O31.33X2 | DX10 | Cont preg aft elctv fetl rdct of 1 fts or more,3rd tri, fts2 | Pregnancy | O3133X2 |
| O31.33X3 | DX10 | Cont preg aft elctv fetl rdct of 1 fts or more,3rd tri, fts3 | Pregnancy | O3133X3 |
| O31.33X4 | DX10 | Cont preg aft elctv fetl rdct of 1 fts or more,3rd tri, fts4 | Pregnancy | O3133X4 |
| O31.33X5 | DX10 | Cont preg aft elctv fetl rdct of 1 fts or more,3rd tri, fts5 | Pregnancy | O3133X5 |
| O31.33X9 | DX10 | Cont preg aft elctv fetl rdct of 1 fts or more, 3rd tri, oth | Pregnancy | O3133X9 |
| O31.8X10 | DX10 | Oth comp specific to multiple gest, first trimester, unsp | Pregnancy | O318X10 |
| O31.8X11 | DX10 | Oth comp specific to multiple gest, first trimester, fetus 1 | Pregnancy | O318X11 |
| O31.8X12 | DX10 | Oth comp specific to multiple gest, first trimester, fetus 2 | Pregnancy | O318X12 |
| O31.8X13 | DX10 | Oth comp specific to multiple gest, first trimester, fetus 3 | Pregnancy | O318X13 |
| O31.8X14 | DX10 | Oth comp specific to multiple gest, first trimester, fetus 4 | Pregnancy | O318X14 |
| O31.8X15 | DX10 | Oth comp specific to multiple gest, first trimester, fetus 5 | Pregnancy | O318X15 |
| O31.8X19 | DX10 | Oth comp specific to multiple gest, first trimester, oth | Pregnancy | O318X19 |
| O31.8X20 | DX10 | Oth comp specific to multiple gest, second trimester, unsp | Pregnancy | O318X20 |
| O31.8X21 | DX10 | Oth comp specific to multiple gest, second tri, fetus 1 | Pregnancy | O318X21 |
| O31.8X22 | DX10 | Oth comp specific to multiple gest, second tri, fetus 2 | Pregnancy | O318X22 |
| O31.8X23 | DX10 | Oth comp specific to multiple gest, second tri, fetus 3 | Pregnancy | O318X23 |
| O31.8X24 | DX10 | Oth comp specific to multiple gest, second tri, fetus 4 | Pregnancy | O318X24 |
| O31.8X25 | DX10 | Oth comp specific to multiple gest, second tri, fetus 5 | Pregnancy | O318X25 |
| O31.8X29 | DX10 | Oth comp specific to multiple gest, second trimester, oth | Pregnancy | O318X29 |
| O31.8X30 | DX10 | Oth comp specific to multiple gest, third trimester, unsp | Pregnancy | O318X30 |
| O31.8X31 | DX10 | Oth comp specific to multiple gest, third trimester, fetus 1 | Pregnancy | O318X31 |
| O31.8X32 | DX10 | Oth comp specific to multiple gest, third trimester, fetus 2 | Pregnancy | O318X32 |
| O31.8X33 | DX10 | Oth comp specific to multiple gest, third trimester, fetus 3 | Pregnancy | O318X33 |
| O31.8X34 | DX10 | Oth comp specific to multiple gest, third trimester, fetus 4 | Pregnancy | O318X34 |

| | | | | |
|---|---|---|---|---|
| O31.8X35 | DX10 | Oth comp specific to multiple gest, third trimester, fetus 5 | Pregnancy | O318X35 |
| O31.8X39 | DX10 | Oth comp specific to multiple gest, third trimester, oth | Pregnancy | O318X39 |
| O31.8X90 | DX10 | Oth comp specific to multiple gest, unsp trimester, unsp | Pregnancy | O318X90 |
| O31.8X91 | DX10 | Oth comp specific to multiple gest, unsp trimester, fetus 1 | Pregnancy | O318X91 |
| O31.8X92 | DX10 | Oth comp specific to multiple gest, unsp trimester, fetus 2 | Pregnancy | O318X92 |
| O31.8X93 | DX10 | Oth comp specific to multiple gest, unsp trimester, fetus 3 | Pregnancy | O318X93 |
| O31.8X94 | DX10 | Oth comp specific to multiple gest, unsp trimester, fetus 4 | Pregnancy | O318X94 |
| O31.8X95 | DX10 | Oth comp specific to multiple gest, unsp trimester, fetus 5 | Pregnancy | O318X95 |
| O31.8X99 | DX10 | Oth comp specific to multiple gestation, unsp trimester, oth | Pregnancy | O318X99 |
| O32.0XX0 | DX10 | Maternal care for unstable lie, not applicable or unsp | Pregnancy | O320XX0 |
| O32.0XX1 | DX10 | Maternal care for unstable lie, fetus 1 | Pregnancy | O320XX1 |
| O32.0XX2 | DX10 | Maternal care for unstable lie, fetus 2 | Pregnancy | O320XX2 |
| O32.0XX3 | DX10 | Maternal care for unstable lie, fetus 3 | Pregnancy | O320XX3 |
| O32.0XX4 | DX10 | Maternal care for unstable lie, fetus 4 | Pregnancy | O320XX4 |
| O32.0XX5 | DX10 | Maternal care for unstable lie, fetus 5 | Pregnancy | O320XX5 |
| O32.0XX9 | DX10 | Maternal care for unstable lie, other fetus | Pregnancy | O320XX9 |
| O32.1XX0 | DX10 | Maternal care for breech presentation, unsp | Pregnancy | O321XX0 |
| O32.1XX1 | DX10 | Maternal care for breech presentation, fetus 1 | Pregnancy | O321XX1 |
| O32.1XX2 | DX10 | Maternal care for breech presentation, fetus 2 | Pregnancy | O321XX2 |
| O32.1XX3 | DX10 | Maternal care for breech presentation, fetus 3 | Pregnancy | O321XX3 |
| O32.1XX4 | DX10 | Maternal care for breech presentation, fetus 4 | Pregnancy | O321XX4 |
| O32.1XX5 | DX10 | Maternal care for breech presentation, fetus 5 | Pregnancy | O321XX5 |
| O32.1XX9 | DX10 | Maternal care for breech presentation, other fetus | Pregnancy | O321XX9 |
| O32.2XX0 | DX10 | Maternal care for transverse and oblique lie, unsp | Pregnancy | O322XX0 |
| O32.2XX1 | DX10 | Maternal care for transverse and oblique lie, fetus 1 | Pregnancy | O322XX1 |
| O32.2XX2 | DX10 | Maternal care for transverse and oblique lie, fetus 2 | Pregnancy | O322XX2 |
| O32.2XX3 | DX10 | Maternal care for transverse and oblique lie, fetus 3 | Pregnancy | O322XX3 |
| O32.2XX4 | DX10 | Maternal care for transverse and oblique lie, fetus 4 | Pregnancy | O322XX4 |
| O32.2XX5 | DX10 | Maternal care for transverse and oblique lie, fetus 5 | Pregnancy | O322XX5 |
| O32.2XX9 | DX10 | Maternal care for transverse and oblique lie, other fetus | Pregnancy | O322XX9 |
| O32.3XX0 | DX10 | Maternal care for face, brow and chin presentation, unsp | Pregnancy | O323XX0 |
| O32.3XX1 | DX10 | Maternal care for face, brow and chin presentation, fetus 1 | Pregnancy | O323XX1 |
| O32.3XX2 | DX10 | Maternal care for face, brow and chin presentation, fetus 2 | Pregnancy | O323XX2 |
| O32.3XX3 | DX10 | Maternal care for face, brow and chin presentation, fetus 3 | Pregnancy | O323XX3 |
| O32.3XX4 | DX10 | Maternal care for face, brow and chin presentation, fetus 4 | Pregnancy | O323XX4 |
| O32.3XX5 | DX10 | Maternal care for face, brow and chin presentation, fetus 5 | Pregnancy | O323XX5 |
| O32.3XX9 | DX10 | Maternal care for face, brow and chin presentation, oth | Pregnancy | O323XX9 |
| O32.4XX0 | DX10 | Maternal care for high head at term, not applicable or unsp | Pregnancy | O324XX0 |
| O32.4XX1 | DX10 | Maternal care for high head at term, fetus 1 | Pregnancy | O324XX1 |
| O32.4XX2 | DX10 | Maternal care for high head at term, fetus 2 | Pregnancy | O324XX2 |
| O32.4XX3 | DX10 | Maternal care for high head at term, fetus 3 | Pregnancy | O324XX3 |
| O32.4XX4 | DX10 | Maternal care for high head at term, fetus 4 | Pregnancy | O324XX4 |
| O32.4XX5 | DX10 | Maternal care for high head at term, fetus 5 | Pregnancy | O324XX5 |
| O32.4XX9 | DX10 | Maternal care for high head at term, other fetus | Pregnancy | O324XX9 |
| O32.6XX0 | DX10 | Maternal care for compound presentation, unsp | Pregnancy | O326XX0 |

| | | | | |
|---|---|---|---|---|
| O32.6XX1 | DX10 | Maternal care for compound presentation, fetus 1 | Pregnancy | O326XX1 |
| O32.6XX2 | DX10 | Maternal care for compound presentation, fetus 2 | Pregnancy | O326XX2 |
| O32.6XX3 | DX10 | Maternal care for compound presentation, fetus 3 | Pregnancy | O326XX3 |
| O32.6XX4 | DX10 | Maternal care for compound presentation, fetus 4 | Pregnancy | O326XX4 |
| O32.6XX5 | DX10 | Maternal care for compound presentation, fetus 5 | Pregnancy | O326XX5 |
| O32.6XX9 | DX10 | Maternal care for compound presentation, other fetus | Pregnancy | O326XX9 |
| O32.8XX0 | DX10 | Maternal care for oth malpresentation of fetus, unsp | Pregnancy | O328XX0 |
| O32.8XX1 | DX10 | Maternal care for other malpresentation of fetus, fetus 1 | Pregnancy | O328XX1 |
| O32.8XX2 | DX10 | Maternal care for other malpresentation of fetus, fetus 2 | Pregnancy | O328XX2 |
| O32.8XX3 | DX10 | Maternal care for other malpresentation of fetus, fetus 3 | Pregnancy | O328XX3 |
| O32.8XX4 | DX10 | Maternal care for other malpresentation of fetus, fetus 4 | Pregnancy | O328XX4 |
| O32.8XX5 | DX10 | Maternal care for other malpresentation of fetus, fetus 5 | Pregnancy | O328XX5 |
| O32.8XX9 | DX10 | Maternal care for oth malpresentation of fetus, other fetus | Pregnancy | O328XX9 |
| O32.9XX0 | DX10 | Maternal care for malpresentation of fetus, unsp, unsp | Pregnancy | O329XX0 |
| O32.9XX1 | DX10 | Maternal care for malpresentation of fetus, unsp, fetus 1 | Pregnancy | O329XX1 |
| O32.9XX2 | DX10 | Maternal care for malpresentation of fetus, unsp, fetus 2 | Pregnancy | O329XX2 |
| O32.9XX3 | DX10 | Maternal care for malpresentation of fetus, unsp, fetus 3 | Pregnancy | O329XX3 |
| O32.9XX4 | DX10 | Maternal care for malpresentation of fetus, unsp, fetus 4 | Pregnancy | O329XX4 |
| O32.9XX5 | DX10 | Maternal care for malpresentation of fetus, unsp, fetus 5 | Pregnancy | O329XX5 |
| O32.9XX9 | DX10 | Maternal care for malpresentation of fetus, unsp, oth fetus | Pregnancy | O329XX9 |
| O33.0 | DX10 | Matern care for disproprtn d/t deformity of matern pelv bone | Pregnancy | O330 |
| O33.1 | DX10 | Matern care for disproprtn d/t generally contracted pelvis | Pregnancy | O331 |
| O33.2 | DX10 | Maternal care for disproprtn due to inlet contrctn of pelvis | Pregnancy | O332 |
| O33.3XX0 | DX10 | Matern care for disproprtn d/t outlet contrctn of pelv, unsp | Pregnancy | O333XX0 |
| O33.3XX1 | DX10 | Matern care for disproprtn d/t outlet contrctn of pelv, fts1 | Pregnancy | O333XX1 |
| O33.3XX2 | DX10 | Matern care for disproprtn d/t outlet contrctn of pelv, fts2 | Pregnancy | O333XX2 |
| O33.3XX3 | DX10 | Matern care for disproprtn d/t outlet contrctn of pelv, fts3 | Pregnancy | O333XX3 |
| O33.3XX4 | DX10 | Matern care for disproprtn d/t outlet contrctn of pelv, fts4 | Pregnancy | O333XX4 |
| O33.3XX5 | DX10 | Matern care for disproprtn d/t outlet contrctn of pelv, fts5 | Pregnancy | O333XX5 |
| O33.3XX9 | DX10 | Matern care for disproprtn d/t outlet contrctn of pelv, oth | Pregnancy | O333XX9 |
| O33.4XX0 | DX10 | Matern care for disproprtn of mix matern & fetl origin, unsp | Pregnancy | O334XX0 |
| O33.4XX1 | DX10 | Matern care for disproprtn of mix matern & fetl origin, fts1 | Pregnancy | O334XX1 |
| O33.4XX2 | DX10 | Matern care for disproprtn of mix matern & fetl origin, fts2 | Pregnancy | O334XX2 |
| O33.4XX3 | DX10 | Matern care for disproprtn of mix matern & fetl origin, fts3 | Pregnancy | O334XX3 |
| O33.4XX4 | DX10 | Matern care for disproprtn of mix matern & fetl origin, fts4 | Pregnancy | O334XX4 |
| O33.4XX5 | DX10 | Matern care for disproprtn of mix matern & fetl origin, fts5 | Pregnancy | O334XX5 |
| O33.4XX9 | DX10 | Matern care for disproprtn of mix matern & fetl origin, oth | Pregnancy | O334XX9 |
| O33.5XX0 | DX10 | Matern care for disproprtn d/t unusually large fetus, unsp | Pregnancy | O335XX0 |
| O33.5XX1 | DX10 | Matern care for disproprtn d/t unusually large fetus, fts1 | Pregnancy | O335XX1 |
| O33.5XX2 | DX10 | Matern care for disproprtn d/t unusually large fetus, fts2 | Pregnancy | O335XX2 |

| | | | | |
|---|---|---|---|---|
| O33.5XX3 | DX10 | Matern care for disproprtn d/t unusually large fetus, fts3 | Pregnancy | O335XX3 |
| O33.5XX4 | DX10 | Matern care for disproprtn d/t unusually large fetus, fts4 | Pregnancy | O335XX4 |
| O33.5XX5 | DX10 | Matern care for disproprtn d/t unusually large fetus, fts5 | Pregnancy | O335XX5 |
| O33.5XX9 | DX10 | Matern care for disproprtn due to unusually large fetus, oth | Pregnancy | O335XX9 |
| O33.6XX0 | DX10 | Matern care for disproprtn due to hydrocephalic fetus, unsp | Pregnancy | O336XX0 |
| O33.6XX1 | DX10 | Matern care for disproprtn due to hydrocephalic fetus, fts1 | Pregnancy | O336XX1 |
| O33.6XX2 | DX10 | Matern care for disproprtn due to hydrocephalic fetus, fts2 | Pregnancy | O336XX2 |
| O33.6XX3 | DX10 | Matern care for disproprtn due to hydrocephalic fetus, fts3 | Pregnancy | O336XX3 |
| O33.6XX4 | DX10 | Matern care for disproprtn due to hydrocephalic fetus, fts4 | Pregnancy | O336XX4 |
| O33.6XX5 | DX10 | Matern care for disproprtn due to hydrocephalic fetus, fts5 | Pregnancy | O336XX5 |
| O33.6XX9 | DX10 | Maternal care for disproprtn due to hydrocephalic fetus, oth | Pregnancy | O336XX9 |
| O33.7 | DX10 | Maternal care for disproportion due to oth fetal deformities | Pregnancy | O337 |
| O33.7XX0 | DX10 | Maternal care for disproportion due to other fetal deformities, not applicable or unspecified | Pregnancy | O337XX0 |
| O33.7XX1 | DX10 | Maternal care for disproportion due to other fetal deformities, fetus 1 | Pregnancy | O337XX1 |
| O33.7XX2 | DX10 | Maternal care for disproportion due to other fetal deformities, fetus 2 | Pregnancy | O337XX2 |
| O33.7XX3 | DX10 | Maternal care for disproportion due to other fetal deformities, fetus 3 | Pregnancy | O337XX3 |
| O33.7XX4 | DX10 | Maternal care for disproportion due to other fetal deformities, fetus 4 | Pregnancy | O337XX4 |
| O33.8 | DX10 | Maternal care for disproportion of other origin | Pregnancy | O338 |
| O33.9 | DX10 | Maternal care for disproportion, unspecified | Pregnancy | O339 |
| O337.XX5 | DX10 | Maternal care for disproportion due to other fetal deformities, fetus 5 | Pregnancy | O337XX5 |
| O337.XX9 | DX10 | Maternal care for disproportion due to other fetal deformities, other fetus | Pregnancy | O337XX9 |
| O34.00 | DX10 | Maternal care for unsp congen malform of uterus, unsp tri | Pregnancy | O3400 |
| O34.01 | DX10 | Maternal care for unsp congen malform of uterus, first tri | Pregnancy | O3401 |
| O34.02 | DX10 | Maternal care for unsp congen malform of uterus, second tri | Pregnancy | O3402 |
| O34.03 | DX10 | Maternal care for unsp congen malform of uterus, third tri | Pregnancy | O3403 |
| O34.10 | DX10 | Maternal care for benign tumor of corpus uteri, unsp tri | Pregnancy | O3410 |
| O34.11 | DX10 | Maternal care for benign tumor of corpus uteri, first tri | Pregnancy | O3411 |
| O34.12 | DX10 | Maternal care for benign tumor of corpus uteri, second tri | Pregnancy | O3412 |
| O34.13 | DX10 | Maternal care for benign tumor of corpus uteri, third tri | Pregnancy | O3413 |
| O34.21 | DX10 | Maternal care for scar from previous cesarean delivery | Pregnancy | O3421 |
| O34.211 | DX10 | Maternal care for low transverse scar from previous cesarean delivery | Pregnancy | O34211 |
| O34.212 | DX10 | Maternal care for vertical scar from previous cesarean delivery | Pregnancy | O34212 |
| O34.219 | DX10 | MATERNAL CARE FOR UNSPECIFIED TYPE SCAR FROM PREV CESAREAN DEL | Pregnancy | O34219 |
| O34.29 | DX10 | Maternal care due to uterine scar from oth previous surgery | Pregnancy | O3429 |
| O34.30 | DX10 | Maternal care for cervical incompetence, unsp trimester | Pregnancy | O3430 |

| | | | | |
|---|---|---|---|---|
| O34.31 | DX10 | Maternal care for cervical incompetence, first trimester | Pregnancy | O3431 |
| O34.32 | DX10 | Maternal care for cervical incompetence, second trimester | Pregnancy | O3432 |
| O34.33 | DX10 | Maternal care for cervical incompetence, third trimester | Pregnancy | O3433 |
| O34.40 | DX10 | Maternal care for oth abnlt of cervix, unsp trimester | Pregnancy | O3440 |
| O34.41 | DX10 | Maternal care for oth abnlt of cervix, first trimester | Pregnancy | O3441 |
| O34.42 | DX10 | Maternal care for oth abnlt of cervix, second trimester | Pregnancy | O3442 |
| O34.43 | DX10 | Maternal care for oth abnlt of cervix, third trimester | Pregnancy | O3443 |
| O34.511 | DX10 | Maternal care for incarceration of gravid uterus, first tri | Pregnancy | O34511 |
| O34.512 | DX10 | Maternal care for incarceration of gravid uterus, second tri | Pregnancy | O34512 |
| O34.513 | DX10 | Maternal care for incarceration of gravid uterus, third tri | Pregnancy | O34513 |
| O34.519 | DX10 | Maternal care for incarceration of gravid uterus, unsp tri | Pregnancy | O34519 |
| O34.521 | DX10 | Maternal care for prolapse of gravid uterus, first trimester | Pregnancy | O34521 |
| O34.522 | DX10 | Maternal care for prolapse of gravid uterus, second tri | Pregnancy | O34522 |
| O34.523 | DX10 | Maternal care for prolapse of gravid uterus, third trimester | Pregnancy | O34523 |
| O34.529 | DX10 | Maternal care for prolapse of gravid uterus, unsp trimester | Pregnancy | O34529 |
| O34.531 | DX10 | Maternal care for retroversion of gravid uterus, first tri | Pregnancy | O34531 |
| O34.532 | DX10 | Maternal care for retroversion of gravid uterus, second tri | Pregnancy | O34532 |
| O34.533 | DX10 | Maternal care for retroversion of gravid uterus, third tri | Pregnancy | O34533 |
| O34.539 | DX10 | Maternal care for retroversion of gravid uterus, unsp tri | Pregnancy | O34539 |
| O34.591 | DX10 | Maternal care for oth abnlt of gravid uterus, first tri | Pregnancy | O34591 |
| O34.592 | DX10 | Maternal care for oth abnlt of gravid uterus, second tri | Pregnancy | O34592 |
| O34.593 | DX10 | Maternal care for oth abnlt of gravid uterus, third tri | Pregnancy | O34593 |
| O34.599 | DX10 | Maternal care for oth abnlt of gravid uterus, unsp trimester | Pregnancy | O34599 |
| O34.60 | DX10 | Maternal care for abnormality of vagina, unsp trimester | Pregnancy | O3460 |
| O34.61 | DX10 | Maternal care for abnormality of vagina, first trimester | Pregnancy | O3461 |
| O34.62 | DX10 | Maternal care for abnormality of vagina, second trimester | Pregnancy | O3462 |
| O34.63 | DX10 | Maternal care for abnormality of vagina, third trimester | Pregnancy | O3463 |
| O34.70 | DX10 | Maternal care for abnlt of vulva and perineum, unsp tri | Pregnancy | O3470 |
| O34.71 | DX10 | Maternal care for abnlt of vulva and perineum, first tri | Pregnancy | O3471 |
| O34.72 | DX10 | Maternal care for abnlt of vulva and perineum, second tri | Pregnancy | O3472 |
| O34.73 | DX10 | Maternal care for abnlt of vulva and perineum, third tri | Pregnancy | O3473 |
| O34.80 | DX10 | Maternal care for oth abnlt of pelvic organs, unsp trimester | Pregnancy | O3480 |
| O34.81 | DX10 | Maternal care for oth abnlt of pelvic organs, first tri | Pregnancy | O3481 |
| O34.82 | DX10 | Maternal care for oth abnlt of pelvic organs, second tri | Pregnancy | O3482 |
| O34.83 | DX10 | Maternal care for oth abnlt of pelvic organs, third tri | Pregnancy | O3483 |
| O34.90 | DX10 | Maternal care for abnlt of pelvic organ, unsp, unsp tri | Pregnancy | O3490 |
| O34.91 | DX10 | Maternal care for abnlt of pelvic organ, unsp, first tri | Pregnancy | O3491 |
| O34.92 | DX10 | Maternal care for abnlt of pelvic organ, unsp, second tri | Pregnancy | O3492 |
| O34.93 | DX10 | Maternal care for abnlt of pelvic organ, unsp, third tri | Pregnancy | O3493 |
| O35.0XX0 | DX10 | Maternal care for (suspected) cnsl malform in fetus, unsp | Pregnancy | O350XX0 |
| O35.0XX1 | DX10 | Maternal care for (suspected) cnsl malform in fetus, fetus 1 | Pregnancy | O350XX1 |
| O35.0XX2 | DX10 | Maternal care for (suspected) cnsl malform in fetus, fetus 2 | Pregnancy | O350XX2 |
| O35.0XX3 | DX10 | Maternal care for (suspected) cnsl malform in fetus, fetus 3 | Pregnancy | O350XX3 |
| O35.0XX4 | DX10 | Maternal care for (suspected) cnsl malform in fetus, fetus 4 | Pregnancy | O350XX4 |
| O35.0XX5 | DX10 | Maternal care for (suspected) cnsl malform in fetus, fetus 5 | Pregnancy | O350XX5 |

| | | | | |
|---|---|---|---|---|
| O35.0XX9 | DX10 | Maternal care for (suspected) cnsl malform in fetus, oth | Pregnancy | O350XX9 |
| O35.1XX0 | DX10 | Maternal care for chromosomal abnormality in fetus, unsp | Pregnancy | O351XX0 |
| O35.1XX1 | DX10 | Maternal care for chromosomal abnormality in fetus, fetus 1 | Pregnancy | O351XX1 |
| O35.1XX2 | DX10 | Maternal care for chromosomal abnormality in fetus, fetus 2 | Pregnancy | O351XX2 |
| O35.1XX3 | DX10 | Maternal care for chromosomal abnormality in fetus, fetus 3 | Pregnancy | O351XX3 |
| O35.1XX4 | DX10 | Maternal care for chromosomal abnormality in fetus, fetus 4 | Pregnancy | O351XX4 |
| O35.1XX5 | DX10 | Maternal care for chromosomal abnormality in fetus, fetus 5 | Pregnancy | O351XX5 |
| O35.1XX9 | DX10 | Maternal care for chromosomal abnormality in fetus, oth | Pregnancy | O351XX9 |
| O35.2XX0 | DX10 | Maternal care for hereditary disease in fetus, unsp | Pregnancy | O352XX0 |
| O35.2XX1 | DX10 | Maternal care for hereditary disease in fetus, fetus 1 | Pregnancy | O352XX1 |
| O35.2XX2 | DX10 | Maternal care for hereditary disease in fetus, fetus 2 | Pregnancy | O352XX2 |
| O35.2XX3 | DX10 | Maternal care for hereditary disease in fetus, fetus 3 | Pregnancy | O352XX3 |
| O35.2XX4 | DX10 | Maternal care for hereditary disease in fetus, fetus 4 | Pregnancy | O352XX4 |
| O35.2XX5 | DX10 | Maternal care for hereditary disease in fetus, fetus 5 | Pregnancy | O352XX5 |
| O35.2XX9 | DX10 | Maternal care for hereditary disease in fetus, oth | Pregnancy | O352XX9 |
| O35.3XX0 | DX10 | Matern care for damag to fts from viral dis in mother, unsp | Pregnancy | O353XX0 |
| O35.3XX1 | DX10 | Matern care for damag to fts from viral dis in mother, fts1 | Pregnancy | O353XX1 |
| O35.3XX2 | DX10 | Matern care for damag to fts from viral dis in mother, fts2 | Pregnancy | O353XX2 |
| O35.3XX3 | DX10 | Matern care for damag to fts from viral dis in mother, fts3 | Pregnancy | O353XX3 |
| O35.3XX4 | DX10 | Matern care for damag to fts from viral dis in mother, fts4 | Pregnancy | O353XX4 |
| O35.3XX5 | DX10 | Matern care for damag to fts from viral dis in mother, fts5 | Pregnancy | O353XX5 |
| O35.3XX9 | DX10 | Matern care for damag to fetus from viral dis in mother, oth | Pregnancy | O353XX9 |
| O35.4XX0 | DX10 | Maternal care for damage to fetus from alcohol, unsp | Pregnancy | O354XX0 |
| O35.4XX1 | DX10 | Maternal care for damage to fetus from alcohol, fetus 1 | Pregnancy | O354XX1 |
| O35.4XX2 | DX10 | Maternal care for damage to fetus from alcohol, fetus 2 | Pregnancy | O354XX2 |
| O35.4XX3 | DX10 | Maternal care for damage to fetus from alcohol, fetus 3 | Pregnancy | O354XX3 |
| O35.4XX4 | DX10 | Maternal care for damage to fetus from alcohol, fetus 4 | Pregnancy | O354XX4 |
| O35.4XX5 | DX10 | Maternal care for damage to fetus from alcohol, fetus 5 | Pregnancy | O354XX5 |
| O35.4XX9 | DX10 | Maternal care for damage to fetus from alcohol, oth | Pregnancy | O354XX9 |
| O35.5XX0 | DX10 | Maternal care for (suspected) damage to fetus by drugs, unsp | Pregnancy | O355XX0 |
| O35.5XX1 | DX10 | Maternal care for damage to fetus by drugs, fetus 1 | Pregnancy | O355XX1 |
| O35.5XX2 | DX10 | Maternal care for damage to fetus by drugs, fetus 2 | Pregnancy | O355XX2 |
| O35.5XX3 | DX10 | Maternal care for damage to fetus by drugs, fetus 3 | Pregnancy | O355XX3 |
| O35.5XX4 | DX10 | Maternal care for damage to fetus by drugs, fetus 4 | Pregnancy | O355XX4 |
| O35.5XX5 | DX10 | Maternal care for damage to fetus by drugs, fetus 5 | Pregnancy | O355XX5 |
| O35.5XX9 | DX10 | Maternal care for (suspected) damage to fetus by drugs, oth | Pregnancy | O355XX9 |
| O35.6XX0 | DX10 | Maternal care for damage to fetus by radiation, unsp | Pregnancy | O356XX0 |
| O35.6XX1 | DX10 | Maternal care for damage to fetus by radiation, fetus 1 | Pregnancy | O356XX1 |
| O35.6XX2 | DX10 | Maternal care for damage to fetus by radiation, fetus 2 | Pregnancy | O356XX2 |
| O35.6XX3 | DX10 | Maternal care for damage to fetus by radiation, fetus 3 | Pregnancy | O356XX3 |
| O35.6XX4 | DX10 | Maternal care for damage to fetus by radiation, fetus 4 | Pregnancy | O356XX4 |
| O35.6XX5 | DX10 | Maternal care for damage to fetus by radiation, fetus 5 | Pregnancy | O356XX5 |
| O35.6XX9 | DX10 | Maternal care for damage to fetus by radiation, oth | Pregnancy | O356XX9 |
| O35.7XX0 | DX10 | Maternal care for damage to fetus by oth medical proc, unsp | Pregnancy | O357XX0 |
| O35.7XX1 | DX10 | Matern care for damage to fetus by oth medical proc, fetus 1 | Pregnancy | O357XX1 |

| | | | | |
|---|---|---|---|---|
| O35.7XX2 | DX10 | Matern care for damage to fetus by oth medical proc, fetus 2 | Pregnancy | O357XX2 |
| O35.7XX3 | DX10 | Matern care for damage to fetus by oth medical proc, fetus 3 | Pregnancy | O357XX3 |
| O35.7XX4 | DX10 | Matern care for damage to fetus by oth medical proc, fetus 4 | Pregnancy | O357XX4 |
| O35.7XX5 | DX10 | Matern care for damage to fetus by oth medical proc, fetus 5 | Pregnancy | O357XX5 |
| O35.7XX9 | DX10 | Maternal care for damage to fetus by oth medical proc, oth | Pregnancy | O357XX9 |
| O35.8XX0 | DX10 | Maternal care for oth fetal abnormality and damage, unsp | Pregnancy | O358XX0 |
| O35.8XX1 | DX10 | Maternal care for oth fetal abnormality and damage, fetus 1 | Pregnancy | O358XX1 |
| O35.8XX2 | DX10 | Maternal care for oth fetal abnormality and damage, fetus 2 | Pregnancy | O358XX2 |
| O35.8XX3 | DX10 | Maternal care for oth fetal abnormality and damage, fetus 3 | Pregnancy | O358XX3 |
| O35.8XX4 | DX10 | Maternal care for oth fetal abnormality and damage, fetus 4 | Pregnancy | O358XX4 |
| O35.8XX5 | DX10 | Maternal care for oth fetal abnormality and damage, fetus 5 | Pregnancy | O358XX5 |
| O35.8XX9 | DX10 | Maternal care for oth fetal abnormality and damage, oth | Pregnancy | O358XX9 |
| O35.9XX0 | DX10 | Maternal care for fetal abnormality and damage, unsp, unsp | Pregnancy | O359XX0 |
| O35.9XX1 | DX10 | Maternal care for fetal abnlt and damage, unsp, fetus 1 | Pregnancy | O359XX1 |
| O35.9XX2 | DX10 | Maternal care for fetal abnlt and damage, unsp, fetus 2 | Pregnancy | O359XX2 |
| O35.9XX3 | DX10 | Maternal care for fetal abnlt and damage, unsp, fetus 3 | Pregnancy | O359XX3 |
| O35.9XX4 | DX10 | Maternal care for fetal abnlt and damage, unsp, fetus 4 | Pregnancy | O359XX4 |
| O35.9XX5 | DX10 | Maternal care for fetal abnlt and damage, unsp, fetus 5 | Pregnancy | O359XX5 |
| O35.9XX9 | DX10 | Maternal care for fetal abnormality and damage, unsp, oth | Pregnancy | O359XX9 |
| O36.0110 | DX10 | Maternal care for anti-D antibodies, first trimester, unsp | Pregnancy | O360110 |
| O36.0111 | DX10 | Maternal care for anti-D antibodies, first tri, fetus 1 | Pregnancy | O360111 |
| O36.0112 | DX10 | Maternal care for anti-D antibodies, first tri, fetus 2 | Pregnancy | O3610112 |
| O36.0113 | DX10 | Maternal care for anti-D antibodies, first tri, fetus 3 | Pregnancy | O360113 |
| O36.0114 | DX10 | Maternal care for anti-D antibodies, first tri, fetus 4 | Pregnancy | O360114 |
| O36.0115 | DX10 | Maternal care for anti-D antibodies, first tri, fetus 5 | Pregnancy | O360115 |
| O36.0119 | DX10 | Maternal care for anti-D antibodies, first trimester, oth | Pregnancy | O360119 |
| O36.0120 | DX10 | Maternal care for anti-D antibodies, second trimester, unsp | Pregnancy | O360120 |
| O36.0121 | DX10 | Maternal care for anti-D antibodies, second tri, fetus 1 | Pregnancy | O360121 |
| O36.0122 | DX10 | Maternal care for anti-D antibodies, second tri, fetus 2 | Pregnancy | O360122 |
| O36.0123 | DX10 | Maternal care for anti-D antibodies, second tri, fetus 3 | Pregnancy | O360123 |
| O36.0124 | DX10 | Maternal care for anti-D antibodies, second tri, fetus 4 | Pregnancy | O360124 |
| O36.0125 | DX10 | Maternal care for anti-D antibodies, second tri, fetus 5 | Pregnancy | O360125 |
| O36.0129 | DX10 | Maternal care for anti-D antibodies, second trimester, oth | Pregnancy | O360129 |
| O36.0130 | DX10 | Maternal care for anti-D antibodies, third trimester, unsp | Pregnancy | O360130 |
| O36.0131 | DX10 | Maternal care for anti-D antibodies, third tri, fetus 1 | Pregnancy | O360131 |
| O36.0132 | DX10 | Maternal care for anti-D antibodies, third tri, fetus 2 | Pregnancy | O360132 |
| O36.0133 | DX10 | Maternal care for anti-D antibodies, third tri, fetus 3 | Pregnancy | O360133 |
| O36.0134 | DX10 | Maternal care for anti-D antibodies, third tri, fetus 4 | Pregnancy | O360134 |
| O36.0135 | DX10 | Maternal care for anti-D antibodies, third tri, fetus 5 | Pregnancy | O360135 |
| O36.0139 | DX10 | Maternal care for anti-D antibodies, third trimester, oth | Pregnancy | O360139 |
| O36.0190 | DX10 | Maternal care for anti-D antibodies, unsp trimester, unsp | Pregnancy | O360190 |
| O36.0191 | DX10 | Maternal care for anti-D antibodies, unsp trimester, fetus 1 | Pregnancy | O360191 |

| | | | | |
|---|---|---|---|---|
| O36.0192 | DX10 | Maternal care for anti-D antibodies, unsp trimester, fetus 2 | Pregnancy | O360192 |
| O36.0193 | DX10 | Maternal care for anti-D antibodies, unsp trimester, fetus 3 | Pregnancy | O360193 |
| O36.0194 | DX10 | Maternal care for anti-D antibodies, unsp trimester, fetus 4 | Pregnancy | O360194 |
| O36.0195 | DX10 | Maternal care for anti-D antibodies, unsp trimester, fetus 5 | Pregnancy | O360195 |
| O36.0199 | DX10 | Maternal care for anti-D antibodies, unsp trimester, oth | Pregnancy | O360199 |
| O36.0910 | DX10 | Maternal care for oth rhesus isoimmun, first trimester, unsp | Pregnancy | O360910 |
| O36.0911 | DX10 | Maternal care for oth rhesus isoimmun, first tri, fetus 1 | Pregnancy | O360911 |
| O36.0912 | DX10 | Maternal care for oth rhesus isoimmun, first tri, fetus 2 | Pregnancy | O360912 |
| O36.0913 | DX10 | Maternal care for oth rhesus isoimmun, first tri, fetus 3 | Pregnancy | O360913 |
| O36.0914 | DX10 | Maternal care for oth rhesus isoimmun, first tri, fetus 4 | Pregnancy | O360914 |
| O36.0915 | DX10 | Maternal care for oth rhesus isoimmun, first tri, fetus 5 | Pregnancy | O360915 |
| O36.0919 | DX10 | Maternal care for oth rhesus isoimmun, first trimester, oth | Pregnancy | O360919 |
| O36.0920 | DX10 | Maternal care for oth rhesus isoimmun, second tri, unsp | Pregnancy | O360920 |
| O36.0921 | DX10 | Maternal care for oth rhesus isoimmun, second tri, fetus 1 | Pregnancy | O360921 |
| O36.0922 | DX10 | Maternal care for oth rhesus isoimmun, second tri, fetus 2 | Pregnancy | O360922 |
| O36.0923 | DX10 | Maternal care for oth rhesus isoimmun, second tri, fetus 3 | Pregnancy | O360923 |
| O36.0924 | DX10 | Maternal care for oth rhesus isoimmun, second tri, fetus 4 | Pregnancy | O360924 |
| O36.0925 | DX10 | Maternal care for oth rhesus isoimmun, second tri, fetus 5 | Pregnancy | O360925 |
| O36.0929 | DX10 | Maternal care for oth rhesus isoimmun, second trimester, oth | Pregnancy | O360929 |
| O36.0930 | DX10 | Maternal care for oth rhesus isoimmun, third trimester, unsp | Pregnancy | O360930 |
| O36.0931 | DX10 | Maternal care for oth rhesus isoimmun, third tri, fetus 1 | Pregnancy | O360931 |
| O36.0932 | DX10 | Maternal care for oth rhesus isoimmun, third tri, fetus 2 | Pregnancy | O360932 |
| O36.0933 | DX10 | Maternal care for oth rhesus isoimmun, third tri, fetus 3 | Pregnancy | O360933 |
| O36.0934 | DX10 | Maternal care for oth rhesus isoimmun, third tri, fetus 4 | Pregnancy | O360934 |
| O36.0935 | DX10 | Maternal care for oth rhesus isoimmun, third tri, fetus 5 | Pregnancy | O360935 |
| O36.0939 | DX10 | Maternal care for oth rhesus isoimmun, third trimester, oth | Pregnancy | O360939 |
| O36.0990 | DX10 | Maternal care for oth rhesus isoimmun, unsp trimester, unsp | Pregnancy | O360990 |
| O36.0991 | DX10 | Maternal care for oth rhesus isoimmun, unsp tri, fetus 1 | Pregnancy | O360991 |
| O36.0992 | DX10 | Maternal care for oth rhesus isoimmun, unsp tri, fetus 2 | Pregnancy | O360992 |
| O36.0993 | DX10 | Maternal care for oth rhesus isoimmun, unsp tri, fetus 3 | Pregnancy | O360993 |
| O36.0994 | DX10 | Maternal care for oth rhesus isoimmun, unsp tri, fetus 4 | Pregnancy | O360994 |
| O36.0995 | DX10 | Maternal care for oth rhesus isoimmun, unsp tri, fetus 5 | Pregnancy | O360995 |
| O36.0999 | DX10 | Maternal care for oth rhesus isoimmun, unsp trimester, oth | Pregnancy | O360999 |
| O36.1110 | DX10 | Maternal care for Anti-A sensitization, first tri, unsp | Pregnancy | O361110 |
| O36.1111 | DX10 | Maternal care for Anti-A sensitization, first tri, fetus 1 | Pregnancy | O361111 |
| O36.1112 | DX10 | Maternal care for Anti-A sensitization, first tri, fetus 2 | Pregnancy | O361112 |
| O36.1113 | DX10 | Maternal care for Anti-A sensitization, first tri, fetus 3 | Pregnancy | O361113 |
| O36.1114 | DX10 | Maternal care for Anti-A sensitization, first tri, fetus 4 | Pregnancy | O361114 |
| O36.1115 | DX10 | Maternal care for Anti-A sensitization, first tri, fetus 5 | Pregnancy | O361115 |
| O36.1119 | DX10 | Maternal care for Anti-A sensitization, first trimester, oth | Pregnancy | O361119 |
| O36.1120 | DX10 | Maternal care for Anti-A sensitization, second tri, unsp | Pregnancy | O361120 |
| O36.1121 | DX10 | Maternal care for Anti-A sensitization, second tri, fetus 1 | Pregnancy | O361121 |
| O36.1122 | DX10 | Maternal care for Anti-A sensitization, second tri, fetus 2 | Pregnancy | O361122 |

| | | | | |
|---|---|---|---|---|
| O36.1123 | DX10 | Maternal care for Anti-A sensitization, second tri, fetus 3 | Pregnancy | O361123 |
| O36.1124 | DX10 | Maternal care for Anti-A sensitization, second tri, fetus 4 | Pregnancy | O361124 |
| O36.1125 | DX10 | Maternal care for Anti-A sensitization, second tri, fetus 5 | Pregnancy | O361125 |
| O36.1129 | DX10 | Maternal care for Anti-A sensitization, second tri, oth | Pregnancy | O361129 |
| O36.1130 | DX10 | Maternal care for Anti-A sensitization, third tri, unsp | Pregnancy | O361130 |
| O36.1131 | DX10 | Maternal care for Anti-A sensitization, third tri, fetus 1 | Pregnancy | O361131 |
| O36.1132 | DX10 | Maternal care for Anti-A sensitization, third tri, fetus 2 | Pregnancy | O361132 |
| O36.1133 | DX10 | Maternal care for Anti-A sensitization, third tri, fetus 3 | Pregnancy | O361133 |
| O36.1134 | DX10 | Maternal care for Anti-A sensitization, third tri, fetus 4 | Pregnancy | O361134 |
| O36.1135 | DX10 | Maternal care for Anti-A sensitization, third tri, fetus 5 | Pregnancy | O361135 |
| O36.1139 | DX10 | Maternal care for Anti-A sensitization, third trimester, oth | Pregnancy | O361139 |
| O36.1190 | DX10 | Maternal care for Anti-A sensitization, unsp trimester, unsp | Pregnancy | O361190 |
| O36.1191 | DX10 | Maternal care for Anti-A sensitization, unsp tri, fetus 1 | Pregnancy | O361191 |
| O36.1192 | DX10 | Maternal care for Anti-A sensitization, unsp tri, fetus 2 | Pregnancy | O361192 |
| O36.1193 | DX10 | Maternal care for Anti-A sensitization, unsp tri, fetus 3 | Pregnancy | O361193 |
| O36.1194 | DX10 | Maternal care for Anti-A sensitization, unsp tri, fetus 4 | Pregnancy | O361194 |
| O36.1195 | DX10 | Maternal care for Anti-A sensitization, unsp tri, fetus 5 | Pregnancy | O361195 |
| O36.1199 | DX10 | Maternal care for Anti-A sensitization, unsp trimester, oth | Pregnancy | O361199 |
| O36.1910 | DX10 | Maternal care for oth isoimmunization, first trimester, unsp | Pregnancy | O361910 |
| O36.1911 | DX10 | Maternal care for oth isoimmun, first trimester, fetus 1 | Pregnancy | O361911 |
| O36.1912 | DX10 | Maternal care for oth isoimmun, first trimester, fetus 2 | Pregnancy | O361912 |
| O36.1913 | DX10 | Maternal care for oth isoimmun, first trimester, fetus 3 | Pregnancy | O361913 |
| O36.1914 | DX10 | Maternal care for oth isoimmun, first trimester, fetus 4 | Pregnancy | O361914 |
| O36.1915 | DX10 | Maternal care for oth isoimmun, first trimester, fetus 5 | Pregnancy | O361915 |
| O36.1919 | DX10 | Maternal care for oth isoimmunization, first trimester, oth | Pregnancy | O361919 |
| O36.1920 | DX10 | Maternal care for oth isoimmun, second trimester, unsp | Pregnancy | O361920 |
| O36.1921 | DX10 | Maternal care for oth isoimmun, second trimester, fetus 1 | Pregnancy | O361921 |
| O36.1922 | DX10 | Maternal care for oth isoimmun, second trimester, fetus 2 | Pregnancy | O361922 |
| O36.1923 | DX10 | Maternal care for oth isoimmun, second trimester, fetus 3 | Pregnancy | O361923 |
| O36.1924 | DX10 | Maternal care for oth isoimmun, second trimester, fetus 4 | Pregnancy | O361924 |
| O36.1925 | DX10 | Maternal care for oth isoimmun, second trimester, fetus 5 | Pregnancy | O361925 |
| O36.1929 | DX10 | Maternal care for oth isoimmunization, second trimester, oth | Pregnancy | O361929 |
| O36.1930 | DX10 | Maternal care for oth isoimmunization, third trimester, unsp | Pregnancy | O361930 |
| O36.1931 | DX10 | Maternal care for oth isoimmun, third trimester, fetus 1 | Pregnancy | O361931 |
| O36.1932 | DX10 | Maternal care for oth isoimmun, third trimester, fetus 2 | Pregnancy | O361932 |
| O36.1933 | DX10 | Maternal care for oth isoimmun, third trimester, fetus 3 | Pregnancy | O361933 |
| O36.1934 | DX10 | Maternal care for oth isoimmun, third trimester, fetus 4 | Pregnancy | O361934 |
| O36.1935 | DX10 | Maternal care for oth isoimmun, third trimester, fetus 5 | Pregnancy | O361935 |
| O36.1939 | DX10 | Maternal care for oth isoimmunization, third trimester, oth | Pregnancy | O361939 |
| O36.1990 | DX10 | Maternal care for oth isoimmunization, unsp trimester, unsp | Pregnancy | O361990 |
| O36.1991 | DX10 | Maternal care for oth isoimmun, unsp trimester, fetus 1 | Pregnancy | O361991 |
| O36.1992 | DX10 | Maternal care for oth isoimmun, unsp trimester, fetus 2 | Pregnancy | O361992 |
| O36.1993 | DX10 | Maternal care for oth isoimmun, unsp trimester, fetus 3 | Pregnancy | O361993 |
| O36.1994 | DX10 | Maternal care for oth isoimmun, unsp trimester, fetus 4 | Pregnancy | O361994 |
| O36.1995 | DX10 | Maternal care for oth isoimmun, unsp trimester, fetus 5 | Pregnancy | O361995 |
| O36.1999 | DX10 | Maternal care for oth isoimmunization, unsp trimester, oth | Pregnancy | O361999 |

| | | | | |
|---|---|---|---|---|
| O36.20X0 | DX10 | Maternal care for hydrops fetalis, unsp trimester, unsp | Pregnancy | O3620X0 |
| O36.20X1 | DX10 | Maternal care for hydrops fetalis, unsp trimester, fetus 1 | Pregnancy | O3620X1 |
| O36.20X2 | DX10 | Maternal care for hydrops fetalis, unsp trimester, fetus 2 | Pregnancy | O3620X2 |
| O36.20X3 | DX10 | Maternal care for hydrops fetalis, unsp trimester, fetus 3 | Pregnancy | O3620X3 |
| O36.20X4 | DX10 | Maternal care for hydrops fetalis, unsp trimester, fetus 4 | Pregnancy | O3620X4 |
| O36.20X5 | DX10 | Maternal care for hydrops fetalis, unsp trimester, fetus 5 | Pregnancy | O3620X5 |
| O36.20X9 | DX10 | Maternal care for hydrops fetalis, unsp trimester, oth fetus | Pregnancy | O3620X9 |
| O36.21X0 | DX10 | Maternal care for hydrops fetalis, first trimester, unsp | Pregnancy | O3621X0 |
| O36.21X1 | DX10 | Maternal care for hydrops fetalis, first trimester, fetus 1 | Pregnancy | O3621X1 |
| O36.21X2 | DX10 | Maternal care for hydrops fetalis, first trimester, fetus 2 | Pregnancy | O3621X2 |
| O36.21X3 | DX10 | Maternal care for hydrops fetalis, first trimester, fetus 3 | Pregnancy | O3621X3 |
| O36.21X4 | DX10 | Maternal care for hydrops fetalis, first trimester, fetus 4 | Pregnancy | O3621X4 |
| O36.21X5 | DX10 | Maternal care for hydrops fetalis, first trimester, fetus 5 | Pregnancy | O3621X5 |
| O36.21X9 | DX10 | Maternal care for hydrops fetalis, first trimester, oth | Pregnancy | O3621X9 |
| O36.22X0 | DX10 | Maternal care for hydrops fetalis, second trimester, unsp | Pregnancy | O3622X0 |
| O36.22X1 | DX10 | Maternal care for hydrops fetalis, second trimester, fetus 1 | Pregnancy | O3622X1 |
| O36.22X2 | DX10 | Maternal care for hydrops fetalis, second trimester, fetus 2 | Pregnancy | O3622X2 |
| O36.22X3 | DX10 | Maternal care for hydrops fetalis, second trimester, fetus 3 | Pregnancy | O3622X3 |
| O36.22X4 | DX10 | Maternal care for hydrops fetalis, second trimester, fetus 4 | Pregnancy | O3622X4 |
| O36.22X5 | DX10 | Maternal care for hydrops fetalis, second trimester, fetus 5 | Pregnancy | O3622X5 |
| O36.22X9 | DX10 | Maternal care for hydrops fetalis, second trimester, oth | Pregnancy | O3622X9 |
| O36.23X0 | DX10 | Maternal care for hydrops fetalis, third trimester, unsp | Pregnancy | O3623X0 |
| O36.23X1 | DX10 | Maternal care for hydrops fetalis, third trimester, fetus 1 | Pregnancy | O3623X1 |
| O36.23X2 | DX10 | Maternal care for hydrops fetalis, third trimester, fetus 2 | Pregnancy | O3623X2 |
| O36.23X3 | DX10 | Maternal care for hydrops fetalis, third trimester, fetus 3 | Pregnancy | O3623X3 |
| O36.23X4 | DX10 | Maternal care for hydrops fetalis, third trimester, fetus 4 | Pregnancy | O3623X4 |
| O36.23X5 | DX10 | Maternal care for hydrops fetalis, third trimester, fetus 5 | Pregnancy | O3623X5 |
| O36.23X9 | DX10 | Maternal care for hydrops fetalis, third trimester, oth | Pregnancy | O3623X9 |
| O36.5110 | DX10 | Matern care for known or susp placntl insuff, 1st tri, unsp | Pregnancy | O365110 |
| O36.5111 | DX10 | Matern care for known or susp placntl insuff, 1st tri, fts1 | Pregnancy | O365111 |
| O36.5112 | DX10 | Matern care for known or susp placntl insuff, 1st tri, fts2 | Pregnancy | O365112 |
| O36.5113 | DX10 | Matern care for known or susp placntl insuff, 1st tri, fts3 | Pregnancy | O365113 |
| O36.5114 | DX10 | Matern care for known or susp placntl insuff, 1st tri, fts4 | Pregnancy | O365114 |
| O36.5115 | DX10 | Matern care for known or susp placntl insuff, 1st tri, fts5 | Pregnancy | O365115 |
| O36.5119 | DX10 | Matern care for known or susp placntl insuff, first tri, oth | Pregnancy | O365119 |
| O36.5120 | DX10 | Matern care for known or susp placntl insuff, 2nd tri, unsp | Pregnancy | O365120 |
| O36.5121 | DX10 | Matern care for known or susp placntl insuff, 2nd tri, fts1 | Pregnancy | O365121 |
| O36.5122 | DX10 | Matern care for known or susp placntl insuff, 2nd tri, fts2 | Pregnancy | O365122 |
| O36.5123 | DX10 | Matern care for known or susp placntl insuff, 2nd tri, fts3 | Pregnancy | O365123 |
| O36.5124 | DX10 | Matern care for known or susp placntl insuff, 2nd tri, fts4 | Pregnancy | O365124 |
| O36.5125 | DX10 | Matern care for known or susp placntl insuff, 2nd tri, fts5 | Pregnancy | O365125 |
| O36.5129 | DX10 | Matern care for known or susp placntl insuff, 2nd tri, oth | Pregnancy | O365129 |
| O36.5130 | DX10 | Matern care for or susp placntl insuff, third tri, unsp | Pregnancy | O365130 |
| O36.5131 | DX10 | Matern care for or susp placntl insuff, third tri, fts1 | Pregnancy | O365131 |
| O36.5132 | DX10 | Matern care for or susp placntl insuff, third tri, fts2 | Pregnancy | O365132 |

| | | | | |
|---|---|---|---|---|
| O36.5133 | DX10 | Matern care for or susp placntl insuff, third tri, fts3 | Pregnancy | O365133 |
| O36.5134 | DX10 | Matern care for or susp placntl insuff, third tri, fts4 | Pregnancy | O365134 |
| O36.5135 | DX10 | Matern care for or susp placntl insuff, third tri, fts5 | Pregnancy | O365135 |
| O36.5139 | DX10 | Matern care for known or susp placntl insuff, third tri, oth | Pregnancy | O365139 |
| O36.5190 | DX10 | Matern care for known or susp placntl insuff, unsp tri, unsp | Pregnancy | O365190 |
| O36.5191 | DX10 | Matern care for known or susp placntl insuff, unsp tri, fts1 | Pregnancy | O365191 |
| O36.5192 | DX10 | Matern care for known or susp placntl insuff, unsp tri, fts2 | Pregnancy | O365192 |
| O36.5193 | DX10 | Matern care for known or susp placntl insuff, unsp tri, fts3 | Pregnancy | O365193 |
| O36.5194 | DX10 | Matern care for known or susp placntl insuff, unsp tri, fts4 | Pregnancy | O365194 |
| O36.5195 | DX10 | Matern care for known or susp placntl insuff, unsp tri, fts5 | Pregnancy | O365195 |
| O36.5199 | DX10 | Matern care for known or susp placntl insuff, unsp tri, oth | Pregnancy | O365199 |
| O36.5910 | DX10 | Matern care for oth or susp poor fetl grth, 1st tri, unsp | Pregnancy | O365910 |
| O36.5911 | DX10 | Matern care for oth or susp poor fetl grth, 1st tri, fts1 | Pregnancy | O365911 |
| O36.5912 | DX10 | Matern care for oth or susp poor fetl grth, 1st tri, fts2 | Pregnancy | O365912 |
| O36.5913 | DX10 | Matern care for oth or susp poor fetl grth, 1st tri, fts3 | Pregnancy | O365913 |
| O36.5914 | DX10 | Matern care for oth or susp poor fetl grth, 1st tri, fts4 | Pregnancy | O365914 |
| O36.5915 | DX10 | Matern care for oth or susp poor fetl grth, 1st tri, fts5 | Pregnancy | O365915 |
| O36.5919 | DX10 | Matern care for oth or susp poor fetl grth, 1st tri, oth | Pregnancy | O365919 |
| O36.5920 | DX10 | Matern care for oth or susp poor fetl grth, 2nd tri, unsp | Pregnancy | O365920 |
| O36.5921 | DX10 | Matern care for oth or susp poor fetl grth, 2nd tri, fts1 | Pregnancy | O365921 |
| O36.5922 | DX10 | Matern care for oth or susp poor fetl grth, 2nd tri, fts2 | Pregnancy | O365922 |
| O36.5923 | DX10 | Matern care for oth or susp poor fetl grth, 2nd tri, fts3 | Pregnancy | O365923 |
| O36.5924 | DX10 | Matern care for oth or susp poor fetl grth, 2nd tri, fts4 | Pregnancy | O365924 |
| O36.5925 | DX10 | Matern care for oth or susp poor fetl grth, 2nd tri, fts5 | Pregnancy | O365925 |
| O36.5929 | DX10 | Matern care for oth or susp poor fetl grth, 2nd tri, oth | Pregnancy | O365929 |
| O36.5930 | DX10 | Matern care for oth or susp poor fetl grth, third tri, unsp | Pregnancy | O365930 |
| O36.5931 | DX10 | Matern care for oth or susp poor fetl grth, third tri, fts1 | Pregnancy | O365931 |
| O36.5932 | DX10 | Matern care for oth or susp poor fetl grth, third tri, fts2 | Pregnancy | O365932 |
| O36.5933 | DX10 | Matern care for oth or susp poor fetl grth, third tri, fts3 | Pregnancy | O365933 |
| O36.5934 | DX10 | Matern care for oth or susp poor fetl grth, third tri, fts4 | Pregnancy | O365934 |
| O36.5935 | DX10 | Matern care for oth or susp poor fetl grth, third tri, fts5 | Pregnancy | O365935 |
| O36.5939 | DX10 | Matern care for oth or susp poor fetl grth, third tri, oth | Pregnancy | O365939 |
| O36.5990 | DX10 | Matern care for oth or susp poor fetl grth, unsp tri, unsp | Pregnancy | O365990 |
| O36.5991 | DX10 | Matern care for oth or susp poor fetl grth, unsp tri, fts1 | Pregnancy | O365991 |
| O36.5992 | DX10 | Matern care for oth or susp poor fetl grth, unsp tri, fts2 | Pregnancy | O365992 |
| O36.5993 | DX10 | Matern care for oth or susp poor fetl grth, unsp tri, fts3 | Pregnancy | O365993 |
| O36.5994 | DX10 | Matern care for oth or susp poor fetl grth, unsp tri, fts4 | Pregnancy | O365994 |
| O36.5995 | DX10 | Matern care for oth or susp poor fetl grth, unsp tri, fts5 | Pregnancy | O365995 |
| O36.5999 | DX10 | Matern care for oth or susp poor fetl grth, unsp tri, oth | Pregnancy | O365999 |
| O36.60X0 | DX10 | Maternal care for excess fetal growth, unsp trimester, unsp | Pregnancy | O3660X0 |
| O36.60X1 | DX10 | Maternal care for excess fetal growth, unsp tri, fetus 1 | Pregnancy | O3660X1 |
| O36.60X2 | DX10 | Maternal care for excess fetal growth, unsp tri, fetus 2 | Pregnancy | O3660X2 |
| O36.60X3 | DX10 | Maternal care for excess fetal growth, unsp tri, fetus 3 | Pregnancy | O3660X3 |
| O36.60X4 | DX10 | Maternal care for excess fetal growth, unsp tri, fetus 4 | Pregnancy | O3660X4 |
| O36.60X5 | DX10 | Maternal care for excess fetal growth, unsp tri, fetus 5 | Pregnancy | O3660X5 |
| O36.60X9 | DX10 | Maternal care for excess fetal growth, unsp trimester, oth | Pregnancy | O3660X9 |
| O36.61X0 | DX10 | Maternal care for excess fetal growth, first trimester, unsp | Pregnancy | O3661X0 |
| O36.61X1 | DX10 | Maternal care for excess fetal growth, first tri, fetus 1 | Pregnancy | O3661X1 |
| O36.61X2 | DX10 | Maternal care for excess fetal growth, first tri, fetus 2 | Pregnancy | O3661X2 |

| | | | | |
|---|---|---|---|---|
| O36.61X3 | DX10 | Maternal care for excess fetal growth, first tri, fetus 3 | Pregnancy | O3661X3 |
| O36.61X4 | DX10 | Maternal care for excess fetal growth, first tri, fetus 4 | Pregnancy | O3661X4 |
| O36.61X5 | DX10 | Maternal care for excess fetal growth, first tri, fetus 5 | Pregnancy | O3661X5 |
| O36.61X9 | DX10 | Maternal care for excess fetal growth, first trimester, oth | Pregnancy | O3661X9 |
| O36.62X0 | DX10 | Maternal care for excess fetal growth, second tri, unsp | Pregnancy | O3662X0 |
| O36.62X1 | DX10 | Maternal care for excess fetal growth, second tri, fetus 1 | Pregnancy | O3662X1 |
| O36.62X2 | DX10 | Maternal care for excess fetal growth, second tri, fetus 2 | Pregnancy | O3662X2 |
| O36.62X3 | DX10 | Maternal care for excess fetal growth, second tri, fetus 3 | Pregnancy | O3662X3 |
| O36.62X4 | DX10 | Maternal care for excess fetal growth, second tri, fetus 4 | Pregnancy | O3662X4 |
| O36.62X5 | DX10 | Maternal care for excess fetal growth, second tri, fetus 5 | Pregnancy | O3662X5 |
| O36.62X9 | DX10 | Maternal care for excess fetal growth, second trimester, oth | Pregnancy | O3662X9 |
| O36.63X0 | DX10 | Maternal care for excess fetal growth, third trimester, unsp | Pregnancy | O3663X0 |
| O36.63X1 | DX10 | Maternal care for excess fetal growth, third tri, fetus 1 | Pregnancy | O3663X1 |
| O36.63X2 | DX10 | Maternal care for excess fetal growth, third tri, fetus 2 | Pregnancy | O3663X2 |
| O36.63X3 | DX10 | Maternal care for excess fetal growth, third tri, fetus 3 | Pregnancy | O3663X3 |
| O36.63X4 | DX10 | Maternal care for excess fetal growth, third tri, fetus 4 | Pregnancy | O3663X4 |
| O36.63X5 | DX10 | Maternal care for excess fetal growth, third tri, fetus 5 | Pregnancy | O3663X5 |
| O36.63X9 | DX10 | Maternal care for excess fetal growth, third trimester, oth | Pregnancy | O3663X9 |
| O36.70X0 | DX10 | Maternal care for viable fetus in abd preg, unsp tri, unsp | Pregnancy | O3670X0 |
| O36.70X1 | DX10 | Matern care for viable fetus in abd preg, unsp tri, fetus 1 | Pregnancy | O3670X1 |
| O36.70X2 | DX10 | Matern care for viable fetus in abd preg, unsp tri, fetus 2 | Pregnancy | O3670X2 |
| O36.70X3 | DX10 | Matern care for viable fetus in abd preg, unsp tri, fetus 3 | Pregnancy | O3670X3 |
| O36.70X4 | DX10 | Matern care for viable fetus in abd preg, unsp tri, fetus 4 | Pregnancy | O3670X4 |
| O36.70X5 | DX10 | Matern care for viable fetus in abd preg, unsp tri, fetus 5 | Pregnancy | O3670X5 |
| O36.70X9 | DX10 | Maternal care for viable fetus in abd preg, unsp tri, oth | Pregnancy | O3670X9 |
| O36.71X0 | DX10 | Maternal care for viable fetus in abd preg, first tri, unsp | Pregnancy | O3671X0 |
| O36.71X1 | DX10 | Matern care for viable fetus in abd preg, first tri, fetus 1 | Pregnancy | O3671X1 |
| O36.71X2 | DX10 | Matern care for viable fetus in abd preg, first tri, fetus 2 | Pregnancy | O3671X2 |
| O36.71X3 | DX10 | Matern care for viable fetus in abd preg, first tri, fetus 3 | Pregnancy | O3671X3 |
| O36.71X4 | DX10 | Matern care for viable fetus in abd preg, first tri, fetus 4 | Pregnancy | O3671X4 |
| O36.71X5 | DX10 | Matern care for viable fetus in abd preg, first tri, fetus 5 | Pregnancy | O3671X5 |
| O36.71X9 | DX10 | Maternal care for viable fetus in abd preg, first tri, oth | Pregnancy | O3671X9 |
| O36.72X0 | DX10 | Maternal care for viable fetus in abd preg, second tri, unsp | Pregnancy | O3672X0 |
| O36.72X1 | DX10 | Matern care for viable fetus in abd preg, second tri, fts1 | Pregnancy | O3672X1 |
| O36.72X2 | DX10 | Matern care for viable fetus in abd preg, second tri, fts2 | Pregnancy | O3672X2 |
| O36.72X3 | DX10 | Matern care for viable fetus in abd preg, second tri, fts3 | Pregnancy | O3672X3 |
| O36.72X4 | DX10 | Matern care for viable fetus in abd preg, second tri, fts4 | Pregnancy | O3672X4 |
| O36.72X5 | DX10 | Matern care for viable fetus in abd preg, second tri, fts5 | Pregnancy | O3672X5 |
| O36.72X9 | DX10 | Maternal care for viable fetus in abd preg, second tri, oth | Pregnancy | O3672X9 |
| O36.73X0 | DX10 | Maternal care for viable fetus in abd preg, third tri, unsp | Pregnancy | O3673X0 |
| O36.73X1 | DX10 | Matern care for viable fetus in abd preg, third tri, fetus 1 | Pregnancy | O3673X1 |
| O36.73X2 | DX10 | Matern care for viable fetus in abd preg, third tri, fetus 2 | Pregnancy | O3673X2 |
| O36.73X3 | DX10 | Matern care for viable fetus in abd preg, third tri, fetus 3 | Pregnancy | O3673X3 |
| O36.73X4 | DX10 | Matern care for viable fetus in abd preg, third tri, fetus 4 | Pregnancy | O3673X4 |
| O36.73X5 | DX10 | Matern care for viable fetus in abd preg, third tri, fetus 5 | Pregnancy | O3673X5 |
| O36.73X9 | DX10 | Maternal care for viable fetus in abd preg, third tri, oth | Pregnancy | O3673X9 |
| O36.80X0 | DX10 | Pregnancy w inconclusive fetal viability, unsp | Pregnancy | O3680X0 |
| O36.80X1 | DX10 | Pregnancy with inconclusive fetal viability, fetus 1 | Pregnancy | O3680X1 |
| O36.80X2 | DX10 | Pregnancy with inconclusive fetal viability, fetus 2 | Pregnancy | O3680X2 |
| O36.80X3 | DX10 | Pregnancy with inconclusive fetal viability, fetus 3 | Pregnancy | O3680X3 |
| O36.80X4 | DX10 | Pregnancy with inconclusive fetal viability, fetus 4 | Pregnancy | O3680X4 |
| O36.80X5 | DX10 | Pregnancy with inconclusive fetal viability, fetus 5 | Pregnancy | O3680X5 |
| O36.80X9 | DX10 | Pregnancy with inconclusive fetal viability, other fetus | Pregnancy | O3680X9 |
| O36.8120 | DX10 | Decreased fetal movements, second trimester, unsp | Pregnancy | O368120 |
| O36.8121 | DX10 | Decreased fetal movements, second trimester, fetus 1 | Pregnancy | O368121 |
| O36.8122 | DX10 | Decreased fetal movements, second trimester, fetus 2 | Pregnancy | O368122 |

| | | | | |
|---|---|---|---|---|
| O36.8123 | DX10 | Decreased fetal movements, second trimester, fetus 3 | Pregnancy | O368123 |
| O36.8124 | DX10 | Decreased fetal movements, second trimester, fetus 4 | Pregnancy | O368124 |
| O36.8125 | DX10 | Decreased fetal movements, second trimester, fetus 5 | Pregnancy | O368125 |
| O36.8129 | DX10 | Decreased fetal movements, second trimester, other fetus | Pregnancy | O368129 |
| O36.8130 | DX10 | Decreased fetal movements, third trimester, unsp | Pregnancy | O368130 |
| O36.8131 | DX10 | Decreased fetal movements, third trimester, fetus 1 | Pregnancy | O368131 |
| O36.8132 | DX10 | Decreased fetal movements, third trimester, fetus 2 | Pregnancy | O368132 |
| O36.8133 | DX10 | Decreased fetal movements, third trimester, fetus 3 | Pregnancy | O368133 |
| O36.8134 | DX10 | Decreased fetal movements, third trimester, fetus 4 | Pregnancy | O368134 |
| O36.8135 | DX10 | Decreased fetal movements, third trimester, fetus 5 | Pregnancy | O368135 |
| O36.8139 | DX10 | Decreased fetal movements, third trimester, other fetus | Pregnancy | O368139 |
| O36.8190 | DX10 | Decreased fetal movements, unsp trimester, unsp | Pregnancy | O368190 |
| O36.8191 | DX10 | Decreased fetal movements, unspecified trimester, fetus 1 | Pregnancy | O368191 |
| O36.8192 | DX10 | Decreased fetal movements, unspecified trimester, fetus 2 | Pregnancy | O368192 |
| O36.8193 | DX10 | Decreased fetal movements, unspecified trimester, fetus 3 | Pregnancy | O368193 |
| O36.8194 | DX10 | Decreased fetal movements, unspecified trimester, fetus 4 | Pregnancy | O368194 |
| O36.8195 | DX10 | Decreased fetal movements, unspecified trimester, fetus 5 | Pregnancy | O368195 |
| O36.8199 | DX10 | Decreased fetal movements, unsp trimester, other fetus | Pregnancy | O368199 |
| O36.8210 | DX10 | Fetal anemia and thrombocytopenia, first trimester, unsp | Pregnancy | O368210 |
| O36.8211 | DX10 | Fetal anemia and thrombocytopenia, first trimester, fetus 1 | Pregnancy | O368211 |
| O36.8212 | DX10 | Fetal anemia and thrombocytopenia, first trimester, fetus 2 | Pregnancy | O368212 |
| O36.8213 | DX10 | Fetal anemia and thrombocytopenia, first trimester, fetus 3 | Pregnancy | O368213 |
| O36.8214 | DX10 | Fetal anemia and thrombocytopenia, first trimester, fetus 4 | Pregnancy | O368214 |
| O36.8215 | DX10 | Fetal anemia and thrombocytopenia, first trimester, fetus 5 | Pregnancy | O368215 |
| O36.8219 | DX10 | Fetal anemia and thrombocytopenia, first trimester, oth | Pregnancy | O368219 |
| O36.8220 | DX10 | Fetal anemia and thrombocytopenia, second trimester, unsp | Pregnancy | O368220 |
| O36.8221 | DX10 | Fetal anemia and thrombocytopenia, second trimester, fetus 1 | Pregnancy | O368221 |
| O36.8222 | DX10 | Fetal anemia and thrombocytopenia, second trimester, fetus 2 | Pregnancy | O368222 |
| O36.8223 | DX10 | Fetal anemia and thrombocytopenia, second trimester, fetus 3 | Pregnancy | O368223 |
| O36.8224 | DX10 | Fetal anemia and thrombocytopenia, second trimester, fetus 4 | Pregnancy | O368224 |
| O36.8225 | DX10 | Fetal anemia and thrombocytopenia, second trimester, fetus 5 | Pregnancy | O368225 |
| O36.8229 | DX10 | Fetal anemia and thrombocytopenia, second trimester, oth | Pregnancy | O368229 |
| O36.8230 | DX10 | Fetal anemia and thrombocytopenia, third trimester, unsp | Pregnancy | O368230 |
| O36.8231 | DX10 | Fetal anemia and thrombocytopenia, third trimester, fetus 1 | Pregnancy | O368231 |
| O36.8232 | DX10 | Fetal anemia and thrombocytopenia, third trimester, fetus 2 | Pregnancy | O368232 |
| O36.8233 | DX10 | Fetal anemia and thrombocytopenia, third trimester, fetus 3 | Pregnancy | O368233 |

| | | | | |
|---|---|---|---|---|
| O36.8234 | DX10 | Fetal anemia and thrombocytopenia, third trimester, fetus 4 | Pregnancy | O368234 |
| O36.8235 | DX10 | Fetal anemia and thrombocytopenia, third trimester, fetus 5 | Pregnancy | O368235 |
| O36.8239 | DX10 | Fetal anemia and thrombocytopenia, third trimester, oth | Pregnancy | O368239 |
| O36.8290 | DX10 | Fetal anemia and thrombocytopenia, unsp trimester, unsp | Pregnancy | O368290 |
| O36.8291 | DX10 | Fetal anemia and thrombocytopenia, unsp trimester, fetus 1 | Pregnancy | O368291 |
| O36.8292 | DX10 | Fetal anemia and thrombocytopenia, unsp trimester, fetus 2 | Pregnancy | O368292 |
| O36.8293 | DX10 | Fetal anemia and thrombocytopenia, unsp trimester, fetus 3 | Pregnancy | O368293 |
| O36.8294 | DX10 | Fetal anemia and thrombocytopenia, unsp trimester, fetus 4 | Pregnancy | O368294 |
| O36.8295 | DX10 | Fetal anemia and thrombocytopenia, unsp trimester, fetus 5 | Pregnancy | O368295 |
| O36.8299 | DX10 | Fetal anemia and thrombocytopenia, unsp trimester, oth fetus | Pregnancy | O368299 |
| O36.8910 | DX10 | Maternal care for oth fetal problems, first trimester, unsp | Pregnancy | O368910 |
| O36.8911 | DX10 | Maternal care for oth fetal problems, first tri, fetus 1 | Pregnancy | O368911 |
| O36.8912 | DX10 | Maternal care for oth fetal problems, first tri, fetus 2 | Pregnancy | O368912 |
| O36.8913 | DX10 | Maternal care for oth fetal problems, first tri, fetus 3 | Pregnancy | O368913 |
| O36.8914 | DX10 | Maternal care for oth fetal problems, first tri, fetus 4 | Pregnancy | O368914 |
| O36.8915 | DX10 | Maternal care for oth fetal problems, first tri, fetus 5 | Pregnancy | O368915 |
| O36.8919 | DX10 | Maternal care for oth fetal problems, first trimester, oth | Pregnancy | O368919 |
| O36.8920 | DX10 | Maternal care for oth fetal problems, second trimester, unsp | Pregnancy | O368920 |
| O36.8921 | DX10 | Maternal care for oth fetal problems, second tri, fetus 1 | Pregnancy | O368921 |
| O36.8922 | DX10 | Maternal care for oth fetal problems, second tri, fetus 2 | Pregnancy | O368922 |
| O36.8923 | DX10 | Maternal care for oth fetal problems, second tri, fetus 3 | Pregnancy | O368923 |
| O36.8924 | DX10 | Maternal care for oth fetal problems, second tri, fetus 4 | Pregnancy | O368924 |
| O36.8925 | DX10 | Maternal care for oth fetal problems, second tri, fetus 5 | Pregnancy | O368925 |
| O36.8929 | DX10 | Maternal care for oth fetal problems, second trimester, oth | Pregnancy | O368929 |
| O36.8930 | DX10 | Maternal care for oth fetal problems, third trimester, unsp | Pregnancy | O368930 |
| O36.8931 | DX10 | Maternal care for oth fetal problems, third tri, fetus 1 | Pregnancy | O368931 |
| O36.8932 | DX10 | Maternal care for oth fetal problems, third tri, fetus 2 | Pregnancy | O368932 |
| O36.8933 | DX10 | Maternal care for oth fetal problems, third tri, fetus 3 | Pregnancy | O368933 |
| O36.8934 | DX10 | Maternal care for oth fetal problems, third tri, fetus 4 | Pregnancy | O368934 |
| O36.8935 | DX10 | Maternal care for oth fetal problems, third tri, fetus 5 | Pregnancy | O368935 |
| O36.8939 | DX10 | Maternal care for oth fetal problems, third trimester, oth | Pregnancy | O368939 |
| O36.8990 | DX10 | Maternal care for oth fetal problems, unsp trimester, unsp | Pregnancy | O368990 |
| O36.8991 | DX10 | Maternal care for oth fetal problems, unsp tri, fetus 1 | Pregnancy | O368991 |
| O36.8992 | DX10 | Maternal care for oth fetal problems, unsp tri, fetus 2 | Pregnancy | O368992 |
| O36.8993 | DX10 | Maternal care for oth fetal problems, unsp tri, fetus 3 | Pregnancy | O368993 |
| O36.8994 | DX10 | Maternal care for oth fetal problems, unsp tri, fetus 4 | Pregnancy | O368994 |
| O36.8995 | DX10 | Maternal care for oth fetal problems, unsp tri, fetus 5 | Pregnancy | O368995 |
| O36.8999 | DX10 | Maternal care for oth fetal problems, unsp trimester, oth | Pregnancy | O368999 |
| O36.90X0 | DX10 | Maternal care for fetal problem, unsp, unsp trimester, unsp | Pregnancy | O3690X0 |
| O36.90X1 | DX10 | Maternal care for fetal problem, unsp, unsp tri, fetus 1 | Pregnancy | O3690X1 |
| O36.90X2 | DX10 | Maternal care for fetal problem, unsp, unsp tri, fetus 2 | Pregnancy | O3690X2 |
| O36.90X3 | DX10 | Maternal care for fetal problem, unsp, unsp tri, fetus 3 | Pregnancy | O3690X3 |
| O36.90X4 | DX10 | Maternal care for fetal problem, unsp, unsp tri, fetus 4 | Pregnancy | O3690X4 |
| O36.90X5 | DX10 | Maternal care for fetal problem, unsp, unsp tri, fetus 5 | Pregnancy | O3690X5 |

| | | | | |
|---|---|---|---|---|
| O36.90X9 | DX10 | Maternal care for fetal problem, unsp, unsp trimester, oth | Pregnancy | O3690X9 |
| O36.91X0 | DX10 | Maternal care for fetal problem, unsp, first trimester, unsp | Pregnancy | O3691X0 |
| O36.91X1 | DX10 | Maternal care for fetal problem, unsp, first tri, fetus 1 | Pregnancy | O3691X1 |
| O36.91X2 | DX10 | Maternal care for fetal problem, unsp, first tri, fetus 2 | Pregnancy | O3691X2 |
| O36.91X3 | DX10 | Maternal care for fetal problem, unsp, first tri, fetus 3 | Pregnancy | O3691X3 |
| O36.91X4 | DX10 | Maternal care for fetal problem, unsp, first tri, fetus 4 | Pregnancy | O3691X4 |
| O36.91X5 | DX10 | Maternal care for fetal problem, unsp, first tri, fetus 5 | Pregnancy | O3691X5 |
| O36.91X9 | DX10 | Maternal care for fetal problem, unsp, first trimester, oth | Pregnancy | O3691X9 |
| O36.92X0 | DX10 | Maternal care for fetal problem, unsp, second tri, unsp | Pregnancy | O3692X0 |
| O36.92X1 | DX10 | Maternal care for fetal problem, unsp, second tri, fetus 1 | Pregnancy | O3692X1 |
| O36.92X2 | DX10 | Maternal care for fetal problem, unsp, second tri, fetus 2 | Pregnancy | O3692X2 |
| O36.92X3 | DX10 | Maternal care for fetal problem, unsp, second tri, fetus 3 | Pregnancy | O3692X3 |
| O36.92X4 | DX10 | Maternal care for fetal problem, unsp, second tri, fetus 4 | Pregnancy | O3692X4 |
| O36.92X5 | DX10 | Maternal care for fetal problem, unsp, second tri, fetus 5 | Pregnancy | O3692X5 |
| O36.92X9 | DX10 | Maternal care for fetal problem, unsp, second trimester, oth | Pregnancy | O3692X9 |
| O36.93X0 | DX10 | Maternal care for fetal problem, unsp, third trimester, unsp | Pregnancy | O3693X0 |
| O36.93X1 | DX10 | Maternal care for fetal problem, unsp, third tri, fetus 1 | Pregnancy | O3693X1 |
| O36.93X2 | DX10 | Maternal care for fetal problem, unsp, third tri, fetus 2 | Pregnancy | O3693X2 |
| O36.93X3 | DX10 | Maternal care for fetal problem, unsp, third tri, fetus 3 | Pregnancy | O3693X3 |
| O36.93X4 | DX10 | Maternal care for fetal problem, unsp, third tri, fetus 4 | Pregnancy | O3693X4 |
| O36.93X5 | DX10 | Maternal care for fetal problem, unsp, third tri, fetus 5 | Pregnancy | O3693X5 |
| O36.93X9 | DX10 | Maternal care for fetal problem, unsp, third trimester, oth | Pregnancy | O3693X9 |
| O40.1XX0 | DX10 | Polyhydramnios, first trimester, not applicable or unsp | Pregnancy | O401XX0 |
| O40.1XX1 | DX10 | Polyhydramnios, first trimester, fetus 1 | Pregnancy | O401XX1 |
| O40.1XX2 | DX10 | Polyhydramnios, first trimester, fetus 2 | Pregnancy | O401XX2 |
| O40.1XX3 | DX10 | Polyhydramnios, first trimester, fetus 3 | Pregnancy | O401XX3 |
| O40.1XX4 | DX10 | Polyhydramnios, first trimester, fetus 4 | Pregnancy | O401XX4 |
| O40.1XX5 | DX10 | Polyhydramnios, first trimester, fetus 5 | Pregnancy | O401XX5 |
| O40.1XX9 | DX10 | Polyhydramnios, first trimester, other fetus | Pregnancy | O401XX9 |
| O40.2XX0 | DX10 | Polyhydramnios, second trimester, not applicable or unsp | Pregnancy | O402XX0 |
| O40.2XX1 | DX10 | Polyhydramnios, second trimester, fetus 1 | Pregnancy | O402XX1 |
| O40.2XX2 | DX10 | Polyhydramnios, second trimester, fetus 2 | Pregnancy | O402XX2 |
| O40.2XX3 | DX10 | Polyhydramnios, second trimester, fetus 3 | Pregnancy | O402XX3 |
| O40.2XX4 | DX10 | Polyhydramnios, second trimester, fetus 4 | Pregnancy | O402XX4 |
| O40.2XX5 | DX10 | Polyhydramnios, second trimester, fetus 5 | Pregnancy | O402XX5 |
| O40.2XX9 | DX10 | Polyhydramnios, second trimester, other fetus | Pregnancy | O402XX9 |
| O40.3XX0 | DX10 | Polyhydramnios, third trimester, not applicable or unsp | Pregnancy | O403XX0 |
| O40.3XX1 | DX10 | Polyhydramnios, third trimester, fetus 1 | Pregnancy | O403XX1 |
| O40.3XX2 | DX10 | Polyhydramnios, third trimester, fetus 2 | Pregnancy | O403XX2 |
| O40.3XX3 | DX10 | Polyhydramnios, third trimester, fetus 3 | Pregnancy | O403XX3 |
| O40.3XX4 | DX10 | Polyhydramnios, third trimester, fetus 4 | Pregnancy | O403XX4 |
| O40.3XX5 | DX10 | Polyhydramnios, third trimester, fetus 5 | Pregnancy | O403XX5 |
| O40.3XX9 | DX10 | Polyhydramnios, third trimester, other fetus | Pregnancy | O403XX9 |
| O40.9XX0 | DX10 | Polyhydramnios, unsp trimester, not applicable or unsp | Pregnancy | O409XX0 |
| O40.9XX1 | DX10 | Polyhydramnios, unspecified trimester, fetus 1 | Pregnancy | O409XX1 |
| O40.9XX2 | DX10 | Polyhydramnios, unspecified trimester, fetus 2 | Pregnancy | O409XX2 |
| O40.9XX3 | DX10 | Polyhydramnios, unspecified trimester, fetus 3 | Pregnancy | O409XX3 |
| O40.9XX4 | DX10 | Polyhydramnios, unspecified trimester, fetus 4 | Pregnancy | O409XX4 |
| O40.9XX5 | DX10 | Polyhydramnios, unspecified trimester, fetus 5 | Pregnancy | O409XX5 |
| O40.9XX9 | DX10 | Polyhydramnios, unspecified trimester, other fetus | Pregnancy | O409XX9 |
| O41.00X0 | DX10 | Oligohydramnios, unsp trimester, not applicable or unsp | Pregnancy | O4100X0 |

| | | | | |
|---|---|---|---|---|
| O41.00X1 | DX10 | Oligohydramnios, unspecified trimester, fetus 1 | Pregnancy | O4100X1 |
| O41.00X2 | DX10 | Oligohydramnios, unspecified trimester, fetus 2 | Pregnancy | O4100X2 |
| O41.00X3 | DX10 | Oligohydramnios, unspecified trimester, fetus 3 | Pregnancy | O4100X3 |
| O41.00X4 | DX10 | Oligohydramnios, unspecified trimester, fetus 4 | Pregnancy | O4100X4 |
| O41.00X5 | DX10 | Oligohydramnios, unspecified trimester, fetus 5 | Pregnancy | O4100X5 |
| O41.00X9 | DX10 | Oligohydramnios, unspecified trimester, other fetus | Pregnancy | O4100X9 |
| O41.01X0 | DX10 | Oligohydramnios, first trimester, not applicable or unsp | Pregnancy | O4101X0 |
| O41.01X1 | DX10 | Oligohydramnios, first trimester, fetus 1 | Pregnancy | O4101X1 |
| O41.01X2 | DX10 | Oligohydramnios, first trimester, fetus 2 | Pregnancy | O4101X2 |
| O41.01X3 | DX10 | Oligohydramnios, first trimester, fetus 3 | Pregnancy | O4101X3 |
| O41.01X4 | DX10 | Oligohydramnios, first trimester, fetus 4 | Pregnancy | O4101X4 |
| O41.01X5 | DX10 | Oligohydramnios, first trimester, fetus 5 | Pregnancy | O4101X5 |
| O41.01X9 | DX10 | Oligohydramnios, first trimester, other fetus | Pregnancy | O4101X9 |
| O41.02X0 | DX10 | Oligohydramnios, second trimester, not applicable or unsp | Pregnancy | O4102X0 |
| O41.02X1 | DX10 | Oligohydramnios, second trimester, fetus 1 | Pregnancy | O4102X1 |
| O41.02X2 | DX10 | Oligohydramnios, second trimester, fetus 2 | Pregnancy | O4102X2 |
| O41.02X3 | DX10 | Oligohydramnios, second trimester, fetus 3 | Pregnancy | O4102X3 |
| O41.02X4 | DX10 | Oligohydramnios, second trimester, fetus 4 | Pregnancy | O4102X4 |
| O41.02X5 | DX10 | Oligohydramnios, second trimester, fetus 5 | Pregnancy | O4102X5 |
| O41.02X9 | DX10 | Oligohydramnios, second trimester, other fetus | Pregnancy | O4102X9 |
| O41.03X0 | DX10 | Oligohydramnios, third trimester, not applicable or unsp | Pregnancy | O4103X0 |
| O41.03X1 | DX10 | Oligohydramnios, third trimester, fetus 1 | Pregnancy | O4103X1 |
| O41.03X2 | DX10 | Oligohydramnios, third trimester, fetus 2 | Pregnancy | O4103X2 |
| O41.03X3 | DX10 | Oligohydramnios, third trimester, fetus 3 | Pregnancy | O4103X3 |
| O41.03X4 | DX10 | Oligohydramnios, third trimester, fetus 4 | Pregnancy | O4103X4 |
| O41.03X5 | DX10 | Oligohydramnios, third trimester, fetus 5 | Pregnancy | O4103X5 |
| O41.03X9 | DX10 | Oligohydramnios, third trimester, other fetus | Pregnancy | O4103X9 |
| O41.1010 | DX10 | Infct of amniotic sac and membrns, unsp, first tri, unsp | Pregnancy | O411010 |
| O41.1011 | DX10 | Infct of amniotic sac and membrns, unsp, first tri, fetus 1 | Pregnancy | O411011 |
| O41.1012 | DX10 | Infct of amniotic sac and membrns, unsp, first tri, fetus 2 | Pregnancy | O411012 |
| O41.1013 | DX10 | Infct of amniotic sac and membrns, unsp, first tri, fetus 3 | Pregnancy | O411013 |
| O41.1014 | DX10 | Infct of amniotic sac and membrns, unsp, first tri, fetus 4 | Pregnancy | O411014 |
| O41.1015 | DX10 | Infct of amniotic sac and membrns, unsp, first tri, fetus 5 | Pregnancy | O411015 |
| O41.1019 | DX10 | Infct of amniotic sac and membrns, unsp, first tri, oth | Pregnancy | O411019 |
| O41.1020 | DX10 | Infct of amniotic sac and membrns, unsp, second tri, unsp | Pregnancy | O411020 |
| O41.1021 | DX10 | Infct of amniotic sac and membrns, unsp, second tri, fetus 1 | Pregnancy | O411021 |
| O41.1022 | DX10 | Infct of amniotic sac and membrns, unsp, second tri, fetus 2 | Pregnancy | O411022 |
| O41.1023 | DX10 | Infct of amniotic sac and membrns, unsp, second tri, fetus 3 | Pregnancy | O411023 |
| O41.1024 | DX10 | Infct of amniotic sac and membrns, unsp, second tri, fetus 4 | Pregnancy | O411024 |
| O41.1025 | DX10 | Infct of amniotic sac and membrns, unsp, second tri, fetus 5 | Pregnancy | O411025 |
| O41.1029 | DX10 | Infct of amniotic sac and membrns, unsp, second tri, oth | Pregnancy | O411029 |
| O41.1030 | DX10 | Infct of amniotic sac and membrns, unsp, third tri, unsp | Pregnancy | O411030 |
| O41.1031 | DX10 | Infct of amniotic sac and membrns, unsp, third tri, fetus 1 | Pregnancy | O411031 |

| | | | | |
|---|---|---|---|---|
| O41.1032 | DX10 | Infct of amniotic sac and membrns, unsp, third tri, fetus 2 | Pregnancy | O411032 |
| O41.1033 | DX10 | Infct of amniotic sac and membrns, unsp, third tri, fetus 3 | Pregnancy | O411033 |
| O41.1034 | DX10 | Infct of amniotic sac and membrns, unsp, third tri, fetus 4 | Pregnancy | O411034 |
| O41.1035 | DX10 | Infct of amniotic sac and membrns, unsp, third tri, fetus 5 | Pregnancy | O411035 |
| O41.1039 | DX10 | Infct of amniotic sac and membrns, unsp, third tri, oth | Pregnancy | O411039 |
| O41.1090 | DX10 | Infct of amniotic sac and membrns, unsp, unsp tri, unsp | Pregnancy | O411090 |
| O41.1091 | DX10 | Infct of amniotic sac and membrns, unsp, unsp tri, fetus 1 | Pregnancy | O411091 |
| O41.1092 | DX10 | Infct of amniotic sac and membrns, unsp, unsp tri, fetus 2 | Pregnancy | O411092 |
| O41.1093 | DX10 | Infct of amniotic sac and membrns, unsp, unsp tri, fetus 3 | Pregnancy | O411093 |
| O41.1094 | DX10 | Infct of amniotic sac and membrns, unsp, unsp tri, fetus 4 | Pregnancy | O411094 |
| O41.1095 | DX10 | Infct of amniotic sac and membrns, unsp, unsp tri, fetus 5 | Pregnancy | O411095 |
| O41.1099 | DX10 | Infct of amniotic sac and membrns, unsp, unsp trimester, oth | Pregnancy | O411099 |
| O41.1210 | DX10 | Chorioamnionitis, first trimester, not applicable or unsp | Pregnancy | O411210 |
| O41.1211 | DX10 | Chorioamnionitis, first trimester, fetus 1 | Pregnancy | O411211 |
| O41.1212 | DX10 | Chorioamnionitis, first trimester, fetus 2 | Pregnancy | O411212 |
| O41.1213 | DX10 | Chorioamnionitis, first trimester, fetus 3 | Pregnancy | O411213 |
| O41.1214 | DX10 | Chorioamnionitis, first trimester, fetus 4 | Pregnancy | O411214 |
| O41.1215 | DX10 | Chorioamnionitis, first trimester, fetus 5 | Pregnancy | O411215 |
| O41.1219 | DX10 | Chorioamnionitis, first trimester, other fetus | Pregnancy | O411219 |
| O41.1220 | DX10 | Chorioamnionitis, second trimester, not applicable or unsp | Pregnancy | O411220 |
| O41.1221 | DX10 | Chorioamnionitis, second trimester, fetus 1 | Pregnancy | O411221 |
| O41.1222 | DX10 | Chorioamnionitis, second trimester, fetus 2 | Pregnancy | O411222 |
| O41.1223 | DX10 | Chorioamnionitis, second trimester, fetus 3 | Pregnancy | O411223 |
| O41.1224 | DX10 | Chorioamnionitis, second trimester, fetus 4 | Pregnancy | O411224 |
| O41.1225 | DX10 | Chorioamnionitis, second trimester, fetus 5 | Pregnancy | O411225 |
| O41.1229 | DX10 | Chorioamnionitis, second trimester, other fetus | Pregnancy | O411229 |
| O41.1230 | DX10 | Chorioamnionitis, third trimester, not applicable or unsp | Pregnancy | O411230 |
| O41.1231 | DX10 | Chorioamnionitis, third trimester, fetus 1 | Pregnancy | O411231 |
| O41.1232 | DX10 | Chorioamnionitis, third trimester, fetus 2 | Pregnancy | O411232 |
| O41.1233 | DX10 | Chorioamnionitis, third trimester, fetus 3 | Pregnancy | O411233 |
| O41.1234 | DX10 | Chorioamnionitis, third trimester, fetus 4 | Pregnancy | O411234 |
| O41.1235 | DX10 | Chorioamnionitis, third trimester, fetus 5 | Pregnancy | O411235 |
| O41.1239 | DX10 | Chorioamnionitis, third trimester, other fetus | Pregnancy | O411239 |
| O41.1290 | DX10 | Chorioamnionitis, unsp trimester, not applicable or unsp | Pregnancy | O411290 |
| O41.1291 | DX10 | Chorioamnionitis, unspecified trimester, fetus 1 | Pregnancy | O411291 |
| O41.1292 | DX10 | Chorioamnionitis, unspecified trimester, fetus 2 | Pregnancy | O411292 |
| O41.1293 | DX10 | Chorioamnionitis, unspecified trimester, fetus 3 | Pregnancy | O411293 |
| O41.1294 | DX10 | Chorioamnionitis, unspecified trimester, fetus 4 | Pregnancy | O411294 |
| O41.1295 | DX10 | Chorioamnionitis, unspecified trimester, fetus 5 | Pregnancy | O411295 |
| O41.1299 | DX10 | Chorioamnionitis, unspecified trimester, other fetus | Pregnancy | O411299 |
| O41.1410 | DX10 | Placentitis, first trimester, not applicable or unspecified | Pregnancy | O411410 |
| O41.1411 | DX10 | Placentitis, first trimester, fetus 1 | Pregnancy | O411411 |
| O41.1412 | DX10 | Placentitis, first trimester, fetus 2 | Pregnancy | O411412 |
| O41.1413 | DX10 | Placentitis, first trimester, fetus 3 | Pregnancy | O411413 |
| O41.1414 | DX10 | Placentitis, first trimester, fetus 4 | Pregnancy | O411414 |

| | | | | |
|---|---|---|---|---|
| O41.1415 | DX10 | Placentitis, first trimester, fetus 5 | Pregnancy | O411415 |
| O41.1419 | DX10 | Placentitis, first trimester, other fetus | Pregnancy | O411419 |
| O41.1420 | DX10 | Placentitis, second trimester, not applicable or unspecified | Pregnancy | O411420 |
| O41.1421 | DX10 | Placentitis, second trimester, fetus 1 | Pregnancy | O411421 |
| O41.1422 | DX10 | Placentitis, second trimester, fetus 2 | Pregnancy | O411422 |
| O41.1423 | DX10 | Placentitis, second trimester, fetus 3 | Pregnancy | O411423 |
| O41.1424 | DX10 | Placentitis, second trimester, fetus 4 | Pregnancy | O411424 |
| O41.1425 | DX10 | Placentitis, second trimester, fetus 5 | Pregnancy | O411425 |
| O41.1429 | DX10 | Placentitis, second trimester, other fetus | Pregnancy | O411429 |
| O41.1430 | DX10 | Placentitis, third trimester, not applicable or unspecified | Pregnancy | O411430 |
| O41.1431 | DX10 | Placentitis, third trimester, fetus 1 | Pregnancy | O411431 |
| O41.1432 | DX10 | Placentitis, third trimester, fetus 2 | Pregnancy | O411432 |
| O41.1433 | DX10 | Placentitis, third trimester, fetus 3 | Pregnancy | O411433 |
| O41.1434 | DX10 | Placentitis, third trimester, fetus 4 | Pregnancy | O411434 |
| O41.1435 | DX10 | Placentitis, third trimester, fetus 5 | Pregnancy | O411435 |
| O41.1439 | DX10 | Placentitis, third trimester, other fetus | Pregnancy | O411439 |
| O41.1490 | DX10 | Placentitis, unsp trimester, not applicable or unspecified | Pregnancy | O411490 |
| O41.1491 | DX10 | Placentitis, unspecified trimester, fetus 1 | Pregnancy | O411491 |
| O41.1492 | DX10 | Placentitis, unspecified trimester, fetus 2 | Pregnancy | O411492 |
| O41.1493 | DX10 | Placentitis, unspecified trimester, fetus 3 | Pregnancy | O411493 |
| O41.1494 | DX10 | Placentitis, unspecified trimester, fetus 4 | Pregnancy | O411494 |
| O41.1495 | DX10 | Placentitis, unspecified trimester, fetus 5 | Pregnancy | O411495 |
| O41.1499 | DX10 | Placentitis, unspecified trimester, other fetus | Pregnancy | O411499 |
| O41.8X10 | DX10 | Oth disrd of amniotic fluid and membrns, first tri, unsp | Pregnancy | O418X10 |
| O41.8X11 | DX10 | Oth disrd of amniotic fluid and membrns, first tri, fetus 1 | Pregnancy | O418X11 |
| O41.8X12 | DX10 | Oth disrd of amniotic fluid and membrns, first tri, fetus 2 | Pregnancy | O418X12 |
| O41.8X13 | DX10 | Oth disrd of amniotic fluid and membrns, first tri, fetus 3 | Pregnancy | O418X13 |
| O41.8X14 | DX10 | Oth disrd of amniotic fluid and membrns, first tri, fetus 4 | Pregnancy | O418X14 |
| O41.8X15 | DX10 | Oth disrd of amniotic fluid and membrns, first tri, fetus 5 | Pregnancy | O418X15 |
| O41.8X19 | DX10 | Oth disrd of amniotic fluid and membrns, first tri, oth | Pregnancy | O418X19 |
| O41.8X20 | DX10 | Oth disrd of amniotic fluid and membrns, second tri, unsp | Pregnancy | O418X20 |
| O41.8X21 | DX10 | Oth disrd of amniotic fluid and membrns, second tri, fetus 1 | Pregnancy | O418X21 |
| O41.8X22 | DX10 | Oth disrd of amniotic fluid and membrns, second tri, fetus 2 | Pregnancy | O418X22 |
| O41.8X23 | DX10 | Oth disrd of amniotic fluid and membrns, second tri, fetus 3 | Pregnancy | O418X23 |
| O41.8X24 | DX10 | Oth disrd of amniotic fluid and membrns, second tri, fetus 4 | Pregnancy | O418X24 |
| O41.8X25 | DX10 | Oth disrd of amniotic fluid and membrns, second tri, fetus 5 | Pregnancy | O418X25 |
| O41.8X29 | DX10 | Oth disrd of amniotic fluid and membrns, second tri, oth | Pregnancy | O418X29 |
| O41.8X30 | DX10 | Oth disrd of amniotic fluid and membrns, third tri, unsp | Pregnancy | O418X30 |
| O41.8X31 | DX10 | Oth disrd of amniotic fluid and membrns, third tri, fetus 1 | Pregnancy | O418X31 |
| O41.8X32 | DX10 | Oth disrd of amniotic fluid and membrns, third tri, fetus 2 | Pregnancy | O418X32 |
| O41.8X33 | DX10 | Oth disrd of amniotic fluid and membrns, third tri, fetus 3 | Pregnancy | O418X33 |
| O41.8X34 | DX10 | Oth disrd of amniotic fluid and membrns, third tri, fetus 4 | Pregnancy | O418X34 |
| O41.8X35 | DX10 | Oth disrd of amniotic fluid and membrns, third tri, fetus 5 | Pregnancy | O418X35 |
| O41.8X39 | DX10 | Oth disrd of amniotic fluid and membrns, third tri, oth | Pregnancy | O418X39 |
| O41.8X90 | DX10 | Oth disrd of amniotic fluid and membrns, unsp tri, unsp | Pregnancy | O418X90 |

| | | | | |
|---|---|---|---|---|
| O41.8X91 | DX10 | Oth disrd of amniotic fluid and membrns, unsp tri, fetus 1 | Pregnancy | O418X91 |
| O41.8X92 | DX10 | Oth disrd of amniotic fluid and membrns, unsp tri, fetus 2 | Pregnancy | O418X92 |
| O41.8X93 | DX10 | Oth disrd of amniotic fluid and membrns, unsp tri, fetus 3 | Pregnancy | O418X93 |
| O41.8X94 | DX10 | Oth disrd of amniotic fluid and membrns, unsp tri, fetus 4 | Pregnancy | O418X94 |
| O41.8X95 | DX10 | Oth disrd of amniotic fluid and membrns, unsp tri, fetus 5 | Pregnancy | O418X95 |
| O41.8X99 | DX10 | Oth disrd of amniotic fluid and membrns, unsp trimester, oth | Pregnancy | O418X99 |
| O41.90X0 | DX10 | Disorder of amniotic fluid and membrns, unsp, unsp tri, unsp | Pregnancy | O4190X0 |
| O41.90X1 | DX10 | Disorder of amnio fluid and membrns, unsp, unsp tri, fetus 1 | Pregnancy | O4190X1 |
| O41.90X2 | DX10 | Disorder of amnio fluid and membrns, unsp, unsp tri, fetus 2 | Pregnancy | O4190X2 |
| O41.90X3 | DX10 | Disorder of amnio fluid and membrns, unsp, unsp tri, fetus 3 | Pregnancy | O4190X3 |
| O41.90X4 | DX10 | Disorder of amnio fluid and membrns, unsp, unsp tri, fetus 4 | Pregnancy | O4190X4 |
| O41.90X5 | DX10 | Disorder of amnio fluid and membrns, unsp, unsp tri, fetus 5 | Pregnancy | O4190X5 |
| O41.90X9 | DX10 | Disorder of amniotic fluid and membrns, unsp, unsp tri, oth | Pregnancy | O4190X9 |
| O41.91X0 | DX10 | Disorder of amnio fluid and membrns, unsp, first tri, unsp | Pregnancy | O4191X0 |
| O41.91X1 | DX10 | Disord of amnio fluid and membrns, unsp, first tri, fetus 1 | Pregnancy | O4191X1 |
| O41.91X2 | DX10 | Disord of amnio fluid and membrns, unsp, first tri, fetus 2 | Pregnancy | O4191X2 |
| O41.91X3 | DX10 | Disord of amnio fluid and membrns, unsp, first tri, fetus 3 | Pregnancy | O4191X3 |
| O41.91X4 | DX10 | Disord of amnio fluid and membrns, unsp, first tri, fetus 4 | Pregnancy | O4191X4 |
| O41.91X5 | DX10 | Disord of amnio fluid and membrns, unsp, first tri, fetus 5 | Pregnancy | O4191X5 |
| O41.91X9 | DX10 | Disorder of amniotic fluid and membrns, unsp, first tri, oth | Pregnancy | O4191X9 |
| O41.92X0 | DX10 | Disorder of amnio fluid and membrns, unsp, second tri, unsp | Pregnancy | O4192X0 |
| O41.92X1 | DX10 | Disord of amnio fluid and membrns, unsp, second tri, fetus 1 | Pregnancy | O4192X1 |
| O41.92X2 | DX10 | Disord of amnio fluid and membrns, unsp, second tri, fetus 2 | Pregnancy | O4192X2 |
| O41.92X3 | DX10 | Disord of amnio fluid and membrns, unsp, second tri, fetus 3 | Pregnancy | O4192X3 |
| O41.92X4 | DX10 | Disord of amnio fluid and membrns, unsp, second tri, fetus 4 | Pregnancy | O4192X4 |
| O41.92X5 | DX10 | Disord of amnio fluid and membrns, unsp, second tri, fetus 5 | Pregnancy | O4192X5 |
| O41.92X9 | DX10 | Disorder of amnio fluid and membrns, unsp, second tri, oth | Pregnancy | O4192X9 |
| O41.93X0 | DX10 | Disorder of amnio fluid and membrns, unsp, third tri, unsp | Pregnancy | O4193X0 |
| O41.93X1 | DX10 | Disord of amnio fluid and membrns, unsp, third tri, fetus 1 | Pregnancy | O4193X1 |

| | | | | |
|---|---|---|---|---|
| O41.93X2 | DX10 | Disord of amnio fluid and membrns, unsp, third tri, fetus 2 | Pregnancy | O4193X2 |
| O41.93X3 | DX10 | Disord of amnio fluid and membrns, unsp, third tri, fetus 3 | Pregnancy | O4193X3 |
| O41.93X4 | DX10 | Disord of amnio fluid and membrns, unsp, third tri, fetus 4 | Pregnancy | O4193X4 |
| O41.93X5 | DX10 | Disord of amnio fluid and membrns, unsp, third tri, fetus 5 | Pregnancy | O4193X5 |
| O41.93X9 | DX10 | Disorder of amniotic fluid and membrns, unsp, third tri, oth | Pregnancy | O4193X9 |
| O42.00 | DX10 | Prem ROM, onset labor w/n 24 hr of rupt, unsp weeks of gest | Pregnancy | O4200 |
| O42.011 | DX10 | Pretrm prem ROM, onset labor w/n 24 hours of rupt, first tri | Pregnancy | O42011 |
| O42.012 | DX10 | Pretrm prem ROM, onset labor w/n 24 hours of rupt, 2nd tri | Pregnancy | O42012 |
| O42.013 | DX10 | Pretrm prem ROM, onset labor w/n 24 hours of rupt, third tri | Pregnancy | O42013 |
| O42.019 | DX10 | Pretrm prem ROM, onset labor w/n 24 hours of rupt, unsp tri | Pregnancy | O42019 |
| O42.02 | DX10 | Full-term prem ROM, onset labor within 24 hours of rupture | Pregnancy | O4202 |
| O42.10 | DX10 | Prem ROM, onset labor > 24 hr fol rupt, unsp weeks of gest | Pregnancy | O4210 |
| O42.111 | DX10 | Pretrm prem ROM, onset labor > 24 hours fol rupt, first tri | Pregnancy | O42111 |
| O42.112 | DX10 | Pretrm prem ROM, onset labor > 24 hours fol rupt, second tri | Pregnancy | O42112 |
| O42.113 | DX10 | Pretrm prem ROM, onset labor > 24 hours fol rupt, third tri | Pregnancy | O42113 |
| O42.119 | DX10 | Pretrm prem ROM, onset labor > 24 hours fol rupt, unsp tri | Pregnancy | O42119 |
| O42.12 | DX10 | Full-term premature ROM, onset labor > 24 hours fol rupture | Pregnancy | O4212 |
| O42.90 | DX10 | Prem ROM, 7th0 betw rupt & onst labr, unsp weeks of gest | Pregnancy | O4290 |
| O42.911 | DX10 | Pretrm prem ROM, unsp time betw rupt and onset labr, 1st tri | Pregnancy | O42911 |
| O42.912 | DX10 | Pretrm prem ROM, unsp time betw rupt and onset labr, 2nd tri | Pregnancy | O42912 |
| O42.913 | DX10 | Pretrm prem ROM, unsp time betw rupt and onst labr, 3rd tri | Pregnancy | O42913 |
| O42.919 | DX10 | Pretrm prem ROM, unsp time betw rupt and onst labr, unsp tri | Pregnancy | O42919 |
| O42.92 | DX10 | Full-term prem ROM, unsp time betw rupture and onset labor | Pregnancy | O4292 |
| O43.011 | DX10 | Fetomaternal placental transfusion syndrome, first trimester | Pregnancy | O43011 |
| O43.012 | DX10 | Fetomaternal placental transfuse syndrome, second trimester | Pregnancy | O43012 |
| O43.013 | DX10 | Fetomaternal placental transfusion syndrome, third trimester | Pregnancy | O43013 |
| O43.019 | DX10 | Fetomaternal placental transfusion syndrome, unsp trimester | Pregnancy | O43019 |
| O43.021 | DX10 | Fetus-to-fetus placental transfuse syndrome, first trimester | Pregnancy | O43021 |
| O43.022 | DX10 | Fetus-to-fetus placntl transfuse syndrome, second trimester | Pregnancy | O43022 |

| | | | | |
|---|---|---|---|---|
| O43.023 | DX10 | Fetus-to-fetus placental transfuse syndrome, third trimester | Pregnancy | O43023 |
| O43.029 | DX10 | Fetus-to-fetus placental transfuse syndrome, unsp trimester | Pregnancy | O43029 |
| O43.101 | DX10 | Malformation of placenta, unspecified, first trimester | Pregnancy | O43101 |
| O43.102 | DX10 | Malformation of placenta, unspecified, second trimester | Pregnancy | O43102 |
| O43.103 | DX10 | Malformation of placenta, unspecified, third trimester | Pregnancy | O43103 |
| O43.109 | DX10 | Malformation of placenta, unspecified, unspecified trimester | Pregnancy | O43109 |
| O43.111 | DX10 | Circumvallate placenta, first trimester | Pregnancy | O43111 |
| O43.112 | DX10 | Circumvallate placenta, second trimester | Pregnancy | O43112 |
| O43.113 | DX10 | Circumvallate placenta, third trimester | Pregnancy | O43113 |
| O43.119 | DX10 | Circumvallate placenta, unspecified trimester | Pregnancy | O43119 |
| O43.121 | DX10 | Velamentous insertion of umbilical cord, first trimester | Pregnancy | O43121 |
| O43.122 | DX10 | Velamentous insertion of umbilical cord, second trimester | Pregnancy | O43122 |
| O43.123 | DX10 | Velamentous insertion of umbilical cord, third trimester | Pregnancy | O43123 |
| O43.129 | DX10 | Velamentous insertion of umbilical cord, unsp trimester | Pregnancy | O43129 |
| O43.191 | DX10 | Other malformation of placenta, first trimester | Pregnancy | O43191 |
| O43.192 | DX10 | Other malformation of placenta, second trimester | Pregnancy | O43192 |
| O43.193 | DX10 | Other malformation of placenta, third trimester | Pregnancy | O43193 |
| O43.199 | DX10 | Other malformation of placenta, unspecified trimester | Pregnancy | O43199 |
| O43.211 | DX10 | Placenta accreta, first trimester | Pregnancy | O43211 |
| O43.212 | DX10 | Placenta accreta, second trimester | Pregnancy | O43212 |
| O43.213 | DX10 | Placenta accreta, third trimester | Pregnancy | O43213 |
| O43.219 | DX10 | Placenta accreta, unspecified trimester | Pregnancy | O43219 |
| O43.221 | DX10 | Placenta increta, first trimester | Pregnancy | O43221 |
| O43.222 | DX10 | Placenta increta, second trimester | Pregnancy | O43222 |
| O43.223 | DX10 | Placenta increta, third trimester | Pregnancy | O43223 |
| O43.229 | DX10 | Placenta increta, unspecified trimester | Pregnancy | O43229 |
| O43.231 | DX10 | Placenta percreta, first trimester | Pregnancy | O43231 |
| O43.232 | DX10 | Placenta percreta, second trimester | Pregnancy | O43232 |
| O43.233 | DX10 | Placenta percreta, third trimester | Pregnancy | O43233 |
| O43.239 | DX10 | Placenta percreta, unspecified trimester | Pregnancy | O43239 |
| O43.811 | DX10 | Placental infarction, first trimester | Pregnancy | O43811 |
| O43.812 | DX10 | Placental infarction, second trimester | Pregnancy | O43812 |
| O43.813 | DX10 | Placental infarction, third trimester | Pregnancy | O43813 |
| O43.819 | DX10 | Placental infarction, unspecified trimester | Pregnancy | O43819 |
| O43.891 | DX10 | Other placental disorders, first trimester | Pregnancy | O43891 |
| O43.892 | DX10 | Other placental disorders, second trimester | Pregnancy | O43892 |
| O43.893 | DX10 | Other placental disorders, third trimester | Pregnancy | O43893 |
| O43.899 | DX10 | Other placental disorders, unspecified trimester | Pregnancy | O43899 |
| O43.90 | DX10 | Unspecified placental disorder, unspecified trimester | Pregnancy | O4390 |
| O43.91 | DX10 | Unspecified placental disorder, first trimester | Pregnancy | O4391 |
| O43.92 | DX10 | Unspecified placental disorder, second trimester | Pregnancy | O4392 |
| O43.93 | DX10 | Unspecified placental disorder, third trimester | Pregnancy | O4393 |
| O44.00 | DX10 | Placenta previa specified as w/o hemorrhage, unsp trimester | Pregnancy | O4400 |
| O44.01 | DX10 | Placenta previa specified as w/o hemorrhage, first trimester | Pregnancy | O4401 |
| O44.02 | DX10 | Placenta previa specified as w/o hemor, second trimester | Pregnancy | O4402 |
| O44.03 | DX10 | Placenta previa specified as w/o hemorrhage, third trimester | Pregnancy | O4403 |
| O44.10 | DX10 | Placenta previa with hemorrhage, unspecified trimester | Pregnancy | O4410 |
| O44.11 | DX10 | Placenta previa with hemorrhage, first trimester | Pregnancy | O4411 |
| O44.12 | DX10 | Placenta previa with hemorrhage, second trimester | Pregnancy | O4412 |
| O44.13 | DX10 | Placenta previa with hemorrhage, third trimester | Pregnancy | O4413 |

| | | | | |
|---|---|---|---|---|
| O44.40 | DX10 | LOW LYING PLACENTA NOS/WO HEMORRHAGE UNS TRIM | Pregnancy | O4440 |
| O45.001 | DX10 | Prem separtn of placenta w coag defect, unsp, first tri | Pregnancy | O45001 |
| O45.002 | DX10 | Prem separtn of placenta w coag defect, unsp, second tri | Pregnancy | O45002 |
| O45.003 | DX10 | Prem separtn of placenta w coag defect, unsp, third tri | Pregnancy | O45003 |
| O45.009 | DX10 | Prem separtn of placenta w coag defect, unsp, unsp trimester | Pregnancy | O45009 |
| O45.011 | DX10 | Prem separtn of placenta w afibrinogenemia, first trimester | Pregnancy | O45011 |
| O45.012 | DX10 | Prem separtn of placenta w afibrinogenemia, second trimester | Pregnancy | O45012 |
| O45.013 | DX10 | Prem separtn of placenta w afibrinogenemia, third trimester | Pregnancy | O45013 |
| O45.019 | DX10 | Prem separtn of placenta w afibrinogenemia, unsp trimester | Pregnancy | O45019 |
| O45.021 | DX10 | Prem separtn of placenta w dissem intravasc coag, first tri | Pregnancy | O45021 |
| O45.022 | DX10 | Prem separtn of placenta w dissem intravasc coag, second tri | Pregnancy | O45022 |
| O45.023 | DX10 | Prem separtn of placenta w dissem intravasc coag, third tri | Pregnancy | O45023 |
| O45.029 | DX10 | Prem separtn of placenta w dissem intravasc coag, unsp tri | Pregnancy | O45029 |
| O45.091 | DX10 | Prem separtn of placenta w oth coag defect, first trimester | Pregnancy | O45091 |
| O45.092 | DX10 | Prem separtn of placenta w oth coag defect, second trimester | Pregnancy | O45092 |
| O45.093 | DX10 | Prem separtn of placenta w oth coag defect, third trimester | Pregnancy | O45093 |
| O45.099 | DX10 | Prem separtn of placenta w oth coag defect, unsp trimester | Pregnancy | O45099 |
| O45.8X1 | DX10 | Other premature separation of placenta, first trimester | Pregnancy | O458X1 |
| O45.8X2 | DX10 | Other premature separation of placenta, second trimester | Pregnancy | O458X2 |
| O45.8X3 | DX10 | Other premature separation of placenta, third trimester | Pregnancy | O458X3 |
| O45.8X9 | DX10 | Other premature separation of placenta, unsp trimester | Pregnancy | O458X9 |
| O45.90 | DX10 | Premature separation of placenta, unsp, unsp trimester | Pregnancy | O4590 |
| O45.91 | DX10 | Premature separation of placenta, unsp, first trimester | Pregnancy | O4591 |
| O45.92 | DX10 | Premature separation of placenta, unsp, second trimester | Pregnancy | O4592 |
| O45.93 | DX10 | Premature separation of placenta, unsp, third trimester | Pregnancy | O4593 |
| O46.001 | DX10 | Antepartum hemorrhage w coag defect, unsp, first trimester | Pregnancy | O46001 |
| O46.002 | DX10 | Antepartum hemorrhage w coag defect, unsp, second trimester | Pregnancy | O46002 |
| O46.003 | DX10 | Antepartum hemorrhage w coag defect, unsp, third trimester | Pregnancy | O46003 |
| O46.009 | DX10 | Antepartum hemorrhage w coag defect, unsp, unsp trimester | Pregnancy | O46009 |
| O46.011 | DX10 | Antepartum hemorrhage with afibrinogenemia, first trimester | Pregnancy | O46011 |
| O46.012 | DX10 | Antepartum hemorrhage with afibrinogenemia, second trimester | Pregnancy | O46012 |
| O46.013 | DX10 | Antepartum hemorrhage with afibrinogenemia, third trimester | Pregnancy | O46013 |
| O46.019 | DX10 | Antepartum hemorrhage with afibrinogenemia, unsp trimester | Pregnancy | O46019 |

| | | | | |
|---|---|---|---|---|
| O46.021 | DX10 | Antepart hemorrhage w dissem intravasc coag, first trimester | Pregnancy | O46021 |
| O46.022 | DX10 | Antepart hemor w dissem intravasc coag, second trimester | Pregnancy | O46022 |
| O46.023 | DX10 | Antepart hemorrhage w dissem intravasc coag, third trimester | Pregnancy | O46023 |
| O46.029 | DX10 | Antepart hemorrhage w dissem intravasc coag, unsp trimester | Pregnancy | O46029 |
| O46.091 | DX10 | Antepartum hemorrhage w oth coag defect, first trimester | Pregnancy | O46091 |
| O46.092 | DX10 | Antepartum hemorrhage w oth coag defect, second trimester | Pregnancy | O46092 |
| O46.093 | DX10 | Antepartum hemorrhage w oth coag defect, third trimester | Pregnancy | O46093 |
| O46.099 | DX10 | Antepartum hemorrhage w oth coag defect, unsp trimester | Pregnancy | O46099 |
| O46.8X1 | DX10 | Other antepartum hemorrhage, first trimester | Pregnancy | O468X1 |
| O46.8X2 | DX10 | Other antepartum hemorrhage, second trimester | Pregnancy | O468X2 |
| O46.8X3 | DX10 | Other antepartum hemorrhage, third trimester | Pregnancy | O468X3 |
| O46.8X9 | DX10 | Other antepartum hemorrhage, unspecified trimester | Pregnancy | O468X9 |
| O46.90 | DX10 | Antepartum hemorrhage, unspecified, unspecified trimester | Pregnancy | O4690 |
| O46.91 | DX10 | Antepartum hemorrhage, unspecified, first trimester | Pregnancy | O4691 |
| O46.92 | DX10 | Antepartum hemorrhage, unspecified, second trimester | Pregnancy | O4692 |
| O46.93 | DX10 | Antepartum hemorrhage, unspecified, third trimester | Pregnancy | O4693 |
| O47.00 | DX10 | False labor before 37 completed weeks of gest, unsp tri | Pregnancy | O4700 |
| O47.02 | DX10 | False labor before 37 completed weeks of gest, second tri | Pregnancy | O4702 |
| O47.03 | DX10 | False labor before 37 completed weeks of gest, third tri | Pregnancy | O4703 |
| O47.1 | DX10 | False labor at or after 37 completed weeks of gestation | Pregnancy | O471 |
| O47.9 | DX10 | False labor, unspecified | Pregnancy | O479 |
| O48.0 | DX10 | Post-term pregnancy | Pregnancy | O480 |
| O48.1 | DX10 | Prolonged pregnancy | Pregnancy | O481 |
| O60.00 | DX10 | Preterm labor without delivery, unspecified trimester | Pregnancy | O6000 |
| O60.02 | DX10 | Preterm labor without delivery, second trimester | Pregnancy | O6002 |
| O60.03 | DX10 | Preterm labor without delivery, third trimester | Pregnancy | O6003 |
| O61.0 | DX10 | Failed medical induction of labor | Pregnancy | O610 |
| O61.1 | DX10 | Failed instrumental induction of labor | Pregnancy | O611 |
| O61.8 | DX10 | Other failed induction of labor | Pregnancy | O618 |
| O61.9 | DX10 | Failed induction of labor, unspecified | Pregnancy | O619 |
| O62.0 | DX10 | Primary inadequate contractions | Pregnancy | O620 |
| O62.1 | DX10 | Secondary uterine inertia | Pregnancy | O621 |
| O62.2 | DX10 | Other uterine inertia | Pregnancy | O622 |
| O62.3 | DX10 | Precipitate labor | Pregnancy | O623 |
| O62.4 | DX10 | Hypertonic, incoordinate, and prolonged uterine contractions | Pregnancy | O624 |
| O62.8 | DX10 | Other abnormalities of forces of labor | Pregnancy | O628 |
| O62.9 | DX10 | Abnormality of forces of labor, unspecified | Pregnancy | O629 |
| O63.0 | DX10 | Prolonged first stage (of labor) | Pregnancy | O630 |
| O63.1 | DX10 | Prolonged second stage (of labor) | Pregnancy | O631 |
| O63.2 | DX10 | Delayed delivery of second twin, triplet, etc. | Pregnancy | O632 |
| O63.9 | DX10 | Long labor, unspecified | Pregnancy | O639 |
| O64.0XX0 | DX10 | Obstructed labor due to incmpl rotation of fetal head, unsp | Pregnancy | O640XX0 |
| O64.0XX1 | DX10 | Obst labor due to incmpl rotation of fetal head, fetus 1 | Pregnancy | O640XX1 |
| O64.0XX2 | DX10 | Obst labor due to incmpl rotation of fetal head, fetus 2 | Pregnancy | O640XX2 |
| O64.0XX3 | DX10 | Obst labor due to incmpl rotation of fetal head, fetus 3 | Pregnancy | O640XX3 |
| O64.0XX4 | DX10 | Obst labor due to incmpl rotation of fetal head, fetus 4 | Pregnancy | O640XX4 |
| O64.0XX5 | DX10 | Obst labor due to incmpl rotation of fetal head, fetus 5 | Pregnancy | O640XX5 |

| | | | | |
|---|---|---|---|---|
| O64.0XX9 | DX10 | Obstructed labor due to incmpl rotation of fetal head, oth | Pregnancy | O640XX9 |
| O64.1XX0 | DX10 | Obstructed labor due to breech presentation, unsp | Pregnancy | O641XX0 |
| O64.1XX1 | DX10 | Obstructed labor due to breech presentation, fetus 1 | Pregnancy | O641XX1 |
| O64.1XX2 | DX10 | Obstructed labor due to breech presentation, fetus 2 | Pregnancy | O641XX2 |
| O64.1XX3 | DX10 | Obstructed labor due to breech presentation, fetus 3 | Pregnancy | O641XX3 |
| O64.1XX4 | DX10 | Obstructed labor due to breech presentation, fetus 4 | Pregnancy | O641XX4 |
| O64.1XX5 | DX10 | Obstructed labor due to breech presentation, fetus 5 | Pregnancy | O641XX5 |
| O64.1XX9 | DX10 | Obstructed labor due to breech presentation, other fetus | Pregnancy | O641XX9 |
| O64.2XX0 | DX10 | Obstructed labor due to face presentation, unsp | Pregnancy | O642XX0 |
| O64.2XX1 | DX10 | Obstructed labor due to face presentation, fetus 1 | Pregnancy | O642XX1 |
| O64.2XX2 | DX10 | Obstructed labor due to face presentation, fetus 2 | Pregnancy | O642XX2 |
| O64.2XX3 | DX10 | Obstructed labor due to face presentation, fetus 3 | Pregnancy | O642XX3 |
| O64.2XX4 | DX10 | Obstructed labor due to face presentation, fetus 4 | Pregnancy | O642XX4 |
| O64.2XX5 | DX10 | Obstructed labor due to face presentation, fetus 5 | Pregnancy | O642XX5 |
| O64.2XX9 | DX10 | Obstructed labor due to face presentation, other fetus | Pregnancy | O642XX9 |
| O64.3XX0 | DX10 | Obstructed labor due to brow presentation, unsp | Pregnancy | O643XX0 |
| O64.3XX1 | DX10 | Obstructed labor due to brow presentation, fetus 1 | Pregnancy | O643XX1 |
| O64.3XX2 | DX10 | Obstructed labor due to brow presentation, fetus 2 | Pregnancy | O643XX2 |
| O64.3XX3 | DX10 | Obstructed labor due to brow presentation, fetus 3 | Pregnancy | O643XX3 |
| O64.3XX4 | DX10 | Obstructed labor due to brow presentation, fetus 4 | Pregnancy | O643XX4 |
| O64.3XX5 | DX10 | Obstructed labor due to brow presentation, fetus 5 | Pregnancy | O643XX5 |
| O64.3XX9 | DX10 | Obstructed labor due to brow presentation, other fetus | Pregnancy | O643XX9 |
| O64.4XX0 | DX10 | Obstructed labor due to shoulder presentation, unsp | Pregnancy | O644XX0 |
| O64.4XX1 | DX10 | Obstructed labor due to shoulder presentation, fetus 1 | Pregnancy | O644XX1 |
| O64.4XX2 | DX10 | Obstructed labor due to shoulder presentation, fetus 2 | Pregnancy | O644XX2 |
| O64.4XX3 | DX10 | Obstructed labor due to shoulder presentation, fetus 3 | Pregnancy | O644XX3 |
| O64.4XX4 | DX10 | Obstructed labor due to shoulder presentation, fetus 4 | Pregnancy | O644XX4 |
| O64.4XX5 | DX10 | Obstructed labor due to shoulder presentation, fetus 5 | Pregnancy | O644XX5 |
| O64.4XX9 | DX10 | Obstructed labor due to shoulder presentation, other fetus | Pregnancy | O644XX9 |
| O64.5XX0 | DX10 | Obstructed labor due to compound presentation, unsp | Pregnancy | O645XX0 |
| O64.5XX1 | DX10 | Obstructed labor due to compound presentation, fetus 1 | Pregnancy | O645XX1 |
| O64.5XX2 | DX10 | Obstructed labor due to compound presentation, fetus 2 | Pregnancy | O645XX2 |
| O64.5XX3 | DX10 | Obstructed labor due to compound presentation, fetus 3 | Pregnancy | O645XX3 |
| O64.5XX4 | DX10 | Obstructed labor due to compound presentation, fetus 4 | Pregnancy | O645XX4 |
| O64.5XX5 | DX10 | Obstructed labor due to compound presentation, fetus 5 | Pregnancy | O645XX5 |
| O64.5XX9 | DX10 | Obstructed labor due to compound presentation, other fetus | Pregnancy | O645XX9 |
| O64.8XX0 | DX10 | Obstructed labor due to oth malposition and malpresent, unsp | Pregnancy | O648XX0 |
| O64.8XX1 | DX10 | Obstructed labor due to oth malpos and malpresent, fetus 1 | Pregnancy | O648XX1 |
| O64.8XX2 | DX10 | Obstructed labor due to oth malpos and malpresent, fetus 2 | Pregnancy | O648XX2 |
| O64.8XX3 | DX10 | Obstructed labor due to oth malpos and malpresent, fetus 3 | Pregnancy | O648XX3 |
| O64.8XX4 | DX10 | Obstructed labor due to oth malpos and malpresent, fetus 4 | Pregnancy | O648XX4 |
| O64.8XX5 | DX10 | Obstructed labor due to oth malpos and malpresent, fetus 5 | Pregnancy | O648XX5 |
| O64.8XX9 | DX10 | Obstructed labor due to oth malposition and malpresent, oth | Pregnancy | O648XX9 |

| | | | | |
|---|---|---|---|---|
| O64.9XX0 | DX10 | Obstructed labor due to malpos and malpresent, unsp, unsp | Pregnancy | O649XX0 |
| O64.9XX1 | DX10 | Obstructed labor due to malpos and malpresent, unsp, fetus 1 | Pregnancy | O649XX1 |
| O64.9XX2 | DX10 | Obstructed labor due to malpos and malpresent, unsp, fetus 2 | Pregnancy | O649XX2 |
| O64.9XX3 | DX10 | Obstructed labor due to malpos and malpresent, unsp, fetus 3 | Pregnancy | O649XX3 |
| O64.9XX4 | DX10 | Obstructed labor due to malpos and malpresent, unsp, fetus 4 | Pregnancy | O649XX4 |
| O64.9XX5 | DX10 | Obstructed labor due to malpos and malpresent, unsp, fetus 5 | Pregnancy | O649XX5 |
| O64.9XX9 | DX10 | Obstructed labor due to malpos and malpresent, unsp, oth | Pregnancy | O649XX9 |
| O65.0 | DX10 | Obstructed labor due to deformed pelvis | Pregnancy | O650 |
| O65.1 | DX10 | Obstructed labor due to generally contracted pelvis | Pregnancy | O651 |
| O65.2 | DX10 | Obstructed labor due to pelvic inlet contraction | Pregnancy | O652 |
| O65.3 | DX10 | Obst labor due to pelvic outlet and mid-cavity contrctn | Pregnancy | O653 |
| O65.4 | DX10 | Obstructed labor due to fetopelvic disproportion, unsp | Pregnancy | O654 |
| O65.5 | DX10 | Obstructed labor due to abnlt of maternal pelvic organs | Pregnancy | O655 |
| O65.8 | DX10 | Obstructed labor due to other maternal pelvic abnormalities | Pregnancy | O658 |
| O65.9 | DX10 | Obstructed labor due to maternal pelvic abnormality, unsp | Pregnancy | O659 |
| O66.0 | DX10 | Obstructed labor due to shoulder dystocia | Pregnancy | O660 |
| O66.1 | DX10 | Obstructed labor due to locked twins | Pregnancy | O661 |
| O66.2 | DX10 | Obstructed labor due to unusually large fetus | Pregnancy | O662 |
| O66.3 | DX10 | Obstructed labor due to other abnormalities of fetus | Pregnancy | O663 |
| O66.40 | DX10 | Failed trial of labor, unspecified | Pregnancy | O6640 |
| O66.41 | DX10 | Failed attempt vaginal birth after previous cesarean del | Pregnancy | O6641 |
| O66.5 | DX10 | Attempted application of vacuum extractor and forceps | Pregnancy | O665 |
| O66.6 | DX10 | Obstructed labor due to other multiple fetuses | Pregnancy | O666 |
| O66.8 | DX10 | Other specified obstructed labor | Pregnancy | O668 |
| O66.9 | DX10 | Obstructed labor, unspecified | Pregnancy | O669 |
| O67.0 | DX10 | Intrapartum hemorrhage with coagulation defect | Pregnancy | O670 |
| O67.8 | DX10 | Other intrapartum hemorrhage | Pregnancy | O678 |
| O67.9 | DX10 | Intrapartum hemorrhage, unspecified | Pregnancy | O679 |
| O68 | DX10 | Labor and delivery comp by abnlt of fetal acid-base balance | Pregnancy | O68 |
| O69.0XX0 | DX10 | Labor and delivery complicated by prolapse of cord, unsp | Pregnancy | O690XX0 |
| O69.0XX1 | DX10 | Labor and delivery complicated by prolapse of cord, fetus 1 | Pregnancy | O690XX1 |
| O69.0XX2 | DX10 | Labor and delivery complicated by prolapse of cord, fetus 2 | Pregnancy | O690XX2 |
| O69.0XX3 | DX10 | Labor and delivery complicated by prolapse of cord, fetus 3 | Pregnancy | O690XX3 |
| O69.0XX4 | DX10 | Labor and delivery complicated by prolapse of cord, fetus 4 | Pregnancy | O690XX4 |
| O69.0XX5 | DX10 | Labor and delivery complicated by prolapse of cord, fetus 5 | Pregnancy | O690XX5 |
| O69.0XX9 | DX10 | Labor and delivery complicated by prolapse of cord, oth | Pregnancy | O690XX9 |
| O69.1XX0 | DX10 | Labor and delivery comp by cord around neck, w comprsn, unsp | Pregnancy | O691XX0 |
| O69.1XX1 | DX10 | Labor and del comp by cord around neck, w comprsn, fetus 1 | Pregnancy | O691XX1 |
| O69.1XX2 | DX10 | Labor and del comp by cord around neck, w comprsn, fetus 2 | Pregnancy | O691XX2 |

| | | | | |
|---|---|---|---|---|
| O69.1XX3 | DX10 | Labor and del comp by cord around neck, w comprsn, fetus 3 | Pregnancy | O691XX3 |
| O69.1XX4 | DX10 | Labor and del comp by cord around neck, w comprsn, fetus 4 | Pregnancy | O691XX4 |
| O69.1XX5 | DX10 | Labor and del comp by cord around neck, w comprsn, fetus 5 | Pregnancy | O691XX5 |
| O69.1XX9 | DX10 | Labor and delivery comp by cord around neck, w comprsn, oth | Pregnancy | O691XX9 |
| O69.2XX0 | DX10 | Labor and del comp by oth cord entangle, w comprsn, unsp | Pregnancy | O692XX0 |
| O69.2XX1 | DX10 | Labor and del comp by oth cord entangle, w comprsn, fetus 1 | Pregnancy | O692XX1 |
| O69.2XX2 | DX10 | Labor and del comp by oth cord entangle, w comprsn, fetus 2 | Pregnancy | O692XX2 |
| O69.2XX3 | DX10 | Labor and del comp by oth cord entangle, w comprsn, fetus 3 | Pregnancy | O692XX3 |
| O69.2XX4 | DX10 | Labor and del comp by oth cord entangle, w comprsn, fetus 4 | Pregnancy | O692XX4 |
| O69.2XX5 | DX10 | Labor and del comp by oth cord entangle, w comprsn, fetus 5 | Pregnancy | O692XX5 |
| O69.2XX9 | DX10 | Labor and delivery comp by oth cord entangle, w comprsn, oth | Pregnancy | O692XX9 |
| O69.3XX0 | DX10 | Labor and delivery complicated by short cord, unsp | Pregnancy | O693XX0 |
| O69.3XX1 | DX10 | Labor and delivery complicated by short cord, fetus 1 | Pregnancy | O693XX1 |
| O69.3XX2 | DX10 | Labor and delivery complicated by short cord, fetus 2 | Pregnancy | O693XX2 |
| O69.3XX3 | DX10 | Labor and delivery complicated by short cord, fetus 3 | Pregnancy | O693XX3 |
| O69.3XX4 | DX10 | Labor and delivery complicated by short cord, fetus 4 | Pregnancy | O693XX4 |
| O69.3XX5 | DX10 | Labor and delivery complicated by short cord, fetus 5 | Pregnancy | O693XX5 |
| O69.3XX9 | DX10 | Labor and delivery complicated by short cord, other fetus | Pregnancy | O693XX9 |
| O69.4XX0 | DX10 | Labor and delivery complicated by vasa previa, unsp | Pregnancy | O694XX0 |
| O69.4XX1 | DX10 | Labor and delivery complicated by vasa previa, fetus 1 | Pregnancy | O694XX1 |
| O69.4XX2 | DX10 | Labor and delivery complicated by vasa previa, fetus 2 | Pregnancy | O694XX2 |
| O69.4XX3 | DX10 | Labor and delivery complicated by vasa previa, fetus 3 | Pregnancy | O694XX3 |
| O69.4XX4 | DX10 | Labor and delivery complicated by vasa previa, fetus 4 | Pregnancy | O694XX4 |
| O69.4XX5 | DX10 | Labor and delivery complicated by vasa previa, fetus 5 | Pregnancy | O694XX5 |
| O69.4XX9 | DX10 | Labor and delivery complicated by vasa previa, other fetus | Pregnancy | O694XX9 |
| O69.5XX0 | DX10 | Labor and delivery comp by vascular lesion of cord, unsp | Pregnancy | O695XX0 |
| O69.5XX1 | DX10 | Labor and delivery comp by vascular lesion of cord, fetus 1 | Pregnancy | O695XX1 |
| O69.5XX2 | DX10 | Labor and delivery comp by vascular lesion of cord, fetus 2 | Pregnancy | O695XX2 |
| O69.5XX3 | DX10 | Labor and delivery comp by vascular lesion of cord, fetus 3 | Pregnancy | O695XX3 |
| O69.5XX4 | DX10 | Labor and delivery comp by vascular lesion of cord, fetus 4 | Pregnancy | O695XX4 |
| O69.5XX5 | DX10 | Labor and delivery comp by vascular lesion of cord, fetus 5 | Pregnancy | O695XX5 |
| O69.5XX9 | DX10 | Labor and delivery comp by vascular lesion of cord, oth | Pregnancy | O695XX9 |
| O69.81X0 | DX10 | Labor and del comp by cord around neck, w/o comprsn, unsp | Pregnancy | O6981X0 |
| O69.81X1 | DX10 | Labor and del comp by cord around neck, w/o comprsn, fetus 1 | Pregnancy | O6981X1 |
| O69.81X2 | DX10 | Labor and del comp by cord around neck, w/o comprsn, fetus 2 | Pregnancy | O6981X2 |
| O69.81X3 | DX10 | Labor and del comp by cord around neck, w/o comprsn, fetus 3 | Pregnancy | O6981X3 |

| | | | | |
|---|---|---|---|---|
| O69.81X4 | DX10 | Labor and del comp by cord around neck, w/o comprsn, fetus 4 | Pregnancy | O6981X4 |
| O69.81X5 | DX10 | Labor and del comp by cord around neck, w/o comprsn, fetus 5 | Pregnancy | O6981X5 |
| O69.81X9 | DX10 | Labor and del comp by cord around neck, w/o comprsn, oth | Pregnancy | O6981X9 |
| O69.82X0 | DX10 | Labor and del comp by oth cord entangle, w/o comprsn, unsp | Pregnancy | O6982X0 |
| O69.82X1 | DX10 | Labor and del comp by oth cord entangle, w/o comprsn, fts1 | Pregnancy | O6982X1 |
| O69.82X2 | DX10 | Labor and del comp by oth cord entangle, w/o comprsn, fts2 | Pregnancy | O6982X2 |
| O69.82X3 | DX10 | Labor and del comp by oth cord entangle, w/o comprsn, fts3 | Pregnancy | O6982X3 |
| O69.82X4 | DX10 | Labor and del comp by oth cord entangle, w/o comprsn, fts4 | Pregnancy | O6982X4 |
| O69.82X5 | DX10 | Labor and del comp by oth cord entangle, w/o comprsn, fts5 | Pregnancy | O6982X5 |
| O69.82X9 | DX10 | Labor and del comp by oth cord entangle, w/o comprsn, oth | Pregnancy | O6982X9 |
| O69.89X0 | DX10 | Labor and delivery complicated by oth cord comp, unsp | Pregnancy | O6989X0 |
| O69.89X1 | DX10 | Labor and delivery complicated by oth cord comp, fetus 1 | Pregnancy | O6989X1 |
| O69.89X2 | DX10 | Labor and delivery complicated by oth cord comp, fetus 2 | Pregnancy | O6989X2 |
| O69.89X3 | DX10 | Labor and delivery complicated by oth cord comp, fetus 3 | Pregnancy | O6989X3 |
| O69.89X4 | DX10 | Labor and delivery complicated by oth cord comp, fetus 4 | Pregnancy | O6989X4 |
| O69.89X5 | DX10 | Labor and delivery complicated by oth cord comp, fetus 5 | Pregnancy | O6989X5 |
| O69.89X9 | DX10 | Labor and delivery complicated by oth cord comp, oth | Pregnancy | O6989X9 |
| O69.9XX0 | DX10 | Labor and delivery complicated by cord comp, unsp, unsp | Pregnancy | O699XX0 |
| O69.9XX1 | DX10 | Labor and delivery complicated by cord comp, unsp, fetus 1 | Pregnancy | O699XX1 |
| O69.9XX2 | DX10 | Labor and delivery complicated by cord comp, unsp, fetus 2 | Pregnancy | O699XX2 |
| O69.9XX3 | DX10 | Labor and delivery complicated by cord comp, unsp, fetus 3 | Pregnancy | O699XX3 |
| O69.9XX4 | DX10 | Labor and delivery complicated by cord comp, unsp, fetus 4 | Pregnancy | O699XX4 |
| O69.9XX5 | DX10 | Labor and delivery complicated by cord comp, unsp, fetus 5 | Pregnancy | O699XX5 |
| O69.9XX9 | DX10 | Labor and delivery complicated by cord comp, unsp, oth | Pregnancy | O699XX9 |
| O70.0 | DX10 | First degree perineal laceration during delivery | Pregnancy | O700 |
| O70.1 | DX10 | Second degree perineal laceration during delivery | Pregnancy | O701 |
| O70.2 | DX10 | Third degree perineal laceration during delivery | Pregnancy | O702 |
| O70.20 | DX10 | Third degree perineal laceration during delivery, unspecified | Pregnancy | O7020 |
| O70.21 | DX10 | Third degree perineal laceration during delivery, IIIa | Pregnancy | O7021 |
| O70.22 | DX10 | Third degree perineal laceration during delivery, IIIb | Pregnancy | O7022 |
| O70.23 | DX10 | Third degree perineal laceration during delivery, IIIc | Pregnancy | O7023 |
| O70.3 | DX10 | Fourth degree perineal laceration during delivery | Pregnancy | O703 |
| O70.4 | DX10 | Anal sphincter tear comp del, not assoc w third degree lac | Pregnancy | O704 |
| O70.9 | DX10 | Perineal laceration during delivery, unspecified | Pregnancy | O709 |

| | | | | |
|---|---|---|---|---|
| O71.00 | DX10 | Rupture of uterus before onset of labor, unsp trimester | Pregnancy | O7100 |
| O71.02 | DX10 | Rupture of uterus before onset of labor, second trimester | Pregnancy | O7102 |
| O71.03 | DX10 | Rupture of uterus before onset of labor, third trimester | Pregnancy | O7103 |
| O71.1 | DX10 | Rupture of uterus during labor | Pregnancy | O711 |
| O71.2 | DX10 | Postpartum inversion of uterus | Pregnancy | O712 |
| O71.3 | DX10 | Obstetric laceration of cervix | Pregnancy | O713 |
| O71.4 | DX10 | Obstetric high vaginal laceration alone | Pregnancy | O714 |
| O71.5 | DX10 | Other obstetric injury to pelvic organs | Pregnancy | O715 |
| O71.6 | DX10 | Obstetric damage to pelvic joints and ligaments | Pregnancy | O716 |
| O71.7 | DX10 | Obstetric hematoma of pelvis | Pregnancy | O717 |
| O71.81 | DX10 | Laceration of uterus, not elsewhere classified | Pregnancy | O7181 |
| O71.82 | DX10 | Other specified trauma to perineum and vulva | Pregnancy | O7182 |
| O71.89 | DX10 | Other specified obstetric trauma | Pregnancy | O7189 |
| O71.9 | DX10 | Obstetric trauma, unspecified | Pregnancy | O719 |
| O72.0 | DX10 | Third-stage hemorrhage | Pregnancy | O720 |
| O72.1 | DX10 | Other immediate postpartum hemorrhage | Pregnancy | O721 |
| O72.2 | DX10 | Delayed and secondary postpartum hemorrhage | Pregnancy | O722 |
| O72.3 | DX10 | Postpartum coagulation defects | Pregnancy | O723 |
| O73.0 | DX10 | Retained placenta without hemorrhage | Pregnancy | O730 |
| O73.1 | DX10 | Retained portions of placenta and membranes, w/o hemorrhage | Pregnancy | O731 |
| O74.0 | DX10 | Aspirat pneumonitis due to anesth during labor and delivery | Pregnancy | O740 |
| O74.1 | DX10 | Oth pulmonary comp of anesthesia during labor and delivery | Pregnancy | O741 |
| O74.2 | DX10 | Cardiac comp of anesthesia during labor and delivery | Pregnancy | O742 |
| O74.3 | DX10 | Cnsl complications of anesthesia during labor and delivery | Pregnancy | O743 |
| O74.4 | DX10 | Toxic reaction to local anesthesia during labor and delivery | Pregnancy | O744 |
| O74.5 | DX10 | Spinal and epidur anesthesia-induced hdache dur labr and del | Pregnancy | O745 |
| O74.6 | DX10 | Oth comp of spinal and epidural anesth during labor and del | Pregnancy | O746 |
| O74.7 | DX10 | Failed or difficult intubation for anesth dur labor and del | Pregnancy | O747 |
| O74.8 | DX10 | Other complications of anesthesia during labor and delivery | Pregnancy | O748 |
| O74.9 | DX10 | Complication of anesthesia during labor and delivery, unsp | Pregnancy | O749 |
| O75.0 | DX10 | Maternal distress during labor and delivery | Pregnancy | O750 |
| O75.1 | DX10 | Shock during or following labor and delivery | Pregnancy | O751 |
| O75.2 | DX10 | Pyrexia during labor, not elsewhere classified | Pregnancy | O752 |
| O75.3 | DX10 | Other infection during labor | Pregnancy | O753 |
| O75.4 | DX10 | Other complications of obstetric surgery and procedures | Pregnancy | O754 |
| O75.81 | DX10 | Maternal exhaustion complicating labor and delivery | Pregnancy | O7581 |
| O75.89 | DX10 | Other specified complications of labor and delivery | Pregnancy | O7589 |
| O75.9 | DX10 | Complication of labor and delivery, unspecified | Pregnancy | O759 |
| O76 | DX10 | Abnlt in fetal heart rate and rhythm comp labor and delivery | Pregnancy | O76 |
| O77.0 | DX10 | Labor and delivery complicated by meconium in amniotic fluid | Pregnancy | O770 |
| O77.1 | DX10 | Fetal stress in labor or delivery due to drug administration | Pregnancy | O771 |
| O77.8 | DX10 | Labor and delivery comp by oth evidence of fetal stress | Pregnancy | O778 |
| O77.9 | DX10 | Labor and delivery complicated by fetal stress, unspecified | Pregnancy | O779 |
| O82 | DX10 | Encounter for cesarean delivery without indication | Pregnancy | O82 |

| | | | | |
|---|---|---|---|---|
| O86.0 | DX10 | Infection of obstetric surgical wound | Pregnancy | O860 |
| O86.11 | DX10 | Cervicitis following delivery | Pregnancy | O8611 |
| O86.12 | DX10 | Endometritis following delivery | Pregnancy | O8612 |
| O86.13 | DX10 | Vaginitis following delivery | Pregnancy | O8613 |
| O86.19 | DX10 | Other infection of genital tract following delivery | Pregnancy | O8619 |
| O86.20 | DX10 | Urinary tract infection following delivery, unspecified | Pregnancy | O8620 |
| O86.21 | DX10 | Infection of kidney following delivery | Pregnancy | O8621 |
| O86.22 | DX10 | Infection of bladder following delivery | Pregnancy | O8622 |
| O86.29 | DX10 | Other urinary tract infection following delivery | Pregnancy | O8629 |
| O86.4 | DX10 | Pyrexia of unknown origin following delivery | Pregnancy | O864 |
| O86.81 | DX10 | Puerperal septic thrombophlebitis | Pregnancy | O8681 |
| O86.89 | DX10 | Other specified puerperal infections | Pregnancy | O8689 |
| O87.0 | DX10 | Superficial thrombophlebitis in the puerperium | Pregnancy | O870 |
| O87.1 | DX10 | Deep phlebothrombosis in the puerperium | Pregnancy | O871 |
| O87.2 | DX10 | Hemorrhoids in the puerperium | Pregnancy | O872 |
| O87.3 | DX10 | Cerebral venous thrombosis in the puerperium | Pregnancy | O873 |
| O87.4 | DX10 | Varicose veins of lower extremity in the puerperium | Pregnancy | O874 |
| O87.8 | DX10 | Other venous complications in the puerperium | Pregnancy | O878 |
| O87.9 | DX10 | Venous complication in the puerperium, unspecified | Pregnancy | O879 |
| O88.011 | DX10 | Air embolism in pregnancy, first trimester | Pregnancy | O88011 |
| O88.012 | DX10 | Air embolism in pregnancy, second trimester | Pregnancy | O88012 |
| O88.013 | DX10 | Air embolism in pregnancy, third trimester | Pregnancy | O88013 |
| O88.019 | DX10 | Air embolism in pregnancy, unspecified trimester | Pregnancy | O88019 |
| O88.02 | DX10 | Air embolism in childbirth | Pregnancy | O8802 |
| O88.03 | DX10 | Air embolism in the puerperium | Pregnancy | O8803 |
| O88.111 | DX10 | Amniotic fluid embolism in pregnancy, first trimester | Pregnancy | O88111 |
| O88.112 | DX10 | Amniotic fluid embolism in pregnancy, second trimester | Pregnancy | O88112 |
| O88.113 | DX10 | Amniotic fluid embolism in pregnancy, third trimester | Pregnancy | O88113 |
| O88.119 | DX10 | Amniotic fluid embolism in pregnancy, unspecified trimester | Pregnancy | O88119 |
| O88.12 | DX10 | Amniotic fluid embolism in childbirth | Pregnancy | O8812 |
| O88.13 | DX10 | Amniotic fluid embolism in the puerperium | Pregnancy | O8813 |
| O88.211 | DX10 | Thromboembolism in pregnancy, first trimester | Pregnancy | O88211 |
| O88.212 | DX10 | Thromboembolism in pregnancy, second trimester | Pregnancy | O88212 |
| O88.213 | DX10 | Thromboembolism in pregnancy, third trimester | Pregnancy | O88213 |
| O88.219 | DX10 | Thromboembolism in pregnancy, unspecified trimester | Pregnancy | O88219 |
| O88.22 | DX10 | Thromboembolism in childbirth | Pregnancy | O8822 |
| O88.23 | DX10 | Thromboembolism in the puerperium | Pregnancy | O8823 |
| O88.311 | DX10 | Pyemic and septic embolism in pregnancy, first trimester | Pregnancy | O88311 |
| O88.312 | DX10 | Pyemic and septic embolism in pregnancy, second trimester | Pregnancy | O88312 |
| O88.313 | DX10 | Pyemic and septic embolism in pregnancy, third trimester | Pregnancy | O88313 |
| O88.319 | DX10 | Pyemic and septic embolism in pregnancy, unsp trimester | Pregnancy | O88319 |
| O88.32 | DX10 | Pyemic and septic embolism in childbirth | Pregnancy | O8832 |
| O88.33 | DX10 | Pyemic and septic embolism in the puerperium | Pregnancy | O8833 |
| O88.811 | DX10 | Other embolism in pregnancy, first trimester | Pregnancy | O88811 |
| O88.812 | DX10 | Other embolism in pregnancy, second trimester | Pregnancy | O88812 |
| O88.813 | DX10 | Other embolism in pregnancy, third trimester | Pregnancy | O88813 |
| O88.819 | DX10 | Other embolism in pregnancy, unspecified trimester | Pregnancy | O88819 |
| O88.82 | DX10 | Other embolism in childbirth | Pregnancy | O8882 |
| O88.83 | DX10 | Other embolism in the puerperium | Pregnancy | O8883 |
| O89.01 | DX10 | Aspiration pneumonitis due to anesth during the puerperium | Pregnancy | O8901 |
| O89.09 | DX10 | Oth pulmonary comp of anesthesia during the puerperium | Pregnancy | O8909 |

| | | | | |
|---|---|---|---|---|
| O89.1 | DX10 | Cardiac complications of anesthesia during the puerperium | Pregnancy | O891 |
| O89.2 | DX10 | Cnsl complications of anesthesia during the puerperium | Pregnancy | O892 |
| O89.3 | DX10 | Toxic reaction to local anesthesia during the puerperium | Pregnancy | O893 |
| O89.4 | DX10 | Spinal and epidur anesthesia-induced hdache during the puerp | Pregnancy | O894 |
| O89.5 | DX10 | Oth comp of spinal and epidural anesth during the puerperium | Pregnancy | O895 |
| O89.6 | DX10 | Failed or difficult intubation for anesth during the puerp | Pregnancy | O896 |
| O89.8 | DX10 | Other complications of anesthesia during the puerperium | Pregnancy | O898 |
| O89.9 | DX10 | Complication of anesthesia during the puerperium, unsp | Pregnancy | O899 |
| O90.0 | DX10 | Disruption of cesarean delivery wound | Pregnancy | O900 |
| O90.1 | DX10 | Disruption of perineal obstetric wound | Pregnancy | O901 |
| O90.2 | DX10 | Hematoma of obstetric wound | Pregnancy | O902 |
| O90.4 | DX10 | Postpartum acute kidney failure | Pregnancy | O904 |
| O90.5 | DX10 | Postpartum thyroiditis | Pregnancy | O905 |
| O90.6 | DX10 | Postpartum mood disturbance | Pregnancy | O906 |
| O90.81 | DX10 | Anemia of the puerperium | Pregnancy | O9081 |
| O90.89 | DX10 | Oth complications of the puerperium, NEC | Pregnancy | O9089 |
| O91.011 | DX10 | Infection of nipple associated w pregnancy, first trimester | Pregnancy | O91011 |
| O91.012 | DX10 | Infection of nipple associated w pregnancy, second trimester | Pregnancy | O91012 |
| O91.013 | DX10 | Infection of nipple associated w pregnancy, third trimester | Pregnancy | O91013 |
| O91.019 | DX10 | Infection of nipple associated w pregnancy, unsp trimester | Pregnancy | O91019 |
| O91.02 | DX10 | Infection of nipple associated with the puerperium | Pregnancy | O9102 |
| O91.03 | DX10 | Infection of nipple associated with lactation | Pregnancy | O9103 |
| O91.111 | DX10 | Abscess of breast associated with pregnancy, first trimester | Pregnancy | O91111 |
| O91.112 | DX10 | Abscess of breast associated w pregnancy, second trimester | Pregnancy | O91112 |
| O91.113 | DX10 | Abscess of breast associated with pregnancy, third trimester | Pregnancy | O91113 |
| O91.119 | DX10 | Abscess of breast associated with pregnancy, unsp trimester | Pregnancy | O91119 |
| O91.12 | DX10 | Abscess of breast associated with the puerperium | Pregnancy | O9112 |
| O91.13 | DX10 | Abscess of breast associated with lactation | Pregnancy | O9113 |
| O91.211 | DX10 | Nonpurulent mastitis associated w pregnancy, first trimester | Pregnancy | O91211 |
| O91.212 | DX10 | Nonpurulent mastitis assoc w pregnancy, second trimester | Pregnancy | O91212 |
| O91.213 | DX10 | Nonpurulent mastitis associated w pregnancy, third trimester | Pregnancy | O91213 |
| O91.219 | DX10 | Nonpurulent mastitis associated w pregnancy, unsp trimester | Pregnancy | O91219 |
| O91.22 | DX10 | Nonpurulent mastitis associated with the puerperium | Pregnancy | O9122 |
| O91.23 | DX10 | Nonpurulent mastitis associated with lactation | Pregnancy | O9123 |
| O92.011 | DX10 | Retracted nipple associated with pregnancy, first trimester | Pregnancy | O92011 |
| O92.012 | DX10 | Retracted nipple associated with pregnancy, second trimester | Pregnancy | O92012 |
| O92.013 | DX10 | Retracted nipple associated with pregnancy, third trimester | Pregnancy | O92013 |

| | | | | |
|---|---|---|---|---|
| O92.019 | DX10 | Retracted nipple associated with pregnancy, unsp trimester | Pregnancy | O92019 |
| O92.02 | DX10 | Retracted nipple associated with the puerperium | Pregnancy | O9202 |
| O92.03 | DX10 | Retracted nipple associated with lactation | Pregnancy | O9203 |
| O92.111 | DX10 | Cracked nipple associated with pregnancy, first trimester | Pregnancy | O92111 |
| O92.112 | DX10 | Cracked nipple associated with pregnancy, second trimester | Pregnancy | O92112 |
| O92.113 | DX10 | Cracked nipple associated with pregnancy, third trimester | Pregnancy | O92113 |
| O92.119 | DX10 | Cracked nipple associated with pregnancy, unsp trimester | Pregnancy | O92119 |
| O92.12 | DX10 | Cracked nipple associated with the puerperium | Pregnancy | O9212 |
| O92.13 | DX10 | Cracked nipple associated with lactation | Pregnancy | O9213 |
| O99.810 | DX10 | Abnormal glucose complicating pregnancy | Pregnancy | O99810 |
| O99.814 | DX10 | Abnormal glucose complicating childbirth | Pregnancy | O99814 |
| O99.815 | DX10 | Abnormal glucose complicating the puerperium | Pregnancy | O99815 |
| O99.820 | DX10 | Streptococcus B carrier state complicating pregnancy | Pregnancy | O99820 |
| O99.824 | DX10 | Streptococcus B carrier state complicating childbirth | Pregnancy | O99824 |
| O99.825 | DX10 | Streptococcus B carrier state complicating the puerperium | Pregnancy | O99825 |
| O99.830 | DX10 | Other infection carrier state complicating pregnancy | Pregnancy | O99830 |
| O99.834 | DX10 | Other infection carrier state complicating childbirth | Pregnancy | O99834 |
| O99.835 | DX10 | Other infection carrier state complicating the puerperium | Pregnancy | O99835 |
| P07.20 | DX10 | P07.20(Extreme immaturity of newborn | Pregnancy | P0720 |
| P07.21 | DX10 | P07.21(Extreme immaturity of NB, gestational age less than 23 completed weeks | Pregnancy | P0721 |
| P07.22 | DX10 | P07.22(Extreme immaturity of NB, gestational age 23 completed weeks | Pregnancy | P0722 |
| P07.23 | DX10 | P07.23(Extreme immaturity of NB, gestational age 24 completed weeks | Pregnancy | P0723 |
| P07.24 | DX10 | P07.24(Extreme immaturity of NB, gestational age 25 completed weeks | Pregnancy | P0724 |
| P07.25 | DX10 | P07.25(Extreme immaturity of NB, gestational age 26 completed weeks | Pregnancy | P0725 |
| P07.26 | DX10 | P07.26(Extreme immaturity of NB, gestational age 27 completed weeks | Pregnancy | P0726 |
| P07.30 | DX10 | P07.30(Preterm newborn, unspecified weeks of gestation | Pregnancy | P0730 |
| P07.31 | DX10 | P07.31(Preterm newborn, gestational age 28 weeks completed | Pregnancy | P0731 |
| P07.32 | DX10 | P07.32(Preterm newborn, gestational age 29 weeks completed | Pregnancy | P0732 |
| P07.33 | DX10 | P07.33(Preterm newborn, gestational age 30 weeks completed | Pregnancy | P0733 |
| P07.34 | DX10 | P07.34(Preterm newborn, gestational age 31 weeks completed | Pregnancy | P0734 |
| P07.35 | DX10 | P07.35(Preterm newborn, gestational age 32 weeks completed | Pregnancy | P0735 |
| P07.36 | DX10 | P07.36(Preterm newborn, gestational age 33 weeks completed | Pregnancy | P0736 |
| P07.37 | DX10 | P07.37(Preterm newborn, gestational age 34 weeks completed | Pregnancy | P0737 |
| P07.38 | DX10 | P07.38(Preterm newborn, gestational age 35 weeks completed | Pregnancy | P0738 |
| P07.39 | DX10 | P07.39(Preterm newborn, gestational age 36 weeks completed | Pregnancy | P0739 |
| P84 | DX10 | Other problems with newborn | Pregnancy | P84 |
| P95 | DX10 | STILLBIRTH | Pregnancy | P95 |
| Z32.01 | DX10 | Encounter for pregnancy test, result positive | Pregnancy | Z3201 |

| | | | | |
|---|---|---|---|---|
| Z33.1 | DX10 | Pregnant state, incidental | Pregnancy | Z331 |
| Z33.2 | DX10 | Encounter for elective termination of pregnancy | Pregnancy | Z332 |
| Z33.3 | DX10 | pregnant state, gestational carrier | Pregnancy | Z333 |
| Z34.00 | DX10 | Encntr for suprvsn of normal first pregnancy, unsp trimester | Pregnancy | Z3400 |
| Z34.01 | DX10 | Encntr for suprvsn of normal first preg, first trimester | Pregnancy | Z3401 |
| Z34.02 | DX10 | Encntr for suprvsn of normal first preg, second trimester | Pregnancy | Z3402 |
| Z34.03 | DX10 | Encntr for suprvsn of normal first preg, third trimester | Pregnancy | Z3403 |
| Z34.80 | DX10 | Encounter for suprvsn of normal pregnancy, unsp trimester | Pregnancy | Z3480 |
| Z34.81 | DX10 | Encounter for suprvsn of normal pregnancy, first trimester | Pregnancy | Z3481 |
| Z34.82 | DX10 | Encounter for suprvsn of normal pregnancy, second trimester | Pregnancy | Z3482 |
| Z34.83 | DX10 | Encounter for suprvsn of normal pregnancy, third trimester | Pregnancy | Z3483 |
| Z34.90 | DX10 | Encntr for suprvsn of normal pregnancy, unsp, unsp trimester | Pregnancy | Z3490 |
| Z34.91 | DX10 | Encntr for suprvsn of normal preg, unsp, first trimester | Pregnancy | Z3491 |
| Z34.92 | DX10 | Encntr for suprvsn of normal preg, unsp, second trimester | Pregnancy | Z3492 |
| Z34.93 | DX10 | Encntr for suprvsn of normal preg, unsp, third trimester | Pregnancy | Z3493 |
| Z36 | DX10 | Encounter for antenatal screening of mother | Pregnancy | Z36 |
| Z38.00 | DX10 | Single liveborn infant, delivered vaginally | Pregnancy | Z3800 |
| Z38.01 | DX10 | Single liveborn infant, delivered by cesarean | Pregnancy | Z3801 |
| Z38.1 | DX10 | Single liveborn infant, born outside hospital | Pregnancy | Z381 |
| Z38.2 | DX10 | Single liveborn infant, unspecified as to place of birth | Pregnancy | Z382 |
| Z38.30 | DX10 | Twin liveborn infant, delivered vaginally | Pregnancy | Z3830 |
| Z38.31 | DX10 | Twin liveborn infant, delivered by cesarean | Pregnancy | Z3831 |
| Z38.4 | DX10 | Twin liveborn infant, born outside hospital | Pregnancy | Z384 |
| Z38.5 | DX10 | Twin liveborn infant, unspecified as to place of birth | Pregnancy | Z385 |
| Z38.61 | DX10 | Triplet liveborn infant, delivered vaginally | Pregnancy | Z3861 |
| Z38.62 | DX10 | Triplet liveborn infant, delivered by cesarean | Pregnancy | Z3862 |
| Z38.63 | DX10 | Quadruplet liveborn infant, delivered vaginally | Pregnancy | Z3863 |
| Z38.64 | DX10 | Quadruplet liveborn infant, delivered by cesarean | Pregnancy | Z3864 |
| Z38.65 | DX10 | Quintuplet liveborn infant, delivered vaginally | Pregnancy | Z3865 |
| Z38.66 | DX10 | Quintuplet liveborn infant, delivered by cesarean | Pregnancy | Z3866 |
| Z38.68 | DX10 | Other multiple liveborn infant, delivered vaginally | Pregnancy | Z3868 |
| Z38.69 | DX10 | Other multiple liveborn infant, delivered by cesarean | Pregnancy | Z3869 |
| Z38.7 | DX10 | Other multiple liveborn infant, born outside hospital | Pregnancy | Z387 |
| Z38.8 | DX10 | Other multiple liveborn infant, unsp as to place of birth | Pregnancy | Z388 |
| Z39.0 | DX10 | Encntr for care and exam of mother immediately after del | Pregnancy | Z390 |
| Z39.1 | DX10 | Encounter for care and examination of lactating mother | Pregnancy | Z391 |
| Z39.2 | DX10 | Encounter for routine postpartum follow-up | Pregnancy | Z392 |
| Z3A.00 | DX10 | weeks of gestation of pregnancy, unspecified | Pregnancy | Z3A00 |
| Z3A.01 | DX10 | less than 8 weeks of gestation of pregnancy | Pregnancy | Z3A01 |
| Z3A.08 | DX10 | 8 weeks gestation of pregnancy | Pregnancy | Z3A08 |
| Z3A.09 | DX10 | 9 weeks gestation of pregnancy | Pregnancy | Z3A09 |
| Z3A.10 | DX10 | 10 weeks gestation of pregnancy | Pregnancy | Z3A10 |
| Z3A.11 | DX10 | 11 weeks gestation of pregnancy | Pregnancy | Z3A11 |
| Z3A.12 | DX10 | 12 weeks gestation of pregnancy | Pregnancy | Z3A12 |
| Z3A.13 | DX10 | 13 weeks gestation of pregnancy | Pregnancy | Z3A13 |
| Z3A.14 | DX10 | 14 weeks gestation of pregnancy | Pregnancy | Z3A14 |
| Z3A.15 | DX10 | 15 weeks gestation of pregnancy | Pregnancy | Z3A15 |
| Z3A.16 | DX10 | 16 weeks gestation of pregnancy | Pregnancy | Z3A16 |
| Z3A.17 | DX10 | 17 weeks gestation of pregnancy | Pregnancy | Z3A17 |
| Z3A.18 | DX10 | 18 weeks gestation of pregnancy | Pregnancy | Z3A18 |
| Z3A.19 | DX10 | 19 weeks gestation of pregnancy | Pregnancy | Z3A19 |

| | | | | |
|---|---|---|---|---|
| Z3A.20 | DX10 | 20 weeks gestation of pregnancy | Pregnancy | Z3A20 |
| Z3A.21 | DX10 | 21 weeks gestation of pregnancy | Pregnancy | Z3A21 |
| Z3A.22 | DX10 | 22 weeks gestation of pregnancy | Pregnancy | Z3A22 |
| Z3A.23 | DX10 | 23 weeks gestation of pregnancy | Pregnancy | Z3A23 |
| Z3A.24 | DX10 | 24 weeks gestation of pregnancy | Pregnancy | Z3A24 |
| Z3A.25 | DX10 | 25 weeks gestation of pregnancy | Pregnancy | Z3A25 |
| Z3A.26 | DX10 | 26 weeks gestation of pregnancy | Pregnancy | Z3A26 |
| Z3A.27 | DX10 | 27 weeks gestation of pregnancy | Pregnancy | Z3A27 |
| Z3A.28 | DX10 | 28 weeks gestation of pregnancy | Pregnancy | Z3A28 |
| Z3A.29 | DX10 | 29 weeks gestation of pregnancy | Pregnancy | Z3A29 |
| Z3A.30 | DX10 | 30 weeks gestation of pregnancy | Pregnancy | Z3A30 |
| Z3A.31 | DX10 | 31 weeks gestation of pregnancy | Pregnancy | Z3A31 |
| Z3A.32 | DX10 | 32 weeks gestation of pregnancy | Pregnancy | Z3A32 |
| Z3A.33 | DX10 | 33 weeks gestation of pregnancy | Pregnancy | Z3A33 |
| Z3A.34 | DX10 | 34 weeks gestation of pregnancy | Pregnancy | Z3A34 |
| Z3A.35 | DX10 | 35 weeks gestation of pregnancy | Pregnancy | Z3A35 |
| Z3A.36 | DX10 | 36 weeks gestation of pregnancy | Pregnancy | Z3A36 |
| Z3A.37 | DX10 | 37 weeks gestation of pregnancy | Pregnancy | Z3A37 |
| Z3A.38 | DX10 | 38 weeks gestation of pregnancy | Pregnancy | Z3A38 |
| Z3A.39 | DX10 | 39 weeks gestation of pregnancy | Pregnancy | Z3A39 |
| Z3A.40 | DX10 | 40 weeks gestation of pregnancy | Pregnancy | Z3A40 |
| Z3A.41 | DX10 | 41 weeks gestation of pregnancy | Pregnancy | Z3A41 |
| Z3A.42 | DX10 | 42 weeks gestation of pregnancy | Pregnancy | Z3A42 |
| Z3A.49 | DX10 | Greater than 42 weeks gestation of pregnancy | Pregnancy | Z3A49 |
| 416* | DX10 | PULMCR_ELIX | Pulmonary_Circulation_Disorder | 416* |
| 4179* | DX10 | PULMCR_ELIX | Pulmonary_Circulation_Disorder | 4179* |
| I26 | DX10 | PULMCR_ELIX | Pulmonary_Circulation_Disorder | I26 |
| I260 | DX10 | PULMCR_ELIX | Pulmonary_Circulation_Disorder | I260 |
| I2601 | DX10 | PULMCR_ELIX | Pulmonary_Circulation_Disorder | I2601 |
| I2602 | DX10 | PULMCR_ELIX | Pulmonary_Circulation_Disorder | I2602 |
| I2609 | DX10 | PULMCR_ELIX | Pulmonary_Circulation_Disorder | I2609 |
| I269 | DX10 | PULMCR_ELIX | Pulmonary_Circulation_Disorder | I269 |
| I2690 | DX10 | PULMCR_ELIX | Pulmonary_Circulation_Disorder | I2690 |
| I2692 | DX10 | PULMCR_ELIX | Pulmonary_Circulation_Disorder | I2692 |
| I2699 | DX10 | PULMCR_ELIX | Pulmonary_Circulation_Disorder | I2699 |
| I27 | DX10 | PULMCR_ELIX | Pulmonary_Circulation_Disorder | I27 |
| I270 | DX10 | PULMCR_ELIX | Pulmonary_Circulation_Disorder | I270 |
| I271 | DX10 | PULMCR_ELIX | Pulmonary_Circulation_Disorder | I271 |
| I272 | DX10 | PULMCR_ELIX | Pulmonary_Circulation_Disorder | I272 |
| I278 | DX10 | PULMCR_ELIX | Pulmonary_Circulation_Disorder | I278 |
| I2781 | DX10 | PULMCR_ELIX | Pulmonary_Circulation_Disorder | I2781 |

| | | | | |
|---|---|---|---|---|
| I2782 | DX10 | PULMCR_ELIX | Pulmonary_Circulation_Disorder | I2782 |
| I2789 | DX10 | PULMCR_ELIX | Pulmonary_Circulation_Disorder | I2789 |
| I279 | DX10 | PULMCR_ELIX | Pulmonary_Circulation_Disorder | I279 |
| I280 | DX10 | PULMCR_ELIX | Pulmonary_Circulation_Disorder | I280 |
| I288 | DX10 | PULMCR_ELIX | Pulmonary_Circulation_Disorder | I288 |
| I289 | DX10 | PULMCR_ELIX | Pulmonary_Circulation_Disorder | I289 |
| 278.01 | DX10 | Morbid obesity | SevereObesity_BMI_ge40 | 27801 |
| E66.01 | DX10 | Morbid (severe) obesity due to excess calories | SevereObesity_BMI_ge40 | E6601 |
| E66.2 | DX10 | Morbid (severe) obesity with alveolar hypoventilation | SevereObesity_BMI_ge40 | E662 |
| V85.41 | DX10 | Body Mass Index 40.0-44.9, adult | SevereObesity_BMI_ge40 | V8541 |
| V85.42 | DX10 | Body Mass Index 45.0-49.9, adult | SevereObesity_BMI_ge40 | V8542 |
| V85.43 | DX10 | Body Mass Index 50.0-59.9, adult | SevereObesity_BMI_ge40 | V8543 |
| V85.44 | DX10 | Body Mass Index 60.0-69.9, adult | SevereObesity_BMI_ge40 | V8544 |
| V85.45 | DX10 | Body Mass Index 70 and over, adult | SevereObesity_BMI_ge40 | V8545 |
| Z68.41 | DX10 | Body mass index (BMI) 40.0-44.9, adult | SevereObesity_BMI_ge40 | Z6841 |
| Z68.42 | DX10 | Body mass index (BMI) 45.0-49.9, adult | SevereObesity_BMI_ge40 | Z6842 |
| Z68.43 | DX10 | Body mass index (BMI) 50.0-59.9, adult | SevereObesity_BMI_ge40 | Z6843 |
| Z68.44 | DX10 | Body mass index (BMI) 60.0-69.9, adult | SevereObesity_BMI_ge40 | Z6844 |
| Z68.45 | DX10 | Body mass index (BMI) 70 or greater, adult | SevereObesity_BMI_ge40 | Z6845 |
| **Z72.0** | DX10 | Tobacco use | Smoking | Z720 |
| **Z87.891** | DX10 | Personal history of nicotine dependence | Smoking | Z87891 |
| F17.210 | DX10 | Nicotine Dependence, Cigarettes, uncomplicated | Smoking | F17210 |
| F17.211 | DX10 | Nicotine Dependence, Cigarettes, in remission | Smoking | F17211 |
| F17.213 | DX10 | Nicotine Dependence, Cigarettes, with withdrawal | Smoking | F17213 |
| F17.218 | DX10 | Nicotine Dependence, Cigarettes, with other nicotine-induced disorders | Smoking | F17218 |
| F17.219 | DX10 | Nicotine Dependence, Cigarettes, with other unspecified nicotine-induced disorders | Smoking | F17219 |
| F17.200 | DX10 | Nicotine Dependence, Unspecified, uncomplicated | Smoking | F17200 |
| F17.201 | DX10 | Nicotine Dependence, Unspecified, in remission | Smoking | F17201 |
| F17.203 | DX10 | Nicotine Dependence, Unspecified, with withdrawal | Smoking | F17203 |
| F17.208 | DX10 | Nicotine Dependence, Unspecified, with other nicotine-induced disorders | Smoking | F17208 |
| F17.209 | DX10 | Nicotine Dependence, Unspecified, with other unspecified nicotine-induced disorders | Smoking | F17209 |
| F17 | DX10 | Nicotine dependence | Smoking | F17 |
| F17.2 | DX10 | Nicotine dependence | Smoking | F172 |
| F17.20 | DX10 | Nicotine dependence, unspecified | Smoking | F1720 |
| F17.21 | DX10 | Nicotine dependence, cigarettes | Smoking | F1721 |

| | | | | |
|---|---|---|---|---|
| T86.1 | DX10 | Complications of kidney transplant | SolidOrgan_Hema_Cell_Transplant | T861 |
| T86.10 | DX10 | Unspecified complication of kidney transplant | SolidOrgan_Hema_Cell_Transplant | T8610 |
| T86.11 | DX10 | Kidney transplant rejection | SolidOrgan_Hema_Cell_Transplant | T8611 |
| T86.12 | DX10 | Kidney transplant failure | SolidOrgan_Hema_Cell_Transplant | T8612 |
| T86.13 | DX10 | Kidney transplant infection | SolidOrgan_Hema_Cell_Transplant | T8613 |
| T86.19 | DX10 | Other complication of kidney transplant | SolidOrgan_Hema_Cell_Transplant | T8619 |
| T86.2 | DX10 | Complications of heart transplant | SolidOrgan_Hema_Cell_Transplant | T862 |
| T86.20 | DX10 | Unspecified complication of heart transplant | SolidOrgan_Hema_Cell_Transplant | T8620 |
| T86.21 | DX10 | Heart transplant rejection | SolidOrgan_Hema_Cell_Transplant | T8621 |
| T86.22 | DX10 | Heart transplant failure | SolidOrgan_Hema_Cell_Transplant | T8622 |
| T86.23 | DX10 | Heart transplant infection | SolidOrgan_Hema_Cell_Transplant | T8623 |
| T86.29 | DX10 | Other complications of heart transplant | SolidOrgan_Hema_Cell_Transplant | T8629 |
| T86.290 | DX10 | Cardiac allograft vasculopathy | SolidOrgan_Hema_Cell_Transplant | T86290 |
| T86.298 | DX10 | Other complications of heart transplant | SolidOrgan_Hema_Cell_Transplant | T86298 |
| T86.3 | DX10 | Complications of heart-lung transplant | SolidOrgan_Hema_Cell_Transplant | T863 |
| T86.30 | DX10 | Unspecified complication of heart-lung transplant | SolidOrgan_Hema_Cell_Transplant | T8630 |
| T86.31 | DX10 | Heart-lung transplant rejection | SolidOrgan_Hema_Cell_Transplant | T8631 |
| T86.32 | DX10 | Heart-lung transplant failure | SolidOrgan_Hema_Cell_Transplant | T8632 |
| T86.33 | DX10 | Heart-lung transplant infection | SolidOrgan_Hema_Cell_Transplant | T8633 |
| T86.39 | DX10 | Other complications of heart-lung transplant | SolidOrgan_Hema_Cell_Transplant | T8639 |
| T86.4 | DX10 | Complications of liver transplant | SolidOrgan_Hema_Cell_Transplant | T864 |
| T86.40 | DX10 | Unspecified complication of liver transplant | SolidOrgan_Hema_Cell_Transplant | T8640 |
| T86.41 | DX10 | Liver transplant rejection | SolidOrgan_Hema_Cell_Transplant | T8641 |
| T86.42 | DX10 | Liver transplant failure | SolidOrgan_Hema_Cell_Transplant | T8642 |
| T86.43 | DX10 | Liver transplant infection | SolidOrgan_Hema_Cell_Transplant | T8643 |
| T86.49 | DX10 | Other complications of liver transplant | SolidOrgan_Hema_Cell_Transplant | T8649 |
| T86.8 | DX10 | Complications of other transplanted organs and tissues | SolidOrgan_Hema_Cell_Transplant | T868 |
| T86.81 | DX10 | Complications of lung transplant | SolidOrgan_Hema_Cell_Transplant | T8681 |
| T86.810 | DX10 | Lung transplant rejection | SolidOrgan_Hema_Cell_Transplant | T86810 |

| | | | | |
|---|---|---|---|---|
| T86.811 | DX10 | Lung transplant failure | SolidOrgan_Hema_Cell_Transplant | T86811 |
| T86.812 | DX10 | Lung transplant infection | SolidOrgan_Hema_Cell_Transplant | T86812 |
| T86.818 | DX10 | Other complications of lung transplant | SolidOrgan_Hema_Cell_Transplant | T86818 |
| T86.819 | DX10 | Unspecified complication of lung transplant | SolidOrgan_Hema_Cell_Transplant | T86819 |
| T86.85 | DX10 | Complication of intestine transplant | SolidOrgan_Hema_Cell_Transplant | T8685 |
| T86.850 | DX10 | Intestine transplant rejection | SolidOrgan_Hema_Cell_Transplant | T86850 |
| T86.851 | DX10 | Intestine transplant failure | SolidOrgan_Hema_Cell_Transplant | T86851 |
| T86.852 | DX10 | Intestine transplant infection | SolidOrgan_Hema_Cell_Transplant | T86852 |
| T86.858 | DX10 | Other complications of intestine transplant | SolidOrgan_Hema_Cell_Transplant | T86858 |
| T86.859 | DX10 | Unspecified complication of intestine transplant | SolidOrgan_Hema_Cell_Transplant | T86859 |
| D47.Z1 | DX10 | Post-transplant lymphoproliferative disorder | SolidOrgan_Hema_Cell_Transplant | D47Z1 |
| Z48.2 | DX10 | Encounter for aftercare following organ transplant | SolidOrgan_Hema_Cell_Transplant | Z482 |
| Z94 | DX10 | Transplanted organ and tissue status | SolidOrgan_Hema_Cell_Transplant | Z94 |
| Z94.0 | DX10 | Kidney transplant status | SolidOrgan_Hema_Cell_Transplant | Z940 |
| Z94.1 | DX10 | Heart transplant status | SolidOrgan_Hema_Cell_Transplant | Z941 |
| Z94.2 | DX10 | Lung transplant status | SolidOrgan_Hema_Cell_Transplant | Z942 |
| Z94.3 | DX10 | Heart and lungs transplant status | SolidOrgan_Hema_Cell_Transplant | Z943 |
| Z94.4 | DX10 | Liver transplant status | SolidOrgan_Hema_Cell_Transplant | Z944 |
| Z94.6 | DX10 | Bone transplant status | SolidOrgan_Hema_Cell_Transplant | Z946 |
| Z94.8 | DX10 | Other transplanted organ and tissue status | SolidOrgan_Hema_Cell_Transplant | Z948 |
| Z94.82 | DX10 | Intestine transplant status | SolidOrgan_Hema_Cell_Transplant | Z9482 |
| Z94.83 | DX10 | Pancreas transplant status | SolidOrgan_Hema_Cell_Transplant | Z9483 |
| Z94.89 | DX10 | Other transplanted organ and tissue status | SolidOrgan_Hema_Cell_Transplant | Z9489 |
| Z94.9 | DX10 | Transplanted organ and tissue status, unspecified | SolidOrgan_Hema_Cell_Transplant | Z949 |
| Z98.85 | DX10 | Transplanted organ removal status | SolidOrgan_Hema_Cell_Transplant | Z9885 |
| 238.77 | DX10 | Post-transplant lymphoproliferative disorder [PTLD] | SolidOrgan_Hema_Cell_Transplant | 23877 |
| 996.81 | DX10 | Complications of transplanted kidney | SolidOrgan_Hema_Cell_Transplant | 99681 |
| 996.82 | DX10 | Complications of transplanted liver | SolidOrgan_Hema_Cell_Transplant | 99682 |
| 996.83 | DX10 | Complications of transplanted heart | SolidOrgan_Hema_Cell_Transplant | 99683 |

| | | | | |
|---|---|---|---|---|
| 996.84 | DX10 | Complications of transplanted lung | SolidOrgan_Hema_Cell_Transplant | 99684 |
| 996.87 | DX10 | Complications of transplanted organ, intestine | SolidOrgan_Hema_Cell_Transplant | 99687 |
| V42.0 | DX10 | Kidney replaced by transplant | SolidOrgan_Hema_Cell_Transplant | V420 |
| V42.1 | DX10 | Heart replaced by transplant | SolidOrgan_Hema_Cell_Transplant | V421 |
| V42.4 | DX10 | Bone replaced by transplant | SolidOrgan_Hema_Cell_Transplant | V424 |
| V42.6 | DX10 | Lung replaced by transplant | SolidOrgan_Hema_Cell_Transplant | V426 |
| V42.7 | DX10 | Liver replaced by transplant | SolidOrgan_Hema_Cell_Transplant | V427 |
| V42.83 | DX10 | Pancreas replaced by transplant | SolidOrgan_Hema_Cell_Transplant | V4283 |
| V42.84 | DX10 | Organ or tissue replaced by transplant, intestines | SolidOrgan_Hema_Cell_Transplant | V4284 |
| V42.89 | DX10 | Other organ or tissue replaced by transplant | SolidOrgan_Hema_Cell_Transplant | V4289 |
| V45.87 | DX10 | Transplanted organ removal status | SolidOrgan_Hema_Cell_Transplant | V4587 |
| A15 | DX10 | Respiratory tuberculosis | Tuberculosis | A15 |
| A15.0 | DX10 | Tuberculosis of lung | Tuberculosis | A150 |
| A15.4 | DX10 | Tuberculosis of intrathoracic lymph nodes | Tuberculosis | A154 |
| A15.5 | DX10 | Tuberculosis of larynx, trachea and bronchus | Tuberculosis | A155 |
| A15.6 | DX10 | Tuberculous pleurisy | Tuberculosis | A156 |
| A15.7 | DX10 | Primary respiratory tuberculosis | Tuberculosis | A157 |
| A15.8 | DX10 | Other respiratory tuberculosis | Tuberculosis | A158 |
| A15.9 | DX10 | Respiratory tuberculosis unspecified | Tuberculosis | A159 |
| A17 | DX10 | Tuberculosis of nervous system | Tuberculosis | A17 |
| A17.0 | DX10 | Tuberculous meningitis | Tuberculosis | A170 |
| A17.1 | DX10 | Meningeal tuberculoma | Tuberculosis | A171 |
| A17.8 | DX10 | Other tuberculosis of nervous system | Tuberculosis | A178 |
| A17.81 | DX10 | Tuberculoma of brain and spinal cord | Tuberculosis | A1781 |
| A17.82 | DX10 | Tuberculous meningoencephalitis | Tuberculosis | A1782 |
| A17.83 | DX10 | Tuberculous neuritis | Tuberculosis | A1783 |
| A17.89 | DX10 | Other tuberculosis of nervous system | Tuberculosis | A1789 |
| A17.9 | DX10 | Tuberculosis of nervous system, unspecified | Tuberculosis | A179 |
| A18 | DX10 | Tuberculosis of other organs | Tuberculosis | A18 |
| A18.0 | DX10 | Tuberculosis of bones and joints | Tuberculosis | A180 |
| A18.01 | DX10 | Tuberculosis of spine | Tuberculosis | A1801 |
| A18.02 | DX10 | Tuberculous arthritis of other joints | Tuberculosis | A1802 |
| A18.03 | DX10 | Tuberculosis of other bones | Tuberculosis | A1803 |
| A18.09 | DX10 | Other musculoskeletal tuberculosis | Tuberculosis | A1809 |
| A18.1 | DX10 | Tuberculosis of genitourinary system | Tuberculosis | A181 |
| A18.10 | DX10 | Tuberculosis of genitourinary system, unspecified | Tuberculosis | A1810 |
| A18.11 | DX10 | Tuberculosis of kidney and ureter | Tuberculosis | A1811 |
| A18.12 | DX10 | Tuberculosis of bladder | Tuberculosis | A1812 |
| A18.13 | DX10 | Tuberculosis of other urinary organs | Tuberculosis | A1813 |
| A18.14 | DX10 | Tuberculosis of prostate | Tuberculosis | A1814 |
| A18.15 | DX10 | Tuberculosis of other male genital organs | Tuberculosis | A1815 |
| A18.16 | DX10 | Tuberculosis of cervix | Tuberculosis | A1816 |
| A18.17 | DX10 | Tuberculous female pelvic inflammatory disease | Tuberculosis | A1817 |
| A18.18 | DX10 | Tuberculosis of other female genital organs | Tuberculosis | A1818 |
| A18.2 | DX10 | Tuberculous peripheral lymphadenopathy | Tuberculosis | A182 |
| A18.3 | DX10 | Tuberculosis of intestines, peritoneum and mesenteric glands | Tuberculosis | A183 |
| A18.31 | DX10 | Tuberculous peritonitis | Tuberculosis | A1831 |

| | | | | |
|---|---|---|---|---|
| A18.32 | DX10 | Tuberculous enteritis | Tuberculosis | A1832 |
| A18.39 | DX10 | Retroperitoneal tuberculosis | Tuberculosis | A1839 |
| A18.4 | DX10 | Tuberculosis of skin and subcutaneous tissue | Tuberculosis | A184 |
| A18.5 | DX10 | Tuberculosis of eye | Tuberculosis | A185 |
| A18.50 | DX10 | Tuberculosis of eye, unspecified | Tuberculosis | A1850 |
| A18.51 | DX10 | Tuberculous episcleritis | Tuberculosis | A1851 |
| A18.52 | DX10 | Tuberculous keratitis | Tuberculosis | A1852 |
| A18.53 | DX10 | Tuberculous chorioretinitis | Tuberculosis | A1853 |
| A18.54 | DX10 | Tuberculous iridocyclitis | Tuberculosis | A1854 |
| A18.59 | DX10 | Other tuberculosis of eye | Tuberculosis | A1859 |
| A18.6 | DX10 | Tuberculosis of (inner) (middle) ear | Tuberculosis | A186 |
| A18.7 | DX10 | Tuberculosis of adrenal glands | Tuberculosis | A187 |
| A18.8 | DX10 | Tuberculosis of other specified organs | Tuberculosis | A188 |
| A18.81 | DX10 | Tuberculosis of thyroid gland | Tuberculosis | A1881 |
| A18.82 | DX10 | Tuberculosis of other endocrine glands | Tuberculosis | A1882 |
| A18.83 | DX10 | Tuberculosis of digestive tract organs, not elsewhere classified | Tuberculosis | A1883 |
| A18.84 | DX10 | Tuberculosis of heart | Tuberculosis | A1884 |
| A18.85 | DX10 | Tuberculosis of spleen | Tuberculosis | A1885 |
| A18.89 | DX10 | Tuberculosis of other sites | Tuberculosis | A1889 |
| A19 | DX10 | Miliary tuberculosis | Tuberculosis | A19 |
| A19.0 | DX10 | Acute miliary tuberculosis of a single specified site | Tuberculosis | A190 |
| A19.1 | DX10 | Acute miliary tuberculosis of multiple sites | Tuberculosis | A191 |
| A19.2 | DX10 | Acute miliary tuberculosis, unspecified | Tuberculosis | A192 |
| A19.8 | DX10 | Other miliary tuberculosis | Tuberculosis | A198 |
| A19.9 | DX10 | Miliary tuberculosis, unspecified | Tuberculosis | A199 |
| J65 | DX10 | Pneumoconiosis associated with tuberculosis | Tuberculosis | J65 |
| O98.01 | DX10 | Tuberculosis complicating pregnancy | Tuberculosis | O9801 |
| O98.02 | DX10 | Tuberculosis complicating childbirth | Tuberculosis | O9802 |
| O98.03 | DX10 | Tuberculosis complicating the puerperium | Tuberculosis | O9803 |
| P37.0 | DX10 | Congenital tuberculosis | Tuberculosis | P370 |
| A16 | DX10 | TB diagnosis without microbiological confirmation | Tuberculosis | A16 |
| A16.0 | DX10 | TB diagnosis without microbiological confirmation | Tuberculosis | A160 |
| A16.1 | DX10 | TB diagnosis without microbiological confirmation | Tuberculosis | A161 |
| A16.2 | DX10 | TB diagnosis without microbiological confirmation | Tuberculosis | A162 |
| A16.3 | DX10 | TB diagnosis without microbiological confirmation | Tuberculosis | A163 |
| A16.4 | DX10 | TB diagnosis without microbiological confirmation | Tuberculosis | A164 |
| A16.5 | DX10 | TB diagnosis without microbiological confirmation | Tuberculosis | A165 |
| A16.7 | DX10 | TB diagnosis without microbiological confirmation | Tuberculosis | A167 |
| A16.8 | DX10 | TB diagnosis without microbiological confirmation | Tuberculosis | A168 |
| A16.9 | DX10 | TB diagnosis without microbiological confirmation | Tuberculosis | A169 |
