## Supplemental Material D for "How age and underlying conditions affect incidence of hospitalization and death after documented COVID-19 diagnosis among individuals aged ≥20 years, a retrospective cohort study using PCORnet, April 2022 – March 2025"

| Code | Codetype | Medication type | Description |
|---|---|---|---|
| 8638 | PR00 | Corticosteriods | prednisolone |
| 6902 | PR00 | Corticosteriods | methylprednisolone |
| 1366551 | PR00 | Corticosteriods | UCERIS |
| 153171 | PR00 | Corticosteriods | ENTOCORT |
| 2273102 | PR00 | Corticosteriods | ORTIKOS |
| 19831 | PR00 | Corticosteriods | BUDESONIDE |
| 1367172 | PR00 | Corticosteriods | BUDESONIDE (11.BETA.,16.ALPHA.(S)) |
| 1244213 | PR00 | Corticosteriods | 24 HR BUDESONIDE 3 MG EXTENDED RELEASE ENTERIC COATED CAPSULE [ENTOCORT] |
| 1244215 | PR00 | Corticosteriods | BUDESONIDE 3 MG DELAYED RELEASE ORAL CAPSULE [ENTOCORT] |
| 1366556 | PR00 | Corticosteriods | 24 HR BUDESONIDE 9 MG EXTENDED RELEASE ORAL TABLET [UCERIS] |
| 1366558 | PR00 | Corticosteriods | BUDESONIDE 9 MG EXTENDED RELEASE ORAL TABLET [UCERIS] |
| 152595 | PR00 | Corticosteriods | 24 HR BUDESONIDE 3 MG EXTENDED RELEASE CAPSULE [ENTOCORT EC] |
| 2273107 | PR00 | Corticosteriods | 24 HR BUDESONIDE 6 MG EXTENDED RELEASE ORAL CAPSULE [ORTIKOS] |
| 2273108 | PR00 | Corticosteriods | BUDESONIDE 6 MG EXTENDED RELEASE ORAL CAPSULE [ORTIKOS] |
| 2273110 | PR00 | Corticosteriods | 24 HR BUDESONIDE 9 MG EXTENDED RELEASE ORAL CAPSULE [ORTIKOS] |
| 2273111 | PR00 | Corticosteriods | BUDESONIDE 9 MG EXTENDED RELEASE ORAL CAPSULE [ORTIKOS] |
| 844427 | PR00 | Corticosteriods | BUDESONIDE 3 MG EXTENDED RELEASE CAPSULE [ENTOCORT EC] |
| 991401 | PR00 | Corticosteriods | 24 HR BUDESONIDE 3 MG EXTENDED RELEASE CAPSULE [ENTOCORT] |
| 991402 | PR00 | Corticosteriods | BUDESONIDE 3 MG EXTENDED RELEASE CAPSULE [ENTOCORT] |
| 1366552 | PR00 | Corticosteriods | BUDESONIDE 9 MG [UCERIS] |
| 2273103 | PR00 | Corticosteriods | BUDESONIDE 6 MG [ORTIKOS] |
| 2273109 | PR00 | Corticosteriods | BUDESONIDE 9 MG [ORTIKOS] |
| 564993 | PR00 | Corticosteriods | BUDESONIDE 3 MG [ENTOCORT EC] |
| 991399 | PR00 | Corticosteriods | BUDESONIDE 3 MG [ENTOCORT] |
| 1244212 | PR00 | Corticosteriods | BUDESONIDE EXTENDED RELEASE ENTERIC COATED CAPSULE [ENTOCORT] |
| 1366553 | PR00 | Corticosteriods | BUDESONIDE EXTENDED RELEASE ORAL TABLET [UCERIS] |
| 1489772 | PR00 | Corticosteriods | BUDESONIDE DELAYED RELEASE ORAL CAPSULE [ENTOCORT] |
| 2273104 | PR00 | Corticosteriods | BUDESONIDE EXTENDED RELEASE ORAL CAPSULE [ORTIKOS] |
| 367142 | PR00 | Corticosteriods | BUDESONIDE 24 HOUR EXTENDED RELEASE CAPSULE [ENTOCORT EC] |
| 991400 | PR00 | Corticosteriods | BUDESONIDE EXTENDED RELEASE CAPSULE [ENTOCORT] |
| 1167031 | PR00 | Corticosteriods | ENTOCORT ORAL PRODUCT |
| 1167032 | PR00 | Corticosteriods | ENTOCORT PILL |
| 1366554 | PR00 | Corticosteriods | UCERIS ORAL PRODUCT |
| 1366555 | PR00 | Corticosteriods | UCERIS PILL |
| 2273105 | PR00 | Corticosteriods | ORTIKOS ORAL PRODUCT |
| 2273106 | PR00 | Corticosteriods | ORTIKOS PILL |
| 1006089 | PR00 | Corticosteriods | BUDESONIDE 3 MG ENTERIC COATED CAPSULE |
| 1244211 | PR00 | Corticosteriods | 24 HR BUDESONIDE 3 MG EXTENDED RELEASE ENTERIC COATED CAPSULE |
| 1244214 | PR00 | Corticosteriods | BUDESONIDE 3 MG DELAYED RELEASE ORAL CAPSULE |
| 1366550 | PR00 | Corticosteriods | 24 HR BUDESONIDE 9 MG EXTENDED RELEASE ORAL TABLET |
| 1366557 | PR00 | Corticosteriods | BUDESONIDE 9 MG EXTENDED RELEASE ORAL TABLET |
| 152594 | PR00 | Corticosteriods | 24 HR BUDESONIDE 3 MG EXTENDED RELEASE CAPSULE |
| 2048810 | PR00 | Corticosteriods | BUDESONIDE 3 MG ORAL CAPSULE |
| 2269305 | PR00 | Corticosteriods | 24 HR BUDESONIDE 6 MG EXTENDED RELEASE ORAL CAPSULE |
| 2269306 | PR00 | Corticosteriods | BUDESONIDE 6 MG EXTENDED RELEASE ORAL CAPSULE |
| 2269307 | PR00 | Corticosteriods | 24 HR BUDESONIDE 9 MG EXTENDED RELEASE ORAL CAPSULE |
| 2269308 | PR00 | Corticosteriods | BUDESONIDE 9 MG EXTENDED RELEASE ORAL CAPSULE |
| 249241 | PR00 | Corticosteriods | BUDESONIDE 3 MG ORAL TABLET |
| 387015 | PR00 | Corticosteriods | BUDESONIDE 3 MG ORAL CAPSULE |
| 484697 | PR00 | Corticosteriods | BUDESONIDE 3 MG EXTENDED RELEASE CAPSULE |
| 844426 | PR00 | Corticosteriods | BUDESONIDE 3 MG EXTENDED RELEASE CAPSULE |
| 1366548 | PR00 | Corticosteriods | BUDESONIDE 9 MG |
| 2269303 | PR00 | Corticosteriods | BUDESONIDE 6 MG |
| 332685 | PR00 | Corticosteriods | BUDESONIDE 3 MG |
| 1006088 | PR00 | Corticosteriods | BUDESONIDE ENTERIC COATED CAPSULE |
| 1244210 | PR00 | Corticosteriods | BUDESONIDE EXTENDED RELEASE ENTERIC COATED CAPSULE |
| 1366549 | PR00 | Corticosteriods | BUDESONIDE EXTENDED RELEASE ORAL TABLET |
| 1489770 | PR00 | Corticosteriods | BUDESONIDE DELAYED RELEASE ORAL CAPSULE |
| 2048809 | PR00 | Corticosteriods | BUDESONIDE ORAL CAPSULE |
| 2269304 | PR00 | Corticosteriods | BUDESONIDE EXTENDED RELEASE ORAL CAPSULE |
| 376186 | PR00 | Corticosteriods | BUDESONIDE ORAL TABLET |
| 379057 | PR00 | Corticosteriods | BUDESONIDE 24 HOUR EXTENDED RELEASE CAPSULE |
| 393140 | PR00 | Corticosteriods | BUDESONIDE ORAL CAPSULE |
| 484696 | PR00 | Corticosteriods | BUDESONIDE EXTENDED RELEASE CAPSULE |
| 1151338 | PR00 | Corticosteriods | BUDESONIDE ORAL PRODUCT |
| 1151339 | PR00 | Corticosteriods | BUDESONIDE PILL |
| 1100388 | PR00 | Corticosteriods | HYBRISIL |
| 203856 | PR00 | Corticosteriods | SOLU-MEDROL |
| 22584 | PR00 | Corticosteriods | DEPO-MEDROL |
| 1358512 | PR00 | Corticosteriods | METHYLPREDNISOLONE ACETATE 20 MG/ML INJECTABLE SUSPENSION [DEPO-MEDROL] |

| | | | |
|---|---|---|---|
| 1743722 | PR00 | Corticosteriods | METHYLPREDNISOLONE 500 MG INJECTION [SOLU-MEDROL] |
| 1743729 | PR00 | Corticosteriods | METHYLPREDNISOLONE 1000 MG INJECTION [SOLU-MEDROL] |
| 207137 | PR00 | Corticosteriods | METHYLPREDNISOLONE 8 MG ORAL TABLET [MEDROL] |
| 1165881 | PR00 | Corticosteriods | A-METHAPRED INJECTABLE PRODUCT |
| 1183797 | PR00 | Corticosteriods | MEDROL PILL |
| 105404 | PR00 | Corticosteriods | METHYLPREDNISOLONE 50 MG/ML INJECTABLE SOLUTION |
| 1357886 | PR00 | Corticosteriods | METHYLPREDNISOLONE 2000 MG INJECTION |
| 1743726 | PR00 | Corticosteriods | METHYLPREDNISOLONE 1000 MG INJECTION |
| 1743855 | PR00 | Corticosteriods | 1 ML METHYLPREDNISOLONE ACETATE 80 MG/ML INJECTION |
| 1012205 | PR00 | Corticosteriods | ASMALPRED PLUS |
| 1191678 | PR00 | Corticosteriods | PREDNISTAB |
| 1493166 | PR00 | Corticosteriods | SOLU-DELTA-CORTEF |
| 152241 | PR00 | Corticosteriods | PREDNESOL |
| 202702 | PR00 | Corticosteriods | MEDROL |
| 202760 | PR00 | Corticosteriods | HYDELTRASOL |
| 203857 | PR00 | Corticosteriods | A-METHAPRED |
| 216258 | PR00 | Corticosteriods | COTOLONE |
| 216464 | PR00 | Corticosteriods | DEPO-PREDATE |
| 216840 | PR00 | Corticosteriods | ECONOPRED |
| 216841 | PR00 | Corticosteriods | ECONOPRED PLUS |
| 217592 | PR00 | Corticosteriods | HYDELTRA-T.B.A. |
| 217898 | PR00 | Corticosteriods | KEY-PRED |
| 217899 | PR00 | Corticosteriods | KEY-PRED SP |
| 218285 | PR00 | Corticosteriods | MEDICORT |
| 219136 | PR00 | Corticosteriods | PEDIAPRED |
| 219282 | PR00 | Corticosteriods | POLY PRED |
| 219341 | PR00 | Corticosteriods | PRED-JECT-50 |
| 219342 | PR00 | Corticosteriods | PREDACORT 50 |
| 219344 | PR00 | Corticosteriods | PREDAJECT-50 |
| 219345 | PR00 | Corticosteriods | PREDALONE 50 |
| 219346 | PR00 | Corticosteriods | PREDATE-50 |
| 219347 | PR00 | Corticosteriods | PREDCOR |
| 219348 | PR00 | Corticosteriods | PREDICORT RP |
| 219349 | PR00 | Corticosteriods | PREDICORT-50 |
| 219356 | PR00 | Corticosteriods | PRELONE |
| 219403 | PR00 | Corticosteriods | PRI-CORTIN 50 |
| 285077 | PR00 | Corticosteriods | ORAPRED |
| 353627 | PR00 | Corticosteriods | CODELSOL |
| 353629 | PR00 | Corticosteriods | STINTISONE |
| 385325 | PR00 | Corticosteriods | CODELSON |
| 541702 | PR00 | Corticosteriods | PREDNORAL |
| 547219 | PR00 | Corticosteriods | PRICORTIN |
| 604201 | PR00 | Corticosteriods | BUBBLI-PRED |
| 792207 | PR00 | Corticosteriods | FLO-PRED |
| 795094 | PR00 | Corticosteriods | MILLIPRED |
| 824886 | PR00 | Corticosteriods | VERIPRED |
| 897060 | PR00 | Corticosteriods | TEMARIL-P |
| 1005831 | PR00 | Corticosteriods | {48 (PREDNISOLONE 5 MG ORAL TABLET [MILLIPRED]) } PACK [MILLIPRED DP 12 DAY] |
| 1235042 | PR00 | Corticosteriods | {21 (PREDNISOLONE 5 MG ORAL TABLET [MILLIPRED]) } PACK [MILLIPRED DP 6 DAY] |
| 834023 | PR00 | Corticosteriods | {21 (METHYLPREDNISOLONE 4 MG ORAL TABLET [MEDROL]) } PACK [MEDROL DOSEPAK] |
| 1005830 | PR00 | Corticosteriods | {48 (PREDNISOLONE 5 MG ORAL TABLET) } PACK |
| 1013114 | PR00 | Corticosteriods | {21 (PREDNISOLONE 5 MG ORAL TABLET) } PACK |
| 762675 | PR00 | Corticosteriods | {21 (METHYLPREDNISOLONE 4 MG ORAL TABLET) } PACK |
| 804173 | PR00 | Corticosteriods | {14 (METHYLPREDNISOLONE 16 MG ORAL TABLET) } PACK |
| 1358612 | PR00 | Corticosteriods | METHYLPREDNISOLONE ACETATE 40 MG/ML INJECTABLE SUSPENSION [DEPO-MEDROL] |
| 1358619 | PR00 | Corticosteriods | METHYLPREDNISOLONE ACETATE 80 MG/ML INJECTABLE SUSPENSION [DEPO-MEDROL] |
| 1743707 | PR00 | Corticosteriods | METHYLPREDNISOLONE 125 MG INJECTION [SOLU-MEDROL] |
| 1743781 | PR00 | Corticosteriods | 1 ML METHYLPREDNISOLONE ACETATE 40 MG/ML INJECTION [DEPO-MEDROL] |
| 1743856 | PR00 | Corticosteriods | 1 ML METHYLPREDNISOLONE ACETATE 80 MG/ML INJECTION [DEPO-MEDROL] |
| 207136 | PR00 | Corticosteriods | METHYLPREDNISOLONE 2 MG ORAL TABLET [MEDROL] |
| 207138 | PR00 | Corticosteriods | METHYLPREDNISOLONE 16 MG ORAL TABLET [MEDROL] |
| 207139 | PR00 | Corticosteriods | METHYLPREDNISOLONE 24 MG ORAL TABLET [MEDROL] |
| 207141 | PR00 | Corticosteriods | METHYLPREDNISOLONE 32 MG ORAL TABLET [MEDROL] |
| 207168 | PR00 | Corticosteriods | METHYLPREDNISOLONE 40 MG/ML INJECTABLE SUSPENSION [DEPO-MEDROL] |
| 207190 | PR00 | Corticosteriods | METHYLPREDNISOLONE 40 MG INJECTION [A-METHAPRED] |
| 207191 | PR00 | Corticosteriods | METHYLPREDNISOLONE 40 MG INJECTION [SOLU-MEDROL] |
| 207192 | PR00 | Corticosteriods | METHYLPREDNISOLONE 125 MG INJECTION [A-METHAPRED] |
| 207193 | PR00 | Corticosteriods | METHYLPREDNISOLONE 62.5 MG/ML INJECTABLE SOLUTION [SOLU-MEDROL] |
| 260330 | PR00 | Corticosteriods | METHYLPREDNISOLONE 4 MG ORAL TABLET [MEDROL] |
| 260409 | PR00 | Corticosteriods | PREDNISOLONE 1 MG/ML ORAL SOLUTION [PEDIAPRED] |
| 544779 | PR00 | Corticosteriods | METHYLPREDNISOLONE 125 MG/ML INJECTABLE SOLUTION [A-METHAPRED] |

| | | | |
|---|---|---|---|
| 647127 | PR00 | Corticosteriods | PREDNISOLONE 15 MG DISINTEGRATING ORAL TABLET [ORAPRED] |
| 668660 | PR00 | Corticosteriods | PREDNISOLONE 30 MG DISINTEGRATING ORAL TABLET [ORAPRED] |
| 808118 | PR00 | Corticosteriods | PREDNISOLONE 5 MG ORAL TABLET [MILLIPRED] |
| 1358611 | PR00 | Corticosteriods | METHYLPREDNISOLONE ACETATE 40 MG/ML [DEPO-MEDROL] |
| 567932 | PR00 | Corticosteriods | METHYLPREDNISOLONE 32 MG [MEDROL] |
| 567980 | PR00 | Corticosteriods | METHYLPREDNISOLONE 125 MG/ML [SOLU-MEDROL] |
| 567982 | PR00 | Corticosteriods | METHYLPREDNISOLONE 40 MG/ML [SOLU-MEDROL] |
| 567984 | PR00 | Corticosteriods | METHYLPREDNISOLONE 62.5 MG/ML [SOLU-MEDROL] |
| 574125 | PR00 | Corticosteriods | METHYLPREDNISOLONE 4 MG [MEDROL] |
| 368772 | PR00 | Corticosteriods | METHYLPREDNISOLONE ORAL TABLET [MEDROL] |
| 1168054 | PR00 | Corticosteriods | BUBBLI-PRED ORAL LIQUID PRODUCT |
| 1168055 | PR00 | Corticosteriods | BUBBLI-PRED ORAL PRODUCT |
| 1169771 | PR00 | Corticosteriods | FLO-PRED ORAL LIQUID PRODUCT |
| 1169772 | PR00 | Corticosteriods | FLO-PRED ORAL PRODUCT |
| 1170721 | PR00 | Corticosteriods | KEY-PRED INJECTABLE PRODUCT |
| 1170722 | PR00 | Corticosteriods | KEY-PRED SP INJECTABLE PRODUCT |
| 1171632 | PR00 | Corticosteriods | COTOLONE INJECTABLE PRODUCT |
| 1171633 | PR00 | Corticosteriods | COTOLONE ORAL PRODUCT |
| 1171634 | PR00 | Corticosteriods | COTOLONE PILL |
| 1172143 | PR00 | Corticosteriods | ASMALPRED PLUS ORAL LIQUID PRODUCT |
| 1172144 | PR00 | Corticosteriods | ASMALPRED PLUS ORAL PRODUCT |
| 1173110 | PR00 | Corticosteriods | DEPO-MEDROL INJECTABLE PRODUCT |
| 1173111 | PR00 | Corticosteriods | DEPO-PREDATE INJECTABLE PRODUCT |
| 1173399 | PR00 | Corticosteriods | HYDELTRA-T.B.A. INJECTABLE PRODUCT |
| 1173400 | PR00 | Corticosteriods | HYDELTRASOL INJECTABLE PRODUCT |
| 1175156 | PR00 | Corticosteriods | CODELSOL INJECTABLE PRODUCT |
| 1175157 | PR00 | Corticosteriods | CODELSON INJECTABLE PRODUCT |
| 1177661 | PR00 | Corticosteriods | PRI-CORTIN 50 INJECTABLE PRODUCT |
| 1178234 | PR00 | Corticosteriods | PRICORTIN INJECTABLE PRODUCT |
| 1179016 | PR00 | Corticosteriods | STINTISONE ORAL PRODUCT |
| 1180193 | PR00 | Corticosteriods | STINTISONE PILL |
| 1181408 | PR00 | Corticosteriods | TEMARIL-P ORAL PRODUCT |
| 1181409 | PR00 | Corticosteriods | TEMARIL-P PILL |
| 1183281 | PR00 | Corticosteriods | MEDICORT INJECTABLE PRODUCT |
| 1183437 | PR00 | Corticosteriods | ORAPRED ORAL LIQUID PRODUCT |
| 1183438 | PR00 | Corticosteriods | ORAPRED ORAL PRODUCT |
| 1183439 | PR00 | Corticosteriods | ORAPRED PILL |
| 1183796 | PR00 | Corticosteriods | MEDROL ORAL PRODUCT |
| 1184005 | PR00 | Corticosteriods | PEDIAPRED ORAL LIQUID PRODUCT |
| 1184006 | PR00 | Corticosteriods | PEDIAPRED ORAL PRODUCT |
| 1184840 | PR00 | Corticosteriods | SOLU-MEDROL INJECTABLE PRODUCT |
| 1184933 | PR00 | Corticosteriods | MILLIPRED ORAL LIQUID PRODUCT |
| 1184934 | PR00 | Corticosteriods | MILLIPRED ORAL PRODUCT |
| 1184935 | PR00 | Corticosteriods | MILLIPRED PILL |
| 1185242 | PR00 | Corticosteriods | PRED-JECT-50 INJECTABLE PRODUCT |
| 1185244 | PR00 | Corticosteriods | PREDACORT 50 INJECTABLE PRODUCT |
| 1185246 | PR00 | Corticosteriods | PREDAJECT-50 INJECTABLE PRODUCT |
| 1185247 | PR00 | Corticosteriods | PREDALONE 50 INJECTABLE PRODUCT |
| 1185248 | PR00 | Corticosteriods | PREDATE-50 INJECTABLE PRODUCT |
| 1185249 | PR00 | Corticosteriods | PREDCOR INJECTABLE PRODUCT |
| 1185252 | PR00 | Corticosteriods | PREDICORT RP INJECTABLE PRODUCT |
| 1185253 | PR00 | Corticosteriods | PREDICORT-50 INJECTABLE PRODUCT |
| 1185254 | PR00 | Corticosteriods | PREDNESOL ORAL PRODUCT |
| 1185255 | PR00 | Corticosteriods | PREDNESOL PILL |
| 1185263 | PR00 | Corticosteriods | PREDNORAL ORAL PRODUCT |
| 1185264 | PR00 | Corticosteriods | PREDNORAL PILL |
| 1185831 | PR00 | Corticosteriods | PRELONE ORAL LIQUID PRODUCT |
| 1185832 | PR00 | Corticosteriods | PRELONE ORAL PRODUCT |
| 1187894 | PR00 | Corticosteriods | VERIPRED ORAL LIQUID PRODUCT |
| 1187895 | PR00 | Corticosteriods | VERIPRED ORAL PRODUCT |
| 1191681 | PR00 | Corticosteriods | PREDNISTAB ORAL PRODUCT |
| 1191682 | PR00 | Corticosteriods | PREDNISTAB PILL |
| 1296584 | PR00 | Corticosteriods | ORAPRED DISINTEGRATING ORAL PRODUCT |
| 1493169 | PR00 | Corticosteriods | SOLU-DELTA-CORTEF INJECTABLE PRODUCT |
| 105403 | PR00 | Corticosteriods | METHYLPREDNISOLONE 125 MG/ML INJECTABLE SOLUTION |
| 105411 | PR00 | Corticosteriods | PREDNISOLONE 2.5 MG ORAL TABLET |
| 1358510 | PR00 | Corticosteriods | METHYLPREDNISOLONE ACETATE 20 MG/ML INJECTABLE SUSPENSION |
| 1358610 | PR00 | Corticosteriods | METHYLPREDNISOLONE ACETATE 40 MG/ML INJECTABLE SUSPENSION |
| 1358617 | PR00 | Corticosteriods | METHYLPREDNISOLONE ACETATE 80 MG/ML INJECTABLE SUSPENSION |
| 1743704 | PR00 | Corticosteriods | METHYLPREDNISOLONE 125 MG INJECTION |
| 1743720 | PR00 | Corticosteriods | METHYLPREDNISOLONE 500 MG INJECTION |

| | | | |
|---|---|---|---|
| 1743779 | PR00 | Corticosteriods | 1 ML METHYLPREDNISOLONE ACETATE 40 MG/ML INJECTION |
| 197970 | PR00 | Corticosteriods | METHYLPREDNISOLONE 24 MG ORAL TABLET |
| 197971 | PR00 | Corticosteriods | METHYLPREDNISOLONE 32 MG ORAL TABLET |
| 197973 | PR00 | Corticosteriods | METHYLPREDNISOLONE 8 MG ORAL TABLET |
| 198142 | PR00 | Corticosteriods | PREDNISOLONE 5 MG ORAL TABLET |
| 199343 | PR00 | Corticosteriods | PREDNISOLONE 1 MG ORAL TABLET |
| 199771 | PR00 | Corticosteriods | METHYLPREDNISOLONE 100 MG ORAL TABLET |
| 208988 | PR00 | Corticosteriods | PREDNISOLONE ACETATE 5 MG ORAL TABLET |
| 249066 | PR00 | Corticosteriods | PREDNISOLONE 5 MG/ML ORAL SOLUTION |
| 259966 | PR00 | Corticosteriods | METHYLPREDNISOLONE 4 MG ORAL TABLET |
| 283077 | PR00 | Corticosteriods | PREDNISOLONE 3 MG/ML ORAL SOLUTION |
| 311659 | PR00 | Corticosteriods | METHYLPREDNISOLONE 40 MG INJECTION |
| 314099 | PR00 | Corticosteriods | METHYLPREDNISOLONE 62.5 MG/ML INJECTABLE SOLUTION |
| 328161 | PR00 | Corticosteriods | METHYLPREDNISOLONE 16 MG ORAL TABLET |
| 346535 | PR00 | Corticosteriods | METHYLPREDNISOLONE 40 MG/ML INJECTABLE SUSPENSION |
| 348428 | PR00 | Corticosteriods | PREDNISOLONE 50 MG ORAL TABLET |
| 429198 | PR00 | Corticosteriods | PREDNISOLONE 2 MG ORAL TABLET |
| 436279 | PR00 | Corticosteriods | PREDNISOLONE 10 MG ORAL TABLET |
| 643123 | PR00 | Corticosteriods | PREDNISOLONE 10 MG DISINTEGRATING ORAL TABLET |
| 643125 | PR00 | Corticosteriods | PREDNISOLONE 15 MG DISINTEGRATING ORAL TABLET |
| 643127 | PR00 | Corticosteriods | PREDNISOLONE 30 MG DISINTEGRATING ORAL TABLET |
| 702306 | PR00 | Corticosteriods | PREDNISOLONE 4 MG/ML ORAL SOLUTION |
| 793099 | PR00 | Corticosteriods | PREDNISOLONE 3 MG/ML ORAL SUSPENSION |
| 794979 | PR00 | Corticosteriods | PREDNISOLONE 2 MG/ML ORAL SOLUTION |
| 1743725 | PR00 | Corticosteriods | METHYLPREDNISOLONE 1000 MG |
| 376656 | PR00 | Corticosteriods | METHYLPREDNISOLONE INJECTABLE SOLUTION |
| 1163486 | PR00 | Corticosteriods | METHYLPREDNISOLONE INJECTABLE PRODUCT |
| 1163488 | PR00 | Corticosteriods | METHYLPREDNISOLONE ORAL PRODUCT |
| 1165759 | PR00 | Corticosteriods | PREDNISOLONE ORAL PRODUCT |
| 1295508 | PR00 | Corticosteriods | PREDNISOLONE DISINTEGRATING ORAL PRODUCT |
| 1303126 | PR00 | Corticosteriods | RAYOS |
| 151587 | PR00 | Corticosteriods | DECORTISYL |
| 219350 | PR00 | Corticosteriods | PREDNICEN-M |
| 220028 | PR00 | Corticosteriods | STERAPRED |
| 220029 | PR00 | Corticosteriods | STERAPRED DS |
| 224927 | PR00 | Corticosteriods | LIQUID PRED |
| 224928 | PR00 | Corticosteriods | METICORTEN |
| 224929 | PR00 | Corticosteriods | ORASONE |
| 227730 | PR00 | Corticosteriods | DELTASONE |
| 352903 | PR00 | Corticosteriods | PREDNICOT |
| 380826 | PR00 | Corticosteriods | ECONOSONE |
| 541812 | PR00 | Corticosteriods | PREDONE |
| 582600 | PR00 | Corticosteriods | PREDNISONE INTENSOL |
| 795855 | PR00 | Corticosteriods | {12 (PREDNISONE 5 MG ORAL TABLET [STERAPRED]) } PACK [STERAPRED UNI-PAK 12 DAY] |
| 795857 | PR00 | Corticosteriods | {6 (PREDNISONE 5 MG ORAL TABLET [STERAPRED]) } PACK [STERAPRED UNI-PAK 6 DAY] |
| 795861 | PR00 | Corticosteriods | {6 (PREDNISONE 10 MG ORAL TABLET [STERAPRED DS]) } PACK [STERAPRED DS UNI-PAK 6 DAY] |
| 845491 | PR00 | Corticosteriods | {48 (PREDNISONE 5 MG ORAL TABLET [STERAPRED]) } PACK [STERAPRED 5 MG 12 DAY UNI-PAK] |
| 845492 | PR00 | Corticosteriods | {21 (PREDNISONE 5 MG ORAL TABLET [STERAPRED]) } PACK [STERAPRED 5 MG UNI-PAK] |
| 845493 | PR00 | Corticosteriods | {48 (PREDNISONE 10 MG ORAL TABLET [STERAPRED DS]) } PACK [STERAPRED DS 12 DAY UNI-PAK] |
| 845494 | PR00 | Corticosteriods | {21 (PREDNISONE 10 MG ORAL TABLET [STERAPRED DS]) } PACK [STERAPRED DS UNI-PAK] |
| 763179 | PR00 | Corticosteriods | {48 (PREDNISONE 5 MG ORAL TABLET) } PACK |
| 763181 | PR00 | Corticosteriods | {21 (PREDNISONE 5 MG ORAL TABLET) } PACK |
| 763183 | PR00 | Corticosteriods | {48 (PREDNISONE 10 MG ORAL TABLET) } PACK |
| 763185 | PR00 | Corticosteriods | {21 (PREDNISONE 10 MG ORAL TABLET) } PACK |
| 795854 | PR00 | Corticosteriods | {12 (PREDNISONE 5 MG ORAL TABLET) } PACK |
| 795856 | PR00 | Corticosteriods | {6 (PREDNISONE 5 MG ORAL TABLET) } PACK |
| 795860 | PR00 | Corticosteriods | {6 (PREDNISONE 10 MG ORAL TABLET) } PACK |
| 834395 | PR00 | Corticosteriods | {25 (PREDNISONE 10 MG ORAL TABLET) } PACK |
| 29523 | PR00 | Corticosteriods | MEPREDNISONE |
| 8640 | PR00 | Corticosteriods | PREDNISONE |
| 1008942 | PR00 | Corticosteriods | CHLORAMPHENICOL / PREDNISONE |
| 815961 | PR00 | Corticosteriods | PIROXICAM / PREDNISONE |
| 1742508 | PR00 | Corticosteriods | MEPREDNISONE DISODIUM PHOSPHATE |
| 105412 | PR00 | Corticosteriods | PREDNISONE 5 MG ORAL TABLET [DECORTISYL] |
| 1303131 | PR00 | Corticosteriods | PREDNISONE 1 MG DELAYED RELEASE ORAL TABLET [RAYOS] |
| 1303134 | PR00 | Corticosteriods | PREDNISONE 2 MG DELAYED RELEASE ORAL TABLET [RAYOS] |
| 1303137 | PR00 | Corticosteriods | PREDNISONE 5 MG DELAYED RELEASE ORAL TABLET [RAYOS] |
| 201082 | PR00 | Corticosteriods | PREDNISONE 1 MG ORAL TABLET [ECONOSONE] |

| | | | |
|---|---|---|---|
| 201083 | PR00 | Corticosteriods | PREDNISONE 5 MG ORAL TABLET [ECONOSONE] |
| 206199 | PR00 | Corticosteriods | PREDNISONE 1 MG/ML ORAL SOLUTION [LIQUID PRED] |
| 206754 | PR00 | Corticosteriods | PREDNISONE 1 MG ORAL TABLET [METICORTEN] |
| 206755 | PR00 | Corticosteriods | PREDNISONE 1 MG ORAL TABLET [ORASONE] |
| 206837 | PR00 | Corticosteriods | PREDNISONE 2.5 MG ORAL TABLET [DELTASONE] |
| 206954 | PR00 | Corticosteriods | PREDNISONE 5 MG ORAL TABLET [DELTASONE] |
| 206955 | PR00 | Corticosteriods | PREDNISONE 5 MG ORAL TABLET [ORASONE] |
| 206956 | PR00 | Corticosteriods | PREDNISONE 5 MG ORAL TABLET [PREDNICEN-M] |
| 206957 | PR00 | Corticosteriods | PREDNISONE 5 MG ORAL TABLET [STERAPRED] |
| 206988 | PR00 | Corticosteriods | PREDNISONE 10 MG ORAL TABLET [DELTASONE] |
| 206989 | PR00 | Corticosteriods | PREDNISONE 10 MG ORAL TABLET [ORASONE] |
| 206990 | PR00 | Corticosteriods | PREDNISONE 10 MG ORAL TABLET [STERAPRED DS] |
| 206997 | PR00 | Corticosteriods | PREDNISONE 20 MG ORAL TABLET [DELTASONE] |
| 206998 | PR00 | Corticosteriods | PREDNISONE 20 MG ORAL TABLET [ORASONE] |
| 207048 | PR00 | Corticosteriods | PREDNISONE 50 MG ORAL TABLET [DELTASONE] |
| 207049 | PR00 | Corticosteriods | PREDNISONE 50 MG ORAL TABLET [ORASONE] |
| 351399 | PR00 | Corticosteriods | PREDNISONE 5 MG ORAL TABLET [PREDNICOT] |
| 351400 | PR00 | Corticosteriods | PREDNISONE 10 MG ORAL TABLET [PREDNICOT] |
| 351401 | PR00 | Corticosteriods | PREDNISONE 20 MG ORAL TABLET [PREDNICOT] |
| 539948 | PR00 | Corticosteriods | PREDNISONE 1 MG/ML SYRUP [LIQUID PRED] |
| 541815 | PR00 | Corticosteriods | PREDNISONE 10 MG ORAL TABLET [PREDONE] |
| 541872 | PR00 | Corticosteriods | PREDNISONE 20 MG ORAL TABLET [PREDONE] |
| 541927 | PR00 | Corticosteriods | PREDNISONE 5 MG ORAL TABLET [LEVOXYL] |
| 582603 | PR00 | Corticosteriods | PREDNISONE 5 MG/ML ORAL SOLUTION [PREDNISONE INTENSOL] |
| 1303127 | PR00 | Corticosteriods | PREDNISONE 1 MG [RAYOS] |
| 1303133 | PR00 | Corticosteriods | PREDNISONE 2 MG [RAYOS] |
| 1303136 | PR00 | Corticosteriods | PREDNISONE 5 MG [RAYOS] |
| 539946 | PR00 | Corticosteriods | PREDNISONE 1 MG/ML [LIQUID PRED] |
| 541813 | PR00 | Corticosteriods | PREDNISONE 10 MG [PREDONE] |
| 541871 | PR00 | Corticosteriods | PREDNISONE 20 MG [PREDONE] |
| 541925 | PR00 | Corticosteriods | PREDNISONE 5 MG [LEVOXYL] |
| 564054 | PR00 | Corticosteriods | PREDNISONE 5 MG [DECORTISYL] |
| 565678 | PR00 | Corticosteriods | PREDNISONE 1 MG [ECONOSONE] |
| 565679 | PR00 | Corticosteriods | PREDNISONE 5 MG [ECONOSONE] |
| 567565 | PR00 | Corticosteriods | PREDNISONE 1 MG [METICORTEN] |
| 567566 | PR00 | Corticosteriods | PREDNISONE 1 MG [ORASONE] |
| 567645 | PR00 | Corticosteriods | PREDNISONE 2.5 MG [DELTASONE] |
| 567755 | PR00 | Corticosteriods | PREDNISONE 5 MG [DELTASONE] |
| 567756 | PR00 | Corticosteriods | PREDNISONE 5 MG [ORASONE] |
| 567757 | PR00 | Corticosteriods | PREDNISONE 5 MG [PREDNICEN-M] |
| 567758 | PR00 | Corticosteriods | PREDNISONE 5 MG [STERAPRED] |
| 567784 | PR00 | Corticosteriods | PREDNISONE 10 MG [DELTASONE] |
| 567785 | PR00 | Corticosteriods | PREDNISONE 10 MG [ORASONE] |
| 567786 | PR00 | Corticosteriods | PREDNISONE 10 MG [STERAPRED DS] |
| 567793 | PR00 | Corticosteriods | PREDNISONE 20 MG [DELTASONE] |
| 567794 | PR00 | Corticosteriods | PREDNISONE 20 MG [ORASONE] |
| 567841 | PR00 | Corticosteriods | PREDNISONE 50 MG [DELTASONE] |
| 567842 | PR00 | Corticosteriods | PREDNISONE 50 MG [ORASONE] |
| 575330 | PR00 | Corticosteriods | PREDNISONE 5 MG [PREDNICOT] |
| 575331 | PR00 | Corticosteriods | PREDNISONE 10 MG [PREDNICOT] |
| 575332 | PR00 | Corticosteriods | PREDNISONE 20 MG [PREDNICOT] |
| 582601 | PR00 | Corticosteriods | PREDNISONE 5 MG/ML [PREDNISONE INTENSOL] |
| 1303128 | PR00 | Corticosteriods | PREDNISONE DELAYED RELEASE ORAL TABLET [RAYOS] |
| 364008 | PR00 | Corticosteriods | PREDNISONE ORAL SOLUTION [LIQUID PRED] |
| 368654 | PR00 | Corticosteriods | PREDNISONE ORAL TABLET [PREDNICOT] |
| 368693 | PR00 | Corticosteriods | PREDNISONE ORAL TABLET [LEVOXYL] |
| 368790 | PR00 | Corticosteriods | PREDNISONE ORAL TABLET [ORASONE] |
| 368806 | PR00 | Corticosteriods | PREDNISONE ORAL TABLET [STERAPRED DS] |
| 368812 | PR00 | Corticosteriods | PREDNISONE ORAL TABLET [STERAPRED] |
| 368813 | PR00 | Corticosteriods | PREDNISONE ORAL TABLET [PREDNICEN-M] |
| 368867 | PR00 | Corticosteriods | PREDNISONE ORAL TABLET [METICORTEN] |
| 380827 | PR00 | Corticosteriods | PREDNISONE ORAL TABLET [ECONOSONE] |
| 380828 | PR00 | Corticosteriods | PREDNISONE ORAL TABLET [DECORTISYL] |
| 539947 | PR00 | Corticosteriods | PREDNISONE SYRUP [LIQUID PRED] |
| 541814 | PR00 | Corticosteriods | PREDNISONE ORAL TABLET [PREDONE] |
| 582602 | PR00 | Corticosteriods | PREDNISONE ORAL SOLUTION [PREDNISONE INTENSOL] |
| 92354 | PR00 | Corticosteriods | PREDNISONE ORAL TABLET [DELTASONE] |
| 1171028 | PR00 | Corticosteriods | DECORTISYL ORAL PRODUCT |
| 1171029 | PR00 | Corticosteriods | DECORTISYL PILL |
| 1171797 | PR00 | Corticosteriods | ECONOSONE ORAL PRODUCT |
| 1171798 | PR00 | Corticosteriods | ECONOSONE PILL |

| | | | |
|---|---|---|---|
| 1172410 | PR00 | Corticosteriods | DELTASONE ORAL PRODUCT |
| 1172411 | PR00 | Corticosteriods | DELTASONE PILL |
| 1176945 | PR00 | Corticosteriods | METICORTEN ORAL PRODUCT |
| 1176946 | PR00 | Corticosteriods | METICORTEN PILL |
| 1178546 | PR00 | Corticosteriods | LIQUID PRED ORAL LIQUID PRODUCT |
| 1178547 | PR00 | Corticosteriods | LIQUID PRED ORAL PRODUCT |
| 1178982 | PR00 | Corticosteriods | STERAPRED DS ORAL PRODUCT |
| 1178983 | PR00 | Corticosteriods | STERAPRED DS PILL |
| 1178984 | PR00 | Corticosteriods | STERAPRED ORAL PRODUCT |
| 1178985 | PR00 | Corticosteriods | STERAPRED PILL |
| 1183442 | PR00 | Corticosteriods | ORASONE ORAL PRODUCT |
| 1183443 | PR00 | Corticosteriods | ORASONE PILL |
| 1185256 | PR00 | Corticosteriods | PREDNICEN-M ORAL PRODUCT |
| 1185257 | PR00 | Corticosteriods | PREDNICEN-M PILL |
| 1185258 | PR00 | Corticosteriods | PREDNICOT ORAL PRODUCT |
| 1185259 | PR00 | Corticosteriods | PREDNICOT PILL |
| 1185261 | PR00 | Corticosteriods | PREDNISONE INTENSOL ORAL LIQUID PRODUCT |
| 1185262 | PR00 | Corticosteriods | PREDNISONE INTENSOL ORAL PRODUCT |
| 1185265 | PR00 | Corticosteriods | PREDONE ORAL PRODUCT |
| 1185266 | PR00 | Corticosteriods | PREDONE PILL |
| 1303129 | PR00 | Corticosteriods | RAYOS ORAL PRODUCT |
| 1303130 | PR00 | Corticosteriods | RAYOS PILL |
| 1303125 | PR00 | Corticosteriods | PREDNISONE 1 MG DELAYED RELEASE ORAL TABLET |
| 1303132 | PR00 | Corticosteriods | PREDNISONE 2 MG DELAYED RELEASE ORAL TABLET |
| 1303135 | PR00 | Corticosteriods | PREDNISONE 5 MG DELAYED RELEASE ORAL TABLET |
| 198144 | PR00 | Corticosteriods | PREDNISONE 1 MG ORAL TABLET |
| 198145 | PR00 | Corticosteriods | PREDNISONE 10 MG ORAL TABLET |
| 198146 | PR00 | Corticosteriods | PREDNISONE 2.5 MG ORAL TABLET |
| 198148 | PR00 | Corticosteriods | PREDNISONE 50 MG ORAL TABLET |
| 205301 | PR00 | Corticosteriods | PREDNISONE 5 MG/ML ORAL SOLUTION |
| 248682 | PR00 | Corticosteriods | MEPREDNISONE 8 MG ORAL TABLET |
| 248702 | PR00 | Corticosteriods | MEPREDNISONE 40 MG ORAL TABLET |
| 248725 | PR00 | Corticosteriods | MEPREDNISONE 4 MG ORAL TABLET |
| 248794 | PR00 | Corticosteriods | MEPREDNISONE 4 MG/ML ORAL SOLUTION |
| 250996 | PR00 | Corticosteriods | PIROXICAM 10 MG / PREDNISONE 2 MG DELAYED RELEASE ORAL TABLET |
| 251947 | PR00 | Corticosteriods | PIROXICAM 10 MG / PREDNISONE 2 MG ORAL TABLET |
| 312615 | PR00 | Corticosteriods | PREDNISONE 20 MG ORAL TABLET |
| 312616 | PR00 | Corticosteriods | PREDNISONE 25 MG ORAL TABLET |
| 312617 | PR00 | Corticosteriods | PREDNISONE 5 MG ORAL TABLET |
| 312618 | PR00 | Corticosteriods | PREDNISONE 1 MG/ML SYRUP |
| 315187 | PR00 | Corticosteriods | PREDNISONE 1 MG/ML ORAL SOLUTION |
| 429331 | PR00 | Corticosteriods | PREDNISONE 8 MG ORAL TABLET |
| 429332 | PR00 | Corticosteriods | PREDNISONE 40 MG ORAL TABLET |
| 746380 | PR00 | Corticosteriods | PREDNISONE 10 MG ORAL CAPSULE |
| 746836 | PR00 | Corticosteriods | PREDNISONE 20 MG ORAL CAPSULE |
| 746837 | PR00 | Corticosteriods | PREDNISONE 5 MG ORAL CAPSULE |
| 316580 | PR00 | Corticosteriods | PREDNISONE 1 MG/ML |
| 316581 | PR00 | Corticosteriods | PREDNISONE 2.5 MG |
| 316582 | PR00 | Corticosteriods | PREDNISONE 20 MG |
| 316583 | PR00 | Corticosteriods | PREDNISONE 25 MG |
| 316584 | PR00 | Corticosteriods | PREDNISONE 5 MG |
| 316585 | PR00 | Corticosteriods | PREDNISONE 5 MG/ML |
| 316586 | PR00 | Corticosteriods | PREDNISONE 50 MG |
| 317475 | PR00 | Corticosteriods | PREDNISONE 1 MG |
| 317663 | PR00 | Corticosteriods | PREDNISONE 10 MG |
| 334021 | PR00 | Corticosteriods | MEPREDNISONE 4 MG/ML |
| 334022 | PR00 | Corticosteriods | MEPREDNISONE 4 MG |
| 334023 | PR00 | Corticosteriods | MEPREDNISONE 40 MG |
| 334024 | PR00 | Corticosteriods | MEPREDNISONE 8 MG |
| 335473 | PR00 | Corticosteriods | PREDNISONE 2 MG |
| 438434 | PR00 | Corticosteriods | PREDNISONE 0.005 MG/MG |
| 451141 | PR00 | Corticosteriods | PREDNISONE 100 MG |
| 451143 | PR00 | Corticosteriods | PREDNISONE 30 MG |
| 451144 | PR00 | Corticosteriods | PREDNISONE 40 MG |
| 451147 | PR00 | Corticosteriods | PREDNISONE 8 MG |
| 1303124 | PR00 | Corticosteriods | PREDNISONE DELAYED RELEASE ORAL TABLET |
| 372772 | PR00 | Corticosteriods | MEPREDNISONE ORAL SOLUTION |
| 372773 | PR00 | Corticosteriods | MEPREDNISONE ORAL TABLET |
| 373495 | PR00 | Corticosteriods | PIROXICAM / PREDNISONE DELAYED RELEASE ORAL TABLET |
| 373583 | PR00 | Corticosteriods | PREDNISONE ORAL SOLUTION |
| 373584 | PR00 | Corticosteriods | PREDNISONE SYRUP |

| | | | |
|---|---|---|---|
| 373585 | PR00 | Corticosteriods | PREDNISONE ORAL TABLET |
| 376283 | PR00 | Corticosteriods | PIROXICAM / PREDNISONE ORAL TABLET |
| 746379 | PR00 | Corticosteriods | PREDNISONE ORAL CAPSULE |
| 1159297 | PR00 | Corticosteriods | PIROXICAM / PREDNISONE ORAL PRODUCT |
| 1159298 | PR00 | Corticosteriods | PIROXICAM / PREDNISONE PILL |
| 1161704 | PR00 | Corticosteriods | PREDNISONE ORAL LIQUID PRODUCT |
| 1161705 | PR00 | Corticosteriods | PREDNISONE ORAL PRODUCT |
| 1161706 | PR00 | Corticosteriods | PREDNISONE PILL |
| 1165700 | PR00 | Corticosteriods | MEPREDNISONE ORAL LIQUID PRODUCT |
| 1165701 | PR00 | Corticosteriods | MEPREDNISONE ORAL PRODUCT |
| 1165702 | PR00 | Corticosteriods | MEPREDNISONE PILL |
| 616011 | PR00 | Immunosuppressants | ORENCIA |
| 607377 | PR00 | Immunosuppressants | ABATACEPT |
| 614391 | PR00 | Immunosuppressants | ABATACEPT |
| 1145932 | PR00 | Immunosuppressants | 1 ML ABATACEPT 125 MG/ML PREFILLED SYRINGE [ORENCIA] |
| 1359480 | PR00 | Immunosuppressants | ABATACEPT 125 MG/ML PREFILLED SYRINGE [ORENCIA] |
| 1799230 | PR00 | Immunosuppressants | 1 ML ABATACEPT 125 MG/ML AUTO-INJECTOR [ORENCIA] |
| 1799232 | PR00 | Immunosuppressants | ABATACEPT 125 MG/ML AUTO-INJECTOR [ORENCIA] |
| 1925255 | PR00 | Immunosuppressants | 0.4 ML ABATACEPT 125 MG/ML PREFILLED SYRINGE [ORENCIA] |
| 1925257 | PR00 | Immunosuppressants | 0.7 ML ABATACEPT 125 MG/ML PREFILLED SYRINGE [ORENCIA] |
| 616018 | PR00 | Immunosuppressants | ABATACEPT 250 MG INJECTION [ORENCIA] |
| 616207 | PR00 | Immunosuppressants | ABATACEPT 10 MG/ML INJECTABLE SOLUTION [ORENCIA] |
| 1145930 | PR00 | Immunosuppressants | ABATACEPT 125 MG/ML [ORENCIA] |
| 1659774 | PR00 | Immunosuppressants | ABATACEPT 250 MG [ORENCIA] |
| 616016 | PR00 | Immunosuppressants | ABATACEPT 25 MG/ML [ORENCIA] |
| 616206 | PR00 | Immunosuppressants | ABATACEPT 10 MG/ML [ORENCIA] |
| 1145931 | PR00 | Immunosuppressants | ABATACEPT PREFILLED SYRINGE [ORENCIA] |
| 1659775 | PR00 | Immunosuppressants | ABATACEPT INJECTION [ORENCIA] |
| 1799229 | PR00 | Immunosuppressants | ABATACEPT AUTO-INJECTOR [ORENCIA] |
| 616017 | PR00 | Immunosuppressants | ABATACEPT INJECTABLE SOLUTION [ORENCIA] |
| 1183963 | PR00 | Immunosuppressants | ORENCIA INJECTABLE PRODUCT |
| 1145929 | PR00 | Immunosuppressants | 1 ML ABATACEPT 125 MG/ML PREFILLED SYRINGE |
| 1360514 | PR00 | Immunosuppressants | ABATACEPT 125 MG/ML PREFILLED SYRINGE |
| 1799228 | PR00 | Immunosuppressants | 1 ML ABATACEPT 125 MG/ML AUTO-INJECTOR |
| 1799231 | PR00 | Immunosuppressants | ABATACEPT 125 MG/ML AUTO-INJECTOR |
| 1925254 | PR00 | Immunosuppressants | 0.4 ML ABATACEPT 125 MG/ML PREFILLED SYRINGE |
| 1925256 | PR00 | Immunosuppressants | 0.7 ML ABATACEPT 125 MG/ML PREFILLED SYRINGE |
| 607381 | PR00 | Immunosuppressants | ABATACEPT 10 MG/ML INJECTABLE SOLUTION |
| 616015 | PR00 | Immunosuppressants | ABATACEPT 250 MG INJECTION |
| 1145927 | PR00 | Immunosuppressants | ABATACEPT 125 MG/ML |
| 1659771 | PR00 | Immunosuppressants | ABATACEPT 250 MG |
| 607379 | PR00 | Immunosuppressants | ABATACEPT 10 MG/ML |
| 616014 | PR00 | Immunosuppressants | ABATACEPT 25 MG/ML |
| 1145928 | PR00 | Immunosuppressants | ABATACEPT PREFILLED SYRINGE |
| 1659772 | PR00 | Immunosuppressants | ABATACEPT INJECTION |
| 1799227 | PR00 | Immunosuppressants | ABATACEPT AUTO-INJECTOR |
| 607380 | PR00 | Immunosuppressants | ABATACEPT INJECTABLE SOLUTION |
| 1163066 | PR00 | Immunosuppressants | ABATACEPT INJECTABLE PRODUCT |
| 353484 | PR00 | Immunosuppressants | HUMIRA |
| 1594352 | PR00 | Immunosuppressants | {6 (0.8 ML ADALIMUMAB 50 MG/ML PREFILLED SYRINGE [HUMIRA]) } PACK [HUMIRA PEDIATRIC CROHN'S DISEASE STARTER PACKAGE (6 COUNT)] |
| 1594358 | PR00 | Immunosuppressants | {3 (0.8 ML ADALIMUMAB 50 MG/ML PREFILLED SYRINGE [HUMIRA]) } PACK [HUMIRA PEDIATRIC CROHN'S DISEASE STARTER PACKAGE (3 COUNT)] |
| 1801278 | PR00 | Immunosuppressants | {4 (0.8 ML ADALIMUMAB 50 MG/ML AUTO-INJECTOR [HUMIRA]) } PACK [HUMIRA PEN - PSORIASIS STARTER PACK] |
| 1855527 | PR00 | Immunosuppressants | {1 (0.4 ML ADALIMUMAB 100 MG/ML PREFILLED SYRINGE [HUMIRA]) / 1 (0.8 ML ADALIMUMAB 100 MG/ML PREFILLED SYRINGE [HUMIRA]) } PACK [HUMIRA PEDIATRIC CROHN'S DISEASE STARTER PACKAGE (2 COUNT)] |
| 1872984 | PR00 | Immunosuppressants | {6 (0.4 ML ADALIMUMAB 100 MG/ML AUTO-INJECTOR [HUMIRA]) } PACK [HUMIRA PEN 40 MG/0.4 ML STARTER PACK - CROHN'S DISEASE] |
| 1872986 | PR00 | Immunosuppressants | {4 (0.4 ML ADALIMUMAB 100 MG/ML AUTO-INJECTOR [HUMIRA]) } PACK [HUMIRA PEN 40 MG/0.4 ML STARTER PACK - PSORIASIS] |
| 1873087 | PR00 | Immunosuppressants | {3 (0.8 ML ADALIMUMAB 100 MG/ML PREFILLED SYRINGE [HUMIRA]) } PACK [HUMIRA PREFILLED SYRINGE 80 MG/0.8 ML STARTER PACK - PEDIATRIC CROHN'S DISEASE] |
| 1921245 | PR00 | Immunosuppressants | {3 (0.8 ML ADALIMUMAB 100 MG/ML AUTO-INJECTOR [HUMIRA]) } PACK [HUMIRA PEN 80 MG/0.8 ML - STARTER PACKAGE FOR CROHN'S DISEASE, ULCERATIVE COLITIS OR HIDRADENITIS SUPPURATIVA] |
| 1921468 | PR00 | Immunosuppressants | {2 (0.4 ML ADALIMUMAB 100 MG/ML AUTO-INJECTOR [HUMIRA]) / 1 (0.8 ML ADALIMUMAB 100 MG/ML AUTO-INJECTOR [HUMIRA]) } PACK [HUMIRA PEN 80 MG/0.8 ML AND 40 MG/0.4 ML - PSORIASIS/UVEITIS STARTER PACKAGE] |

| | | | |
|---|---|---|---|
| 825170 | PR00 | Immunosuppressants | {6 (0.8 ML ADALIMUMAB 50 MG/ML AUTO-INJECTOR [HUMIRA]) } PACK [HUMIRA PEN - CROHN'S DISEASE STARTER PACK] |
| 1594357 | PR00 | Immunosuppressants | {3 (0.8 ML ADALIMUMAB 50 MG/ML PREFILLED SYRINGE) } PACK |
| 1656703 | PR00 | Immunosuppressants | {6 (0.8 ML ADALIMUMAB 50 MG/ML AUTO-INJECTOR) } PACK |
| 1801277 | PR00 | Immunosuppressants | {4 (0.8 ML ADALIMUMAB 50 MG/ML AUTO-INJECTOR) } PACK |
| 1855526 | PR00 | Immunosuppressants | {1 (0.4 ML ADALIMUMAB 100 MG/ML PREFILLED SYRINGE) / 1 (0.8 ML ADALIMUMAB 100 MG/ML PREFILLED SYRINGE) } PACK |
| 1872983 | PR00 | Immunosuppressants | {6 (0.4 ML ADALIMUMAB 100 MG/ML AUTO-INJECTOR) } PACK |
| 1872985 | PR00 | Immunosuppressants | {4 (0.4 ML ADALIMUMAB 100 MG/ML AUTO-INJECTOR) } PACK |
| 1873086 | PR00 | Immunosuppressants | {3 (0.8 ML ADALIMUMAB 100 MG/ML PREFILLED SYRINGE) } PACK |
| 1921244 | PR00 | Immunosuppressants | {3 (0.8 ML ADALIMUMAB 100 MG/ML AUTO-INJECTOR) } PACK |
| 1921467 | PR00 | Immunosuppressants | {2 (0.4 ML ADALIMUMAB 100 MG/ML AUTO-INJECTOR) / 1 (0.8 ML ADALIMUMAB 100 MG/ML AUTO-INJECTOR) } PACK |
| 825169 | PR00 | Immunosuppressants | {6 (0.8 ML ADALIMUMAB 50 MG/ML PREFILLED SYRINGE) } PACK |
| 327361 | PR00 | Immunosuppressants | ADALIMUMAB |
| 2103477 | PR00 | Immunosuppressants | ADALIMUMAB-ADBM |
| 2103479 | PR00 | Immunosuppressants | ADALIMUMAB-ATTO |
| 1359755 | PR00 | Immunosuppressants | ADALIMUMAB 50 MG/ML PREFILLED SYRINGE [HUMIRA] |
| 1551888 | PR00 | Immunosuppressants | 0.2 ML ADALIMUMAB 50 MG/ML PREFILLED SYRINGE [HUMIRA] |
| 1655728 | PR00 | Immunosuppressants | 0.8 ML ADALIMUMAB 50 MG/ML AUTO-INJECTOR [HUMIRA] |
| 1655730 | PR00 | Immunosuppressants | ADALIMUMAB 50 MG/ML AUTO-INJECTOR [HUMIRA] |
| 1658078 | PR00 | Immunosuppressants | ADALIMUMAB 50 MG/ML INJECTION [HUMIRA] |
| 1726846 | PR00 | Immunosuppressants | 0.4 ML ADALIMUMAB 100 MG/ML PREFILLED SYRINGE [HUMIRA] |
| 1726848 | PR00 | Immunosuppressants | ADALIMUMAB 100 MG/ML PREFILLED SYRINGE [HUMIRA] |
| 1855524 | PR00 | Immunosuppressants | 0.8 ML ADALIMUMAB 100 MG/ML PREFILLED SYRINGE [HUMIRA] |
| 1872980 | PR00 | Immunosuppressants | 0.4 ML ADALIMUMAB 100 MG/ML AUTO-INJECTOR [HUMIRA] |
| 1872982 | PR00 | Immunosuppressants | ADALIMUMAB 100 MG/ML AUTO-INJECTOR [HUMIRA] |
| 1921010 | PR00 | Immunosuppressants | 0.1 ML ADALIMUMAB 100 MG/ML PREFILLED SYRINGE [HUMIRA] |
| 1921017 | PR00 | Immunosuppressants | 0.2 ML ADALIMUMAB 100 MG/ML PREFILLED SYRINGE [HUMIRA] |
| 1921240 | PR00 | Immunosuppressants | 0.8 ML ADALIMUMAB 100 MG/ML AUTO-INJECTOR [HUMIRA] |
| 1921242 | PR00 | Immunosuppressants | ADALIMUMAB 100 MG/ML PEN INJECTOR [HUMIRA] |
| 1921466 | PR00 | Immunosuppressants | 0.4 ML ADALIMUMAB 100 MG/ML PEN INJECTOR [HUMIRA] |
| 352334 | PR00 | Immunosuppressants | 0.8 ML ADALIMUMAB 50 MG/ML INJECTION [HUMIRA] |
| 727705 | PR00 | Immunosuppressants | 0.8 ML ADALIMUMAB 50 MG/ML PREFILLED SYRINGE [HUMIRA] |
| 763565 | PR00 | Immunosuppressants | 0.4 ML ADALIMUMAB 50 MG/ML PREFILLED SYRINGE [HUMIRA] |
| 1726845 | PR00 | Immunosuppressants | ADALIMUMAB 100 MG/ML [HUMIRA] |
| 576023 | PR00 | Immunosuppressants | ADALIMUMAB 50 MG/ML [HUMIRA] |
| 1655727 | PR00 | Immunosuppressants | ADALIMUMAB AUTO-INJECTOR [HUMIRA] |
| 1656698 | PR00 | Immunosuppressants | ADALIMUMAB INJECTION [HUMIRA] |
| 1921239 | PR00 | Immunosuppressants | ADALIMUMAB PEN INJECTOR [HUMIRA] |
| 363562 | PR00 | Immunosuppressants | ADALIMUMAB INJECTABLE SOLUTION [HUMIRA] |
| 727704 | PR00 | Immunosuppressants | ADALIMUMAB PREFILLED SYRINGE [HUMIRA] |
| 1171992 | PR00 | Immunosuppressants | HUMIRA INJECTABLE PRODUCT |
| 1360097 | PR00 | Immunosuppressants | ADALIMUMAB 50 MG/ML PREFILLED SYRINGE |
| 1551887 | PR00 | Immunosuppressants | 0.2 ML ADALIMUMAB 50 MG/ML PREFILLED SYRINGE |
| 1655726 | PR00 | Immunosuppressants | 0.8 ML ADALIMUMAB 50 MG/ML AUTO-INJECTOR |
| 1655729 | PR00 | Immunosuppressants | ADALIMUMAB 50 MG/ML AUTO-INJECTOR |
| 1658077 | PR00 | Immunosuppressants | ADALIMUMAB 50 MG/ML INJECTION |
| 1726844 | PR00 | Immunosuppressants | 0.4 ML ADALIMUMAB 100 MG/ML PREFILLED SYRINGE |
| 1726847 | PR00 | Immunosuppressants | ADALIMUMAB 100 MG/ML PREFILLED SYRINGE |
| 1855523 | PR00 | Immunosuppressants | 0.8 ML ADALIMUMAB 100 MG/ML PREFILLED SYRINGE |
| 1872979 | PR00 | Immunosuppressants | 0.4 ML ADALIMUMAB 100 MG/ML AUTO-INJECTOR |
| 1872981 | PR00 | Immunosuppressants | ADALIMUMAB 100 MG/ML AUTO-INJECTOR |
| 1921009 | PR00 | Immunosuppressants | 0.1 ML ADALIMUMAB 100 MG/ML PREFILLED SYRINGE |
| 1921016 | PR00 | Immunosuppressants | 0.2 ML ADALIMUMAB 100 MG/ML PREFILLED SYRINGE |
| 1921238 | PR00 | Immunosuppressants | 0.8 ML ADALIMUMAB 100 MG/ML AUTO-INJECTOR |
| 1921241 | PR00 | Immunosuppressants | ADALIMUMAB 100 MG/ML PEN INJECTOR |
| 1921465 | PR00 | Immunosuppressants | 0.4 ML ADALIMUMAB 100 MG/ML PEN INJECTOR |
| 351290 | PR00 | Immunosuppressants | 0.8 ML ADALIMUMAB 50 MG/ML INJECTION |
| 727703 | PR00 | Immunosuppressants | 0.8 ML ADALIMUMAB 50 MG/ML PREFILLED SYRINGE |
| 763564 | PR00 | Immunosuppressants | 0.4 ML ADALIMUMAB 50 MG/ML PREFILLED SYRINGE |
| 1726843 | PR00 | Immunosuppressants | ADALIMUMAB 100 MG/ML |
| 358817 | PR00 | Immunosuppressants | ADALIMUMAB 50 MG/ML |
| 1655725 | PR00 | Immunosuppressants | ADALIMUMAB AUTO-INJECTOR |
| 1656695 | PR00 | Immunosuppressants | ADALIMUMAB INJECTION |
| 1921237 | PR00 | Immunosuppressants | ADALIMUMAB PEN INJECTOR |
| 378758 | PR00 | Immunosuppressants | ADALIMUMAB INJECTABLE SOLUTION |
| 727702 | PR00 | Immunosuppressants | ADALIMUMAB PREFILLED SYRINGE |
| 1163691 | PR00 | Immunosuppressants | ADALIMUMAB INJECTABLE PRODUCT |
| 1594659 | PR00 | Immunosuppressants | LEMTRADA |
| 284679 | PR00 | Immunosuppressants | CAMPATH |

| | | | |
|---|---|---|---|
| 117055 | PR00 | Immunosuppressants | ALEMTUZUMAB |
| 1594663 | PR00 | Immunosuppressants | 1.2 ML ALEMTUZUMAB 10 MG/ML INJECTION [LEMTRADA] |
| 1656635 | PR00 | Immunosuppressants | ALEMTUZUMAB 30 MG/ML INJECTION [CAMPATH] |
| 1656640 | PR00 | Immunosuppressants | ALEMTUZUMAB 10 MG/ML INJECTION [LEMTRADA] |
| 351967 | PR00 | Immunosuppressants | ALEMTUZUMAB 10 MG/ML INJECTABLE SOLUTION [CAMPATH] |
| 543438 | PR00 | Immunosuppressants | ALEMTUZUMAB 30 MG/ML INJECTABLE SOLUTION [CAMPATH] |
| 828267 | PR00 | Immunosuppressants | 1 ML ALEMTUZUMAB 30 MG/ML INJECTION [CAMPATH] |
| 1594660 | PR00 | Immunosuppressants | ALEMTUZUMAB 10 MG/ML [LEMTRADA] |
| 1656631 | PR00 | Immunosuppressants | ALEMTUZUMAB 30 MG/ML [CAMPATH] |
| 543437 | PR00 | Immunosuppressants | ALEMTUZUMAB 30 MG/ML [CAMPATH] |
| 575719 | PR00 | Immunosuppressants | ALEMTUZUMAB 10 MG/ML [CAMPATH] |
| 828266 | PR00 | Immunosuppressants | ALEMTUZUMAB 0.3 MG/ML [CAMPATH] |
| 1594661 | PR00 | Immunosuppressants | ALEMTUZUMAB INJECTABLE SOLUTION [LEMTRADA] |
| 1656632 | PR00 | Immunosuppressants | ALEMTUZUMAB INJECTION [CAMPATH] |
| 1656637 | PR00 | Immunosuppressants | ALEMTUZUMAB INJECTION [LEMTRADA] |
| 362780 | PR00 | Immunosuppressants | ALEMTUZUMAB INJECTABLE SOLUTION [CAMPATH] |
| 1172298 | PR00 | Immunosuppressants | CAMPATH INJECTABLE PRODUCT |
| 1594662 | PR00 | Immunosuppressants | LEMTRADA INJECTABLE PRODUCT |
| 1594658 | PR00 | Immunosuppressants | 1.2 ML ALEMTUZUMAB 10 MG/ML INJECTION |
| 1656634 | PR00 | Immunosuppressants | ALEMTUZUMAB 30 MG/ML INJECTION |
| 1656639 | PR00 | Immunosuppressants | ALEMTUZUMAB 10 MG/ML INJECTION |
| 284064 | PR00 | Immunosuppressants | ALEMTUZUMAB 10 MG/ML INJECTABLE SOLUTION |
| 485516 | PR00 | Immunosuppressants | ALEMTUZUMAB 30 MG/ML INJECTABLE SOLUTION |
| 828265 | PR00 | Immunosuppressants | 1 ML ALEMTUZUMAB 30 MG/ML INJECTION |
| 1594657 | PR00 | Immunosuppressants | ALEMTUZUMAB 10 MG/ML |
| 1656628 | PR00 | Immunosuppressants | ALEMTUZUMAB 30 MG/ML |
| 343434 | PR00 | Immunosuppressants | ALEMTUZUMAB 10 MG/ML |
| 485515 | PR00 | Immunosuppressants | ALEMTUZUMAB 30 MG/ML |
| 828264 | PR00 | Immunosuppressants | ALEMTUZUMAB 0.3 MG/ML |
| 1656629 | PR00 | Immunosuppressants | ALEMTUZUMAB INJECTION |
| 378757 | PR00 | Immunosuppressants | ALEMTUZUMAB INJECTABLE SOLUTION |
| 1164265 | PR00 | Immunosuppressants | ALEMTUZUMAB INJECTABLE PRODUCT |
| 1112977 | PR00 | Immunosuppressants | NULOJIX |
| 1112973 | PR00 | Immunosuppressants | BELATACEPT |
| 1112980 | PR00 | Immunosuppressants | BELATACEPT 250 MG INJECTION [NULOJIX] |
| 1112978 | PR00 | Immunosuppressants | BELATACEPT 25 MG/ML [NULOJIX] |
| 1804973 | PR00 | Immunosuppressants | BELATACEPT 250 MG [NULOJIX] |
| 1112979 | PR00 | Immunosuppressants | BELATACEPT INJECTABLE SOLUTION [NULOJIX] |
| 1804974 | PR00 | Immunosuppressants | BELATACEPT INJECTION [NULOJIX] |
| 1178807 | PR00 | Immunosuppressants | NULOJIX INJECTABLE PRODUCT |
| 1112976 | PR00 | Immunosuppressants | BELATACEPT 250 MG INJECTION |
| 1112974 | PR00 | Immunosuppressants | BELATACEPT 25 MG/ML |
| 1804970 | PR00 | Immunosuppressants | BELATACEPT 250 MG |
| 1112975 | PR00 | Immunosuppressants | BELATACEPT INJECTABLE SOLUTION |
| 1804971 | PR00 | Immunosuppressants | BELATACEPT INJECTION |
| 1155316 | PR00 | Immunosuppressants | BELATACEPT INJECTABLE PRODUCT |
| 1092441 | PR00 | Immunosuppressants | BENLYSTA |
| 1092437 | PR00 | Immunosuppressants | BELIMUMAB |
| 1092444 | PR00 | Immunosuppressants | BELIMUMAB 80 MG/ML INJECTABLE SOLUTION [BENLYSTA] |
| 1656398 | PR00 | Immunosuppressants | BELIMUMAB 120 MG INJECTION [BENLYSTA] |
| 1656402 | PR00 | Immunosuppressants | BELIMUMAB 400 MG INJECTION [BENLYSTA] |
| 1939299 | PR00 | Immunosuppressants | 1 ML BELIMUMAB 200 MG/ML AUTO-INJECTOR [BENLYSTA] |
| 1939301 | PR00 | Immunosuppressants | BELIMUMAB 200 MG/ML AUTO-INJECTOR [BENLYSTA] |
| 1939348 | PR00 | Immunosuppressants | 1 ML BELIMUMAB 200 MG/ML PREFILLED SYRINGE [BENLYSTA] |
| 1939350 | PR00 | Immunosuppressants | BELIMUMAB 200 MG/ML PREFILLED SYRINGE [BENLYSTA] |
| 1092442 | PR00 | Immunosuppressants | BELIMUMAB 80 MG/ML [BENLYSTA] |
| 1656396 | PR00 | Immunosuppressants | BELIMUMAB 120 MG [BENLYSTA] |
| 1656401 | PR00 | Immunosuppressants | BELIMUMAB 400 MG [BENLYSTA] |
| 1939297 | PR00 | Immunosuppressants | BELIMUMAB 200 MG/ML [BENLYSTA] |
| 1092443 | PR00 | Immunosuppressants | BELIMUMAB INJECTABLE SOLUTION [BENLYSTA] |
| 1656397 | PR00 | Immunosuppressants | BELIMUMAB INJECTION [BENLYSTA] |
| 1939298 | PR00 | Immunosuppressants | BELIMUMAB AUTO-INJECTOR [BENLYSTA] |
| 1939347 | PR00 | Immunosuppressants | BELIMUMAB PREFILLED SYRINGE [BENLYSTA] |
| 1171464 | PR00 | Immunosuppressants | BENLYSTA INJECTABLE PRODUCT |
| 1092440 | PR00 | Immunosuppressants | BELIMUMAB 80 MG/ML INJECTABLE SOLUTION |
| 1656395 | PR00 | Immunosuppressants | BELIMUMAB 120 MG INJECTION |
| 1656400 | PR00 | Immunosuppressants | BELIMUMAB 400 MG INJECTION |
| 1939296 | PR00 | Immunosuppressants | 1 ML BELIMUMAB 200 MG/ML AUTO-INJECTOR |
| 1939300 | PR00 | Immunosuppressants | BELIMUMAB 200 MG/ML AUTO-INJECTOR |
| 1939346 | PR00 | Immunosuppressants | 1 ML BELIMUMAB 200 MG/ML PREFILLED SYRINGE |
| 1939349 | PR00 | Immunosuppressants | BELIMUMAB 200 MG/ML PREFILLED SYRINGE |

| | | | |
|---|---|---|---|
| 1092438 | PR00 | Immunosuppressants | BELIMUMAB 80 MG/ML |
| 1656393 | PR00 | Immunosuppressants | BELIMUMAB 120 MG |
| 1656399 | PR00 | Immunosuppressants | BELIMUMAB 400 MG |
| 1939294 | PR00 | Immunosuppressants | BELIMUMAB 200 MG/ML |
| 1092439 | PR00 | Immunosuppressants | BELIMUMAB INJECTABLE SOLUTION |
| 1656394 | PR00 | Immunosuppressants | BELIMUMAB INJECTION |
| 1939295 | PR00 | Immunosuppressants | BELIMUMAB AUTO-INJECTOR |
| 1939345 | PR00 | Immunosuppressants | BELIMUMAB PREFILLED SYRINGE |
| 1159402 | PR00 | Immunosuppressants | BELIMUMAB INJECTABLE PRODUCT |
| 853495 | PR00 | Immunosuppressants | ILARIS |
| 853491 | PR00 | Immunosuppressants | CANAKINUMAB |
| 1864326 | PR00 | Immunosuppressants | 1 ML CANAKINUMAB 150 MG/ML INJECTION [ILARIS] |
| 1864328 | PR00 | Immunosuppressants | CANAKINUMAB 150 MG/ML INJECTION [ILARIS] |
| 853498 | PR00 | Immunosuppressants | CANAKINUMAB 180 MG INJECTION [ILARIS] |
| 1655974 | PR00 | Immunosuppressants | CANAKINUMAB 180 MG [ILARIS] |
| 1864325 | PR00 | Immunosuppressants | CANAKINUMAB 150 MG/ML [ILARIS] |
| 853496 | PR00 | Immunosuppressants | CANAKINUMAB 150 MG/ML [ILARIS] |
| 1655975 | PR00 | Immunosuppressants | CANAKINUMAB INJECTION [ILARIS] |
| 853497 | PR00 | Immunosuppressants | CANAKINUMAB INJECTABLE SOLUTION [ILARIS] |
| 1167137 | PR00 | Immunosuppressants | ILARIS INJECTABLE PRODUCT |
| 1864324 | PR00 | Immunosuppressants | 1 ML CANAKINUMAB 150 MG/ML INJECTION |
| 1864327 | PR00 | Immunosuppressants | CANAKINUMAB 150 MG/ML INJECTION |
| 853494 | PR00 | Immunosuppressants | CANAKINUMAB 180 MG INJECTION |
| 1655971 | PR00 | Immunosuppressants | CANAKINUMAB 180 MG |
| 1864323 | PR00 | Immunosuppressants | CANAKINUMAB 150 MG/ML |
| 853492 | PR00 | Immunosuppressants | CANAKINUMAB 150 MG/ML |
| 1655972 | PR00 | Immunosuppressants | CANAKINUMAB INJECTION |
| 853493 | PR00 | Immunosuppressants | CANAKINUMAB INJECTABLE SOLUTION |
| 1156606 | PR00 | Immunosuppressants | CANAKINUMAB INJECTABLE PRODUCT |
| 795082 | PR00 | Immunosuppressants | CIMZIA |
| 709271 | PR00 | Immunosuppressants | CERTOLIZUMAB PEGOL |
| 1359935 | PR00 | Immunosuppressants | CERTOLIZUMAB PEGOL 200 MG/ML PREFILLED SYRINGE [CIMZIA] |
| 795085 | PR00 | Immunosuppressants | CERTOLIZUMAB PEGOL 200 MG INJECTION [CIMZIA] |
| 849599 | PR00 | Immunosuppressants | 1 ML CERTOLIZUMAB PEGOL 200 MG/ML PREFILLED SYRINGE [CIMZIA] |
| 1653730 | PR00 | Immunosuppressants | CERTOLIZUMAB PEGOL 200 MG [CIMZIA] |
| 795083 | PR00 | Immunosuppressants | CERTOLIZUMAB PEGOL 200 MG/ML [CIMZIA] |
| 1653731 | PR00 | Immunosuppressants | CERTOLIZUMAB PEGOL INJECTION [CIMZIA] |
| 795084 | PR00 | Immunosuppressants | CERTOLIZUMAB PEGOL INJECTABLE SOLUTION [CIMZIA] |
| 849598 | PR00 | Immunosuppressants | CERTOLIZUMAB PEGOL PREFILLED SYRINGE [CIMZIA] |
| 1170885 | PR00 | Immunosuppressants | CIMZIA INJECTABLE PRODUCT |
| 1360081 | PR00 | Immunosuppressants | CERTOLIZUMAB PEGOL 200 MG/ML PREFILLED SYRINGE |
| 795081 | PR00 | Immunosuppressants | CERTOLIZUMAB PEGOL 200 MG INJECTION |
| 849597 | PR00 | Immunosuppressants | 1 ML CERTOLIZUMAB PEGOL 200 MG/ML PREFILLED SYRINGE |
| 1653727 | PR00 | Immunosuppressants | CERTOLIZUMAB PEGOL 200 MG |
| 795079 | PR00 | Immunosuppressants | CERTOLIZUMAB PEGOL 200 MG/ML |
| 1653728 | PR00 | Immunosuppressants | CERTOLIZUMAB PEGOL INJECTION |
| 795080 | PR00 | Immunosuppressants | CERTOLIZUMAB PEGOL INJECTABLE SOLUTION |
| 849596 | PR00 | Immunosuppressants | CERTOLIZUMAB PEGOL PREFILLED SYRINGE |
| 1161226 | PR00 | Immunosuppressants | CERTOLIZUMAB PEGOL INJECTABLE PRODUCT |
| 202994 | PR00 | Immunosuppressants | LEUKERAN |
| 2346 | PR00 | Immunosuppressants | CHLORAMBUCIL |
| 105553 | PR00 | Immunosuppressants | CHLORAMBUCIL 2 MG ORAL TABLET [LEUKERAN] |
| 105554 | PR00 | Immunosuppressants | CHLORAMBUCIL 5 MG ORAL TABLET [LEUKERAN] |
| 564080 | PR00 | Immunosuppressants | CHLORAMBUCIL 2 MG [LEUKERAN] |
| 564081 | PR00 | Immunosuppressants | CHLORAMBUCIL 5 MG [LEUKERAN] |
| 368349 | PR00 | Immunosuppressants | CHLORAMBUCIL ORAL TABLET [LEUKERAN] |
| 1166066 | PR00 | Immunosuppressants | LEUKERAN ORAL PRODUCT |
| 1166067 | PR00 | Immunosuppressants | LEUKERAN PILL |
| 197462 | PR00 | Immunosuppressants | CHLORAMBUCIL 2 MG ORAL TABLET |
| 199311 | PR00 | Immunosuppressants | CHLORAMBUCIL 5 MG ORAL TABLET |
| 249898 | PR00 | Immunosuppressants | CHLORAMBUCIL 10 MG ORAL TABLET |
| 411288 | PR00 | Immunosuppressants | CHLORAMBUCIL 2 MG ORAL CAPSULE |
| 315626 | PR00 | Immunosuppressants | CHLORAMBUCIL 2 MG |
| 331665 | PR00 | Immunosuppressants | CHLORAMBUCIL 5 MG |
| 332752 | PR00 | Immunosuppressants | CHLORAMBUCIL 10 MG |
| 371375 | PR00 | Immunosuppressants | CHLORAMBUCIL ORAL TABLET |
| 438413 | PR00 | Immunosuppressants | CHLORAMBUCIL ORAL CAPSULE |
| 1153038 | PR00 | Immunosuppressants | CHLORAMBUCIL ORAL PRODUCT |
| 1153039 | PR00 | Immunosuppressants | CHLORAMBUCIL PILL |
| 680973 | PR00 | Immunosuppressants | SOLIRIS |
| 591781 | PR00 | Immunosuppressants | ECULIZUMAB |

| | | | |
|---|---|---|---|
| 1655952 | PR00 | Immunosuppressants | ECULIZUMAB 10 MG/ML INJECTION [SOLIRIS] |
| 700387 | PR00 | Immunosuppressants | 30 ML ECULIZUMAB 10 MG/ML INJECTION [SOLIRIS] |
| 700385 | PR00 | Immunosuppressants | ECULIZUMAB 10 MG/ML [SOLIRIS] |
| 1655949 | PR00 | Immunosuppressants | ECULIZUMAB INJECTION [SOLIRIS] |
| 700386 | PR00 | Immunosuppressants | ECULIZUMAB INJECTABLE SOLUTION [SOLIRIS] |
| 1184826 | PR00 | Immunosuppressants | SOLIRIS INJECTABLE PRODUCT |
| 1655951 | PR00 | Immunosuppressants | ECULIZUMAB 10 MG/ML INJECTION |
| 700384 | PR00 | Immunosuppressants | 30 ML ECULIZUMAB 10 MG/ML INJECTION |
| 700382 | PR00 | Immunosuppressants | ECULIZUMAB 10 MG/ML |
| 1655947 | PR00 | Immunosuppressants | ECULIZUMAB INJECTION |
| 700383 | PR00 | Immunosuppressants | ECULIZUMAB INJECTABLE SOLUTION |
| 1161281 | PR00 | Immunosuppressants | ECULIZUMAB INJECTABLE PRODUCT |
| 216891 | PR00 | Immunosuppressants | ENBREL |
| 214555 | PR00 | Immunosuppressants | ETANERCEPT |
| 2103480 | PR00 | Immunosuppressants | ETANERCEPT-SZZS |
| 1360430 | PR00 | Immunosuppressants | ETANERCEPT 50 MG/ML PREFILLED SYRINGE [ENBREL] |
| 1653225 | PR00 | Immunosuppressants | 1 ML ETANERCEPT 50 MG/ML AUTO-INJECTOR [ENBREL] |
| 1653227 | PR00 | Immunosuppressants | ETANERCEPT 50 MG/ML AUTO-INJECTOR [ENBREL] |
| 2182340 | PR00 | Immunosuppressants | 1 ML ETANERCEPT 50 MG/ML CARTRIDGE [ENBREL] |
| 2182342 | PR00 | Immunosuppressants | ETANERCEPT 50 MG/ML CARTRIDGE [ENBREL] |
| 261105 | PR00 | Immunosuppressants | ETANERCEPT 25 MG/ML INJECTABLE SOLUTION [ENBREL] |
| 582673 | PR00 | Immunosuppressants | ETANERCEPT 50 MG/ML INJECTABLE SOLUTION [ENBREL] |
| 802652 | PR00 | Immunosuppressants | 1 ML ETANERCEPT 50 MG/ML PREFILLED SYRINGE [ENBREL] |
| 802654 | PR00 | Immunosuppressants | 1 ML ETANERCEPT 50 MG/ML PREFILLED SYRINGE [ENBREL] |
| 809159 | PR00 | Immunosuppressants | 0.5 ML ETANERCEPT 50 MG/ML PREFILLED SYRINGE [ENBREL] |
| 574352 | PR00 | Immunosuppressants | ETANERCEPT 25 MG/ML [ENBREL] |
| 582672 | PR00 | Immunosuppressants | ETANERCEPT 50 MG/ML [ENBREL] |
| 1653224 | PR00 | Immunosuppressants | ETANERCEPT AUTO-INJECTOR [ENBREL] |
| 2182339 | PR00 | Immunosuppressants | ETANERCEPT CARTRIDGE [ENBREL] |
| 363197 | PR00 | Immunosuppressants | ETANERCEPT INJECTABLE SOLUTION [ENBREL] |
| 802651 | PR00 | Immunosuppressants | ETANERCEPT PREFILLED SYRINGE [ENBREL] |
| 1175957 | PR00 | Immunosuppressants | ENBREL INJECTABLE PRODUCT |
| 1359949 | PR00 | Immunosuppressants | ETANERCEPT 50 MG/ML PREFILLED SYRINGE |
| 1653223 | PR00 | Immunosuppressants | 1 ML ETANERCEPT 50 MG/ML AUTO-INJECTOR |
| 1653226 | PR00 | Immunosuppressants | ETANERCEPT 50 MG/ML AUTO-INJECTOR |
| 2182338 | PR00 | Immunosuppressants | 1 ML ETANERCEPT 50 MG/ML CARTRIDGE |
| 2182341 | PR00 | Immunosuppressants | ETANERCEPT 50 MG/ML CARTRIDGE |
| 253014 | PR00 | Immunosuppressants | ETANERCEPT 25 MG/ML INJECTABLE SOLUTION |
| 582671 | PR00 | Immunosuppressants | ETANERCEPT 50 MG/ML INJECTABLE SOLUTION |
| 727757 | PR00 | Immunosuppressants | 1 ML ETANERCEPT 50 MG/ML PREFILLED SYRINGE |
| 727904 | PR00 | Immunosuppressants | 0.51 ML ETANERCEPT 20 MG/ML PREFILLED SYRINGE |
| 802653 | PR00 | Immunosuppressants | 1 ML ETANERCEPT 50 MG/ML PREFILLED SYRINGE |
| 809158 | PR00 | Immunosuppressants | 0.5 ML ETANERCEPT 50 MG/ML PREFILLED SYRINGE |
| 342977 | PR00 | Immunosuppressants | ETANERCEPT 25 MG/ML |
| 582670 | PR00 | Immunosuppressants | ETANERCEPT 50 MG/ML |
| 727903 | PR00 | Immunosuppressants | ETANERCEPT 20 MG/ML |
| 1653222 | PR00 | Immunosuppressants | ETANERCEPT AUTO-INJECTOR |
| 2182337 | PR00 | Immunosuppressants | ETANERCEPT CARTRIDGE |
| 385098 | PR00 | Immunosuppressants | ETANERCEPT INJECTABLE SOLUTION |
| 727756 | PR00 | Immunosuppressants | ETANERCEPT PREFILLED SYRINGE |
| 1156809 | PR00 | Immunosuppressants | ETANERCEPT INJECTABLE PRODUCT |
| 848161 | PR00 | Immunosuppressants | SIMPONI |
| 819300 | PR00 | Immunosuppressants | GOLIMUMAB |
| 1360436 | PR00 | Immunosuppressants | GOLIMUMAB 100 MG/ML PREFILLED SYRINGE [SIMPONI] |
| 1431647 | PR00 | Immunosuppressants | 4 ML GOLIMUMAB 12.5 MG/ML INJECTION [SIMPONI] |
| 1482814 | PR00 | Immunosuppressants | 1 ML GOLIMUMAB 100 MG/ML PREFILLED SYRINGE [SIMPONI] |
| 1653144 | PR00 | Immunosuppressants | 0.5 ML GOLIMUMAB 100 MG/ML AUTO-INJECTOR [SIMPONI] |
| 1653146 | PR00 | Immunosuppressants | GOLIMUMAB 100 MG/ML AUTO-INJECTOR [SIMPONI] |
| 1653166 | PR00 | Immunosuppressants | 1 ML GOLIMUMAB 100 MG/ML AUTO-INJECTOR [SIMPONI] |
| 1653574 | PR00 | Immunosuppressants | GOLIMUMAB 12.5 MG/ML INJECTION [SIMPONI] |
| 848164 | PR00 | Immunosuppressants | 0.5 ML GOLIMUMAB 100 MG/ML PREFILLED SYRINGE [SIMPONI] |
| 1431644 | PR00 | Immunosuppressants | GOLIMUMAB 12.5 MG/ML [SIMPONI] |
| 848162 | PR00 | Immunosuppressants | GOLIMUMAB 100 MG/ML [SIMPONI] |
| 1431645 | PR00 | Immunosuppressants | GOLIMUMAB INJECTABLE SOLUTION [SIMPONI ARIA] |
| 1653143 | PR00 | Immunosuppressants | GOLIMUMAB AUTO-INJECTOR [SIMPONI] |
| 1653571 | PR00 | Immunosuppressants | GOLIMUMAB INJECTION [SIMPONI] |
| 848163 | PR00 | Immunosuppressants | GOLIMUMAB PREFILLED SYRINGE [SIMPONI] |
| 1182643 | PR00 | Immunosuppressants | SIMPONI INJECTABLE PRODUCT |
| 1359486 | PR00 | Immunosuppressants | GOLIMUMAB 100 MG/ML PREFILLED SYRINGE |
| 1431642 | PR00 | Immunosuppressants | 4 ML GOLIMUMAB 12.5 MG/ML INJECTION |
| 1482813 | PR00 | Immunosuppressants | 1 ML GOLIMUMAB 100 MG/ML PREFILLED SYRINGE |

| | | | |
|---|---|---|---|
| 1653142 | PR00 | Immunosuppressants | 0.5 ML GOLIMUMAB 100 MG/ML AUTO-INJECTOR |
| 1653145 | PR00 | Immunosuppressants | GOLIMUMAB 100 MG/ML AUTO-INJECTOR |
| 1653165 | PR00 | Immunosuppressants | 1 ML GOLIMUMAB 100 MG/ML AUTO-INJECTOR |
| 1653573 | PR00 | Immunosuppressants | GOLIMUMAB 12.5 MG/ML INJECTION |
| 848160 | PR00 | Immunosuppressants | 0.5 ML GOLIMUMAB 100 MG/ML PREFILLED SYRINGE |
| 1431639 | PR00 | Immunosuppressants | GOLIMUMAB 12.5 MG/ML |
| 848158 | PR00 | Immunosuppressants | GOLIMUMAB 100 MG/ML |
| 1431641 | PR00 | Immunosuppressants | GOLIMUMAB INJECTABLE SOLUTION |
| 1653141 | PR00 | Immunosuppressants | GOLIMUMAB AUTO-INJECTOR |
| 1653569 | PR00 | Immunosuppressants | GOLIMUMAB INJECTION |
| 848159 | PR00 | Immunosuppressants | GOLIMUMAB PREFILLED SYRINGE |
| 1157808 | PR00 | Immunosuppressants | GOLIMUMAB INJECTABLE PRODUCT |
| 1790542 | PR00 | Immunosuppressants | INFLECTRA |
| 1927286 | PR00 | Immunosuppressants | RENFLEXIS |
| 219610 | PR00 | Immunosuppressants | REMICADE |
| 2266526 | PR00 | Immunosuppressants | AVSOLA |
| 191831 | PR00 | Immunosuppressants | INFLIXIMAB |
| 1790539 | PR00 | Immunosuppressants | INFLIXIMAB-DYYB |
| 1927283 | PR00 | Immunosuppressants | INFLIXIMAB-ABDA |
| 2103476 | PR00 | Immunosuppressants | INFLIXIMAB-QBTX |
| 2266523 | PR00 | Immunosuppressants | INFLIXIMAB-AXXQ |
| 1790546 | PR00 | Immunosuppressants | INFLIXIMAB-DYYB 100 MG INJECTION [INFLECTRA] |
| 1927290 | PR00 | Immunosuppressants | INFLIXIMAB-ABDA 100 MG INJECTION [RENFLEXIS] |
| 213361 | PR00 | Immunosuppressants | INFLIXIMAB 100 MG INJECTION [REMICADE] |
| 2266530 | PR00 | Immunosuppressants | INFLIXIMAB-AXXQ 100 MG INJECTION [AVSOLA] |
| 1655943 | PR00 | Immunosuppressants | INFLIXIMAB 100 MG [REMICADE] |
| 1790543 | PR00 | Immunosuppressants | INFLIXIMAB-DYYB 100 MG [INFLECTRA] |
| 1927287 | PR00 | Immunosuppressants | INFLIXIMAB-ABDA 100 MG [RENFLEXIS] |
| 2266527 | PR00 | Immunosuppressants | INFLIXIMAB-AXXQ 100 MG [AVSOLA] |
| 573255 | PR00 | Immunosuppressants | INFLIXIMAB 10 MG/ML [REMICADE] |
| 1655944 | PR00 | Immunosuppressants | INFLIXIMAB INJECTION [REMICADE] |
| 1790544 | PR00 | Immunosuppressants | INFLIXIMAB INJECTION [INFLECTRA] |
| 1927288 | PR00 | Immunosuppressants | INFLIXIMAB INJECTION [RENFLEXIS] |
| 2266528 | PR00 | Immunosuppressants | INFLIXIMAB INJECTION [AVSOLA] |
| 363437 | PR00 | Immunosuppressants | INFLIXIMAB INJECTABLE SOLUTION [REMICADE] |
| 1180726 | PR00 | Immunosuppressants | REMICADE INJECTABLE PRODUCT |
| 1790545 | PR00 | Immunosuppressants | INFLECTRA INJECTABLE PRODUCT |
| 1927289 | PR00 | Immunosuppressants | RENFLEXIS INJECTABLE PRODUCT |
| 2266529 | PR00 | Immunosuppressants | AVSOLA INJECTABLE PRODUCT |
| 1790541 | PR00 | Immunosuppressants | INFLIXIMAB-DYYB 100 MG INJECTION |
| 1927285 | PR00 | Immunosuppressants | INFLIXIMAB-ABDA 100 MG INJECTION |
| 2266525 | PR00 | Immunosuppressants | INFLIXIMAB-AXXQ 100 MG INJECTION |
| 310994 | PR00 | Immunosuppressants | INFLIXIMAB 100 MG INJECTION |
| 1655940 | PR00 | Immunosuppressants | INFLIXIMAB 100 MG |
| 1790540 | PR00 | Immunosuppressants | INFLIXIMAB-DYYB 100 MG |
| 1927284 | PR00 | Immunosuppressants | INFLIXIMAB-ABDA 100 MG |
| 2266524 | PR00 | Immunosuppressants | INFLIXIMAB-AXXQ 100 MG |
| 358693 | PR00 | Immunosuppressants | INFLIXIMAB 10 MG/ML |
| 1655941 | PR00 | Immunosuppressants | INFLIXIMAB INJECTION |
| 376873 | PR00 | Immunosuppressants | INFLIXIMAB INJECTABLE SOLUTION |
| 1160692 | PR00 | Immunosuppressants | INFLIXIMAB INJECTABLE PRODUCT |
| 215393 | PR00 | Immunosuppressants | ARAVA |
| 27169 | PR00 | Immunosuppressants | LEFLUNOMIDE |
| 213377 | PR00 | Immunosuppressants | LEFLUNOMIDE 10 MG ORAL TABLET [ARAVA] |
| 213379 | PR00 | Immunosuppressants | LEFLUNOMIDE 20 MG ORAL TABLET [ARAVA] |
| 213380 | PR00 | Immunosuppressants | LEFLUNOMIDE 100 MG ORAL TABLET [ARAVA] |
| 573271 | PR00 | Immunosuppressants | LEFLUNOMIDE 10 MG [ARAVA] |
| 573273 | PR00 | Immunosuppressants | LEFLUNOMIDE 20 MG [ARAVA] |
| 573274 | PR00 | Immunosuppressants | LEFLUNOMIDE 100 MG [ARAVA] |
| 368264 | PR00 | Immunosuppressants | LEFLUNOMIDE ORAL TABLET [ARAVA] |
| 1170095 | PR00 | Immunosuppressants | ARAVA ORAL PRODUCT |
| 1170096 | PR00 | Immunosuppressants | ARAVA PILL |
| 205284 | PR00 | Immunosuppressants | LEFLUNOMIDE 10 MG ORAL TABLET |
| 205285 | PR00 | Immunosuppressants | LEFLUNOMIDE 20 MG ORAL TABLET |
| 205286 | PR00 | Immunosuppressants | LEFLUNOMIDE 100 MG ORAL TABLET |
| 316133 | PR00 | Immunosuppressants | LEFLUNOMIDE 10 MG |
| 316134 | PR00 | Immunosuppressants | LEFLUNOMIDE 20 MG |
| 317399 | PR00 | Immunosuppressants | LEFLUNOMIDE 100 MG |
| 372570 | PR00 | Immunosuppressants | LEFLUNOMIDE ORAL TABLET |
| 1162566 | PR00 | Immunosuppressants | LEFLUNOMIDE ORAL PRODUCT |
| 1162567 | PR00 | Immunosuppressants | LEFLUNOMIDE PILL |

| | | | |
|---|---|---|---|
| 1876381 | PR00 | Immunosuppressants | OCREVUS |
| 1876366 | PR00 | Immunosuppressants | OCRELIZUMAB |
| 1876385 | PR00 | Immunosuppressants | 10 ML OCRELIZUMAB 30 MG/ML INJECTION [OCREVUS] |
| 1876387 | PR00 | Immunosuppressants | OCRELIZUMAB 30 MG/ML INJECTION [OCREVUS] |
| 1876382 | PR00 | Immunosuppressants | OCRELIZUMAB 30 MG/ML [OCREVUS] |
| 1876383 | PR00 | Immunosuppressants | OCRELIZUMAB INJECTION [OCREVUS] |
| 1876384 | PR00 | Immunosuppressants | OCREVUS INJECTABLE PRODUCT |
| 1876380 | PR00 | Immunosuppressants | 10 ML OCRELIZUMAB 30 MG/ML INJECTION |
| 1876386 | PR00 | Immunosuppressants | OCRELIZUMAB 30 MG/ML INJECTION |
| 1876377 | PR00 | Immunosuppressants | OCRELIZUMAB 30 MG/ML |
| 1876379 | PR00 | Immunosuppressants | OCRELIZUMAB INJECTION |
| 1876378 | PR00 | Immunosuppressants | OCRELIZUMAB INJECTABLE PRODUCT |
| 876301 | PR00 | Immunosuppressants | ARZERRA |
| 712566 | PR00 | Immunosuppressants | OFATUMUMAB |
| 1658044 | PR00 | Immunosuppressants | 50 ML OFATUMUMAB 20 MG/ML INJECTION [ARZERRA] |
| 1658046 | PR00 | Immunosuppressants | OFATUMUMAB 20 MG/ML INJECTION [ARZERRA] |
| 1658048 | PR00 | Immunosuppressants | 5 ML OFATUMUMAB 20 MG/ML INJECTION [ARZERRA] |
| 876304 | PR00 | Immunosuppressants | OFATUMUMAB 10 MG/ML INJECTABLE SOLUTION [ARZERRA] |
| 877012 | PR00 | Immunosuppressants | OFATUMUMAB 20 MG/ML INJECTABLE SOLUTION [ARZERRA] |
| 876302 | PR00 | Immunosuppressants | OFATUMUMAB 10 MG/ML [ARZERRA] |
| 877011 | PR00 | Immunosuppressants | OFATUMUMAB 20 MG/ML [ARZERRA] |
| 1658043 | PR00 | Immunosuppressants | OFATUMUMAB INJECTION [ARZERRA] |
| 876303 | PR00 | Immunosuppressants | OFATUMUMAB INJECTABLE SOLUTION [ARZERRA] |
| 1171445 | PR00 | Immunosuppressants | ARZERRA INJECTABLE PRODUCT |
| 1658042 | PR00 | Immunosuppressants | 50 ML OFATUMUMAB 20 MG/ML INJECTION |
| 1658045 | PR00 | Immunosuppressants | OFATUMUMAB 20 MG/ML INJECTION |
| 1658047 | PR00 | Immunosuppressants | 5 ML OFATUMUMAB 20 MG/ML INJECTION |
| 876300 | PR00 | Immunosuppressants | OFATUMUMAB 10 MG/ML INJECTABLE SOLUTION |
| 877010 | PR00 | Immunosuppressants | OFATUMUMAB 20 MG/ML INJECTABLE SOLUTION |
| 876298 | PR00 | Immunosuppressants | OFATUMUMAB 10 MG/ML |
| 877009 | PR00 | Immunosuppressants | OFATUMUMAB 20 MG/ML |
| 1658041 | PR00 | Immunosuppressants | OFATUMUMAB INJECTION |
| 876299 | PR00 | Immunosuppressants | OFATUMUMAB INJECTABLE SOLUTION |
| 1159554 | PR00 | Immunosuppressants | OFATUMUMAB INJECTABLE PRODUCT |
| 763454 | PR00 | Immunosuppressants | ARCALYST |
| 763450 | PR00 | Immunosuppressants | RILONACEPT |
| 763457 | PR00 | Immunosuppressants | RILONACEPT 220 MG INJECTION [ARCALYST] |
| 1724261 | PR00 | Immunosuppressants | RILONACEPT 220 MG [ARCALYST] |
| 763455 | PR00 | Immunosuppressants | RILONACEPT 80 MG/ML [ARCALYST] |
| 1724262 | PR00 | Immunosuppressants | RILONACEPT INJECTION [ARCALYST] |
| 763456 | PR00 | Immunosuppressants | RILONACEPT INJECTABLE SOLUTION [ARCALYST] |
| 1170102 | PR00 | Immunosuppressants | ARCALYST INJECTABLE PRODUCT |
| 763453 | PR00 | Immunosuppressants | RILONACEPT 220 MG INJECTION |
| 1724258 | PR00 | Immunosuppressants | RILONACEPT 220 MG |
| 763451 | PR00 | Immunosuppressants | RILONACEPT 80 MG/ML |
| 1724259 | PR00 | Immunosuppressants | RILONACEPT INJECTION |
| 763452 | PR00 | Immunosuppressants | RILONACEPT INJECTABLE SOLUTION |
| 1157438 | PR00 | Immunosuppressants | RILONACEPT INJECTABLE PRODUCT |
| 1927886 | PR00 | Immunosuppressants | RITUXAN HYCELA |
| 2105827 | PR00 | Immunosuppressants | TRUXIMA |
| 226754 | PR00 | Immunosuppressants | RITUXAN |
| 2273513 | PR00 | Immunosuppressants | RUXIENCE |
| 121191 | PR00 | Immunosuppressants | RITUXIMAB |
| 1927884 | PR00 | Immunosuppressants | HYALURONIDASE / RITUXIMAB |
| 2105824 | PR00 | Immunosuppressants | RITUXIMAB-ABBS |
| 2273510 | PR00 | Immunosuppressants | RITUXIMAB-PVVR |
| 1657864 | PR00 | Immunosuppressants | 10 ML RITUXIMAB 10 MG/ML INJECTION [RITUXAN] |
| 1657866 | PR00 | Immunosuppressants | RITUXIMAB 10 MG/ML INJECTION [RITUXAN] |
| 1657868 | PR00 | Immunosuppressants | 50 ML RITUXIMAB 10 MG/ML INJECTION [RITUXAN] |
| 1927890 | PR00 | Immunosuppressants | 11.7 ML HYALURONIDASE, HUMAN RECOMBINANT 2000 UNT/ML / RITUXIMAB 120 MG/ML INJECTION [RITUXAN HYCELA] |
| 1927892 | PR00 | Immunosuppressants | HYALURONIDASE, HUMAN RECOMBINANT 2000 UNT/ML / RITUXIMAB 120 MG/ML INJECTION [RITUXAN HYCELA] |
| 1927894 | PR00 | Immunosuppressants | 13.4 ML HYALURONIDASE, HUMAN RECOMBINANT 2000 UNT/ML / RITUXIMAB 120 MG/ML INJECTION [RITUXAN HYCELA] |
| 2105831 | PR00 | Immunosuppressants | 10 ML RITUXIMAB-ABBS 10 MG/ML INJECTION [TRUXIMA] |
| 2105833 | PR00 | Immunosuppressants | RITUXIMAB-ABBS 10 MG/ML INJECTION [TRUXIMA] |
| 2105835 | PR00 | Immunosuppressants | 50 ML RITUXIMAB-ABBS 10 MG/ML INJECTION [TRUXIMA] |
| 213126 | PR00 | Immunosuppressants | RITUXIMAB 10 MG/ML INJECTABLE SOLUTION [RITUXAN] |
| 2273517 | PR00 | Immunosuppressants | 10 ML RITUXIMAB-PVVR 10 MG/ML INJECTION [RUXIENCE] |
| 2273519 | PR00 | Immunosuppressants | RITUXIMAB-PVVR 10 MG/ML INJECTION [RUXIENCE] |

| | | | |
|---|---|---|---|
| 2273521 | PR00 | Immunosuppressants | 50 ML RITUXIMAB-PVVR 10 MG/ML INJECTION [RUXIENCE] |
| 1927887 | PR00 | Immunosuppressants | HYALURONIDASE, HUMAN RECOMBINANT 2000 UNT/ML / RITUXIMAB 120 MG/ML [RITUXAN HYCELA] |
| 2105828 | PR00 | Immunosuppressants | RITUXIMAB-ABBS 10 MG/ML [TRUXIMA] |
| 2273514 | PR00 | Immunosuppressants | RITUXIMAB-PVVR 10 MG/ML [RUXIENCE] |
| 573051 | PR00 | Immunosuppressants | RITUXIMAB 10 MG/ML [RITUXAN] |
| 1657863 | PR00 | Immunosuppressants | RITUXIMAB INJECTION [RITUXAN] |
| 1927888 | PR00 | Immunosuppressants | HYALURONIDASE / RITUXIMAB INJECTION [RITUXAN HYCELA] |
| 2105829 | PR00 | Immunosuppressants | RITUXIMAB INJECTION [TRUXIMA] |
| 2273515 | PR00 | Immunosuppressants | RITUXIMAB INJECTION [RUXIENCE] |
| 362907 | PR00 | Immunosuppressants | RITUXIMAB INJECTABLE SOLUTION [RITUXAN] |
| 1185345 | PR00 | Immunosuppressants | RITUXAN INJECTABLE PRODUCT |
| 1927889 | PR00 | Immunosuppressants | RITUXAN HYCELA INJECTABLE PRODUCT |
| 2105830 | PR00 | Immunosuppressants | TRUXIMA INJECTABLE PRODUCT |
| 2273516 | PR00 | Immunosuppressants | RUXIENCE INJECTABLE PRODUCT |
| 1657862 | PR00 | Immunosuppressants | 10 ML RITUXIMAB 10 MG/ML INJECTION |
| 1657865 | PR00 | Immunosuppressants | RITUXIMAB 10 MG/ML INJECTION |
| 1657867 | PR00 | Immunosuppressants | 50 ML RITUXIMAB 10 MG/ML INJECTION |
| 1927885 | PR00 | Immunosuppressants | 11.7 ML HYALURONIDASE, HUMAN RECOMBINANT 2000 UNT/ML / RITUXIMAB 120 MG/ML INJECTION |
| 1927891 | PR00 | Immunosuppressants | HYALURONIDASE, HUMAN RECOMBINANT 2000 UNT/ML / RITUXIMAB 120 MG/ML INJECTION |
| 1927893 | PR00 | Immunosuppressants | 13.4 ML HYALURONIDASE, HUMAN RECOMBINANT 2000 UNT/ML / RITUXIMAB 120 MG/ML INJECTION |
| 2105826 | PR00 | Immunosuppressants | 10 ML RITUXIMAB-ABBS 10 MG/ML INJECTION |
| 2105832 | PR00 | Immunosuppressants | RITUXIMAB-ABBS 10 MG/ML INJECTION |
| 2105834 | PR00 | Immunosuppressants | 50 ML RITUXIMAB-ABBS 10 MG/ML INJECTION |
| 2273512 | PR00 | Immunosuppressants | 10 ML RITUXIMAB-PVVR 10 MG/ML INJECTION |
| 2273518 | PR00 | Immunosuppressants | RITUXIMAB-PVVR 10 MG/ML INJECTION |
| 2273520 | PR00 | Immunosuppressants | 50 ML RITUXIMAB-PVVR 10 MG/ML INJECTION |
| 242435 | PR00 | Immunosuppressants | RITUXIMAB 10 MG/ML INJECTABLE SOLUTION |
| 1927881 | PR00 | Immunosuppressants | RITUXIMAB 120 MG/ML |
| 2105825 | PR00 | Immunosuppressants | RITUXIMAB-ABBS 10 MG/ML |
| 2273511 | PR00 | Immunosuppressants | RITUXIMAB-PVVR 10 MG/ML |
| 316648 | PR00 | Immunosuppressants | RITUXIMAB 10 MG/ML |
| 1657861 | PR00 | Immunosuppressants | RITUXIMAB INJECTION |
| 1927883 | PR00 | Immunosuppressants | HYALURONIDASE / RITUXIMAB INJECTION |
| 377259 | PR00 | Immunosuppressants | RITUXIMAB INJECTABLE SOLUTION |
| 1157967 | PR00 | Immunosuppressants | RITUXIMAB INJECTABLE PRODUCT |
| 1927882 | PR00 | Immunosuppressants | HYALURONIDASE / RITUXIMAB INJECTABLE PRODUCT |
| 1599793 | PR00 | Immunosuppressants | COSENTYX |
| 1599788 | PR00 | Immunosuppressants | SECUKINUMAB |
| 1599797 | PR00 | Immunosuppressants | 1 ML SECUKINUMAB 150 MG/ML PREFILLED SYRINGE [COSENTYX] |
| 1599799 | PR00 | Immunosuppressants | SECUKINUMAB 150 MG/ML PREFILLED SYRINGE [COSENTYX] |
| 1653243 | PR00 | Immunosuppressants | 1 ML SECUKINUMAB 150 MG/ML AUTO-INJECTOR [COSENTYX] |
| 1653245 | PR00 | Immunosuppressants | SECUKINUMAB 150 MG/ML AUTO-INJECTOR [COSENTYX] |
| 1599794 | PR00 | Immunosuppressants | SECUKINUMAB 150 MG/ML [COSENTYX] |
| 1599795 | PR00 | Immunosuppressants | SECUKINUMAB PREFILLED SYRINGE [COSENTYX] |
| 1653242 | PR00 | Immunosuppressants | SECUKINUMAB AUTO-INJECTOR [COSENTYX] |
| 1599796 | PR00 | Immunosuppressants | COSENTYX INJECTABLE PRODUCT |
| 1599792 | PR00 | Immunosuppressants | 1 ML SECUKINUMAB 150 MG/ML PREFILLED SYRINGE |
| 1599798 | PR00 | Immunosuppressants | SECUKINUMAB 150 MG/ML PREFILLED SYRINGE |
| 1653241 | PR00 | Immunosuppressants | 1 ML SECUKINUMAB 150 MG/ML AUTO-INJECTOR |
| 1653244 | PR00 | Immunosuppressants | SECUKINUMAB 150 MG/ML AUTO-INJECTOR |
| 1599789 | PR00 | Immunosuppressants | SECUKINUMAB 150 MG/ML |
| 1599791 | PR00 | Immunosuppressants | SECUKINUMAB PREFILLED SYRINGE |
| 1653240 | PR00 | Immunosuppressants | SECUKINUMAB AUTO-INJECTOR |
| 1599790 | PR00 | Immunosuppressants | SECUKINUMAB INJECTABLE PRODUCT |
| 895761 | PR00 | Immunosuppressants | ACTEMRA |
| 612865 | PR00 | Immunosuppressants | TOCILIZUMAB |
| 1441530 | PR00 | Immunosuppressants | 0.9 ML TOCILIZUMAB 180 MG/ML PREFILLED SYRINGE [ACTEMRA] |
| 1441532 | PR00 | Immunosuppressants | TOCILIZUMAB 180 MG/ML PREFILLED SYRINGE [ACTEMRA] |
| 1657976 | PR00 | Immunosuppressants | 4 ML TOCILIZUMAB 20 MG/ML INJECTION [ACTEMRA] |
| 1657978 | PR00 | Immunosuppressants | TOCILIZUMAB 20 MG/ML INJECTION [ACTEMRA] |
| 1657980 | PR00 | Immunosuppressants | 10 ML TOCILIZUMAB 20 MG/ML INJECTION [ACTEMRA] |
| 1657982 | PR00 | Immunosuppressants | 20 ML TOCILIZUMAB 20 MG/ML INJECTION [ACTEMRA] |
| 2106075 | PR00 | Immunosuppressants | 0.9 ML TOCILIZUMAB 180 MG/ML AUTO-INJECTOR [ACTEMRA] |
| 2106077 | PR00 | Immunosuppressants | TOCILIZUMAB 180 MG/ML AUTO-INJECTOR [ACTEMRA] |
| 895764 | PR00 | Immunosuppressants | TOCILIZUMAB 20 MG/ML INJECTABLE SOLUTION [ACTEMRA] |
| 1441528 | PR00 | Immunosuppressants | TOCILIZUMAB 180 MG/ML [ACTEMRA] |
| 895762 | PR00 | Immunosuppressants | TOCILIZUMAB 20 MG/ML [ACTEMRA] |

| | | | |
|---|---|---|---|
| 1441529 | PR00 | Immunosuppressants | TOCILIZUMAB PREFILLED SYRINGE [ACTEMRA] |
| 1657975 | PR00 | Immunosuppressants | TOCILIZUMAB INJECTION [ACTEMRA] |
| 2106074 | PR00 | Immunosuppressants | TOCILIZUMAB AUTO-INJECTOR [ACTEMRA] |
| 895763 | PR00 | Immunosuppressants | TOCILIZUMAB INJECTABLE SOLUTION [ACTEMRA] |
| 1169167 | PR00 | Immunosuppressants | ACTEMRA INJECTABLE PRODUCT |
| 1441527 | PR00 | Immunosuppressants | 0.9 ML TOCILIZUMAB 180 MG/ML PREFILLED SYRINGE |
| 1441531 | PR00 | Immunosuppressants | TOCILIZUMAB 180 MG/ML PREFILLED SYRINGE |
| 1657974 | PR00 | Immunosuppressants | 4 ML TOCILIZUMAB 20 MG/ML INJECTION |
| 1657977 | PR00 | Immunosuppressants | TOCILIZUMAB 20 MG/ML INJECTION |
| 1657979 | PR00 | Immunosuppressants | 10 ML TOCILIZUMAB 20 MG/ML INJECTION |
| 1657981 | PR00 | Immunosuppressants | 20 ML TOCILIZUMAB 20 MG/ML INJECTION |
| 2106073 | PR00 | Immunosuppressants | 0.9 ML TOCILIZUMAB 180 MG/ML AUTO-INJECTOR |
| 2106076 | PR00 | Immunosuppressants | TOCILIZUMAB 180 MG/ML AUTO-INJECTOR |
| 895760 | PR00 | Immunosuppressants | TOCILIZUMAB 20 MG/ML INJECTABLE SOLUTION |
| 1441525 | PR00 | Immunosuppressants | TOCILIZUMAB 180 MG/ML |
| 895758 | PR00 | Immunosuppressants | TOCILIZUMAB 20 MG/ML |
| 1441526 | PR00 | Immunosuppressants | TOCILIZUMAB PREFILLED SYRINGE |
| 1657973 | PR00 | Immunosuppressants | TOCILIZUMAB INJECTION |
| 2106072 | PR00 | Immunosuppressants | TOCILIZUMAB AUTO-INJECTOR |
| 895759 | PR00 | Immunosuppressants | TOCILIZUMAB INJECTABLE SOLUTION |
| 1162729 | PR00 | Immunosuppressants | TOCILIZUMAB INJECTABLE PRODUCT |
| 1357542 | PR00 | Immunosuppressants | XELJANZ |
| 1357536 | PR00 | Immunosuppressants | TOFACITINIB |
| 1358492 | PR00 | Immunosuppressants | TOFACITINIB CITRATE |
| 1357547 | PR00 | Immunosuppressants | TOFACITINIB 5 MG ORAL TABLET [XELJANZ] |
| 1741049 | PR00 | Immunosuppressants | 24 HR TOFACITINIB 11 MG EXTENDED RELEASE ORAL TABLET [XELJANZ] |
| 1741051 | PR00 | Immunosuppressants | TOFACITINIB 11 MG EXTENDED RELEASE ORAL TABLET [XELJANZ] |
| 2048568 | PR00 | Immunosuppressants | TOFACITINIB 10 MG ORAL TABLET [XELJANZ] |
| 2273115 | PR00 | Immunosuppressants | 24 HR TOFACITINIB 22 MG EXTENDED RELEASE ORAL TABLET [XELJANZ] |
| 2273117 | PR00 | Immunosuppressants | TOFACITINIB 22 MG EXTENDED RELEASE ORAL TABLET [XELJANZ] |
| 1357543 | PR00 | Immunosuppressants | TOFACITINIB 5 MG [XELJANZ] |
| 1741047 | PR00 | Immunosuppressants | TOFACITINIB 11 MG [XELJANZ] |
| 2048567 | PR00 | Immunosuppressants | TOFACITINIB 10 MG [XELJANZ] |
| 2273114 | PR00 | Immunosuppressants | TOFACITINIB 22 MG [XELJANZ] |
| 1357544 | PR00 | Immunosuppressants | TOFACITINIB ORAL TABLET [XELJANZ] |
| 1741048 | PR00 | Immunosuppressants | TOFACITINIB EXTENDED RELEASE ORAL TABLET [XELJANZ] |
| 1357545 | PR00 | Immunosuppressants | XELJANZ ORAL PRODUCT |
| 1357546 | PR00 | Immunosuppressants | XELJANZ PILL |
| 1357541 | PR00 | Immunosuppressants | TOFACITINIB 5 MG ORAL TABLET |
| 1741046 | PR00 | Immunosuppressants | 24 HR TOFACITINIB 11 MG EXTENDED RELEASE ORAL TABLET |
| 1741050 | PR00 | Immunosuppressants | TOFACITINIB 11 MG EXTENDED RELEASE ORAL TABLET |
| 2048566 | PR00 | Immunosuppressants | TOFACITINIB 10 MG ORAL TABLET |
| 2273113 | PR00 | Immunosuppressants | 24 HR TOFACITINIB 22 MG EXTENDED RELEASE ORAL TABLET |
| 2273116 | PR00 | Immunosuppressants | TOFACITINIB 22 MG EXTENDED RELEASE ORAL TABLET |
| 1357537 | PR00 | Immunosuppressants | TOFACITINIB 5 MG |
| 1741044 | PR00 | Immunosuppressants | TOFACITINIB 11 MG |
| 2048565 | PR00 | Immunosuppressants | TOFACITINIB 10 MG |
| 2273112 | PR00 | Immunosuppressants | TOFACITINIB 22 MG |
| 1357540 | PR00 | Immunosuppressants | TOFACITINIB ORAL TABLET |
| 1741045 | PR00 | Immunosuppressants | TOFACITINIB EXTENDED RELEASE ORAL TABLET |
| 1357538 | PR00 | Immunosuppressants | TOFACITINIB ORAL PRODUCT |
| 1357539 | PR00 | Immunosuppressants | TOFACITINIB PILL |
| 151397 | PR00 | Immunosuppressants | AZAMUNE |
| 151888 | PR00 | Immunosuppressants | IMMUNOPRIN |
| 152156 | PR00 | Immunosuppressants | OPRISINE |
| 202559 | PR00 | Immunosuppressants | IMURAN |
| 360562 | PR00 | Immunosuppressants | BERKAPRINE |
| 404872 | PR00 | Immunosuppressants | AZASAN |
| 1256 | PR00 | Immunosuppressants | AZATHIOPRINE |
| 267476 | PR00 | Immunosuppressants | AZATHIOPRINE SODIUM |
| 105609 | PR00 | Immunosuppressants | AZATHIOPRINE 50 MG ORAL TABLET [AZAMUNE] |
| 105610 | PR00 | Immunosuppressants | AZATHIOPRINE 25 MG ORAL TABLET [IMURAN] |
| 105611 | PR00 | Immunosuppressants | AZATHIOPRINE 50 MG ORAL TABLET [IMURAN] |
| 105612 | PR00 | Immunosuppressants | AZATHIOPRINE 50 MG ORAL TABLET [IMMUNOPRIN] |
| 105613 | PR00 | Immunosuppressants | AZATHIOPRINE 50 MG ORAL TABLET [BERKAPRINE] |
| 108922 | PR00 | Immunosuppressants | AZATHIOPRINE 50 MG ORAL TABLET [OPRISINE] |
| 153130 | PR00 | Immunosuppressants | AZATHIOPRINE 10 MG ORAL TABLET [IMURAN] |
| 213563 | PR00 | Immunosuppressants | AZATHIOPRINE 10 MG/ML INJECTABLE SOLUTION [IMURAN] |
| 404351 | PR00 | Immunosuppressants | AZATHIOPRINE 50 MG ORAL TABLET [AZASAN] |
| 404475 | PR00 | Immunosuppressants | AZATHIOPRINE 75 MG ORAL TABLET [AZASAN] |
| 404476 | PR00 | Immunosuppressants | AZATHIOPRINE 100 MG ORAL TABLET [AZASAN] |

| | | | |
|---|---|---|---|
| 701338 | PR00 | Immunosuppressants | AZATHIOPRINE 25 MG ORAL TABLET [AZASAN] |
| 564118 | PR00 | Immunosuppressants | AZATHIOPRINE 50 MG [AZAMUNE] |
| 564119 | PR00 | Immunosuppressants | AZATHIOPRINE 25 MG [IMURAN] |
| 564120 | PR00 | Immunosuppressants | AZATHIOPRINE 50 MG [IMURAN] |
| 564121 | PR00 | Immunosuppressants | AZATHIOPRINE 50 MG [IMMUNOPRIN] |
| 564122 | PR00 | Immunosuppressants | AZATHIOPRINE 50 MG [BERKAPRINE] |
| 564655 | PR00 | Immunosuppressants | AZATHIOPRINE 50 MG [OPRISINE] |
| 565180 | PR00 | Immunosuppressants | AZATHIOPRINE 10 MG [IMURAN] |
| 573446 | PR00 | Immunosuppressants | AZATHIOPRINE 10 MG/ML [IMURAN] |
| 576332 | PR00 | Immunosuppressants | AZATHIOPRINE 50 MG [AZASAN] |
| 576426 | PR00 | Immunosuppressants | AZATHIOPRINE 75 MG [AZASAN] |
| 576427 | PR00 | Immunosuppressants | AZATHIOPRINE 100 MG [AZASAN] |
| 701337 | PR00 | Immunosuppressants | AZATHIOPRINE 25 MG [AZASAN] |
| 368214 | PR00 | Immunosuppressants | AZATHIOPRINE ORAL TABLET [IMURAN] |
| 368521 | PR00 | Immunosuppressants | AZATHIOPRINE ORAL TABLET [BERKAPRINE] |
| 369525 | PR00 | Immunosuppressants | AZATHIOPRINE ORAL TABLET [AZAMUNE] |
| 369534 | PR00 | Immunosuppressants | AZATHIOPRINE ORAL TABLET [IMMUNOPRIN] |
| 403109 | PR00 | Immunosuppressants | AZATHIOPRINE ORAL TABLET [OPRISINE] |
| 405819 | PR00 | Immunosuppressants | AZATHIOPRINE ORAL TABLET [AZASAN] |
| 92955 | PR00 | Immunosuppressants | AZATHIOPRINE INJECTABLE SOLUTION [IMURAN] |
| 1167775 | PR00 | Immunosuppressants | IMMUNOPRIN ORAL PRODUCT |
| 1167776 | PR00 | Immunosuppressants | IMMUNOPRIN PILL |
| 1172006 | PR00 | Immunosuppressants | IMURAN ORAL PRODUCT |
| 1172007 | PR00 | Immunosuppressants | IMURAN PILL |
| 1172211 | PR00 | Immunosuppressants | BERKAPRINE ORAL PRODUCT |
| 1172212 | PR00 | Immunosuppressants | BERKAPRINE PILL |
| 1176316 | PR00 | Immunosuppressants | AZAMUNE ORAL PRODUCT |
| 1176317 | PR00 | Immunosuppressants | AZAMUNE PILL |
| 1176318 | PR00 | Immunosuppressants | AZASAN ORAL PRODUCT |
| 1176319 | PR00 | Immunosuppressants | AZASAN PILL |
| 1182311 | PR00 | Immunosuppressants | OPRISINE ORAL PRODUCT |
| 1182312 | PR00 | Immunosuppressants | OPRISINE PILL |
| 153129 | PR00 | Immunosuppressants | AZATHIOPRINE 10 MG ORAL TABLET |
| 197388 | PR00 | Immunosuppressants | AZATHIOPRINE 50 MG ORAL TABLET |
| 199310 | PR00 | Immunosuppressants | AZATHIOPRINE 25 MG ORAL TABLET |
| 239983 | PR00 | Immunosuppressants | AZATHIOPRINE 100 MG INJECTION |
| 359228 | PR00 | Immunosuppressants | AZATHIOPRINE 100 MG ORAL TABLET |
| 359229 | PR00 | Immunosuppressants | AZATHIOPRINE 75 MG ORAL TABLET |
| 410148 | PR00 | Immunosuppressants | AZATHIOPRINE 250 MG ORAL CAPSULE |
| 315447 | PR00 | Immunosuppressants | AZATHIOPRINE 50 MG |
| 329313 | PR00 | Immunosuppressants | AZATHIOPRINE 25 MG |
| 333493 | PR00 | Immunosuppressants | AZATHIOPRINE 10 MG/ML |
| 360265 | PR00 | Immunosuppressants | AZATHIOPRINE 75 MG |
| 360266 | PR00 | Immunosuppressants | AZATHIOPRINE 100 MG |
| 385562 | PR00 | Immunosuppressants | AZATHIOPRINE 10 MG |
| 434684 | PR00 | Immunosuppressants | AZATHIOPRINE 250 MG |
| 1789839 | PR00 | Immunosuppressants | AZATHIOPRINE INJECTION |
| 370973 | PR00 | Immunosuppressants | AZATHIOPRINE ORAL TABLET |
| 375463 | PR00 | Immunosuppressants | AZATHIOPRINE INJECTABLE SOLUTION |
| 434685 | PR00 | Immunosuppressants | AZATHIOPRINE ORAL CAPSULE |
| 1155000 | PR00 | Immunosuppressants | AZATHIOPRINE INJECTABLE PRODUCT |
| 1155001 | PR00 | Immunosuppressants | AZATHIOPRINE ORAL PRODUCT |
| 1155002 | PR00 | Immunosuppressants | AZATHIOPRINE PILL |
| 219912 | PR00 | Immunosuppressants | SIMULECT |
| 196102 | PR00 | Immunosuppressants | BASILIXIMAB |
| 1656646 | PR00 | Immunosuppressants | BASILIXIMAB 10 MG INJECTION [SIMULECT] |
| 1656650 | PR00 | Immunosuppressants | BASILIXIMAB 20 MG INJECTION [SIMULECT] |
| 227146 | PR00 | Immunosuppressants | BASILIXIMAB 4 MG/ML INJECTABLE SOLUTION [SIMULECT] |
| 1656644 | PR00 | Immunosuppressants | BASILIXIMAB 10 MG [SIMULECT] |
| 1656649 | PR00 | Immunosuppressants | BASILIXIMAB 20 MG [SIMULECT] |
| 574069 | PR00 | Immunosuppressants | BASILIXIMAB 4 MG/ML [SIMULECT] |
| 1656645 | PR00 | Immunosuppressants | BASILIXIMAB INJECTION [SIMULECT] |
| 362559 | PR00 | Immunosuppressants | BASILIXIMAB INJECTABLE SOLUTION [SIMULECT] |
| 1182652 | PR00 | Immunosuppressants | SIMULECT INJECTABLE PRODUCT |
| 1656643 | PR00 | Immunosuppressants | BASILIXIMAB 10 MG INJECTION |
| 1656648 | PR00 | Immunosuppressants | BASILIXIMAB 20 MG INJECTION |
| 313873 | PR00 | Immunosuppressants | BASILIXIMAB 10 MG INJECTABLE SOLUTION |
| 359248 | PR00 | Immunosuppressants | BASILIXIMAB 10 MG INJECTABLE SOLUTION |
| 1656641 | PR00 | Immunosuppressants | BASILIXIMAB 10 MG |
| 1656647 | PR00 | Immunosuppressants | BASILIXIMAB 20 MG |
| 333494 | PR00 | Immunosuppressants | BASILIXIMAB 10 MG |

| | | | |
|---|---|---|---|
| 360267 | PR00 | Immunosuppressants | BASILIXIMAB 10 MG |
| 1656642 | PR00 | Immunosuppressants | BASILIXIMAB INJECTION |
| 377149 | PR00 | Immunosuppressants | BASILIXIMAB INJECTABLE SOLUTION |
| 1159396 | PR00 | Immunosuppressants | BASILIXIMAB INJECTABLE PRODUCT |
| 202816 | PR00 | Immunosuppressants | SANDIMMUNE |
| 202817 | PR00 | Immunosuppressants | NEORAL |
| 219800 | PR00 | Immunosuppressants | SANGCYA |
| 261530 | PR00 | Immunosuppressants | GENGRAF |
| 349643 | PR00 | Immunosuppressants | EON BRAND OF CYCLOSPORINE |
| 349644 | PR00 | Immunosuppressants | SIDMAK BRAND OF CYCLOSPORINE |
| 583287 | PR00 | Immunosuppressants | CYSLOSPORINE |
| 3008 | PR00 | Immunosuppressants | CYCLOSPORINE |
| 236077 | PR00 | Immunosuppressants | CYCLOSPORINE, MODIFIED |
| 108843 | PR00 | Immunosuppressants | CYCLOSPORINE 100 MG ORAL CAPSULE [NEORAL] |
| 108844 | PR00 | Immunosuppressants | CYCLOSPORINE 50 MG ORAL CAPSULE [NEORAL] |
| 108845 | PR00 | Immunosuppressants | CYCLOSPORINE 25 MG ORAL CAPSULE [NEORAL] |
| 1732369 | PR00 | Immunosuppressants | CYCLOSPORINE 50 MG/ML INJECTION [SANDIMMUNE] |
| 1797521 | PR00 | Immunosuppressants | CYCLOSPORINE, MODIFIED 50 MG ORAL CAPSULE [GENGRAF] |
| 197552 | PR00 | Immunosuppressants | CYCLOSPORINE 100 MG ORAL CAPSULE [SANDIMMUNE] |
| 212810 | PR00 | Immunosuppressants | CYCLOSPORINE 25 MG ORAL CAPSULE [SANDIMMUNE] |
| 212821 | PR00 | Immunosuppressants | CYCLOSPORINE 50 MG ORAL CAPSULE [SANDIMMUNE] |
| 212844 | PR00 | Immunosuppressants | 5 ML CYCLOSPORINE 50 MG/ML INJECTION [SANDIMMUNE] |
| 212956 | PR00 | Immunosuppressants | CYCLOSPORINE 100 MG/ML ORAL SOLUTION [NEORAL] |
| 212957 | PR00 | Immunosuppressants | CYCLOSPORINE 100 MG/ML ORAL SOLUTION [SANDIMMUNE] |
| 212958 | PR00 | Immunosuppressants | CYCLOSPORINE 100 MG/ML ORAL SOLUTION [SANGCYA] |
| 309627 | PR00 | Immunosuppressants | CYCLOSPORINE 100 MG ORAL CAPSULE [EON BRAND OF CYCLOSPORINE] |
| 309628 | PR00 | Immunosuppressants | CYCLOSPORINE 100 MG ORAL CAPSULE [GENGRAF] |
| 309629 | PR00 | Immunosuppressants | CYCLOSPORINE 25 MG ORAL CAPSULE [GENGRAF] |
| 309630 | PR00 | Immunosuppressants | CYCLOSPORINE 100 MG ORAL CAPSULE [SIDMAK BRAND OF CYCLOSPORINE] |
| 309631 | PR00 | Immunosuppressants | CYCLOSPORINE 25 MG ORAL CAPSULE [SIDMAK BRAND OF CYCLOSPORINE] |
| 313937 | PR00 | Immunosuppressants | CYCLOSPORINE 25 MG ORAL CAPSULE [EON BRAND OF CYCLOSPORINE] |
| 358934 | PR00 | Immunosuppressants | CYCLOSPORINE 100 MG/ML ORAL SOLUTION [GENGRAF] |
| 358935 | PR00 | Immunosuppressants | CYCLOSPORINE 100 MG/ML ORAL SOLUTION [SIDMAK BRAND OF CYCLOSPORINE] |
| 583290 | PR00 | Immunosuppressants | CYCLOSPORINE 100 MG ORAL CAPSULE [CYSLOSPORINE] |
| 835835 | PR00 | Immunosuppressants | CYCLOSPORINE, MODIFIED 100 MG ORAL CAPSULE [GENGRAF] |
| 835892 | PR00 | Immunosuppressants | CYCLOSPORINE, MODIFIED 100 MG/ML ORAL SOLUTION [GENGRAF] |
| 835896 | PR00 | Immunosuppressants | CYCLOSPORINE, MODIFIED 25 MG ORAL CAPSULE [GENGRAF] |
| 835899 | PR00 | Immunosuppressants | CYCLOSPORINE, MODIFIED 100 MG/ML ORAL SOLUTION [NEORAL] |
| 835909 | PR00 | Immunosuppressants | CYCLOSPORINE, MODIFIED 25 MG ORAL CAPSULE [NEORAL] |
| 835911 | PR00 | Immunosuppressants | CYCLOSPORINE, MODIFIED 100 MG ORAL CAPSULE [NEORAL] |
| 1797520 | PR00 | Immunosuppressants | CYCLOSPORINE, MODIFIED 50 MG [GENGRAF] |
| 486331 | PR00 | Immunosuppressants | CYCLOSPORINE 100 MG/ML [SANDIMMUNE] |
| 564618 | PR00 | Immunosuppressants | CYCLOSPORINE 100 MG [NEORAL] |
| 564619 | PR00 | Immunosuppressants | CYCLOSPORINE 50 MG [NEORAL] |
| 564620 | PR00 | Immunosuppressants | CYCLOSPORINE 25 MG [NEORAL] |
| 565430 | PR00 | Immunosuppressants | CYCLOSPORINE 100 MG [SANDIMMUNE] |
| 572889 | PR00 | Immunosuppressants | CYCLOSPORINE 25 MG [SANDIMMUNE] |
| 572895 | PR00 | Immunosuppressants | CYCLOSPORINE 50 MG [SANDIMMUNE] |
| 572915 | PR00 | Immunosuppressants | CYCLOSPORINE 50 MG/ML [SANDIMMUNE] |
| 572918 | PR00 | Immunosuppressants | CYCLOSPORINE 100 MG/ML [NEORAL] |
| 572919 | PR00 | Immunosuppressants | CYCLOSPORINE 100 MG/ML [SANGCYA] |
| 575103 | PR00 | Immunosuppressants | CYCLOSPORINE 100 MG [EON BRAND OF CYCLOSPORINE] |
| 575104 | PR00 | Immunosuppressants | CYCLOSPORINE 100 MG [GENGRAF] |
| 575105 | PR00 | Immunosuppressants | CYCLOSPORINE 25 MG [GENGRAF] |
| 575106 | PR00 | Immunosuppressants | CYCLOSPORINE 100 MG [SIDMAK BRAND OF CYCLOSPORINE] |
| 575107 | PR00 | Immunosuppressants | CYCLOSPORINE 25 MG [SIDMAK BRAND OF CYCLOSPORINE] |
| 575200 | PR00 | Immunosuppressants | CYCLOSPORINE 25 MG [EON BRAND OF CYCLOSPORINE] |
| 576050 | PR00 | Immunosuppressants | CYCLOSPORINE 100 MG/ML [GENGRAF] |
| 576051 | PR00 | Immunosuppressants | CYCLOSPORINE 100 MG/ML [SIDMAK BRAND OF CYCLOSPORINE] |
| 583288 | PR00 | Immunosuppressants | CYCLOSPORINE 100 MG [CYSLOSPORINE] |
| 835787 | PR00 | Immunosuppressants | CYCLOSPORINE, MODIFIED 100 MG [GENGRAF] |
| 835887 | PR00 | Immunosuppressants | CYCLOSPORINE, MODIFIED 100 MG/ML [GENGRAF] |
| 835895 | PR00 | Immunosuppressants | CYCLOSPORINE, MODIFIED 25 MG [GENGRAF] |
| 835897 | PR00 | Immunosuppressants | CYCLOSPORINE, MODIFIED 100 MG/ML [NEORAL] |
| 835900 | PR00 | Immunosuppressants | CYCLOSPORINE, MODIFIED 100 MG [NEORAL] |
| 835901 | PR00 | Immunosuppressants | CYCLOSPORINE, MODIFIED 25 MG [NEORAL] |
| 1732366 | PR00 | Immunosuppressants | CYCLOSPORINE INJECTION [SANDIMMUNE] |
| 363913 | PR00 | Immunosuppressants | CYCLOSPORINE ORAL SOLUTION [SIDMAK BRAND OF CYCLOSPORINE] |
| 363914 | PR00 | Immunosuppressants | CYCLOSPORINE ORAL SOLUTION [GENGRAF] |
| 363942 | PR00 | Immunosuppressants | CYCLOSPORINE ORAL SOLUTION [NEORAL] |
| 364418 | PR00 | Immunosuppressants | CYCLOSPORINE ORAL SOLUTION [SANGCYA] |

| | | | |
|---|---|---|---|
| 366172 | PR00 | Immunosuppressants | CYCLOSPORINE ORAL CAPSULE [SIDMAK BRAND OF CYCLOSPORINE] |
| 366173 | PR00 | Immunosuppressants | CYCLOSPORINE ORAL CAPSULE [GENGRAF] |
| 366174 | PR00 | Immunosuppressants | CYCLOSPORINE ORAL CAPSULE [EON BRAND OF CYCLOSPORINE] |
| 366479 | PR00 | Immunosuppressants | CYCLOSPORINE ORAL CAPSULE [NEORAL] |
| 366488 | PR00 | Immunosuppressants | CYCLOSPORINE ORAL CAPSULE [SANDIMMUNE] |
| 583289 | PR00 | Immunosuppressants | CYCLOSPORINE ORAL CAPSULE [CYSLOSPORINE] |
| 94034 | PR00 | Immunosuppressants | CYCLOSPORINE INJECTABLE SOLUTION [SANDIMMUNE] |
| 94035 | PR00 | Immunosuppressants | CYCLOSPORINE ORAL SOLUTION [SANDIMMUNE] |
| 1169824 | PR00 | Immunosuppressants | GENGRAF ORAL LIQUID PRODUCT |
| 1169825 | PR00 | Immunosuppressants | GENGRAF ORAL PRODUCT |
| 1169826 | PR00 | Immunosuppressants | GENGRAF PILL |
| 1174462 | PR00 | Immunosuppressants | CYSLOSPORINE ORAL PRODUCT |
| 1174463 | PR00 | Immunosuppressants | CYSLOSPORINE PILL |
| 1181672 | PR00 | Immunosuppressants | NEORAL ORAL LIQUID PRODUCT |
| 1181673 | PR00 | Immunosuppressants | NEORAL ORAL PRODUCT |
| 1181674 | PR00 | Immunosuppressants | NEORAL PILL |
| 1185408 | PR00 | Immunosuppressants | SANDIMMUNE INJECTABLE PRODUCT |
| 1185409 | PR00 | Immunosuppressants | SANDIMMUNE ORAL LIQUID PRODUCT |
| 1185410 | PR00 | Immunosuppressants | SANDIMMUNE ORAL PRODUCT |
| 1185411 | PR00 | Immunosuppressants | SANDIMMUNE PILL |
| 1185971 | PR00 | Immunosuppressants | SANGCYA ORAL LIQUID PRODUCT |
| 1185972 | PR00 | Immunosuppressants | SANGCYA ORAL PRODUCT |
| 1732368 | PR00 | Immunosuppressants | CYCLOSPORINE 50 MG/ML INJECTION |
| 197553 | PR00 | Immunosuppressants | CYCLOSPORINE 25 MG ORAL CAPSULE |
| 205168 | PR00 | Immunosuppressants | CYCLOSPORINE 50 MG ORAL CAPSULE |
| 205175 | PR00 | Immunosuppressants | 5 ML CYCLOSPORINE 50 MG/ML INJECTION |
| 226692 | PR00 | Immunosuppressants | CYCLOSPORINE 10 MG ORAL CAPSULE |
| 241834 | PR00 | Immunosuppressants | CYCLOSPORINE, MODIFIED 100 MG ORAL CAPSULE |
| 309632 | PR00 | Immunosuppressants | CYCLOSPORINE 100 MG/ML ORAL SOLUTION |
| 328160 | PR00 | Immunosuppressants | CYCLOSPORINE 100 MG ORAL CAPSULE |
| 348431 | PR00 | Immunosuppressants | CYCLOSPORINE 1000 MG/ML ORAL SOLUTION |
| 670177 | PR00 | Immunosuppressants | CYCLOSPORINE 50 MG/ML ORAL SOLUTION |
| 835886 | PR00 | Immunosuppressants | CYCLOSPORINE, MODIFIED 100 MG/ML ORAL SOLUTION |
| 835894 | PR00 | Immunosuppressants | CYCLOSPORINE, MODIFIED 25 MG ORAL CAPSULE |
| 835919 | PR00 | Immunosuppressants | CYCLOSPORINE, MODIFIED 10 MG ORAL CAPSULE |
| 835925 | PR00 | Immunosuppressants | CYCLOSPORINE, MODIFIED 50 MG ORAL CAPSULE |
| 315749 | PR00 | Immunosuppressants | CYCLOSPORINE 100 MG |
| 315750 | PR00 | Immunosuppressants | CYCLOSPORINE 100 MG/ML |
| 315751 | PR00 | Immunosuppressants | CYCLOSPORINE 25 MG |
| 315752 | PR00 | Immunosuppressants | CYCLOSPORINE 50 MG |
| 315753 | PR00 | Immunosuppressants | CYCLOSPORINE 50 MG/ML |
| 334294 | PR00 | Immunosuppressants | CYCLOSPORINE, MODIFIED 100 MG |
| 358763 | PR00 | Immunosuppressants | CYCLOSPORINE 1000 MG/ML |
| 385602 | PR00 | Immunosuppressants | CYCLOSPORINE 10 MG |
| 835885 | PR00 | Immunosuppressants | CYCLOSPORINE, MODIFIED 100 MG/ML |
| 835893 | PR00 | Immunosuppressants | CYCLOSPORINE, MODIFIED 25 MG |
| 835918 | PR00 | Immunosuppressants | CYCLOSPORINE, MODIFIED 10 MG |
| 835924 | PR00 | Immunosuppressants | CYCLOSPORINE, MODIFIED 50 MG |
| 1732364 | PR00 | Immunosuppressants | CYCLOSPORINE INJECTION |
| 371666 | PR00 | Immunosuppressants | CYCLOSPORINE ORAL CAPSULE |
| 371667 | PR00 | Immunosuppressants | CYCLOSPORINE ORAL SOLUTION |
| 374573 | PR00 | Immunosuppressants | CYCLOSPORINE INJECTABLE SOLUTION |
| 1156186 | PR00 | Immunosuppressants | CYCLOSPORINE INJECTABLE PRODUCT |
| 1156188 | PR00 | Immunosuppressants | CYCLOSPORINE ORAL LIQUID PRODUCT |
| 1156189 | PR00 | Immunosuppressants | CYCLOSPORINE ORAL PRODUCT |
| 1156190 | PR00 | Immunosuppressants | CYCLOSPORINE PILL |
| 1795335 | PR00 | Immunosuppressants | ZINBRYTA |
| 81770 | PR00 | Immunosuppressants | ZENAPAX |
| 190353 | PR00 | Immunosuppressants | DACLIZUMAB |
| 1795339 | PR00 | Immunosuppressants | 1 ML DACLIZUMAB 150 MG/ML PREFILLED SYRINGE [ZINBRYTA] |
| 1795341 | PR00 | Immunosuppressants | DACLIZUMAB 150 MG/ML PREFILLED SYRINGE [ZINBRYTA] |
| 1925211 | PR00 | Immunosuppressants | 1 ML DACLIZUMAB 150 MG/ML AUTO-INJECTOR [ZINBRYTA] |
| 1925213 | PR00 | Immunosuppressants | DACLIZUMAB 150 MG/ML AUTO-INJECTOR [ZINBRYTA] |
| 213177 | PR00 | Immunosuppressants | DACLIZUMAB 5 MG/ML INJECTABLE SOLUTION [ZENAPAX] |
| 1795336 | PR00 | Immunosuppressants | DACLIZUMAB 150 MG/ML [ZINBRYTA] |
| 573100 | PR00 | Immunosuppressants | DACLIZUMAB 5 MG/ML [ZENAPAX] |
| 1795337 | PR00 | Immunosuppressants | DACLIZUMAB PREFILLED SYRINGE [ZINBRYTA] |
| 1925210 | PR00 | Immunosuppressants | DACLIZUMAB AUTO-INJECTOR [ZINBRYTA] |
| 362855 | PR00 | Immunosuppressants | DACLIZUMAB INJECTABLE SOLUTION [ZENAPAX] |
| 1187246 | PR00 | Immunosuppressants | ZENAPAX INJECTABLE PRODUCT |
| 1795338 | PR00 | Immunosuppressants | ZINBRYTA INJECTABLE PRODUCT |

| | | | |
|---|---|---|---|
| 1795334 | PR00 | Immunosuppressants | 1 ML DACLIZUMAB 150 MG/ML PREFILLED SYRINGE |
| 1795340 | PR00 | Immunosuppressants | DACLIZUMAB 150 MG/ML PREFILLED SYRINGE |
| 1925209 | PR00 | Immunosuppressants | 1 ML DACLIZUMAB 150 MG/ML AUTO-INJECTOR |
| 1925212 | PR00 | Immunosuppressants | DACLIZUMAB 150 MG/ML AUTO-INJECTOR |
| 200171 | PR00 | Immunosuppressants | DACLIZUMAB 5 MG/ML INJECTABLE SOLUTION |
| 1795332 | PR00 | Immunosuppressants | DACLIZUMAB 150 MG/ML |
| 315755 | PR00 | Immunosuppressants | DACLIZUMAB 5 MG/ML |
| 1795333 | PR00 | Immunosuppressants | DACLIZUMAB PREFILLED SYRINGE |
| 1925208 | PR00 | Immunosuppressants | DACLIZUMAB AUTO-INJECTOR |
| 377159 | PR00 | Immunosuppressants | DACLIZUMAB INJECTABLE SOLUTION |
| 1151834 | PR00 | Immunosuppressants | DACLIZUMAB INJECTABLE PRODUCT |
| 2104612 | PR00 | Immunosuppressants | GAMIFANT |
| 2104604 | PR00 | Immunosuppressants | EMAPALUMAB |
| 2104603 | PR00 | Immunosuppressants | EMAPALUMAB-LZSG |
| 2104616 | PR00 | Immunosuppressants | 2 ML EMAPALUMAB-LZSG 5 MG/ML INJECTION [GAMIFANT] |
| 2104618 | PR00 | Immunosuppressants | EMAPALUMAB-LZSG 5 MG/ML INJECTION [GAMIFANT] |
| 2104622 | PR00 | Immunosuppressants | 10 ML EMAPALUMAB-LZSG 5 MG/ML INJECTION [GAMIFANT] |
| 2104613 | PR00 | Immunosuppressants | EMAPALUMAB-LZSG 5 MG/ML [GAMIFANT] |
| 2104614 | PR00 | Immunosuppressants | EMAPALUMAB INJECTION [GAMIFANT] |
| 2104615 | PR00 | Immunosuppressants | GAMIFANT INJECTABLE PRODUCT |
| 2104611 | PR00 | Immunosuppressants | 2 ML EMAPALUMAB-LZSG 5 MG/ML INJECTION |
| 2104617 | PR00 | Immunosuppressants | EMAPALUMAB-LZSG 5 MG/ML INJECTION |
| 2104621 | PR00 | Immunosuppressants | 10 ML EMAPALUMAB-LZSG 5 MG/ML INJECTION |
| 2104608 | PR00 | Immunosuppressants | EMAPALUMAB-LZSG 5 MG/ML |
| 2104610 | PR00 | Immunosuppressants | EMAPALUMAB INJECTION |
| 2104609 | PR00 | Immunosuppressants | EMAPALUMAB INJECTABLE PRODUCT |
| 845509 | PR00 | Immunosuppressants | AFINITOR |
| 977428 | PR00 | Immunosuppressants | ZORTRESS |
| 845508 | PR00 | Immunosuppressants | {28 (EVEROLIMUS 10 MG ORAL TABLET) } PACK |
| 845516 | PR00 | Immunosuppressants | {28 (EVEROLIMUS 5 MG ORAL TABLET) } PACK |
| 141704 | PR00 | Immunosuppressants | EVEROLIMUS |
| 1119402 | PR00 | Immunosuppressants | EVEROLIMUS 7.5 MG ORAL TABLET [AFINITOR] |
| 1310138 | PR00 | Immunosuppressants | EVEROLIMUS 2 MG TABLET FOR ORAL SUSPENSION [AFINITOR] |
| 1310144 | PR00 | Immunosuppressants | EVEROLIMUS 3 MG TABLET FOR ORAL SUSPENSION [AFINITOR] |
| 1310147 | PR00 | Immunosuppressants | EVEROLIMUS 5 MG TABLET FOR ORAL SUSPENSION [AFINITOR] |
| 2056897 | PR00 | Immunosuppressants | EVEROLIMUS 1 MG ORAL TABLET [ZORTRESS] |
| 845512 | PR00 | Immunosuppressants | EVEROLIMUS 10 MG ORAL TABLET [AFINITOR] |
| 845518 | PR00 | Immunosuppressants | EVEROLIMUS 5 MG ORAL TABLET [AFINITOR] |
| 977431 | PR00 | Immunosuppressants | EVEROLIMUS 0.25 MG ORAL TABLET [ZORTRESS] |
| 977436 | PR00 | Immunosuppressants | EVEROLIMUS 0.5 MG ORAL TABLET [ZORTRESS] |
| 977440 | PR00 | Immunosuppressants | EVEROLIMUS 0.75 MG ORAL TABLET [ZORTRESS] |
| 998191 | PR00 | Immunosuppressants | EVEROLIMUS 2.5 MG ORAL TABLET [AFINITOR] |
| 1119401 | PR00 | Immunosuppressants | EVEROLIMUS 7.5 MG [AFINITOR] |
| 1310135 | PR00 | Immunosuppressants | EVEROLIMUS 0.08 MG/ML [AFINITOR] |
| 1310143 | PR00 | Immunosuppressants | EVEROLIMUS 0.12 MG/ML [AFINITOR] |
| 1310146 | PR00 | Immunosuppressants | EVEROLIMUS 0.2 MG/ML [AFINITOR] |
| 1869515 | PR00 | Immunosuppressants | EVEROLIMUS 2 MG [AFINITOR] |
| 1869520 | PR00 | Immunosuppressants | EVEROLIMUS 3 MG [AFINITOR] |
| 2056896 | PR00 | Immunosuppressants | EVEROLIMUS 1 MG [ZORTRESS] |
| 845510 | PR00 | Immunosuppressants | EVEROLIMUS 10 MG [AFINITOR] |
| 845517 | PR00 | Immunosuppressants | EVEROLIMUS 5 MG [AFINITOR] |
| 977429 | PR00 | Immunosuppressants | EVEROLIMUS 0.25 MG [ZORTRESS] |
| 977435 | PR00 | Immunosuppressants | EVEROLIMUS 0.5 MG [ZORTRESS] |
| 977439 | PR00 | Immunosuppressants | EVEROLIMUS 0.75 MG [ZORTRESS] |
| 998190 | PR00 | Immunosuppressants | EVEROLIMUS 2.5 MG [AFINITOR] |
| 1310136 | PR00 | Immunosuppressants | EVEROLIMUS ORAL SUSPENSION [AFINITOR] |
| 1869516 | PR00 | Immunosuppressants | EVEROLIMUS TABLET FOR ORAL SUSPENSION [AFINITOR] |
| 845511 | PR00 | Immunosuppressants | EVEROLIMUS ORAL TABLET [AFINITOR] |
| 977430 | PR00 | Immunosuppressants | EVEROLIMUS ORAL TABLET [ZORTRESS] |
| 1172066 | PR00 | Immunosuppressants | AFINITOR ORAL PRODUCT |
| 1172067 | PR00 | Immunosuppressants | AFINITOR PILL |
| 1186316 | PR00 | Immunosuppressants | ZORTRESS ORAL PRODUCT |
| 1186317 | PR00 | Immunosuppressants | ZORTRESS PILL |
| 1310137 | PR00 | Immunosuppressants | AFINITOR ORAL LIQUID PRODUCT |
| 1119400 | PR00 | Immunosuppressants | EVEROLIMUS 7.5 MG ORAL TABLET |
| 1308428 | PR00 | Immunosuppressants | EVEROLIMUS 2 MG TABLET FOR ORAL SUSPENSION |
| 1308430 | PR00 | Immunosuppressants | EVEROLIMUS 3 MG TABLET FOR ORAL SUSPENSION |
| 1308432 | PR00 | Immunosuppressants | EVEROLIMUS 5 MG TABLET FOR ORAL SUSPENSION |
| 2056895 | PR00 | Immunosuppressants | EVEROLIMUS 1 MG ORAL TABLET |
| 845507 | PR00 | Immunosuppressants | EVEROLIMUS 10 MG ORAL TABLET |
| 845515 | PR00 | Immunosuppressants | EVEROLIMUS 5 MG ORAL TABLET |

| | | | |
|---|---|---|---|
| 977427 | PR00 | Immunosuppressants | EVEROLIMUS 0.25 MG ORAL TABLET |
| 977434 | PR00 | Immunosuppressants | EVEROLIMUS 0.5 MG ORAL TABLET |
| 977438 | PR00 | Immunosuppressants | EVEROLIMUS 0.75 MG ORAL TABLET |
| 998189 | PR00 | Immunosuppressants | EVEROLIMUS 2.5 MG ORAL TABLET |
| 1119399 | PR00 | Immunosuppressants | EVEROLIMUS 7.5 MG |
| 1308425 | PR00 | Immunosuppressants | EVEROLIMUS 0.08 MG/ML |
| 1308429 | PR00 | Immunosuppressants | EVEROLIMUS 0.12 MG/ML |
| 1308431 | PR00 | Immunosuppressants | EVEROLIMUS 0.2 MG/ML |
| 1869512 | PR00 | Immunosuppressants | EVEROLIMUS 2 MG |
| 1869518 | PR00 | Immunosuppressants | EVEROLIMUS 3 MG |
| 2056894 | PR00 | Immunosuppressants | EVEROLIMUS 1 MG |
| 845505 | PR00 | Immunosuppressants | EVEROLIMUS 10 MG |
| 845514 | PR00 | Immunosuppressants | EVEROLIMUS 5 MG |
| 977426 | PR00 | Immunosuppressants | EVEROLIMUS 0.25 MG |
| 977433 | PR00 | Immunosuppressants | EVEROLIMUS 0.5 MG |
| 977437 | PR00 | Immunosuppressants | EVEROLIMUS 0.75 MG |
| 998188 | PR00 | Immunosuppressants | EVEROLIMUS 2.5 MG |
| 1308427 | PR00 | Immunosuppressants | EVEROLIMUS ORAL SUSPENSION |
| 1869513 | PR00 | Immunosuppressants | EVEROLIMUS TABLET FOR ORAL SUSPENSION |
| 845506 | PR00 | Immunosuppressants | EVEROLIMUS ORAL TABLET |
| 1163786 | PR00 | Immunosuppressants | EVEROLIMUS ORAL PRODUCT |
| 1163787 | PR00 | Immunosuppressants | EVEROLIMUS PILL |
| 1308426 | PR00 | Immunosuppressants | EVEROLIMUS ORAL LIQUID PRODUCT |
| 1536485 | PR00 | Immunosuppressants | PURIXAN |
| 219513 | PR00 | Immunosuppressants | PURINETHOL |
| 103 | PR00 | Immunosuppressants | MERCAPTOPURINE |
| 1546028 | PR00 | Immunosuppressants | MERCAPTOPURINE ANHYDROUS |
| 1536490 | PR00 | Immunosuppressants | MERCAPTOPURINE 20 MG/ML ORAL SUSPENSION [PURIXAN] |
| 206788 | PR00 | Immunosuppressants | MERCAPTOPURINE 50 MG ORAL TABLET [PURINETHOL] |
| 1536486 | PR00 | Immunosuppressants | MERCAPTOPURINE 20 MG/ML [PURIXAN] |
| 567599 | PR00 | Immunosuppressants | MERCAPTOPURINE 50 MG [PURINETHOL] |
| 1536487 | PR00 | Immunosuppressants | MERCAPTOPURINE ORAL SUSPENSION [PURIXAN] |
| 368861 | PR00 | Immunosuppressants | MERCAPTOPURINE ORAL TABLET [PURINETHOL] |
| 1183583 | PR00 | Immunosuppressants | PURINETHOL ORAL PRODUCT |
| 1183584 | PR00 | Immunosuppressants | PURINETHOL PILL |
| 1536488 | PR00 | Immunosuppressants | PURIXAN ORAL LIQUID PRODUCT |
| 1536489 | PR00 | Immunosuppressants | PURIXAN ORAL PRODUCT |
| 1536484 | PR00 | Immunosuppressants | MERCAPTOPURINE 20 MG/ML ORAL SUSPENSION |
| 197931 | PR00 | Immunosuppressants | MERCAPTOPURINE 50 MG ORAL TABLET |
| 199766 | PR00 | Immunosuppressants | MERCAPTOPURINE 10 MG ORAL TABLET |
| 1536481 | PR00 | Immunosuppressants | MERCAPTOPURINE 20 MG/ML |
| 316246 | PR00 | Immunosuppressants | MERCAPTOPURINE 50 MG |
| 332204 | PR00 | Immunosuppressants | MERCAPTOPURINE 10 MG |
| 1536483 | PR00 | Immunosuppressants | MERCAPTOPURINE ORAL SUSPENSION |
| 372782 | PR00 | Immunosuppressants | MERCAPTOPURINE ORAL TABLET |
| 1165710 | PR00 | Immunosuppressants | MERCAPTOPURINE ORAL PRODUCT |
| 1165711 | PR00 | Immunosuppressants | MERCAPTOPURINE PILL |
| 1536482 | PR00 | Immunosuppressants | MERCAPTOPURINE ORAL LIQUID PRODUCT |
| 1441403 | PR00 | Immunosuppressants | OTREXUP |
| 152008 | PR00 | Immunosuppressants | MAXTREX |
| 1544379 | PR00 | Immunosuppressants | RASUVO |
| 1921593 | PR00 | Immunosuppressants | XATMEP |
| 284900 | PR00 | Immunosuppressants | TREXALL |
| 346424 | PR00 | Immunosuppressants | EMTEXATE |
| 491604 | PR00 | Immunosuppressants | RHEUMATREX |
| 1150118 | PR00 | Immunosuppressants | {8 (METHOTREXATE 2.5 MG ORAL TABLET [RHEUMATREX]) } PACK [RHEUMATREX DOSE PACK 8] |
| 1541216 | PR00 | Immunosuppressants | {12 (METHOTREXATE 2.5 MG ORAL TABLET [RHEUMATREX]) } PACK [RHEUMATREX DOSE PACK 12] |
| 1541218 | PR00 | Immunosuppressants | {16 (METHOTREXATE 2.5 MG ORAL TABLET [RHEUMATREX]) } PACK [RHEUMATREX DOSE PACK 16] |
| 1541221 | PR00 | Immunosuppressants | {20 (METHOTREXATE 2.5 MG ORAL TABLET [RHEUMATREX]) } PACK [RHEUMATREX DOSE PACK 20] |
| 1541224 | PR00 | Immunosuppressants | {24 (METHOTREXATE 2.5 MG ORAL TABLET [RHEUMATREX]) } PACK [RHEUMATREX DOSE PACK 24] |
| 1541226 | PR00 | Immunosuppressants | {28 (METHOTREXATE 2.5 MG ORAL TABLET [RHEUMATREX]) } PACK [RHEUMATREX DOSE PACK 28] |
| 1541228 | PR00 | Immunosuppressants | {32 (METHOTREXATE 2.5 MG ORAL TABLET [RHEUMATREX]) } PACK [RHEUMATREX DOSE PACK 32] |
| 1150117 | PR00 | Immunosuppressants | {8 (METHOTREXATE 2.5 MG ORAL TABLET) } PACK |
| 1541215 | PR00 | Immunosuppressants | {12 (METHOTREXATE 2.5 MG ORAL TABLET) } PACK |

| | | | |
|---|---|---|---|
| 1541217 | PR00 | Immunosuppressants | {16 (METHOTREXATE 2.5 MG ORAL TABLET) } PACK |
| 1541220 | PR00 | Immunosuppressants | {20 (METHOTREXATE 2.5 MG ORAL TABLET) } PACK |
| 1541222 | PR00 | Immunosuppressants | {24 (METHOTREXATE 2.5 MG ORAL TABLET) } PACK |
| 1541225 | PR00 | Immunosuppressants | {28 (METHOTREXATE 2.5 MG ORAL TABLET) } PACK |
| 1541227 | PR00 | Immunosuppressants | {32 (METHOTREXATE 2.5 MG ORAL TABLET) } PACK |
| 6851 | PR00 | Immunosuppressants | METHOTREXATE |
| 287734 | PR00 | Immunosuppressants | METHOTREXATE SODIUM |
| 102953 | PR00 | Immunosuppressants | METHOTREXATE 10 MG ORAL TABLET [EMTEXATE] |
| 102954 | PR00 | Immunosuppressants | METHOTREXATE 2.5 MG/ML INJECTABLE SOLUTION [EMTEXATE] |
| 102955 | PR00 | Immunosuppressants | METHOTREXATE 25 MG/ML INJECTABLE SOLUTION [EMTEXATE] |
| 102956 | PR00 | Immunosuppressants | METHOTREXATE 100 MG/ML INJECTABLE SOLUTION [EMTEXATE] |
| 102957 | PR00 | Immunosuppressants | METHOTREXATE 50 MG/ML INJECTABLE SOLUTION [EMTEXATE] |
| 102958 | PR00 | Immunosuppressants | METHOTREXATE 2.5 MG/ML INJECTABLE SOLUTION [MAXTREX] |
| 102959 | PR00 | Immunosuppressants | METHOTREXATE 25 MG/ML INJECTABLE SOLUTION [MAXTREX] |
| 105587 | PR00 | Immunosuppressants | METHOTREXATE 2.5 MG ORAL TABLET [MAXTREX] |
| 105588 | PR00 | Immunosuppressants | METHOTREXATE 10 MG ORAL TABLET [MAXTREX] |
| 1441407 | PR00 | Immunosuppressants | 0.4 ML METHOTREXATE 25 MG/ML AUTO-INJECTOR [OTREXUP] |
| 1441409 | PR00 | Immunosuppressants | METHOTREXATE 25 MG/ML AUTO-INJECTOR [OTREXUP] |
| 1441413 | PR00 | Immunosuppressants | 0.4 ML METHOTREXATE 37.5 MG/ML AUTO-INJECTOR [OTREXUP] |
| 1441415 | PR00 | Immunosuppressants | METHOTREXATE 37.5 MG/ML AUTO-INJECTOR [OTREXUP] |
| 1441418 | PR00 | Immunosuppressants | 0.4 ML METHOTREXATE 50 MG/ML AUTO-INJECTOR [OTREXUP] |
| 1441420 | PR00 | Immunosuppressants | METHOTREXATE 50 MG/ML AUTO-INJECTOR [OTREXUP] |
| 1441424 | PR00 | Immunosuppressants | 0.4 ML METHOTREXATE 62.5 MG/ML AUTO-INJECTOR [OTREXUP] |
| 1441426 | PR00 | Immunosuppressants | METHOTREXATE 62.5 MG/ML AUTO-INJECTOR [OTREXUP] |
| 1544383 | PR00 | Immunosuppressants | 0.2 ML METHOTREXATE 50 MG/ML AUTO-INJECTOR [RASUVO] |
| 1544384 | PR00 | Immunosuppressants | METHOTREXATE 50 MG/ML AUTO-INJECTOR [RASUVO] |
| 1544386 | PR00 | Immunosuppressants | 0.25 ML METHOTREXATE 50 MG/ML AUTO-INJECTOR [RASUVO] |
| 1544388 | PR00 | Immunosuppressants | 0.3 ML METHOTREXATE 50 MG/ML AUTO-INJECTOR [RASUVO] |
| 1544390 | PR00 | Immunosuppressants | 0.35 ML METHOTREXATE 50 MG/ML AUTO-INJECTOR [RASUVO] |
| 1544394 | PR00 | Immunosuppressants | 0.4 ML METHOTREXATE 50 MG/ML AUTO-INJECTOR [RASUVO] |
| 1544396 | PR00 | Immunosuppressants | 0.45 ML METHOTREXATE 50 MG/ML AUTO-INJECTOR [RASUVO] |
| 1544398 | PR00 | Immunosuppressants | 0.5 ML METHOTREXATE 50 MG/ML AUTO-INJECTOR [RASUVO] |
| 1544400 | PR00 | Immunosuppressants | 0.55 ML METHOTREXATE 50 MG/ML AUTO-INJECTOR [RASUVO] |
| 1544402 | PR00 | Immunosuppressants | 0.6 ML METHOTREXATE 50 MG/ML AUTO-INJECTOR [RASUVO] |
| 1544404 | PR00 | Immunosuppressants | 0.15 ML METHOTREXATE 50 MG/ML AUTO-INJECTOR [RASUVO] |
| 1594759 | PR00 | Immunosuppressants | 0.4 ML METHOTREXATE 18.8 MG/ML AUTO-INJECTOR [OTREXUP] |
| 1594761 | PR00 | Immunosuppressants | METHOTREXATE 18.8 MG/ML AUTO-INJECTOR [OTREXUP] |
| 1747181 | PR00 | Immunosuppressants | 0.4 ML METHOTREXATE 31.3 MG/ML AUTO-INJECTOR [OTREXUP] |
| 1747183 | PR00 | Immunosuppressants | METHOTREXATE 31.3 MG/ML AUTO-INJECTOR [OTREXUP] |
| 1747187 | PR00 | Immunosuppressants | 0.4 ML METHOTREXATE 43.8 MG/ML AUTO-INJECTOR [OTREXUP] |
| 1747189 | PR00 | Immunosuppressants | METHOTREXATE 43.8 MG/ML AUTO-INJECTOR [OTREXUP] |
| 1747194 | PR00 | Immunosuppressants | 0.4 ML METHOTREXATE 56.3 MG/ML AUTO-INJECTOR [OTREXUP] |
| 1747196 | PR00 | Immunosuppressants | METHOTREXATE 56.3 MG/ML AUTO-INJECTOR [OTREXUP] |
| 1921598 | PR00 | Immunosuppressants | METHOTREXATE 2.5 MG/ML ORAL SOLUTION [XATMEP] |
| 207033 | PR00 | Immunosuppressants | METHOTREXATE 25 MG/ML INJECTABLE SOLUTION [FOLEX PFS] |
| 284592 | PR00 | Immunosuppressants | METHOTREXATE 5 MG ORAL TABLET [TREXALL] |
| 284593 | PR00 | Immunosuppressants | METHOTREXATE 10 MG ORAL TABLET [TREXALL] |
| 284594 | PR00 | Immunosuppressants | METHOTREXATE 7.5 MG ORAL TABLET [TREXALL] |
| 284595 | PR00 | Immunosuppressants | METHOTREXATE 15 MG ORAL TABLET [TREXALL] |
| 579782 | PR00 | Immunosuppressants | METHOTREXATE 2.5 MG ORAL TABLET [RHEUMATREX] |
| 1441404 | PR00 | Immunosuppressants | METHOTREXATE 25 MG/ML [OTREXUP] |
| 1441412 | PR00 | Immunosuppressants | METHOTREXATE 37.5 MG/ML [OTREXUP] |
| 1441417 | PR00 | Immunosuppressants | METHOTREXATE 50 MG/ML [OTREXUP] |
| 1441423 | PR00 | Immunosuppressants | METHOTREXATE 62.5 MG/ML [OTREXUP] |
| 1544380 | PR00 | Immunosuppressants | METHOTREXATE 50 MG/ML [RASUVO] |
| 1594758 | PR00 | Immunosuppressants | METHOTREXATE 18.75 MG/ML [OTREXUP] |
| 1659721 | PR00 | Immunosuppressants | METHOTREXATE 18.8 MG/ML [OTREXUP] |
| 1747180 | PR00 | Immunosuppressants | METHOTREXATE 31.3 MG/ML [OTREXUP] |
| 1747186 | PR00 | Immunosuppressants | METHOTREXATE 43.8 MG/ML [OTREXUP] |
| 1747193 | PR00 | Immunosuppressants | METHOTREXATE 56.3 MG/ML [OTREXUP] |
| 1921594 | PR00 | Immunosuppressants | METHOTREXATE 2.5 MG/ML [XATMEP] |
| 563189 | PR00 | Immunosuppressants | METHOTREXATE 10 MG [EMTEXATE] |
| 563190 | PR00 | Immunosuppressants | METHOTREXATE 2.5 MG/ML [EMTEXATE] |
| 563191 | PR00 | Immunosuppressants | METHOTREXATE 25 MG/ML [EMTEXATE] |
| 563192 | PR00 | Immunosuppressants | METHOTREXATE 100 MG/ML [EMTEXATE] |
| 563193 | PR00 | Immunosuppressants | METHOTREXATE 50 MG/ML [EMTEXATE] |
| 563194 | PR00 | Immunosuppressants | METHOTREXATE 2.5 MG/ML [MAXTREX] |
| 563195 | PR00 | Immunosuppressants | METHOTREXATE 25 MG/ML [MAXTREX] |
| 564104 | PR00 | Immunosuppressants | METHOTREXATE 2.5 MG [MAXTREX] |
| 564105 | PR00 | Immunosuppressants | METHOTREXATE 10 MG [MAXTREX] |
| 567829 | PR00 | Immunosuppressants | METHOTREXATE 25 MG/ML [FOLEX PFS] |

| | | | |
|---|---|---|---|
| 575019 | PR00 | Immunosuppressants | METHOTREXATE 5 MG [TREXALL] |
| 575020 | PR00 | Immunosuppressants | METHOTREXATE 10 MG [TREXALL] |
| 575021 | PR00 | Immunosuppressants | METHOTREXATE 7.5 MG [TREXALL] |
| 575022 | PR00 | Immunosuppressants | METHOTREXATE 15 MG [TREXALL] |
| 579780 | PR00 | Immunosuppressants | METHOTREXATE 2.5 MG [RHEUMATREX] |
| 1441405 | PR00 | Immunosuppressants | METHOTREXATE PREFILLED SYRINGE [OTREXUP] |
| 1544381 | PR00 | Immunosuppressants | METHOTREXATE PREFILLED SYRINGE [RASUVO] |
| 1655205 | PR00 | Immunosuppressants | METHOTREXATE AUTO-INJECTOR [OTREXUP] |
| 1655252 | PR00 | Immunosuppressants | METHOTREXATE AUTO-INJECTOR [RASUVO] |
| 1921595 | PR00 | Immunosuppressants | METHOTREXATE ORAL SOLUTION [XATMEP] |
| 362547 | PR00 | Immunosuppressants | METHOTREXATE INJECTABLE SOLUTION [EMTEXATE] |
| 362736 | PR00 | Immunosuppressants | METHOTREXATE INJECTABLE SOLUTION [MAXTREX] |
| 363432 | PR00 | Immunosuppressants | METHOTREXATE INJECTABLE SOLUTION [FOLEX PFS] |
| 368138 | PR00 | Immunosuppressants | METHOTREXATE ORAL TABLET [TREXALL] |
| 368588 | PR00 | Immunosuppressants | METHOTREXATE ORAL TABLET [MAXTREX] |
| 368665 | PR00 | Immunosuppressants | METHOTREXATE ORAL TABLET [EMTEXATE] |
| 579781 | PR00 | Immunosuppressants | METHOTREXATE ORAL TABLET [RHEUMATREX] |
| 1175944 | PR00 | Immunosuppressants | EMTEXATE INJECTABLE PRODUCT |
| 1175945 | PR00 | Immunosuppressants | EMTEXATE ORAL PRODUCT |
| 1175946 | PR00 | Immunosuppressants | EMTEXATE PILL |
| 1179677 | PR00 | Immunosuppressants | TREXALL ORAL PRODUCT |
| 1179678 | PR00 | Immunosuppressants | TREXALL PILL |
| 1182738 | PR00 | Immunosuppressants | MAXTREX INJECTABLE PRODUCT |
| 1182739 | PR00 | Immunosuppressants | MAXTREX ORAL PRODUCT |
| 1182740 | PR00 | Immunosuppressants | MAXTREX PILL |
| 1183068 | PR00 | Immunosuppressants | RHEUMATREX ORAL PRODUCT |
| 1183069 | PR00 | Immunosuppressants | RHEUMATREX PILL |
| 1441406 | PR00 | Immunosuppressants | OTREXUP INJECTABLE PRODUCT |
| 1544382 | PR00 | Immunosuppressants | RASUVO INJECTABLE PRODUCT |
| 1921596 | PR00 | Immunosuppressants | XATMEP ORAL LIQUID PRODUCT |
| 1921597 | PR00 | Immunosuppressants | XATMEP ORAL PRODUCT |
| 105585 | PR00 | Immunosuppressants | METHOTREXATE 2.5 MG ORAL TABLET |
| 105586 | PR00 | Immunosuppressants | METHOTREXATE 10 MG ORAL TABLET |
| 105589 | PR00 | Immunosuppressants | METHOTREXATE 100 MG/ML INJECTABLE SOLUTION |
| 1441402 | PR00 | Immunosuppressants | 0.4 ML METHOTREXATE 25 MG/ML AUTO-INJECTOR |
| 1441408 | PR00 | Immunosuppressants | METHOTREXATE 25 MG/ML AUTO-INJECTOR |
| 1441411 | PR00 | Immunosuppressants | 0.4 ML METHOTREXATE 37.5 MG/ML AUTO-INJECTOR |
| 1441414 | PR00 | Immunosuppressants | METHOTREXATE 37.5 MG/ML AUTO-INJECTOR |
| 1441416 | PR00 | Immunosuppressants | 0.4 ML METHOTREXATE 50 MG/ML AUTO-INJECTOR |
| 1441419 | PR00 | Immunosuppressants | METHOTREXATE 50 MG/ML AUTO-INJECTOR |
| 1441422 | PR00 | Immunosuppressants | 0.4 ML METHOTREXATE 62.5 MG/ML AUTO-INJECTOR |
| 1441425 | PR00 | Immunosuppressants | METHOTREXATE 62.5 MG/ML AUTO-INJECTOR |
| 1544378 | PR00 | Immunosuppressants | 0.2 ML METHOTREXATE 50 MG/ML AUTO-INJECTOR |
| 1544385 | PR00 | Immunosuppressants | 0.25 ML METHOTREXATE 50 MG/ML AUTO-INJECTOR |
| 1544387 | PR00 | Immunosuppressants | 0.3 ML METHOTREXATE 50 MG/ML AUTO-INJECTOR |
| 1544389 | PR00 | Immunosuppressants | 0.35 ML METHOTREXATE 50 MG/ML AUTO-INJECTOR |
| 1544395 | PR00 | Immunosuppressants | 0.45 ML METHOTREXATE 50 MG/ML AUTO-INJECTOR |
| 1544397 | PR00 | Immunosuppressants | 0.5 ML METHOTREXATE 50 MG/ML AUTO-INJECTOR |
| 1544399 | PR00 | Immunosuppressants | 0.55 ML METHOTREXATE 50 MG/ML AUTO-INJECTOR |
| 1544401 | PR00 | Immunosuppressants | 0.6 ML METHOTREXATE 50 MG/ML AUTO-INJECTOR |
| 1544403 | PR00 | Immunosuppressants | 0.15 ML METHOTREXATE 50 MG/ML AUTO-INJECTOR |
| 1594757 | PR00 | Immunosuppressants | 0.4 ML METHOTREXATE 18.8 MG/ML AUTO-INJECTOR |
| 1594760 | PR00 | Immunosuppressants | METHOTREXATE 18.8 MG/ML AUTO-INJECTOR |
| 1655956 | PR00 | Immunosuppressants | 40 ML METHOTREXATE 25 MG/ML INJECTION |
| 1655957 | PR00 | Immunosuppressants | METHOTREXATE 25 MG/ML INJECTION |
| 1655959 | PR00 | Immunosuppressants | 10 ML METHOTREXATE 25 MG/ML INJECTION |
| 1655960 | PR00 | Immunosuppressants | 2 ML METHOTREXATE 25 MG/ML INJECTION |
| 1655967 | PR00 | Immunosuppressants | 4 ML METHOTREXATE 25 MG/ML INJECTION |
| 1655968 | PR00 | Immunosuppressants | 8 ML METHOTREXATE 25 MG/ML INJECTION |
| 1747179 | PR00 | Immunosuppressants | 0.4 ML METHOTREXATE 31.3 MG/ML AUTO-INJECTOR |
| 1747182 | PR00 | Immunosuppressants | METHOTREXATE 31.3 MG/ML AUTO-INJECTOR |
| 1747185 | PR00 | Immunosuppressants | 0.4 ML METHOTREXATE 43.8 MG/ML AUTO-INJECTOR |
| 1747188 | PR00 | Immunosuppressants | METHOTREXATE 43.8 MG/ML AUTO-INJECTOR |
| 1747192 | PR00 | Immunosuppressants | 0.4 ML METHOTREXATE 56.3 MG/ML AUTO-INJECTOR |
| 1747195 | PR00 | Immunosuppressants | METHOTREXATE 56.3 MG/ML AUTO-INJECTOR |
| 1921592 | PR00 | Immunosuppressants | METHOTREXATE 2.5 MG/ML ORAL SOLUTION |
| 1946772 | PR00 | Immunosuppressants | METHOTREXATE 25 MG/ML INJECTABLE SOLUTION |
| 245256 | PR00 | Immunosuppressants | METHOTREXATE 10 MG/ML INJECTABLE SOLUTION |
| 250936 | PR00 | Immunosuppressants | METHOTREXATE 500 MG INJECTABLE SOLUTION |
| 283510 | PR00 | Immunosuppressants | METHOTREXATE 15 MG ORAL TABLET |
| 283511 | PR00 | Immunosuppressants | METHOTREXATE 5 MG ORAL TABLET |

| | | | |
|---|---|---|---|
| 283671 | PR00 | Immunosuppressants | METHOTREXATE 7.5 MG ORAL TABLET |
| 311624 | PR00 | Immunosuppressants | METHOTREXATE 15 MG/ML INJECTABLE SOLUTION |
| 311625 | PR00 | Immunosuppressants | METHOTREXATE 1000 MG INJECTION |
| 311626 | PR00 | Immunosuppressants | METHOTREXATE 12.5 MG/ML INJECTABLE SOLUTION |
| 311627 | PR00 | Immunosuppressants | METHOTREXATE 25 MG/ML INJECTABLE SOLUTION |
| 315148 | PR00 | Immunosuppressants | METHOTREXATE 2.5 MG/ML INJECTABLE SOLUTION |
| 415045 | PR00 | Immunosuppressants | METHOTREXATE 5 MG/ML INJECTABLE SOLUTION |
| 415046 | PR00 | Immunosuppressants | METHOTREXATE 7.5 MG/ML INJECTABLE SOLUTION |
| 415063 | PR00 | Immunosuppressants | METHOTREXATE 7.46 MG/ML INJECTABLE SOLUTION |
| 1441410 | PR00 | Immunosuppressants | METHOTREXATE 37.5 MG/ML |
| 1441421 | PR00 | Immunosuppressants | METHOTREXATE 62.5 MG/ML |
| 1594756 | PR00 | Immunosuppressants | METHOTREXATE 18.75 MG/ML |
| 1656681 | PR00 | Immunosuppressants | METHOTREXATE 1000 MG |
| 1659719 | PR00 | Immunosuppressants | METHOTREXATE 18.8 MG/ML |
| 1747178 | PR00 | Immunosuppressants | METHOTREXATE 31.3 MG/ML |
| 1747184 | PR00 | Immunosuppressants | METHOTREXATE 43.8 MG/ML |
| 1747191 | PR00 | Immunosuppressants | METHOTREXATE 56.3 MG/ML |
| 328404 | PR00 | Immunosuppressants | METHOTREXATE 50 MG/ML |
| 328406 | PR00 | Immunosuppressants | METHOTREXATE 25 MG/ML |
| 328407 | PR00 | Immunosuppressants | METHOTREXATE 2.5 MG |
| 330028 | PR00 | Immunosuppressants | METHOTREXATE 2.5 MG/ML |
| 331568 | PR00 | Immunosuppressants | METHOTREXATE 10 MG |
| 331569 | PR00 | Immunosuppressants | METHOTREXATE 15 MG |
| 332454 | PR00 | Immunosuppressants | METHOTREXATE 5 MG |
| 332455 | PR00 | Immunosuppressants | METHOTREXATE 7.5 MG |
| 334051 | PR00 | Immunosuppressants | METHOTREXATE 10 MG/ML |
| 334421 | PR00 | Immunosuppressants | METHOTREXATE 500 MG |
| 346335 | PR00 | Immunosuppressants | METHOTREXATE 12.5 MG/ML |
| 360547 | PR00 | Immunosuppressants | METHOTREXATE 100 MG/ML |
| 380984 | PR00 | Immunosuppressants | METHOTREXATE 15 MG/ML |
| 451557 | PR00 | Immunosuppressants | METHOTREXATE 7.46 MG/ML |
| 451938 | PR00 | Immunosuppressants | METHOTREXATE 5 MG/ML |
| 451941 | PR00 | Immunosuppressants | METHOTREXATE 7.5 MG/ML |
| 1441401 | PR00 | Immunosuppressants | METHOTREXATE PREFILLED SYRINGE |
| 1655203 | PR00 | Immunosuppressants | METHOTREXATE AUTO-INJECTOR |
| 1655955 | PR00 | Immunosuppressants | METHOTREXATE INJECTION |
| 1921591 | PR00 | Immunosuppressants | METHOTREXATE ORAL SOLUTION |
| 372835 | PR00 | Immunosuppressants | METHOTREXATE INJECTABLE SOLUTION |
| 372836 | PR00 | Immunosuppressants | METHOTREXATE ORAL TABLET |
| 1162235 | PR00 | Immunosuppressants | METHOTREXATE INJECTABLE PRODUCT |
| 1162236 | PR00 | Immunosuppressants | METHOTREXATE ORAL PRODUCT |
| 1162237 | PR00 | Immunosuppressants | METHOTREXATE PILL |
| 1921590 | PR00 | Immunosuppressants | METHOTREXATE ORAL LIQUID PRODUCT |
| 227302 | PR00 | Immunosuppressants | ORTHOCLONE OKT3 |
| 42405 | PR00 | Immunosuppressants | MUROMONAB-CD3 |
| 206950 | PR00 | Immunosuppressants | MUROMONAB-CD3 1 MG/ML INJECTABLE SOLUTION [ORTHOCLONE OKT3] |
| 567751 | PR00 | Immunosuppressants | MUROMONAB-CD3 1 MG/ML [ORTHOCLONE OKT3] |
| 402708 | PR00 | Immunosuppressants | MUROMONAB-CD3 INJECTABLE SOLUTION [ORTHOCLONE OKT3] |
| 1184532 | PR00 | Immunosuppressants | ORTHOCLONE OKT3 INJECTABLE PRODUCT |
| 239984 | PR00 | Immunosuppressants | MUROMONAB-CD3 1 MG/ML INJECTABLE SOLUTION |
| 317432 | PR00 | Immunosuppressants | MUROMONAB-CD3 1 MG/ML |
| 372973 | PR00 | Immunosuppressants | MUROMONAB-CD3 INJECTABLE SOLUTION |
| 1156388 | PR00 | Immunosuppressants | MUROMONAB-CD3 INJECTABLE PRODUCT |
| 152633 | PR00 | Immunosuppressants | CELLCEPT |
| 265323 | PR00 | Immunosuppressants | MYCOPHENOLATE |
| 68149 | PR00 | Immunosuppressants | MYCOPHENOLATE MOFETIL |
| 237112 | PR00 | Immunosuppressants | MYCOPHENOLATE MOFETIL HYDROCHLORIDE |
| 408142 | PR00 | Immunosuppressants | MYCOPHENOLATE SODIUM |
| 152591 | PR00 | Immunosuppressants | MYCOPHENOLATE 250 MG ORAL CAPSULE [CELLCEPT] |
| 153754 | PR00 | Immunosuppressants | MYCOPHENOLATE 500 MG ORAL TABLET [CELLCEPT] |
| 213394 | PR00 | Immunosuppressants | MYCOPHENOLATE MOFETIL 500 MG INJECTION [CELLCEPT] |
| 261236 | PR00 | Immunosuppressants | MYCOPHENOLATE 200 MG/ML ORAL SUSPENSION [CELLCEPT] |
| 579573 | PR00 | Immunosuppressants | MYCOPHENOLATE 180 MG ENTERIC COATED TABLET [MYFORTIC] |
| 579576 | PR00 | Immunosuppressants | MYCOPHENOLATE 360 MG ENTERIC COATED TABLET [MYFORTIC] |
| 616435 | PR00 | Immunosuppressants | MYCOPHENOLATE MOFETIL 500 MG ORAL TABLET [CELLCEPT] |
| 616444 | PR00 | Immunosuppressants | MYCOPHENOLATE MOFETIL 200 MG/ML ORAL SUSPENSION [CELLCEPT] |
| 616447 | PR00 | Immunosuppressants | MYCOPHENOLATE MOFETIL 250 MG ORAL CAPSULE [CELLCEPT] |
| 616449 | PR00 | Immunosuppressants | MYCOPHENOLATE 360 MG ENTERIC COATED TABLET [MYFORTIC] |
| 616450 | PR00 | Immunosuppressants | MYCOPHENOLATE 180 MG ENTERIC COATED TABLET [MYFORTIC] |
| 541580 | PR00 | Immunosuppressants | MYCOPHENOLATE 180 MG [MYFORTIC] |
| 541585 | PR00 | Immunosuppressants | MYCOPHENOLATE 360 MG [MYFORTIC] |

| | | | |
|---|---|---|---|
| 564992 | PR00 | Immunosuppressants | MYCOPHENOLATE 250 MG [CELLCEPT] |
| 565378 | PR00 | Immunosuppressants | MYCOPHENOLATE 500 MG [CELLCEPT] |
| 573288 | PR00 | Immunosuppressants | MYCOPHENOLATE MOFETIL 6 MG/ML [CELLCEPT] |
| 574465 | PR00 | Immunosuppressants | MYCOPHENOLATE 200 MG/ML [CELLCEPT] |
| 579571 | PR00 | Immunosuppressants | MYCOPHENOLATE 180 MG [MYFORTIC] |
| 579574 | PR00 | Immunosuppressants | MYCOPHENOLATE 360 MG [MYFORTIC] |
| 616433 | PR00 | Immunosuppressants | MYCOPHENOLATE MOFETIL 500 MG [CELLCEPT] |
| 616442 | PR00 | Immunosuppressants | MYCOPHENOLATE MOFETIL 200 MG/ML [CELLCEPT] |
| 616445 | PR00 | Immunosuppressants | MYCOPHENOLATE MOFETIL 250 MG [CELLCEPT] |
| 1726179 | PR00 | Immunosuppressants | MYCOPHENOLATE MOFETIL INJECTION [CELLCEPT] |
| 363166 | PR00 | Immunosuppressants | MYCOPHENOLATE MOFETIL INJECTABLE SOLUTION [CELLCEPT] |
| 365853 | PR00 | Immunosuppressants | MYCOPHENOLATE ORAL SUSPENSION [CELLCEPT] |
| 366524 | PR00 | Immunosuppressants | MYCOPHENOLATE ORAL CAPSULE [CELLCEPT] |
| 369377 | PR00 | Immunosuppressants | MYCOPHENOLATE ORAL TABLET [CELLCEPT] |
| 579572 | PR00 | Immunosuppressants | MYCOPHENOLATE ENTERIC COATED TABLET [MYFORTIC] |
| 579575 | PR00 | Immunosuppressants | MYCOPHENOLATE ENTERIC COATED TABLET [MYFORTIC] |
| 616434 | PR00 | Immunosuppressants | MYCOPHENOLATE MOFETIL ORAL TABLET [CELLCEPT] |
| 616443 | PR00 | Immunosuppressants | MYCOPHENOLATE MOFETIL ORAL SUSPENSION [CELLCEPT] |
| 616446 | PR00 | Immunosuppressants | MYCOPHENOLATE MOFETIL ORAL CAPSULE [CELLCEPT] |
| 616448 | PR00 | Immunosuppressants | MYCOPHENOLATE ENTERIC COATED TABLET [MYFORTIC] |
| 1166213 | PR00 | Immunosuppressants | CELLCEPT INJECTABLE PRODUCT |
| 1166214 | PR00 | Immunosuppressants | CELLCEPT ORAL LIQUID PRODUCT |
| 1166215 | PR00 | Immunosuppressants | CELLCEPT ORAL PRODUCT |
| 1166216 | PR00 | Immunosuppressants | CELLCEPT PILL |
| 199058 | PR00 | Immunosuppressants | MYCOPHENOLATE MOFETIL 250 MG ORAL CAPSULE |
| 200060 | PR00 | Immunosuppressants | MYCOPHENOLATE MOFETIL 500 MG ORAL TABLET |
| 311880 | PR00 | Immunosuppressants | MYCOPHENOLATE MOFETIL 200 MG/ML ORAL SUSPENSION |
| 414999 | PR00 | Immunosuppressants | MYCOPHENOLATE MOFETIL 25 MG/ML INJECTABLE SOLUTION |
| 433833 | PR00 | Immunosuppressants | MYCOPHENOLATE 180 MG ENTERIC COATED TABLET |
| 433834 | PR00 | Immunosuppressants | MYCOPHENOLATE 360 MG ENTERIC COATED TABLET |
| 562143 | PR00 | Immunosuppressants | MYCOPHENOLATE 250 MG ORAL CAPSULE |
| 562175 | PR00 | Immunosuppressants | MYCOPHENOLATE 500 MG ORAL TABLET |
| 562701 | PR00 | Immunosuppressants | MYCOPHENOLATE 200 MG/ML ORAL SUSPENSION |
| 579569 | PR00 | Immunosuppressants | MYCOPHENOLATE 180 MG ENTERIC COATED TABLET |
| 596873 | PR00 | Immunosuppressants | MYCOPHENOLATE 180 MG EXTENDED RELEASE ENTERIC COATED TABLET |
| 670049 | PR00 | Immunosuppressants | MYCOPHENOLATE 180 MG ENTERIC COATED TABLET |
| 670051 | PR00 | Immunosuppressants | MYCOPHENOLATE 360 MG ENTERIC COATED TABLET |
| 316316 | PR00 | Immunosuppressants | MYCOPHENOLATE MOFETIL 250 MG |
| 316317 | PR00 | Immunosuppressants | MYCOPHENOLATE MOFETIL 500 MG |
| 331587 | PR00 | Immunosuppressants | MYCOPHENOLATE MOFETIL 200 MG/ML |
| 332547 | PR00 | Immunosuppressants | MYCOPHENOLATE MOFETIL 6 MG/ML |
| 353404 | PR00 | Immunosuppressants | MYCOPHENOLATE 250 MG |
| 353412 | PR00 | Immunosuppressants | MYCOPHENOLATE 500 MG |
| 353414 | PR00 | Immunosuppressants | MYCOPHENOLATE 200 MG/ML |
| 446560 | PR00 | Immunosuppressants | MYCOPHENOLATE 180 MG |
| 446562 | PR00 | Immunosuppressants | MYCOPHENOLATE 360 MG |
| 452791 | PR00 | Immunosuppressants | MYCOPHENOLATE MOFETIL 25 MG/ML |
| 485018 | PR00 | Immunosuppressants | MYCOPHENOLATE 180 MG |
| 485022 | PR00 | Immunosuppressants | MYCOPHENOLATE 360 MG |
| 579567 | PR00 | Immunosuppressants | MYCOPHENOLATE 180 MG |
| 670048 | PR00 | Immunosuppressants | MYCOPHENOLATE 180 MG |
| 670050 | PR00 | Immunosuppressants | MYCOPHENOLATE 360 MG |
| 1726176 | PR00 | Immunosuppressants | MYCOPHENOLATE MOFETIL INJECTION |
| 374667 | PR00 | Immunosuppressants | MYCOPHENOLATE MOFETIL ORAL TABLET |
| 374670 | PR00 | Immunosuppressants | MYCOPHENOLATE MOFETIL ORAL SUSPENSION |
| 376670 | PR00 | Immunosuppressants | MYCOPHENOLATE MOFETIL INJECTABLE SOLUTION |
| 379060 | PR00 | Immunosuppressants | MYCOPHENOLATE MOFETIL ORAL CAPSULE |
| 446561 | PR00 | Immunosuppressants | MYCOPHENOLATE ENTERIC COATED TABLET |
| 562142 | PR00 | Immunosuppressants | MYCOPHENOLATE ORAL CAPSULE |
| 562174 | PR00 | Immunosuppressants | MYCOPHENOLATE ORAL TABLET |
| 562700 | PR00 | Immunosuppressants | MYCOPHENOLATE ORAL SUSPENSION |
| 579568 | PR00 | Immunosuppressants | MYCOPHENOLATE ENTERIC COATED TABLET |
| 596872 | PR00 | Immunosuppressants | MYCOPHENOLATE EXTENDED RELEASE ENTERIC COATED TABLET |
| 1157869 | PR00 | Immunosuppressants | MYCOPHENOLATE MOFETIL INJECTABLE PRODUCT |
| 1157870 | PR00 | Immunosuppressants | MYCOPHENOLATE MOFETIL ORAL LIQUID PRODUCT |
| 1157871 | PR00 | Immunosuppressants | MYCOPHENOLATE MOFETIL ORAL PRODUCT |
| 1157872 | PR00 | Immunosuppressants | MYCOPHENOLATE MOFETIL PILL |
| 258355 | PR00 | Immunosuppressants | RAPAMUNE |
| 35302 | PR00 | Immunosuppressants | SIROLIMUS |
| 1724309 | PR00 | Immunosuppressants | TEMSIROLIMUS 25 MG/ML INJECTION [TORISEL] |
| 351901 | PR00 | Immunosuppressants | SIROLIMUS 1 MG/ML ORAL SOLUTION [RAPAMUNE] |

| | | | |
|---|---|---|---|
| 351989 | PR00 | Immunosuppressants | SIROLIMUS 1 MG ORAL TABLET [RAPAMUNE] |
| 404432 | PR00 | Immunosuppressants | SIROLIMUS 2 MG ORAL TABLET [RAPAMUNE] |
| 905160 | PR00 | Immunosuppressants | SIROLIMUS 0.5 MG ORAL TABLET [RAPAMUNE] |
| 575659 | PR00 | Immunosuppressants | SIROLIMUS 1 MG/ML [RAPAMUNE] |
| 575733 | PR00 | Immunosuppressants | SIROLIMUS 1 MG [RAPAMUNE] |
| 576390 | PR00 | Immunosuppressants | SIROLIMUS 2 MG [RAPAMUNE] |
| 905159 | PR00 | Immunosuppressants | SIROLIMUS 0.5 MG [RAPAMUNE] |
| 1724306 | PR00 | Immunosuppressants | TEMSIROLIMUS INJECTION [TORISEL] |
| 364904 | PR00 | Immunosuppressants | SIROLIMUS ORAL SOLUTION [RAPAMUNE] |
| 368672 | PR00 | Immunosuppressants | SIROLIMUS ORAL TABLET [RAPAMUNE] |
| 1177701 | PR00 | Immunosuppressants | RAPAMUNE ORAL LIQUID PRODUCT |
| 1177702 | PR00 | Immunosuppressants | RAPAMUNE ORAL PRODUCT |
| 1177703 | PR00 | Immunosuppressants | RAPAMUNE PILL |
| 1178590 | PR00 | Immunosuppressants | TORISEL INJECTABLE PRODUCT |
| 314230 | PR00 | Immunosuppressants | SIROLIMUS 1 MG/ML ORAL SOLUTION |
| 349208 | PR00 | Immunosuppressants | SIROLIMUS 1 MG ORAL TABLET |
| 360110 | PR00 | Immunosuppressants | SIROLIMUS 2 MG ORAL TABLET |
| 905158 | PR00 | Immunosuppressants | SIROLIMUS 0.5 MG ORAL TABLET |
| 333296 | PR00 | Immunosuppressants | SIROLIMUS 1 MG/ML |
| 350512 | PR00 | Immunosuppressants | SIROLIMUS 1 MG |
| 360318 | PR00 | Immunosuppressants | SIROLIMUS 2 MG |
| 905157 | PR00 | Immunosuppressants | SIROLIMUS 0.5 MG |
| 1724304 | PR00 | Immunosuppressants | TEMSIROLIMUS INJECTION |
| 375278 | PR00 | Immunosuppressants | SIROLIMUS ORAL SOLUTION |
| 378277 | PR00 | Immunosuppressants | SIROLIMUS ORAL TABLET |
| 1158757 | PR00 | Immunosuppressants | SIROLIMUS ORAL LIQUID PRODUCT |
| 1158758 | PR00 | Immunosuppressants | SIROLIMUS ORAL PRODUCT |
| 1158759 | PR00 | Immunosuppressants | SIROLIMUS PILL |
| 1161488 | PR00 | Immunosuppressants | TEMSIROLIMUS INJECTABLE PRODUCT |
| 1235236 | PR00 | Immunosuppressants | HECORIA |
| 1431972 | PR00 | Immunosuppressants | ASTAGRAF |
| 1664435 | PR00 | Immunosuppressants | ENVARSUS |
| 196463 | PR00 | Immunosuppressants | PROGRAF |
| 42316 | PR00 | Immunosuppressants | TACROLIMUS |
| 235991 | PR00 | Immunosuppressants | ANHYDROUS TACROLIMUS |
| 484288 | PR00 | Immunosuppressants | TACROLIMUS MONOHYDRATE |
| 108513 | PR00 | Immunosuppressants | TACROLIMUS 1 MG ORAL CAPSULE [PROGRAF] |
| 108514 | PR00 | Immunosuppressants | TACROLIMUS 5 MG ORAL CAPSULE [PROGRAF] |
| 108515 | PR00 | Immunosuppressants | 1 ML TACROLIMUS 5 MG/ML INJECTION [PROGRAF] |
| 1235241 | PR00 | Immunosuppressants | TACROLIMUS 0.5 MG ORAL CAPSULE [HECORIA] |
| 1235243 | PR00 | Immunosuppressants | TACROLIMUS 1 MG ORAL CAPSULE [HECORIA] |
| 1235245 | PR00 | Immunosuppressants | TACROLIMUS 5 MG ORAL CAPSULE [HECORIA] |
| 1431977 | PR00 | Immunosuppressants | 24 HR TACROLIMUS 0.5 MG EXTENDED RELEASE ORAL CAPSULE [ASTAGRAF] |
| 1431979 | PR00 | Immunosuppressants | TACROLIMUS 0.5 MG EXTENDED RELEASE ORAL CAPSULE [ASTAGRAF] |
| 1431982 | PR00 | Immunosuppressants | 24 HR TACROLIMUS 1 MG EXTENDED RELEASE ORAL CAPSULE [ASTAGRAF] |
| 1431984 | PR00 | Immunosuppressants | TACROLIMUS 1 MG EXTENDED RELEASE ORAL CAPSULE [ASTAGRAF] |
| 1431987 | PR00 | Immunosuppressants | 24 HR TACROLIMUS 5 MG EXTENDED RELEASE ORAL CAPSULE [ASTAGRAF] |
| 1431989 | PR00 | Immunosuppressants | TACROLIMUS 5 MG EXTENDED RELEASE ORAL CAPSULE [ASTAGRAF] |
| 1664440 | PR00 | Immunosuppressants | 24 HR TACROLIMUS 4 MG EXTENDED RELEASE ORAL TABLET [ENVARSUS] |
| 1664442 | PR00 | Immunosuppressants | TACROLIMUS 4 MG EXTENDED RELEASE ORAL TABLET [ENVARSUS] |
| 1664458 | PR00 | Immunosuppressants | 24 HR TACROLIMUS 0.75 MG EXTENDED RELEASE ORAL TABLET [ENVARSUS] |
| 1664460 | PR00 | Immunosuppressants | TACROLIMUS 0.75 MG EXTENDED RELEASE ORAL TABLET [ENVARSUS] |
| 1664463 | PR00 | Immunosuppressants | 24 HR TACROLIMUS 1 MG EXTENDED RELEASE ORAL TABLET [ENVARSUS] |
| 1664465 | PR00 | Immunosuppressants | TACROLIMUS 1 MG EXTENDED RELEASE ORAL TABLET [ENVARSUS] |
| 1738339 | PR00 | Immunosuppressants | TACROLIMUS 5 MG/ML INJECTION [PROGRAF] |
| 2052609 | PR00 | Immunosuppressants | TACROLIMUS 0.2 MG GRANULES FOR ORAL SUSPENSION [PROGRAF] |
| 2052704 | PR00 | Immunosuppressants | TACROLIMUS 1 MG GRANULES FOR ORAL SUSPENSION [PROGRAF] |
| 261134 | PR00 | Immunosuppressants | TACROLIMUS 0.5 MG ORAL CAPSULE [PROGRAF] |
| 1235237 | PR00 | Immunosuppressants | TACROLIMUS 0.5 MG [HECORIA] |
| 1235242 | PR00 | Immunosuppressants | TACROLIMUS 1 MG [HECORIA] |
| 1235244 | PR00 | Immunosuppressants | TACROLIMUS 5 MG [HECORIA] |
| 1431973 | PR00 | Immunosuppressants | TACROLIMUS 0.5 MG [ASTAGRAF] |
| 1431981 | PR00 | Immunosuppressants | TACROLIMUS 1 MG [ASTAGRAF] |
| 1431986 | PR00 | Immunosuppressants | TACROLIMUS 5 MG [ASTAGRAF] |
| 1664436 | PR00 | Immunosuppressants | TACROLIMUS 4 MG [ENVARSUS] |
| 1664457 | PR00 | Immunosuppressants | TACROLIMUS 0.75 MG [ENVARSUS] |
| 1664462 | PR00 | Immunosuppressants | TACROLIMUS 1 MG [ENVARSUS] |
| 2052606 | PR00 | Immunosuppressants | TACROLIMUS 0.2 MG [PROGRAF] |
| 564557 | PR00 | Immunosuppressants | TACROLIMUS 1 MG [PROGRAF] |
| 564558 | PR00 | Immunosuppressants | TACROLIMUS 5 MG [PROGRAF] |
| 564559 | PR00 | Immunosuppressants | TACROLIMUS 5 MG/ML [PROGRAF] |

| | | | |
|---|---|---|---|
| 574378 | PR00 | Immunosuppressants | TACROLIMUS 0.5 MG [PROGRAF] |
| 1235238 | PR00 | Immunosuppressants | TACROLIMUS ORAL CAPSULE [HECORIA] |
| 1431974 | PR00 | Immunosuppressants | TACROLIMUS EXTENDED RELEASE ORAL CAPSULE [ASTAGRAF] |
| 1664437 | PR00 | Immunosuppressants | TACROLIMUS EXTENDED RELEASE ORAL TABLET [ENVARSUS] |
| 1738336 | PR00 | Immunosuppressants | TACROLIMUS INJECTION [PROGRAF] |
| 2052607 | PR00 | Immunosuppressants | TACROLIMUS GRANULES FOR ORAL SUSPENSION [PROGRAF] |
| 362879 | PR00 | Immunosuppressants | TACROLIMUS INJECTABLE SOLUTION [PROGRAF] |
| 366273 | PR00 | Immunosuppressants | TACROLIMUS ORAL CAPSULE [PROGRAF] |
| 1180662 | PR00 | Immunosuppressants | PROGRAF INJECTABLE PRODUCT |
| 1180663 | PR00 | Immunosuppressants | PROGRAF ORAL PRODUCT |
| 1180664 | PR00 | Immunosuppressants | PROGRAF PILL |
| 1235239 | PR00 | Immunosuppressants | HECORIA ORAL PRODUCT |
| 1235240 | PR00 | Immunosuppressants | HECORIA PILL |
| 1431975 | PR00 | Immunosuppressants | ASTAGRAF ORAL PRODUCT |
| 1431976 | PR00 | Immunosuppressants | ASTAGRAF PILL |
| 1664438 | PR00 | Immunosuppressants | ENVARSUS ORAL PRODUCT |
| 1664439 | PR00 | Immunosuppressants | ENVARSUS PILL |
| 2052608 | PR00 | Immunosuppressants | PROGRAF GRANULE PRODUCT |
| 1431971 | PR00 | Immunosuppressants | 24 HR TACROLIMUS 0.5 MG EXTENDED RELEASE ORAL CAPSULE |
| 1431978 | PR00 | Immunosuppressants | TACROLIMUS 0.5 MG EXTENDED RELEASE ORAL CAPSULE |
| 1431980 | PR00 | Immunosuppressants | 24 HR TACROLIMUS 1 MG EXTENDED RELEASE ORAL CAPSULE |
| 1431983 | PR00 | Immunosuppressants | TACROLIMUS 1 MG EXTENDED RELEASE ORAL CAPSULE |
| 1431985 | PR00 | Immunosuppressants | 24 HR TACROLIMUS 5 MG EXTENDED RELEASE ORAL CAPSULE |
| 1431988 | PR00 | Immunosuppressants | TACROLIMUS 5 MG EXTENDED RELEASE ORAL CAPSULE |
| 1664434 | PR00 | Immunosuppressants | 24 HR TACROLIMUS 4 MG EXTENDED RELEASE ORAL TABLET |
| 1664441 | PR00 | Immunosuppressants | TACROLIMUS 4 MG EXTENDED RELEASE ORAL TABLET |
| 1664456 | PR00 | Immunosuppressants | 24 HR TACROLIMUS 0.75 MG EXTENDED RELEASE ORAL TABLET |
| 1664459 | PR00 | Immunosuppressants | TACROLIMUS 0.75 MG EXTENDED RELEASE ORAL TABLET |
| 1664461 | PR00 | Immunosuppressants | 24 HR TACROLIMUS 1 MG EXTENDED RELEASE ORAL TABLET |
| 1664464 | PR00 | Immunosuppressants | TACROLIMUS 1 MG EXTENDED RELEASE ORAL TABLET |
| 1738338 | PR00 | Immunosuppressants | TACROLIMUS 5 MG/ML INJECTION |
| 198377 | PR00 | Immunosuppressants | TACROLIMUS 1 MG ORAL CAPSULE |
| 198378 | PR00 | Immunosuppressants | TACROLIMUS 5 MG ORAL CAPSULE |
| 198379 | PR00 | Immunosuppressants | 1 ML TACROLIMUS 5 MG/ML INJECTION |
| 2052605 | PR00 | Immunosuppressants | TACROLIMUS 0.2 MG GRANULES FOR ORAL SUSPENSION |
| 2052703 | PR00 | Immunosuppressants | TACROLIMUS 1 MG GRANULES FOR ORAL SUSPENSION |
| 427807 | PR00 | Immunosuppressants | TACROLIMUS 5 MG ORAL TABLET |
| 427808 | PR00 | Immunosuppressants | TACROLIMUS 1 MG ORAL TABLET |
| 1599039 | PR00 | Immunosuppressants | TACROLIMUS 0.5 MG/ML |
| 1664432 | PR00 | Immunosuppressants | TACROLIMUS 4 MG |
| 1664455 | PR00 | Immunosuppressants | TACROLIMUS 0.75 MG |
| 2052503 | PR00 | Immunosuppressants | TACROLIMUS 0.2 MG |
| 316758 | PR00 | Immunosuppressants | TACROLIMUS 1 MG |
| 316759 | PR00 | Immunosuppressants | TACROLIMUS 5 MG |
| 317510 | PR00 | Immunosuppressants | TACROLIMUS 5 MG/ML |
| 330404 | PR00 | Immunosuppressants | TACROLIMUS 0.5 MG |
| 331549 | PR00 | Immunosuppressants | TACROLIMUS 0.001 MG/MG |
| 331550 | PR00 | Immunosuppressants | TACROLIMUS 0.0003 MG/MG |
| 1431970 | PR00 | Immunosuppressants | TACROLIMUS EXTENDED RELEASE ORAL CAPSULE |
| 1664433 | PR00 | Immunosuppressants | TACROLIMUS EXTENDED RELEASE ORAL TABLET |
| 1738334 | PR00 | Immunosuppressants | TACROLIMUS INJECTION |
| 2052604 | PR00 | Immunosuppressants | TACROLIMUS GRANULES FOR ORAL SUSPENSION |
| 374015 | PR00 | Immunosuppressants | TACROLIMUS ORAL CAPSULE |
| 377119 | PR00 | Immunosuppressants | TACROLIMUS INJECTABLE SOLUTION |
| 451875 | PR00 | Immunosuppressants | TACROLIMUS ORAL TABLET |
| 1163655 | PR00 | Immunosuppressants | TACROLIMUS INJECTABLE PRODUCT |
| 1164202 | PR00 | Immunosuppressants | TACROLIMUS ORAL PRODUCT |
| 1164203 | PR00 | Immunosuppressants | TACROLIMUS PILL |
| 2052603 | PR00 | Immunosuppressants | TACROLIMUS GRANULE PRODUCT |
| 723805 | PR00 | Immunosuppressants | TORISEL |
| 657797 | PR00 | Immunosuppressants | TEMSIROLIMUS |
| 725108 | PR00 | Immunosuppressants | 1.2 ML TEMSIROLIMUS 25 MG/ML INJECTION [TORISEL] |
| 725106 | PR00 | Immunosuppressants | TEMSIROLIMUS 25 MG/ML [TORISEL] |
| 725107 | PR00 | Immunosuppressants | TEMSIROLIMUS INJECTABLE SOLUTION [TORISEL] |
| 1724308 | PR00 | Immunosuppressants | TEMSIROLIMUS 25 MG/ML INJECTION |
| 722289 | PR00 | Immunosuppressants | 1.2 ML TEMSIROLIMUS 25 MG/ML INJECTION |
| 993785 | PR00 | Immunosuppressants | TEMSIROLIMUS 10 MG/ML INJECTABLE SOLUTION |
| 722287 | PR00 | Immunosuppressants | TEMSIROLIMUS 25 MG/ML |
| 993784 | PR00 | Immunosuppressants | TEMSIROLIMUS 10 MG/ML |
| 722288 | PR00 | Immunosuppressants | TEMSIROLIMUS INJECTABLE SOLUTION |
| 72435 | PR00 | Immunosuppressants | anakinra |

| | | | |
|---|---|---|---|
| 352734 | PR00 | Immunosuppressants | Kineret |
| 727711 | PR00 | Immunosuppressants | 0.67 ML anakinra 149 MG/ML Prefilled Syringe |
| 727714 | PR00 | Immunosuppressants | 0.67 ML anakinra 149 MG/ML Prefilled Syringe [Kineret] |
| 727708 | PR00 | Immunosuppressants | anakinra Prefilled Syringe |
| 727710 | PR00 | Immunosuppressants | anakinra 149 MG/ML |
| 727712 | PR00 | Immunosuppressants | anakinra 149 MG/ML [Kineret] |
| 727713 | PR00 | Immunosuppressants | anakinra Prefilled Syringe [Kineret] |
| 1157701 | PR00 | Immunosuppressants | anakinra Injectable Product |
| 1170741 | PR00 | Immunosuppressants | Kineret Injectable Product |
| 1012899 | PR00 | Immunosuppressants | fingolimod 0.5 MG |
| 1193337 | PR00 | Immunosuppressants | ruxolitinib 10 MG |
| 1193341 | PR00 | Immunosuppressants | ruxolitinib 5 MG |
| 1193345 | PR00 | Immunosuppressants | ruxolitinib 15 MG |
| 1193349 | PR00 | Immunosuppressants | ruxolitinib 20 MG |
| 1193353 | PR00 | Immunosuppressants | ruxolitinib 25 MG |
| 1442698 | PR00 | Immunosuppressants | obinutuzumab 25 MG/ML |
| 1654079 | PR00 | Immunosuppressants | ustekinumab 90 MG/ML |
| 1720606 | PR00 | Immunosuppressants | mepolizumab 100 MG |
| 1745108 | PR00 | Immunosuppressants | ixekizumab 80 MG/ML |
| 1745114 | PR00 | Immunosuppressants | ixekizumab 80 MG/ML |
| 1811257 | PR00 | Immunosuppressants | ustekinumab 5 MG/ML |
| 1872261 | PR00 | Immunosuppressants | brodalumab 140 MG/ML |
| 1876406 | PR00 | Immunosuppressants | dupilumab 150 MG/ML |
| 1923338 | PR00 | Immunosuppressants | sarilumab 132 MG/ML |
| 1923347 | PR00 | Immunosuppressants | sarilumab 175 MG/ML |
| 1928689 | PR00 | Immunosuppressants | guselkumab 100 MG/ML |
| 1942959 | PR00 | Immunosuppressants | inotuzumab ozogamicin 0.9 MG |
| 1989109 | PR00 | Immunosuppressants | benralizumab 30 MG/ML |
| 2003757 | PR00 | Immunosuppressants | sarilumab 132 MG/ML |
| 2003767 | PR00 | Immunosuppressants | sarilumab 175 MG/ML |
| 2045493 | PR00 | Immunosuppressants | fingolimod 0.25 MG |
| 2055026 | PR00 | Immunosuppressants | Cyclosporine 0.9 MG/ML |
| 2099302 | PR00 | Immunosuppressants | dupilumab 175 MG/ML |
| 2118741 | PR00 | Immunosuppressants | guselkumab 100 MG/ML |
| 2170993 | PR00 | Immunosuppressants | mepolizumab 100 MG/ML |
| 2173822 | PR00 | Immunosuppressants | mepolizumab 100 MG/ML |
| 2196900 | PR00 | Immunosuppressants | upadacitinib 15 MG |
| 311881 | PR00 | Immunosuppressants | mycophenolate mofetil 500 MG |
| 313190 | PR00 | Immunosuppressants | Tacrolimus 0.5 MG |
| 402092 | PR00 | Immunosuppressants | Cyclosporine 0.5 MG/ML |
| 402240 | PR00 | Immunosuppressants | efalizumab 100 MG/ML |
| 603541 | PR00 | Immunosuppressants | natalizumab 20 MG/ML |
| 795724 | PR00 | Immunosuppressants | Methohexital 50 MG/ML |
| 853354 | PR00 | Immunosuppressants | ustekinumab 90 MG/ML |
| 853356 | PR00 | Immunosuppressants | ustekinumab 90 MG/ML |
| 865174 | PR00 | Immunosuppressants | ustekinumab 90 MG/ML |
| 1012891 | PR00 | Immunosuppressants | fingolimod hydrochloride |
| 1012892 | PR00 | Immunosuppressants | fingolimod |
| 1012893 | PR00 | Immunosuppressants | fingolimod 0.5 MG |
| 1012894 | PR00 | Immunosuppressants | fingolimod Oral Capsule |
| 1012895 | PR00 | Immunosuppressants | fingolimod 0.5 MG |
| 1012897 | PR00 | Immunosuppressants | fingolimod 0.5 MG |
| 1012898 | PR00 | Immunosuppressants | fingolimod Oral Capsule |
| 1158395 | PR00 | Immunosuppressants | natalizumab Injectable Product |
| 1161283 | PR00 | Immunosuppressants | efalizumab Injectable Product |
| 1163983 | PR00 | Immunosuppressants | ustekinumab Injectable Product |
| 1165017 | PR00 | Immunosuppressants | fingolimod Oral Product |
| 1165018 | PR00 | Immunosuppressants | fingolimod Pill |
| 1193325 | PR00 | Immunosuppressants | ruxolitinib phosphate |
| 1193326 | PR00 | Immunosuppressants | ruxolitinib |
| 1193327 | PR00 | Immunosuppressants | ruxolitinib 10 MG |
| 1193328 | PR00 | Immunosuppressants | ruxolitinib Oral Product |
| 1193329 | PR00 | Immunosuppressants | ruxolitinib Pill |
| 1193330 | PR00 | Immunosuppressants | ruxolitinib Oral Tablet |
| 1193331 | PR00 | Immunosuppressants | ruxolitinib 10 MG |
| 1193333 | PR00 | Immunosuppressants | ruxolitinib 10 MG |
| 1193334 | PR00 | Immunosuppressants | ruxolitinib Oral Tablet |
| 1193338 | PR00 | Immunosuppressants | ruxolitinib 5 MG |
| 1193339 | PR00 | Immunosuppressants | ruxolitinib 5 MG |
| 1193340 | PR00 | Immunosuppressants | ruxolitinib 5 MG |
| 1193342 | PR00 | Immunosuppressants | ruxolitinib 15 MG |

| | | | |
|---|---|---|---|
| 1193343 | PR00 | Immunosuppressants | ruxolitinib 15 MG |
| 1193344 | PR00 | Immunosuppressants | ruxolitinib 15 MG |
| 1193346 | PR00 | Immunosuppressants | ruxolitinib 20 MG |
| 1193347 | PR00 | Immunosuppressants | ruxolitinib 20 MG |
| 1193348 | PR00 | Immunosuppressants | ruxolitinib 20 MG |
| 1193350 | PR00 | Immunosuppressants | ruxolitinib 25 MG |
| 1193351 | PR00 | Immunosuppressants | ruxolitinib 25 MG |
| 1193352 | PR00 | Immunosuppressants | ruxolitinib 25 MG |
| 1359534 | PR00 | Immunosuppressants | ustekinumab 90 MG/ML |
| 1359943 | PR00 | Immunosuppressants | ustekinumab 90 MG/ML |
| 1442690 | PR00 | Immunosuppressants | obinutuzumab 25 MG/ML |
| 1442691 | PR00 | Immunosuppressants | obinutuzumab Injectable Product |
| 1442692 | PR00 | Immunosuppressants | obinutuzumab Injectable Solution |
| 1442693 | PR00 | Immunosuppressants | obinutuzumab 25 MG/ML |
| 1442695 | PR00 | Immunosuppressants | obinutuzumab 25 MG/ML |
| 1442696 | PR00 | Immunosuppressants | obinutuzumab Injectable Solution |
| 1654076 | PR00 | Immunosuppressants | ustekinumab Injection |
| 1654077 | PR00 | Immunosuppressants | ustekinumab 90 MG/ML |
| 1654078 | PR00 | Immunosuppressants | ustekinumab Injection |
| 1654080 | PR00 | Immunosuppressants | ustekinumab 90 MG/ML |
| 1654081 | PR00 | Immunosuppressants | ustekinumab 90 MG/ML |
| 1654082 | PR00 | Immunosuppressants | ustekinumab 90 MG/ML |
| 1654083 | PR00 | Immunosuppressants | ustekinumab 90 MG/ML |
| 1656270 | PR00 | Immunosuppressants | natalizumab Injection |
| 1656272 | PR00 | Immunosuppressants | natalizumab Injection |
| 1656274 | PR00 | Immunosuppressants | natalizumab 20 MG/ML |
| 1656275 | PR00 | Immunosuppressants | natalizumab 20 MG/ML |
| 1656294 | PR00 | Immunosuppressants | obinutuzumab Injection |
| 1656296 | PR00 | Immunosuppressants | obinutuzumab Injection |
| 1656298 | PR00 | Immunosuppressants | obinutuzumab 25 MG/ML |
| 1656299 | PR00 | Immunosuppressants | obinutuzumab 25 MG/ML |
| 1720597 | PR00 | Immunosuppressants | mepolizumab |
| 1720598 | PR00 | Immunosuppressants | mepolizumab 100 MG |
| 1720599 | PR00 | Immunosuppressants | mepolizumab Injectable Product |
| 1720600 | PR00 | Immunosuppressants | mepolizumab Injection |
| 1720601 | PR00 | Immunosuppressants | mepolizumab 100 MG |
| 1720603 | PR00 | Immunosuppressants | mepolizumab 100 MG |
| 1720604 | PR00 | Immunosuppressants | mepolizumab Injection |
| 1745099 | PR00 | Immunosuppressants | ixekizumab |
| 1745100 | PR00 | Immunosuppressants | ixekizumab 80 MG/ML |
| 1745101 | PR00 | Immunosuppressants | ixekizumab Injectable Product |
| 1745102 | PR00 | Immunosuppressants | ixekizumab Auto-Injector |
| 1745103 | PR00 | Immunosuppressants | ixekizumab 80 MG/ML |
| 1745105 | PR00 | Immunosuppressants | ixekizumab 80 MG/ML |
| 1745106 | PR00 | Immunosuppressants | ixekizumab Auto-Injector |
| 1745109 | PR00 | Immunosuppressants | ixekizumab 80 MG/ML |
| 1745110 | PR00 | Immunosuppressants | ixekizumab 80 MG/ML |
| 1745111 | PR00 | Immunosuppressants | ixekizumab Prefilled Syringe |
| 1745112 | PR00 | Immunosuppressants | ixekizumab 80 MG/ML |
| 1745113 | PR00 | Immunosuppressants | ixekizumab Prefilled Syringe |
| 1745115 | PR00 | Immunosuppressants | ixekizumab 80 MG/ML |
| 1745116 | PR00 | Immunosuppressants | ixekizumab 80 MG/ML |
| 1811254 | PR00 | Immunosuppressants | ustekinumab 5 MG/ML |
| 1811255 | PR00 | Immunosuppressants | ustekinumab 5 MG/ML |
| 1811256 | PR00 | Immunosuppressants | ustekinumab 5 MG/ML |
| 1811258 | PR00 | Immunosuppressants | ustekinumab 5 MG/ML |
| 1811259 | PR00 | Immunosuppressants | ustekinumab 5 MG/ML |
| 1872251 | PR00 | Immunosuppressants | brodalumab |
| 1872253 | PR00 | Immunosuppressants | brodalumab 140 MG/ML |
| 1872254 | PR00 | Immunosuppressants | brodalumab Injectable Product |
| 1872255 | PR00 | Immunosuppressants | brodalumab Prefilled Syringe |
| 1872256 | PR00 | Immunosuppressants | brodalumab 140 MG/ML |
| 1872258 | PR00 | Immunosuppressants | brodalumab 140 MG/ML |
| 1872259 | PR00 | Immunosuppressants | brodalumab Prefilled Syringe |
| 1872262 | PR00 | Immunosuppressants | brodalumab 140 MG/ML |
| 1872263 | PR00 | Immunosuppressants | brodalumab 140 MG/ML |
| 1876376 | PR00 | Immunosuppressants | dupilumab |
| 1876398 | PR00 | Immunosuppressants | dupilumab 150 MG/ML |
| 1876399 | PR00 | Immunosuppressants | dupilumab Injectable Product |
| 1876400 | PR00 | Immunosuppressants | dupilumab Prefilled Syringe |
| 1876401 | PR00 | Immunosuppressants | dupilumab 150 MG/ML |

| | | | |
|---|---|---|---|
| 1876403 | PR00 | Immunosuppressants | dupilumab 150 MG/ML |
| 1876404 | PR00 | Immunosuppressants | dupilumab Prefilled Syringe |
| 1876407 | PR00 | Immunosuppressants | dupilumab 150 MG/ML |
| 1876408 | PR00 | Immunosuppressants | dupilumab 150 MG/ML |
| 1923319 | PR00 | Immunosuppressants | sarilumab |
| 1923330 | PR00 | Immunosuppressants | sarilumab 132 MG/ML |
| 1923331 | PR00 | Immunosuppressants | sarilumab Injectable Product |
| 1923332 | PR00 | Immunosuppressants | sarilumab Prefilled Syringe |
| 1923333 | PR00 | Immunosuppressants | sarilumab 132 MG/ML |
| 1923335 | PR00 | Immunosuppressants | sarilumab 132 MG/ML |
| 1923336 | PR00 | Immunosuppressants | sarilumab Prefilled Syringe |
| 1923339 | PR00 | Immunosuppressants | sarilumab 132 MG/ML |
| 1923340 | PR00 | Immunosuppressants | sarilumab 132 MG/ML |
| 1923344 | PR00 | Immunosuppressants | sarilumab 175 MG/ML |
| 1923345 | PR00 | Immunosuppressants | sarilumab 175 MG/ML |
| 1923346 | PR00 | Immunosuppressants | sarilumab 175 MG/ML |
| 1923348 | PR00 | Immunosuppressants | sarilumab 175 MG/ML |
| 1923349 | PR00 | Immunosuppressants | sarilumab 175 MG/ML |
| 1928588 | PR00 | Immunosuppressants | guselkumab |
| 1928589 | PR00 | Immunosuppressants | guselkumab 100 MG/ML |
| 1928590 | PR00 | Immunosuppressants | guselkumab Injectable Product |
| 1928594 | PR00 | Immunosuppressants | guselkumab 100 MG/ML |
| 1928685 | PR00 | Immunosuppressants | guselkumab Prefilled Syringe |
| 1928686 | PR00 | Immunosuppressants | guselkumab 100 MG/ML |
| 1928687 | PR00 | Immunosuppressants | guselkumab Prefilled Syringe |
| 1928690 | PR00 | Immunosuppressants | guselkumab 100 MG/ML |
| 1928691 | PR00 | Immunosuppressants | guselkumab 100 MG/ML |
| 1942950 | PR00 | Immunosuppressants | inotuzumab ozogamicin |
| 1942951 | PR00 | Immunosuppressants | inotuzumab ozogamicin 0.9 MG |
| 1942952 | PR00 | Immunosuppressants | inotuzumab ozogamicin Injectable Product |
| 1942953 | PR00 | Immunosuppressants | inotuzumab ozogamicin Injection |
| 1942954 | PR00 | Immunosuppressants | inotuzumab ozogamicin 0.9 MG |
| 1942956 | PR00 | Immunosuppressants | inotuzumab ozogamicin 0.9 MG |
| 1942957 | PR00 | Immunosuppressants | inotuzumab ozogamicin Injection |
| 1989100 | PR00 | Immunosuppressants | benralizumab |
| 1989101 | PR00 | Immunosuppressants | benralizumab 30 MG/ML |
| 1989102 | PR00 | Immunosuppressants | benralizumab Injectable Product |
| 1989103 | PR00 | Immunosuppressants | benralizumab Prefilled Syringe |
| 1989104 | PR00 | Immunosuppressants | benralizumab 30 MG/ML |
| 1989106 | PR00 | Immunosuppressants | benralizumab 30 MG/ML |
| 1989107 | PR00 | Immunosuppressants | benralizumab Prefilled Syringe |
| 1989110 | PR00 | Immunosuppressants | benralizumab 30 MG/ML |
| 1989111 | PR00 | Immunosuppressants | benralizumab 30 MG/ML |
| 2003754 | PR00 | Immunosuppressants | sarilumab Auto-Injector |
| 2003755 | PR00 | Immunosuppressants | sarilumab 132 MG/ML |
| 2003756 | PR00 | Immunosuppressants | sarilumab Auto-Injector |
| 2003758 | PR00 | Immunosuppressants | sarilumab 132 MG/ML |
| 2003759 | PR00 | Immunosuppressants | sarilumab 132 MG/ML |
| 2003766 | PR00 | Immunosuppressants | sarilumab 175 MG/ML |
| 2003768 | PR00 | Immunosuppressants | sarilumab 175 MG/ML |
| 2003769 | PR00 | Immunosuppressants | sarilumab 175 MG/ML |
| 2045490 | PR00 | Immunosuppressants | fingolimod 0.25 MG |
| 2045491 | PR00 | Immunosuppressants | fingolimod 0.25 MG |
| 2045492 | PR00 | Immunosuppressants | fingolimod 0.25 MG |
| 2099299 | PR00 | Immunosuppressants | dupilumab 200 MG/ML |
| 2099300 | PR00 | Immunosuppressants | dupilumab 175 MG/ML |
| 2099301 | PR00 | Immunosuppressants | dupilumab 200 MG/ML |
| 2099303 | PR00 | Immunosuppressants | dupilumab 200 MG/ML |
| 2099304 | PR00 | Immunosuppressants | dupilumab 200 MG/ML |
| 2101124 | PR00 | Immunosuppressants | dupilumab 175 MG/ML |
| 2101126 | PR00 | Immunosuppressants | dupilumab 175 MG/ML |
| 2101128 | PR00 | Immunosuppressants | dupilumab 175 MG/ML |
| 2101129 | PR00 | Immunosuppressants | dupilumab 175 MG/ML |
| 2118738 | PR00 | Immunosuppressants | guselkumab Auto-Injector |
| 2118739 | PR00 | Immunosuppressants | guselkumab 100 MG/ML |
| 2118740 | PR00 | Immunosuppressants | guselkumab Auto-Injector |
| 2118742 | PR00 | Immunosuppressants | guselkumab 100 MG/ML |
| 2118743 | PR00 | Immunosuppressants | guselkumab 100 MG/ML |
| 2170988 | PR00 | Immunosuppressants | mepolizumab 100 MG/ML |
| 2170989 | PR00 | Immunosuppressants | mepolizumab Auto-Injector |
| 2170990 | PR00 | Immunosuppressants | mepolizumab 100 MG/ML |

| | | | |
|---|---|---|---|
| 2170991 | PR00 | Immunosuppressants | mepolizumab 100 MG/ML |
| 2170992 | PR00 | Immunosuppressants | mepolizumab Auto-Injector |
| 2170994 | PR00 | Immunosuppressants | mepolizumab 100 MG/ML |
| 2170995 | PR00 | Immunosuppressants | mepolizumab 100 MG/ML |
| 2173819 | PR00 | Immunosuppressants | mepolizumab Prefilled Syringe |
| 2173820 | PR00 | Immunosuppressants | mepolizumab 100 MG/ML |
| 2173821 | PR00 | Immunosuppressants | mepolizumab Prefilled Syringe |
| 2173823 | PR00 | Immunosuppressants | mepolizumab 100 MG/ML |
| 2173824 | PR00 | Immunosuppressants | mepolizumab 100 MG/ML |
| 2196092 | PR00 | Immunosuppressants | upadacitinib |
| 2196890 | PR00 | Immunosuppressants | upadacitinib 15 MG |
| 2196891 | PR00 | Immunosuppressants | upadacitinib Oral Product |
| 2196892 | PR00 | Immunosuppressants | upadacitinib Pill |
| 2196893 | PR00 | Immunosuppressants | upadacitinib Extended Release Oral Tablet |
| 2196894 | PR00 | Immunosuppressants | upadacitinib 15 MG |
| 2196896 | PR00 | Immunosuppressants | upadacitinib 15 MG |
| 2196897 | PR00 | Immunosuppressants | upadacitinib Extended Release Oral Tablet |
| 2196901 | PR00 | Immunosuppressants | upadacitinib 15 MG |
| 2196902 | PR00 | Immunosuppressants | upadacitinib 15 MG |
| 262323 | PR00 | Immunosuppressants | ibritumomab tiuxetan |
| 354770 | PR00 | Immunosuppressants | natalizumab |
| 356988 | PR00 | Immunosuppressants | efalizumab |
| 402239 | PR00 | Immunosuppressants | efalizumab 100 MG/ML |
| 402350 | PR00 | Immunosuppressants | efalizumab 100 MG/ML |
| 402351 | PR00 | Immunosuppressants | efalizumab Injectable Solution |
| 402369 | PR00 | Immunosuppressants | efalizumab Injectable Solution |
| 477484 | PR00 | Immunosuppressants | natalizumab 20 MG/ML |
| 477485 | PR00 | Immunosuppressants | natalizumab 20 MG/ML |
| 477487 | PR00 | Immunosuppressants | natalizumab Injectable Solution |
| 576113 | PR00 | Immunosuppressants | efalizumab 100 MG/ML |
| 603539 | PR00 | Immunosuppressants | natalizumab 20 MG/ML |
| 603540 | PR00 | Immunosuppressants | natalizumab Injectable Solution |
| 847083 | PR00 | Immunosuppressants | ustekinumab |
| 853348 | PR00 | Immunosuppressants | ustekinumab 90 MG/ML |
| 853349 | PR00 | Immunosuppressants | ustekinumab Prefilled Syringe |
| 853350 | PR00 | Immunosuppressants | ustekinumab 90 MG/ML |
| 853352 | PR00 | Immunosuppressants | ustekinumab 90 MG/ML |
| 853353 | PR00 | Immunosuppressants | ustekinumab Prefilled Syringe |
| 853355 | PR00 | Immunosuppressants | ustekinumab 90 MG/ML |
| 865171 | PR00 | Immunosuppressants | ustekinumab Injectable Solution |
| 865172 | PR00 | Immunosuppressants | ustekinumab 90 MG/ML |
| 865173 | PR00 | Immunosuppressants | ustekinumab Injectable Solution |
| 974779 | PR00 | Immunosuppressants | obinutuzumab |
| 1170524 | PR00 | Immunosuppressants | Gilenya Oral Product |
| 1170525 | PR00 | Immunosuppressants | Gilenya Pill |
| 1178283 | PR00 | Immunosuppressants | Raptiva Injectable Product |
| 1178973 | PR00 | Immunosuppressants | Stelara Injectable Product |
| 1187514 | PR00 | Immunosuppressants | Tysabri Injectable Product |
| 1193335 | PR00 | Immunosuppressants | Jakafi Oral Product |
| 1193336 | PR00 | Immunosuppressants | Jakafi Pill |
| 1366322 | PR00 | Immunosuppressants | Ceratonia siliqua whole extract |
| 1442697 | PR00 | Immunosuppressants | Gazyva Injectable Product |
| 1720605 | PR00 | Immunosuppressants | Nucala Injectable Product |
| 1745107 | PR00 | Immunosuppressants | Taltz Injectable Product |
| 1872260 | PR00 | Immunosuppressants | Siliq Injectable Product |
| 1876405 | PR00 | Immunosuppressants | Dupixent Injectable Product |
| 1923337 | PR00 | Immunosuppressants | Kevzara Injectable Product |
| 1928596 | PR00 | Immunosuppressants | Tremfya Injectable Product |
| 1942958 | PR00 | Immunosuppressants | Besponsa Injectable Product |
| 1989108 | PR00 | Immunosuppressants | Fasenra Injectable Product |
| 2047245 | PR00 | Immunosuppressants | Olumiant Oral Product |
| 2047246 | PR00 | Immunosuppressants | Olumiant Pill |
| 2196898 | PR00 | Immunosuppressants | Rinvoq Oral Product |
| 2196899 | PR00 | Immunosuppressants | Rinvoq Pill |
| 1182522 | PR00 | Immunosuppressants | Revlimid Oral Product |
| 1182529 | PR00 | Immunosuppressants | Revlimid Pill |
| 1242232 | PR00 | Immunosuppressants | lenalidomide 2.5 MG |
| 1242233 | PR00 | Immunosuppressants | lenalidomide 2.5 MG |
| 1429366 | PR00 | Immunosuppressants | lenalidomide 20 MG |
| 1429367 | PR00 | Immunosuppressants | lenalidomide 20 MG |
| 337535 | PR00 | Immunosuppressants | (NA) |

| | | | |
|---|---|---|---|
| 616112 | PR00 | Immunosuppressants | lenalidomide 10 MG |
| 616113 | PR00 | Immunosuppressants | lenalidomide Oral Capsule |
| 616114 | PR00 | Immunosuppressants | lenalidomide 10 MG |
| 616115 | PR00 | Immunosuppressants | lenalidomide 5 MG |
| 616116 | PR00 | Immunosuppressants | lenalidomide 5 MG |
| 643713 | PR00 | Immunosuppressants | lenalidomide 15 MG |
| 643714 | PR00 | Immunosuppressants | lenalidomide 15 MG |
| 643721 | PR00 | Immunosuppressants | lenalidomide 25 MG |
| 643722 | PR00 | Immunosuppressants | lenalidomide 25 MG |
| 10432 | PR00 | Immunosuppressants | Thalidomide |
| 1164873 | PR00 | Immunosuppressants | Thalidomide Oral Product |
| 1164874 | PR00 | Immunosuppressants | Thalidomide Pill |
| 200390 | PR00 | Immunosuppressants | Thalidomide 50 MG |
| 213360 | PR00 | Immunosuppressants | Thalidomide 50 MG |
| 316784 | PR00 | Immunosuppressants | Thalidomide 50 MG |
| 360176 | PR00 | Immunosuppressants | Thalidomide 100 MG |
| 360177 | PR00 | Immunosuppressants | Thalidomide 200 MG |
| 360321 | PR00 | Immunosuppressants | Thalidomide 100 MG |
| 360322 | PR00 | Immunosuppressants | Thalidomide 200 MG |
| 366451 | PR00 | Immunosuppressants | Thalidomide Oral Capsule |
| 374072 | PR00 | Immunosuppressants | Thalidomide Oral Capsule |
| 404449 | PR00 | Immunosuppressants | Thalidomide 100 MG |
| 404450 | PR00 | Immunosuppressants | Thalidomide 200 MG |
| 432527 | PR00 | Immunosuppressants | Thalidomide 100 MG |
| 452059 | PR00 | Immunosuppressants | Thalidomide Oral Tablet |
| 573254 | PR00 | Immunosuppressants | Thalidomide 50 MG |
| 576406 | PR00 | Immunosuppressants | Thalidomide 100 MG |
| 576407 | PR00 | Immunosuppressants | Thalidomide 200 MG |
| 700415 | PR00 | Immunosuppressants | Thalidomide 150 MG |
| 700416 | PR00 | Immunosuppressants | Thalidomide 150 MG |
| 700417 | PR00 | Immunosuppressants | Thalidomide 150 MG |
| 700418 | PR00 | Immunosuppressants | Thalidomide 150 MG |
| 107050 | PR00 | Immunosuppressants | Rabbit anti-human T-lymphocyte globulin 25 MG |
| 1177887 | PR00 | Immunosuppressants | Thymoglobulin Injectable Product |
| 1870637 | PR00 | Immunosuppressants | Rabbit anti-human T-lymphocyte globulin 25 MG |
| 1870638 | PR00 | Immunosuppressants | Rabbit anti-human T-lymphocyte globulin Injection |
| 225741 | PR00 | Immunosuppressants | (NA) |
| 363507 | PR00 | Immunosuppressants | Rabbit anti-human T-lymphocyte globulin Injectable Solution |
| 564411 | PR00 | Immunosuppressants | Rabbit anti-human T-lymphocyte globulin 5 MG/ML |
| 616011 | MA00 | Immunosuppressants | ORENCIA |
| 607377 | MA00 | Immunosuppressants | ABATACEPT |
| 614391 | MA00 | Immunosuppressants | ABATACEPT |
| 1145932 | MA00 | Immunosuppressants | 1 ML ABATACEPT 125 MG/ML PREFILLED SYRINGE [ORENCIA] |
| 1359480 | MA00 | Immunosuppressants | ABATACEPT 125 MG/ML PREFILLED SYRINGE [ORENCIA] |
| 1799230 | MA00 | Immunosuppressants | 1 ML ABATACEPT 125 MG/ML AUTO-INJECTOR [ORENCIA] |
| 1799232 | MA00 | Immunosuppressants | ABATACEPT 125 MG/ML AUTO-INJECTOR [ORENCIA] |
| 1925255 | MA00 | Immunosuppressants | 0.4 ML ABATACEPT 125 MG/ML PREFILLED SYRINGE [ORENCIA] |
| 1925257 | MA00 | Immunosuppressants | 0.7 ML ABATACEPT 125 MG/ML PREFILLED SYRINGE [ORENCIA] |
| 616018 | MA00 | Immunosuppressants | ABATACEPT 250 MG INJECTION [ORENCIA] |
| 616207 | MA00 | Immunosuppressants | ABATACEPT 10 MG/ML INJECTABLE SOLUTION [ORENCIA] |
| 1145930 | MA00 | Immunosuppressants | ABATACEPT 125 MG/ML [ORENCIA] |
| 1659774 | MA00 | Immunosuppressants | ABATACEPT 250 MG [ORENCIA] |
| 616016 | MA00 | Immunosuppressants | ABATACEPT 25 MG/ML [ORENCIA] |
| 616206 | MA00 | Immunosuppressants | ABATACEPT 10 MG/ML [ORENCIA] |
| 1145931 | MA00 | Immunosuppressants | ABATACEPT PREFILLED SYRINGE [ORENCIA] |
| 1659775 | MA00 | Immunosuppressants | ABATACEPT INJECTION [ORENCIA] |
| 1799229 | MA00 | Immunosuppressants | ABATACEPT AUTO-INJECTOR [ORENCIA] |
| 616017 | MA00 | Immunosuppressants | ABATACEPT INJECTABLE SOLUTION [ORENCIA] |
| 1183963 | MA00 | Immunosuppressants | ORENCIA INJECTABLE PRODUCT |
| 1145929 | MA00 | Immunosuppressants | 1 ML ABATACEPT 125 MG/ML PREFILLED SYRINGE |
| 1360514 | MA00 | Immunosuppressants | ABATACEPT 125 MG/ML PREFILLED SYRINGE |
| 1799228 | MA00 | Immunosuppressants | 1 ML ABATACEPT 125 MG/ML AUTO-INJECTOR |
| 1799231 | MA00 | Immunosuppressants | ABATACEPT 125 MG/ML AUTO-INJECTOR |
| 1925254 | MA00 | Immunosuppressants | 0.4 ML ABATACEPT 125 MG/ML PREFILLED SYRINGE |
| 1925256 | MA00 | Immunosuppressants | 0.7 ML ABATACEPT 125 MG/ML PREFILLED SYRINGE |
| 607381 | MA00 | Immunosuppressants | ABATACEPT 10 MG/ML INJECTABLE SOLUTION |
| 616015 | MA00 | Immunosuppressants | ABATACEPT 250 MG INJECTION |
| 1145927 | MA00 | Immunosuppressants | ABATACEPT 125 MG/ML |
| 1659771 | MA00 | Immunosuppressants | ABATACEPT 250 MG |
| 607379 | MA00 | Immunosuppressants | ABATACEPT 10 MG/ML |
| 616014 | MA00 | Immunosuppressants | ABATACEPT 25 MG/ML |

| | | | |
|---|---|---|---|
| 1145928 | MA00 | Immunosuppressants | ABATACEPT PREFILLED SYRINGE |
| 1659772 | MA00 | Immunosuppressants | ABATACEPT INJECTION |
| 1799227 | MA00 | Immunosuppressants | ABATACEPT AUTO-INJECTOR |
| 607380 | MA00 | Immunosuppressants | ABATACEPT INJECTABLE SOLUTION |
| 1163066 | MA00 | Immunosuppressants | ABATACEPT INJECTABLE PRODUCT |
| 353484 | MA00 | Immunosuppressants | HUMIRA |
| 1594352 | MA00 | Immunosuppressants | {6 (0.8 ML ADALIMUMAB 50 MG/ML PREFILLED SYRINGE [HUMIRA]) } PACK [HUMIRA PEDIATRIC CROHN'S DISEASE STARTER PACKAGE (6 COUNT)] |
| 1594358 | MA00 | Immunosuppressants | {3 (0.8 ML ADALIMUMAB 50 MG/ML PREFILLED SYRINGE [HUMIRA]) } PACK [HUMIRA PEDIATRIC CROHN'S DISEASE STARTER PACKAGE (3 COUNT)] |
| 1801278 | MA00 | Immunosuppressants | {4 (0.8 ML ADALIMUMAB 50 MG/ML AUTO-INJECTOR [HUMIRA]) } PACK [HUMIRA PEN - PSORIASIS STARTER PACK] |
| 1855527 | MA00 | Immunosuppressants | {1 (0.4 ML ADALIMUMAB 100 MG/ML PREFILLED SYRINGE [HUMIRA]) / 1 (0.8 ML ADALIMUMAB 100 MG/ML PREFILLED SYRINGE [HUMIRA]) } PACK [HUMIRA PEDIATRIC CROHN'S DISEASE STARTER PACKAGE (2 COUNT)] |
| 1872984 | MA00 | Immunosuppressants | {6 (0.4 ML ADALIMUMAB 100 MG/ML AUTO-INJECTOR [HUMIRA]) } PACK [HUMIRA PEN 40 MG/0.4 ML STARTER PACK - CROHN'S DISEASE] |
| 1872986 | MA00 | Immunosuppressants | {4 (0.4 ML ADALIMUMAB 100 MG/ML AUTO-INJECTOR [HUMIRA]) } PACK [HUMIRA PEN 40 MG/0.4 ML STARTER PACK - PSORIASIS] |
| 1873087 | MA00 | Immunosuppressants | {3 (0.8 ML ADALIMUMAB 100 MG/ML PREFILLED SYRINGE [HUMIRA]) } PACK [HUMIRA PREFILLED SYRINGE 80 MG/0.8 ML STARTER PACK - PEDIATRIC CROHN'S DISEASE] |
| 1921245 | MA00 | Immunosuppressants | {3 (0.8 ML ADALIMUMAB 100 MG/ML AUTO-INJECTOR [HUMIRA]) } PACK [HUMIRA PEN 80 MG/0.8 ML - STARTER PACKAGE FOR CROHN'S DISEASE, ULCERATIVE COLITIS OR HIDRADENITIS SUPPURATIVA] |
| 1921468 | MA00 | Immunosuppressants | {2 (0.4 ML ADALIMUMAB 100 MG/ML AUTO-INJECTOR [HUMIRA]) / 1 (0.8 ML ADALIMUMAB 100 MG/ML AUTO-INJECTOR [HUMIRA]) } PACK [HUMIRA PEN 80 MG/0.8 ML AND 40 MG/0.4 ML - PSORIASIS/UVEITIS STARTER PACKAGE] |
| 825170 | MA00 | Immunosuppressants | {6 (0.8 ML ADALIMUMAB 50 MG/ML AUTO-INJECTOR [HUMIRA]) } PACK [HUMIRA PEN - CROHN'S DISEASE STARTER PACK] |
| 1594357 | MA00 | Immunosuppressants | {3 (0.8 ML ADALIMUMAB 50 MG/ML PREFILLED SYRINGE) } PACK |
| 1656703 | MA00 | Immunosuppressants | {6 (0.8 ML ADALIMUMAB 50 MG/ML AUTO-INJECTOR) } PACK |
| 1801277 | MA00 | Immunosuppressants | {4 (0.8 ML ADALIMUMAB 50 MG/ML AUTO-INJECTOR) } PACK |
| 1855526 | MA00 | Immunosuppressants | {1 (0.4 ML ADALIMUMAB 100 MG/ML PREFILLED SYRINGE) / 1 (0.8 ML ADALIMUMAB 100 MG/ML PREFILLED SYRINGE) } PACK |
| 1872983 | MA00 | Immunosuppressants | {6 (0.4 ML ADALIMUMAB 100 MG/ML AUTO-INJECTOR) } PACK |
| 1872985 | MA00 | Immunosuppressants | {4 (0.4 ML ADALIMUMAB 100 MG/ML AUTO-INJECTOR) } PACK |
| 1873086 | MA00 | Immunosuppressants | {3 (0.8 ML ADALIMUMAB 100 MG/ML PREFILLED SYRINGE) } PACK |
| 1921244 | MA00 | Immunosuppressants | {3 (0.8 ML ADALIMUMAB 100 MG/ML AUTO-INJECTOR) } PACK |
| 1921467 | MA00 | Immunosuppressants | {2 (0.4 ML ADALIMUMAB 100 MG/ML AUTO-INJECTOR) / 1 (0.8 ML ADALIMUMAB 100 MG/ML AUTO-INJECTOR) } PACK |
| 825169 | MA00 | Immunosuppressants | {6 (0.8 ML ADALIMUMAB 50 MG/ML PREFILLED SYRINGE) } PACK |
| 327361 | MA00 | Immunosuppressants | ADALIMUMAB |
| 2103477 | MA00 | Immunosuppressants | ADALIMUMAB-ADBM |
| 2103479 | MA00 | Immunosuppressants | ADALIMUMAB-ATTO |
| 1359755 | MA00 | Immunosuppressants | ADALIMUMAB 50 MG/ML PREFILLED SYRINGE [HUMIRA] |
| 1551888 | MA00 | Immunosuppressants | 0.2 ML ADALIMUMAB 50 MG/ML PREFILLED SYRINGE [HUMIRA] |
| 1655728 | MA00 | Immunosuppressants | 0.8 ML ADALIMUMAB 50 MG/ML AUTO-INJECTOR [HUMIRA] |
| 1655730 | MA00 | Immunosuppressants | ADALIMUMAB 50 MG/ML AUTO-INJECTOR [HUMIRA] |
| 1658078 | MA00 | Immunosuppressants | ADALIMUMAB 50 MG/ML INJECTION [HUMIRA] |
| 1726846 | MA00 | Immunosuppressants | 0.4 ML ADALIMUMAB 100 MG/ML PREFILLED SYRINGE [HUMIRA] |
| 1726848 | MA00 | Immunosuppressants | ADALIMUMAB 100 MG/ML PREFILLED SYRINGE [HUMIRA] |
| 1855524 | MA00 | Immunosuppressants | 0.8 ML ADALIMUMAB 100 MG/ML PREFILLED SYRINGE [HUMIRA] |
| 1872980 | MA00 | Immunosuppressants | 0.4 ML ADALIMUMAB 100 MG/ML AUTO-INJECTOR [HUMIRA] |
| 1872982 | MA00 | Immunosuppressants | ADALIMUMAB 100 MG/ML AUTO-INJECTOR [HUMIRA] |
| 1921010 | MA00 | Immunosuppressants | 0.1 ML ADALIMUMAB 100 MG/ML PREFILLED SYRINGE [HUMIRA] |
| 1921017 | MA00 | Immunosuppressants | 0.2 ML ADALIMUMAB 100 MG/ML PREFILLED SYRINGE [HUMIRA] |
| 1921240 | MA00 | Immunosuppressants | 0.8 ML ADALIMUMAB 100 MG/ML AUTO-INJECTOR [HUMIRA] |
| 1921242 | MA00 | Immunosuppressants | ADALIMUMAB 100 MG/ML PEN INJECTOR [HUMIRA] |
| 1921466 | MA00 | Immunosuppressants | 0.4 ML ADALIMUMAB 100 MG/ML PEN INJECTOR [HUMIRA] |
| 352334 | MA00 | Immunosuppressants | 0.8 ML ADALIMUMAB 50 MG/ML INJECTION [HUMIRA] |
| 727705 | MA00 | Immunosuppressants | 0.8 ML ADALIMUMAB 50 MG/ML PREFILLED SYRINGE [HUMIRA] |
| 763565 | MA00 | Immunosuppressants | 0.4 ML ADALIMUMAB 50 MG/ML PREFILLED SYRINGE [HUMIRA] |
| 1726845 | MA00 | Immunosuppressants | ADALIMUMAB 100 MG/ML [HUMIRA] |
| 576023 | MA00 | Immunosuppressants | ADALIMUMAB 50 MG/ML [HUMIRA] |
| 1655727 | MA00 | Immunosuppressants | ADALIMUMAB AUTO-INJECTOR [HUMIRA] |
| 1656698 | MA00 | Immunosuppressants | ADALIMUMAB INJECTION [HUMIRA] |
| 1921239 | MA00 | Immunosuppressants | ADALIMUMAB PEN INJECTOR [HUMIRA] |
| 363562 | MA00 | Immunosuppressants | ADALIMUMAB INJECTABLE SOLUTION [HUMIRA] |
| 727704 | MA00 | Immunosuppressants | ADALIMUMAB PREFILLED SYRINGE [HUMIRA] |
| 1171992 | MA00 | Immunosuppressants | HUMIRA INJECTABLE PRODUCT |
| 1360097 | MA00 | Immunosuppressants | ADALIMUMAB 50 MG/ML PREFILLED SYRINGE |

| | | | |
|---|---|---|---|
| 1551887 | MA00 | Immunosuppressants | 0.2 ML ADALIMUMAB 50 MG/ML PREFILLED SYRINGE |
| 1655726 | MA00 | Immunosuppressants | 0.8 ML ADALIMUMAB 50 MG/ML AUTO-INJECTOR |
| 1655729 | MA00 | Immunosuppressants | ADALIMUMAB 50 MG/ML AUTO-INJECTOR |
| 1658077 | MA00 | Immunosuppressants | ADALIMUMAB 50 MG/ML INJECTION |
| 1726844 | MA00 | Immunosuppressants | 0.4 ML ADALIMUMAB 100 MG/ML PREFILLED SYRINGE |
| 1726847 | MA00 | Immunosuppressants | ADALIMUMAB 100 MG/ML PREFILLED SYRINGE |
| 1855523 | MA00 | Immunosuppressants | 0.8 ML ADALIMUMAB 100 MG/ML PREFILLED SYRINGE |
| 1872979 | MA00 | Immunosuppressants | 0.4 ML ADALIMUMAB 100 MG/ML AUTO-INJECTOR |
| 1872981 | MA00 | Immunosuppressants | ADALIMUMAB 100 MG/ML AUTO-INJECTOR |
| 1921009 | MA00 | Immunosuppressants | 0.1 ML ADALIMUMAB 100 MG/ML PREFILLED SYRINGE |
| 1921016 | MA00 | Immunosuppressants | 0.2 ML ADALIMUMAB 100 MG/ML PREFILLED SYRINGE |
| 1921238 | MA00 | Immunosuppressants | 0.8 ML ADALIMUMAB 100 MG/ML AUTO-INJECTOR |
| 1921241 | MA00 | Immunosuppressants | ADALIMUMAB 100 MG/ML PEN INJECTOR |
| 1921465 | MA00 | Immunosuppressants | 0.4 ML ADALIMUMAB 100 MG/ML PEN INJECTOR |
| 351290 | MA00 | Immunosuppressants | 0.8 ML ADALIMUMAB 50 MG/ML INJECTION |
| 727703 | MA00 | Immunosuppressants | 0.8 ML ADALIMUMAB 50 MG/ML PREFILLED SYRINGE |
| 763564 | MA00 | Immunosuppressants | 0.4 ML ADALIMUMAB 50 MG/ML PREFILLED SYRINGE |
| 1726843 | MA00 | Immunosuppressants | ADALIMUMAB 100 MG/ML |
| 358817 | MA00 | Immunosuppressants | ADALIMUMAB 50 MG/ML |
| 1655725 | MA00 | Immunosuppressants | ADALIMUMAB AUTO-INJECTOR |
| 1656695 | MA00 | Immunosuppressants | ADALIMUMAB INJECTION |
| 1921237 | MA00 | Immunosuppressants | ADALIMUMAB PEN INJECTOR |
| 378758 | MA00 | Immunosuppressants | ADALIMUMAB INJECTABLE SOLUTION |
| 727702 | MA00 | Immunosuppressants | ADALIMUMAB PREFILLED SYRINGE |
| 1163691 | MA00 | Immunosuppressants | ADALIMUMAB INJECTABLE PRODUCT |
| 1594659 | MA00 | Immunosuppressants | LEMTRADA |
| 284679 | MA00 | Immunosuppressants | CAMPATH |
| 117055 | MA00 | Immunosuppressants | ALEMTUZUMAB |
| 1594663 | MA00 | Immunosuppressants | 1.2 ML ALEMTUZUMAB 10 MG/ML INJECTION [LEMTRADA] |
| 1656635 | MA00 | Immunosuppressants | ALEMTUZUMAB 30 MG/ML INJECTION [CAMPATH] |
| 1656640 | MA00 | Immunosuppressants | ALEMTUZUMAB 10 MG/ML INJECTION [LEMTRADA] |
| 351967 | MA00 | Immunosuppressants | ALEMTUZUMAB 10 MG/ML INJECTABLE SOLUTION [CAMPATH] |
| 543438 | MA00 | Immunosuppressants | ALEMTUZUMAB 30 MG/ML INJECTABLE SOLUTION [CAMPATH] |
| 828267 | MA00 | Immunosuppressants | 1 ML ALEMTUZUMAB 30 MG/ML INJECTION [CAMPATH] |
| 1594660 | MA00 | Immunosuppressants | ALEMTUZUMAB 10 MG/ML [LEMTRADA] |
| 1656631 | MA00 | Immunosuppressants | ALEMTUZUMAB 30 MG/ML [CAMPATH] |
| 543437 | MA00 | Immunosuppressants | ALEMTUZUMAB 30 MG/ML [CAMPATH] |
| 575719 | MA00 | Immunosuppressants | ALEMTUZUMAB 10 MG/ML [CAMPATH] |
| 828266 | MA00 | Immunosuppressants | ALEMTUZUMAB 0.3 MG/ML [CAMPATH] |
| 1594661 | MA00 | Immunosuppressants | ALEMTUZUMAB INJECTABLE SOLUTION [LEMTRADA] |
| 1656632 | MA00 | Immunosuppressants | ALEMTUZUMAB INJECTION [CAMPATH] |
| 1656637 | MA00 | Immunosuppressants | ALEMTUZUMAB INJECTION [LEMTRADA] |
| 362780 | MA00 | Immunosuppressants | ALEMTUZUMAB INJECTABLE SOLUTION [CAMPATH] |
| 1172298 | MA00 | Immunosuppressants | CAMPATH INJECTABLE PRODUCT |
| 1594662 | MA00 | Immunosuppressants | LEMTRADA INJECTABLE PRODUCT |
| 1594658 | MA00 | Immunosuppressants | 1.2 ML ALEMTUZUMAB 10 MG/ML INJECTION |
| 1656634 | MA00 | Immunosuppressants | ALEMTUZUMAB 30 MG/ML INJECTION |
| 1656639 | MA00 | Immunosuppressants | ALEMTUZUMAB 10 MG/ML INJECTION |
| 284064 | MA00 | Immunosuppressants | ALEMTUZUMAB 10 MG/ML INJECTABLE SOLUTION |
| 485516 | MA00 | Immunosuppressants | ALEMTUZUMAB 30 MG/ML INJECTABLE SOLUTION |
| 828265 | MA00 | Immunosuppressants | 1 ML ALEMTUZUMAB 30 MG/ML INJECTION |
| 1594657 | MA00 | Immunosuppressants | ALEMTUZUMAB 10 MG/ML |
| 1656628 | MA00 | Immunosuppressants | ALEMTUZUMAB 30 MG/ML |
| 343434 | MA00 | Immunosuppressants | ALEMTUZUMAB 10 MG/ML |
| 485515 | MA00 | Immunosuppressants | ALEMTUZUMAB 30 MG/ML |
| 828264 | MA00 | Immunosuppressants | ALEMTUZUMAB 0.3 MG/ML |
| 1656629 | MA00 | Immunosuppressants | ALEMTUZUMAB INJECTION |
| 378757 | MA00 | Immunosuppressants | ALEMTUZUMAB INJECTABLE SOLUTION |
| 1164265 | MA00 | Immunosuppressants | ALEMTUZUMAB INJECTABLE PRODUCT |
| 1112977 | MA00 | Immunosuppressants | NULOJIX |
| 1112973 | MA00 | Immunosuppressants | BELATACEPT |
| 1112980 | MA00 | Immunosuppressants | BELATACEPT 250 MG INJECTION [NULOJIX] |
| 1112978 | MA00 | Immunosuppressants | BELATACEPT 25 MG/ML [NULOJIX] |
| 1804973 | MA00 | Immunosuppressants | BELATACEPT 250 MG [NULOJIX] |
| 1112979 | MA00 | Immunosuppressants | BELATACEPT INJECTABLE SOLUTION [NULOJIX] |
| 1804974 | MA00 | Immunosuppressants | BELATACEPT INJECTION [NULOJIX] |
| 1178807 | MA00 | Immunosuppressants | NULOJIX INJECTABLE PRODUCT |
| 1112976 | MA00 | Immunosuppressants | BELATACEPT 250 MG INJECTION |
| 1112974 | MA00 | Immunosuppressants | BELATACEPT 25 MG/ML |
| 1804970 | MA00 | Immunosuppressants | BELATACEPT 250 MG |
| 1112975 | MA00 | Immunosuppressants | BELATACEPT INJECTABLE SOLUTION |

| | | | |
|---|---|---|---|
| 1804971 | MA00 | Immunosuppressants | BELATACEPT INJECTION |
| 1155316 | MA00 | Immunosuppressants | BELATACEPT INJECTABLE PRODUCT |
| 1092441 | MA00 | Immunosuppressants | BENLYSTA |
| 1092437 | MA00 | Immunosuppressants | BELIMUMAB |
| 1092444 | MA00 | Immunosuppressants | BELIMUMAB 80 MG/ML INJECTABLE SOLUTION [BENLYSTA] |
| 1656398 | MA00 | Immunosuppressants | BELIMUMAB 120 MG INJECTION [BENLYSTA] |
| 1656402 | MA00 | Immunosuppressants | BELIMUMAB 400 MG INJECTION [BENLYSTA] |
| 1939299 | MA00 | Immunosuppressants | 1 ML BELIMUMAB 200 MG/ML AUTO-INJECTOR [BENLYSTA] |
| 1939301 | MA00 | Immunosuppressants | BELIMUMAB 200 MG/ML AUTO-INJECTOR [BENLYSTA] |
| 1939348 | MA00 | Immunosuppressants | 1 ML BELIMUMAB 200 MG/ML PREFILLED SYRINGE [BENLYSTA] |
| 1939350 | MA00 | Immunosuppressants | BELIMUMAB 200 MG/ML PREFILLED SYRINGE [BENLYSTA] |
| 1092442 | MA00 | Immunosuppressants | BELIMUMAB 80 MG/ML [BENLYSTA] |
| 1656396 | MA00 | Immunosuppressants | BELIMUMAB 120 MG [BENLYSTA] |
| 1656401 | MA00 | Immunosuppressants | BELIMUMAB 400 MG [BENLYSTA] |
| 1939297 | MA00 | Immunosuppressants | BELIMUMAB 200 MG/ML [BENLYSTA] |
| 1092443 | MA00 | Immunosuppressants | BELIMUMAB INJECTABLE SOLUTION [BENLYSTA] |
| 1656397 | MA00 | Immunosuppressants | BELIMUMAB INJECTION [BENLYSTA] |
| 1939298 | MA00 | Immunosuppressants | BELIMUMAB AUTO-INJECTOR [BENLYSTA] |
| 1939347 | MA00 | Immunosuppressants | BELIMUMAB PREFILLED SYRINGE [BENLYSTA] |
| 1171464 | MA00 | Immunosuppressants | BENLYSTA INJECTABLE PRODUCT |
| 1092440 | MA00 | Immunosuppressants | BELIMUMAB 80 MG/ML INJECTABLE SOLUTION |
| 1656395 | MA00 | Immunosuppressants | BELIMUMAB 120 MG INJECTION |
| 1656400 | MA00 | Immunosuppressants | BELIMUMAB 400 MG INJECTION |
| 1939296 | MA00 | Immunosuppressants | 1 ML BELIMUMAB 200 MG/ML AUTO-INJECTOR |
| 1939300 | MA00 | Immunosuppressants | BELIMUMAB 200 MG/ML AUTO-INJECTOR |
| 1939346 | MA00 | Immunosuppressants | 1 ML BELIMUMAB 200 MG/ML PREFILLED SYRINGE |
| 1939349 | MA00 | Immunosuppressants | BELIMUMAB 200 MG/ML PREFILLED SYRINGE |
| 1092438 | MA00 | Immunosuppressants | BELIMUMAB 80 MG/ML |
| 1656393 | MA00 | Immunosuppressants | BELIMUMAB 120 MG |
| 1656399 | MA00 | Immunosuppressants | BELIMUMAB 400 MG |
| 1939294 | MA00 | Immunosuppressants | BELIMUMAB 200 MG/ML |
| 1092439 | MA00 | Immunosuppressants | BELIMUMAB INJECTABLE SOLUTION |
| 1656394 | MA00 | Immunosuppressants | BELIMUMAB INJECTION |
| 1939295 | MA00 | Immunosuppressants | BELIMUMAB AUTO-INJECTOR |
| 1939345 | MA00 | Immunosuppressants | BELIMUMAB PREFILLED SYRINGE |
| 1159402 | MA00 | Immunosuppressants | BELIMUMAB INJECTABLE PRODUCT |
| 853495 | MA00 | Immunosuppressants | ILARIS |
| 853491 | MA00 | Immunosuppressants | CANAKINUMAB |
| 1864326 | MA00 | Immunosuppressants | 1 ML CANAKINUMAB 150 MG/ML INJECTION [ILARIS] |
| 1864328 | MA00 | Immunosuppressants | CANAKINUMAB 150 MG/ML INJECTION [ILARIS] |
| 853498 | MA00 | Immunosuppressants | CANAKINUMAB 180 MG INJECTION [ILARIS] |
| 1655974 | MA00 | Immunosuppressants | CANAKINUMAB 180 MG [ILARIS] |
| 1864325 | MA00 | Immunosuppressants | CANAKINUMAB 150 MG/ML [ILARIS] |
| 853496 | MA00 | Immunosuppressants | CANAKINUMAB 150 MG/ML [ILARIS] |
| 1655975 | MA00 | Immunosuppressants | CANAKINUMAB INJECTION [ILARIS] |
| 853497 | MA00 | Immunosuppressants | CANAKINUMAB INJECTABLE SOLUTION [ILARIS] |
| 1167137 | MA00 | Immunosuppressants | ILARIS INJECTABLE PRODUCT |
| 1864324 | MA00 | Immunosuppressants | 1 ML CANAKINUMAB 150 MG/ML INJECTION |
| 1864327 | MA00 | Immunosuppressants | CANAKINUMAB 150 MG/ML INJECTION |
| 853494 | MA00 | Immunosuppressants | CANAKINUMAB 180 MG INJECTION |
| 1655971 | MA00 | Immunosuppressants | CANAKINUMAB 180 MG |
| 1864323 | MA00 | Immunosuppressants | CANAKINUMAB 150 MG/ML |
| 853492 | MA00 | Immunosuppressants | CANAKINUMAB 150 MG/ML |
| 1655972 | MA00 | Immunosuppressants | CANAKINUMAB INJECTION |
| 853493 | MA00 | Immunosuppressants | CANAKINUMAB INJECTABLE SOLUTION |
| 1156606 | MA00 | Immunosuppressants | CANAKINUMAB INJECTABLE PRODUCT |
| 795082 | MA00 | Immunosuppressants | CIMZIA |
| 709271 | MA00 | Immunosuppressants | CERTOLIZUMAB PEGOL |
| 1359935 | MA00 | Immunosuppressants | CERTOLIZUMAB PEGOL 200 MG/ML PREFILLED SYRINGE [CIMZIA] |
| 795085 | MA00 | Immunosuppressants | CERTOLIZUMAB PEGOL 200 MG INJECTION [CIMZIA] |
| 849599 | MA00 | Immunosuppressants | 1 ML CERTOLIZUMAB PEGOL 200 MG/ML PREFILLED SYRINGE [CIMZIA] |
| 1653730 | MA00 | Immunosuppressants | CERTOLIZUMAB PEGOL 200 MG [CIMZIA] |
| 795083 | MA00 | Immunosuppressants | CERTOLIZUMAB PEGOL 200 MG/ML [CIMZIA] |
| 1653731 | MA00 | Immunosuppressants | CERTOLIZUMAB PEGOL INJECTION [CIMZIA] |
| 795084 | MA00 | Immunosuppressants | CERTOLIZUMAB PEGOL INJECTABLE SOLUTION [CIMZIA] |
| 849598 | MA00 | Immunosuppressants | CERTOLIZUMAB PEGOL PREFILLED SYRINGE [CIMZIA] |
| 1170885 | MA00 | Immunosuppressants | CIMZIA INJECTABLE PRODUCT |
| 1360081 | MA00 | Immunosuppressants | CERTOLIZUMAB PEGOL 200 MG/ML PREFILLED SYRINGE |
| 795081 | MA00 | Immunosuppressants | CERTOLIZUMAB PEGOL 200 MG INJECTION |
| 849597 | MA00 | Immunosuppressants | 1 ML CERTOLIZUMAB PEGOL 200 MG/ML PREFILLED SYRINGE |
| 1653727 | MA00 | Immunosuppressants | CERTOLIZUMAB PEGOL 200 MG |

| | | | |
|---|---|---|---|
| 795079 | MA00 | Immunosuppressants | CERTOLIZUMAB PEGOL 200 MG/ML |
| 1653728 | MA00 | Immunosuppressants | CERTOLIZUMAB PEGOL INJECTION |
| 795080 | MA00 | Immunosuppressants | CERTOLIZUMAB PEGOL INJECTABLE SOLUTION |
| 849596 | MA00 | Immunosuppressants | CERTOLIZUMAB PEGOL PREFILLED SYRINGE |
| 1161226 | MA00 | Immunosuppressants | CERTOLIZUMAB PEGOL INJECTABLE PRODUCT |
| 202994 | MA00 | Immunosuppressants | LEUKERAN |
| 2346 | MA00 | Immunosuppressants | CHLORAMBUCIL |
| 105553 | MA00 | Immunosuppressants | CHLORAMBUCIL 2 MG ORAL TABLET [LEUKERAN] |
| 105554 | MA00 | Immunosuppressants | CHLORAMBUCIL 5 MG ORAL TABLET [LEUKERAN] |
| 564080 | MA00 | Immunosuppressants | CHLORAMBUCIL 2 MG [LEUKERAN] |
| 564081 | MA00 | Immunosuppressants | CHLORAMBUCIL 5 MG [LEUKERAN] |
| 368349 | MA00 | Immunosuppressants | CHLORAMBUCIL ORAL TABLET [LEUKERAN] |
| 1166066 | MA00 | Immunosuppressants | LEUKERAN ORAL PRODUCT |
| 1166067 | MA00 | Immunosuppressants | LEUKERAN PILL |
| 197462 | MA00 | Immunosuppressants | CHLORAMBUCIL 2 MG ORAL TABLET |
| 199311 | MA00 | Immunosuppressants | CHLORAMBUCIL 5 MG ORAL TABLET |
| 249898 | MA00 | Immunosuppressants | CHLORAMBUCIL 10 MG ORAL TABLET |
| 411288 | MA00 | Immunosuppressants | CHLORAMBUCIL 2 MG ORAL CAPSULE |
| 315626 | MA00 | Immunosuppressants | CHLORAMBUCIL 2 MG |
| 331665 | MA00 | Immunosuppressants | CHLORAMBUCIL 5 MG |
| 332752 | MA00 | Immunosuppressants | CHLORAMBUCIL 10 MG |
| 371375 | MA00 | Immunosuppressants | CHLORAMBUCIL ORAL TABLET |
| 438413 | MA00 | Immunosuppressants | CHLORAMBUCIL ORAL CAPSULE |
| 1153038 | MA00 | Immunosuppressants | CHLORAMBUCIL ORAL PRODUCT |
| 1153039 | MA00 | Immunosuppressants | CHLORAMBUCIL PILL |
| 680973 | MA00 | Immunosuppressants | SOLIRIS |
| 591781 | MA00 | Immunosuppressants | ECULIZUMAB |
| 1655952 | MA00 | Immunosuppressants | ECULIZUMAB 10 MG/ML INJECTION [SOLIRIS] |
| 700387 | MA00 | Immunosuppressants | 30 ML ECULIZUMAB 10 MG/ML INJECTION [SOLIRIS] |
| 700385 | MA00 | Immunosuppressants | ECULIZUMAB 10 MG/ML [SOLIRIS] |
| 1655949 | MA00 | Immunosuppressants | ECULIZUMAB INJECTION [SOLIRIS] |
| 700386 | MA00 | Immunosuppressants | ECULIZUMAB INJECTABLE SOLUTION [SOLIRIS] |
| 1184826 | MA00 | Immunosuppressants | SOLIRIS INJECTABLE PRODUCT |
| 1655951 | MA00 | Immunosuppressants | ECULIZUMAB 10 MG/ML INJECTION |
| 700384 | MA00 | Immunosuppressants | 30 ML ECULIZUMAB 10 MG/ML INJECTION |
| 700382 | MA00 | Immunosuppressants | ECULIZUMAB 10 MG/ML |
| 1655947 | MA00 | Immunosuppressants | ECULIZUMAB INJECTION |
| 700383 | MA00 | Immunosuppressants | ECULIZUMAB INJECTABLE SOLUTION |
| 1161281 | MA00 | Immunosuppressants | ECULIZUMAB INJECTABLE PRODUCT |
| 216891 | MA00 | Immunosuppressants | ENBREL |
| 214555 | MA00 | Immunosuppressants | ETANERCEPT |
| 2103480 | MA00 | Immunosuppressants | ETANERCEPT-SZZS |
| 1360430 | MA00 | Immunosuppressants | ETANERCEPT 50 MG/ML PREFILLED SYRINGE [ENBREL] |
| 1653225 | MA00 | Immunosuppressants | 1 ML ETANERCEPT 50 MG/ML AUTO-INJECTOR [ENBREL] |
| 1653227 | MA00 | Immunosuppressants | ETANERCEPT 50 MG/ML AUTO-INJECTOR [ENBREL] |
| 2182340 | MA00 | Immunosuppressants | 1 ML ETANERCEPT 50 MG/ML CARTRIDGE [ENBREL] |
| 2182342 | MA00 | Immunosuppressants | ETANERCEPT 50 MG/ML CARTRIDGE [ENBREL] |
| 261105 | MA00 | Immunosuppressants | ETANERCEPT 25 MG/ML INJECTABLE SOLUTION [ENBREL] |
| 582673 | MA00 | Immunosuppressants | ETANERCEPT 50 MG/ML INJECTABLE SOLUTION [ENBREL] |
| 802652 | MA00 | Immunosuppressants | 1 ML ETANERCEPT 50 MG/ML PREFILLED SYRINGE [ENBREL] |
| 802654 | MA00 | Immunosuppressants | 1 ML ETANERCEPT 50 MG/ML PREFILLED SYRINGE [ENBREL] |
| 809159 | MA00 | Immunosuppressants | 0.5 ML ETANERCEPT 50 MG/ML PREFILLED SYRINGE [ENBREL] |
| 574352 | MA00 | Immunosuppressants | ETANERCEPT 25 MG/ML [ENBREL] |
| 582672 | MA00 | Immunosuppressants | ETANERCEPT 50 MG/ML [ENBREL] |
| 1653224 | MA00 | Immunosuppressants | ETANERCEPT AUTO-INJECTOR [ENBREL] |
| 2182339 | MA00 | Immunosuppressants | ETANERCEPT CARTRIDGE [ENBREL] |
| 363197 | MA00 | Immunosuppressants | ETANERCEPT INJECTABLE SOLUTION [ENBREL] |
| 802651 | MA00 | Immunosuppressants | ETANERCEPT PREFILLED SYRINGE [ENBREL] |
| 1175957 | MA00 | Immunosuppressants | ENBREL INJECTABLE PRODUCT |
| 1359949 | MA00 | Immunosuppressants | ETANERCEPT 50 MG/ML PREFILLED SYRINGE |
| 1653223 | MA00 | Immunosuppressants | 1 ML ETANERCEPT 50 MG/ML AUTO-INJECTOR |
| 1653226 | MA00 | Immunosuppressants | ETANERCEPT 50 MG/ML AUTO-INJECTOR |
| 2182338 | MA00 | Immunosuppressants | 1 ML ETANERCEPT 50 MG/ML CARTRIDGE |
| 2182341 | MA00 | Immunosuppressants | ETANERCEPT 50 MG/ML CARTRIDGE |
| 253014 | MA00 | Immunosuppressants | ETANERCEPT 25 MG/ML INJECTABLE SOLUTION |
| 582671 | MA00 | Immunosuppressants | ETANERCEPT 50 MG/ML INJECTABLE SOLUTION |
| 727757 | MA00 | Immunosuppressants | 1 ML ETANERCEPT 50 MG/ML PREFILLED SYRINGE |
| 727904 | MA00 | Immunosuppressants | 0.51 ML ETANERCEPT 20 MG/ML PREFILLED SYRINGE |
| 802653 | MA00 | Immunosuppressants | 1 ML ETANERCEPT 50 MG/ML PREFILLED SYRINGE |
| 809158 | MA00 | Immunosuppressants | 0.5 ML ETANERCEPT 50 MG/ML PREFILLED SYRINGE |
| 342977 | MA00 | Immunosuppressants | ETANERCEPT 25 MG/ML |

| | | | |
|---|---|---|---|
| 582670 | MA00 | Immunosuppressants | ETANERCEPT 50 MG/ML |
| 727903 | MA00 | Immunosuppressants | ETANERCEPT 20 MG/ML |
| 1653222 | MA00 | Immunosuppressants | ETANERCEPT AUTO-INJECTOR |
| 2182337 | MA00 | Immunosuppressants | ETANERCEPT CARTRIDGE |
| 385098 | MA00 | Immunosuppressants | ETANERCEPT INJECTABLE SOLUTION |
| 727756 | MA00 | Immunosuppressants | ETANERCEPT PREFILLED SYRINGE |
| 1156809 | MA00 | Immunosuppressants | ETANERCEPT INJECTABLE PRODUCT |
| 848161 | MA00 | Immunosuppressants | SIMPONI |
| 819300 | MA00 | Immunosuppressants | GOLIMUMAB |
| 1360436 | MA00 | Immunosuppressants | GOLIMUMAB 100 MG/ML PREFILLED SYRINGE [SIMPONI] |
| 1431647 | MA00 | Immunosuppressants | 4 ML GOLIMUMAB 12.5 MG/ML INJECTION [SIMPONI] |
| 1482814 | MA00 | Immunosuppressants | 1 ML GOLIMUMAB 100 MG/ML PREFILLED SYRINGE [SIMPONI] |
| 1653144 | MA00 | Immunosuppressants | 0.5 ML GOLIMUMAB 100 MG/ML AUTO-INJECTOR [SIMPONI] |
| 1653146 | MA00 | Immunosuppressants | GOLIMUMAB 100 MG/ML AUTO-INJECTOR [SIMPONI] |
| 1653166 | MA00 | Immunosuppressants | 1 ML GOLIMUMAB 100 MG/ML AUTO-INJECTOR [SIMPONI] |
| 1653574 | MA00 | Immunosuppressants | GOLIMUMAB 12.5 MG/ML INJECTION [SIMPONI] |
| 848164 | MA00 | Immunosuppressants | 0.5 ML GOLIMUMAB 100 MG/ML PREFILLED SYRINGE [SIMPONI] |
| 1431644 | MA00 | Immunosuppressants | GOLIMUMAB 12.5 MG/ML [SIMPONI] |
| 848162 | MA00 | Immunosuppressants | GOLIMUMAB 100 MG/ML [SIMPONI] |
| 1431645 | MA00 | Immunosuppressants | GOLIMUMAB INJECTABLE SOLUTION [SIMPONI ARIA] |
| 1653143 | MA00 | Immunosuppressants | GOLIMUMAB AUTO-INJECTOR [SIMPONI] |
| 1653571 | MA00 | Immunosuppressants | GOLIMUMAB INJECTION [SIMPONI] |
| 848163 | MA00 | Immunosuppressants | GOLIMUMAB PREFILLED SYRINGE [SIMPONI] |
| 1182643 | MA00 | Immunosuppressants | SIMPONI INJECTABLE PRODUCT |
| 1359486 | MA00 | Immunosuppressants | GOLIMUMAB 100 MG/ML PREFILLED SYRINGE |
| 1431642 | MA00 | Immunosuppressants | 4 ML GOLIMUMAB 12.5 MG/ML INJECTION |
| 1482813 | MA00 | Immunosuppressants | 1 ML GOLIMUMAB 100 MG/ML PREFILLED SYRINGE |
| 1653142 | MA00 | Immunosuppressants | 0.5 ML GOLIMUMAB 100 MG/ML AUTO-INJECTOR |
| 1653145 | MA00 | Immunosuppressants | GOLIMUMAB 100 MG/ML AUTO-INJECTOR |
| 1653165 | MA00 | Immunosuppressants | 1 ML GOLIMUMAB 100 MG/ML AUTO-INJECTOR |
| 1653573 | MA00 | Immunosuppressants | GOLIMUMAB 12.5 MG/ML INJECTION |
| 848160 | MA00 | Immunosuppressants | 0.5 ML GOLIMUMAB 100 MG/ML PREFILLED SYRINGE |
| 1431639 | MA00 | Immunosuppressants | GOLIMUMAB 12.5 MG/ML |
| 848158 | MA00 | Immunosuppressants | GOLIMUMAB 100 MG/ML |
| 1431641 | MA00 | Immunosuppressants | GOLIMUMAB INJECTABLE SOLUTION |
| 1653141 | MA00 | Immunosuppressants | GOLIMUMAB AUTO-INJECTOR |
| 1653569 | MA00 | Immunosuppressants | GOLIMUMAB INJECTION |
| 848159 | MA00 | Immunosuppressants | GOLIMUMAB PREFILLED SYRINGE |
| 1157808 | MA00 | Immunosuppressants | GOLIMUMAB INJECTABLE PRODUCT |
| 1790542 | MA00 | Immunosuppressants | INFLECTRA |
| 1927286 | MA00 | Immunosuppressants | RENFLEXIS |
| 219610 | MA00 | Immunosuppressants | REMICADE |
| 2266526 | MA00 | Immunosuppressants | AVSOLA |
| 191831 | MA00 | Immunosuppressants | INFLIXIMAB |
| 1790539 | MA00 | Immunosuppressants | INFLIXIMAB-DYYB |
| 1927283 | MA00 | Immunosuppressants | INFLIXIMAB-ABDA |
| 2103476 | MA00 | Immunosuppressants | INFLIXIMAB-QBTX |
| 2266523 | MA00 | Immunosuppressants | INFLIXIMAB-AXXQ |
| 1790546 | MA00 | Immunosuppressants | INFLIXIMAB-DYYB 100 MG INJECTION [INFLECTRA] |
| 1927290 | MA00 | Immunosuppressants | INFLIXIMAB-ABDA 100 MG INJECTION [RENFLEXIS] |
| 213361 | MA00 | Immunosuppressants | INFLIXIMAB 100 MG INJECTION [REMICADE] |
| 2266530 | MA00 | Immunosuppressants | INFLIXIMAB-AXXQ 100 MG INJECTION [AVSOLA] |
| 1655943 | MA00 | Immunosuppressants | INFLIXIMAB 100 MG [REMICADE] |
| 1790543 | MA00 | Immunosuppressants | INFLIXIMAB-DYYB 100 MG [INFLECTRA] |
| 1927287 | MA00 | Immunosuppressants | INFLIXIMAB-ABDA 100 MG [RENFLEXIS] |
| 2266527 | MA00 | Immunosuppressants | INFLIXIMAB-AXXQ 100 MG [AVSOLA] |
| 573255 | MA00 | Immunosuppressants | INFLIXIMAB 10 MG/ML [REMICADE] |
| 1655944 | MA00 | Immunosuppressants | INFLIXIMAB INJECTION [REMICADE] |
| 1790544 | MA00 | Immunosuppressants | INFLIXIMAB INJECTION [INFLECTRA] |
| 1927288 | MA00 | Immunosuppressants | INFLIXIMAB INJECTION [RENFLEXIS] |
| 2266528 | MA00 | Immunosuppressants | INFLIXIMAB INJECTION [AVSOLA] |
| 363437 | MA00 | Immunosuppressants | INFLIXIMAB INJECTABLE SOLUTION [REMICADE] |
| 1180726 | MA00 | Immunosuppressants | REMICADE INJECTABLE PRODUCT |
| 1790545 | MA00 | Immunosuppressants | INFLECTRA INJECTABLE PRODUCT |
| 1927289 | MA00 | Immunosuppressants | RENFLEXIS INJECTABLE PRODUCT |
| 2266529 | MA00 | Immunosuppressants | AVSOLA INJECTABLE PRODUCT |
| 1790541 | MA00 | Immunosuppressants | INFLIXIMAB-DYYB 100 MG INJECTION |
| 1927285 | MA00 | Immunosuppressants | INFLIXIMAB-ABDA 100 MG INJECTION |
| 2266525 | MA00 | Immunosuppressants | INFLIXIMAB-AXXQ 100 MG INJECTION |
| 310994 | MA00 | Immunosuppressants | INFLIXIMAB 100 MG INJECTION |
| 1655940 | MA00 | Immunosuppressants | INFLIXIMAB 100 MG |

| | | | |
|---|---|---|---|
| 1790540 | MA00 | Immunosuppressants | INFLIXIMAB-DYYB 100 MG |
| 1927284 | MA00 | Immunosuppressants | INFLIXIMAB-ABDA 100 MG |
| 2266524 | MA00 | Immunosuppressants | INFLIXIMAB-AXXQ 100 MG |
| 358693 | MA00 | Immunosuppressants | INFLIXIMAB 10 MG/ML |
| 1655941 | MA00 | Immunosuppressants | INFLIXIMAB INJECTION |
| 376873 | MA00 | Immunosuppressants | INFLIXIMAB INJECTABLE SOLUTION |
| 1160692 | MA00 | Immunosuppressants | INFLIXIMAB INJECTABLE PRODUCT |
| 215393 | MA00 | Immunosuppressants | ARAVA |
| 27169 | MA00 | Immunosuppressants | LEFLUNOMIDE |
| 213377 | MA00 | Immunosuppressants | LEFLUNOMIDE 10 MG ORAL TABLET [ARAVA] |
| 213379 | MA00 | Immunosuppressants | LEFLUNOMIDE 20 MG ORAL TABLET [ARAVA] |
| 213380 | MA00 | Immunosuppressants | LEFLUNOMIDE 100 MG ORAL TABLET [ARAVA] |
| 573271 | MA00 | Immunosuppressants | LEFLUNOMIDE 10 MG [ARAVA] |
| 573273 | MA00 | Immunosuppressants | LEFLUNOMIDE 20 MG [ARAVA] |
| 573274 | MA00 | Immunosuppressants | LEFLUNOMIDE 100 MG [ARAVA] |
| 368264 | MA00 | Immunosuppressants | LEFLUNOMIDE ORAL TABLET [ARAVA] |
| 1170095 | MA00 | Immunosuppressants | ARAVA ORAL PRODUCT |
| 1170096 | MA00 | Immunosuppressants | ARAVA PILL |
| 205284 | MA00 | Immunosuppressants | LEFLUNOMIDE 10 MG ORAL TABLET |
| 205285 | MA00 | Immunosuppressants | LEFLUNOMIDE 20 MG ORAL TABLET |
| 205286 | MA00 | Immunosuppressants | LEFLUNOMIDE 100 MG ORAL TABLET |
| 316133 | MA00 | Immunosuppressants | LEFLUNOMIDE 10 MG |
| 316134 | MA00 | Immunosuppressants | LEFLUNOMIDE 20 MG |
| 317399 | MA00 | Immunosuppressants | LEFLUNOMIDE 100 MG |
| 372570 | MA00 | Immunosuppressants | LEFLUNOMIDE ORAL TABLET |
| 1162566 | MA00 | Immunosuppressants | LEFLUNOMIDE ORAL PRODUCT |
| 1162567 | MA00 | Immunosuppressants | LEFLUNOMIDE PILL |
| 1876381 | MA00 | Immunosuppressants | OCREVUS |
| 1876366 | MA00 | Immunosuppressants | OCRELIZUMAB |
| 1876385 | MA00 | Immunosuppressants | 10 ML OCRELIZUMAB 30 MG/ML INJECTION [OCREVUS] |
| 1876387 | MA00 | Immunosuppressants | OCRELIZUMAB 30 MG/ML INJECTION [OCREVUS] |
| 1876382 | MA00 | Immunosuppressants | OCRELIZUMAB 30 MG/ML [OCREVUS] |
| 1876383 | MA00 | Immunosuppressants | OCRELIZUMAB INJECTION [OCREVUS] |
| 1876384 | MA00 | Immunosuppressants | OCREVUS INJECTABLE PRODUCT |
| 1876380 | MA00 | Immunosuppressants | 10 ML OCRELIZUMAB 30 MG/ML INJECTION |
| 1876386 | MA00 | Immunosuppressants | OCRELIZUMAB 30 MG/ML INJECTION |
| 1876377 | MA00 | Immunosuppressants | OCRELIZUMAB 30 MG/ML |
| 1876379 | MA00 | Immunosuppressants | OCRELIZUMAB INJECTION |
| 1876378 | MA00 | Immunosuppressants | OCRELIZUMAB INJECTABLE PRODUCT |
| 876301 | MA00 | Immunosuppressants | ARZERRA |
| 712566 | MA00 | Immunosuppressants | OFATUMUMAB |
| 1658044 | MA00 | Immunosuppressants | 50 ML OFATUMUMAB 20 MG/ML INJECTION [ARZERRA] |
| 1658046 | MA00 | Immunosuppressants | OFATUMUMAB 20 MG/ML INJECTION [ARZERRA] |
| 1658048 | MA00 | Immunosuppressants | 5 ML OFATUMUMAB 20 MG/ML INJECTION [ARZERRA] |
| 876304 | MA00 | Immunosuppressants | OFATUMUMAB 10 MG/ML INJECTABLE SOLUTION [ARZERRA] |
| 877012 | MA00 | Immunosuppressants | OFATUMUMAB 20 MG/ML INJECTABLE SOLUTION [ARZERRA] |
| 876302 | MA00 | Immunosuppressants | OFATUMUMAB 10 MG/ML [ARZERRA] |
| 877011 | MA00 | Immunosuppressants | OFATUMUMAB 20 MG/ML [ARZERRA] |
| 1658043 | MA00 | Immunosuppressants | OFATUMUMAB INJECTION [ARZERRA] |
| 876303 | MA00 | Immunosuppressants | OFATUMUMAB INJECTABLE SOLUTION [ARZERRA] |
| 1171445 | MA00 | Immunosuppressants | ARZERRA INJECTABLE PRODUCT |
| 1658042 | MA00 | Immunosuppressants | 50 ML OFATUMUMAB 20 MG/ML INJECTION |
| 1658045 | MA00 | Immunosuppressants | OFATUMUMAB 20 MG/ML INJECTION |
| 1658047 | MA00 | Immunosuppressants | 5 ML OFATUMUMAB 20 MG/ML INJECTION |
| 876300 | MA00 | Immunosuppressants | OFATUMUMAB 10 MG/ML INJECTABLE SOLUTION |
| 877010 | MA00 | Immunosuppressants | OFATUMUMAB 20 MG/ML INJECTABLE SOLUTION |
| 876298 | MA00 | Immunosuppressants | OFATUMUMAB 10 MG/ML |
| 877009 | MA00 | Immunosuppressants | OFATUMUMAB 20 MG/ML |
| 1658041 | MA00 | Immunosuppressants | OFATUMUMAB INJECTION |
| 876299 | MA00 | Immunosuppressants | OFATUMUMAB INJECTABLE SOLUTION |
| 1159554 | MA00 | Immunosuppressants | OFATUMUMAB INJECTABLE PRODUCT |
| 763454 | MA00 | Immunosuppressants | ARCALYST |
| 763450 | MA00 | Immunosuppressants | RILONACEPT |
| 763457 | MA00 | Immunosuppressants | RILONACEPT 220 MG INJECTION [ARCALYST] |
| 1724261 | MA00 | Immunosuppressants | RILONACEPT 220 MG [ARCALYST] |
| 763455 | MA00 | Immunosuppressants | RILONACEPT 80 MG/ML [ARCALYST] |
| 1724262 | MA00 | Immunosuppressants | RILONACEPT INJECTION [ARCALYST] |
| 763456 | MA00 | Immunosuppressants | RILONACEPT INJECTABLE SOLUTION [ARCALYST] |
| 1170102 | MA00 | Immunosuppressants | ARCALYST INJECTABLE PRODUCT |
| 763453 | MA00 | Immunosuppressants | RILONACEPT 220 MG INJECTION |
| 1724258 | MA00 | Immunosuppressants | RILONACEPT 220 MG |

| | | | |
|---|---|---|---|
| 763451 | MA00 | Immunosuppressants | RILONACEPT 80 MG/ML |
| 1724259 | MA00 | Immunosuppressants | RILONACEPT INJECTION |
| 763452 | MA00 | Immunosuppressants | RILONACEPT INJECTABLE SOLUTION |
| 1157438 | MA00 | Immunosuppressants | RILONACEPT INJECTABLE PRODUCT |
| 1927886 | MA00 | Immunosuppressants | RITUXAN HYCELA |
| 2105827 | MA00 | Immunosuppressants | TRUXIMA |
| 226754 | MA00 | Immunosuppressants | RITUXAN |
| 2273513 | MA00 | Immunosuppressants | RUXIENCE |
| 121191 | MA00 | Immunosuppressants | RITUXIMAB |
| 1927884 | MA00 | Immunosuppressants | HYALURONIDASE / RITUXIMAB |
| 2105824 | MA00 | Immunosuppressants | RITUXIMAB-ABBS |
| 2273510 | MA00 | Immunosuppressants | RITUXIMAB-PVVR |
| 1657864 | MA00 | Immunosuppressants | 10 ML RITUXIMAB 10 MG/ML INJECTION [RITUXAN] |
| 1657866 | MA00 | Immunosuppressants | RITUXIMAB 10 MG/ML INJECTION [RITUXAN] |
| 1657868 | MA00 | Immunosuppressants | 50 ML RITUXIMAB 10 MG/ML INJECTION [RITUXAN] |
| 1927890 | MA00 | Immunosuppressants | 11.7 ML HYALURONIDASE, HUMAN RECOMBINANT 2000 UNT/ML / RITUXIMAB 120 MG/ML INJECTION [RITUXAN HYCELA] |
| 1927892 | MA00 | Immunosuppressants | HYALURONIDASE, HUMAN RECOMBINANT 2000 UNT/ML / RITUXIMAB 120 MG/ML INJECTION [RITUXAN HYCELA] |
| 1927894 | MA00 | Immunosuppressants | 13.4 ML HYALURONIDASE, HUMAN RECOMBINANT 2000 UNT/ML / RITUXIMAB 120 MG/ML INJECTION [RITUXAN HYCELA] |
| 2105831 | MA00 | Immunosuppressants | 10 ML RITUXIMAB-ABBS 10 MG/ML INJECTION [TRUXIMA] |
| 2105833 | MA00 | Immunosuppressants | RITUXIMAB-ABBS 10 MG/ML INJECTION [TRUXIMA] |
| 2105835 | MA00 | Immunosuppressants | 50 ML RITUXIMAB-ABBS 10 MG/ML INJECTION [TRUXIMA] |
| 213126 | MA00 | Immunosuppressants | RITUXIMAB 10 MG/ML INJECTABLE SOLUTION [RITUXAN] |
| 2273517 | MA00 | Immunosuppressants | 10 ML RITUXIMAB-PVVR 10 MG/ML INJECTION [RUXIENCE] |
| 2273519 | MA00 | Immunosuppressants | RITUXIMAB-PVVR 10 MG/ML INJECTION [RUXIENCE] |
| 2273521 | MA00 | Immunosuppressants | 50 ML RITUXIMAB-PVVR 10 MG/ML INJECTION [RUXIENCE] |
| 1927887 | MA00 | Immunosuppressants | HYALURONIDASE, HUMAN RECOMBINANT 2000 UNT/ML / RITUXIMAB 120 MG/ML [RITUXAN HYCELA] |
| 2105828 | MA00 | Immunosuppressants | RITUXIMAB-ABBS 10 MG/ML [TRUXIMA] |
| 2273514 | MA00 | Immunosuppressants | RITUXIMAB-PVVR 10 MG/ML [RUXIENCE] |
| 573051 | MA00 | Immunosuppressants | RITUXIMAB 10 MG/ML [RITUXAN] |
| 1657863 | MA00 | Immunosuppressants | RITUXIMAB INJECTION [RITUXAN] |
| 1927888 | MA00 | Immunosuppressants | HYALURONIDASE / RITUXIMAB INJECTION [RITUXAN HYCELA] |
| 2105829 | MA00 | Immunosuppressants | RITUXIMAB INJECTION [TRUXIMA] |
| 2273515 | MA00 | Immunosuppressants | RITUXIMAB INJECTION [RUXIENCE] |
| 362907 | MA00 | Immunosuppressants | RITUXIMAB INJECTABLE SOLUTION [RITUXAN] |
| 1185345 | MA00 | Immunosuppressants | RITUXAN INJECTABLE PRODUCT |
| 1927889 | MA00 | Immunosuppressants | RITUXAN HYCELA INJECTABLE PRODUCT |
| 2105830 | MA00 | Immunosuppressants | TRUXIMA INJECTABLE PRODUCT |
| 2273516 | MA00 | Immunosuppressants | RUXIENCE INJECTABLE PRODUCT |
| 1657862 | MA00 | Immunosuppressants | 10 ML RITUXIMAB 10 MG/ML INJECTION |
| 1657865 | MA00 | Immunosuppressants | RITUXIMAB 10 MG/ML INJECTION |
| 1657867 | MA00 | Immunosuppressants | 50 ML RITUXIMAB 10 MG/ML INJECTION |
| 1927885 | MA00 | Immunosuppressants | 11.7 ML HYALURONIDASE, HUMAN RECOMBINANT 2000 UNT/ML / RITUXIMAB 120 MG/ML INJECTION |
| 1927891 | MA00 | Immunosuppressants | HYALURONIDASE, HUMAN RECOMBINANT 2000 UNT/ML / RITUXIMAB 120 MG/ML INJECTION |
| 1927893 | MA00 | Immunosuppressants | 13.4 ML HYALURONIDASE, HUMAN RECOMBINANT 2000 UNT/ML / RITUXIMAB 120 MG/ML INJECTION |
| 2105826 | MA00 | Immunosuppressants | 10 ML RITUXIMAB-ABBS 10 MG/ML INJECTION |
| 2105832 | MA00 | Immunosuppressants | RITUXIMAB-ABBS 10 MG/ML INJECTION |
| 2105834 | MA00 | Immunosuppressants | 50 ML RITUXIMAB-ABBS 10 MG/ML INJECTION |
| 2273512 | MA00 | Immunosuppressants | 10 ML RITUXIMAB-PVVR 10 MG/ML INJECTION |
| 2273518 | MA00 | Immunosuppressants | RITUXIMAB-PVVR 10 MG/ML INJECTION |
| 2273520 | MA00 | Immunosuppressants | 50 ML RITUXIMAB-PVVR 10 MG/ML INJECTION |
| 242435 | MA00 | Immunosuppressants | RITUXIMAB 10 MG/ML INJECTABLE SOLUTION |
| 1927881 | MA00 | Immunosuppressants | RITUXIMAB 120 MG/ML |
| 2105825 | MA00 | Immunosuppressants | RITUXIMAB-ABBS 10 MG/ML |
| 2273511 | MA00 | Immunosuppressants | RITUXIMAB-PVVR 10 MG/ML |
| 316648 | MA00 | Immunosuppressants | RITUXIMAB 10 MG/ML |
| 1657861 | MA00 | Immunosuppressants | RITUXIMAB INJECTION |
| 1927883 | MA00 | Immunosuppressants | HYALURONIDASE / RITUXIMAB INJECTION |
| 377259 | MA00 | Immunosuppressants | RITUXIMAB INJECTABLE SOLUTION |
| 1157967 | MA00 | Immunosuppressants | RITUXIMAB INJECTABLE PRODUCT |
| 1927882 | MA00 | Immunosuppressants | HYALURONIDASE / RITUXIMAB INJECTABLE PRODUCT |
| 1599793 | MA00 | Immunosuppressants | COSENTYX |
| 1599788 | MA00 | Immunosuppressants | SECUKINUMAB |
| 1599797 | MA00 | Immunosuppressants | 1 ML SECUKINUMAB 150 MG/ML PREFILLED SYRINGE [COSENTYX] |
| 1599799 | MA00 | Immunosuppressants | SECUKINUMAB 150 MG/ML PREFILLED SYRINGE [COSENTYX] |

| | | | |
|---|---|---|---|
| 1653243 | MA00 | Immunosuppressants | 1 ML SECUKINUMAB 150 MG/ML AUTO-INJECTOR [COSENTYX] |
| 1653245 | MA00 | Immunosuppressants | SECUKINUMAB 150 MG/ML AUTO-INJECTOR [COSENTYX] |
| 1599794 | MA00 | Immunosuppressants | SECUKINUMAB 150 MG/ML [COSENTYX] |
| 1599795 | MA00 | Immunosuppressants | SECUKINUMAB PREFILLED SYRINGE [COSENTYX] |
| 1653242 | MA00 | Immunosuppressants | SECUKINUMAB AUTO-INJECTOR [COSENTYX] |
| 1599796 | MA00 | Immunosuppressants | COSENTYX INJECTABLE PRODUCT |
| 1599792 | MA00 | Immunosuppressants | 1 ML SECUKINUMAB 150 MG/ML PREFILLED SYRINGE |
| 1599798 | MA00 | Immunosuppressants | SECUKINUMAB 150 MG/ML PREFILLED SYRINGE |
| 1653241 | MA00 | Immunosuppressants | 1 ML SECUKINUMAB 150 MG/ML AUTO-INJECTOR |
| 1653244 | MA00 | Immunosuppressants | SECUKINUMAB 150 MG/ML AUTO-INJECTOR |
| 1599789 | MA00 | Immunosuppressants | SECUKINUMAB 150 MG/ML |
| 1599791 | MA00 | Immunosuppressants | SECUKINUMAB PREFILLED SYRINGE |
| 1653240 | MA00 | Immunosuppressants | SECUKINUMAB AUTO-INJECTOR |
| 1599790 | MA00 | Immunosuppressants | SECUKINUMAB INJECTABLE PRODUCT |
| 895761 | MA00 | Immunosuppressants | ACTEMRA |
| 612865 | MA00 | Immunosuppressants | TOCILIZUMAB |
| 1441530 | MA00 | Immunosuppressants | 0.9 ML TOCILIZUMAB 180 MG/ML PREFILLED SYRINGE [ACTEMRA] |
| 1441532 | MA00 | Immunosuppressants | TOCILIZUMAB 180 MG/ML PREFILLED SYRINGE [ACTEMRA] |
| 1657976 | MA00 | Immunosuppressants | 4 ML TOCILIZUMAB 20 MG/ML INJECTION [ACTEMRA] |
| 1657978 | MA00 | Immunosuppressants | TOCILIZUMAB 20 MG/ML INJECTION [ACTEMRA] |
| 1657980 | MA00 | Immunosuppressants | 10 ML TOCILIZUMAB 20 MG/ML INJECTION [ACTEMRA] |
| 1657982 | MA00 | Immunosuppressants | 20 ML TOCILIZUMAB 20 MG/ML INJECTION [ACTEMRA] |
| 2106075 | MA00 | Immunosuppressants | 0.9 ML TOCILIZUMAB 180 MG/ML AUTO-INJECTOR [ACTEMRA] |
| 2106077 | MA00 | Immunosuppressants | TOCILIZUMAB 180 MG/ML AUTO-INJECTOR [ACTEMRA] |
| 895764 | MA00 | Immunosuppressants | TOCILIZUMAB 20 MG/ML INJECTABLE SOLUTION [ACTEMRA] |
| 1441528 | MA00 | Immunosuppressants | TOCILIZUMAB 180 MG/ML [ACTEMRA] |
| 895762 | MA00 | Immunosuppressants | TOCILIZUMAB 20 MG/ML [ACTEMRA] |
| 1441529 | MA00 | Immunosuppressants | TOCILIZUMAB PREFILLED SYRINGE [ACTEMRA] |
| 1657975 | MA00 | Immunosuppressants | TOCILIZUMAB INJECTION [ACTEMRA] |
| 2106074 | MA00 | Immunosuppressants | TOCILIZUMAB AUTO-INJECTOR [ACTEMRA] |
| 895763 | MA00 | Immunosuppressants | TOCILIZUMAB INJECTABLE SOLUTION [ACTEMRA] |
| 1169167 | MA00 | Immunosuppressants | ACTEMRA INJECTABLE PRODUCT |
| 1441527 | MA00 | Immunosuppressants | 0.9 ML TOCILIZUMAB 180 MG/ML PREFILLED SYRINGE |
| 1441531 | MA00 | Immunosuppressants | TOCILIZUMAB 180 MG/ML PREFILLED SYRINGE |
| 1657974 | MA00 | Immunosuppressants | 4 ML TOCILIZUMAB 20 MG/ML INJECTION |
| 1657977 | MA00 | Immunosuppressants | TOCILIZUMAB 20 MG/ML INJECTION |
| 1657979 | MA00 | Immunosuppressants | 10 ML TOCILIZUMAB 20 MG/ML INJECTION |
| 1657981 | MA00 | Immunosuppressants | 20 ML TOCILIZUMAB 20 MG/ML INJECTION |
| 2106073 | MA00 | Immunosuppressants | 0.9 ML TOCILIZUMAB 180 MG/ML AUTO-INJECTOR |
| 2106076 | MA00 | Immunosuppressants | TOCILIZUMAB 180 MG/ML AUTO-INJECTOR |
| 895760 | MA00 | Immunosuppressants | TOCILIZUMAB 20 MG/ML INJECTABLE SOLUTION |
| 1441525 | MA00 | Immunosuppressants | TOCILIZUMAB 180 MG/ML |
| 895758 | MA00 | Immunosuppressants | TOCILIZUMAB 20 MG/ML |
| 1441526 | MA00 | Immunosuppressants | TOCILIZUMAB PREFILLED SYRINGE |
| 1657973 | MA00 | Immunosuppressants | TOCILIZUMAB INJECTION |
| 2106072 | MA00 | Immunosuppressants | TOCILIZUMAB AUTO-INJECTOR |
| 895759 | MA00 | Immunosuppressants | TOCILIZUMAB INJECTABLE SOLUTION |
| 1162729 | MA00 | Immunosuppressants | TOCILIZUMAB INJECTABLE PRODUCT |
| 1357542 | MA00 | Immunosuppressants | XELJANZ |
| 1357536 | MA00 | Immunosuppressants | TOFACITINIB |
| 1358492 | MA00 | Immunosuppressants | TOFACITINIB CITRATE |
| 1357547 | MA00 | Immunosuppressants | TOFACITINIB 5 MG ORAL TABLET [XELJANZ] |
| 1741049 | MA00 | Immunosuppressants | 24 HR TOFACITINIB 11 MG EXTENDED RELEASE ORAL TABLET [XELJANZ] |
| 1741051 | MA00 | Immunosuppressants | TOFACITINIB 11 MG EXTENDED RELEASE ORAL TABLET [XELJANZ] |
| 2048568 | MA00 | Immunosuppressants | TOFACITINIB 10 MG ORAL TABLET [XELJANZ] |
| 2273115 | MA00 | Immunosuppressants | 24 HR TOFACITINIB 22 MG EXTENDED RELEASE ORAL TABLET [XELJANZ] |
| 2273117 | MA00 | Immunosuppressants | TOFACITINIB 22 MG EXTENDED RELEASE ORAL TABLET [XELJANZ] |
| 1357543 | MA00 | Immunosuppressants | TOFACITINIB 5 MG [XELJANZ] |
| 1741047 | MA00 | Immunosuppressants | TOFACITINIB 11 MG [XELJANZ] |
| 2048567 | MA00 | Immunosuppressants | TOFACITINIB 10 MG [XELJANZ] |
| 2273114 | MA00 | Immunosuppressants | TOFACITINIB 22 MG [XELJANZ] |
| 1357544 | MA00 | Immunosuppressants | TOFACITINIB ORAL TABLET [XELJANZ] |
| 1741048 | MA00 | Immunosuppressants | TOFACITINIB EXTENDED RELEASE ORAL TABLET [XELJANZ] |
| 1357545 | MA00 | Immunosuppressants | XELJANZ ORAL PRODUCT |
| 1357546 | MA00 | Immunosuppressants | XELJANZ PILL |
| 1357541 | MA00 | Immunosuppressants | TOFACITINIB 5 MG ORAL TABLET |
| 1741046 | MA00 | Immunosuppressants | 24 HR TOFACITINIB 11 MG EXTENDED RELEASE ORAL TABLET |
| 1741050 | MA00 | Immunosuppressants | TOFACITINIB 11 MG EXTENDED RELEASE ORAL TABLET |
| 2048566 | MA00 | Immunosuppressants | TOFACITINIB 10 MG ORAL TABLET |
| 2273113 | MA00 | Immunosuppressants | 24 HR TOFACITINIB 22 MG EXTENDED RELEASE ORAL TABLET |
| 2273116 | MA00 | Immunosuppressants | TOFACITINIB 22 MG EXTENDED RELEASE ORAL TABLET |

| | | | |
|---|---|---|---|
| 1357537 | MA00 | Immunosuppressants | TOFACITINIB 5 MG |
| 1741044 | MA00 | Immunosuppressants | TOFACITINIB 11 MG |
| 2048565 | MA00 | Immunosuppressants | TOFACITINIB 10 MG |
| 2273112 | MA00 | Immunosuppressants | TOFACITINIB 22 MG |
| 1357540 | MA00 | Immunosuppressants | TOFACITINIB ORAL TABLET |
| 1741045 | MA00 | Immunosuppressants | TOFACITINIB EXTENDED RELEASE ORAL TABLET |
| 1357538 | MA00 | Immunosuppressants | TOFACITINIB ORAL PRODUCT |
| 1357539 | MA00 | Immunosuppressants | TOFACITINIB PILL |
| 151397 | MA00 | Immunosuppressants | AZAMUNE |
| 151888 | MA00 | Immunosuppressants | IMMUNOPRIN |
| 152156 | MA00 | Immunosuppressants | OPRISINE |
| 202559 | MA00 | Immunosuppressants | IMURAN |
| 360562 | MA00 | Immunosuppressants | BERKAPRINE |
| 404872 | MA00 | Immunosuppressants | AZASAN |
| 1256 | MA00 | Immunosuppressants | AZATHIOPRINE |
| 267476 | MA00 | Immunosuppressants | AZATHIOPRINE SODIUM |
| 105609 | MA00 | Immunosuppressants | AZATHIOPRINE 50 MG ORAL TABLET [AZAMUNE] |
| 105610 | MA00 | Immunosuppressants | AZATHIOPRINE 25 MG ORAL TABLET [IMURAN] |
| 105611 | MA00 | Immunosuppressants | AZATHIOPRINE 50 MG ORAL TABLET [IMURAN] |
| 105612 | MA00 | Immunosuppressants | AZATHIOPRINE 50 MG ORAL TABLET [IMMUNOPRIN] |
| 105613 | MA00 | Immunosuppressants | AZATHIOPRINE 50 MG ORAL TABLET [BERKAPRINE] |
| 108922 | MA00 | Immunosuppressants | AZATHIOPRINE 50 MG ORAL TABLET [OPRISINE] |
| 153130 | MA00 | Immunosuppressants | AZATHIOPRINE 10 MG ORAL TABLET [IMURAN] |
| 213563 | MA00 | Immunosuppressants | AZATHIOPRINE 10 MG/ML INJECTABLE SOLUTION [IMURAN] |
| 404351 | MA00 | Immunosuppressants | AZATHIOPRINE 50 MG ORAL TABLET [AZASAN] |
| 404475 | MA00 | Immunosuppressants | AZATHIOPRINE 75 MG ORAL TABLET [AZASAN] |
| 404476 | MA00 | Immunosuppressants | AZATHIOPRINE 100 MG ORAL TABLET [AZASAN] |
| 701338 | MA00 | Immunosuppressants | AZATHIOPRINE 25 MG ORAL TABLET [AZASAN] |
| 564118 | MA00 | Immunosuppressants | AZATHIOPRINE 50 MG [AZAMUNE] |
| 564119 | MA00 | Immunosuppressants | AZATHIOPRINE 25 MG [IMURAN] |
| 564120 | MA00 | Immunosuppressants | AZATHIOPRINE 50 MG [IMURAN] |
| 564121 | MA00 | Immunosuppressants | AZATHIOPRINE 50 MG [IMMUNOPRIN] |
| 564122 | MA00 | Immunosuppressants | AZATHIOPRINE 50 MG [BERKAPRINE] |
| 564655 | MA00 | Immunosuppressants | AZATHIOPRINE 50 MG [OPRISINE] |
| 565180 | MA00 | Immunosuppressants | AZATHIOPRINE 10 MG [IMURAN] |
| 573446 | MA00 | Immunosuppressants | AZATHIOPRINE 10 MG/ML [IMURAN] |
| 576332 | MA00 | Immunosuppressants | AZATHIOPRINE 50 MG [AZASAN] |
| 576426 | MA00 | Immunosuppressants | AZATHIOPRINE 75 MG [AZASAN] |
| 576427 | MA00 | Immunosuppressants | AZATHIOPRINE 100 MG [AZASAN] |
| 701337 | MA00 | Immunosuppressants | AZATHIOPRINE 25 MG [AZASAN] |
| 368214 | MA00 | Immunosuppressants | AZATHIOPRINE ORAL TABLET [IMURAN] |
| 368521 | MA00 | Immunosuppressants | AZATHIOPRINE ORAL TABLET [BERKAPRINE] |
| 369525 | MA00 | Immunosuppressants | AZATHIOPRINE ORAL TABLET [AZAMUNE] |
| 369534 | MA00 | Immunosuppressants | AZATHIOPRINE ORAL TABLET [IMMUNOPRIN] |
| 403109 | MA00 | Immunosuppressants | AZATHIOPRINE ORAL TABLET [OPRISINE] |
| 405819 | MA00 | Immunosuppressants | AZATHIOPRINE ORAL TABLET [AZASAN] |
| 92955 | MA00 | Immunosuppressants | AZATHIOPRINE INJECTABLE SOLUTION [IMURAN] |
| 1167775 | MA00 | Immunosuppressants | IMMUNOPRIN ORAL PRODUCT |
| 1167776 | MA00 | Immunosuppressants | IMMUNOPRIN PILL |
| 1172006 | MA00 | Immunosuppressants | IMURAN ORAL PRODUCT |
| 1172007 | MA00 | Immunosuppressants | IMURAN PILL |
| 1172211 | MA00 | Immunosuppressants | BERKAPRINE ORAL PRODUCT |
| 1172212 | MA00 | Immunosuppressants | BERKAPRINE PILL |
| 1176316 | MA00 | Immunosuppressants | AZAMUNE ORAL PRODUCT |
| 1176317 | MA00 | Immunosuppressants | AZAMUNE PILL |
| 1176318 | MA00 | Immunosuppressants | AZASAN ORAL PRODUCT |
| 1176319 | MA00 | Immunosuppressants | AZASAN PILL |
| 1182311 | MA00 | Immunosuppressants | OPRISINE ORAL PRODUCT |
| 1182312 | MA00 | Immunosuppressants | OPRISINE PILL |
| 153129 | MA00 | Immunosuppressants | AZATHIOPRINE 10 MG ORAL TABLET |
| 197388 | MA00 | Immunosuppressants | AZATHIOPRINE 50 MG ORAL TABLET |
| 199310 | MA00 | Immunosuppressants | AZATHIOPRINE 25 MG ORAL TABLET |
| 239983 | MA00 | Immunosuppressants | AZATHIOPRINE 100 MG INJECTION |
| 359228 | MA00 | Immunosuppressants | AZATHIOPRINE 100 MG ORAL TABLET |
| 359229 | MA00 | Immunosuppressants | AZATHIOPRINE 75 MG ORAL TABLET |
| 410148 | MA00 | Immunosuppressants | AZATHIOPRINE 250 MG ORAL CAPSULE |
| 315447 | MA00 | Immunosuppressants | AZATHIOPRINE 50 MG |
| 329313 | MA00 | Immunosuppressants | AZATHIOPRINE 25 MG |
| 333493 | MA00 | Immunosuppressants | AZATHIOPRINE 10 MG/ML |
| 360265 | MA00 | Immunosuppressants | AZATHIOPRINE 75 MG |
| 360266 | MA00 | Immunosuppressants | AZATHIOPRINE 100 MG |

| | | | |
|---|---|---|---|
| 385562 | MA00 | Immunosuppressants | AZATHIOPRINE 10 MG |
| 434684 | MA00 | Immunosuppressants | AZATHIOPRINE 250 MG |
| 1789839 | MA00 | Immunosuppressants | AZATHIOPRINE INJECTION |
| 370973 | MA00 | Immunosuppressants | AZATHIOPRINE ORAL TABLET |
| 375463 | MA00 | Immunosuppressants | AZATHIOPRINE INJECTABLE SOLUTION |
| 434685 | MA00 | Immunosuppressants | AZATHIOPRINE ORAL CAPSULE |
| 1155000 | MA00 | Immunosuppressants | AZATHIOPRINE INJECTABLE PRODUCT |
| 1155001 | MA00 | Immunosuppressants | AZATHIOPRINE ORAL PRODUCT |
| 1155002 | MA00 | Immunosuppressants | AZATHIOPRINE PILL |
| 219912 | MA00 | Immunosuppressants | SIMULECT |
| 196102 | MA00 | Immunosuppressants | BASILIXIMAB |
| 1656646 | MA00 | Immunosuppressants | BASILIXIMAB 10 MG INJECTION [SIMULECT] |
| 1656650 | MA00 | Immunosuppressants | BASILIXIMAB 20 MG INJECTION [SIMULECT] |
| 227146 | MA00 | Immunosuppressants | BASILIXIMAB 4 MG/ML INJECTABLE SOLUTION [SIMULECT] |
| 1656644 | MA00 | Immunosuppressants | BASILIXIMAB 10 MG [SIMULECT] |
| 1656649 | MA00 | Immunosuppressants | BASILIXIMAB 20 MG [SIMULECT] |
| 574069 | MA00 | Immunosuppressants | BASILIXIMAB 4 MG/ML [SIMULECT] |
| 1656645 | MA00 | Immunosuppressants | BASILIXIMAB INJECTION [SIMULECT] |
| 362559 | MA00 | Immunosuppressants | BASILIXIMAB INJECTABLE SOLUTION [SIMULECT] |
| 1182652 | MA00 | Immunosuppressants | SIMULECT INJECTABLE PRODUCT |
| 1656643 | MA00 | Immunosuppressants | BASILIXIMAB 10 MG INJECTION |
| 1656648 | MA00 | Immunosuppressants | BASILIXIMAB 20 MG INJECTION |
| 313873 | MA00 | Immunosuppressants | BASILIXIMAB 10 MG INJECTABLE SOLUTION |
| 359248 | MA00 | Immunosuppressants | BASILIXIMAB 10 MG INJECTABLE SOLUTION |
| 1656641 | MA00 | Immunosuppressants | BASILIXIMAB 10 MG |
| 1656647 | MA00 | Immunosuppressants | BASILIXIMAB 20 MG |
| 333494 | MA00 | Immunosuppressants | BASILIXIMAB 10 MG |
| 360267 | MA00 | Immunosuppressants | BASILIXIMAB 10 MG |
| 1656642 | MA00 | Immunosuppressants | BASILIXIMAB INJECTION |
| 377149 | MA00 | Immunosuppressants | BASILIXIMAB INJECTABLE SOLUTION |
| 1159396 | MA00 | Immunosuppressants | BASILIXIMAB INJECTABLE PRODUCT |
| 202816 | MA00 | Immunosuppressants | SANDIMMUNE |
| 202817 | MA00 | Immunosuppressants | NEORAL |
| 219800 | MA00 | Immunosuppressants | SANGCYA |
| 261530 | MA00 | Immunosuppressants | GENGRAF |
| 349643 | MA00 | Immunosuppressants | EON BRAND OF CYCLOSPORINE |
| 349644 | MA00 | Immunosuppressants | SIDMAK BRAND OF CYCLOSPORINE |
| 583287 | MA00 | Immunosuppressants | CYSLOSPORINE |
| 3008 | MA00 | Immunosuppressants | CYCLOSPORINE |
| 236077 | MA00 | Immunosuppressants | CYCLOSPORINE, MODIFIED |
| 108843 | MA00 | Immunosuppressants | CYCLOSPORINE 100 MG ORAL CAPSULE [NEORAL] |
| 108844 | MA00 | Immunosuppressants | CYCLOSPORINE 50 MG ORAL CAPSULE [NEORAL] |
| 108845 | MA00 | Immunosuppressants | CYCLOSPORINE 25 MG ORAL CAPSULE [NEORAL] |
| 1732369 | MA00 | Immunosuppressants | CYCLOSPORINE 50 MG/ML INJECTION [SANDIMMUNE] |
| 1797521 | MA00 | Immunosuppressants | CYCLOSPORINE, MODIFIED 50 MG ORAL CAPSULE [GENGRAF] |
| 197552 | MA00 | Immunosuppressants | CYCLOSPORINE 100 MG ORAL CAPSULE [SANDIMMUNE] |
| 212810 | MA00 | Immunosuppressants | CYCLOSPORINE 25 MG ORAL CAPSULE [SANDIMMUNE] |
| 212821 | MA00 | Immunosuppressants | CYCLOSPORINE 50 MG ORAL CAPSULE [SANDIMMUNE] |
| 212844 | MA00 | Immunosuppressants | 5 ML CYCLOSPORINE 50 MG/ML INJECTION [SANDIMMUNE] |
| 212956 | MA00 | Immunosuppressants | CYCLOSPORINE 100 MG/ML ORAL SOLUTION [NEORAL] |
| 212957 | MA00 | Immunosuppressants | CYCLOSPORINE 100 MG/ML ORAL SOLUTION [SANDIMMUNE] |
| 212958 | MA00 | Immunosuppressants | CYCLOSPORINE 100 MG/ML ORAL SOLUTION [SANGCYA] |
| 309627 | MA00 | Immunosuppressants | CYCLOSPORINE 100 MG ORAL CAPSULE [EON BRAND OF CYCLOSPORINE] |
| 309628 | MA00 | Immunosuppressants | CYCLOSPORINE 100 MG ORAL CAPSULE [GENGRAF] |
| 309629 | MA00 | Immunosuppressants | CYCLOSPORINE 25 MG ORAL CAPSULE [GENGRAF] |
| 309630 | MA00 | Immunosuppressants | CYCLOSPORINE 100 MG ORAL CAPSULE [SIDMAK BRAND OF CYCLOSPORINE] |
| 309631 | MA00 | Immunosuppressants | CYCLOSPORINE 25 MG ORAL CAPSULE [SIDMAK BRAND OF CYCLOSPORINE] |
| 313937 | MA00 | Immunosuppressants | CYCLOSPORINE 25 MG ORAL CAPSULE [EON BRAND OF CYCLOSPORINE] |
| 358934 | MA00 | Immunosuppressants | CYCLOSPORINE 100 MG/ML ORAL SOLUTION [GENGRAF] |
| 358935 | MA00 | Immunosuppressants | CYCLOSPORINE 100 MG/ML ORAL SOLUTION [SIDMAK BRAND OF CYCLOSPORINE] |
| 583290 | MA00 | Immunosuppressants | CYCLOSPORINE 100 MG ORAL CAPSULE [CYSLOSPORINE] |
| 835835 | MA00 | Immunosuppressants | CYCLOSPORINE, MODIFIED 100 MG ORAL CAPSULE [GENGRAF] |
| 835892 | MA00 | Immunosuppressants | CYCLOSPORINE, MODIFIED 100 MG/ML ORAL SOLUTION [GENGRAF] |
| 835896 | MA00 | Immunosuppressants | CYCLOSPORINE, MODIFIED 25 MG ORAL CAPSULE [GENGRAF] |
| 835899 | MA00 | Immunosuppressants | CYCLOSPORINE, MODIFIED 100 MG/ML ORAL SOLUTION [NEORAL] |
| 835909 | MA00 | Immunosuppressants | CYCLOSPORINE, MODIFIED 25 MG ORAL CAPSULE [NEORAL] |
| 835911 | MA00 | Immunosuppressants | CYCLOSPORINE, MODIFIED 100 MG ORAL CAPSULE [NEORAL] |
| 1797520 | MA00 | Immunosuppressants | CYCLOSPORINE, MODIFIED 50 MG [GENGRAF] |
| 486331 | MA00 | Immunosuppressants | CYCLOSPORINE 100 MG/ML [SANDIMMUNE] |
| 564618 | MA00 | Immunosuppressants | CYCLOSPORINE 100 MG [NEORAL] |
| 564619 | MA00 | Immunosuppressants | CYCLOSPORINE 50 MG [NEORAL] |

| | | | |
|---|---|---|---|
| 564620 | MA00 | Immunosuppressants | CYCLOSPORINE 25 MG [NEORAL] |
| 565430 | MA00 | Immunosuppressants | CYCLOSPORINE 100 MG [SANDIMMUNE] |
| 572889 | MA00 | Immunosuppressants | CYCLOSPORINE 25 MG [SANDIMMUNE] |
| 572895 | MA00 | Immunosuppressants | CYCLOSPORINE 50 MG [SANDIMMUNE] |
| 572915 | MA00 | Immunosuppressants | CYCLOSPORINE 50 MG/ML [SANDIMMUNE] |
| 572918 | MA00 | Immunosuppressants | CYCLOSPORINE 100 MG/ML [NEORAL] |
| 572919 | MA00 | Immunosuppressants | CYCLOSPORINE 100 MG/ML [SANGCYA] |
| 575103 | MA00 | Immunosuppressants | CYCLOSPORINE 100 MG [EON BRAND OF CYCLOSPORINE] |
| 575104 | MA00 | Immunosuppressants | CYCLOSPORINE 100 MG [GENGRAF] |
| 575105 | MA00 | Immunosuppressants | CYCLOSPORINE 25 MG [GENGRAF] |
| 575106 | MA00 | Immunosuppressants | CYCLOSPORINE 100 MG [SIDMAK BRAND OF CYCLOSPORINE] |
| 575107 | MA00 | Immunosuppressants | CYCLOSPORINE 25 MG [SIDMAK BRAND OF CYCLOSPORINE] |
| 575200 | MA00 | Immunosuppressants | CYCLOSPORINE 25 MG [EON BRAND OF CYCLOSPORINE] |
| 576050 | MA00 | Immunosuppressants | CYCLOSPORINE 100 MG/ML [GENGRAF] |
| 576051 | MA00 | Immunosuppressants | CYCLOSPORINE 100 MG/ML [SIDMAK BRAND OF CYCLOSPORINE] |
| 583288 | MA00 | Immunosuppressants | CYCLOSPORINE 100 MG [CYSLOSPORINE] |
| 835787 | MA00 | Immunosuppressants | CYCLOSPORINE, MODIFIED 100 MG [GENGRAF] |
| 835887 | MA00 | Immunosuppressants | CYCLOSPORINE, MODIFIED 100 MG/ML [GENGRAF] |
| 835895 | MA00 | Immunosuppressants | CYCLOSPORINE, MODIFIED 25 MG [GENGRAF] |
| 835897 | MA00 | Immunosuppressants | CYCLOSPORINE, MODIFIED 100 MG/ML [NEORAL] |
| 835900 | MA00 | Immunosuppressants | CYCLOSPORINE, MODIFIED 100 MG [NEORAL] |
| 835901 | MA00 | Immunosuppressants | CYCLOSPORINE, MODIFIED 25 MG [NEORAL] |
| 1732366 | MA00 | Immunosuppressants | CYCLOSPORINE INJECTION [SANDIMMUNE] |
| 363913 | MA00 | Immunosuppressants | CYCLOSPORINE ORAL SOLUTION [SIDMAK BRAND OF CYCLOSPORINE] |
| 363914 | MA00 | Immunosuppressants | CYCLOSPORINE ORAL SOLUTION [GENGRAF] |
| 363942 | MA00 | Immunosuppressants | CYCLOSPORINE ORAL SOLUTION [NEORAL] |
| 364418 | MA00 | Immunosuppressants | CYCLOSPORINE ORAL SOLUTION [SANGCYA] |
| 366172 | MA00 | Immunosuppressants | CYCLOSPORINE ORAL CAPSULE [SIDMAK BRAND OF CYCLOSPORINE] |
| 366173 | MA00 | Immunosuppressants | CYCLOSPORINE ORAL CAPSULE [GENGRAF] |
| 366174 | MA00 | Immunosuppressants | CYCLOSPORINE ORAL CAPSULE [EON BRAND OF CYCLOSPORINE] |
| 366479 | MA00 | Immunosuppressants | CYCLOSPORINE ORAL CAPSULE [NEORAL] |
| 366488 | MA00 | Immunosuppressants | CYCLOSPORINE ORAL CAPSULE [SANDIMMUNE] |
| 583289 | MA00 | Immunosuppressants | CYCLOSPORINE ORAL CAPSULE [CYSLOSPORINE] |
| 94034 | MA00 | Immunosuppressants | CYCLOSPORINE INJECTABLE SOLUTION [SANDIMMUNE] |
| 94035 | MA00 | Immunosuppressants | CYCLOSPORINE ORAL SOLUTION [SANDIMMUNE] |
| 1169824 | MA00 | Immunosuppressants | GENGRAF ORAL LIQUID PRODUCT |
| 1169825 | MA00 | Immunosuppressants | GENGRAF ORAL PRODUCT |
| 1169826 | MA00 | Immunosuppressants | GENGRAF PILL |
| 1174462 | MA00 | Immunosuppressants | CYSLOSPORINE ORAL PRODUCT |
| 1174463 | MA00 | Immunosuppressants | CYSLOSPORINE PILL |
| 1181672 | MA00 | Immunosuppressants | NEORAL ORAL LIQUID PRODUCT |
| 1181673 | MA00 | Immunosuppressants | NEORAL ORAL PRODUCT |
| 1181674 | MA00 | Immunosuppressants | NEORAL PILL |
| 1185408 | MA00 | Immunosuppressants | SANDIMMUNE INJECTABLE PRODUCT |
| 1185409 | MA00 | Immunosuppressants | SANDIMMUNE ORAL LIQUID PRODUCT |
| 1185410 | MA00 | Immunosuppressants | SANDIMMUNE ORAL PRODUCT |
| 1185411 | MA00 | Immunosuppressants | SANDIMMUNE PILL |
| 1185971 | MA00 | Immunosuppressants | SANGCYA ORAL LIQUID PRODUCT |
| 1185972 | MA00 | Immunosuppressants | SANGCYA ORAL PRODUCT |
| 1732368 | MA00 | Immunosuppressants | CYCLOSPORINE 50 MG/ML INJECTION |
| 197553 | MA00 | Immunosuppressants | CYCLOSPORINE 25 MG ORAL CAPSULE |
| 205168 | MA00 | Immunosuppressants | CYCLOSPORINE 50 MG ORAL CAPSULE |
| 205175 | MA00 | Immunosuppressants | 5 ML CYCLOSPORINE 50 MG/ML INJECTION |
| 226692 | MA00 | Immunosuppressants | CYCLOSPORINE 10 MG ORAL CAPSULE |
| 241834 | MA00 | Immunosuppressants | CYCLOSPORINE, MODIFIED 100 MG ORAL CAPSULE |
| 309632 | MA00 | Immunosuppressants | CYCLOSPORINE 100 MG/ML ORAL SOLUTION |
| 328160 | MA00 | Immunosuppressants | CYCLOSPORINE 100 MG ORAL CAPSULE |
| 348431 | MA00 | Immunosuppressants | CYCLOSPORINE 1000 MG/ML ORAL SOLUTION |
| 670177 | MA00 | Immunosuppressants | CYCLOSPORINE 50 MG/ML ORAL SOLUTION |
| 835886 | MA00 | Immunosuppressants | CYCLOSPORINE, MODIFIED 100 MG/ML ORAL SOLUTION |
| 835894 | MA00 | Immunosuppressants | CYCLOSPORINE, MODIFIED 25 MG ORAL CAPSULE |
| 835919 | MA00 | Immunosuppressants | CYCLOSPORINE, MODIFIED 10 MG ORAL CAPSULE |
| 835925 | MA00 | Immunosuppressants | CYCLOSPORINE, MODIFIED 50 MG ORAL CAPSULE |
| 315749 | MA00 | Immunosuppressants | CYCLOSPORINE 100 MG |
| 315750 | MA00 | Immunosuppressants | CYCLOSPORINE 100 MG/ML |
| 315751 | MA00 | Immunosuppressants | CYCLOSPORINE 25 MG |
| 315752 | MA00 | Immunosuppressants | CYCLOSPORINE 50 MG |
| 315753 | MA00 | Immunosuppressants | CYCLOSPORINE 50 MG/ML |
| 334294 | MA00 | Immunosuppressants | CYCLOSPORINE, MODIFIED 100 MG |
| 358763 | MA00 | Immunosuppressants | CYCLOSPORINE 1000 MG/ML |
| 385602 | MA00 | Immunosuppressants | CYCLOSPORINE 10 MG |

| | | | |
|---|---|---|---|
| 835885 | MA00 | Immunosuppressants | CYCLOSPORINE, MODIFIED 100 MG/ML |
| 835893 | MA00 | Immunosuppressants | CYCLOSPORINE, MODIFIED 25 MG |
| 835918 | MA00 | Immunosuppressants | CYCLOSPORINE, MODIFIED 10 MG |
| 835924 | MA00 | Immunosuppressants | CYCLOSPORINE, MODIFIED 50 MG |
| 1732364 | MA00 | Immunosuppressants | CYCLOSPORINE INJECTION |
| 371666 | MA00 | Immunosuppressants | CYCLOSPORINE ORAL CAPSULE |
| 371667 | MA00 | Immunosuppressants | CYCLOSPORINE ORAL SOLUTION |
| 374573 | MA00 | Immunosuppressants | CYCLOSPORINE INJECTABLE SOLUTION |
| 1156186 | MA00 | Immunosuppressants | CYCLOSPORINE INJECTABLE PRODUCT |
| 1156188 | MA00 | Immunosuppressants | CYCLOSPORINE ORAL LIQUID PRODUCT |
| 1156189 | MA00 | Immunosuppressants | CYCLOSPORINE ORAL PRODUCT |
| 1156190 | MA00 | Immunosuppressants | CYCLOSPORINE PILL |
| 1795335 | MA00 | Immunosuppressants | ZINBRYTA |
| 81770 | MA00 | Immunosuppressants | ZENAPAX |
| 190353 | MA00 | Immunosuppressants | DACLIZUMAB |
| 1795339 | MA00 | Immunosuppressants | 1 ML DACLIZUMAB 150 MG/ML PREFILLED SYRINGE [ZINBRYTA] |
| 1795341 | MA00 | Immunosuppressants | DACLIZUMAB 150 MG/ML PREFILLED SYRINGE [ZINBRYTA] |
| 1925211 | MA00 | Immunosuppressants | 1 ML DACLIZUMAB 150 MG/ML AUTO-INJECTOR [ZINBRYTA] |
| 1925213 | MA00 | Immunosuppressants | DACLIZUMAB 150 MG/ML AUTO-INJECTOR [ZINBRYTA] |
| 213177 | MA00 | Immunosuppressants | DACLIZUMAB 5 MG/ML INJECTABLE SOLUTION [ZENAPAX] |
| 1795336 | MA00 | Immunosuppressants | DACLIZUMAB 150 MG/ML [ZINBRYTA] |
| 573100 | MA00 | Immunosuppressants | DACLIZUMAB 5 MG/ML [ZENAPAX] |
| 1795337 | MA00 | Immunosuppressants | DACLIZUMAB PREFILLED SYRINGE [ZINBRYTA] |
| 1925210 | MA00 | Immunosuppressants | DACLIZUMAB AUTO-INJECTOR [ZINBRYTA] |
| 362855 | MA00 | Immunosuppressants | DACLIZUMAB INJECTABLE SOLUTION [ZENAPAX] |
| 1187246 | MA00 | Immunosuppressants | ZENAPAX INJECTABLE PRODUCT |
| 1795338 | MA00 | Immunosuppressants | ZINBRYTA INJECTABLE PRODUCT |
| 1795334 | MA00 | Immunosuppressants | 1 ML DACLIZUMAB 150 MG/ML PREFILLED SYRINGE |
| 1795340 | MA00 | Immunosuppressants | DACLIZUMAB 150 MG/ML PREFILLED SYRINGE |
| 1925209 | MA00 | Immunosuppressants | 1 ML DACLIZUMAB 150 MG/ML AUTO-INJECTOR |
| 1925212 | MA00 | Immunosuppressants | DACLIZUMAB 150 MG/ML AUTO-INJECTOR |
| 200171 | MA00 | Immunosuppressants | DACLIZUMAB 5 MG/ML INJECTABLE SOLUTION |
| 1795332 | MA00 | Immunosuppressants | DACLIZUMAB 150 MG/ML |
| 315755 | MA00 | Immunosuppressants | DACLIZUMAB 5 MG/ML |
| 1795333 | MA00 | Immunosuppressants | DACLIZUMAB PREFILLED SYRINGE |
| 1925208 | MA00 | Immunosuppressants | DACLIZUMAB AUTO-INJECTOR |
| 377159 | MA00 | Immunosuppressants | DACLIZUMAB INJECTABLE SOLUTION |
| 1151834 | MA00 | Immunosuppressants | DACLIZUMAB INJECTABLE PRODUCT |
| 2104612 | MA00 | Immunosuppressants | GAMIFANT |
| 2104604 | MA00 | Immunosuppressants | EMAPALUMAB |
| 2104603 | MA00 | Immunosuppressants | EMAPALUMAB-LZSG |
| 2104616 | MA00 | Immunosuppressants | 2 ML EMAPALUMAB-LZSG 5 MG/ML INJECTION [GAMIFANT] |
| 2104618 | MA00 | Immunosuppressants | EMAPALUMAB-LZSG 5 MG/ML INJECTION [GAMIFANT] |
| 2104622 | MA00 | Immunosuppressants | 10 ML EMAPALUMAB-LZSG 5 MG/ML INJECTION [GAMIFANT] |
| 2104613 | MA00 | Immunosuppressants | EMAPALUMAB-LZSG 5 MG/ML [GAMIFANT] |
| 2104614 | MA00 | Immunosuppressants | EMAPALUMAB INJECTION [GAMIFANT] |
| 2104615 | MA00 | Immunosuppressants | GAMIFANT INJECTABLE PRODUCT |
| 2104611 | MA00 | Immunosuppressants | 2 ML EMAPALUMAB-LZSG 5 MG/ML INJECTION |
| 2104617 | MA00 | Immunosuppressants | EMAPALUMAB-LZSG 5 MG/ML INJECTION |
| 2104621 | MA00 | Immunosuppressants | 10 ML EMAPALUMAB-LZSG 5 MG/ML INJECTION |
| 2104608 | MA00 | Immunosuppressants | EMAPALUMAB-LZSG 5 MG/ML |
| 2104610 | MA00 | Immunosuppressants | EMAPALUMAB INJECTION |
| 2104609 | MA00 | Immunosuppressants | EMAPALUMAB INJECTABLE PRODUCT |
| 845509 | MA00 | Immunosuppressants | AFINITOR |
| 977428 | MA00 | Immunosuppressants | ZORTRESS |
| 845508 | MA00 | Immunosuppressants | {28 (EVEROLIMUS 10 MG ORAL TABLET) } PACK |
| 845516 | MA00 | Immunosuppressants | {28 (EVEROLIMUS 5 MG ORAL TABLET) } PACK |
| 141704 | MA00 | Immunosuppressants | EVEROLIMUS |
| 1119402 | MA00 | Immunosuppressants | EVEROLIMUS 7.5 MG ORAL TABLET [AFINITOR] |
| 1310138 | MA00 | Immunosuppressants | EVEROLIMUS 2 MG TABLET FOR ORAL SUSPENSION [AFINITOR] |
| 1310144 | MA00 | Immunosuppressants | EVEROLIMUS 3 MG TABLET FOR ORAL SUSPENSION [AFINITOR] |
| 1310147 | MA00 | Immunosuppressants | EVEROLIMUS 5 MG TABLET FOR ORAL SUSPENSION [AFINITOR] |
| 2056897 | MA00 | Immunosuppressants | EVEROLIMUS 1 MG ORAL TABLET [ZORTRESS] |
| 845512 | MA00 | Immunosuppressants | EVEROLIMUS 10 MG ORAL TABLET [AFINITOR] |
| 845518 | MA00 | Immunosuppressants | EVEROLIMUS 5 MG ORAL TABLET [AFINITOR] |
| 977431 | MA00 | Immunosuppressants | EVEROLIMUS 0.25 MG ORAL TABLET [ZORTRESS] |
| 977436 | MA00 | Immunosuppressants | EVEROLIMUS 0.5 MG ORAL TABLET [ZORTRESS] |
| 977440 | MA00 | Immunosuppressants | EVEROLIMUS 0.75 MG ORAL TABLET [ZORTRESS] |
| 998191 | MA00 | Immunosuppressants | EVEROLIMUS 2.5 MG ORAL TABLET [AFINITOR] |
| 1119401 | MA00 | Immunosuppressants | EVEROLIMUS 7.5 MG [AFINITOR] |
| 1310135 | MA00 | Immunosuppressants | EVEROLIMUS 0.08 MG/ML [AFINITOR] |

| | | | |
|---|---|---|---|
| 1310143 | MA00 | Immunosuppressants | EVEROLIMUS 0.12 MG/ML [AFINITOR] |
| 1310146 | MA00 | Immunosuppressants | EVEROLIMUS 0.2 MG/ML [AFINITOR] |
| 1869515 | MA00 | Immunosuppressants | EVEROLIMUS 2 MG [AFINITOR] |
| 1869520 | MA00 | Immunosuppressants | EVEROLIMUS 3 MG [AFINITOR] |
| 2056896 | MA00 | Immunosuppressants | EVEROLIMUS 1 MG [ZORTRESS] |
| 845510 | MA00 | Immunosuppressants | EVEROLIMUS 10 MG [AFINITOR] |
| 845517 | MA00 | Immunosuppressants | EVEROLIMUS 5 MG [AFINITOR] |
| 977429 | MA00 | Immunosuppressants | EVEROLIMUS 0.25 MG [ZORTRESS] |
| 977435 | MA00 | Immunosuppressants | EVEROLIMUS 0.5 MG [ZORTRESS] |
| 977439 | MA00 | Immunosuppressants | EVEROLIMUS 0.75 MG [ZORTRESS] |
| 998190 | MA00 | Immunosuppressants | EVEROLIMUS 2.5 MG [AFINITOR] |
| 1310136 | MA00 | Immunosuppressants | EVEROLIMUS ORAL SUSPENSION [AFINITOR] |
| 1869516 | MA00 | Immunosuppressants | EVEROLIMUS TABLET FOR ORAL SUSPENSION [AFINITOR] |
| 845511 | MA00 | Immunosuppressants | EVEROLIMUS ORAL TABLET [AFINITOR] |
| 977430 | MA00 | Immunosuppressants | EVEROLIMUS ORAL TABLET [ZORTRESS] |
| 1172066 | MA00 | Immunosuppressants | AFINITOR ORAL PRODUCT |
| 1172067 | MA00 | Immunosuppressants | AFINITOR PILL |
| 1186316 | MA00 | Immunosuppressants | ZORTRESS ORAL PRODUCT |
| 1186317 | MA00 | Immunosuppressants | ZORTRESS PILL |
| 1310137 | MA00 | Immunosuppressants | AFINITOR ORAL LIQUID PRODUCT |
| 1119400 | MA00 | Immunosuppressants | EVEROLIMUS 7.5 MG ORAL TABLET |
| 1308428 | MA00 | Immunosuppressants | EVEROLIMUS 2 MG TABLET FOR ORAL SUSPENSION |
| 1308430 | MA00 | Immunosuppressants | EVEROLIMUS 3 MG TABLET FOR ORAL SUSPENSION |
| 1308432 | MA00 | Immunosuppressants | EVEROLIMUS 5 MG TABLET FOR ORAL SUSPENSION |
| 2056895 | MA00 | Immunosuppressants | EVEROLIMUS 1 MG ORAL TABLET |
| 845507 | MA00 | Immunosuppressants | EVEROLIMUS 10 MG ORAL TABLET |
| 845515 | MA00 | Immunosuppressants | EVEROLIMUS 5 MG ORAL TABLET |
| 977427 | MA00 | Immunosuppressants | EVEROLIMUS 0.25 MG ORAL TABLET |
| 977434 | MA00 | Immunosuppressants | EVEROLIMUS 0.5 MG ORAL TABLET |
| 977438 | MA00 | Immunosuppressants | EVEROLIMUS 0.75 MG ORAL TABLET |
| 998189 | MA00 | Immunosuppressants | EVEROLIMUS 2.5 MG ORAL TABLET |
| 1119399 | MA00 | Immunosuppressants | EVEROLIMUS 7.5 MG |
| 1308425 | MA00 | Immunosuppressants | EVEROLIMUS 0.08 MG/ML |
| 1308429 | MA00 | Immunosuppressants | EVEROLIMUS 0.12 MG/ML |
| 1308431 | MA00 | Immunosuppressants | EVEROLIMUS 0.2 MG/ML |
| 1869512 | MA00 | Immunosuppressants | EVEROLIMUS 2 MG |
| 1869518 | MA00 | Immunosuppressants | EVEROLIMUS 3 MG |
| 2056894 | MA00 | Immunosuppressants | EVEROLIMUS 1 MG |
| 845505 | MA00 | Immunosuppressants | EVEROLIMUS 10 MG |
| 845514 | MA00 | Immunosuppressants | EVEROLIMUS 5 MG |
| 977426 | MA00 | Immunosuppressants | EVEROLIMUS 0.25 MG |
| 977433 | MA00 | Immunosuppressants | EVEROLIMUS 0.5 MG |
| 977437 | MA00 | Immunosuppressants | EVEROLIMUS 0.75 MG |
| 998188 | MA00 | Immunosuppressants | EVEROLIMUS 2.5 MG |
| 1308427 | MA00 | Immunosuppressants | EVEROLIMUS ORAL SUSPENSION |
| 1869513 | MA00 | Immunosuppressants | EVEROLIMUS TABLET FOR ORAL SUSPENSION |
| 845506 | MA00 | Immunosuppressants | EVEROLIMUS ORAL TABLET |
| 1163786 | MA00 | Immunosuppressants | EVEROLIMUS ORAL PRODUCT |
| 1163787 | MA00 | Immunosuppressants | EVEROLIMUS PILL |
| 1308426 | MA00 | Immunosuppressants | EVEROLIMUS ORAL LIQUID PRODUCT |
| 1536485 | MA00 | Immunosuppressants | PURIXAN |
| 219513 | MA00 | Immunosuppressants | PURINETHOL |
| 103 | MA00 | Immunosuppressants | MERCAPTOPURINE |
| 1546028 | MA00 | Immunosuppressants | MERCAPTOPURINE ANHYDROUS |
| 1536490 | MA00 | Immunosuppressants | MERCAPTOPURINE 20 MG/ML ORAL SUSPENSION [PURIXAN] |
| 206788 | MA00 | Immunosuppressants | MERCAPTOPURINE 50 MG ORAL TABLET [PURINETHOL] |
| 1536486 | MA00 | Immunosuppressants | MERCAPTOPURINE 20 MG/ML [PURIXAN] |
| 567599 | MA00 | Immunosuppressants | MERCAPTOPURINE 50 MG [PURINETHOL] |
| 1536487 | MA00 | Immunosuppressants | MERCAPTOPURINE ORAL SUSPENSION [PURIXAN] |
| 368861 | MA00 | Immunosuppressants | MERCAPTOPURINE ORAL TABLET [PURINETHOL] |
| 1183583 | MA00 | Immunosuppressants | PURINETHOL ORAL PRODUCT |
| 1183584 | MA00 | Immunosuppressants | PURINETHOL PILL |
| 1536488 | MA00 | Immunosuppressants | PURIXAN ORAL LIQUID PRODUCT |
| 1536489 | MA00 | Immunosuppressants | PURIXAN ORAL PRODUCT |
| 1536484 | MA00 | Immunosuppressants | MERCAPTOPURINE 20 MG/ML ORAL SUSPENSION |
| 197931 | MA00 | Immunosuppressants | MERCAPTOPURINE 50 MG ORAL TABLET |
| 199766 | MA00 | Immunosuppressants | MERCAPTOPURINE 10 MG ORAL TABLET |
| 1536481 | MA00 | Immunosuppressants | MERCAPTOPURINE 20 MG/ML |
| 316246 | MA00 | Immunosuppressants | MERCAPTOPURINE 50 MG |
| 332204 | MA00 | Immunosuppressants | MERCAPTOPURINE 10 MG |
| 1536483 | MA00 | Immunosuppressants | MERCAPTOPURINE ORAL SUSPENSION |

| | | | |
|---|---|---|---|
| 372782 | MA00 | Immunosuppressants | MERCAPTOPURINE ORAL TABLET |
| 1165710 | MA00 | Immunosuppressants | MERCAPTOPURINE ORAL PRODUCT |
| 1165711 | MA00 | Immunosuppressants | MERCAPTOPURINE PILL |
| 1536482 | MA00 | Immunosuppressants | MERCAPTOPURINE ORAL LIQUID PRODUCT |
| 1441403 | MA00 | Immunosuppressants | OTREXUP |
| 152008 | MA00 | Immunosuppressants | MAXTREX |
| 1544379 | MA00 | Immunosuppressants | RASUVO |
| 1921593 | MA00 | Immunosuppressants | XATMEP |
| 284900 | MA00 | Immunosuppressants | TREXALL |
| 346424 | MA00 | Immunosuppressants | EMTEXATE |
| 491604 | MA00 | Immunosuppressants | RHEUMATREX |
| 1150118 | MA00 | Immunosuppressants | {8 (METHOTREXATE 2.5 MG ORAL TABLET [RHEUMATREX]) } PACK [RHEUMATREX DOSE PACK 8] |
| 1541216 | MA00 | Immunosuppressants | {12 (METHOTREXATE 2.5 MG ORAL TABLET [RHEUMATREX]) } PACK [RHEUMATREX DOSE PACK 12] |
| 1541218 | MA00 | Immunosuppressants | {16 (METHOTREXATE 2.5 MG ORAL TABLET [RHEUMATREX]) } PACK [RHEUMATREX DOSE PACK 16] |
| 1541221 | MA00 | Immunosuppressants | {20 (METHOTREXATE 2.5 MG ORAL TABLET [RHEUMATREX]) } PACK [RHEUMATREX DOSE PACK 20] |
| 1541224 | MA00 | Immunosuppressants | {24 (METHOTREXATE 2.5 MG ORAL TABLET [RHEUMATREX]) } PACK [RHEUMATREX DOSE PACK 24] |
| 1541226 | MA00 | Immunosuppressants | {28 (METHOTREXATE 2.5 MG ORAL TABLET [RHEUMATREX]) } PACK [RHEUMATREX DOSE PACK 28] |
| 1541228 | MA00 | Immunosuppressants | {32 (METHOTREXATE 2.5 MG ORAL TABLET [RHEUMATREX]) } PACK [RHEUMATREX DOSE PACK 32] |
| 1150117 | MA00 | Immunosuppressants | {8 (METHOTREXATE 2.5 MG ORAL TABLET) } PACK |
| 1541215 | MA00 | Immunosuppressants | {12 (METHOTREXATE 2.5 MG ORAL TABLET) } PACK |
| 1541217 | MA00 | Immunosuppressants | {16 (METHOTREXATE 2.5 MG ORAL TABLET) } PACK |
| 1541220 | MA00 | Immunosuppressants | {20 (METHOTREXATE 2.5 MG ORAL TABLET) } PACK |
| 1541222 | MA00 | Immunosuppressants | {24 (METHOTREXATE 2.5 MG ORAL TABLET) } PACK |
| 1541225 | MA00 | Immunosuppressants | {28 (METHOTREXATE 2.5 MG ORAL TABLET) } PACK |
| 1541227 | MA00 | Immunosuppressants | {32 (METHOTREXATE 2.5 MG ORAL TABLET) } PACK |
| 6851 | MA00 | Immunosuppressants | METHOTREXATE |
| 287734 | MA00 | Immunosuppressants | METHOTREXATE SODIUM |
| 102953 | MA00 | Immunosuppressants | METHOTREXATE 10 MG ORAL TABLET [EMTEXATE] |
| 102954 | MA00 | Immunosuppressants | METHOTREXATE 2.5 MG/ML INJECTABLE SOLUTION [EMTEXATE] |
| 102955 | MA00 | Immunosuppressants | METHOTREXATE 25 MG/ML INJECTABLE SOLUTION [EMTEXATE] |
| 102956 | MA00 | Immunosuppressants | METHOTREXATE 100 MG/ML INJECTABLE SOLUTION [EMTEXATE] |
| 102957 | MA00 | Immunosuppressants | METHOTREXATE 50 MG/ML INJECTABLE SOLUTION [EMTEXATE] |
| 102958 | MA00 | Immunosuppressants | METHOTREXATE 2.5 MG/ML INJECTABLE SOLUTION [MAXTREX] |
| 102959 | MA00 | Immunosuppressants | METHOTREXATE 25 MG/ML INJECTABLE SOLUTION [MAXTREX] |
| 105587 | MA00 | Immunosuppressants | METHOTREXATE 2.5 MG ORAL TABLET [MAXTREX] |
| 105588 | MA00 | Immunosuppressants | METHOTREXATE 10 MG ORAL TABLET [MAXTREX] |
| 1441407 | MA00 | Immunosuppressants | 0.4 ML METHOTREXATE 25 MG/ML AUTO-INJECTOR [OTREXUP] |
| 1441409 | MA00 | Immunosuppressants | METHOTREXATE 25 MG/ML AUTO-INJECTOR [OTREXUP] |
| 1441413 | MA00 | Immunosuppressants | 0.4 ML METHOTREXATE 37.5 MG/ML AUTO-INJECTOR [OTREXUP] |
| 1441415 | MA00 | Immunosuppressants | METHOTREXATE 37.5 MG/ML AUTO-INJECTOR [OTREXUP] |
| 1441418 | MA00 | Immunosuppressants | 0.4 ML METHOTREXATE 50 MG/ML AUTO-INJECTOR [OTREXUP] |
| 1441420 | MA00 | Immunosuppressants | METHOTREXATE 50 MG/ML AUTO-INJECTOR [OTREXUP] |
| 1441424 | MA00 | Immunosuppressants | 0.4 ML METHOTREXATE 62.5 MG/ML AUTO-INJECTOR [OTREXUP] |
| 1441426 | MA00 | Immunosuppressants | METHOTREXATE 62.5 MG/ML AUTO-INJECTOR [OTREXUP] |
| 1544383 | MA00 | Immunosuppressants | 0.2 ML METHOTREXATE 50 MG/ML AUTO-INJECTOR [RASUVO] |
| 1544384 | MA00 | Immunosuppressants | METHOTREXATE 50 MG/ML AUTO-INJECTOR [RASUVO] |
| 1544386 | MA00 | Immunosuppressants | 0.25 ML METHOTREXATE 50 MG/ML AUTO-INJECTOR [RASUVO] |
| 1544388 | MA00 | Immunosuppressants | 0.3 ML METHOTREXATE 50 MG/ML AUTO-INJECTOR [RASUVO] |
| 1544390 | MA00 | Immunosuppressants | 0.35 ML METHOTREXATE 50 MG/ML AUTO-INJECTOR [RASUVO] |
| 1544394 | MA00 | Immunosuppressants | 0.4 ML METHOTREXATE 50 MG/ML AUTO-INJECTOR [RASUVO] |
| 1544396 | MA00 | Immunosuppressants | 0.45 ML METHOTREXATE 50 MG/ML AUTO-INJECTOR [RASUVO] |
| 1544398 | MA00 | Immunosuppressants | 0.5 ML METHOTREXATE 50 MG/ML AUTO-INJECTOR [RASUVO] |
| 1544400 | MA00 | Immunosuppressants | 0.55 ML METHOTREXATE 50 MG/ML AUTO-INJECTOR [RASUVO] |
| 1544402 | MA00 | Immunosuppressants | 0.6 ML METHOTREXATE 50 MG/ML AUTO-INJECTOR [RASUVO] |
| 1544404 | MA00 | Immunosuppressants | 0.15 ML METHOTREXATE 50 MG/ML AUTO-INJECTOR [RASUVO] |
| 1594759 | MA00 | Immunosuppressants | 0.4 ML METHOTREXATE 18.8 MG/ML AUTO-INJECTOR [OTREXUP] |
| 1594761 | MA00 | Immunosuppressants | METHOTREXATE 18.8 MG/ML AUTO-INJECTOR [OTREXUP] |
| 1747181 | MA00 | Immunosuppressants | 0.4 ML METHOTREXATE 31.3 MG/ML AUTO-INJECTOR [OTREXUP] |
| 1747183 | MA00 | Immunosuppressants | METHOTREXATE 31.3 MG/ML AUTO-INJECTOR [OTREXUP] |
| 1747187 | MA00 | Immunosuppressants | 0.4 ML METHOTREXATE 43.8 MG/ML AUTO-INJECTOR [OTREXUP] |
| 1747189 | MA00 | Immunosuppressants | METHOTREXATE 43.8 MG/ML AUTO-INJECTOR [OTREXUP] |
| 1747194 | MA00 | Immunosuppressants | 0.4 ML METHOTREXATE 56.3 MG/ML AUTO-INJECTOR [OTREXUP] |
| 1747196 | MA00 | Immunosuppressants | METHOTREXATE 56.3 MG/ML AUTO-INJECTOR [OTREXUP] |
| 1921598 | MA00 | Immunosuppressants | METHOTREXATE 2.5 MG/ML ORAL SOLUTION [XATMEP] |

| | | | |
|---|---|---|---|
| 207033 | MA00 | Immunosuppressants | METHOTREXATE 25 MG/ML INJECTABLE SOLUTION [FOLEX PFS] |
| 284592 | MA00 | Immunosuppressants | METHOTREXATE 5 MG ORAL TABLET [TREXALL] |
| 284593 | MA00 | Immunosuppressants | METHOTREXATE 10 MG ORAL TABLET [TREXALL] |
| 284594 | MA00 | Immunosuppressants | METHOTREXATE 7.5 MG ORAL TABLET [TREXALL] |
| 284595 | MA00 | Immunosuppressants | METHOTREXATE 15 MG ORAL TABLET [TREXALL] |
| 579782 | MA00 | Immunosuppressants | METHOTREXATE 2.5 MG ORAL TABLET [RHEUMATREX] |
| 1441404 | MA00 | Immunosuppressants | METHOTREXATE 25 MG/ML [OTREXUP] |
| 1441412 | MA00 | Immunosuppressants | METHOTREXATE 37.5 MG/ML [OTREXUP] |
| 1441417 | MA00 | Immunosuppressants | METHOTREXATE 50 MG/ML [OTREXUP] |
| 1441423 | MA00 | Immunosuppressants | METHOTREXATE 62.5 MG/ML [OTREXUP] |
| 1544380 | MA00 | Immunosuppressants | METHOTREXATE 50 MG/ML [RASUVO] |
| 1594758 | MA00 | Immunosuppressants | METHOTREXATE 18.75 MG/ML [OTREXUP] |
| 1659721 | MA00 | Immunosuppressants | METHOTREXATE 18.8 MG/ML [OTREXUP] |
| 1747180 | MA00 | Immunosuppressants | METHOTREXATE 31.3 MG/ML [OTREXUP] |
| 1747186 | MA00 | Immunosuppressants | METHOTREXATE 43.8 MG/ML [OTREXUP] |
| 1747193 | MA00 | Immunosuppressants | METHOTREXATE 56.3 MG/ML [OTREXUP] |
| 1921594 | MA00 | Immunosuppressants | METHOTREXATE 2.5 MG/ML [XATMEP] |
| 563189 | MA00 | Immunosuppressants | METHOTREXATE 10 MG [EMTEXATE] |
| 563190 | MA00 | Immunosuppressants | METHOTREXATE 2.5 MG/ML [EMTEXATE] |
| 563191 | MA00 | Immunosuppressants | METHOTREXATE 25 MG/ML [EMTEXATE] |
| 563192 | MA00 | Immunosuppressants | METHOTREXATE 100 MG/ML [EMTEXATE] |
| 563193 | MA00 | Immunosuppressants | METHOTREXATE 50 MG/ML [EMTEXATE] |
| 563194 | MA00 | Immunosuppressants | METHOTREXATE 2.5 MG/ML [MAXTREX] |
| 563195 | MA00 | Immunosuppressants | METHOTREXATE 25 MG/ML [MAXTREX] |
| 564104 | MA00 | Immunosuppressants | METHOTREXATE 2.5 MG [MAXTREX] |
| 564105 | MA00 | Immunosuppressants | METHOTREXATE 10 MG [MAXTREX] |
| 567829 | MA00 | Immunosuppressants | METHOTREXATE 25 MG/ML [FOLEX PFS] |
| 575019 | MA00 | Immunosuppressants | METHOTREXATE 5 MG [TREXALL] |
| 575020 | MA00 | Immunosuppressants | METHOTREXATE 10 MG [TREXALL] |
| 575021 | MA00 | Immunosuppressants | METHOTREXATE 7.5 MG [TREXALL] |
| 575022 | MA00 | Immunosuppressants | METHOTREXATE 15 MG [TREXALL] |
| 579780 | MA00 | Immunosuppressants | METHOTREXATE 2.5 MG [RHEUMATREX] |
| 1441405 | MA00 | Immunosuppressants | METHOTREXATE PREFILLED SYRINGE [OTREXUP] |
| 1544381 | MA00 | Immunosuppressants | METHOTREXATE PREFILLED SYRINGE [RASUVO] |
| 1655205 | MA00 | Immunosuppressants | METHOTREXATE AUTO-INJECTOR [OTREXUP] |
| 1655252 | MA00 | Immunosuppressants | METHOTREXATE AUTO-INJECTOR [RASUVO] |
| 1921595 | MA00 | Immunosuppressants | METHOTREXATE ORAL SOLUTION [XATMEP] |
| 362547 | MA00 | Immunosuppressants | METHOTREXATE INJECTABLE SOLUTION [EMTEXATE] |
| 362736 | MA00 | Immunosuppressants | METHOTREXATE INJECTABLE SOLUTION [MAXTREX] |
| 363432 | MA00 | Immunosuppressants | METHOTREXATE INJECTABLE SOLUTION [FOLEX PFS] |
| 368138 | MA00 | Immunosuppressants | METHOTREXATE ORAL TABLET [TREXALL] |
| 368588 | MA00 | Immunosuppressants | METHOTREXATE ORAL TABLET [MAXTREX] |
| 368665 | MA00 | Immunosuppressants | METHOTREXATE ORAL TABLET [EMTEXATE] |
| 579781 | MA00 | Immunosuppressants | METHOTREXATE ORAL TABLET [RHEUMATREX] |
| 1175944 | MA00 | Immunosuppressants | EMTEXATE INJECTABLE PRODUCT |
| 1175945 | MA00 | Immunosuppressants | EMTEXATE ORAL PRODUCT |
| 1175946 | MA00 | Immunosuppressants | EMTEXATE PILL |
| 1179677 | MA00 | Immunosuppressants | TREXALL ORAL PRODUCT |
| 1179678 | MA00 | Immunosuppressants | TREXALL PILL |
| 1182738 | MA00 | Immunosuppressants | MAXTREX INJECTABLE PRODUCT |
| 1182739 | MA00 | Immunosuppressants | MAXTREX ORAL PRODUCT |
| 1182740 | MA00 | Immunosuppressants | MAXTREX PILL |
| 1183068 | MA00 | Immunosuppressants | RHEUMATREX ORAL PRODUCT |
| 1183069 | MA00 | Immunosuppressants | RHEUMATREX PILL |
| 1441406 | MA00 | Immunosuppressants | OTREXUP INJECTABLE PRODUCT |
| 1544382 | MA00 | Immunosuppressants | RASUVO INJECTABLE PRODUCT |
| 1921596 | MA00 | Immunosuppressants | XATMEP ORAL LIQUID PRODUCT |
| 1921597 | MA00 | Immunosuppressants | XATMEP ORAL PRODUCT |
| 105585 | MA00 | Immunosuppressants | METHOTREXATE 2.5 MG ORAL TABLET |
| 105586 | MA00 | Immunosuppressants | METHOTREXATE 10 MG ORAL TABLET |
| 105589 | MA00 | Immunosuppressants | METHOTREXATE 100 MG/ML INJECTABLE SOLUTION |
| 1441402 | MA00 | Immunosuppressants | 0.4 ML METHOTREXATE 25 MG/ML AUTO-INJECTOR |
| 1441408 | MA00 | Immunosuppressants | METHOTREXATE 25 MG/ML AUTO-INJECTOR |
| 1441411 | MA00 | Immunosuppressants | 0.4 ML METHOTREXATE 37.5 MG/ML AUTO-INJECTOR |
| 1441414 | MA00 | Immunosuppressants | METHOTREXATE 37.5 MG/ML AUTO-INJECTOR |
| 1441416 | MA00 | Immunosuppressants | 0.4 ML METHOTREXATE 50 MG/ML AUTO-INJECTOR |
| 1441419 | MA00 | Immunosuppressants | METHOTREXATE 50 MG/ML AUTO-INJECTOR |
| 1441422 | MA00 | Immunosuppressants | 0.4 ML METHOTREXATE 62.5 MG/ML AUTO-INJECTOR |
| 1441425 | MA00 | Immunosuppressants | METHOTREXATE 62.5 MG/ML AUTO-INJECTOR |
| 1544378 | MA00 | Immunosuppressants | 0.2 ML METHOTREXATE 50 MG/ML AUTO-INJECTOR |
| 1544385 | MA00 | Immunosuppressants | 0.25 ML METHOTREXATE 50 MG/ML AUTO-INJECTOR |

| | | | |
|---|---|---|---|
| 1544387 | MA00 | Immunosuppressants | 0.3 ML METHOTREXATE 50 MG/ML AUTO-INJECTOR |
| 1544389 | MA00 | Immunosuppressants | 0.35 ML METHOTREXATE 50 MG/ML AUTO-INJECTOR |
| 1544395 | MA00 | Immunosuppressants | 0.45 ML METHOTREXATE 50 MG/ML AUTO-INJECTOR |
| 1544397 | MA00 | Immunosuppressants | 0.5 ML METHOTREXATE 50 MG/ML AUTO-INJECTOR |
| 1544399 | MA00 | Immunosuppressants | 0.55 ML METHOTREXATE 50 MG/ML AUTO-INJECTOR |
| 1544401 | MA00 | Immunosuppressants | 0.6 ML METHOTREXATE 50 MG/ML AUTO-INJECTOR |
| 1544403 | MA00 | Immunosuppressants | 0.15 ML METHOTREXATE 50 MG/ML AUTO-INJECTOR |
| 1594757 | MA00 | Immunosuppressants | 0.4 ML METHOTREXATE 18.8 MG/ML AUTO-INJECTOR |
| 1594760 | MA00 | Immunosuppressants | METHOTREXATE 18.8 MG/ML AUTO-INJECTOR |
| 1655956 | MA00 | Immunosuppressants | 40 ML METHOTREXATE 25 MG/ML INJECTION |
| 1655957 | MA00 | Immunosuppressants | METHOTREXATE 25 MG/ML INJECTION |
| 1655959 | MA00 | Immunosuppressants | 10 ML METHOTREXATE 25 MG/ML INJECTION |
| 1655960 | MA00 | Immunosuppressants | 2 ML METHOTREXATE 25 MG/ML INJECTION |
| 1655967 | MA00 | Immunosuppressants | 4 ML METHOTREXATE 25 MG/ML INJECTION |
| 1655968 | MA00 | Immunosuppressants | 8 ML METHOTREXATE 25 MG/ML INJECTION |
| 1747179 | MA00 | Immunosuppressants | 0.4 ML METHOTREXATE 31.3 MG/ML AUTO-INJECTOR |
| 1747182 | MA00 | Immunosuppressants | METHOTREXATE 31.3 MG/ML AUTO-INJECTOR |
| 1747185 | MA00 | Immunosuppressants | 0.4 ML METHOTREXATE 43.8 MG/ML AUTO-INJECTOR |
| 1747188 | MA00 | Immunosuppressants | METHOTREXATE 43.8 MG/ML AUTO-INJECTOR |
| 1747192 | MA00 | Immunosuppressants | 0.4 ML METHOTREXATE 56.3 MG/ML AUTO-INJECTOR |
| 1747195 | MA00 | Immunosuppressants | METHOTREXATE 56.3 MG/ML AUTO-INJECTOR |
| 1921592 | MA00 | Immunosuppressants | METHOTREXATE 2.5 MG/ML ORAL SOLUTION |
| 1946772 | MA00 | Immunosuppressants | METHOTREXATE 25 MG/ML INJECTABLE SOLUTION |
| 245256 | MA00 | Immunosuppressants | METHOTREXATE 10 MG/ML INJECTABLE SOLUTION |
| 250936 | MA00 | Immunosuppressants | METHOTREXATE 500 MG INJECTABLE SOLUTION |
| 283510 | MA00 | Immunosuppressants | METHOTREXATE 15 MG ORAL TABLET |
| 283511 | MA00 | Immunosuppressants | METHOTREXATE 5 MG ORAL TABLET |
| 283671 | MA00 | Immunosuppressants | METHOTREXATE 7.5 MG ORAL TABLET |
| 311624 | MA00 | Immunosuppressants | METHOTREXATE 15 MG/ML INJECTABLE SOLUTION |
| 311625 | MA00 | Immunosuppressants | METHOTREXATE 1000 MG INJECTION |
| 311626 | MA00 | Immunosuppressants | METHOTREXATE 12.5 MG/ML INJECTABLE SOLUTION |
| 311627 | MA00 | Immunosuppressants | METHOTREXATE 25 MG/ML INJECTABLE SOLUTION |
| 315148 | MA00 | Immunosuppressants | METHOTREXATE 2.5 MG/ML INJECTABLE SOLUTION |
| 415045 | MA00 | Immunosuppressants | METHOTREXATE 5 MG/ML INJECTABLE SOLUTION |
| 415046 | MA00 | Immunosuppressants | METHOTREXATE 7.5 MG/ML INJECTABLE SOLUTION |
| 415063 | MA00 | Immunosuppressants | METHOTREXATE 7.46 MG/ML INJECTABLE SOLUTION |
| 1441410 | MA00 | Immunosuppressants | METHOTREXATE 37.5 MG/ML |
| 1441421 | MA00 | Immunosuppressants | METHOTREXATE 62.5 MG/ML |
| 1594756 | MA00 | Immunosuppressants | METHOTREXATE 18.75 MG/ML |
| 1656681 | MA00 | Immunosuppressants | METHOTREXATE 1000 MG |
| 1659719 | MA00 | Immunosuppressants | METHOTREXATE 18.8 MG/ML |
| 1747178 | MA00 | Immunosuppressants | METHOTREXATE 31.3 MG/ML |
| 1747184 | MA00 | Immunosuppressants | METHOTREXATE 43.8 MG/ML |
| 1747191 | MA00 | Immunosuppressants | METHOTREXATE 56.3 MG/ML |
| 328404 | MA00 | Immunosuppressants | METHOTREXATE 50 MG/ML |
| 328406 | MA00 | Immunosuppressants | METHOTREXATE 25 MG/ML |
| 328407 | MA00 | Immunosuppressants | METHOTREXATE 2.5 MG |
| 330028 | MA00 | Immunosuppressants | METHOTREXATE 2.5 MG/ML |
| 331568 | MA00 | Immunosuppressants | METHOTREXATE 10 MG |
| 331569 | MA00 | Immunosuppressants | METHOTREXATE 15 MG |
| 332454 | MA00 | Immunosuppressants | METHOTREXATE 5 MG |
| 332455 | MA00 | Immunosuppressants | METHOTREXATE 7.5 MG |
| 334051 | MA00 | Immunosuppressants | METHOTREXATE 10 MG/ML |
| 334421 | MA00 | Immunosuppressants | METHOTREXATE 500 MG |
| 346335 | MA00 | Immunosuppressants | METHOTREXATE 12.5 MG/ML |
| 360547 | MA00 | Immunosuppressants | METHOTREXATE 100 MG/ML |
| 380984 | MA00 | Immunosuppressants | METHOTREXATE 15 MG/ML |
| 451557 | MA00 | Immunosuppressants | METHOTREXATE 7.46 MG/ML |
| 451938 | MA00 | Immunosuppressants | METHOTREXATE 5 MG/ML |
| 451941 | MA00 | Immunosuppressants | METHOTREXATE 7.5 MG/ML |
| 1441401 | MA00 | Immunosuppressants | METHOTREXATE PREFILLED SYRINGE |
| 1655203 | MA00 | Immunosuppressants | METHOTREXATE AUTO-INJECTOR |
| 1655955 | MA00 | Immunosuppressants | METHOTREXATE INJECTION |
| 1921591 | MA00 | Immunosuppressants | METHOTREXATE ORAL SOLUTION |
| 372835 | MA00 | Immunosuppressants | METHOTREXATE INJECTABLE SOLUTION |
| 372836 | MA00 | Immunosuppressants | METHOTREXATE ORAL TABLET |
| 1162235 | MA00 | Immunosuppressants | METHOTREXATE INJECTABLE PRODUCT |
| 1162236 | MA00 | Immunosuppressants | METHOTREXATE ORAL PRODUCT |
| 1162237 | MA00 | Immunosuppressants | METHOTREXATE PILL |
| 1921590 | MA00 | Immunosuppressants | METHOTREXATE ORAL LIQUID PRODUCT |
| 227302 | MA00 | Immunosuppressants | ORTHOCLONE OKT3 |

| | | | |
|---|---|---|---|
| 42405 | MA00 | Immunosuppressants | MUROMONAB-CD3 |
| 206950 | MA00 | Immunosuppressants | MUROMONAB-CD3 1 MG/ML INJECTABLE SOLUTION [ORTHOCLONE OKT3] |
| 567751 | MA00 | Immunosuppressants | MUROMONAB-CD3 1 MG/ML [ORTHOCLONE OKT3] |
| 402708 | MA00 | Immunosuppressants | MUROMONAB-CD3 INJECTABLE SOLUTION [ORTHOCLONE OKT3] |
| 1184532 | MA00 | Immunosuppressants | ORTHOCLONE OKT3 INJECTABLE PRODUCT |
| 239984 | MA00 | Immunosuppressants | MUROMONAB-CD3 1 MG/ML INJECTABLE SOLUTION |
| 317432 | MA00 | Immunosuppressants | MUROMONAB-CD3 1 MG/ML |
| 372973 | MA00 | Immunosuppressants | MUROMONAB-CD3 INJECTABLE SOLUTION |
| 1156388 | MA00 | Immunosuppressants | MUROMONAB-CD3 INJECTABLE PRODUCT |
| 152633 | MA00 | Immunosuppressants | CELLCEPT |
| 265323 | MA00 | Immunosuppressants | MYCOPHENOLATE |
| 68149 | MA00 | Immunosuppressants | MYCOPHENOLATE MOFETIL |
| 237112 | MA00 | Immunosuppressants | MYCOPHENOLATE MOFETIL HYDROCHLORIDE |
| 408142 | MA00 | Immunosuppressants | MYCOPHENOLATE SODIUM |
| 152591 | MA00 | Immunosuppressants | MYCOPHENOLATE 250 MG ORAL CAPSULE [CELLCEPT] |
| 153754 | MA00 | Immunosuppressants | MYCOPHENOLATE 500 MG ORAL TABLET [CELLCEPT] |
| 213394 | MA00 | Immunosuppressants | MYCOPHENOLATE MOFETIL 500 MG INJECTION [CELLCEPT] |
| 261236 | MA00 | Immunosuppressants | MYCOPHENOLATE 200 MG/ML ORAL SUSPENSION [CELLCEPT] |
| 579573 | MA00 | Immunosuppressants | MYCOPHENOLATE 180 MG ENTERIC COATED TABLET [MYFORTIC] |
| 579576 | MA00 | Immunosuppressants | MYCOPHENOLATE 360 MG ENTERIC COATED TABLET [MYFORTIC] |
| 616435 | MA00 | Immunosuppressants | MYCOPHENOLATE MOFETIL 500 MG ORAL TABLET [CELLCEPT] |
| 616444 | MA00 | Immunosuppressants | MYCOPHENOLATE MOFETIL 200 MG/ML ORAL SUSPENSION [CELLCEPT] |
| 616447 | MA00 | Immunosuppressants | MYCOPHENOLATE MOFETIL 250 MG ORAL CAPSULE [CELLCEPT] |
| 616449 | MA00 | Immunosuppressants | MYCOPHENOLATE 360 MG ENTERIC COATED TABLET [MYFORTIC] |
| 616450 | MA00 | Immunosuppressants | MYCOPHENOLATE 180 MG ENTERIC COATED TABLET [MYFORTIC] |
| 541580 | MA00 | Immunosuppressants | MYCOPHENOLATE 180 MG [MYFORTIC] |
| 541585 | MA00 | Immunosuppressants | MYCOPHENOLATE 360 MG [MYFORTIC] |
| 564992 | MA00 | Immunosuppressants | MYCOPHENOLATE 250 MG [CELLCEPT] |
| 565378 | MA00 | Immunosuppressants | MYCOPHENOLATE 500 MG [CELLCEPT] |
| 573288 | MA00 | Immunosuppressants | MYCOPHENOLATE MOFETIL 6 MG/ML [CELLCEPT] |
| 574465 | MA00 | Immunosuppressants | MYCOPHENOLATE 200 MG/ML [CELLCEPT] |
| 579571 | MA00 | Immunosuppressants | MYCOPHENOLATE 180 MG [MYFORTIC] |
| 579574 | MA00 | Immunosuppressants | MYCOPHENOLATE 360 MG [MYFORTIC] |
| 616433 | MA00 | Immunosuppressants | MYCOPHENOLATE MOFETIL 500 MG [CELLCEPT] |
| 616442 | MA00 | Immunosuppressants | MYCOPHENOLATE MOFETIL 200 MG/ML [CELLCEPT] |
| 616445 | MA00 | Immunosuppressants | MYCOPHENOLATE MOFETIL 250 MG [CELLCEPT] |
| 1726179 | MA00 | Immunosuppressants | MYCOPHENOLATE MOFETIL INJECTION [CELLCEPT] |
| 363166 | MA00 | Immunosuppressants | MYCOPHENOLATE MOFETIL INJECTABLE SOLUTION [CELLCEPT] |
| 365853 | MA00 | Immunosuppressants | MYCOPHENOLATE ORAL SUSPENSION [CELLCEPT] |
| 366524 | MA00 | Immunosuppressants | MYCOPHENOLATE ORAL CAPSULE [CELLCEPT] |
| 369377 | MA00 | Immunosuppressants | MYCOPHENOLATE ORAL TABLET [CELLCEPT] |
| 579572 | MA00 | Immunosuppressants | MYCOPHENOLATE ENTERIC COATED TABLET [MYFORTIC] |
| 579575 | MA00 | Immunosuppressants | MYCOPHENOLATE ENTERIC COATED TABLET [MYFORTIC] |
| 616434 | MA00 | Immunosuppressants | MYCOPHENOLATE MOFETIL ORAL TABLET [CELLCEPT] |
| 616443 | MA00 | Immunosuppressants | MYCOPHENOLATE MOFETIL ORAL SUSPENSION [CELLCEPT] |
| 616446 | MA00 | Immunosuppressants | MYCOPHENOLATE MOFETIL ORAL CAPSULE [CELLCEPT] |
| 616448 | MA00 | Immunosuppressants | MYCOPHENOLATE ENTERIC COATED TABLET [MYFORTIC] |
| 1166213 | MA00 | Immunosuppressants | CELLCEPT INJECTABLE PRODUCT |
| 1166214 | MA00 | Immunosuppressants | CELLCEPT ORAL LIQUID PRODUCT |
| 1166215 | MA00 | Immunosuppressants | CELLCEPT ORAL PRODUCT |
| 1166216 | MA00 | Immunosuppressants | CELLCEPT PILL |
| 199058 | MA00 | Immunosuppressants | MYCOPHENOLATE MOFETIL 250 MG ORAL CAPSULE |
| 200060 | MA00 | Immunosuppressants | MYCOPHENOLATE MOFETIL 500 MG ORAL TABLET |
| 311880 | MA00 | Immunosuppressants | MYCOPHENOLATE MOFETIL 200 MG/ML ORAL SUSPENSION |
| 414999 | MA00 | Immunosuppressants | MYCOPHENOLATE MOFETIL 25 MG/ML INJECTABLE SOLUTION |
| 433833 | MA00 | Immunosuppressants | MYCOPHENOLATE 180 MG ENTERIC COATED TABLET |
| 433834 | MA00 | Immunosuppressants | MYCOPHENOLATE 360 MG ENTERIC COATED TABLET |
| 562143 | MA00 | Immunosuppressants | MYCOPHENOLATE 250 MG ORAL CAPSULE |
| 562175 | MA00 | Immunosuppressants | MYCOPHENOLATE 500 MG ORAL TABLET |
| 562701 | MA00 | Immunosuppressants | MYCOPHENOLATE 200 MG/ML ORAL SUSPENSION |
| 579569 | MA00 | Immunosuppressants | MYCOPHENOLATE 180 MG ENTERIC COATED TABLET |
| 596873 | MA00 | Immunosuppressants | MYCOPHENOLATE 180 MG EXTENDED RELEASE ENTERIC COATED TABLET |
| 670049 | MA00 | Immunosuppressants | MYCOPHENOLATE 180 MG ENTERIC COATED TABLET |
| 670051 | MA00 | Immunosuppressants | MYCOPHENOLATE 360 MG ENTERIC COATED TABLET |
| 316316 | MA00 | Immunosuppressants | MYCOPHENOLATE MOFETIL 250 MG |
| 316317 | MA00 | Immunosuppressants | MYCOPHENOLATE MOFETIL 500 MG |
| 331587 | MA00 | Immunosuppressants | MYCOPHENOLATE MOFETIL 200 MG/ML |
| 332547 | MA00 | Immunosuppressants | MYCOPHENOLATE MOFETIL 6 MG/ML |
| 353404 | MA00 | Immunosuppressants | MYCOPHENOLATE 250 MG |
| 353412 | MA00 | Immunosuppressants | MYCOPHENOLATE 500 MG |
| 353414 | MA00 | Immunosuppressants | MYCOPHENOLATE 200 MG/ML |

| | | | |
|---|---|---|---|
| 446560 | MA00 | Immunosuppressants | MYCOPHENOLATE 180 MG |
| 446562 | MA00 | Immunosuppressants | MYCOPHENOLATE 360 MG |
| 452791 | MA00 | Immunosuppressants | MYCOPHENOLATE MOFETIL 25 MG/ML |
| 485018 | MA00 | Immunosuppressants | MYCOPHENOLATE 180 MG |
| 485022 | MA00 | Immunosuppressants | MYCOPHENOLATE 360 MG |
| 579567 | MA00 | Immunosuppressants | MYCOPHENOLATE 180 MG |
| 670048 | MA00 | Immunosuppressants | MYCOPHENOLATE 180 MG |
| 670050 | MA00 | Immunosuppressants | MYCOPHENOLATE 360 MG |
| 1726176 | MA00 | Immunosuppressants | MYCOPHENOLATE MOFETIL INJECTION |
| 374667 | MA00 | Immunosuppressants | MYCOPHENOLATE MOFETIL ORAL TABLET |
| 374670 | MA00 | Immunosuppressants | MYCOPHENOLATE MOFETIL ORAL SUSPENSION |
| 376670 | MA00 | Immunosuppressants | MYCOPHENOLATE MOFETIL INJECTABLE SOLUTION |
| 379060 | MA00 | Immunosuppressants | MYCOPHENOLATE MOFETIL ORAL CAPSULE |
| 446561 | MA00 | Immunosuppressants | MYCOPHENOLATE ENTERIC COATED TABLET |
| 562142 | MA00 | Immunosuppressants | MYCOPHENOLATE ORAL CAPSULE |
| 562174 | MA00 | Immunosuppressants | MYCOPHENOLATE ORAL TABLET |
| 562700 | MA00 | Immunosuppressants | MYCOPHENOLATE ORAL SUSPENSION |
| 579568 | MA00 | Immunosuppressants | MYCOPHENOLATE ENTERIC COATED TABLET |
| 596872 | MA00 | Immunosuppressants | MYCOPHENOLATE EXTENDED RELEASE ENTERIC COATED TABLET |
| 1157869 | MA00 | Immunosuppressants | MYCOPHENOLATE MOFETIL INJECTABLE PRODUCT |
| 1157870 | MA00 | Immunosuppressants | MYCOPHENOLATE MOFETIL ORAL LIQUID PRODUCT |
| 1157871 | MA00 | Immunosuppressants | MYCOPHENOLATE MOFETIL ORAL PRODUCT |
| 1157872 | MA00 | Immunosuppressants | MYCOPHENOLATE MOFETIL PILL |
| 258355 | MA00 | Immunosuppressants | RAPAMUNE |
| 35302 | MA00 | Immunosuppressants | SIROLIMUS |
| 1724309 | MA00 | Immunosuppressants | TEMSIROLIMUS 25 MG/ML INJECTION [TORISEL] |
| 351901 | MA00 | Immunosuppressants | SIROLIMUS 1 MG/ML ORAL SOLUTION [RAPAMUNE] |
| 351989 | MA00 | Immunosuppressants | SIROLIMUS 1 MG ORAL TABLET [RAPAMUNE] |
| 404432 | MA00 | Immunosuppressants | SIROLIMUS 2 MG ORAL TABLET [RAPAMUNE] |
| 905160 | MA00 | Immunosuppressants | SIROLIMUS 0.5 MG ORAL TABLET [RAPAMUNE] |
| 575659 | MA00 | Immunosuppressants | SIROLIMUS 1 MG/ML [RAPAMUNE] |
| 575733 | MA00 | Immunosuppressants | SIROLIMUS 1 MG [RAPAMUNE] |
| 576390 | MA00 | Immunosuppressants | SIROLIMUS 2 MG [RAPAMUNE] |
| 905159 | MA00 | Immunosuppressants | SIROLIMUS 0.5 MG [RAPAMUNE] |
| 1724306 | MA00 | Immunosuppressants | TEMSIROLIMUS INJECTION [TORISEL] |
| 364904 | MA00 | Immunosuppressants | SIROLIMUS ORAL SOLUTION [RAPAMUNE] |
| 368672 | MA00 | Immunosuppressants | SIROLIMUS ORAL TABLET [RAPAMUNE] |
| 1177701 | MA00 | Immunosuppressants | RAPAMUNE ORAL LIQUID PRODUCT |
| 1177702 | MA00 | Immunosuppressants | RAPAMUNE ORAL PRODUCT |
| 1177703 | MA00 | Immunosuppressants | RAPAMUNE PILL |
| 1178590 | MA00 | Immunosuppressants | TORISEL INJECTABLE PRODUCT |
| 314230 | MA00 | Immunosuppressants | SIROLIMUS 1 MG/ML ORAL SOLUTION |
| 349208 | MA00 | Immunosuppressants | SIROLIMUS 1 MG ORAL TABLET |
| 360110 | MA00 | Immunosuppressants | SIROLIMUS 2 MG ORAL TABLET |
| 905158 | MA00 | Immunosuppressants | SIROLIMUS 0.5 MG ORAL TABLET |
| 333296 | MA00 | Immunosuppressants | SIROLIMUS 1 MG/ML |
| 350512 | MA00 | Immunosuppressants | SIROLIMUS 1 MG |
| 360318 | MA00 | Immunosuppressants | SIROLIMUS 2 MG |
| 905157 | MA00 | Immunosuppressants | SIROLIMUS 0.5 MG |
| 1724304 | MA00 | Immunosuppressants | TEMSIROLIMUS INJECTION |
| 375278 | MA00 | Immunosuppressants | SIROLIMUS ORAL SOLUTION |
| 378277 | MA00 | Immunosuppressants | SIROLIMUS ORAL TABLET |
| 1158757 | MA00 | Immunosuppressants | SIROLIMUS ORAL LIQUID PRODUCT |
| 1158758 | MA00 | Immunosuppressants | SIROLIMUS ORAL PRODUCT |
| 1158759 | MA00 | Immunosuppressants | SIROLIMUS PILL |
| 1161488 | MA00 | Immunosuppressants | TEMSIROLIMUS INJECTABLE PRODUCT |
| 1235236 | MA00 | Immunosuppressants | HECORIA |
| 1431972 | MA00 | Immunosuppressants | ASTAGRAF |
| 1664435 | MA00 | Immunosuppressants | ENVARSUS |
| 196463 | MA00 | Immunosuppressants | PROGRAF |
| 42316 | MA00 | Immunosuppressants | TACROLIMUS |
| 235991 | MA00 | Immunosuppressants | ANHYDROUS TACROLIMUS |
| 484288 | MA00 | Immunosuppressants | TACROLIMUS MONOHYDRATE |
| 108513 | MA00 | Immunosuppressants | TACROLIMUS 1 MG ORAL CAPSULE [PROGRAF] |
| 108514 | MA00 | Immunosuppressants | TACROLIMUS 5 MG ORAL CAPSULE [PROGRAF] |
| 108515 | MA00 | Immunosuppressants | 1 ML TACROLIMUS 5 MG/ML INJECTION [PROGRAF] |
| 1235241 | MA00 | Immunosuppressants | TACROLIMUS 0.5 MG ORAL CAPSULE [HECORIA] |
| 1235243 | MA00 | Immunosuppressants | TACROLIMUS 1 MG ORAL CAPSULE [HECORIA] |
| 1235245 | MA00 | Immunosuppressants | TACROLIMUS 5 MG ORAL CAPSULE [HECORIA] |
| 1431977 | MA00 | Immunosuppressants | 24 HR TACROLIMUS 0.5 MG EXTENDED RELEASE ORAL CAPSULE [ASTAGRAF] |
| 1431979 | MA00 | Immunosuppressants | TACROLIMUS 0.5 MG EXTENDED RELEASE ORAL CAPSULE [ASTAGRAF] |

| | | | |
|---|---|---|---|
| 1431982 | MA00 | Immunosuppressants | 24 HR TACROLIMUS 1 MG EXTENDED RELEASE ORAL CAPSULE [ASTAGRAF] |
| 1431984 | MA00 | Immunosuppressants | TACROLIMUS 1 MG EXTENDED RELEASE ORAL CAPSULE [ASTAGRAF] |
| 1431987 | MA00 | Immunosuppressants | 24 HR TACROLIMUS 5 MG EXTENDED RELEASE ORAL CAPSULE [ASTAGRAF] |
| 1431989 | MA00 | Immunosuppressants | TACROLIMUS 5 MG EXTENDED RELEASE ORAL CAPSULE [ASTAGRAF] |
| 1664440 | MA00 | Immunosuppressants | 24 HR TACROLIMUS 4 MG EXTENDED RELEASE ORAL TABLET [ENVARSUS] |
| 1664442 | MA00 | Immunosuppressants | TACROLIMUS 4 MG EXTENDED RELEASE ORAL TABLET [ENVARSUS] |
| 1664458 | MA00 | Immunosuppressants | 24 HR TACROLIMUS 0.75 MG EXTENDED RELEASE ORAL TABLET [ENVARSUS] |
| 1664460 | MA00 | Immunosuppressants | TACROLIMUS 0.75 MG EXTENDED RELEASE ORAL TABLET [ENVARSUS] |
| 1664463 | MA00 | Immunosuppressants | 24 HR TACROLIMUS 1 MG EXTENDED RELEASE ORAL TABLET [ENVARSUS] |
| 1664465 | MA00 | Immunosuppressants | TACROLIMUS 1 MG EXTENDED RELEASE ORAL TABLET [ENVARSUS] |
| 1738339 | MA00 | Immunosuppressants | TACROLIMUS 5 MG/ML INJECTION [PROGRAF] |
| 2052609 | MA00 | Immunosuppressants | TACROLIMUS 0.2 MG GRANULES FOR ORAL SUSPENSION [PROGRAF] |
| 2052704 | MA00 | Immunosuppressants | TACROLIMUS 1 MG GRANULES FOR ORAL SUSPENSION [PROGRAF] |
| 261134 | MA00 | Immunosuppressants | TACROLIMUS 0.5 MG ORAL CAPSULE [PROGRAF] |
| 1235237 | MA00 | Immunosuppressants | TACROLIMUS 0.5 MG [HECORIA] |
| 1235242 | MA00 | Immunosuppressants | TACROLIMUS 1 MG [HECORIA] |
| 1235244 | MA00 | Immunosuppressants | TACROLIMUS 5 MG [HECORIA] |
| 1431973 | MA00 | Immunosuppressants | TACROLIMUS 0.5 MG [ASTAGRAF] |
| 1431981 | MA00 | Immunosuppressants | TACROLIMUS 1 MG [ASTAGRAF] |
| 1431986 | MA00 | Immunosuppressants | TACROLIMUS 5 MG [ASTAGRAF] |
| 1664436 | MA00 | Immunosuppressants | TACROLIMUS 4 MG [ENVARSUS] |
| 1664457 | MA00 | Immunosuppressants | TACROLIMUS 0.75 MG [ENVARSUS] |
| 1664462 | MA00 | Immunosuppressants | TACROLIMUS 1 MG [ENVARSUS] |
| 2052606 | MA00 | Immunosuppressants | TACROLIMUS 0.2 MG [PROGRAF] |
| 564557 | MA00 | Immunosuppressants | TACROLIMUS 1 MG [PROGRAF] |
| 564558 | MA00 | Immunosuppressants | TACROLIMUS 5 MG [PROGRAF] |
| 564559 | MA00 | Immunosuppressants | TACROLIMUS 5 MG/ML [PROGRAF] |
| 574378 | MA00 | Immunosuppressants | TACROLIMUS 0.5 MG [PROGRAF] |
| 1235238 | MA00 | Immunosuppressants | TACROLIMUS ORAL CAPSULE [HECORIA] |
| 1431974 | MA00 | Immunosuppressants | TACROLIMUS EXTENDED RELEASE ORAL CAPSULE [ASTAGRAF] |
| 1664437 | MA00 | Immunosuppressants | TACROLIMUS EXTENDED RELEASE ORAL TABLET [ENVARSUS] |
| 1738336 | MA00 | Immunosuppressants | TACROLIMUS INJECTION [PROGRAF] |
| 2052607 | MA00 | Immunosuppressants | TACROLIMUS GRANULES FOR ORAL SUSPENSION [PROGRAF] |
| 362879 | MA00 | Immunosuppressants | TACROLIMUS INJECTABLE SOLUTION [PROGRAF] |
| 366273 | MA00 | Immunosuppressants | TACROLIMUS ORAL CAPSULE [PROGRAF] |
| 1180662 | MA00 | Immunosuppressants | PROGRAF INJECTABLE PRODUCT |
| 1180663 | MA00 | Immunosuppressants | PROGRAF ORAL PRODUCT |
| 1180664 | MA00 | Immunosuppressants | PROGRAF PILL |
| 1235239 | MA00 | Immunosuppressants | HECORIA ORAL PRODUCT |
| 1235240 | MA00 | Immunosuppressants | HECORIA PILL |
| 1431975 | MA00 | Immunosuppressants | ASTAGRAF ORAL PRODUCT |
| 1431976 | MA00 | Immunosuppressants | ASTAGRAF PILL |
| 1664438 | MA00 | Immunosuppressants | ENVARSUS ORAL PRODUCT |
| 1664439 | MA00 | Immunosuppressants | ENVARSUS PILL |
| 2052608 | MA00 | Immunosuppressants | PROGRAF GRANULE PRODUCT |
| 1431971 | MA00 | Immunosuppressants | 24 HR TACROLIMUS 0.5 MG EXTENDED RELEASE ORAL CAPSULE |
| 1431978 | MA00 | Immunosuppressants | TACROLIMUS 0.5 MG EXTENDED RELEASE ORAL CAPSULE |
| 1431980 | MA00 | Immunosuppressants | 24 HR TACROLIMUS 1 MG EXTENDED RELEASE ORAL CAPSULE |
| 1431983 | MA00 | Immunosuppressants | TACROLIMUS 1 MG EXTENDED RELEASE ORAL CAPSULE |
| 1431985 | MA00 | Immunosuppressants | 24 HR TACROLIMUS 5 MG EXTENDED RELEASE ORAL CAPSULE |
| 1431988 | MA00 | Immunosuppressants | TACROLIMUS 5 MG EXTENDED RELEASE ORAL CAPSULE |
| 1664434 | MA00 | Immunosuppressants | 24 HR TACROLIMUS 4 MG EXTENDED RELEASE ORAL TABLET |
| 1664441 | MA00 | Immunosuppressants | TACROLIMUS 4 MG EXTENDED RELEASE ORAL TABLET |
| 1664456 | MA00 | Immunosuppressants | 24 HR TACROLIMUS 0.75 MG EXTENDED RELEASE ORAL TABLET |
| 1664459 | MA00 | Immunosuppressants | TACROLIMUS 0.75 MG EXTENDED RELEASE ORAL TABLET |
| 1664461 | MA00 | Immunosuppressants | 24 HR TACROLIMUS 1 MG EXTENDED RELEASE ORAL TABLET |
| 1664464 | MA00 | Immunosuppressants | TACROLIMUS 1 MG EXTENDED RELEASE ORAL TABLET |
| 1738338 | MA00 | Immunosuppressants | TACROLIMUS 5 MG/ML INJECTION |
| 198377 | MA00 | Immunosuppressants | TACROLIMUS 1 MG ORAL CAPSULE |
| 198378 | MA00 | Immunosuppressants | TACROLIMUS 5 MG ORAL CAPSULE |
| 198379 | MA00 | Immunosuppressants | 1 ML TACROLIMUS 5 MG/ML INJECTION |
| 2052605 | MA00 | Immunosuppressants | TACROLIMUS 0.2 MG GRANULES FOR ORAL SUSPENSION |
| 2052703 | MA00 | Immunosuppressants | TACROLIMUS 1 MG GRANULES FOR ORAL SUSPENSION |
| 427807 | MA00 | Immunosuppressants | TACROLIMUS 5 MG ORAL TABLET |
| 427808 | MA00 | Immunosuppressants | TACROLIMUS 1 MG ORAL TABLET |
| 1599039 | MA00 | Immunosuppressants | TACROLIMUS 0.5 MG/ML |
| 1664432 | MA00 | Immunosuppressants | TACROLIMUS 4 MG |
| 1664455 | MA00 | Immunosuppressants | TACROLIMUS 0.75 MG |
| 2052503 | MA00 | Immunosuppressants | TACROLIMUS 0.2 MG |
| 316758 | MA00 | Immunosuppressants | TACROLIMUS 1 MG |
| 316759 | MA00 | Immunosuppressants | TACROLIMUS 5 MG |

| | | | |
|---|---|---|---|
| 317510 | MA00 | Immunosuppressants | TACROLIMUS 5 MG/ML |
| 330404 | MA00 | Immunosuppressants | TACROLIMUS 0.5 MG |
| 331549 | MA00 | Immunosuppressants | TACROLIMUS 0.001 MG/MG |
| 331550 | MA00 | Immunosuppressants | TACROLIMUS 0.0003 MG/MG |
| 1431970 | MA00 | Immunosuppressants | TACROLIMUS EXTENDED RELEASE ORAL CAPSULE |
| 1664433 | MA00 | Immunosuppressants | TACROLIMUS EXTENDED RELEASE ORAL TABLET |
| 1738334 | MA00 | Immunosuppressants | TACROLIMUS INJECTION |
| 2052604 | MA00 | Immunosuppressants | TACROLIMUS GRANULES FOR ORAL SUSPENSION |
| 374015 | MA00 | Immunosuppressants | TACROLIMUS ORAL CAPSULE |
| 377119 | MA00 | Immunosuppressants | TACROLIMUS INJECTABLE SOLUTION |
| 451875 | MA00 | Immunosuppressants | TACROLIMUS ORAL TABLET |
| 1163655 | MA00 | Immunosuppressants | TACROLIMUS INJECTABLE PRODUCT |
| 1164202 | MA00 | Immunosuppressants | TACROLIMUS ORAL PRODUCT |
| 1164203 | MA00 | Immunosuppressants | TACROLIMUS PILL |
| 2052603 | MA00 | Immunosuppressants | TACROLIMUS GRANULE PRODUCT |
| 723805 | MA00 | Immunosuppressants | TORISEL |
| 657797 | MA00 | Immunosuppressants | TEMSIROLIMUS |
| 725108 | MA00 | Immunosuppressants | 1.2 ML TEMSIROLIMUS 25 MG/ML INJECTION [TORISEL] |
| 725106 | MA00 | Immunosuppressants | TEMSIROLIMUS 25 MG/ML [TORISEL] |
| 725107 | MA00 | Immunosuppressants | TEMSIROLIMUS INJECTABLE SOLUTION [TORISEL] |
| 1724308 | MA00 | Immunosuppressants | TEMSIROLIMUS 25 MG/ML INJECTION |
| 722289 | MA00 | Immunosuppressants | 1.2 ML TEMSIROLIMUS 25 MG/ML INJECTION |
| 993785 | MA00 | Immunosuppressants | TEMSIROLIMUS 10 MG/ML INJECTABLE SOLUTION |
| 722287 | MA00 | Immunosuppressants | TEMSIROLIMUS 25 MG/ML |
| 993784 | MA00 | Immunosuppressants | TEMSIROLIMUS 10 MG/ML |
| 722288 | MA00 | Immunosuppressants | TEMSIROLIMUS INJECTABLE SOLUTION |
| 72435 | MA00 | Immunosuppressants | anakinra |
| 352734 | MA00 | Immunosuppressants | Kineret |
| 727711 | MA00 | Immunosuppressants | 0.67 ML anakinra 149 MG/ML Prefilled Syringe |
| 727714 | MA00 | Immunosuppressants | 0.67 ML anakinra 149 MG/ML Prefilled Syringe [Kineret] |
| 727708 | MA00 | Immunosuppressants | anakinra Prefilled Syringe |
| 727710 | MA00 | Immunosuppressants | anakinra 149 MG/ML |
| 727712 | MA00 | Immunosuppressants | anakinra 149 MG/ML [Kineret] |
| 727713 | MA00 | Immunosuppressants | anakinra Prefilled Syringe [Kineret] |
| 1157701 | MA00 | Immunosuppressants | anakinra Injectable Product |
| 1170741 | MA00 | Immunosuppressants | Kineret Injectable Product |
| 3218811 | MA11 | Immunosuppressants | abatacept |
| 3281411 | MA11 | Immunosuppressants | abatacept |
| 3218831 | MA11 | Immunosuppressants | abatacept |
| 3281811 | MA11 | Immunosuppressants | abatacept |
| 3218851 | MA11 | Immunosuppressants | abatacept |
| 3218713 | MA11 | Immunosuppressants | abatacept/maltose |
| 3218710 | MA11 | Immunosuppressants | abatacept/maltose |
| 74061602 | MA11 | Immunosuppressants | adalimumab |
| 74006702 | MA11 | Immunosuppressants | adalimumab |
| 74433901 | MA11 | Immunosuppressants | adalimumab |
| 74012474 | MA11 | Immunosuppressants | adalimumab |
| 74153903 | MA11 | Immunosuppressants | adalimumab |
| 74055471 | MA11 | Immunosuppressants | adalimumab |
| 74433971 | MA11 | Immunosuppressants | adalimumab |
| 74433974 | MA11 | Immunosuppressants | adalimumab |
| 74055402 | MA11 | Immunosuppressants | adalimumab |
| 74433906 | MA11 | Immunosuppressants | adalimumab |
| 74379906 | MA11 | Immunosuppressants | adalimumab |
| 74024302 | MA11 | Immunosuppressants | adalimumab |
| 74254003 | MA11 | Immunosuppressants | adalimumab |
| 74433902 | MA11 | Immunosuppressants | adalimumab |
| 54868482200 | MA11 | Immunosuppressants | adalimumab |
| 74081702 | MA11 | Immunosuppressants | adalimumab |
| 74012403 | MA11 | Immunosuppressants | adalimumab |
| 74379903 | MA11 | Immunosuppressants | adalimumab |
| 74379902 | MA11 | Immunosuppressants | adalimumab |
| 74634702 | MA11 | Immunosuppressants | adalimumab |
| 54569552400 | MA11 | Immunosuppressants | adalimumab |
| 74937402 | MA11 | Immunosuppressants | adalimumab |
| 74433907 | MA11 | Immunosuppressants | adalimumab |
| 58468035703 | MA11 | Immunosuppressants | alemtuzumab |
| 50419035510 | MA11 | Immunosuppressants | alemtuzumab |
| 58468020001 | MA11 | Immunosuppressants | alemtuzumab |
| 50419035703 | MA11 | Immunosuppressants | alemtuzumab |
| 58468035701 | MA11 | Immunosuppressants | alemtuzumab |

| | | | |
|---|---|---|---|
| 50419035512 | MA11 | Immunosuppressants | alemtuzumab |
| 3037113 | MA11 | Immunosuppressants | belatacept |
| 49401008835 | MA11 | Immunosuppressants | belimumab |
| 49401010101 | MA11 | Immunosuppressants | belimumab |
| 49401010201 | MA11 | Immunosuppressants | belimumab |
| 49401008801 | MA11 | Immunosuppressants | belimumab |
| 49401008842 | MA11 | Immunosuppressants | belimumab |
| 49401008847 | MA11 | Immunosuppressants | belimumab |
| 78073461 | MA11 | Immunosuppressants | canakinumab/PF |
| 78058261 | MA11 | Immunosuppressants | canakinumab/PF |
| 50474071079 | MA11 | Immunosuppressants | certolizumab pegol |
| 50474070062 | MA11 | Immunosuppressants | certolizumab pegol |
| 50474071081 | MA11 | Immunosuppressants | certolizumab pegol |
| 38779115603 | MA11 | Immunosuppressants | chlorambucil |
| 54868112602 | MA11 | Immunosuppressants | chlorambucil |
| 54868112605 | MA11 | Immunosuppressants | chlorambucil |
| 38779115606 | MA11 | Immunosuppressants | chlorambucil |
| 38779115600 | MA11 | Immunosuppressants | chlorambucil |
| 54868112600 | MA11 | Immunosuppressants | chlorambucil |
| 76388063525 | MA11 | Immunosuppressants | chlorambucil |
| 46144015825 | MA11 | Immunosuppressants | chlorambucil |
| 46144015801 | MA11 | Immunosuppressants | chlorambucil |
| 51927217700 | MA11 | Immunosuppressants | chlorambucil |
| 76388063550 | MA11 | Immunosuppressants | chlorambucil |
| 51927446700 | MA11 | Immunosuppressants | chlorambucil |
| 38779115607 | MA11 | Immunosuppressants | chlorambucil |
| 173063535 | MA11 | Immunosuppressants | chlorambucil |
| 54868112601 | MA11 | Immunosuppressants | chlorambucil |
| 54868112603 | MA11 | Immunosuppressants | chlorambucil |
| 54868112604 | MA11 | Immunosuppressants | chlorambucil |
| 25682000101 | MA11 | Immunosuppressants | eculizumab |
| 58406044504 | MA11 | Immunosuppressants | etanercept |
| 58406044501 | MA11 | Immunosuppressants | etanercept |
| 54868478200 | MA11 | Immunosuppressants | etanercept |
| 58406003204 | MA11 | Immunosuppressants | etanercept |
| 58406001001 | MA11 | Immunosuppressants | etanercept |
| 58406004404 | MA11 | Immunosuppressants | etanercept |
| 58406045604 | MA11 | Immunosuppressants | etanercept |
| 58406045504 | MA11 | Immunosuppressants | etanercept |
| 58406001004 | MA11 | Immunosuppressants | etanercept |
| 58406043501 | MA11 | Immunosuppressants | etanercept |
| 58406045501 | MA11 | Immunosuppressants | etanercept |
| 58406002104 | MA11 | Immunosuppressants | etanercept |
| 58406003201 | MA11 | Immunosuppressants | etanercept |
| 54868544400 | MA11 | Immunosuppressants | etanercept |
| 58406042534 | MA11 | Immunosuppressants | etanercept |
| 58406045601 | MA11 | Immunosuppressants | etanercept |
| 58406043504 | MA11 | Immunosuppressants | etanercept |
| 58406002101 | MA11 | Immunosuppressants | etanercept |
| 58406042541 | MA11 | Immunosuppressants | etanercept |
| 58406004401 | MA11 | Immunosuppressants | etanercept |
| 57894035001 | MA11 | Immunosuppressants | golimumab |
| 57894007001 | MA11 | Immunosuppressants | golimumab |
| 57894007101 | MA11 | Immunosuppressants | golimumab |
| 57894007002 | MA11 | Immunosuppressants | golimumab |
| 57894007102 | MA11 | Immunosuppressants | golimumab |
| 57894003001 | MA11 | Immunosuppressants | infliximab |
| 6430501 | MA11 | Immunosuppressants | infliximab-abda |
| 6430502 | MA11 | Immunosuppressants | infliximab-abda |
| 69080901 | MA11 | Immunosuppressants | infliximab-dyyb |
| 49884088811 | MA11 | Immunosuppressants | leflunomide |
| 63629126301 | MA11 | Immunosuppressants | leflunomide |
| 66993016030 | MA11 | Immunosuppressants | leflunomide |
| 54868231900 | MA11 | Immunosuppressants | leflunomide |
| 60429032030 | MA11 | Immunosuppressants | leflunomide |
| 60505250201 | MA11 | Immunosuppressants | leflunomide |
| 781505731 | MA11 | Immunosuppressants | leflunomide |
| 68115081730 | MA11 | Immunosuppressants | leflunomide |
| 555035201 | MA11 | Immunosuppressants | leflunomide |
| 70710115703 | MA11 | Immunosuppressants | leflunomide |
| 54868438500 | MA11 | Immunosuppressants | leflunomide |

| | | | |
|---|---|---|---|
| 93017456 | MA11 | Immunosuppressants | leflunomide |
| 10702027703 | MA11 | Immunosuppressants | leflunomide |
| 54868490200 | MA11 | Immunosuppressants | leflunomide |
| 60505250203 | MA11 | Immunosuppressants | leflunomide |
| 955173530 | MA11 | Immunosuppressants | leflunomide |
| 60505250303 | MA11 | Immunosuppressants | leflunomide |
| 42291042130 | MA11 | Immunosuppressants | leflunomide |
| 23155004403 | MA11 | Immunosuppressants | leflunomide |
| 88216130 | MA11 | Immunosuppressants | leflunomide |
| 49884088905 | MA11 | Immunosuppressants | leflunomide |
| 62332006130 | MA11 | Immunosuppressants | leflunomide |
| 54868617000 | MA11 | Immunosuppressants | leflunomide |
| 88216030 | MA11 | Immunosuppressants | leflunomide |
| 555035101 | MA11 | Immunosuppressants | leflunomide |
| 13811067830 | MA11 | Immunosuppressants | leflunomide |
| 955173730 | MA11 | Immunosuppressants | leflunomide |
| 60505250301 | MA11 | Immunosuppressants | leflunomide |
| 42291042030 | MA11 | Immunosuppressants | leflunomide |
| 88216203 | MA11 | Immunosuppressants | leflunomide |
| 23155004303 | MA11 | Immunosuppressants | leflunomide |
| 93017356 | MA11 | Immunosuppressants | leflunomide |
| 49884088911 | MA11 | Immunosuppressants | leflunomide |
| 49884088805 | MA11 | Immunosuppressants | leflunomide |
| 70710115803 | MA11 | Immunosuppressants | leflunomide |
| 60429031930 | MA11 | Immunosuppressants | leflunomide |
| 16714033101 | MA11 | Immunosuppressants | leflunomide |
| 10702027803 | MA11 | Immunosuppressants | leflunomide |
| 66993016130 | MA11 | Immunosuppressants | leflunomide |
| 13811067730 | MA11 | Immunosuppressants | leflunomide |
| 16714032101 | MA11 | Immunosuppressants | leflunomide |
| 781505631 | MA11 | Immunosuppressants | leflunomide |
| 62332006230 | MA11 | Immunosuppressants | leflunomide |
| 50242015001 | MA11 | Immunosuppressants | ocrelizumab |
| 173082102 | MA11 | Immunosuppressants | ofatumumab |
| 173080802 | MA11 | Immunosuppressants | ofatumumab |
| 173082133 | MA11 | Immunosuppressants | ofatumumab |
| 78066913 | MA11 | Immunosuppressants | ofatumumab |
| 173080805 | MA11 | Immunosuppressants | ofatumumab |
| 78069061 | MA11 | Immunosuppressants | ofatumumab |
| 173082101 | MA11 | Immunosuppressants | ofatumumab |
| 78066961 | MA11 | Immunosuppressants | ofatumumab |
| 61755000101 | MA11 | Immunosuppressants | rilonacept |
| 50242005121 | MA11 | Immunosuppressants | rituximab |
| 50242005306 | MA11 | Immunosuppressants | rituximab |
| 50242005110 | MA11 | Immunosuppressants | rituximab |
| 63459010450 | MA11 | Immunosuppressants | rituximab-abbs |
| 63459010310 | MA11 | Immunosuppressants | rituximab-abbs |
| 69024901 | MA11 | Immunosuppressants | rituximab-pvvr |
| 69023801 | MA11 | Immunosuppressants | rituximab-pvvr |
| 50242010801 | MA11 | Immunosuppressants | rituximab/hyaluronidase, human recombinant |
| 50242010901 | MA11 | Immunosuppressants | rituximab/hyaluronidase, human recombinant |
| 78063998 | MA11 | Immunosuppressants | secukinumab |
| 78063997 | MA11 | Immunosuppressants | secukinumab |
| 78063941 | MA11 | Immunosuppressants | secukinumab |
| 78063968 | MA11 | Immunosuppressants | secukinumab |
| 50242013801 | MA11 | Immunosuppressants | tocilizumab |
| 50242013701 | MA11 | Immunosuppressants | tocilizumab |
| 50242013501 | MA11 | Immunosuppressants | tocilizumab |
| 50242013601 | MA11 | Immunosuppressants | tocilizumab |
| 50242014301 | MA11 | Immunosuppressants | tocilizumab |
| 69050230 | MA11 | Immunosuppressants | tofacitinib citrate |
| 69050130 | MA11 | Immunosuppressants | tofacitinib citrate |
| 69050114 | MA11 | Immunosuppressants | tofacitinib citrate |
| 69100101 | MA11 | Immunosuppressants | tofacitinib citrate |
| 69100201 | MA11 | Immunosuppressants | tofacitinib citrate |
| 71610030660 | MA11 | Immunosuppressants | azathioprine |
| 38779031204 | MA11 | Immunosuppressants | azathioprine |
| 51552077902 | MA11 | Immunosuppressants | azathioprine |
| 49452078303 | MA11 | Immunosuppressants | azathioprine |
| 65483059010 | MA11 | Immunosuppressants | azathioprine |
| 49452078301 | MA11 | Immunosuppressants | azathioprine |

| | | | |
|---|---|---|---|
| 378100501 | MA11 | Immunosuppressants | azathioprine |
| 68382000301 | MA11 | Immunosuppressants | azathioprine |
| 51079062006 | MA11 | Immunosuppressants | azathioprine |
| 54868531003 | MA11 | Immunosuppressants | azathioprine |
| 43353068609 | MA11 | Immunosuppressants | azathioprine |
| 60976059755 | MA11 | Immunosuppressants | azathioprine |
| 49452078302 | MA11 | Immunosuppressants | azathioprine |
| 54868092101 | MA11 | Immunosuppressants | azathioprine |
| 52372072803 | MA11 | Immunosuppressants | azathioprine |
| 62991218901 | MA11 | Immunosuppressants | azathioprine |
| 54868531000 | MA11 | Immunosuppressants | azathioprine |
| 49452078304 | MA11 | Immunosuppressants | azathioprine |
| 54808425 | MA11 | Immunosuppressants | azathioprine |
| 38779031225 | MA11 | Immunosuppressants | azathioprine |
| 49452078202 | MA11 | Immunosuppressants | azathioprine |
| 43353074560 | MA11 | Immunosuppressants | azathioprine |
| 62991218904 | MA11 | Immunosuppressants | azathioprine |
| 52959007900 | MA11 | Immunosuppressants | azathioprine |
| 38779031206 | MA11 | Immunosuppressants | azathioprine |
| 66591024141 | MA11 | Immunosuppressants | azathioprine |
| 406200301 | MA11 | Immunosuppressants | azathioprine |
| 38779031211 | MA11 | Immunosuppressants | azathioprine |
| 52372072802 | MA11 | Immunosuppressants | azathioprine |
| 38779031203 | MA11 | Immunosuppressants | azathioprine |
| 68382000305 | MA11 | Immunosuppressants | azathioprine |
| 51079062001 | MA11 | Immunosuppressants | azathioprine |
| 65649024141 | MA11 | Immunosuppressants | azathioprine |
| 68462050201 | MA11 | Immunosuppressants | azathioprine |
| 65649023141 | MA11 | Immunosuppressants | azathioprine |
| 68084022901 | MA11 | Immunosuppressants | azathioprine |
| 51407018201 | MA11 | Immunosuppressants | azathioprine |
| 781507501 | MA11 | Immunosuppressants | azathioprine |
| 52372072804 | MA11 | Immunosuppressants | azathioprine |
| 69238107601 | MA11 | Immunosuppressants | azathioprine |
| 68084022911 | MA11 | Immunosuppressants | azathioprine |
| 43353074509 | MA11 | Immunosuppressants | azathioprine |
| 49452078203 | MA11 | Immunosuppressants | azathioprine |
| 51552077905 | MA11 | Immunosuppressants | azathioprine |
| 49452078201 | MA11 | Immunosuppressants | azathioprine |
| 57866902101 | MA11 | Immunosuppressants | azathioprine |
| 66591023141 | MA11 | Immunosuppressants | azathioprine |
| 781105901 | MA11 | Immunosuppressants | azathioprine |
| 66591022141 | MA11 | Immunosuppressants | azathioprine |
| 23490511009 | MA11 | Immunosuppressants | azathioprine |
| 62991218903 | MA11 | Immunosuppressants | azathioprine |
| 54868092102 | MA11 | Immunosuppressants | azathioprine |
| 66479030110 | MA11 | Immunosuppressants | azathioprine |
| 60219107601 | MA11 | Immunosuppressants | azathioprine |
| 43353068660 | MA11 | Immunosuppressants | azathioprine |
| 54868531006 | MA11 | Immunosuppressants | azathioprine |
| 173059755 | MA11 | Immunosuppressants | azathioprine |
| 49452078305 | MA11 | Immunosuppressants | azathioprine |
| 54868531002 | MA11 | Immunosuppressants | azathioprine |
| 21695048475 | MA11 | Immunosuppressants | azathioprine |
| 71610012409 | MA11 | Immunosuppressants | azathioprine |
| 54868531004 | MA11 | Immunosuppressants | azathioprine |
| 54868531001 | MA11 | Immunosuppressants | azathioprine |
| 54408425 | MA11 | Immunosuppressants | azathioprine |
| 38779031215 | MA11 | Immunosuppressants | azathioprine |
| 51927225800 | MA11 | Immunosuppressants | azathioprine |
| 54766059010 | MA11 | Immunosuppressants | azathioprine |
| 51552077904 | MA11 | Immunosuppressants | azathioprine |
| 52372072801 | MA11 | Immunosuppressants | azathioprine |
| 54868092104 | MA11 | Immunosuppressants | azathioprine |
| 60976059871 | MA11 | Immunosuppressants | azathioprine sodium |
| 143956601 | MA11 | Immunosuppressants | azathioprine sodium |
| 65483055101 | MA11 | Immunosuppressants | azathioprine sodium |
| 173059871 | MA11 | Immunosuppressants | azathioprine sodium |
| 55390060020 | MA11 | Immunosuppressants | azathioprine sodium |
| 78039361 | MA11 | Immunosuppressants | basiliximab |
| 78033184 | MA11 | Immunosuppressants | basiliximab |

| | | | |
|---|---|---|---|
| 78024061 | MA11 | Immunosuppressants | cyclosporine |
| 51552066301 | MA11 | Immunosuppressants | cyclosporine |
| 63307013105 | MA11 | Immunosuppressants | cyclosporine |
| 63307013111 | MA11 | Immunosuppressants | cyclosporine |
| 51552066302 | MA11 | Immunosuppressants | cyclosporine |
| 54868552200 | MA11 | Immunosuppressants | cyclosporine |
| 62991153305 | MA11 | Immunosuppressants | cyclosporine |
| 62991153301 | MA11 | Immunosuppressants | cyclosporine |
| 60505013400 | MA11 | Immunosuppressants | cyclosporine |
| 38779066004 | MA11 | Immunosuppressants | cyclosporine |
| 51927319600 | MA11 | Immunosuppressants | cyclosporine |
| 78024115 | MA11 | Immunosuppressants | cyclosporine |
| 38779066005 | MA11 | Immunosuppressants | cyclosporine |
| 574086610 | MA11 | Immunosuppressants | cyclosporine |
| 51552066309 | MA11 | Immunosuppressants | cyclosporine |
| 38779066001 | MA11 | Immunosuppressants | cyclosporine |
| 62584082721 | MA11 | Immunosuppressants | cyclosporine |
| 70457982601 | MA11 | Immunosuppressants | cyclosporine |
| 70457982603 | MA11 | Immunosuppressants | cyclosporine |
| 68084092125 | MA11 | Immunosuppressants | cyclosporine |
| 63307013137 | MA11 | Immunosuppressants | cyclosporine |
| 51552066306 | MA11 | Immunosuppressants | cyclosporine |
| 46144032325 | MA11 | Immunosuppressants | cyclosporine |
| 395810819 | MA11 | Immunosuppressants | cyclosporine |
| 78010901 | MA11 | Immunosuppressants | cyclosporine |
| 70457982602 | MA11 | Immunosuppressants | cyclosporine |
| 63307013101 | MA11 | Immunosuppressants | cyclosporine |
| 78024015 | MA11 | Immunosuppressants | cyclosporine |
| 68084087995 | MA11 | Immunosuppressants | cyclosporine |
| 38779066008 | MA11 | Immunosuppressants | cyclosporine |
| 78011022 | MA11 | Immunosuppressants | cyclosporine |
| 395810835 | MA11 | Immunosuppressants | cyclosporine |
| 63307013115 | MA11 | Immunosuppressants | cyclosporine |
| 55390012210 | MA11 | Immunosuppressants | cyclosporine |
| 68084087925 | MA11 | Immunosuppressants | cyclosporine |
| 51552066305 | MA11 | Immunosuppressants | cyclosporine |
| 62584082711 | MA11 | Immunosuppressants | cyclosporine |
| 517086601 | MA11 | Immunosuppressants | cyclosporine |
| 395810862 | MA11 | Immunosuppressants | cyclosporine |
| 517086610 | MA11 | Immunosuppressants | cyclosporine |
| 78010961 | MA11 | Immunosuppressants | cyclosporine |
| 51552066304 | MA11 | Immunosuppressants | cyclosporine |
| 60505013300 | MA11 | Immunosuppressants | cyclosporine |
| 62991153302 | MA11 | Immunosuppressants | cyclosporine |
| 68084092195 | MA11 | Immunosuppressants | cyclosporine |
| 38779066006 | MA11 | Immunosuppressants | cyclosporine |
| 63370005710 | MA11 | Immunosuppressants | cyclosporine |
| 78024161 | MA11 | Immunosuppressants | cyclosporine |
| 49452241102 | MA11 | Immunosuppressants | cyclosporine |
| 60432014050 | MA11 | Immunosuppressants | cyclosporine |
| 38779066003 | MA11 | Immunosuppressants | cyclosporine |
| 60505463203 | MA11 | Immunosuppressants | cyclosporine, modified |
| 172731100 | MA11 | Immunosuppressants | cyclosporine, modified |
| 591222455 | MA11 | Immunosuppressants | cyclosporine, modified |
| 74310932 | MA11 | Immunosuppressants | cyclosporine, modified |
| 591222354 | MA11 | Immunosuppressants | cyclosporine, modified |
| 74647932 | MA11 | Immunosuppressants | cyclosporine, modified |
| 60505463003 | MA11 | Immunosuppressants | cyclosporine, modified |
| 50111092043 | MA11 | Immunosuppressants | cyclosporine, modified |
| 93574165 | MA11 | Immunosuppressants | cyclosporine, modified |
| 51862046047 | MA11 | Immunosuppressants | cyclosporine, modified |
| 74310832 | MA11 | Immunosuppressants | cyclosporine, modified |
| 172731146 | MA11 | Immunosuppressants | cyclosporine, modified |
| 93574019 | MA11 | Immunosuppressants | cyclosporine, modified |
| 93574119 | MA11 | Immunosuppressants | cyclosporine, modified |
| 185093287 | MA11 | Immunosuppressants | cyclosporine, modified |
| 78024661 | MA11 | Immunosuppressants | cyclosporine, modified |
| 62053053905 | MA11 | Immunosuppressants | cyclosporine, modified |
| 185093386 | MA11 | Immunosuppressants | cyclosporine, modified |
| 172731000 | MA11 | Immunosuppressants | cyclosporine, modified |
| 172731320 | MA11 | Immunosuppressants | cyclosporine, modified |

| | | | |
|---|---|---|---|
| 185093230 | MA11 | Immunosuppressants | cyclosporine, modified |
| 78024615 | MA11 | Immunosuppressants | cyclosporine, modified |
| 50111088542 | MA11 | Immunosuppressants | cyclosporine, modified |
| 78027422 | MA11 | Immunosuppressants | cyclosporine, modified |
| 74054130 | MA11 | Immunosuppressants | cyclosporine, modified |
| 51862046001 | MA11 | Immunosuppressants | cyclosporine, modified |
| 172731046 | MA11 | Immunosuppressants | cyclosporine, modified |
| 50111090943 | MA11 | Immunosuppressants | cyclosporine, modified |
| 78024815 | MA11 | Immunosuppressants | cyclosporine, modified |
| 51862045801 | MA11 | Immunosuppressants | cyclosporine, modified |
| 54868623200 | MA11 | Immunosuppressants | cyclosporine, modified |
| 51862045847 | MA11 | Immunosuppressants | cyclosporine, modified |
| 172731200 | MA11 | Immunosuppressants | cyclosporine, modified |
| 74646332 | MA11 | Immunosuppressants | cyclosporine, modified |
| 185093330 | MA11 | Immunosuppressants | cyclosporine, modified |
| 172731246 | MA11 | Immunosuppressants | cyclosporine, modified |
| 93574265 | MA11 | Immunosuppressants | cyclosporine, modified |
| 185093387 | MA11 | Immunosuppressants | cyclosporine, modified |
| 93574065 | MA11 | Immunosuppressants | cyclosporine, modified |
| 74726950 | MA11 | Immunosuppressants | cyclosporine, modified |
| 591222315 | MA11 | Immunosuppressants | cyclosporine, modified |
| 60505035401 | MA11 | Immunosuppressants | cyclosporine, modified |
| 591222215 | MA11 | Immunosuppressants | cyclosporine, modified |
| 60505463103 | MA11 | Immunosuppressants | cyclosporine, modified |
| 78024861 | MA11 | Immunosuppressants | cyclosporine, modified |
| 93574219 | MA11 | Immunosuppressants | cyclosporine, modified |
| 4050109 | MA11 | Immunosuppressants | daclizumab |
| 74003301 | MA11 | Immunosuppressants | daclizumab |
| 54569494200 | MA11 | Immunosuppressants | daclizumab |
| 72171050501 | MA11 | Immunosuppressants | emapalumab-lzsg |
| 72171050101 | MA11 | Immunosuppressants | emapalumab-lzsg |
| 49884011991 | MA11 | Immunosuppressants | everolimus |
| 78056751 | MA11 | Immunosuppressants | everolimus |
| 78062861 | MA11 | Immunosuppressants | everolimus |
| 78059451 | MA11 | Immunosuppressants | everolimus |
| 78042261 | MA11 | Immunosuppressants | everolimus |
| 78056761 | MA11 | Immunosuppressants | everolimus |
| 49884012791 | MA11 | Immunosuppressants | everolimus |
| 78062761 | MA11 | Immunosuppressants | everolimus |
| 78056661 | MA11 | Immunosuppressants | everolimus |
| 78062061 | MA11 | Immunosuppressants | everolimus |
| 78062051 | MA11 | Immunosuppressants | everolimus |
| 78062751 | MA11 | Immunosuppressants | everolimus |
| 78042220 | MA11 | Immunosuppressants | everolimus |
| 78041520 | MA11 | Immunosuppressants | everolimus |
| 78056651 | MA11 | Immunosuppressants | everolimus |
| 49884012752 | MA11 | Immunosuppressants | everolimus |
| 78062851 | MA11 | Immunosuppressants | everolimus |
| 78041761 | MA11 | Immunosuppressants | everolimus |
| 78041561 | MA11 | Immunosuppressants | everolimus |
| 49884012552 | MA11 | Immunosuppressants | everolimus |
| 78041461 | MA11 | Immunosuppressants | everolimus |
| 49884012591 | MA11 | Immunosuppressants | everolimus |
| 49884011952 | MA11 | Immunosuppressants | everolimus |
| 78041420 | MA11 | Immunosuppressants | everolimus |
| 78062651 | MA11 | Immunosuppressants | everolimus |
| 78041720 | MA11 | Immunosuppressants | everolimus |
| 78059461 | MA11 | Immunosuppressants | everolimus |
| 78062661 | MA11 | Immunosuppressants | everolimus |
| 54458127 | MA11 | Immunosuppressants | mercaptopurine |
| 62484002001 | MA11 | Immunosuppressants | mercaptopurine |
| 57844052206 | MA11 | Immunosuppressants | mercaptopurine |
| 173080765 | MA11 | Immunosuppressants | mercaptopurine |
| 68084032521 | MA11 | Immunosuppressants | mercaptopurine |
| 38779142703 | MA11 | Immunosuppressants | mercaptopurine |
| 38779142706 | MA11 | Immunosuppressants | mercaptopurine |
| 38779142704 | MA11 | Immunosuppressants | mercaptopurine |
| 68084032511 | MA11 | Immunosuppressants | mercaptopurine |
| 62991289902 | MA11 | Immunosuppressants | mercaptopurine |
| 60311045001 | MA11 | Immunosuppressants | mercaptopurine |
| 69076091302 | MA11 | Immunosuppressants | mercaptopurine |

| | | | |
|---|---|---|---|
| 49884092204 | MA11 | Immunosuppressants | mercaptopurine |
| 49452446302 | MA11 | Immunosuppressants | mercaptopurine |
| 57844052252 | MA11 | Immunosuppressants | mercaptopurine |
| 49884092202 | MA11 | Immunosuppressants | mercaptopurine |
| 173080725 | MA11 | Immunosuppressants | mercaptopurine |
| 378354725 | MA11 | Immunosuppressants | mercaptopurine |
| 51927200000 | MA11 | Immunosuppressants | mercaptopurine |
| 93551006 | MA11 | Immunosuppressants | mercaptopurine |
| 62484002002 | MA11 | Immunosuppressants | mercaptopurine |
| 62991289901 | MA11 | Immunosuppressants | mercaptopurine |
| 378354752 | MA11 | Immunosuppressants | mercaptopurine |
| 54458111 | MA11 | Immunosuppressants | mercaptopurine |
| 54868528201 | MA11 | Immunosuppressants | mercaptopurine |
| 57844052207 | MA11 | Immunosuppressants | mercaptopurine |
| 62484000202 | MA11 | Immunosuppressants | mercaptopurine |
| 54868528200 | MA11 | Immunosuppressants | mercaptopurine |
| 54868528202 | MA11 | Immunosuppressants | mercaptopurine |
| 68258910301 | MA11 | Immunosuppressants | mercaptopurine |
| 62991289903 | MA11 | Immunosuppressants | mercaptopurine |
| 69076091325 | MA11 | Immunosuppressants | mercaptopurine |
| 63370015410 | MA11 | Immunosuppressants | methotrexate |
| 38779003504 | MA11 | Immunosuppressants | methotrexate |
| 49452460102 | MA11 | Immunosuppressants | methotrexate |
| 62991120001 | MA11 | Immunosuppressants | methotrexate |
| 51552105401 | MA11 | Immunosuppressants | methotrexate |
| 38779003515 | MA11 | Immunosuppressants | methotrexate |
| 38779003510 | MA11 | Immunosuppressants | methotrexate |
| 38779003503 | MA11 | Immunosuppressants | methotrexate |
| 49452460104 | MA11 | Immunosuppressants | methotrexate |
| 38779003525 | MA11 | Immunosuppressants | methotrexate |
| 52652200106 | MA11 | Immunosuppressants | methotrexate |
| 49452460002 | MA11 | Immunosuppressants | methotrexate |
| 63370015415 | MA11 | Immunosuppressants | methotrexate |
| 49452460101 | MA11 | Immunosuppressants | methotrexate |
| 62991120004 | MA11 | Immunosuppressants | methotrexate |
| 62991120002 | MA11 | Immunosuppressants | methotrexate |
| 38779003511 | MA11 | Immunosuppressants | methotrexate |
| 52652200101 | MA11 | Immunosuppressants | methotrexate |
| 63370015425 | MA11 | Immunosuppressants | methotrexate |
| 51927156500 | MA11 | Immunosuppressants | methotrexate |
| 49452460003 | MA11 | Immunosuppressants | methotrexate |
| 51552105409 | MA11 | Immunosuppressants | methotrexate |
| 49452460103 | MA11 | Immunosuppressants | methotrexate |
| 49452460001 | MA11 | Immunosuppressants | methotrexate |
| 38779003506 | MA11 | Immunosuppressants | methotrexate |
| 55289092430 | MA11 | Immunosuppressants | methotrexate sodium |
| 61703035037 | MA11 | Immunosuppressants | methotrexate sodium |
| 67253058046 | MA11 | Immunosuppressants | methotrexate sodium |
| 21695011100 | MA11 | Immunosuppressants | methotrexate sodium |
| 67253032010 | MA11 | Immunosuppressants | methotrexate sodium |
| 50090029409 | MA11 | Immunosuppressants | methotrexate sodium |
| 51285036901 | MA11 | Immunosuppressants | methotrexate sodium |
| 69238142306 | MA11 | Immunosuppressants | methotrexate sodium |
| 51285036601 | MA11 | Immunosuppressants | methotrexate sodium |
| 21695011130 | MA11 | Immunosuppressants | methotrexate sodium |
| 54855003 | MA11 | Immunosuppressants | methotrexate sodium |
| 51285036801 | MA11 | Immunosuppressants | methotrexate sodium |
| 10019094101 | MA11 | Immunosuppressants | methotrexate sodium |
| 51079067005 | MA11 | Immunosuppressants | methotrexate sodium |
| 63629147202 | MA11 | Immunosuppressants | methotrexate sodium |
| 63323012310 | MA11 | Immunosuppressants | methotrexate sodium |
| 51285050902 | MA11 | Immunosuppressants | methotrexate sodium |
| 54855006 | MA11 | Immunosuppressants | methotrexate sodium |
| 555057246 | MA11 | Immunosuppressants | methotrexate sodium |
| 54855010 | MA11 | Immunosuppressants | methotrexate sodium |
| 43063043930 | MA11 | Immunosuppressants | methotrexate sodium |
| 555057245 | MA11 | Immunosuppressants | methotrexate sodium |
| 42291059401 | MA11 | Immunosuppressants | methotrexate sodium |
| 5450709 | MA11 | Immunosuppressants | methotrexate sodium |
| 61703040707 | MA11 | Immunosuppressants | methotrexate sodium |
| 60429008036 | MA11 | Immunosuppressants | methotrexate sodium |

| | | | |
|---|---|---|---|
| 51285036701 | MA11 | Immunosuppressants | methotrexate sodium |
| 54855005 | MA11 | Immunosuppressants | methotrexate sodium |
| 54868382605 | MA11 | Immunosuppressants | methotrexate sodium |
| 66479013509 | MA11 | Immunosuppressants | methotrexate sodium |
| 904601260 | MA11 | Immunosuppressants | methotrexate sodium |
| 54868382606 | MA11 | Immunosuppressants | methotrexate sodium |
| 54455025 | MA11 | Immunosuppressants | methotrexate sodium |
| 67253058045 | MA11 | Immunosuppressants | methotrexate sodium |
| 67253058043 | MA11 | Immunosuppressants | methotrexate sodium |
| 63629147201 | MA11 | Immunosuppressants | methotrexate sodium |
| 54569531600 | MA11 | Immunosuppressants | methotrexate sodium |
| 58406068114 | MA11 | Immunosuppressants | methotrexate sodium |
| 47335023583 | MA11 | Immunosuppressants | methotrexate sodium |
| 42254011030 | MA11 | Immunosuppressants | methotrexate sodium |
| 54868382600 | MA11 | Immunosuppressants | methotrexate sodium |
| 54868382602 | MA11 | Immunosuppressants | methotrexate sodium |
| 5450704 | MA11 | Immunosuppressants | methotrexate sodium |
| 58406068117 | MA11 | Immunosuppressants | methotrexate sodium |
| 61703035010 | MA11 | Immunosuppressants | methotrexate sodium |
| 61703035038 | MA11 | Immunosuppressants | methotrexate sodium |
| 61703035009 | MA11 | Immunosuppressants | methotrexate sodium |
| 904174960 | MA11 | Immunosuppressants | methotrexate sodium |
| 904174973 | MA11 | Immunosuppressants | methotrexate sodium |
| 555092701 | MA11 | Immunosuppressants | methotrexate sodium |
| 67253032036 | MA11 | Immunosuppressants | methotrexate sodium |
| 555094501 | MA11 | Immunosuppressants | methotrexate sodium |
| 63323012302 | MA11 | Immunosuppressants | methotrexate sodium |
| 54868382607 | MA11 | Immunosuppressants | methotrexate sodium |
| 66479013501 | MA11 | Immunosuppressants | methotrexate sodium |
| 378001450 | MA11 | Immunosuppressants | methotrexate sodium |
| 59911587401 | MA11 | Immunosuppressants | methotrexate sodium |
| 54868382608 | MA11 | Immunosuppressants | methotrexate sodium |
| 62701094099 | MA11 | Immunosuppressants | methotrexate sodium |
| 49999038036 | MA11 | Immunosuppressants | methotrexate sodium |
| 62701094036 | MA11 | Immunosuppressants | methotrexate sodium |
| 51079067001 | MA11 | Immunosuppressants | methotrexate sodium |
| 68382077501 | MA11 | Immunosuppressants | methotrexate sodium |
| 54855007 | MA11 | Immunosuppressants | methotrexate sodium |
| 61703040858 | MA11 | Immunosuppressants | methotrexate sodium |
| 67253058044 | MA11 | Immunosuppressants | methotrexate sodium |
| 555057235 | MA11 | Immunosuppressants | methotrexate sodium |
| 54868382603 | MA11 | Immunosuppressants | methotrexate sodium |
| 555092901 | MA11 | Immunosuppressants | methotrexate sodium |
| 54868382601 | MA11 | Immunosuppressants | methotrexate sodium |
| 182153995 | MA11 | Immunosuppressants | methotrexate sodium |
| 5450723 | MA11 | Immunosuppressants | methotrexate sodium |
| 54569181809 | MA11 | Immunosuppressants | methotrexate sodium |
| 182153901 | MA11 | Immunosuppressants | methotrexate sodium |
| 49999038024 | MA11 | Immunosuppressants | methotrexate sodium |
| 51407012101 | MA11 | Immunosuppressants | methotrexate sodium |
| 23490588900 | MA11 | Immunosuppressants | methotrexate sodium |
| 5450791 | MA11 | Immunosuppressants | methotrexate sodium |
| 50090234509 | MA11 | Immunosuppressants | methotrexate sodium |
| 54868382609 | MA11 | Immunosuppressants | methotrexate sodium |
| 61703040822 | MA11 | Immunosuppressants | methotrexate sodium |
| 555092801 | MA11 | Immunosuppressants | methotrexate sodium |
| 61703040732 | MA11 | Immunosuppressants | methotrexate sodium |
| 67253058042 | MA11 | Immunosuppressants | methotrexate sodium |
| 54868479600 | MA11 | Immunosuppressants | methotrexate sodium |
| 47335023596 | MA11 | Immunosuppressants | methotrexate sodium |
| 603449921 | MA11 | Immunosuppressants | methotrexate sodium |
| 5450707 | MA11 | Immunosuppressants | methotrexate sodium |
| 5450705 | MA11 | Immunosuppressants | methotrexate sodium |
| 555057202 | MA11 | Immunosuppressants | methotrexate sodium |
| 54455015 | MA11 | Immunosuppressants | methotrexate sodium |
| 54569181800 | MA11 | Immunosuppressants | methotrexate sodium |
| 555057248 | MA11 | Immunosuppressants | methotrexate sodium |
| 52959024400 | MA11 | Immunosuppressants | methotrexate sodium |
| 54868382604 | MA11 | Immunosuppressants | methotrexate sodium |
| 555057249 | MA11 | Immunosuppressants | methotrexate sodium |
| 69238142301 | MA11 | Immunosuppressants | methotrexate sodium |

| | | | |
|---|---|---|---|
| 68115063200 | MA11 | Immunosuppressants | methotrexate sodium |
| 378001401 | MA11 | Immunosuppressants | methotrexate sodium |
| 555057247 | MA11 | Immunosuppressants | methotrexate sodium |
| 54855025 | MA11 | Immunosuppressants | methotrexate sodium |
| 50090341802 | MA11 | Immunosuppressants | methotrexate sodium |
| 10139006202 | MA11 | Immunosuppressants | methotrexate sodium/PF |
| 69020410 | MA11 | Immunosuppressants | methotrexate sodium/PF |
| 54868471600 | MA11 | Immunosuppressants | methotrexate sodium/PF |
| 55390003210 | MA11 | Immunosuppressants | methotrexate sodium/PF |
| 63323012140 | MA11 | Immunosuppressants | methotrexate sodium/PF |
| 69014702 | MA11 | Immunosuppressants | methotrexate sodium/PF |
| 61703040813 | MA11 | Immunosuppressants | methotrexate sodium/PF |
| 67457022140 | MA11 | Immunosuppressants | methotrexate sodium/PF |
| 703367193 | MA11 | Immunosuppressants | methotrexate sodium/PF |
| 703367101 | MA11 | Immunosuppressants | methotrexate sodium/PF |
| 67457048604 | MA11 | Immunosuppressants | methotrexate sodium/PF |
| 55390003310 | MA11 | Immunosuppressants | methotrexate sodium/PF |
| 703367801 | MA11 | Immunosuppressants | methotrexate sodium/PF |
| 703367301 | MA11 | Immunosuppressants | methotrexate sodium/PF |
| 703367881 | MA11 | Immunosuppressants | methotrexate sodium/PF |
| 69020401 | MA11 | Immunosuppressants | methotrexate sodium/PF |
| 16729027703 | MA11 | Immunosuppressants | methotrexate sodium/PF |
| 54868017301 | MA11 | Immunosuppressants | methotrexate sodium/PF |
| 143951901 | MA11 | Immunosuppressants | methotrexate sodium/PF |
| 703367591 | MA11 | Immunosuppressants | methotrexate sodium/PF |
| 63323012250 | MA11 | Immunosuppressants | methotrexate sodium/PF |
| 10139006240 | MA11 | Immunosuppressants | methotrexate sodium/PF |
| 66479013929 | MA11 | Immunosuppressants | methotrexate sodium/PF |
| 55390003410 | MA11 | Immunosuppressants | methotrexate sodium/PF |
| 63323012259 | MA11 | Immunosuppressants | methotrexate sodium/PF |
| 67457022110 | MA11 | Immunosuppressants | methotrexate sodium/PF |
| 61703040832 | MA11 | Immunosuppressants | methotrexate sodium/PF |
| 69014601 | MA11 | Immunosuppressants | methotrexate sodium/PF |
| 63323012110 | MA11 | Immunosuppressants | methotrexate sodium/PF |
| 69018102 | MA11 | Immunosuppressants | methotrexate sodium/PF |
| 67457046721 | MA11 | Immunosuppressants | methotrexate sodium/PF |
| 63323012102 | MA11 | Immunosuppressants | methotrexate sodium/PF |
| 67457046799 | MA11 | Immunosuppressants | methotrexate sodium/PF |
| 67457048508 | MA11 | Immunosuppressants | methotrexate sodium/PF |
| 10139006210 | MA11 | Immunosuppressants | methotrexate sodium/PF |
| 143951910 | MA11 | Immunosuppressants | methotrexate sodium/PF |
| 703367191 | MA11 | Immunosuppressants | methotrexate sodium/PF |
| 69014801 | MA11 | Immunosuppressants | methotrexate sodium/PF |
| 58406068318 | MA11 | Immunosuppressants | methotrexate sodium/PF |
| 66479013613 | MA11 | Immunosuppressants | methotrexate sodium/PF |
| 63323012104 | MA11 | Immunosuppressants | methotrexate sodium/PF |
| 67457046610 | MA11 | Immunosuppressants | methotrexate sodium/PF |
| 16729027735 | MA11 | Immunosuppressants | methotrexate sodium/PF |
| 61703040807 | MA11 | Immunosuppressants | methotrexate sodium/PF |
| 66479013611 | MA11 | Immunosuppressants | methotrexate sodium/PF |
| 143983001 | MA11 | Immunosuppressants | methotrexate sodium/PF |
| 58406067105 | MA11 | Immunosuppressants | methotrexate sodium/PF |
| 16729027730 | MA11 | Immunosuppressants | methotrexate sodium/PF |
| 69018101 | MA11 | Immunosuppressants | methotrexate sodium/PF |
| 66479013721 | MA11 | Immunosuppressants | methotrexate sodium/PF |
| 67457048699 | MA11 | Immunosuppressants | methotrexate sodium/PF |
| 69014701 | MA11 | Immunosuppressants | methotrexate sodium/PF |
| 703367501 | MA11 | Immunosuppressants | methotrexate sodium/PF |
| 66758004101 | MA11 | Immunosuppressants | methotrexate sodium/PF |
| 58406068315 | MA11 | Immunosuppressants | methotrexate sodium/PF |
| 69014901 | MA11 | Immunosuppressants | methotrexate sodium/PF |
| 55390003110 | MA11 | Immunosuppressants | methotrexate sodium/PF |
| 69014602 | MA11 | Immunosuppressants | methotrexate sodium/PF |
| 66758004002 | MA11 | Immunosuppressants | methotrexate sodium/PF |
| 67457048599 | MA11 | Immunosuppressants | methotrexate sodium/PF |
| 67457022102 | MA11 | Immunosuppressants | methotrexate sodium/PF |
| 58406067301 | MA11 | Immunosuppressants | methotrexate sodium/PF |
| 61703040841 | MA11 | Immunosuppressants | methotrexate sodium/PF |
| 54569452500 | MA11 | Immunosuppressants | methotrexate sodium/PF |
| 55390014301 | MA11 | Immunosuppressants | methotrexate sodium/PF |
| 10019094001 | MA11 | Immunosuppressants | methotrexate sodium/PF |

| | | | |
|---|---|---|---|
| 66758004008 | MA11 | Immunosuppressants | methotrexate sodium/PF |
| 66758004001 | MA11 | Immunosuppressants | methotrexate sodium/PF |
| 58406068316 | MA11 | Immunosuppressants | methotrexate sodium/PF |
| 703367103 | MA11 | Immunosuppressants | methotrexate sodium/PF |
| 61703040804 | MA11 | Immunosuppressants | methotrexate sodium/PF |
| 67457048040 | MA11 | Immunosuppressants | methotrexate sodium/PF |
| 61703040825 | MA11 | Immunosuppressants | methotrexate sodium/PF |
| 10019094002 | MA11 | Immunosuppressants | methotrexate sodium/PF |
| 66479013619 | MA11 | Immunosuppressants | methotrexate sodium/PF |
| 63323012108 | MA11 | Immunosuppressants | methotrexate sodium/PF |
| 54436002502 | MA11 | Immunosuppressants | methotrexate/PF |
| 54436002204 | MA11 | Immunosuppressants | methotrexate/PF |
| 54436001504 | MA11 | Immunosuppressants | methotrexate/PF |
| 54436002002 | MA11 | Immunosuppressants | methotrexate/PF |
| 59137052000 | MA11 | Immunosuppressants | methotrexate/PF |
| 54436001704 | MA11 | Immunosuppressants | methotrexate/PF |
| 54436001002 | MA11 | Immunosuppressants | methotrexate/PF |
| 59137051504 | MA11 | Immunosuppressants | methotrexate/PF |
| 59137051500 | MA11 | Immunosuppressants | methotrexate/PF |
| 59137054500 | MA11 | Immunosuppressants | methotrexate/PF |
| 59137053504 | MA11 | Immunosuppressants | methotrexate/PF |
| 54436002504 | MA11 | Immunosuppressants | methotrexate/PF |
| 54436001202 | MA11 | Immunosuppressants | methotrexate/PF |
| 59137053000 | MA11 | Immunosuppressants | methotrexate/PF |
| 59137052004 | MA11 | Immunosuppressants | methotrexate/PF |
| 54436001204 | MA11 | Immunosuppressants | methotrexate/PF |
| 59137055000 | MA11 | Immunosuppressants | methotrexate/PF |
| 54436007504 | MA11 | Immunosuppressants | methotrexate/PF |
| 54436001004 | MA11 | Immunosuppressants | methotrexate/PF |
| 54436002004 | MA11 | Immunosuppressants | methotrexate/PF |
| 59137052500 | MA11 | Immunosuppressants | methotrexate/PF |
| 54436001502 | MA11 | Immunosuppressants | methotrexate/PF |
| 59137051000 | MA11 | Immunosuppressants | methotrexate/PF |
| 59137054504 | MA11 | Immunosuppressants | methotrexate/PF |
| 54436002202 | MA11 | Immunosuppressants | methotrexate/PF |
| 54436001702 | MA11 | Immunosuppressants | methotrexate/PF |
| 59137050504 | MA11 | Immunosuppressants | methotrexate/PF |
| 59137055004 | MA11 | Immunosuppressants | methotrexate/PF |
| 59137052504 | MA11 | Immunosuppressants | methotrexate/PF |
| 59137053500 | MA11 | Immunosuppressants | methotrexate/PF |
| 59137050500 | MA11 | Immunosuppressants | methotrexate/PF |
| 54436007502 | MA11 | Immunosuppressants | methotrexate/PF |
| 59137053004 | MA11 | Immunosuppressants | methotrexate/PF |
| 59137053001 | MA11 | Immunosuppressants | methotrexate/PF |
| 59137051004 | MA11 | Immunosuppressants | methotrexate/PF |
| 59137054000 | MA11 | Immunosuppressants | methotrexate/PF |
| 59137054004 | MA11 | Immunosuppressants | methotrexate/PF |
| 59676010101 | MA11 | Immunosuppressants | muromonab-CD3 |
| 4025901 | MA11 | Immunosuppressants | mycophenolate mofetil |
| 71610003370 | MA11 | Immunosuppressants | mycophenolate mofetil |
| 68084017801 | MA11 | Immunosuppressants | mycophenolate mofetil |
| 64380072506 | MA11 | Immunosuppressants | mycophenolate mofetil |
| 70748018602 | MA11 | Immunosuppressants | mycophenolate mofetil |
| 68382013110 | MA11 | Immunosuppressants | mycophenolate mofetil |
| 378447205 | MA11 | Immunosuppressants | mycophenolate mofetil |
| 68084058701 | MA11 | Immunosuppressants | mycophenolate mofetil |
| 54016325 | MA11 | Immunosuppressants | mycophenolate mofetil |
| 67263040701 | MA11 | Immunosuppressants | mycophenolate mofetil |
| 68258905201 | MA11 | Immunosuppressants | mycophenolate mofetil |
| 68382013005 | MA11 | Immunosuppressants | mycophenolate mofetil |
| 60429007005 | MA11 | Immunosuppressants | mycophenolate mofetil |
| 71610003360 | MA11 | Immunosuppressants | mycophenolate mofetil |
| 68084058801 | MA11 | Immunosuppressants | mycophenolate mofetil |
| 16729001901 | MA11 | Immunosuppressants | mycophenolate mofetil |
| 59762070301 | MA11 | Immunosuppressants | mycophenolate mofetil |
| 67877022501 | MA11 | Immunosuppressants | mycophenolate mofetil |
| 60951073585 | MA11 | Immunosuppressants | mycophenolate mofetil |
| 71610003390 | MA11 | Immunosuppressants | mycophenolate mofetil |
| 60951073470 | MA11 | Immunosuppressants | mycophenolate mofetil |
| 60505296801 | MA11 | Immunosuppressants | mycophenolate mofetil |
| 93733405 | MA11 | Immunosuppressants | mycophenolate mofetil |

| | | | |
|---|---|---|---|
| 68084017701 | MA11 | Immunosuppressants | mycophenolate mofetil |
| 68382013019 | MA11 | Immunosuppressants | mycophenolate mofetil |
| 60505296701 | MA11 | Immunosuppressants | mycophenolate mofetil |
| 781206705 | MA11 | Immunosuppressants | mycophenolate mofetil |
| 21695017100 | MA11 | Immunosuppressants | mycophenolate mofetil |
| 67877022505 | MA11 | Immunosuppressants | mycophenolate mofetil |
| 68084080101 | MA11 | Immunosuppressants | mycophenolate mofetil |
| 70748018601 | MA11 | Immunosuppressants | mycophenolate mofetil |
| 68084079511 | MA11 | Immunosuppressants | mycophenolate mofetil |
| 93733493 | MA11 | Immunosuppressants | mycophenolate mofetil |
| 59762070203 | MA11 | Immunosuppressants | mycophenolate mofetil |
| 64380072507 | MA11 | Immunosuppressants | mycophenolate mofetil |
| 68084017811 | MA11 | Immunosuppressants | mycophenolate mofetil |
| 59762070201 | MA11 | Immunosuppressants | mycophenolate mofetil |
| 378447201 | MA11 | Immunosuppressants | mycophenolate mofetil |
| 93747701 | MA11 | Immunosuppressants | mycophenolate mofetil |
| 16729009416 | MA11 | Immunosuppressants | mycophenolate mofetil |
| 781517505 | MA11 | Immunosuppressants | mycophenolate mofetil |
| 93733401 | MA11 | Immunosuppressants | mycophenolate mofetil |
| 781206789 | MA11 | Immunosuppressants | mycophenolate mofetil |
| 60429005905 | MA11 | Immunosuppressants | mycophenolate mofetil |
| 60505296807 | MA11 | Immunosuppressants | mycophenolate mofetil |
| 60951073570 | MA11 | Immunosuppressants | mycophenolate mofetil |
| 4025943 | MA11 | Immunosuppressants | mycophenolate mofetil |
| 68084079501 | MA11 | Immunosuppressants | mycophenolate mofetil |
| 67877026601 | MA11 | Immunosuppressants | mycophenolate mofetil |
| 60429007001 | MA11 | Immunosuppressants | mycophenolate mofetil |
| 68382013010 | MA11 | Immunosuppressants | mycophenolate mofetil |
| 378225005 | MA11 | Immunosuppressants | mycophenolate mofetil |
| 51079072120 | MA11 | Immunosuppressants | mycophenolate mofetil |
| 71610003375 | MA11 | Immunosuppressants | mycophenolate mofetil |
| 71610003394 | MA11 | Immunosuppressants | mycophenolate mofetil |
| 59762070302 | MA11 | Immunosuppressants | mycophenolate mofetil |
| 68382013001 | MA11 | Immunosuppressants | mycophenolate mofetil |
| 51079072101 | MA11 | Immunosuppressants | mycophenolate mofetil |
| 60951073485 | MA11 | Immunosuppressants | mycophenolate mofetil |
| 16729001916 | MA11 | Immunosuppressants | mycophenolate mofetil |
| 4026129 | MA11 | Immunosuppressants | mycophenolate mofetil |
| 68084080111 | MA11 | Immunosuppressants | mycophenolate mofetil |
| 54016625 | MA11 | Immunosuppressants | mycophenolate mofetil |
| 16729009401 | MA11 | Immunosuppressants | mycophenolate mofetil |
| 781517501 | MA11 | Immunosuppressants | mycophenolate mofetil |
| 4025905 | MA11 | Immunosuppressants | mycophenolate mofetil |
| 781206701 | MA11 | Immunosuppressants | mycophenolate mofetil |
| 54016629 | MA11 | Immunosuppressants | mycophenolate mofetil |
| 378225001 | MA11 | Immunosuppressants | mycophenolate mofetil |
| 49999093630 | MA11 | Immunosuppressants | mycophenolate mofetil |
| 54016329 | MA11 | Immunosuppressants | mycophenolate mofetil |
| 60505296805 | MA11 | Immunosuppressants | mycophenolate mofetil |
| 68084017711 | MA11 | Immunosuppressants | mycophenolate mofetil |
| 71610003353 | MA11 | Immunosuppressants | mycophenolate mofetil |
| 51079037920 | MA11 | Immunosuppressants | mycophenolate mofetil |
| 50268058115 | MA11 | Immunosuppressants | mycophenolate mofetil |
| 68382013105 | MA11 | Immunosuppressants | mycophenolate mofetil |
| 51079037901 | MA11 | Immunosuppressants | mycophenolate mofetil |
| 66689030708 | MA11 | Immunosuppressants | mycophenolate mofetil |
| 4026001 | MA11 | Immunosuppressants | mycophenolate mofetil |
| 49999093600 | MA11 | Immunosuppressants | mycophenolate mofetil |
| 64380072607 | MA11 | Immunosuppressants | mycophenolate mofetil |
| 49999093730 | MA11 | Immunosuppressants | mycophenolate mofetil |
| 60429005901 | MA11 | Immunosuppressants | mycophenolate mofetil |
| 93747705 | MA11 | Immunosuppressants | mycophenolate mofetil |
| 68084058811 | MA11 | Immunosuppressants | mycophenolate mofetil |
| 60687043801 | MA11 | Immunosuppressants | mycophenolate mofetil |
| 59762070303 | MA11 | Immunosuppressants | mycophenolate mofetil |
| 71610003392 | MA11 | Immunosuppressants | mycophenolate mofetil |
| 4026043 | MA11 | Immunosuppressants | mycophenolate mofetil |
| 71610003398 | MA11 | Immunosuppressants | mycophenolate mofetil |
| 71610003380 | MA11 | Immunosuppressants | mycophenolate mofetil |
| 93733419 | MA11 | Immunosuppressants | mycophenolate mofetil |
| 68258907301 | MA11 | Immunosuppressants | mycophenolate mofetil |

| | | | |
|---|---|---|---|
| 60505296705 | MA11 | Immunosuppressants | mycophenolate mofetil |
| 67877026605 | MA11 | Immunosuppressants | mycophenolate mofetil |
| 50268058111 | MA11 | Immunosuppressants | mycophenolate mofetil |
| 60505296707 | MA11 | Immunosuppressants | mycophenolate mofetil |
| 67877023022 | MA11 | Immunosuppressants | mycophenolate mofetil |
| 68084058711 | MA11 | Immunosuppressants | mycophenolate mofetil |
| 60687043811 | MA11 | Immunosuppressants | mycophenolate mofetil |
| 64380072606 | MA11 | Immunosuppressants | mycophenolate mofetil |
| 68382013101 | MA11 | Immunosuppressants | mycophenolate mofetil |
| 17478042240 | MA11 | Immunosuppressants | mycophenolate mofetil HCl |
| 42023017204 | MA11 | Immunosuppressants | mycophenolate mofetil HCl |
| 4029830 | MA11 | Immunosuppressants | mycophenolate mofetil HCl |
| 4029809 | MA11 | Immunosuppressants | mycophenolate mofetil HCl |
| 60429001612 | MA11 | Immunosuppressants | mycophenolate sodium |
| 51079050820 | MA11 | Immunosuppressants | mycophenolate sodium |
| 67263037812 | MA11 | Immunosuppressants | mycophenolate sodium |
| 51079050920 | MA11 | Immunosuppressants | mycophenolate sodium |
| 378420178 | MA11 | Immunosuppressants | mycophenolate sodium |
| 68084091825 | MA11 | Immunosuppressants | mycophenolate sodium |
| 68084090711 | MA11 | Immunosuppressants | mycophenolate sodium |
| 68084091895 | MA11 | Immunosuppressants | mycophenolate sodium |
| 51079050801 | MA11 | Immunosuppressants | mycophenolate sodium |
| 904678604 | MA11 | Immunosuppressants | mycophenolate sodium |
| 51079050901 | MA11 | Immunosuppressants | mycophenolate sodium |
| 78038566 | MA11 | Immunosuppressants | mycophenolate sodium |
| 904678561 | MA11 | Immunosuppressants | mycophenolate sodium |
| 60429001712 | MA11 | Immunosuppressants | mycophenolate sodium |
| 78038666 | MA11 | Immunosuppressants | mycophenolate sodium |
| 60505296607 | MA11 | Immunosuppressants | mycophenolate sodium |
| 904678504 | MA11 | Immunosuppressants | mycophenolate sodium |
| 904678661 | MA11 | Immunosuppressants | mycophenolate sodium |
| 93703189 | MA11 | Immunosuppressants | mycophenolate sodium |
| 378420278 | MA11 | Immunosuppressants | mycophenolate sodium |
| 60505296507 | MA11 | Immunosuppressants | mycophenolate sodium |
| 93703289 | MA11 | Immunosuppressants | mycophenolate sodium |
| 16729018929 | MA11 | Immunosuppressants | mycophenolate sodium |
| 67263037912 | MA11 | Immunosuppressants | mycophenolate sodium |
| 16729026129 | MA11 | Immunosuppressants | mycophenolate sodium |
| 68084090721 | MA11 | Immunosuppressants | mycophenolate sodium |
| 8104105 | MA11 | Immunosuppressants | sirolimus |
| 55111065401 | MA11 | Immunosuppressants | sirolimus |
| 8103007 | MA11 | Immunosuppressants | sirolimus |
| 8104005 | MA11 | Immunosuppressants | sirolimus |
| 8103002 | MA11 | Immunosuppressants | sirolimus |
| 68084091595 | MA11 | Immunosuppressants | sirolimus |
| 8103110 | MA11 | Immunosuppressants | sirolimus |
| 8103006 | MA11 | Immunosuppressants | sirolimus |
| 69238159403 | MA11 | Immunosuppressants | sirolimus |
| 8103005 | MA11 | Immunosuppressants | sirolimus |
| 8104010 | MA11 | Immunosuppressants | sirolimus |
| 68084091525 | MA11 | Immunosuppressants | sirolimus |
| 68084095695 | MA11 | Immunosuppressants | sirolimus |
| 8103001 | MA11 | Immunosuppressants | sirolimus |
| 59762100101 | MA11 | Immunosuppressants | sirolimus |
| 50268071811 | MA11 | Immunosuppressants | sirolimus |
| 8104110 | MA11 | Immunosuppressants | sirolimus |
| 68084095625 | MA11 | Immunosuppressants | sirolimus |
| 68382052001 | MA11 | Immunosuppressants | sirolimus |
| 8103105 | MA11 | Immunosuppressants | sirolimus |
| 8103014 | MA11 | Immunosuppressants | sirolimus |
| 59762120506 | MA11 | Immunosuppressants | sirolimus |
| 8103015 | MA11 | Immunosuppressants | sirolimus |
| 66689034702 | MA11 | Immunosuppressants | sirolimus |
| 35356028000 | MA11 | Immunosuppressants | sirolimus |
| 69238159406 | MA11 | Immunosuppressants | sirolimus |
| 8103205 | MA11 | Immunosuppressants | sirolimus |
| 8103003 | MA11 | Immunosuppressants | sirolimus |
| 59762100201 | MA11 | Immunosuppressants | sirolimus |
| 8103004 | MA11 | Immunosuppressants | sirolimus |
| 59762120504 | MA11 | Immunosuppressants | sirolimus |
| 50268071813 | MA11 | Immunosuppressants | sirolimus |

| | | | |
|---|---|---|---|
| 55111065301 | MA11 | Immunosuppressants | sirolimus |
| 59762100301 | MA11 | Immunosuppressants | sirolimus |
| 8104205 | MA11 | Immunosuppressants | sirolimus |
| 55111052601 | MA11 | Immunosuppressants | tacrolimus |
| 68992304003 | MA11 | Immunosuppressants | tacrolimus |
| 16729004101 | MA11 | Immunosuppressants | tacrolimus |
| 55111052701 | MA11 | Immunosuppressants | tacrolimus |
| 781210401 | MA11 | Immunosuppressants | tacrolimus |
| 16729004301 | MA11 | Immunosuppressants | tacrolimus |
| 51079002820 | MA11 | Immunosuppressants | tacrolimus |
| 68992301003 | MA11 | Immunosuppressants | tacrolimus |
| 68084045001 | MA11 | Immunosuppressants | tacrolimus |
| 51079081820 | MA11 | Immunosuppressants | tacrolimus |
| 67544120553 | MA11 | Immunosuppressants | tacrolimus |
| 68992301001 | MA11 | Immunosuppressants | tacrolimus |
| 62175038237 | MA11 | Immunosuppressants | tacrolimus |
| 43353031709 | MA11 | Immunosuppressants | tacrolimus |
| 55111052501 | MA11 | Immunosuppressants | tacrolimus |
| 469061773 | MA11 | Immunosuppressants | tacrolimus |
| 64380072106 | MA11 | Immunosuppressants | tacrolimus |
| 591335901 | MA11 | Immunosuppressants | tacrolimus |
| 378204701 | MA11 | Immunosuppressants | tacrolimus |
| 469068773 | MA11 | Immunosuppressants | tacrolimus |
| 43353017809 | MA11 | Immunosuppressants | tacrolimus |
| 43353031753 | MA11 | Immunosuppressants | tacrolimus |
| 68258909901 | MA11 | Immunosuppressants | tacrolimus |
| 469065771 | MA11 | Immunosuppressants | tacrolimus |
| 469061710 | MA11 | Immunosuppressants | tacrolimus |
| 469065711 | MA11 | Immunosuppressants | tacrolimus |
| 469133050 | MA11 | Immunosuppressants | tacrolimus |
| 78061805 | MA11 | Immunosuppressants | tacrolimus |
| 60429037801 | MA11 | Immunosuppressants | tacrolimus |
| 51079081720 | MA11 | Immunosuppressants | tacrolimus |
| 16729004201 | MA11 | Immunosuppressants | tacrolimus |
| 781930401 | MA11 | Immunosuppressants | tacrolimus |
| 67544120580 | MA11 | Immunosuppressants | tacrolimus |
| 50742020701 | MA11 | Immunosuppressants | tacrolimus |
| 469064773 | MA11 | Immunosuppressants | tacrolimus |
| 51079081801 | MA11 | Immunosuppressants | tacrolimus |
| 68992304001 | MA11 | Immunosuppressants | tacrolimus |
| 62175038037 | MA11 | Immunosuppressants | tacrolimus |
| 469061711 | MA11 | Immunosuppressants | tacrolimus |
| 469060767 | MA11 | Immunosuppressants | tacrolimus |
| 51079002801 | MA11 | Immunosuppressants | tacrolimus |
| 62175038137 | MA11 | Immunosuppressants | tacrolimus |
| 469301601 | MA11 | Immunosuppressants | tacrolimus |
| 378204705 | MA11 | Immunosuppressants | tacrolimus |
| 469065773 | MA11 | Immunosuppressants | tacrolimus |
| 64380072206 | MA11 | Immunosuppressants | tacrolimus |
| 469123050 | MA11 | Immunosuppressants | tacrolimus |
| 469065710 | MA11 | Immunosuppressants | tacrolimus |
| 69452015520 | MA11 | Immunosuppressants | tacrolimus |
| 43353031716 | MA11 | Immunosuppressants | tacrolimus |
| 904662361 | MA11 | Immunosuppressants | tacrolimus |
| 378204505 | MA11 | Immunosuppressants | tacrolimus |
| 69452015420 | MA11 | Immunosuppressants | tacrolimus |
| 68992307501 | MA11 | Immunosuppressants | tacrolimus |
| 68084044901 | MA11 | Immunosuppressants | tacrolimus |
| 68084045101 | MA11 | Immunosuppressants | tacrolimus |
| 60429037901 | MA11 | Immunosuppressants | tacrolimus |
| 68084045111 | MA11 | Immunosuppressants | tacrolimus |
| 50742020901 | MA11 | Immunosuppressants | tacrolimus |
| 43353017860 | MA11 | Immunosuppressants | tacrolimus |
| 378204501 | MA11 | Immunosuppressants | tacrolimus |
| 67263039601 | MA11 | Immunosuppressants | tacrolimus |
| 378204601 | MA11 | Immunosuppressants | tacrolimus |
| 904642561 | MA11 | Immunosuppressants | tacrolimus |
| 78061605 | MA11 | Immunosuppressants | tacrolimus |
| 469067773 | MA11 | Immunosuppressants | tacrolimus |
| 68084044911 | MA11 | Immunosuppressants | tacrolimus |
| 50090224500 | MA11 | Immunosuppressants | tacrolimus |

| | | | |
|---|---|---|---|
| 469060773 | MA11 | Immunosuppressants | tacrolimus |
| 904662461 | MA11 | Immunosuppressants | tacrolimus |
| 781210201 | MA11 | Immunosuppressants | tacrolimus |
| 43353017853 | MA11 | Immunosuppressants | tacrolimus |
| 64380072006 | MA11 | Immunosuppressants | tacrolimus |
| 43353017880 | MA11 | Immunosuppressants | tacrolimus |
| 43353031770 | MA11 | Immunosuppressants | tacrolimus |
| 378204605 | MA11 | Immunosuppressants | tacrolimus |
| 68084045011 | MA11 | Immunosuppressants | tacrolimus |
| 781930201 | MA11 | Immunosuppressants | tacrolimus |
| 469061771 | MA11 | Immunosuppressants | tacrolimus |
| 78061705 | MA11 | Immunosuppressants | tacrolimus |
| 50742020801 | MA11 | Immunosuppressants | tacrolimus |
| 43353031780 | MA11 | Immunosuppressants | tacrolimus |
| 8117901 | MA11 | Immunosuppressants | temsirolimus |
| 16729022361 | MA11 | Immunosuppressants | temsirolimus |
| 8117905 | MA11 | Immunosuppressants | temsirolimus |
| 16729022130 | MA11 | Immunosuppressants | temsirolimus |
| 1012899 | MA00 | Immunosuppressants | fingolimod 0.5 MG |
| 1193337 | MA00 | Immunosuppressants | ruxolitinib 10 MG |
| 1193341 | MA00 | Immunosuppressants | ruxolitinib 5 MG |
| 1193345 | MA00 | Immunosuppressants | ruxolitinib 15 MG |
| 1193349 | MA00 | Immunosuppressants | ruxolitinib 20 MG |
| 1193353 | MA00 | Immunosuppressants | ruxolitinib 25 MG |
| 1442698 | MA00 | Immunosuppressants | obinutuzumab 25 MG/ML |
| 1654079 | MA00 | Immunosuppressants | ustekinumab 90 MG/ML |
| 1720606 | MA00 | Immunosuppressants | mepolizumab 100 MG |
| 1745108 | MA00 | Immunosuppressants | ixekizumab 80 MG/ML |
| 1745114 | MA00 | Immunosuppressants | ixekizumab 80 MG/ML |
| 1811257 | MA00 | Immunosuppressants | ustekinumab 5 MG/ML |
| 1872261 | MA00 | Immunosuppressants | brodalumab 140 MG/ML |
| 1876406 | MA00 | Immunosuppressants | dupilumab 150 MG/ML |
| 1923338 | MA00 | Immunosuppressants | sarilumab 132 MG/ML |
| 1923347 | MA00 | Immunosuppressants | sarilumab 175 MG/ML |
| 1928689 | MA00 | Immunosuppressants | guselkumab 100 MG/ML |
| 1942959 | MA00 | Immunosuppressants | inotuzumab ozogamicin 0.9 MG |
| 1989109 | MA00 | Immunosuppressants | benralizumab 30 MG/ML |
| 2003757 | MA00 | Immunosuppressants | sarilumab 132 MG/ML |
| 2003767 | MA00 | Immunosuppressants | sarilumab 175 MG/ML |
| 2045493 | MA00 | Immunosuppressants | fingolimod 0.25 MG |
| 2055026 | MA00 | Immunosuppressants | Cyclosporine 0.9 MG/ML |
| 2099302 | MA00 | Immunosuppressants | dupilumab 175 MG/ML |
| 2118741 | MA00 | Immunosuppressants | guselkumab 100 MG/ML |
| 2170993 | MA00 | Immunosuppressants | mepolizumab 100 MG/ML |
| 2173822 | MA00 | Immunosuppressants | mepolizumab 100 MG/ML |
| 2196900 | MA00 | Immunosuppressants | upadacitinib 15 MG |
| 311881 | MA00 | Immunosuppressants | mycophenolate mofetil 500 MG |
| 313190 | MA00 | Immunosuppressants | Tacrolimus 0.5 MG |
| 402092 | MA00 | Immunosuppressants | Cyclosporine 0.5 MG/ML |
| 402240 | MA00 | Immunosuppressants | efalizumab 100 MG/ML |
| 603541 | MA00 | Immunosuppressants | natalizumab 20 MG/ML |
| 795724 | MA00 | Immunosuppressants | Methohexital 50 MG/ML |
| 853354 | MA00 | Immunosuppressants | ustekinumab 90 MG/ML |
| 853356 | MA00 | Immunosuppressants | ustekinumab 90 MG/ML |
| 865174 | MA00 | Immunosuppressants | ustekinumab 90 MG/ML |
| 1012891 | MA00 | Immunosuppressants | fingolimod hydrochloride |
| 1012892 | MA00 | Immunosuppressants | fingolimod |
| 1012893 | MA00 | Immunosuppressants | fingolimod 0.5 MG |
| 1012894 | MA00 | Immunosuppressants | fingolimod Oral Capsule |
| 1012895 | MA00 | Immunosuppressants | fingolimod 0.5 MG |
| 1012897 | MA00 | Immunosuppressants | fingolimod 0.5 MG |
| 1012898 | MA00 | Immunosuppressants | fingolimod Oral Capsule |
| 1158395 | MA00 | Immunosuppressants | natalizumab Injectable Product |
| 1161283 | MA00 | Immunosuppressants | efalizumab Injectable Product |
| 1163983 | MA00 | Immunosuppressants | ustekinumab Injectable Product |
| 1165017 | MA00 | Immunosuppressants | fingolimod Oral Product |
| 1165018 | MA00 | Immunosuppressants | fingolimod Pill |
| 1193325 | MA00 | Immunosuppressants | ruxolitinib phosphate |
| 1193326 | MA00 | Immunosuppressants | ruxolitinib |
| 1193327 | MA00 | Immunosuppressants | ruxolitinib 10 MG |
| 1193328 | MA00 | Immunosuppressants | ruxolitinib Oral Product |

| | | | |
|---|---|---|---|
| 1193329 | MA00 | Immunosuppressants | ruxolitinib Pill |
| 1193330 | MA00 | Immunosuppressants | ruxolitinib Oral Tablet |
| 1193331 | MA00 | Immunosuppressants | ruxolitinib 10 MG |
| 1193333 | MA00 | Immunosuppressants | ruxolitinib 10 MG |
| 1193334 | MA00 | Immunosuppressants | ruxolitinib Oral Tablet |
| 1193338 | MA00 | Immunosuppressants | ruxolitinib 5 MG |
| 1193339 | MA00 | Immunosuppressants | ruxolitinib 5 MG |
| 1193340 | MA00 | Immunosuppressants | ruxolitinib 5 MG |
| 1193342 | MA00 | Immunosuppressants | ruxolitinib 15 MG |
| 1193343 | MA00 | Immunosuppressants | ruxolitinib 15 MG |
| 1193344 | MA00 | Immunosuppressants | ruxolitinib 15 MG |
| 1193346 | MA00 | Immunosuppressants | ruxolitinib 20 MG |
| 1193347 | MA00 | Immunosuppressants | ruxolitinib 20 MG |
| 1193348 | MA00 | Immunosuppressants | ruxolitinib 20 MG |
| 1193350 | MA00 | Immunosuppressants | ruxolitinib 25 MG |
| 1193351 | MA00 | Immunosuppressants | ruxolitinib 25 MG |
| 1193352 | MA00 | Immunosuppressants | ruxolitinib 25 MG |
| 1359534 | MA00 | Immunosuppressants | ustekinumab 90 MG/ML |
| 1359943 | MA00 | Immunosuppressants | ustekinumab 90 MG/ML |
| 1442690 | MA00 | Immunosuppressants | obinutuzumab 25 MG/ML |
| 1442691 | MA00 | Immunosuppressants | obinutuzumab Injectable Product |
| 1442692 | MA00 | Immunosuppressants | obinutuzumab Injectable Solution |
| 1442693 | MA00 | Immunosuppressants | obinutuzumab 25 MG/ML |
| 1442695 | MA00 | Immunosuppressants | obinutuzumab 25 MG/ML |
| 1442696 | MA00 | Immunosuppressants | obinutuzumab Injectable Solution |
| 1654076 | MA00 | Immunosuppressants | ustekinumab Injection |
| 1654077 | MA00 | Immunosuppressants | ustekinumab 90 MG/ML |
| 1654078 | MA00 | Immunosuppressants | ustekinumab Injection |
| 1654080 | MA00 | Immunosuppressants | ustekinumab 90 MG/ML |
| 1654081 | MA00 | Immunosuppressants | ustekinumab 90 MG/ML |
| 1654082 | MA00 | Immunosuppressants | ustekinumab 90 MG/ML |
| 1654083 | MA00 | Immunosuppressants | ustekinumab 90 MG/ML |
| 1656270 | MA00 | Immunosuppressants | natalizumab Injection |
| 1656272 | MA00 | Immunosuppressants | natalizumab Injection |
| 1656274 | MA00 | Immunosuppressants | natalizumab 20 MG/ML |
| 1656275 | MA00 | Immunosuppressants | natalizumab 20 MG/ML |
| 1656294 | MA00 | Immunosuppressants | obinutuzumab Injection |
| 1656296 | MA00 | Immunosuppressants | obinutuzumab Injection |
| 1656298 | MA00 | Immunosuppressants | obinutuzumab 25 MG/ML |
| 1656299 | MA00 | Immunosuppressants | obinutuzumab 25 MG/ML |
| 1720597 | MA00 | Immunosuppressants | mepolizumab |
| 1720598 | MA00 | Immunosuppressants | mepolizumab 100 MG |
| 1720599 | MA00 | Immunosuppressants | mepolizumab Injectable Product |
| 1720600 | MA00 | Immunosuppressants | mepolizumab Injection |
| 1720601 | MA00 | Immunosuppressants | mepolizumab 100 MG |
| 1720603 | MA00 | Immunosuppressants | mepolizumab 100 MG |
| 1720604 | MA00 | Immunosuppressants | mepolizumab Injection |
| 1745099 | MA00 | Immunosuppressants | ixekizumab |
| 1745100 | MA00 | Immunosuppressants | ixekizumab 80 MG/ML |
| 1745101 | MA00 | Immunosuppressants | ixekizumab Injectable Product |
| 1745102 | MA00 | Immunosuppressants | ixekizumab Auto-Injector |
| 1745103 | MA00 | Immunosuppressants | ixekizumab 80 MG/ML |
| 1745105 | MA00 | Immunosuppressants | ixekizumab 80 MG/ML |
| 1745106 | MA00 | Immunosuppressants | ixekizumab Auto-Injector |
| 1745109 | MA00 | Immunosuppressants | ixekizumab 80 MG/ML |
| 1745110 | MA00 | Immunosuppressants | ixekizumab 80 MG/ML |
| 1745111 | MA00 | Immunosuppressants | ixekizumab Prefilled Syringe |
| 1745112 | MA00 | Immunosuppressants | ixekizumab 80 MG/ML |
| 1745113 | MA00 | Immunosuppressants | ixekizumab Prefilled Syringe |
| 1745115 | MA00 | Immunosuppressants | ixekizumab 80 MG/ML |
| 1745116 | MA00 | Immunosuppressants | ixekizumab 80 MG/ML |
| 1811254 | MA00 | Immunosuppressants | ustekinumab 5 MG/ML |
| 1811255 | MA00 | Immunosuppressants | ustekinumab 5 MG/ML |
| 1811256 | MA00 | Immunosuppressants | ustekinumab 5 MG/ML |
| 1811258 | MA00 | Immunosuppressants | ustekinumab 5 MG/ML |
| 1811259 | MA00 | Immunosuppressants | ustekinumab 5 MG/ML |
| 1872251 | MA00 | Immunosuppressants | brodalumab |
| 1872253 | MA00 | Immunosuppressants | brodalumab 140 MG/ML |
| 1872254 | MA00 | Immunosuppressants | brodalumab Injectable Product |
| 1872255 | MA00 | Immunosuppressants | brodalumab Prefilled Syringe |
| 1872256 | MA00 | Immunosuppressants | brodalumab 140 MG/ML |

| | | | |
|---|---|---|---|
| 1872258 | MA00 | Immunosuppressants | brodalumab 140 MG/ML |
| 1872259 | MA00 | Immunosuppressants | brodalumab Prefilled Syringe |
| 1872262 | MA00 | Immunosuppressants | brodalumab 140 MG/ML |
| 1872263 | MA00 | Immunosuppressants | brodalumab 140 MG/ML |
| 1876376 | MA00 | Immunosuppressants | dupilumab |
| 1876398 | MA00 | Immunosuppressants | dupilumab 150 MG/ML |
| 1876399 | MA00 | Immunosuppressants | dupilumab Injectable Product |
| 1876400 | MA00 | Immunosuppressants | dupilumab Prefilled Syringe |
| 1876401 | MA00 | Immunosuppressants | dupilumab 150 MG/ML |
| 1876403 | MA00 | Immunosuppressants | dupilumab 150 MG/ML |
| 1876404 | MA00 | Immunosuppressants | dupilumab Prefilled Syringe |
| 1876407 | MA00 | Immunosuppressants | dupilumab 150 MG/ML |
| 1876408 | MA00 | Immunosuppressants | dupilumab 150 MG/ML |
| 1923319 | MA00 | Immunosuppressants | sarilumab |
| 1923330 | MA00 | Immunosuppressants | sarilumab 132 MG/ML |
| 1923331 | MA00 | Immunosuppressants | sarilumab Injectable Product |
| 1923332 | MA00 | Immunosuppressants | sarilumab Prefilled Syringe |
| 1923333 | MA00 | Immunosuppressants | sarilumab 132 MG/ML |
| 1923335 | MA00 | Immunosuppressants | sarilumab 132 MG/ML |
| 1923336 | MA00 | Immunosuppressants | sarilumab Prefilled Syringe |
| 1923339 | MA00 | Immunosuppressants | sarilumab 132 MG/ML |
| 1923340 | MA00 | Immunosuppressants | sarilumab 132 MG/ML |
| 1923344 | MA00 | Immunosuppressants | sarilumab 175 MG/ML |
| 1923345 | MA00 | Immunosuppressants | sarilumab 175 MG/ML |
| 1923346 | MA00 | Immunosuppressants | sarilumab 175 MG/ML |
| 1923348 | MA00 | Immunosuppressants | sarilumab 175 MG/ML |
| 1923349 | MA00 | Immunosuppressants | sarilumab 175 MG/ML |
| 1928588 | MA00 | Immunosuppressants | guselkumab |
| 1928589 | MA00 | Immunosuppressants | guselkumab 100 MG/ML |
| 1928590 | MA00 | Immunosuppressants | guselkumab Injectable Product |
| 1928594 | MA00 | Immunosuppressants | guselkumab 100 MG/ML |
| 1928685 | MA00 | Immunosuppressants | guselkumab Prefilled Syringe |
| 1928686 | MA00 | Immunosuppressants | guselkumab 100 MG/ML |
| 1928687 | MA00 | Immunosuppressants | guselkumab Prefilled Syringe |
| 1928690 | MA00 | Immunosuppressants | guselkumab 100 MG/ML |
| 1928691 | MA00 | Immunosuppressants | guselkumab 100 MG/ML |
| 1942950 | MA00 | Immunosuppressants | inotuzumab ozogamicin |
| 1942951 | MA00 | Immunosuppressants | inotuzumab ozogamicin 0.9 MG |
| 1942952 | MA00 | Immunosuppressants | inotuzumab ozogamicin Injectable Product |
| 1942953 | MA00 | Immunosuppressants | inotuzumab ozogamicin Injection |
| 1942954 | MA00 | Immunosuppressants | inotuzumab ozogamicin 0.9 MG |
| 1942956 | MA00 | Immunosuppressants | inotuzumab ozogamicin 0.9 MG |
| 1942957 | MA00 | Immunosuppressants | inotuzumab ozogamicin Injection |
| 1989100 | MA00 | Immunosuppressants | benralizumab |
| 1989101 | MA00 | Immunosuppressants | benralizumab 30 MG/ML |
| 1989102 | MA00 | Immunosuppressants | benralizumab Injectable Product |
| 1989103 | MA00 | Immunosuppressants | benralizumab Prefilled Syringe |
| 1989104 | MA00 | Immunosuppressants | benralizumab 30 MG/ML |
| 1989106 | MA00 | Immunosuppressants | benralizumab 30 MG/ML |
| 1989107 | MA00 | Immunosuppressants | benralizumab Prefilled Syringe |
| 1989110 | MA00 | Immunosuppressants | benralizumab 30 MG/ML |
| 1989111 | MA00 | Immunosuppressants | benralizumab 30 MG/ML |
| 2003754 | MA00 | Immunosuppressants | sarilumab Auto-Injector |
| 2003755 | MA00 | Immunosuppressants | sarilumab 132 MG/ML |
| 2003756 | MA00 | Immunosuppressants | sarilumab Auto-Injector |
| 2003758 | MA00 | Immunosuppressants | sarilumab 132 MG/ML |
| 2003759 | MA00 | Immunosuppressants | sarilumab 132 MG/ML |
| 2003766 | MA00 | Immunosuppressants | sarilumab 175 MG/ML |
| 2003768 | MA00 | Immunosuppressants | sarilumab 175 MG/ML |
| 2003769 | MA00 | Immunosuppressants | sarilumab 175 MG/ML |
| 2045490 | MA00 | Immunosuppressants | fingolimod 0.25 MG |
| 2045491 | MA00 | Immunosuppressants | fingolimod 0.25 MG |
| 2045492 | MA00 | Immunosuppressants | fingolimod 0.25 MG |
| 2099299 | MA00 | Immunosuppressants | dupilumab 200 MG/ML |
| 2099300 | MA00 | Immunosuppressants | dupilumab 175 MG/ML |
| 2099301 | MA00 | Immunosuppressants | dupilumab 200 MG/ML |
| 2099303 | MA00 | Immunosuppressants | dupilumab 200 MG/ML |
| 2099304 | MA00 | Immunosuppressants | dupilumab 200 MG/ML |
| 2101124 | MA00 | Immunosuppressants | dupilumab 175 MG/ML |
| 2101126 | MA00 | Immunosuppressants | dupilumab 175 MG/ML |
| 2101128 | MA00 | Immunosuppressants | dupilumab 175 MG/ML |

| | | | |
|---|---|---|---|
| 2101129 | MA00 | Immunosuppressants | dupilumab 175 MG/ML |
| 2118738 | MA00 | Immunosuppressants | guselkumab Auto-Injector |
| 2118739 | MA00 | Immunosuppressants | guselkumab 100 MG/ML |
| 2118740 | MA00 | Immunosuppressants | guselkumab Auto-Injector |
| 2118742 | MA00 | Immunosuppressants | guselkumab 100 MG/ML |
| 2118743 | MA00 | Immunosuppressants | guselkumab 100 MG/ML |
| 2170988 | MA00 | Immunosuppressants | mepolizumab 100 MG/ML |
| 2170989 | MA00 | Immunosuppressants | mepolizumab Auto-Injector |
| 2170990 | MA00 | Immunosuppressants | mepolizumab 100 MG/ML |
| 2170991 | MA00 | Immunosuppressants | mepolizumab 100 MG/ML |
| 2170992 | MA00 | Immunosuppressants | mepolizumab Auto-Injector |
| 2170994 | MA00 | Immunosuppressants | mepolizumab 100 MG/ML |
| 2170995 | MA00 | Immunosuppressants | mepolizumab 100 MG/ML |
| 2173819 | MA00 | Immunosuppressants | mepolizumab Prefilled Syringe |
| 2173820 | MA00 | Immunosuppressants | mepolizumab 100 MG/ML |
| 2173821 | MA00 | Immunosuppressants | mepolizumab Prefilled Syringe |
| 2173823 | MA00 | Immunosuppressants | mepolizumab 100 MG/ML |
| 2173824 | MA00 | Immunosuppressants | mepolizumab 100 MG/ML |
| 2196092 | MA00 | Immunosuppressants | upadacitinib |
| 2196890 | MA00 | Immunosuppressants | upadacitinib 15 MG |
| 2196891 | MA00 | Immunosuppressants | upadacitinib Oral Product |
| 2196892 | MA00 | Immunosuppressants | upadacitinib Pill |
| 2196893 | MA00 | Immunosuppressants | upadacitinib Extended Release Oral Tablet |
| 2196894 | MA00 | Immunosuppressants | upadacitinib 15 MG |
| 2196896 | MA00 | Immunosuppressants | upadacitinib 15 MG |
| 2196897 | MA00 | Immunosuppressants | upadacitinib Extended Release Oral Tablet |
| 2196901 | MA00 | Immunosuppressants | upadacitinib 15 MG |
| 2196902 | MA00 | Immunosuppressants | upadacitinib 15 MG |
| 262323 | MA00 | Immunosuppressants | ibritumomab tiuxetan |
| 354770 | MA00 | Immunosuppressants | natalizumab |
| 356988 | MA00 | Immunosuppressants | efalizumab |
| 402239 | MA00 | Immunosuppressants | efalizumab 100 MG/ML |
| 402350 | MA00 | Immunosuppressants | efalizumab 100 MG/ML |
| 402351 | MA00 | Immunosuppressants | efalizumab Injectable Solution |
| 402369 | MA00 | Immunosuppressants | efalizumab Injectable Solution |
| 477484 | MA00 | Immunosuppressants | natalizumab 20 MG/ML |
| 477485 | MA00 | Immunosuppressants | natalizumab 20 MG/ML |
| 477487 | MA00 | Immunosuppressants | natalizumab Injectable Solution |
| 576113 | MA00 | Immunosuppressants | efalizumab 100 MG/ML |
| 603539 | MA00 | Immunosuppressants | natalizumab 20 MG/ML |
| 603540 | MA00 | Immunosuppressants | natalizumab Injectable Solution |
| 847083 | MA00 | Immunosuppressants | ustekinumab |
| 853348 | MA00 | Immunosuppressants | ustekinumab 90 MG/ML |
| 853349 | MA00 | Immunosuppressants | ustekinumab Prefilled Syringe |
| 853350 | MA00 | Immunosuppressants | ustekinumab 90 MG/ML |
| 853352 | MA00 | Immunosuppressants | ustekinumab 90 MG/ML |
| 853353 | MA00 | Immunosuppressants | ustekinumab Prefilled Syringe |
| 853355 | MA00 | Immunosuppressants | ustekinumab 90 MG/ML |
| 865171 | MA00 | Immunosuppressants | ustekinumab Injectable Solution |
| 865172 | MA00 | Immunosuppressants | ustekinumab 90 MG/ML |
| 865173 | MA00 | Immunosuppressants | ustekinumab Injectable Solution |
| 974779 | MA00 | Immunosuppressants | obinutuzumab |
| 1170524 | MA00 | Immunosuppressants | Gilenya Oral Product |
| 1170525 | MA00 | Immunosuppressants | Gilenya Pill |
| 1178283 | MA00 | Immunosuppressants | Raptiva Injectable Product |
| 1178973 | MA00 | Immunosuppressants | Stelara Injectable Product |
| 1187514 | MA00 | Immunosuppressants | Tysabri Injectable Product |
| 1193335 | MA00 | Immunosuppressants | Jakafi Oral Product |
| 1193336 | MA00 | Immunosuppressants | Jakafi Pill |
| 1366322 | MA00 | Immunosuppressants | Ceratonia siliqua whole extract |
| 1442697 | MA00 | Immunosuppressants | Gazyva Injectable Product |
| 1720605 | MA00 | Immunosuppressants | Nucala Injectable Product |
| 1745107 | MA00 | Immunosuppressants | Taltz Injectable Product |
| 1872260 | MA00 | Immunosuppressants | Siliq Injectable Product |
| 1876405 | MA00 | Immunosuppressants | Dupixent Injectable Product |
| 1923337 | MA00 | Immunosuppressants | Kevzara Injectable Product |
| 1928596 | MA00 | Immunosuppressants | Tremfya Injectable Product |
| 1942958 | MA00 | Immunosuppressants | Besponsa Injectable Product |
| 1989108 | MA00 | Immunosuppressants | Fasenra Injectable Product |
| 2047245 | MA00 | Immunosuppressants | Olumiant Oral Product |
| 2047246 | MA00 | Immunosuppressants | Olumiant Pill |

| | | | |
|---|---|---|---|
| 2196898 | MA00 | Immunosuppressants | Rinvoq Oral Product |
| 2196899 | MA00 | Immunosuppressants | Rinvoq Pill |
| 72789012930 | MA11 | Immunosuppressants | azathioprine |
| 67877049301 | MA11 | Immunosuppressants | azathioprine |
| 66658051001 | MA11 | Immunosuppressants | emapalumab-lzsg |
| 66658050501 | MA11 | Immunosuppressants | emapalumab-lzsg |
| 58406005504 | MA11 | Immunosuppressants | etanercept |
| 378309732 | MA11 | Immunosuppressants | everolimus |
| 51991082299 | MA11 | Immunosuppressants | everolimus |
| 378309632 | MA11 | Immunosuppressants | everolimus |
| 54047221 | MA11 | Immunosuppressants | everolimus |
| 93776619 | MA11 | Immunosuppressants | everolimus |
| 93776719 | MA11 | Immunosuppressants | everolimus |
| 51991082399 | MA11 | Immunosuppressants | everolimus |
| 54047121 | MA11 | Immunosuppressants | everolimus |
| 378309885 | MA11 | Immunosuppressants | everolimus |
| 54048114 | MA11 | Immunosuppressants | everolimus |
| 51991082128 | MA11 | Immunosuppressants | everolimus |
| 93776819 | MA11 | Immunosuppressants | everolimus |
| 54048014 | MA11 | Immunosuppressants | everolimus |
| 54049714 | MA11 | Immunosuppressants | everolimus |
| 54047021 | MA11 | Immunosuppressants | everolimus |
| 78060789 | MA11 | Immunosuppressants | fingolimod HCl |
| 78096589 | MA11 | Immunosuppressants | fingolimod HCl |
| 50268047815 | MA11 | Immunosuppressants | leflunomide |
| 70748012906 | MA11 | Immunosuppressants | leflunomide |
| 50268047715 | MA11 | Immunosuppressants | leflunomide |
| 70748013006 | MA11 | Immunosuppressants | leflunomide |
| 16729048601 | MA11 | Immunosuppressants | methotrexate sodium |
| 42291050501 | MA11 | Immunosuppressants | methotrexate sodium |
| 59651018201 | MA11 | Immunosuppressants | methotrexate sodium |
| 143936701 | MA11 | Immunosuppressants | methotrexate sodium/PF |
| 66220081722 | MA11 | Immunosuppressants | methotrexate/PF |
| 66220081022 | MA11 | Immunosuppressants | methotrexate/PF |
| 66220080711 | MA11 | Immunosuppressants | methotrexate/PF |
| 66220081522 | MA11 | Immunosuppressants | methotrexate/PF |
| 66220081211 | MA11 | Immunosuppressants | methotrexate/PF |
| 66220082511 | MA11 | Immunosuppressants | methotrexate/PF |
| 66220082222 | MA11 | Immunosuppressants | methotrexate/PF |
| 66220082022 | MA11 | Immunosuppressants | methotrexate/PF |
| 60687049401 | MA11 | Immunosuppressants | mycophenolate mofetil |
| 904707861 | MA11 | Immunosuppressants | mycophenolate mofetil |
| 70748026201 | MA11 | Immunosuppressants | mycophenolate mofetil |
| 904707461 | MA11 | Immunosuppressants | mycophenolate mofetil |
| 71288080321 | MA11 | Immunosuppressants | mycophenolate mofetil HCl |
| 67457038600 | MA11 | Immunosuppressants | mycophenolate mofetil HCl |
| 17478095740 | MA11 | Immunosuppressants | mycophenolate mofetil HCl |
| 50268056011 | MA11 | Immunosuppressants | mycophenolate sodium |
| 50268055912 | MA11 | Immunosuppressants | mycophenolate sodium |
| 70748021816 | MA11 | Immunosuppressants | mycophenolate sodium |
| 70748021716 | MA11 | Immunosuppressants | mycophenolate sodium |
| 73604091404 | MA11 | Immunosuppressants | rilonacept |
| 24592201 | MA11 | Immunosuppressants | sarilumab |
| 24591001 | MA11 | Immunosuppressants | sarilumab |
| 24590801 | MA11 | Immunosuppressants | sarilumab |
| 24592001 | MA11 | Immunosuppressants | sarilumab |
| 68462068401 | MA11 | Immunosuppressants | sirolimus |
| 67877074601 | MA11 | Immunosuppressants | sirolimus |
| 68462068201 | MA11 | Immunosuppressants | sirolimus |
| 60505619702 | MA11 | Immunosuppressants | sirolimus |
| 68462068301 | MA11 | Immunosuppressants | sirolimus |
| 67877074701 | MA11 | Immunosuppressants | sirolimus |
| 67877074801 | MA11 | Immunosuppressants | sirolimus |
| 16714009801 | MA11 | Immunosuppressants | tacrolimus |
| 60429037701 | MA11 | Immunosuppressants | tacrolimus |
| 16714010001 | MA11 | Immunosuppressants | tacrolimus |
| 16714009901 | MA11 | Immunosuppressants | tacrolimus |
| 42291075401 | MA11 | Immunosuppressants | tacrolimus |
| 70748022001 | MA11 | Immunosuppressants | tacrolimus |
| 68462068601 | MA11 | Immunosuppressants | tacrolimus |
| 51079081701 | MA11 | Immunosuppressants | tacrolimus |

| | | | |
|---|---|---|---|
| 69452015320 | MA11 | Immunosuppressants | tacrolimus |
| 42291075301 | MA11 | Immunosuppressants | tacrolimus |
| 70748022101 | MA11 | Immunosuppressants | tacrolimus |
| 70377001511 | MA11 | Immunosuppressants | tacrolimus |
| 781210301 | MA11 | Immunosuppressants | tacrolimus |
| 68462068501 | MA11 | Immunosuppressants | tacrolimus |
| 67877028001 | MA11 | Immunosuppressants | tacrolimus |
| 904709761 | MA11 | Immunosuppressants | tacrolimus |
| 67877027801 | MA11 | Immunosuppressants | tacrolimus |
| 70377001411 | MA11 | Immunosuppressants | tacrolimus |
| 70377001611 | MA11 | Immunosuppressants | tacrolimus |
| 67877027901 | MA11 | Immunosuppressants | tacrolimus |
| 54288013501 | MA11 | Immunosuppressants | tacrolimus |
| 42291075201 | MA11 | Immunosuppressants | tacrolimus |
| 70748021901 | MA11 | Immunosuppressants | tacrolimus |
| 54288013201 | MA11 | Immunosuppressants | tacrolimus |
| 69102901 | MA11 | Immunosuppressants | tofacitinib citrate |
| 74230670 | MA11 | Immunosuppressants | upadacitinib |
| 51991082228 | MA11 | Immunosuppressants | everolimus |
| 378309832 | MA11 | Immunosuppressants | everolimus |
| 51991082199 | MA11 | Immunosuppressants | everolimus |
| 51991082328 | MA11 | Immunosuppressants | everolimus |
| 66220081011 | MA11 | Immunosuppressants | methotrexate/PF |
| 66220081511 | MA11 | Immunosuppressants | methotrexate/PF |
| 50242007001 | MA11 | Immunosuppressants | obinutuzumab |
| 78100768 | MA11 | Immunosuppressants | ofatumumab |
| 55513022401 | MA11 | Immunosuppressants | rituximab-arrx |
| 55513032601 | MA11 | Immunosuppressants | rituximab-arrx |
| 50881002060 | MA11 | Immunosuppressants | ruxolitinib phosphate |
| 50881000560 | MA11 | Immunosuppressants | ruxolitinib phosphate |
| 50881001060 | MA11 | Immunosuppressants | ruxolitinib phosphate |
| 50881002560 | MA11 | Immunosuppressants | ruxolitinib phosphate |
| 50881001560 | MA11 | Immunosuppressants | ruxolitinib phosphate |
| 72611078001 | MA11 | Immunosuppressants | temsirolimus |
| 72611078502 | MA11 | Immunosuppressants | temsirolimus |
| 65219020505 | MA11 | Immunosuppressants | temsirolimus |
| 65219020005 | MA11 | Immunosuppressants | temsirolimus |
| 57894005427 | MA11 | Immunosuppressants | ustekinumab |
| 57894006002 | MA11 | Immunosuppressants | ustekinumab |
| 57894006003 | MA11 | Immunosuppressants | ustekinumab |
| 57894006103 | MA11 | Immunosuppressants | ustekinumab |
| 57894006104 | MA11 | Immunosuppressants | ustekinumab |
| 57894006004 | MA11 | Immunosuppressants | ustekinumab |
| 57894005416 | MA11 | Immunosuppressants | ustekinumab |
| 74230630 | MA11 | Immunosuppressants | upadacitinib |
| 74231030 | MA11 | Immunosuppressants | upadacitinib |
| 64406000801 | MA11 | Immunosuppressants | natalizumab |
| 50242005801 | MA11 | Immunosuppressants | Raptiva |
| 50242005804 | MA11 | Immunosuppressants | Raptiva |
| 2144501 | MA11 | Immunosuppressants | ixekizumab |
| 2144509 | MA11 | Immunosuppressants | ixekizumab |
| 2144511 | MA11 | Immunosuppressants | ixekizumab |
| 2144527 | MA11 | Immunosuppressants | ixekizumab |
| 2772401 | MA11 | Immunosuppressants | ixekizumab |
| 2772409 | MA11 | Immunosuppressants | ixekizumab |
| 2772411 | MA11 | Immunosuppressants | ixekizumab |
| 2772427 | MA11 | Immunosuppressants | ixekizumab |
| 8010001 | MA11 | Immunosuppressants | inotuzumab ozogamicin |
| 24591100 | MA11 | Immunosuppressants | dupilumab |
| 24591102 | MA11 | Immunosuppressants | dupilumab |
| 24591400 | MA11 | Immunosuppressants | dupilumab |
| 24591401 | MA11 | Immunosuppressants | dupilumab |
| 24591402 | MA11 | Immunosuppressants | dupilumab |
| 24591500 | MA11 | Immunosuppressants | dupilumab |
| 24591501 | MA11 | Immunosuppressants | dupilumab |
| 24591502 | MA11 | Immunosuppressants | dupilumab |
| 24591520 | MA11 | Immunosuppressants | dupilumab |
| 24591800 | MA11 | Immunosuppressants | dupilumab |
| 24591801 | MA11 | Immunosuppressants | dupilumab |
| 24591802 | MA11 | Immunosuppressants | dupilumab |
| 24591900 | MA11 | Immunosuppressants | dupilumab |

| | | | |
|---|---|---|---|
| 24591901 | MA11 | Immunosuppressants | dupilumab |
| 24591902 | MA11 | Immunosuppressants | dupilumab |
| 24591920 | MA11 | Immunosuppressants | dupilumab |
| 78060715 | MA11 | Immunosuppressants | fingolimod HCl |
| 78060751 | MA11 | Immunosuppressants | fingolimod HCl |
| 173088101 | MA11 | Immunosuppressants | mepolizumab |
| 173089201 | MA11 | Immunosuppressants | mepolizumab |
| 173089242 | MA11 | Immunosuppressants | mepolizumab |
| 173089261 | MA11 | Immunosuppressants | mepolizumab |
| 187000400 | MA11 | Immunosuppressants | brodalumab |
| 187000402 | MA11 | Immunosuppressants | brodalumab |
| 310173030 | MA11 | Immunosuppressants | benralizumab |
| 310173085 | MA11 | Immunosuppressants | benralizumab |
| 310183030 | MA11 | Immunosuppressants | benralizumab |
| 310183085 | MA11 | Immunosuppressants | benralizumab |
| 50881000705 | MA11 | Immunosuppressants | ruxolitinib phosphate |
| 57894064001 | MA11 | Immunosuppressants | guselkumab |
| 57894064011 | MA11 | Immunosuppressants | guselkumab |
| 58468008001 | MA11 | Immunosuppressants | anti-thymocyte globulin,rabbit |
| 62053053425 | MA11 | Immunosuppressants | anti-thymocyte globulin,rabbit |
| 59572040200 | MA11 | Immunosuppressants | lenalidomide |
| 59572040228 | MA11 | Immunosuppressants | lenalidomide |
| 59572040500 | MA11 | Immunosuppressants | lenalidomide |
| 59572040528 | MA11 | Immunosuppressants | lenalidomide |
| 59572040530 | MA11 | Immunosuppressants | lenalidomide |
| 59572041000 | MA11 | Immunosuppressants | lenalidomide |
| 59572041028 | MA11 | Immunosuppressants | lenalidomide |
| 59572041030 | MA11 | Immunosuppressants | lenalidomide |
| 59572041500 | MA11 | Immunosuppressants | lenalidomide |
| 59572041521 | MA11 | Immunosuppressants | lenalidomide |
| 59572042000 | MA11 | Immunosuppressants | lenalidomide |
| 59572042021 | MA11 | Immunosuppressants | lenalidomide |
| 59572042500 | MA11 | Immunosuppressants | lenalidomide |
| 59572042521 | MA11 | Immunosuppressants | lenalidomide |
| 59572042525 | MA11 | Immunosuppressants | lenalidomide |
| 59572020514 | MA11 | Immunosuppressants | thalidomide |
| 59572020517 | MA11 | Immunosuppressants | thalidomide |
| 59572020594 | MA11 | Immunosuppressants | thalidomide |
| 59572020597 | MA11 | Immunosuppressants | thalidomide |
| 59572021015 | MA11 | Immunosuppressants | thalidomide |
| 59572021095 | MA11 | Immunosuppressants | thalidomide |
| 59572021513 | MA11 | Immunosuppressants | thalidomide |
| 59572021593 | MA11 | Immunosuppressants | thalidomide |
| 59572022016 | MA11 | Immunosuppressants | thalidomide |
| 59572022096 | MA11 | Immunosuppressants | thalidomide |
| 59572010592 | MA11 | Immunosuppressants | thalidomide |
| 1182522 | MA00 | Immunosuppressants | Revlimid Oral Product |
| 1182529 | MA00 | Immunosuppressants | Revlimid Pill |
| 1242232 | MA00 | Immunosuppressants | lenalidomide 2.5 MG |
| 1242233 | MA00 | Immunosuppressants | lenalidomide 2.5 MG |
| 1429366 | MA00 | Immunosuppressants | lenalidomide 20 MG |
| 1429367 | MA00 | Immunosuppressants | lenalidomide 20 MG |
| 337535 | MA00 | Immunosuppressants | (NA) |
| 616112 | MA00 | Immunosuppressants | lenalidomide 10 MG |
| 616113 | MA00 | Immunosuppressants | lenalidomide Oral Capsule |
| 616114 | MA00 | Immunosuppressants | lenalidomide 10 MG |
| 616115 | MA00 | Immunosuppressants | lenalidomide 5 MG |
| 616116 | MA00 | Immunosuppressants | lenalidomide 5 MG |
| 643713 | MA00 | Immunosuppressants | lenalidomide 15 MG |
| 643714 | MA00 | Immunosuppressants | lenalidomide 15 MG |
| 643721 | MA00 | Immunosuppressants | lenalidomide 25 MG |
| 643722 | MA00 | Immunosuppressants | lenalidomide 25 MG |
| 10432 | MA00 | Immunosuppressants | Thalidomide |
| 1164873 | MA00 | Immunosuppressants | Thalidomide Oral Product |
| 1164874 | MA00 | Immunosuppressants | Thalidomide Pill |
| 200390 | MA00 | Immunosuppressants | Thalidomide 50 MG |
| 213360 | MA00 | Immunosuppressants | Thalidomide 50 MG |
| 316784 | MA00 | Immunosuppressants | Thalidomide 50 MG |
| 360176 | MA00 | Immunosuppressants | Thalidomide 100 MG |
| 360177 | MA00 | Immunosuppressants | Thalidomide 200 MG |
| 360321 | MA00 | Immunosuppressants | Thalidomide 100 MG |

| | | | |
|---|---|---|---|
| 360322 | MA00 | Immunosuppressants | Thalidomide 200 MG |
| 366451 | MA00 | Immunosuppressants | Thalidomide Oral Capsule |
| 374072 | MA00 | Immunosuppressants | Thalidomide Oral Capsule |
| 404449 | MA00 | Immunosuppressants | Thalidomide 100 MG |
| 404450 | MA00 | Immunosuppressants | Thalidomide 200 MG |
| 432527 | MA00 | Immunosuppressants | Thalidomide 100 MG |
| 452059 | MA00 | Immunosuppressants | Thalidomide Oral Tablet |
| 573254 | MA00 | Immunosuppressants | Thalidomide 50 MG |
| 576406 | MA00 | Immunosuppressants | Thalidomide 100 MG |
| 576407 | MA00 | Immunosuppressants | Thalidomide 200 MG |
| 700415 | MA00 | Immunosuppressants | Thalidomide 150 MG |
| 700416 | MA00 | Immunosuppressants | Thalidomide 150 MG |
| 700417 | MA00 | Immunosuppressants | Thalidomide 150 MG |
| 700418 | MA00 | Immunosuppressants | Thalidomide 150 MG |
| 107050 | MA00 | Immunosuppressants | Rabbit anti-human T-lymphocyte globulin 25 MG |
| 1177887 | MA00 | Immunosuppressants | Thymoglobulin Injectable Product |
| 1870637 | MA00 | Immunosuppressants | Rabbit anti-human T-lymphocyte globulin 25 MG |
| 1870638 | MA00 | Immunosuppressants | Rabbit anti-human T-lymphocyte globulin Injection |
| 225741 | MA00 | Immunosuppressants | (NA) |
| 363507 | MA00 | Immunosuppressants | Rabbit anti-human T-lymphocyte globulin Injectable Solution |
| 564411 | MA00 | Immunosuppressants | Rabbit anti-human T-lymphocyte globulin 5 MG/ML |
| 8638 | MA00 | Corticosteriods | prednisolone |
| 6902 | MA00 | Corticosteriods | methylprednisolone |
| 1366551 | MA00 | Corticosteriods | UCERIS |
| 153171 | MA00 | Corticosteriods | ENTOCORT |
| 2273102 | MA00 | Corticosteriods | ORTIKOS |
| 19831 | MA00 | Corticosteriods | BUDESONIDE |
| 1367172 | MA00 | Corticosteriods | BUDESONIDE (11.BETA.,16.ALPHA.(S)) |
| 1244213 | MA00 | Corticosteriods | 24 HR BUDESONIDE 3 MG EXTENDED RELEASE ENTERIC COATED CAPSULE [ENTOCORT] |
| 1244215 | MA00 | Corticosteriods | BUDESONIDE 3 MG DELAYED RELEASE ORAL CAPSULE [ENTOCORT] |
| 1366556 | MA00 | Corticosteriods | 24 HR BUDESONIDE 9 MG EXTENDED RELEASE ORAL TABLET [UCERIS] |
| 1366558 | MA00 | Corticosteriods | BUDESONIDE 9 MG EXTENDED RELEASE ORAL TABLET [UCERIS] |
| 152595 | MA00 | Corticosteriods | 24 HR BUDESONIDE 3 MG EXTENDED RELEASE CAPSULE [ENTOCORT EC] |
| 2273107 | MA00 | Corticosteriods | 24 HR BUDESONIDE 6 MG EXTENDED RELEASE ORAL CAPSULE [ORTIKOS] |
| 2273108 | MA00 | Corticosteriods | BUDESONIDE 6 MG EXTENDED RELEASE ORAL CAPSULE [ORTIKOS] |
| 2273110 | MA00 | Corticosteriods | 24 HR BUDESONIDE 9 MG EXTENDED RELEASE ORAL CAPSULE [ORTIKOS] |
| 2273111 | MA00 | Corticosteriods | BUDESONIDE 9 MG EXTENDED RELEASE ORAL CAPSULE [ORTIKOS] |
| 844427 | MA00 | Corticosteriods | BUDESONIDE 3 MG EXTENDED RELEASE CAPSULE [ENTOCORT EC] |
| 991401 | MA00 | Corticosteriods | 24 HR BUDESONIDE 3 MG EXTENDED RELEASE CAPSULE [ENTOCORT] |
| 991402 | MA00 | Corticosteriods | BUDESONIDE 3 MG EXTENDED RELEASE CAPSULE [ENTOCORT] |
| 1366552 | MA00 | Corticosteriods | BUDESONIDE 9 MG [UCERIS] |
| 2273103 | MA00 | Corticosteriods | BUDESONIDE 6 MG [ORTIKOS] |
| 2273109 | MA00 | Corticosteriods | BUDESONIDE 9 MG [ORTIKOS] |
| 564993 | MA00 | Corticosteriods | BUDESONIDE 3 MG [ENTOCORT EC] |
| 991399 | MA00 | Corticosteriods | BUDESONIDE 3 MG [ENTOCORT] |
| 1244212 | MA00 | Corticosteriods | BUDESONIDE EXTENDED RELEASE ENTERIC COATED CAPSULE [ENTOCORT] |
| 1366553 | MA00 | Corticosteriods | BUDESONIDE EXTENDED RELEASE ORAL TABLET [UCERIS] |
| 1489772 | MA00 | Corticosteriods | BUDESONIDE DELAYED RELEASE ORAL CAPSULE [ENTOCORT] |
| 2273104 | MA00 | Corticosteriods | BUDESONIDE EXTENDED RELEASE ORAL CAPSULE [ORTIKOS] |
| 367142 | MA00 | Corticosteriods | BUDESONIDE 24 HOUR EXTENDED RELEASE CAPSULE [ENTOCORT EC] |
| 991400 | MA00 | Corticosteriods | BUDESONIDE EXTENDED RELEASE CAPSULE [ENTOCORT] |
| 1167031 | MA00 | Corticosteriods | ENTOCORT ORAL PRODUCT |
| 1167032 | MA00 | Corticosteriods | ENTOCORT PILL |
| 1366554 | MA00 | Corticosteriods | UCERIS ORAL PRODUCT |
| 1366555 | MA00 | Corticosteriods | UCERIS PILL |
| 2273105 | MA00 | Corticosteriods | ORTIKOS ORAL PRODUCT |
| 2273106 | MA00 | Corticosteriods | ORTIKOS PILL |
| 1006089 | MA00 | Corticosteriods | BUDESONIDE 3 MG ENTERIC COATED CAPSULE |
| 1244211 | MA00 | Corticosteriods | 24 HR BUDESONIDE 3 MG EXTENDED RELEASE ENTERIC COATED CAPSULE |
| 1244214 | MA00 | Corticosteriods | BUDESONIDE 3 MG DELAYED RELEASE ORAL CAPSULE |
| 1366550 | MA00 | Corticosteriods | 24 HR BUDESONIDE 9 MG EXTENDED RELEASE ORAL TABLET |
| 1366557 | MA00 | Corticosteriods | BUDESONIDE 9 MG EXTENDED RELEASE ORAL TABLET |
| 152594 | MA00 | Corticosteriods | 24 HR BUDESONIDE 3 MG EXTENDED RELEASE CAPSULE |
| 2048810 | MA00 | Corticosteriods | BUDESONIDE 3 MG ORAL CAPSULE |
| 2269305 | MA00 | Corticosteriods | 24 HR BUDESONIDE 6 MG EXTENDED RELEASE ORAL CAPSULE |
| 2269306 | MA00 | Corticosteriods | BUDESONIDE 6 MG EXTENDED RELEASE ORAL CAPSULE |
| 2269307 | MA00 | Corticosteriods | 24 HR BUDESONIDE 9 MG EXTENDED RELEASE ORAL CAPSULE |
| 2269308 | MA00 | Corticosteriods | BUDESONIDE 9 MG EXTENDED RELEASE ORAL CAPSULE |
| 249241 | MA00 | Corticosteriods | BUDESONIDE 3 MG ORAL TABLET |
| 387015 | MA00 | Corticosteriods | BUDESONIDE 3 MG ORAL CAPSULE |
| 484697 | MA00 | Corticosteriods | BUDESONIDE 3 MG EXTENDED RELEASE CAPSULE |

| | | | |
|---|---|---|---|
| 844426 | MA00 | Corticosteriods | BUDESONIDE 3 MG EXTENDED RELEASE CAPSULE |
| 1366548 | MA00 | Corticosteriods | BUDESONIDE 9 MG |
| 2269303 | MA00 | Corticosteriods | BUDESONIDE 6 MG |
| 332685 | MA00 | Corticosteriods | BUDESONIDE 3 MG |
| 1006088 | MA00 | Corticosteriods | BUDESONIDE ENTERIC COATED CAPSULE |
| 1244210 | MA00 | Corticosteriods | BUDESONIDE EXTENDED RELEASE ENTERIC COATED CAPSULE |
| 1366549 | MA00 | Corticosteriods | BUDESONIDE EXTENDED RELEASE ORAL TABLET |
| 1489770 | MA00 | Corticosteriods | BUDESONIDE DELAYED RELEASE ORAL CAPSULE |
| 2048809 | MA00 | Corticosteriods | BUDESONIDE ORAL CAPSULE |
| 2269304 | MA00 | Corticosteriods | BUDESONIDE EXTENDED RELEASE ORAL CAPSULE |
| 376186 | MA00 | Corticosteriods | BUDESONIDE ORAL TABLET |
| 379057 | MA00 | Corticosteriods | BUDESONIDE 24 HOUR EXTENDED RELEASE CAPSULE |
| 393140 | MA00 | Corticosteriods | BUDESONIDE ORAL CAPSULE |
| 484696 | MA00 | Corticosteriods | BUDESONIDE EXTENDED RELEASE CAPSULE |
| 1151338 | MA00 | Corticosteriods | BUDESONIDE ORAL PRODUCT |
| 1151339 | MA00 | Corticosteriods | BUDESONIDE PILL |
| 1100388 | MA00 | Corticosteriods | HYBRISIL |
| 203856 | MA00 | Corticosteriods | SOLU-MEDROL |
| 22584 | MA00 | Corticosteriods | DEPO-MEDROL |
| 1358512 | MA00 | Corticosteriods | METHYLPREDNISOLONE ACETATE 20 MG/ML INJECTABLE SUSPENSION [DEPO-MEDROL] |
| 1743722 | MA00 | Corticosteriods | METHYLPREDNISOLONE 500 MG INJECTION [SOLU-MEDROL] |
| 1743729 | MA00 | Corticosteriods | METHYLPREDNISOLONE 1000 MG INJECTION [SOLU-MEDROL] |
| 207137 | MA00 | Corticosteriods | METHYLPREDNISOLONE 8 MG ORAL TABLET [MEDROL] |
| 1165881 | MA00 | Corticosteriods | A-METHAPRED INJECTABLE PRODUCT |
| 1183797 | MA00 | Corticosteriods | MEDROL PILL |
| 105404 | MA00 | Corticosteriods | METHYLPREDNISOLONE 50 MG/ML INJECTABLE SOLUTION |
| 1357886 | MA00 | Corticosteriods | METHYLPREDNISOLONE 2000 MG INJECTION |
| 1743726 | MA00 | Corticosteriods | METHYLPREDNISOLONE 1000 MG INJECTION |
| 1743855 | MA00 | Corticosteriods | 1 ML METHYLPREDNISOLONE ACETATE 80 MG/ML INJECTION |
| 1012205 | MA00 | Corticosteriods | ASMALPRED PLUS |
| 1191678 | MA00 | Corticosteriods | PREDNISTAB |
| 1493166 | MA00 | Corticosteriods | SOLU-DELTA-CORTEF |
| 152241 | MA00 | Corticosteriods | PREDNESOL |
| 202702 | MA00 | Corticosteriods | MEDROL |
| 202760 | MA00 | Corticosteriods | HYDELTRASOL |
| 203857 | MA00 | Corticosteriods | A-METHAPRED |
| 216258 | MA00 | Corticosteriods | COTOLONE |
| 216464 | MA00 | Corticosteriods | DEPO-PREDATE |
| 216840 | MA00 | Corticosteriods | ECONOPRED |
| 216841 | MA00 | Corticosteriods | ECONOPRED PLUS |
| 217592 | MA00 | Corticosteriods | HYDELTRA-T.B.A. |
| 217898 | MA00 | Corticosteriods | KEY-PRED |
| 217899 | MA00 | Corticosteriods | KEY-PRED SP |
| 218285 | MA00 | Corticosteriods | MEDICORT |
| 219136 | MA00 | Corticosteriods | PEDIAPRED |
| 219282 | MA00 | Corticosteriods | POLY PRED |
| 219341 | MA00 | Corticosteriods | PRED-JECT-50 |
| 219342 | MA00 | Corticosteriods | PREDACORT 50 |
| 219344 | MA00 | Corticosteriods | PREDAJECT-50 |
| 219345 | MA00 | Corticosteriods | PREDALONE 50 |
| 219346 | MA00 | Corticosteriods | PREDATE-50 |
| 219347 | MA00 | Corticosteriods | PREDCOR |
| 219348 | MA00 | Corticosteriods | PREDICORT RP |
| 219349 | MA00 | Corticosteriods | PREDICORT-50 |
| 219356 | MA00 | Corticosteriods | PRELONE |
| 219403 | MA00 | Corticosteriods | PRI-CORTIN 50 |
| 285077 | MA00 | Corticosteriods | ORAPRED |
| 353627 | MA00 | Corticosteriods | CODELSOL |
| 353629 | MA00 | Corticosteriods | STINTISONE |
| 385325 | MA00 | Corticosteriods | CODELSON |
| 541702 | MA00 | Corticosteriods | PREDNORAL |
| 547219 | MA00 | Corticosteriods | PRICORTIN |
| 604201 | MA00 | Corticosteriods | BUBBLI-PRED |
| 792207 | MA00 | Corticosteriods | FLO-PRED |
| 795094 | MA00 | Corticosteriods | MILLIPRED |
| 824886 | MA00 | Corticosteriods | VERIPRED |
| 897060 | MA00 | Corticosteriods | TEMARIL-P |
| 1005831 | MA00 | Corticosteriods | {48 (PREDNISOLONE 5 MG ORAL TABLET [MILLIPRED]) } PACK [MILLIPRED DP 12 DAY] |
| 1235042 | MA00 | Corticosteriods | {21 (PREDNISOLONE 5 MG ORAL TABLET [MILLIPRED]) } PACK [MILLIPRED DP 6 DAY] |
| 834023 | MA00 | Corticosteriods | {21 (METHYLPREDNISOLONE 4 MG ORAL TABLET [MEDROL]) } PACK [MEDROL DOSEPAK] |
| 1005830 | MA00 | Corticosteriods | {48 (PREDNISOLONE 5 MG ORAL TABLET) } PACK |

| | | | |
|---|---|---|---|
| 1013114 | MA00 | Corticosteriods | {21 (PREDNISOLONE 5 MG ORAL TABLET) } PACK |
| 762675 | MA00 | Corticosteriods | {21 (METHYLPREDNISOLONE 4 MG ORAL TABLET) } PACK |
| 804173 | MA00 | Corticosteriods | {14 (METHYLPREDNISOLONE 16 MG ORAL TABLET) } PACK |
| 1358612 | MA00 | Corticosteriods | METHYLPREDNISOLONE ACETATE 40 MG/ML INJECTABLE SUSPENSION [DEPO-MEDROL] |
| 1358619 | MA00 | Corticosteriods | METHYLPREDNISOLONE ACETATE 80 MG/ML INJECTABLE SUSPENSION [DEPO-MEDROL] |
| 1743707 | MA00 | Corticosteriods | METHYLPREDNISOLONE 125 MG INJECTION [SOLU-MEDROL] |
| 1743781 | MA00 | Corticosteriods | 1 ML METHYLPREDNISOLONE ACETATE 40 MG/ML INJECTION [DEPO-MEDROL] |
| 1743856 | MA00 | Corticosteriods | 1 ML METHYLPREDNISOLONE ACETATE 80 MG/ML INJECTION [DEPO-MEDROL] |
| 207136 | MA00 | Corticosteriods | METHYLPREDNISOLONE 2 MG ORAL TABLET [MEDROL] |
| 207138 | MA00 | Corticosteriods | METHYLPREDNISOLONE 16 MG ORAL TABLET [MEDROL] |
| 207139 | MA00 | Corticosteriods | METHYLPREDNISOLONE 24 MG ORAL TABLET [MEDROL] |
| 207141 | MA00 | Corticosteriods | METHYLPREDNISOLONE 32 MG ORAL TABLET [MEDROL] |
| 207168 | MA00 | Corticosteriods | METHYLPREDNISOLONE 40 MG/ML INJECTABLE SUSPENSION [DEPO-MEDROL] |
| 207190 | MA00 | Corticosteriods | METHYLPREDNISOLONE 40 MG INJECTION [A-METHAPRED] |
| 207191 | MA00 | Corticosteriods | METHYLPREDNISOLONE 40 MG INJECTION [SOLU-MEDROL] |
| 207192 | MA00 | Corticosteriods | METHYLPREDNISOLONE 125 MG INJECTION [A-METHAPRED] |
| 207193 | MA00 | Corticosteriods | METHYLPREDNISOLONE 62.5 MG/ML INJECTABLE SOLUTION [SOLU-MEDROL] |
| 260330 | MA00 | Corticosteriods | METHYLPREDNISOLONE 4 MG ORAL TABLET [MEDROL] |
| 260409 | MA00 | Corticosteriods | PREDNISOLONE 1 MG/ML ORAL SOLUTION [PEDIAPRED] |
| 544779 | MA00 | Corticosteriods | METHYLPREDNISOLONE 125 MG/ML INJECTABLE SOLUTION [A-METHAPRED] |
| 647127 | MA00 | Corticosteriods | PREDNISOLONE 15 MG DISINTEGRATING ORAL TABLET [ORAPRED] |
| 668660 | MA00 | Corticosteriods | PREDNISOLONE 30 MG DISINTEGRATING ORAL TABLET [ORAPRED] |
| 808118 | MA00 | Corticosteriods | PREDNISOLONE 5 MG ORAL TABLET [MILLIPRED] |
| 1358611 | MA00 | Corticosteriods | METHYLPREDNISOLONE ACETATE 40 MG/ML [DEPO-MEDROL] |
| 567932 | MA00 | Corticosteriods | METHYLPREDNISOLONE 32 MG [MEDROL] |
| 567980 | MA00 | Corticosteriods | METHYLPREDNISOLONE 125 MG/ML [SOLU-MEDROL] |
| 567982 | MA00 | Corticosteriods | METHYLPREDNISOLONE 40 MG/ML [SOLU-MEDROL] |
| 567984 | MA00 | Corticosteriods | METHYLPREDNISOLONE 62.5 MG/ML [SOLU-MEDROL] |
| 574125 | MA00 | Corticosteriods | METHYLPREDNISOLONE 4 MG [MEDROL] |
| 368772 | MA00 | Corticosteriods | METHYLPREDNISOLONE ORAL TABLET [MEDROL] |
| 1168054 | MA00 | Corticosteriods | BUBBLI-PRED ORAL LIQUID PRODUCT |
| 1168055 | MA00 | Corticosteriods | BUBBLI-PRED ORAL PRODUCT |
| 1169771 | MA00 | Corticosteriods | FLO-PRED ORAL LIQUID PRODUCT |
| 1169772 | MA00 | Corticosteriods | FLO-PRED ORAL PRODUCT |
| 1170721 | MA00 | Corticosteriods | KEY-PRED INJECTABLE PRODUCT |
| 1170722 | MA00 | Corticosteriods | KEY-PRED SP INJECTABLE PRODUCT |
| 1171632 | MA00 | Corticosteriods | COTOLONE INJECTABLE PRODUCT |
| 1171633 | MA00 | Corticosteriods | COTOLONE ORAL PRODUCT |
| 1171634 | MA00 | Corticosteriods | COTOLONE PILL |
| 1172143 | MA00 | Corticosteriods | ASMALPRED PLUS ORAL LIQUID PRODUCT |
| 1172144 | MA00 | Corticosteriods | ASMALPRED PLUS ORAL PRODUCT |
| 1173110 | MA00 | Corticosteriods | DEPO-MEDROL INJECTABLE PRODUCT |
| 1173111 | MA00 | Corticosteriods | DEPO-PREDATE INJECTABLE PRODUCT |
| 1173399 | MA00 | Corticosteriods | HYDELTRA-T.B.A. INJECTABLE PRODUCT |
| 1173400 | MA00 | Corticosteriods | HYDELTRASOL INJECTABLE PRODUCT |
| 1175156 | MA00 | Corticosteriods | CODELSOL INJECTABLE PRODUCT |
| 1175157 | MA00 | Corticosteriods | CODELSON INJECTABLE PRODUCT |
| 1177661 | MA00 | Corticosteriods | PRI-CORTIN 50 INJECTABLE PRODUCT |
| 1178234 | MA00 | Corticosteriods | PRICORTIN INJECTABLE PRODUCT |
| 1179016 | MA00 | Corticosteriods | STINTISONE ORAL PRODUCT |
| 1180193 | MA00 | Corticosteriods | STINTISONE PILL |
| 1181408 | MA00 | Corticosteriods | TEMARIL-P ORAL PRODUCT |
| 1181409 | MA00 | Corticosteriods | TEMARIL-P PILL |
| 1183281 | MA00 | Corticosteriods | MEDICORT INJECTABLE PRODUCT |
| 1183437 | MA00 | Corticosteriods | ORAPRED ORAL LIQUID PRODUCT |
| 1183438 | MA00 | Corticosteriods | ORAPRED ORAL PRODUCT |
| 1183439 | MA00 | Corticosteriods | ORAPRED PILL |
| 1183796 | MA00 | Corticosteriods | MEDROL ORAL PRODUCT |
| 1184005 | MA00 | Corticosteriods | PEDIAPRED ORAL LIQUID PRODUCT |
| 1184006 | MA00 | Corticosteriods | PEDIAPRED ORAL PRODUCT |
| 1184840 | MA00 | Corticosteriods | SOLU-MEDROL INJECTABLE PRODUCT |
| 1184933 | MA00 | Corticosteriods | MILLIPRED ORAL LIQUID PRODUCT |
| 1184934 | MA00 | Corticosteriods | MILLIPRED ORAL PRODUCT |
| 1184935 | MA00 | Corticosteriods | MILLIPRED PILL |
| 1185242 | MA00 | Corticosteriods | PRED-JECT-50 INJECTABLE PRODUCT |
| 1185244 | MA00 | Corticosteriods | PREDACORT 50 INJECTABLE PRODUCT |
| 1185246 | MA00 | Corticosteriods | PREDAJECT-50 INJECTABLE PRODUCT |
| 1185247 | MA00 | Corticosteriods | PREDALONE 50 INJECTABLE PRODUCT |
| 1185248 | MA00 | Corticosteriods | PREDATE-50 INJECTABLE PRODUCT |
| 1185249 | MA00 | Corticosteriods | PREDCOR INJECTABLE PRODUCT |
| 1185252 | MA00 | Corticosteriods | PREDICORT RP INJECTABLE PRODUCT |

| | | | |
|---|---|---|---|
| 1185253 | MA00 | Corticosteriods | PREDICORT-50 INJECTABLE PRODUCT |
| 1185254 | MA00 | Corticosteriods | PREDNESOL ORAL PRODUCT |
| 1185255 | MA00 | Corticosteriods | PREDNESOL PILL |
| 1185263 | MA00 | Corticosteriods | PREDNORAL ORAL PRODUCT |
| 1185264 | MA00 | Corticosteriods | PREDNORAL PILL |
| 1185831 | MA00 | Corticosteriods | PRELONE ORAL LIQUID PRODUCT |
| 1185832 | MA00 | Corticosteriods | PRELONE ORAL PRODUCT |
| 1187894 | MA00 | Corticosteriods | VERIPRED ORAL LIQUID PRODUCT |
| 1187895 | MA00 | Corticosteriods | VERIPRED ORAL PRODUCT |
| 1191681 | MA00 | Corticosteriods | PREDNISTAB ORAL PRODUCT |
| 1191682 | MA00 | Corticosteriods | PREDNISTAB PILL |
| 1296584 | MA00 | Corticosteriods | ORAPRED DISINTEGRATING ORAL PRODUCT |
| 1493169 | MA00 | Corticosteriods | SOLU-DELTA-CORTEF INJECTABLE PRODUCT |
| 105403 | MA00 | Corticosteriods | METHYLPREDNISOLONE 125 MG/ML INJECTABLE SOLUTION |
| 105411 | MA00 | Corticosteriods | PREDNISOLONE 2.5 MG ORAL TABLET |
| 1358510 | MA00 | Corticosteriods | METHYLPREDNISOLONE ACETATE 20 MG/ML INJECTABLE SUSPENSION |
| 1358610 | MA00 | Corticosteriods | METHYLPREDNISOLONE ACETATE 40 MG/ML INJECTABLE SUSPENSION |
| 1358617 | MA00 | Corticosteriods | METHYLPREDNISOLONE ACETATE 80 MG/ML INJECTABLE SUSPENSION |
| 1743704 | MA00 | Corticosteriods | METHYLPREDNISOLONE 125 MG INJECTION |
| 1743720 | MA00 | Corticosteriods | METHYLPREDNISOLONE 500 MG INJECTION |
| 1743779 | MA00 | Corticosteriods | 1 ML METHYLPREDNISOLONE ACETATE 40 MG/ML INJECTION |
| 197970 | MA00 | Corticosteriods | METHYLPREDNISOLONE 24 MG ORAL TABLET |
| 197971 | MA00 | Corticosteriods | METHYLPREDNISOLONE 32 MG ORAL TABLET |
| 197973 | MA00 | Corticosteriods | METHYLPREDNISOLONE 8 MG ORAL TABLET |
| 198142 | MA00 | Corticosteriods | PREDNISOLONE 5 MG ORAL TABLET |
| 199343 | MA00 | Corticosteriods | PREDNISOLONE 1 MG ORAL TABLET |
| 199771 | MA00 | Corticosteriods | METHYLPREDNISOLONE 100 MG ORAL TABLET |
| 208988 | MA00 | Corticosteriods | PREDNISOLONE ACETATE 5 MG ORAL TABLET |
| 249066 | MA00 | Corticosteriods | PREDNISOLONE 5 MG/ML ORAL SOLUTION |
| 259966 | MA00 | Corticosteriods | METHYLPREDNISOLONE 4 MG ORAL TABLET |
| 283077 | MA00 | Corticosteriods | PREDNISOLONE 3 MG/ML ORAL SOLUTION |
| 311659 | MA00 | Corticosteriods | METHYLPREDNISOLONE 40 MG INJECTION |
| 314099 | MA00 | Corticosteriods | METHYLPREDNISOLONE 62.5 MG/ML INJECTABLE SOLUTION |
| 328161 | MA00 | Corticosteriods | METHYLPREDNISOLONE 16 MG ORAL TABLET |
| 346535 | MA00 | Corticosteriods | METHYLPREDNISOLONE 40 MG/ML INJECTABLE SUSPENSION |
| 348428 | MA00 | Corticosteriods | PREDNISOLONE 50 MG ORAL TABLET |
| 429198 | MA00 | Corticosteriods | PREDNISOLONE 2 MG ORAL TABLET |
| 436279 | MA00 | Corticosteriods | PREDNISOLONE 10 MG ORAL TABLET |
| 643123 | MA00 | Corticosteriods | PREDNISOLONE 10 MG DISINTEGRATING ORAL TABLET |
| 643125 | MA00 | Corticosteriods | PREDNISOLONE 15 MG DISINTEGRATING ORAL TABLET |
| 643127 | MA00 | Corticosteriods | PREDNISOLONE 30 MG DISINTEGRATING ORAL TABLET |
| 702306 | MA00 | Corticosteriods | PREDNISOLONE 4 MG/ML ORAL SOLUTION |
| 793099 | MA00 | Corticosteriods | PREDNISOLONE 3 MG/ML ORAL SUSPENSION |
| 794979 | MA00 | Corticosteriods | PREDNISOLONE 2 MG/ML ORAL SOLUTION |
| 1743725 | MA00 | Corticosteriods | METHYLPREDNISOLONE 1000 MG |
| 376656 | MA00 | Corticosteriods | METHYLPREDNISOLONE INJECTABLE SOLUTION |
| 1163486 | MA00 | Corticosteriods | METHYLPREDNISOLONE INJECTABLE PRODUCT |
| 1163488 | MA00 | Corticosteriods | METHYLPREDNISOLONE ORAL PRODUCT |
| 1165759 | MA00 | Corticosteriods | PREDNISOLONE ORAL PRODUCT |
| 1295508 | MA00 | Corticosteriods | PREDNISOLONE DISINTEGRATING ORAL PRODUCT |
| 1303126 | MA00 | Corticosteriods | RAYOS |
| 151587 | MA00 | Corticosteriods | DECORTISYL |
| 219350 | MA00 | Corticosteriods | PREDNICEN-M |
| 220028 | MA00 | Corticosteriods | STERAPRED |
| 220029 | MA00 | Corticosteriods | STERAPRED DS |
| 224927 | MA00 | Corticosteriods | LIQUID PRED |
| 224928 | MA00 | Corticosteriods | METICORTEN |
| 224929 | MA00 | Corticosteriods | ORASONE |
| 227730 | MA00 | Corticosteriods | DELTASONE |
| 352903 | MA00 | Corticosteriods | PREDNICOT |
| 380826 | MA00 | Corticosteriods | ECONOSONE |
| 541812 | MA00 | Corticosteriods | PREDONE |
| 582600 | MA00 | Corticosteriods | PREDNISONE INTENSOL |
| 795855 | MA00 | Corticosteriods | {12 (PREDNISONE 5 MG ORAL TABLET [STERAPRED]) } PACK [STERAPRED UNI-PAK 12 DAY] |
| 795857 | MA00 | Corticosteriods | {6 (PREDNISONE 5 MG ORAL TABLET [STERAPRED]) } PACK [STERAPRED UNI-PAK 6 DAY] |
| 795861 | MA00 | Corticosteriods | {6 (PREDNISONE 10 MG ORAL TABLET [STERAPRED DS]) } PACK [STERAPRED DS UNI-PAK 6 DAY] |
| 845491 | MA00 | Corticosteriods | {48 (PREDNISONE 5 MG ORAL TABLET [STERAPRED]) } PACK [STERAPRED 5 MG 12 DAY UNI-PAK] |
| 845492 | MA00 | Corticosteriods | {21 (PREDNISONE 5 MG ORAL TABLET [STERAPRED]) } PACK [STERAPRED 5 MG UNI-PAK] |

| | | | |
|---|---|---|---|
| 845493 | MA00 | Corticosteriods | {48 (PREDNISONE 10 MG ORAL TABLET [STERAPRED DS]) } PACK [STERAPRED DS 12 DAY UNI-PAK] |
| 845494 | MA00 | Corticosteriods | {21 (PREDNISONE 10 MG ORAL TABLET [STERAPRED DS]) } PACK [STERAPRED DS UNI-PAK] |
| 763179 | MA00 | Corticosteriods | {48 (PREDNISONE 5 MG ORAL TABLET) } PACK |
| 763181 | MA00 | Corticosteriods | {21 (PREDNISONE 5 MG ORAL TABLET) } PACK |
| 763183 | MA00 | Corticosteriods | {48 (PREDNISONE 10 MG ORAL TABLET) } PACK |
| 763185 | MA00 | Corticosteriods | {21 (PREDNISONE 10 MG ORAL TABLET) } PACK |
| 795854 | MA00 | Corticosteriods | {12 (PREDNISONE 5 MG ORAL TABLET) } PACK |
| 795856 | MA00 | Corticosteriods | {6 (PREDNISONE 5 MG ORAL TABLET) } PACK |
| 795860 | MA00 | Corticosteriods | {6 (PREDNISONE 10 MG ORAL TABLET) } PACK |
| 834395 | MA00 | Corticosteriods | {25 (PREDNISONE 10 MG ORAL TABLET) } PACK |
| 29523 | MA00 | Corticosteriods | MEPREDNISONE |
| 8640 | MA00 | Corticosteriods | PREDNISONE |
| 1008942 | MA00 | Corticosteriods | CHLORAMPHENICOL / PREDNISONE |
| 815961 | MA00 | Corticosteriods | PIROXICAM / PREDNISONE |
| 1742508 | MA00 | Corticosteriods | MEPREDNISONE DISODIUM PHOSPHATE |
| 105412 | MA00 | Corticosteriods | PREDNISONE 5 MG ORAL TABLET [DECORTISYL] |
| 1303131 | MA00 | Corticosteriods | PREDNISONE 1 MG DELAYED RELEASE ORAL TABLET [RAYOS] |
| 1303134 | MA00 | Corticosteriods | PREDNISONE 2 MG DELAYED RELEASE ORAL TABLET [RAYOS] |
| 1303137 | MA00 | Corticosteriods | PREDNISONE 5 MG DELAYED RELEASE ORAL TABLET [RAYOS] |
| 201082 | MA00 | Corticosteriods | PREDNISONE 1 MG ORAL TABLET [ECONOSONE] |
| 201083 | MA00 | Corticosteriods | PREDNISONE 5 MG ORAL TABLET [ECONOSONE] |
| 206199 | MA00 | Corticosteriods | PREDNISONE 1 MG/ML ORAL SOLUTION [LIQUID PRED] |
| 206754 | MA00 | Corticosteriods | PREDNISONE 1 MG ORAL TABLET [METICORTEN] |
| 206755 | MA00 | Corticosteriods | PREDNISONE 1 MG ORAL TABLET [ORASONE] |
| 206837 | MA00 | Corticosteriods | PREDNISONE 2.5 MG ORAL TABLET [DELTASONE] |
| 206954 | MA00 | Corticosteriods | PREDNISONE 5 MG ORAL TABLET [DELTASONE] |
| 206955 | MA00 | Corticosteriods | PREDNISONE 5 MG ORAL TABLET [ORASONE] |
| 206956 | MA00 | Corticosteriods | PREDNISONE 5 MG ORAL TABLET [PREDNICEN-M] |
| 206957 | MA00 | Corticosteriods | PREDNISONE 5 MG ORAL TABLET [STERAPRED] |
| 206988 | MA00 | Corticosteriods | PREDNISONE 10 MG ORAL TABLET [DELTASONE] |
| 206989 | MA00 | Corticosteriods | PREDNISONE 10 MG ORAL TABLET [ORASONE] |
| 206990 | MA00 | Corticosteriods | PREDNISONE 10 MG ORAL TABLET [STERAPRED DS] |
| 206997 | MA00 | Corticosteriods | PREDNISONE 20 MG ORAL TABLET [DELTASONE] |
| 206998 | MA00 | Corticosteriods | PREDNISONE 20 MG ORAL TABLET [ORASONE] |
| 207048 | MA00 | Corticosteriods | PREDNISONE 50 MG ORAL TABLET [DELTASONE] |
| 207049 | MA00 | Corticosteriods | PREDNISONE 50 MG ORAL TABLET [ORASONE] |
| 351399 | MA00 | Corticosteriods | PREDNISONE 5 MG ORAL TABLET [PREDNICOT] |
| 351400 | MA00 | Corticosteriods | PREDNISONE 10 MG ORAL TABLET [PREDNICOT] |
| 351401 | MA00 | Corticosteriods | PREDNISONE 20 MG ORAL TABLET [PREDNICOT] |
| 539948 | MA00 | Corticosteriods | PREDNISONE 1 MG/ML SYRUP [LIQUID PRED] |
| 541815 | MA00 | Corticosteriods | PREDNISONE 10 MG ORAL TABLET [PREDONE] |
| 541872 | MA00 | Corticosteriods | PREDNISONE 20 MG ORAL TABLET [PREDONE] |
| 541927 | MA00 | Corticosteriods | PREDNISONE 5 MG ORAL TABLET [LEVOXYL] |
| 582603 | MA00 | Corticosteriods | PREDNISONE 5 MG/ML ORAL SOLUTION [PREDNISONE INTENSOL] |
| 1303127 | MA00 | Corticosteriods | PREDNISONE 1 MG [RAYOS] |
| 1303133 | MA00 | Corticosteriods | PREDNISONE 2 MG [RAYOS] |
| 1303136 | MA00 | Corticosteriods | PREDNISONE 5 MG [RAYOS] |
| 539946 | MA00 | Corticosteriods | PREDNISONE 1 MG/ML [LIQUID PRED] |
| 541813 | MA00 | Corticosteriods | PREDNISONE 10 MG [PREDONE] |
| 541871 | MA00 | Corticosteriods | PREDNISONE 20 MG [PREDONE] |
| 541925 | MA00 | Corticosteriods | PREDNISONE 5 MG [LEVOXYL] |
| 564054 | MA00 | Corticosteriods | PREDNISONE 5 MG [DECORTISYL] |
| 565678 | MA00 | Corticosteriods | PREDNISONE 1 MG [ECONOSONE] |
| 565679 | MA00 | Corticosteriods | PREDNISONE 5 MG [ECONOSONE] |
| 567565 | MA00 | Corticosteriods | PREDNISONE 1 MG [METICORTEN] |
| 567566 | MA00 | Corticosteriods | PREDNISONE 1 MG [ORASONE] |
| 567645 | MA00 | Corticosteriods | PREDNISONE 2.5 MG [DELTASONE] |
| 567755 | MA00 | Corticosteriods | PREDNISONE 5 MG [DELTASONE] |
| 567756 | MA00 | Corticosteriods | PREDNISONE 5 MG [ORASONE] |
| 567757 | MA00 | Corticosteriods | PREDNISONE 5 MG [PREDNICEN-M] |
| 567758 | MA00 | Corticosteriods | PREDNISONE 5 MG [STERAPRED] |
| 567784 | MA00 | Corticosteriods | PREDNISONE 10 MG [DELTASONE] |
| 567785 | MA00 | Corticosteriods | PREDNISONE 10 MG [ORASONE] |
| 567786 | MA00 | Corticosteriods | PREDNISONE 10 MG [STERAPRED DS] |
| 567793 | MA00 | Corticosteriods | PREDNISONE 20 MG [DELTASONE] |
| 567794 | MA00 | Corticosteriods | PREDNISONE 20 MG [ORASONE] |
| 567841 | MA00 | Corticosteriods | PREDNISONE 50 MG [DELTASONE] |
| 567842 | MA00 | Corticosteriods | PREDNISONE 50 MG [ORASONE] |
| 575330 | MA00 | Corticosteriods | PREDNISONE 5 MG [PREDNICOT] |
| 575331 | MA00 | Corticosteriods | PREDNISONE 10 MG [PREDNICOT] |

| | | | |
|---|---|---|---|
| 575332 | MA00 | Corticosteriods | PREDNISONE 20 MG [PREDNICOT] |
| 582601 | MA00 | Corticosteriods | PREDNISONE 5 MG/ML [PREDNISONE INTENSOL] |
| 1303128 | MA00 | Corticosteriods | PREDNISONE DELAYED RELEASE ORAL TABLET [RAYOS] |
| 364008 | MA00 | Corticosteriods | PREDNISONE ORAL SOLUTION [LIQUID PRED] |
| 368654 | MA00 | Corticosteriods | PREDNISONE ORAL TABLET [PREDNICOT] |
| 368693 | MA00 | Corticosteriods | PREDNISONE ORAL TABLET [LEVOXYL] |
| 368790 | MA00 | Corticosteriods | PREDNISONE ORAL TABLET [ORASONE] |
| 368806 | MA00 | Corticosteriods | PREDNISONE ORAL TABLET [STERAPRED DS] |
| 368812 | MA00 | Corticosteriods | PREDNISONE ORAL TABLET [STERAPRED] |
| 368813 | MA00 | Corticosteriods | PREDNISONE ORAL TABLET [PREDNICEN-M] |
| 368867 | MA00 | Corticosteriods | PREDNISONE ORAL TABLET [METICORTEN] |
| 380827 | MA00 | Corticosteriods | PREDNISONE ORAL TABLET [ECONOSONE] |
| 380828 | MA00 | Corticosteriods | PREDNISONE ORAL TABLET [DECORTISYL] |
| 539947 | MA00 | Corticosteriods | PREDNISONE SYRUP [LIQUID PRED] |
| 541814 | MA00 | Corticosteriods | PREDNISONE ORAL TABLET [PREDONE] |
| 582602 | MA00 | Corticosteriods | PREDNISONE ORAL SOLUTION [PREDNISONE INTENSOL] |
| 92354 | MA00 | Corticosteriods | PREDNISONE ORAL TABLET [DELTASONE] |
| 1171028 | MA00 | Corticosteriods | DECORTISYL ORAL PRODUCT |
| 1171029 | MA00 | Corticosteriods | DECORTISYL PILL |
| 1171797 | MA00 | Corticosteriods | ECONOSONE ORAL PRODUCT |
| 1171798 | MA00 | Corticosteriods | ECONOSONE PILL |
| 1172410 | MA00 | Corticosteriods | DELTASONE ORAL PRODUCT |
| 1172411 | MA00 | Corticosteriods | DELTASONE PILL |
| 1176945 | MA00 | Corticosteriods | METICORTEN ORAL PRODUCT |
| 1176946 | MA00 | Corticosteriods | METICORTEN PILL |
| 1178546 | MA00 | Corticosteriods | LIQUID PRED ORAL LIQUID PRODUCT |
| 1178547 | MA00 | Corticosteriods | LIQUID PRED ORAL PRODUCT |
| 1178982 | MA00 | Corticosteriods | STERAPRED DS ORAL PRODUCT |
| 1178983 | MA00 | Corticosteriods | STERAPRED DS PILL |
| 1178984 | MA00 | Corticosteriods | STERAPRED ORAL PRODUCT |
| 1178985 | MA00 | Corticosteriods | STERAPRED PILL |
| 1183442 | MA00 | Corticosteriods | ORASONE ORAL PRODUCT |
| 1183443 | MA00 | Corticosteriods | ORASONE PILL |
| 1185256 | MA00 | Corticosteriods | PREDNICEN-M ORAL PRODUCT |
| 1185257 | MA00 | Corticosteriods | PREDNICEN-M PILL |
| 1185258 | MA00 | Corticosteriods | PREDNICOT ORAL PRODUCT |
| 1185259 | MA00 | Corticosteriods | PREDNICOT PILL |
| 1185261 | MA00 | Corticosteriods | PREDNISONE INTENSOL ORAL LIQUID PRODUCT |
| 1185262 | MA00 | Corticosteriods | PREDNISONE INTENSOL ORAL PRODUCT |
| 1185265 | MA00 | Corticosteriods | PREDONE ORAL PRODUCT |
| 1185266 | MA00 | Corticosteriods | PREDONE PILL |
| 1303129 | MA00 | Corticosteriods | RAYOS ORAL PRODUCT |
| 1303130 | MA00 | Corticosteriods | RAYOS PILL |
| 1303125 | MA00 | Corticosteriods | PREDNISONE 1 MG DELAYED RELEASE ORAL TABLET |
| 1303132 | MA00 | Corticosteriods | PREDNISONE 2 MG DELAYED RELEASE ORAL TABLET |
| 1303135 | MA00 | Corticosteriods | PREDNISONE 5 MG DELAYED RELEASE ORAL TABLET |
| 198144 | MA00 | Corticosteriods | PREDNISONE 1 MG ORAL TABLET |
| 198145 | MA00 | Corticosteriods | PREDNISONE 10 MG ORAL TABLET |
| 198146 | MA00 | Corticosteriods | PREDNISONE 2.5 MG ORAL TABLET |
| 198148 | MA00 | Corticosteriods | PREDNISONE 50 MG ORAL TABLET |
| 205301 | MA00 | Corticosteriods | PREDNISONE 5 MG/ML ORAL SOLUTION |
| 248682 | MA00 | Corticosteriods | MEPREDNISONE 8 MG ORAL TABLET |
| 248702 | MA00 | Corticosteriods | MEPREDNISONE 40 MG ORAL TABLET |
| 248725 | MA00 | Corticosteriods | MEPREDNISONE 4 MG ORAL TABLET |
| 248794 | MA00 | Corticosteriods | MEPREDNISONE 4 MG/ML ORAL SOLUTION |
| 250996 | MA00 | Corticosteriods | PIROXICAM 10 MG / PREDNISONE 2 MG DELAYED RELEASE ORAL TABLET |
| 251947 | MA00 | Corticosteriods | PIROXICAM 10 MG / PREDNISONE 2 MG ORAL TABLET |
| 312615 | MA00 | Corticosteriods | PREDNISONE 20 MG ORAL TABLET |
| 312616 | MA00 | Corticosteriods | PREDNISONE 25 MG ORAL TABLET |
| 312617 | MA00 | Corticosteriods | PREDNISONE 5 MG ORAL TABLET |
| 312618 | MA00 | Corticosteriods | PREDNISONE 1 MG/ML SYRUP |
| 315187 | MA00 | Corticosteriods | PREDNISONE 1 MG/ML ORAL SOLUTION |
| 429331 | MA00 | Corticosteriods | PREDNISONE 8 MG ORAL TABLET |
| 429332 | MA00 | Corticosteriods | PREDNISONE 40 MG ORAL TABLET |
| 746380 | MA00 | Corticosteriods | PREDNISONE 10 MG ORAL CAPSULE |
| 746836 | MA00 | Corticosteriods | PREDNISONE 20 MG ORAL CAPSULE |
| 746837 | MA00 | Corticosteriods | PREDNISONE 5 MG ORAL CAPSULE |
| 316580 | MA00 | Corticosteriods | PREDNISONE 1 MG/ML |
| 316581 | MA00 | Corticosteriods | PREDNISONE 2.5 MG |
| 316582 | MA00 | Corticosteriods | PREDNISONE 20 MG |
| 316583 | MA00 | Corticosteriods | PREDNISONE 25 MG |

| | | | |
|---|---|---|---|
| 316584 | MA00 | Corticosteriods | PREDNISONE 5 MG |
| 316585 | MA00 | Corticosteriods | PREDNISONE 5 MG/ML |
| 316586 | MA00 | Corticosteriods | PREDNISONE 50 MG |
| 317475 | MA00 | Corticosteriods | PREDNISONE 1 MG |
| 317663 | MA00 | Corticosteriods | PREDNISONE 10 MG |
| 334021 | MA00 | Corticosteriods | MEPREDNISONE 4 MG/ML |
| 334022 | MA00 | Corticosteriods | MEPREDNISONE 4 MG |
| 334023 | MA00 | Corticosteriods | MEPREDNISONE 40 MG |
| 334024 | MA00 | Corticosteriods | MEPREDNISONE 8 MG |
| 335473 | MA00 | Corticosteriods | PREDNISONE 2 MG |
| 438434 | MA00 | Corticosteriods | PREDNISONE 0.005 MG/MG |
| 451141 | MA00 | Corticosteriods | PREDNISONE 100 MG |
| 451143 | MA00 | Corticosteriods | PREDNISONE 30 MG |
| 451144 | MA00 | Corticosteriods | PREDNISONE 40 MG |
| 451147 | MA00 | Corticosteriods | PREDNISONE 8 MG |
| 1303124 | MA00 | Corticosteriods | PREDNISONE DELAYED RELEASE ORAL TABLET |
| 372772 | MA00 | Corticosteriods | MEPREDNISONE ORAL SOLUTION |
| 372773 | MA00 | Corticosteriods | MEPREDNISONE ORAL TABLET |
| 373495 | MA00 | Corticosteriods | PIROXICAM / PREDNISONE DELAYED RELEASE ORAL TABLET |
| 373583 | MA00 | Corticosteriods | PREDNISONE ORAL SOLUTION |
| 373584 | MA00 | Corticosteriods | PREDNISONE SYRUP |
| 373585 | MA00 | Corticosteriods | PREDNISONE ORAL TABLET |
| 376283 | MA00 | Corticosteriods | PIROXICAM / PREDNISONE ORAL TABLET |
| 746379 | MA00 | Corticosteriods | PREDNISONE ORAL CAPSULE |
| 1159297 | MA00 | Corticosteriods | PIROXICAM / PREDNISONE ORAL PRODUCT |
| 1159298 | MA00 | Corticosteriods | PIROXICAM / PREDNISONE PILL |
| 1161704 | MA00 | Corticosteriods | PREDNISONE ORAL LIQUID PRODUCT |
| 1161705 | MA00 | Corticosteriods | PREDNISONE ORAL PRODUCT |
| 1161706 | MA00 | Corticosteriods | PREDNISONE PILL |
| 1165700 | MA00 | Corticosteriods | MEPREDNISONE ORAL LIQUID PRODUCT |
| 1165701 | MA00 | Corticosteriods | MEPREDNISONE ORAL PRODUCT |
| 1165702 | MA00 | Corticosteriods | MEPREDNISONE PILL |
| 51407022430 | MA11 | Corticosteriods | budesonide |
| 51862058201 | MA11 | Corticosteriods | budesonide |
| 68682030930 | MA11 | Corticosteriods | budesonide |
| 378715505 | MA11 | Corticosteriods | budesonide |
| 591251030 | MA11 | Corticosteriods | budesonide |
| 574985510 | MA11 | Corticosteriods | budesonide |
| 60687042195 | MA11 | Corticosteriods | budesonide |
| 54868491002 | MA11 | Corticosteriods | budesonide |
| 64980025501 | MA11 | Corticosteriods | budesonide |
| 60687042125 | MA11 | Corticosteriods | budesonide |
| 51862058001 | MA11 | Corticosteriods | budesonide |
| 574985010 | MA11 | Corticosteriods | budesonide |
| 93744501 | MA11 | Corticosteriods | budesonide |
| 378715501 | MA11 | Corticosteriods | budesonide |
| 68012030930 | MA11 | Corticosteriods | budesonide |
| 51079002003 | MA11 | Corticosteriods | budesonide |
| 16714082901 | MA11 | Corticosteriods | budesonide |
| 49884050101 | MA11 | Corticosteriods | budesonide |
| 65162077810 | MA11 | Corticosteriods | budesonide |
| 68382072001 | MA11 | Corticosteriods | budesonide |
| 186070210 | MA11 | Corticosteriods | budesonide |
| 54868491001 | MA11 | Corticosteriods | budesonide |
| 55700059090 | MA11 | Corticosteriods | budesonide |
| 54868491000 | MA11 | Corticosteriods | budesonide |
| 51079002001 | MA11 | Corticosteriods | budesonide |
| 67263034301 | MA11 | Corticosteriods | budesonide |
| 65483070210 | MA11 | Corticosteriods | budesonide |
| 47781014601 | MA11 | Corticosteriods | budesonide |
| 54868491003 | MA11 | Corticosteriods | budesonide |
| 60429026401 | MA11 | Corticosteriods | budesonide |
| 59762005001 | MA11 | Corticosteriods | methylprednisolone |
| 23629004701 | MA11 | Corticosteriods | methylprednisolone |
| 904217519 | MA11 | Corticosteriods | methylprednisolone |
| 247136530 | MA11 | Corticosteriods | methylprednisolone |
| 35356019421 | MA11 | Corticosteriods | methylprednisolone |
| 677183101 | MA11 | Corticosteriods | methylprednisolone |
| 55887055450 | MA11 | Corticosteriods | methylprednisolone |
| 59762332701 | MA11 | Corticosteriods | methylprednisolone |
| 68382091911 | MA11 | Corticosteriods | methylprednisolone |

| | | | |
|---|---|---|---|
| 66116066221 | MA11 | Corticosteriods | methylprednisolone |
| 68387017001 | MA11 | Corticosteriods | methylprednisolone |
| 62269035124 | MA11 | Corticosteriods | methylprednisolone |
| 62269035121 | MA11 | Corticosteriods | methylprednisolone |
| 18837008621 | MA11 | Corticosteriods | methylprednisolone |
| 55175110301 | MA11 | Corticosteriods | methylprednisolone |
| 58016471901 | MA11 | Corticosteriods | methylprednisolone |
| 54868291300 | MA11 | Corticosteriods | methylprednisolone |
| 9005602 | MA11 | Corticosteriods | methylprednisolone |
| 54868291303 | MA11 | Corticosteriods | methylprednisolone |
| 677056513 | MA11 | Corticosteriods | methylprednisolone |
| 59762444002 | MA11 | Corticosteriods | methylprednisolone |
| 9005605 | MA11 | Corticosteriods | methylprednisolone |
| 51285030121 | MA11 | Corticosteriods | methylprednisolone |
| 54868662401 | MA11 | Corticosteriods | methylprednisolone |
| 440776021 | MA11 | Corticosteriods | methylprednisolone |
| 57866403701 | MA11 | Corticosteriods | methylprednisolone |
| 54569451600 | MA11 | Corticosteriods | methylprednisolone |
| 68084014901 | MA11 | Corticosteriods | methylprednisolone |
| 55289064930 | MA11 | Corticosteriods | methylprednisolone |
| 9017601 | MA11 | Corticosteriods | methylprednisolone |
| 63739016110 | MA11 | Corticosteriods | methylprednisolone |
| 247136501 | MA11 | Corticosteriods | methylprednisolone |
| 60346060820 | MA11 | Corticosteriods | methylprednisolone |
| 603459315 | MA11 | Corticosteriods | methylprednisolone |
| 54868077601 | MA11 | Corticosteriods | methylprednisolone |
| 72189000821 | MA11 | Corticosteriods | methylprednisolone |
| 57866710001 | MA11 | Corticosteriods | methylprednisolone |
| 9007301 | MA11 | Corticosteriods | methylprednisolone |
| 63629391001 | MA11 | Corticosteriods | methylprednisolone |
| 52544079001 | MA11 | Corticosteriods | methylprednisolone |
| 49999015321 | MA11 | Corticosteriods | methylprednisolone |
| 60346060844 | MA11 | Corticosteriods | methylprednisolone |
| 603459321 | MA11 | Corticosteriods | methylprednisolone |
| 23629004710 | MA11 | Corticosteriods | methylprednisolone |
| 781502207 | MA11 | Corticosteriods | methylprednisolone |
| 54868495201 | MA11 | Corticosteriods | methylprednisolone |
| 59746000109 | MA11 | Corticosteriods | methylprednisolone |
| 67263040086 | MA11 | Corticosteriods | methylprednisolone |
| 50090309400 | MA11 | Corticosteriods | methylprednisolone |
| 50742018901 | MA11 | Corticosteriods | methylprednisolone |
| 67253036021 | MA11 | Corticosteriods | methylprednisolone |
| 63739016115 | MA11 | Corticosteriods | methylprednisolone |
| 42806040021 | MA11 | Corticosteriods | methylprednisolone |
| 66993084225 | MA11 | Corticosteriods | methylprednisolone |
| 51991018831 | MA11 | Corticosteriods | methylprednisolone |
| 59762444003 | MA11 | Corticosteriods | methylprednisolone |
| 54868291301 | MA11 | Corticosteriods | methylprednisolone |
| 68115096021 | MA11 | Corticosteriods | methylprednisolone |
| 50436403701 | MA11 | Corticosteriods | methylprednisolone |
| 68084014911 | MA11 | Corticosteriods | methylprednisolone |
| 403130418 | MA11 | Corticosteriods | methylprednisolone |
| 527129607 | MA11 | Corticosteriods | methylprednisolone |
| 904657461 | MA11 | Corticosteriods | methylprednisolone |
| 904217560 | MA11 | Corticosteriods | methylprednisolone |
| 59762004901 | MA11 | Corticosteriods | methylprednisolone |
| 49884049001 | MA11 | Corticosteriods | methylprednisolone |
| 23490590201 | MA11 | Corticosteriods | methylprednisolone |
| 182105003 | MA11 | Corticosteriods | methylprednisolone |
| 42806040001 | MA11 | Corticosteriods | methylprednisolone |
| 59746000103 | MA11 | Corticosteriods | methylprednisolone |
| 21695008021 | MA11 | Corticosteriods | methylprednisolone |
| 72647033104 | MA11 | Corticosteriods | methylprednisolone |
| 55289064998 | MA11 | Corticosteriods | methylprednisolone |
| 59746000206 | MA11 | Corticosteriods | methylprednisolone |
| 54569458000 | MA11 | Corticosteriods | methylprednisolone |
| 247001221 | MA11 | Corticosteriods | methylprednisolone |
| 10544032221 | MA11 | Corticosteriods | methylprednisolone |
| 555030138 | MA11 | Corticosteriods | methylprednisolone |
| 42549052221 | MA11 | Corticosteriods | methylprednisolone |
| 49999015330 | MA11 | Corticosteriods | methylprednisolone |

| | | | |
|---|---|---|---|
| 55045351301 | MA11 | Corticosteriods | methylprednisolone |
| 364046721 | MA11 | Corticosteriods | methylprednisolone |
| 247136520 | MA11 | Corticosteriods | methylprednisolone |
| 58016200101 | MA11 | Corticosteriods | methylprednisolone |
| 68382091634 | MA11 | Corticosteriods | methylprednisolone |
| 60346060810 | MA11 | Corticosteriods | methylprednisolone |
| 52959010000 | MA11 | Corticosteriods | methylprednisolone |
| 66993084021 | MA11 | Corticosteriods | methylprednisolone |
| 9002201 | MA11 | Corticosteriods | methylprednisolone |
| 49884049059 | MA11 | Corticosteriods | methylprednisolone |
| 9004902 | MA11 | Corticosteriods | methylprednisolone |
| 591079001 | MA11 | Corticosteriods | methylprednisolone |
| 254421613 | MA11 | Corticosteriods | methylprednisolone |
| 55887054425 | MA11 | Corticosteriods | methylprednisolone |
| 35356076321 | MA11 | Corticosteriods | methylprednisolone |
| 54868495200 | MA11 | Corticosteriods | methylprednisolone |
| 63187016021 | MA11 | Corticosteriods | methylprednisolone |
| 591079021 | MA11 | Corticosteriods | methylprednisolone |
| 9005604 | MA11 | Corticosteriods | methylprednisolone |
| 54569103600 | MA11 | Corticosteriods | methylprednisolone |
| 66267096121 | MA11 | Corticosteriods | methylprednisolone |
| 247136521 | MA11 | Corticosteriods | methylprednisolone |
| 50090263800 | MA11 | Corticosteriods | methylprednisolone |
| 59746000106 | MA11 | Corticosteriods | methylprednisolone |
| 58016200401 | MA11 | Corticosteriods | methylprednisolone |
| 55045181108 | MA11 | Corticosteriods | methylprednisolone |
| 68001000501 | MA11 | Corticosteriods | methylprednisolone |
| 781502201 | MA11 | Corticosteriods | methylprednisolone |
| 59746000314 | MA11 | Corticosteriods | methylprednisolone |
| 9015501 | MA11 | Corticosteriods | methylprednisolone |
| 51285030102 | MA11 | Corticosteriods | methylprednisolone |
| 63304059122 | MA11 | Corticosteriods | methylprednisolone |
| 50742018921 | MA11 | Corticosteriods | methylprednisolone |
| 364046701 | MA11 | Corticosteriods | methylprednisolone |
| 182105001 | MA11 | Corticosteriods | methylprednisolone |
| 51991018801 | MA11 | Corticosteriods | methylprednisolone |
| 677183113 | MA11 | Corticosteriods | methylprednisolone |
| 33358024121 | MA11 | Corticosteriods | methylprednisolone |
| 68382091818 | MA11 | Corticosteriods | methylprednisolone |
| 254421628 | MA11 | Corticosteriods | methylprednisolone |
| 68030403701 | MA11 | Corticosteriods | methylprednisolone |
| 63874041321 | MA11 | Corticosteriods | methylprednisolone |
| 66993084002 | MA11 | Corticosteriods | methylprednisolone |
| 68382091601 | MA11 | Corticosteriods | methylprednisolone |
| 63187080221 | MA11 | Corticosteriods | methylprednisolone |
| 677056501 | MA11 | Corticosteriods | methylprednisolone |
| 72647033101 | MA11 | Corticosteriods | methylprednisolone |
| 9002001 | MA11 | Corticosteriods | methylprednisolone |
| 71205005421 | MA11 | Corticosteriods | methylprednisolone |
| 59746000204 | MA11 | Corticosteriods | methylprednisolone |
| 54569032700 | MA11 | Corticosteriods | methylprednisolone |
| 63739013310 | MA11 | Corticosteriods | methylprednisolone |
| 55887095321 | MA11 | Corticosteriods | methylprednisolone |
| 9005603 | MA11 | Corticosteriods | methylprednisolone |
| 50090009200 | MA11 | Corticosteriods | methylprednisolone |
| 61919014921 | MA11 | Corticosteriods | methylprednisolone |
| 59762005101 | MA11 | Corticosteriods | methylprednisolone |
| 67253036010 | MA11 | Corticosteriods | methylprednisolone |
| 16590014921 | MA11 | Corticosteriods | methylprednisolone |
| 59746001504 | MA11 | Corticosteriods | methylprednisolone |
| 55045125909 | MA11 | Corticosteriods | methylprednisolone |
| 51991018821 | MA11 | Corticosteriods | methylprednisolone |
| 904691461 | MA11 | Corticosteriods | methylprednisolone |
| 59762332702 | MA11 | Corticosteriods | methylprednisolone |
| 52544079021 | MA11 | Corticosteriods | methylprednisolone |
| 555030102 | MA11 | Corticosteriods | methylprednisolone |
| 68382091711 | MA11 | Corticosteriods | methylprednisolone |
| 54868291302 | MA11 | Corticosteriods | methylprednisolone |
| 35356048310 | MA11 | Corticosteriods | methylprednisolone acetate |
| 54868118500 | MA11 | Corticosteriods | methylprednisolone acetate |
| 54569221300 | MA11 | Corticosteriods | methylprednisolone acetate |

| | | | |
|---|---|---|---|
| 9347503 | MA11 | Corticosteriods | methylprednisolone acetate |
| 9307301 | MA11 | Corticosteriods | methylprednisolone acetate |
| 68258889805 | MA11 | Corticosteriods | methylprednisolone acetate |
| 35356048405 | MA11 | Corticosteriods | methylprednisolone acetate |
| 9028051 | MA11 | Corticosteriods | methylprednisolone acetate |
| 54868059000 | MA11 | Corticosteriods | methylprednisolone acetate |
| 9347522 | MA11 | Corticosteriods | methylprednisolone acetate |
| 55045324205 | MA11 | Corticosteriods | methylprednisolone acetate |
| 54569337400 | MA11 | Corticosteriods | methylprednisolone acetate |
| 703003101 | MA11 | Corticosteriods | methylprednisolone acetate |
| 781313295 | MA11 | Corticosteriods | methylprednisolone acetate |
| 54569190101 | MA11 | Corticosteriods | methylprednisolone acetate |
| 21695085005 | MA11 | Corticosteriods | methylprednisolone acetate |
| 9307323 | MA11 | Corticosteriods | methylprednisolone acetate |
| 58016489301 | MA11 | Corticosteriods | methylprednisolone acetate |
| 21695095205 | MA11 | Corticosteriods | methylprednisolone acetate |
| 54868199400 | MA11 | Corticosteriods | methylprednisolone acetate |
| 21695095201 | MA11 | Corticosteriods | methylprednisolone acetate |
| 781313775 | MA11 | Corticosteriods | methylprednisolone acetate |
| 781313670 | MA11 | Corticosteriods | methylprednisolone acetate |
| 703006301 | MA11 | Corticosteriods | methylprednisolone acetate |
| 54868389601 | MA11 | Corticosteriods | methylprednisolone acetate |
| 54868118501 | MA11 | Corticosteriods | methylprednisolone acetate |
| 53225365001 | MA11 | Corticosteriods | methylprednisolone acetate |
| 57866389501 | MA11 | Corticosteriods | methylprednisolone acetate |
| 9307322 | MA11 | Corticosteriods | methylprednisolone acetate |
| 9307303 | MA11 | Corticosteriods | methylprednisolone acetate |
| 54569512500 | MA11 | Corticosteriods | methylprednisolone acetate |
| 9028003 | MA11 | Corticosteriods | methylprednisolone acetate |
| 703004501 | MA11 | Corticosteriods | methylprednisolone acetate |
| 9027401 | MA11 | Corticosteriods | methylprednisolone acetate |
| 21695084910 | MA11 | Corticosteriods | methylprednisolone acetate |
| 54868389602 | MA11 | Corticosteriods | methylprednisolone acetate |
| 58016993401 | MA11 | Corticosteriods | methylprednisolone acetate |
| 54569394600 | MA11 | Corticosteriods | methylprednisolone acetate |
| 12280025605 | MA11 | Corticosteriods | methylprednisolone acetate |
| 9030602 | MA11 | Corticosteriods | methylprednisolone acetate |
| 54569426500 | MA11 | Corticosteriods | methylprednisolone acetate |
| 402106810 | MA11 | Corticosteriods | methylprednisolone acetate |
| 703004301 | MA11 | Corticosteriods | methylprednisolone acetate |
| 703005101 | MA11 | Corticosteriods | methylprednisolone acetate |
| 12280025601 | MA11 | Corticosteriods | methylprednisolone acetate |
| 54868389600 | MA11 | Corticosteriods | methylprednisolone acetate |
| 54569223200 | MA11 | Corticosteriods | methylprednisolone acetate |
| 781313171 | MA11 | Corticosteriods | methylprednisolone acetate |
| 781313271 | MA11 | Corticosteriods | methylprednisolone acetate |
| 703005104 | MA11 | Corticosteriods | methylprednisolone acetate |
| 9030612 | MA11 | Corticosteriods | methylprednisolone acetate |
| 49836001117 | MA11 | Corticosteriods | methylprednisolone acetate |
| 21695095210 | MA11 | Corticosteriods | methylprednisolone acetate |
| 9347501 | MA11 | Corticosteriods | methylprednisolone acetate |
| 49836001418 | MA11 | Corticosteriods | methylprednisolone acetate |
| 35356008301 | MA11 | Corticosteriods | methylprednisolone acetate |
| 9347523 | MA11 | Corticosteriods | methylprednisolone acetate |
| 55045324301 | MA11 | Corticosteriods | methylprednisolone acetate |
| 35356017805 | MA11 | Corticosteriods | methylprednisolone acetate |
| 703003104 | MA11 | Corticosteriods | methylprednisolone acetate |
| 781313195 | MA11 | Corticosteriods | methylprednisolone acetate |
| 9028052 | MA11 | Corticosteriods | methylprednisolone acetate |
| 9028002 | MA11 | Corticosteriods | methylprednisolone acetate |
| 71283062502 | MA11 | Corticosteriods | methylprednisolone acetate in sodium chloride,iso-osmotic/PF |
| 71283063402 | MA11 | Corticosteriods | methylprednisolone acetate in sodium chloride,iso-osmotic/PF |
| 71283063601 | MA11 | Corticosteriods | methylprednisolone acetate in sterile water for injection |
| 71283061901 | MA11 | Corticosteriods | methylprednisolone acetate in sterile water for injection |
| 62991163502 | MA11 | Corticosteriods | methylprednisolone acetate, micronized |
| 38779014403 | MA11 | Corticosteriods | methylprednisolone acetate, micronized |
| 51552095805 | MA11 | Corticosteriods | methylprednisolone acetate, micronized |
| 55812023903 | MA11 | Corticosteriods | methylprednisolone acetate, micronized |
| 52372828202 | MA11 | Corticosteriods | methylprednisolone acetate, micronized |
| 52372828203 | MA11 | Corticosteriods | methylprednisolone acetate, micronized |
| 62991163501 | MA11 | Corticosteriods | methylprednisolone acetate, micronized |

| | | | |
|---|---|---|---|
| 55812023902 | MA11 | Corticosteriods | methylprednisolone acetate, micronized |
| 38779014405 | MA11 | Corticosteriods | methylprednisolone acetate, micronized |
| 38779014404 | MA11 | Corticosteriods | methylprednisolone acetate, micronized |
| 38779014425 | MA11 | Corticosteriods | methylprednisolone acetate, micronized |
| 51552095802 | MA11 | Corticosteriods | methylprednisolone acetate, micronized |
| 52372828210 | MA11 | Corticosteriods | methylprednisolone acetate, micronized |
| 52372828250 | MA11 | Corticosteriods | methylprednisolone acetate, micronized |
| 52372828204 | MA11 | Corticosteriods | methylprednisolone acetate, micronized |
| 49452468801 | MA11 | Corticosteriods | methylprednisolone acetate, micronized |
| 52372828207 | MA11 | Corticosteriods | methylprednisolone acetate, micronized |
| 62991163505 | MA11 | Corticosteriods | methylprednisolone acetate, micronized |
| 55812023904 | MA11 | Corticosteriods | methylprednisolone acetate, micronized |
| 52372828206 | MA11 | Corticosteriods | methylprednisolone acetate, micronized |
| 38779014406 | MA11 | Corticosteriods | methylprednisolone acetate, micronized |
| 38779014415 | MA11 | Corticosteriods | methylprednisolone acetate, micronized |
| 10695004110 | MA11 | Corticosteriods | methylprednisolone acetate, micronized |
| 52372828205 | MA11 | Corticosteriods | methylprednisolone acetate, micronized |
| 51552095804 | MA11 | Corticosteriods | methylprednisolone acetate, micronized |
| 49452468802 | MA11 | Corticosteriods | methylprednisolone acetate, micronized |
| 38779014409 | MA11 | Corticosteriods | methylprednisolone acetate, micronized |
| 49452468803 | MA11 | Corticosteriods | methylprednisolone acetate, micronized |
| 51927133200 | MA11 | Corticosteriods | methylprednisolone acetate, micronized |
| 38779014408 | MA11 | Corticosteriods | methylprednisolone acetate, micronized |
| 55812023901 | MA11 | Corticosteriods | methylprednisolone acetate, micronized |
| 52372828201 | MA11 | Corticosteriods | methylprednisolone acetate, micronized |
| 55812023905 | MA11 | Corticosteriods | methylprednisolone acetate, micronized |
| 38779014411 | MA11 | Corticosteriods | methylprednisolone acetate, micronized |
| 62991163503 | MA11 | Corticosteriods | methylprednisolone acetate, micronized |
| 62991163504 | MA11 | Corticosteriods | methylprednisolone acetate, micronized |
| 76420052001 | MA11 | Corticosteriods | methylprednisolone acetate/bupivacaine HCl |
| 71283062301 | MA11 | Corticosteriods | methylprednisolone acetate/bupivacaine HCl in sterile water |
| 71283062401 | MA11 | Corticosteriods | methylprednisolone acetate/bupivacaine HCl in sterile water |
| 49836001219 | MA11 | Corticosteriods | methylprednisolone acetate/norflurane/HFC 245fa |
| 49836001520 | MA11 | Corticosteriods | methylprednisolone acetate/norflurane/HFC 245fa |
| 76420026001 | MA11 | Corticosteriods | methylprednisolone acetate/norflurane/HFC 245fa |
| 76420026201 | MA11 | Corticosteriods | methylprednisolone acetate/norflurane/HFC 245fa |
| 70121100001 | MA11 | Corticosteriods | methylprednisolone sodium succinate |
| 409568502 | MA11 | Corticosteriods | methylprednisolone sodium succinate |
| 25021080705 | MA11 | Corticosteriods | methylprednisolone sodium succinate |
| 9069801 | MA11 | Corticosteriods | methylprednisolone sodium succinate |
| 9085001 | MA11 | Corticosteriods | methylprednisolone sodium succinate |
| 9011319 | MA11 | Corticosteriods | methylprednisolone sodium succinate |
| 55390021010 | MA11 | Corticosteriods | methylprednisolone sodium succinate |
| 54868076800 | MA11 | Corticosteriods | methylprednisolone sodium succinate |
| 409568523 | MA11 | Corticosteriods | methylprednisolone sodium succinate |
| 409568401 | MA11 | Corticosteriods | methylprednisolone sodium succinate |
| 9019009 | MA11 | Corticosteriods | methylprednisolone sodium succinate |
| 25021080810 | MA11 | Corticosteriods | methylprednisolone sodium succinate |
| 9011312 | MA11 | Corticosteriods | methylprednisolone sodium succinate |
| 63323026530 | MA11 | Corticosteriods | methylprednisolone sodium succinate |
| 12280023201 | MA11 | Corticosteriods | methylprednisolone sodium succinate |
| 74568401 | MA11 | Corticosteriods | methylprednisolone sodium succinate |
| 409568423 | MA11 | Corticosteriods | methylprednisolone sodium succinate |
| 9075801 | MA11 | Corticosteriods | methylprednisolone sodium succinate |
| 58016945201 | MA11 | Corticosteriods | methylprednisolone sodium succinate |
| 409321805 | MA11 | Corticosteriods | methylprednisolone sodium succinate |
| 74563108 | MA11 | Corticosteriods | methylprednisolone sodium succinate |
| 74568502 | MA11 | Corticosteriods | methylprednisolone sodium succinate |
| 25021081030 | MA11 | Corticosteriods | methylprednisolone sodium succinate |
| 54569213600 | MA11 | Corticosteriods | methylprednisolone sodium succinate |
| 143985101 | MA11 | Corticosteriods | methylprednisolone sodium succinate |
| 55390021901 | MA11 | Corticosteriods | methylprednisolone sodium succinate |
| 70121100101 | MA11 | Corticosteriods | methylprednisolone sodium succinate |
| 63323025503 | MA11 | Corticosteriods | methylprednisolone sodium succinate |
| 641250641 | MA11 | Corticosteriods | methylprednisolone sodium succinate |
| 9079601 | MA11 | Corticosteriods | methylprednisolone sodium succinate |
| 70121100005 | MA11 | Corticosteriods | methylprednisolone sodium succinate |
| 54868363701 | MA11 | Corticosteriods | methylprednisolone sodium succinate |
| 9338901 | MA11 | Corticosteriods | methylprednisolone sodium succinate |
| 74563004 | MA11 | Corticosteriods | methylprednisolone sodium succinate |
| 409321705 | MA11 | Corticosteriods | methylprednisolone sodium succinate |

| | | | |
|---|---|---|---|
| 641250541 | MA11 | Corticosteriods | methylprednisolone sodium succinate |
| 55390021801 | MA11 | Corticosteriods | methylprednisolone sodium succinate |
| 70121100105 | MA11 | Corticosteriods | methylprednisolone sodium succinate |
| 55390025901 | MA11 | Corticosteriods | methylprednisolone sodium succinate |
| 54868363700 | MA11 | Corticosteriods | methylprednisolone sodium succinate |
| 143985001 | MA11 | Corticosteriods | methylprednisolone sodium succinate |
| 21695058710 | MA11 | Corticosteriods | methylprednisolone sodium succinate |
| 51927355200 | MA11 | Corticosteriods | methylprednisolone sodium succinate |
| 54868362300 | MA11 | Corticosteriods | methylprednisolone sodium succinate |
| 63323025803 | MA11 | Corticosteriods | methylprednisolone sodium succinate |
| 55390025801 | MA11 | Corticosteriods | methylprednisolone sodium succinate |
| 9076502 | MA11 | Corticosteriods | methylprednisolone sodium succinate |
| 55390020910 | MA11 | Corticosteriods | methylprednisolone sodium succinate |
| 9069802 | MA11 | Corticosteriods | methylprednisolone sodium succinate |
| 9019016 | MA11 | Corticosteriods | methylprednisolone sodium succinate |
| 54569393400 | MA11 | Corticosteriods | methylprednisolone sodium succinate/PF |
| 9004704 | MA11 | Corticosteriods | methylprednisolone sodium succinate/PF |
| 9004722 | MA11 | Corticosteriods | methylprednisolone sodium succinate/PF |
| 9003905 | MA11 | Corticosteriods | methylprednisolone sodium succinate/PF |
| 9004726 | MA11 | Corticosteriods | methylprednisolone sodium succinate/PF |
| 9000302 | MA11 | Corticosteriods | methylprednisolone sodium succinate/PF |
| 55045350901 | MA11 | Corticosteriods | methylprednisolone sodium succinate/PF |
| 9001820 | MA11 | Corticosteriods | methylprednisolone sodium succinate/PF |
| 54569393401 | MA11 | Corticosteriods | methylprednisolone sodium succinate/PF |
| 9004725 | MA11 | Corticosteriods | methylprednisolone sodium succinate/PF |
| 9003906 | MA11 | Corticosteriods | methylprednisolone sodium succinate/PF |
| 9003932 | MA11 | Corticosteriods | methylprednisolone sodium succinate/PF |
| 9003930 | MA11 | Corticosteriods | methylprednisolone sodium succinate/PF |
| 9004727 | MA11 | Corticosteriods | methylprednisolone sodium succinate/PF |
| 55154394105 | MA11 | Corticosteriods | methylprednisolone sodium succinate/PF |
| 54569155500 | MA11 | Corticosteriods | methylprednisolone sodium succinate/PF |
| 54868619900 | MA11 | Corticosteriods | methylprednisolone sodium succinate/PF |
| 9003933 | MA11 | Corticosteriods | methylprednisolone sodium succinate/PF |
| 55154394005 | MA11 | Corticosteriods | methylprednisolone sodium succinate/PF |
| 9004703 | MA11 | Corticosteriods | methylprednisolone sodium succinate/PF |
| 9003928 | MA11 | Corticosteriods | methylprednisolone sodium succinate/PF |
| 54569155501 | MA11 | Corticosteriods | methylprednisolone sodium succinate/PF |
| 49452468603 | MA11 | Corticosteriods | methylprednisolone, micronized |
| 62991290301 | MA11 | Corticosteriods | methylprednisolone, micronized |
| 51552060304 | MA11 | Corticosteriods | methylprednisolone, micronized |
| 52372818105 | MA11 | Corticosteriods | methylprednisolone, micronized |
| 38779014211 | MA11 | Corticosteriods | methylprednisolone, micronized |
| 49452468601 | MA11 | Corticosteriods | methylprednisolone, micronized |
| 38779014225 | MA11 | Corticosteriods | methylprednisolone, micronized |
| 62991290303 | MA11 | Corticosteriods | methylprednisolone, micronized |
| 51927228700 | MA11 | Corticosteriods | methylprednisolone, micronized |
| 49452468602 | MA11 | Corticosteriods | methylprednisolone, micronized |
| 62991290302 | MA11 | Corticosteriods | methylprednisolone, micronized |
| 38779014203 | MA11 | Corticosteriods | methylprednisolone, micronized |
| 38779014215 | MA11 | Corticosteriods | methylprednisolone, micronized |
| 51552060305 | MA11 | Corticosteriods | methylprednisolone, micronized |
| 52372818125 | MA11 | Corticosteriods | methylprednisolone, micronized |
| 38779014206 | MA11 | Corticosteriods | methylprednisolone, micronized |
| 51552060302 | MA11 | Corticosteriods | methylprednisolone, micronized |
| 38779014204 | MA11 | Corticosteriods | methylprednisolone, micronized |
| 17317047705 | MA11 | Corticosteriods | prednisolone |
| 49452598003 | MA11 | Corticosteriods | prednisolone |
| 451220104 | MA11 | Corticosteriods | prednisolone |
| 23490614402 | MA11 | Corticosteriods | prednisolone |
| 527120105 | MA11 | Corticosteriods | prednisolone |
| 54569401200 | MA11 | Corticosteriods | prednisolone |
| 93611816 | MA11 | Corticosteriods | prednisolone |
| 54569401201 | MA11 | Corticosteriods | prednisolone |
| 38779015015 | MA11 | Corticosteriods | prednisolone |
| 66440015016 | MA11 | Corticosteriods | prednisolone |
| 63629186201 | MA11 | Corticosteriods | prednisolone |
| 378342548 | MA11 | Corticosteriods | prednisolone |
| 472025008 | MA11 | Corticosteriods | prednisolone |
| 66440015008 | MA11 | Corticosteriods | prednisolone |
| 55175184406 | MA11 | Corticosteriods | prednisolone |
| 57866432702 | MA11 | Corticosteriods | prednisolone |

| | | | |
|---|---|---|---|
| 50383004248 | MA11 | Corticosteriods | prednisolone |
| 50090065500 | MA11 | Corticosteriods | prednisolone |
| 60432013708 | MA11 | Corticosteriods | prednisolone |
| 16477050501 | MA11 | Corticosteriods | prednisolone |
| 68071153208 | MA11 | Corticosteriods | prednisolone |
| 58177091007 | MA11 | Corticosteriods | prednisolone |
| 59196001048 | MA11 | Corticosteriods | prednisolone |
| 527120110 | MA11 | Corticosteriods | prednisolone |
| 58177091203 | MA11 | Corticosteriods | prednisolone |
| 38779015001 | MA11 | Corticosteriods | prednisolone |
| 54569480700 | MA11 | Corticosteriods | prednisolone |
| 52959062260 | MA11 | Corticosteriods | prednisolone |
| 51552002625 | MA11 | Corticosteriods | prednisolone |
| 603157058 | MA11 | Corticosteriods | prednisolone |
| 93611887 | MA11 | Corticosteriods | prednisolone |
| 54569401202 | MA11 | Corticosteriods | prednisolone |
| 527120101 | MA11 | Corticosteriods | prednisolone |
| 121068708 | MA11 | Corticosteriods | prednisolone |
| 472021208 | MA11 | Corticosteriods | prednisolone |
| 52544052038 | MA11 | Corticosteriods | prednisolone |
| 54868474900 | MA11 | Corticosteriods | prednisolone |
| 59196001024 | MA11 | Corticosteriods | prednisolone |
| 17317047708 | MA11 | Corticosteriods | prednisolone |
| 472025016 | MA11 | Corticosteriods | prednisolone |
| 364021701 | MA11 | Corticosteriods | prednisolone |
| 364021702 | MA11 | Corticosteriods | prednisolone |
| 49999033508 | MA11 | Corticosteriods | prednisolone |
| 451150016 | MA11 | Corticosteriods | prednisolone |
| 58016484301 | MA11 | Corticosteriods | prednisolone |
| 23594050521 | MA11 | Corticosteriods | prednisolone |
| 50383004224 | MA11 | Corticosteriods | prednisolone |
| 54569401203 | MA11 | Corticosteriods | prednisolone |
| 49999033524 | MA11 | Corticosteriods | prednisolone |
| 23490614503 | MA11 | Corticosteriods | prednisolone |
| 16477050521 | MA11 | Corticosteriods | prednisolone |
| 591505901 | MA11 | Corticosteriods | prednisolone |
| 38779015025 | MA11 | Corticosteriods | prednisolone |
| 54868474901 | MA11 | Corticosteriods | prednisolone |
| 38739015008 | MA11 | Corticosteriods | prednisolone |
| 55175184203 | MA11 | Corticosteriods | prednisolone |
| 60432013716 | MA11 | Corticosteriods | prednisolone |
| 54868474800 | MA11 | Corticosteriods | prednisolone |
| 49452598001 | MA11 | Corticosteriods | prednisolone |
| 67870010316 | MA11 | Corticosteriods | prednisolone |
| 68115089808 | MA11 | Corticosteriods | prednisolone |
| 49452598002 | MA11 | Corticosteriods | prednisolone |
| 55289095204 | MA11 | Corticosteriods | prednisolone |
| 66116017482 | MA11 | Corticosteriods | prednisolone |
| 33358029108 | MA11 | Corticosteriods | prednisolone |
| 67870010308 | MA11 | Corticosteriods | prednisolone |
| 49999092901 | MA11 | Corticosteriods | prednisolone |
| 54569401204 | MA11 | Corticosteriods | prednisolone |
| 23490614403 | MA11 | Corticosteriods | prednisolone |
| 378342524 | MA11 | Corticosteriods | prednisolone |
| 23490614502 | MA11 | Corticosteriods | prednisolone |
| 55045182502 | MA11 | Corticosteriods | prednisolone |
| 23490614501 | MA11 | Corticosteriods | prednisolone |
| 68258898702 | MA11 | Corticosteriods | prednisolone |
| 23490614401 | MA11 | Corticosteriods | prednisolone |
| 23594050548 | MA11 | Corticosteriods | prednisolone |
| 591052038 | MA11 | Corticosteriods | prednisolone |
| 68094052458 | MA11 | Corticosteriods | prednisolone |
| 55289095202 | MA11 | Corticosteriods | prednisolone |
| 51552002699 | MA11 | Corticosteriods | prednisolone |
| 52544052016 | MA11 | Corticosteriods | prednisolone |
| 54569482701 | MA11 | Corticosteriods | prednisolone |
| 16477050748 | MA11 | Corticosteriods | prednisolone |
| 121068705 | MA11 | Corticosteriods | prednisolone |
| 472021216 | MA11 | Corticosteriods | prednisolone |
| 603157056 | MA11 | Corticosteriods | prednisolone |
| 21695036516 | MA11 | Corticosteriods | prednisolone |

| | | | |
|---|---|---|---|
| 23594050501 | MA11 | Corticosteriods | prednisolone |
| 57866432701 | MA11 | Corticosteriods | prednisolone |
| 16477050548 | MA11 | Corticosteriods | prednisolone |
| 58177091005 | MA11 | Corticosteriods | prednisolone |
| 17317047701 | MA11 | Corticosteriods | prednisolone |
| 879080108 | MA11 | Corticosteriods | prednisolone |
| 591505910 | MA11 | Corticosteriods | prednisolone |
| 879080116 | MA11 | Corticosteriods | prednisolone |
| 54868322000 | MA11 | Corticosteriods | prednisolone |
| 603156756 | MA11 | Corticosteriods | prednisolone |
| 51927114800 | MA11 | Corticosteriods | prednisolone |
| 54569482700 | MA11 | Corticosteriods | prednisolone |
| 23594050550 | MA11 | Corticosteriods | prednisolone |
| 23594050502 | MA11 | Corticosteriods | prednisolone |
| 451150008 | MA11 | Corticosteriods | prednisolone |
| 21695036508 | MA11 | Corticosteriods | prednisolone |
| 603156758 | MA11 | Corticosteriods | prednisolone |
| 38779015010 | MA11 | Corticosteriods | prednisolone |
| 121068716 | MA11 | Corticosteriods | prednisolone |
| 55175184206 | MA11 | Corticosteriods | prednisolone |
| 684011510 | MA11 | Corticosteriods | prednisolone acetate |
| 49452599001 | MA11 | Corticosteriods | prednisolone acetate |
| 785801510 | MA11 | Corticosteriods | prednisolone acetate |
| 314069530 | MA11 | Corticosteriods | prednisolone acetate |
| 463102010 | MA11 | Corticosteriods | prednisolone acetate |
| 49452599002 | MA11 | Corticosteriods | prednisolone acetate |
| 51672133803 | MA11 | Corticosteriods | prednisolone acetate |
| 217840408 | MA11 | Corticosteriods | prednisolone acetate |
| 588535270 | MA11 | Corticosteriods | prednisolone acetate |
| 51552002725 | MA11 | Corticosteriods | prednisolone acetate |
| 17317089008 | MA11 | Corticosteriods | prednisolone acetate |
| 49452599003 | MA11 | Corticosteriods | prednisolone acetate |
| 182093966 | MA11 | Corticosteriods | prednisolone acetate |
| 54569216100 | MA11 | Corticosteriods | prednisolone acetate |
| 463101930 | MA11 | Corticosteriods | prednisolone acetate |
| 17317089001 | MA11 | Corticosteriods | prednisolone acetate |
| 17317089005 | MA11 | Corticosteriods | prednisolone acetate |
| 314069770 | MA11 | Corticosteriods | prednisolone acetate |
| 51552002799 | MA11 | Corticosteriods | prednisolone acetate |
| 38779015203 | MA11 | Corticosteriods | prednisolone acetate, micronized |
| 38779015210 | MA11 | Corticosteriods | prednisolone acetate, micronized |
| 38779015208 | MA11 | Corticosteriods | prednisolone acetate, micronized |
| 62991169201 | MA11 | Corticosteriods | prednisolone acetate, micronized |
| 62991169202 | MA11 | Corticosteriods | prednisolone acetate, micronized |
| 38779015215 | MA11 | Corticosteriods | prednisolone acetate, micronized |
| 38779015209 | MA11 | Corticosteriods | prednisolone acetate, micronized |
| 38779015201 | MA11 | Corticosteriods | prednisolone acetate, micronized |
| 38779015205 | MA11 | Corticosteriods | prednisolone acetate, micronized |
| 38779015204 | MA11 | Corticosteriods | prednisolone acetate, micronized |
| 51927132500 | MA11 | Corticosteriods | prednisolone acetate, micronized |
| 38779015225 | MA11 | Corticosteriods | prednisolone acetate, micronized |
| 38779256205 | MA11 | Corticosteriods | prednisolone acetate, micronized |
| 62991169203 | MA11 | Corticosteriods | prednisolone acetate, micronized |
| 49452599004 | MA11 | Corticosteriods | prednisolone acetate, micronized |
| 51552002702 | MA11 | Corticosteriods | prednisolone acetate, micronized |
| 51552002705 | MA11 | Corticosteriods | prednisolone acetate, micronized |
| 51552002704 | MA11 | Corticosteriods | prednisolone acetate, micronized |
| 121077708 | MA11 | Corticosteriods | prednisolone sodium phosphate |
| 58016414401 | MA11 | Corticosteriods | prednisolone sodium phosphate |
| 54868172000 | MA11 | Corticosteriods | prednisolone sodium phosphate |
| 62991120506 | MA11 | Corticosteriods | prednisolone sodium phosphate |
| 59212070048 | MA11 | Corticosteriods | prednisolone sodium phosphate |
| 62991120502 | MA11 | Corticosteriods | prednisolone sodium phosphate |
| 70166054905 | MA11 | Corticosteriods | prednisolone sodium phosphate |
| 69101041001 | MA11 | Corticosteriods | prednisolone sodium phosphate |
| 54569520401 | MA11 | Corticosteriods | prednisolone sodium phosphate |
| 68188048202 | MA11 | Corticosteriods | prednisolone sodium phosphate |
| 59630070014 | MA11 | Corticosteriods | prednisolone sodium phosphate |
| 59196001224 | MA11 | Corticosteriods | prednisolone sodium phosphate |
| 50090095500 | MA11 | Corticosteriods | prednisolone sodium phosphate |
| 51552073505 | MA11 | Corticosteriods | prednisolone sodium phosphate |

| | | | |
|---|---|---|---|
| 65580025101 | MA11 | Corticosteriods | prednisolone sodium phosphate |
| 38779015303 | MA11 | Corticosteriods | prednisolone sodium phosphate |
| 59439045502 | MA11 | Corticosteriods | prednisolone sodium phosphate |
| 16477051008 | MA11 | Corticosteriods | prednisolone sodium phosphate |
| 121077308 | MA11 | Corticosteriods | prednisolone sodium phosphate |
| 68791010004 | MA11 | Corticosteriods | prednisolone sodium phosphate |
| 66993084551 | MA11 | Corticosteriods | prednisolone sodium phosphate |
| 23594051008 | MA11 | Corticosteriods | prednisolone sodium phosphate |
| 68791010404 | MA11 | Corticosteriods | prednisolone sodium phosphate |
| 66993084651 | MA11 | Corticosteriods | prednisolone sodium phosphate |
| 62991120504 | MA11 | Corticosteriods | prednisolone sodium phosphate |
| 17856075905 | MA11 | Corticosteriods | prednisolone sodium phosphate |
| 68135045502 | MA11 | Corticosteriods | prednisolone sodium phosphate |
| 58177093205 | MA11 | Corticosteriods | prednisolone sodium phosphate |
| 51552073502 | MA11 | Corticosteriods | prednisolone sodium phosphate |
| 54868524200 | MA11 | Corticosteriods | prednisolone sodium phosphate |
| 62991120503 | MA11 | Corticosteriods | prednisolone sodium phosphate |
| 50383017708 | MA11 | Corticosteriods | prednisolone sodium phosphate |
| 38779015301 | MA11 | Corticosteriods | prednisolone sodium phosphate |
| 38779015305 | MA11 | Corticosteriods | prednisolone sodium phosphate |
| 53014025001 | MA11 | Corticosteriods | prednisolone sodium phosphate |
| 178058201 | MA11 | Corticosteriods | prednisolone sodium phosphate |
| 65162066788 | MA11 | Corticosteriods | prednisolone sodium phosphate |
| 59196001104 | MA11 | Corticosteriods | prednisolone sodium phosphate |
| 68188048402 | MA11 | Corticosteriods | prednisolone sodium phosphate |
| 121071120 | MA11 | Corticosteriods | prednisolone sodium phosphate |
| 378471022 | MA11 | Corticosteriods | prednisolone sodium phosphate |
| 121071105 | MA11 | Corticosteriods | prednisolone sodium phosphate |
| 65162066790 | MA11 | Corticosteriods | prednisolone sodium phosphate |
| 68188048002 | MA11 | Corticosteriods | prednisolone sodium phosphate |
| 13925050104 | MA11 | Corticosteriods | prednisolone sodium phosphate |
| 178058208 | MA11 | Corticosteriods | prednisolone sodium phosphate |
| 574014804 | MA11 | Corticosteriods | prednisolone sodium phosphate |
| 378471522 | MA11 | Corticosteriods | prednisolone sodium phosphate |
| 38779015315 | MA11 | Corticosteriods | prednisolone sodium phosphate |
| 121071110 | MA11 | Corticosteriods | prednisolone sodium phosphate |
| 42254010208 | MA11 | Corticosteriods | prednisolone sodium phosphate |
| 59630070248 | MA11 | Corticosteriods | prednisolone sodium phosphate |
| 54569647800 | MA11 | Corticosteriods | prednisolone sodium phosphate |
| 403161118 | MA11 | Corticosteriods | prednisolone sodium phosphate |
| 38779015304 | MA11 | Corticosteriods | prednisolone sodium phosphate |
| 121075908 | MA11 | Corticosteriods | prednisolone sodium phosphate |
| 603938856 | MA11 | Corticosteriods | prednisolone sodium phosphate |
| 21695040508 | MA11 | Corticosteriods | prednisolone sodium phosphate |
| 38779015308 | MA11 | Corticosteriods | prednisolone sodium phosphate |
| 51552073504 | MA11 | Corticosteriods | prednisolone sodium phosphate |
| 59630070148 | MA11 | Corticosteriods | prednisolone sodium phosphate |
| 51927446500 | MA11 | Corticosteriods | prednisolone sodium phosphate |
| 35356048804 | MA11 | Corticosteriods | prednisolone sodium phosphate |
| 55045288508 | MA11 | Corticosteriods | prednisolone sodium phosphate |
| 59630071010 | MA11 | Corticosteriods | prednisolone sodium phosphate |
| 23594091508 | MA11 | Corticosteriods | prednisolone sodium phosphate |
| 121071104 | MA11 | Corticosteriods | prednisolone sodium phosphate |
| 13925016604 | MA11 | Corticosteriods | prednisolone sodium phosphate |
| 55175400401 | MA11 | Corticosteriods | prednisolone sodium phosphate |
| 59212070148 | MA11 | Corticosteriods | prednisolone sodium phosphate |
| 54569520400 | MA11 | Corticosteriods | prednisolone sodium phosphate |
| 54569574900 | MA11 | Corticosteriods | prednisolone sodium phosphate |
| 59630070048 | MA11 | Corticosteriods | prednisolone sodium phosphate |
| 59630070114 | MA11 | Corticosteriods | prednisolone sodium phosphate |
| 68135045503 | MA11 | Corticosteriods | prednisolone sodium phosphate |
| 60432021208 | MA11 | Corticosteriods | prednisolone sodium phosphate |
| 38779015310 | MA11 | Corticosteriods | prednisolone sodium phosphate |
| 42799081301 | MA11 | Corticosteriods | prednisolone sodium phosphate |
| 54868095400 | MA11 | Corticosteriods | prednisolone sodium phosphate |
| 60432008904 | MA11 | Corticosteriods | prednisolone sodium phosphate |
| 66993084451 | MA11 | Corticosteriods | prednisolone sodium phosphate |
| 54569574901 | MA11 | Corticosteriods | prednisolone sodium phosphate |
| 50383004004 | MA11 | Corticosteriods | prednisolone sodium phosphate |
| 59630070214 | MA11 | Corticosteriods | prednisolone sodium phosphate |
| 66993084462 | MA11 | Corticosteriods | prednisolone sodium phosphate |

| | | | |
|---|---|---|---|
| 59212070248 | MA11 | Corticosteriods | prednisolone sodium phosphate |
| 66993084562 | MA11 | Corticosteriods | prednisolone sodium phosphate |
| 55045288500 | MA11 | Corticosteriods | prednisolone sodium phosphate |
| 54569133500 | MA11 | Corticosteriods | prednisolone sodium phosphate |
| 54868082100 | MA11 | Corticosteriods | prednisolone sodium phosphate |
| 49452599501 | MA11 | Corticosteriods | prednisolone sodium phosphate |
| 42799081201 | MA11 | Corticosteriods | prednisolone sodium phosphate |
| 44523018208 | MA11 | Corticosteriods | prednisolone sodium phosphate |
| 51927155000 | MA11 | Corticosteriods | prednisolone sodium phosphate |
| 59630071008 | MA11 | Corticosteriods | prednisolone sodium phosphate |
| 55289051102 | MA11 | Corticosteriods | prednisolone sodium phosphate |
| 63717091508 | MA11 | Corticosteriods | prednisolone sodium phosphate |
| 38779015325 | MA11 | Corticosteriods | prednisolone sodium phosphate |
| 378473022 | MA11 | Corticosteriods | prednisolone sodium phosphate |
| 71384055203 | MA11 | Corticosteriods | prednisolone sodium phosphate/bromfenac sodium |
| 23589007093 | MA11 | Corticosteriods | Prednisolone Sodium Phosphate/Peak Flow Meter |
| 23589006793 | MA11 | Corticosteriods | Prednisolone Sodium Phosphate/Peak Flow Meter |
| 62991125701 | MA11 | Corticosteriods | prednisolone, micronized |
| 62991125702 | MA11 | Corticosteriods | prednisolone, micronized |
| 51552002605 | MA11 | Corticosteriods | prednisolone, micronized |
| 38779015009 | MA11 | Corticosteriods | prednisolone, micronized |
| 38779015005 | MA11 | Corticosteriods | prednisolone, micronized |
| 38779015003 | MA11 | Corticosteriods | prednisolone, micronized |
| 51552002604 | MA11 | Corticosteriods | prednisolone, micronized |
| 38779015008 | MA11 | Corticosteriods | prednisolone, micronized |
| 51552002602 | MA11 | Corticosteriods | prednisolone, micronized |
| 38779015004 | MA11 | Corticosteriods | prednisolone, micronized |
| 62991125704 | MA11 | Corticosteriods | prednisolone, micronized |
| 247007220 | MA11 | Corticosteriods | prednisone |
| 60346009412 | MA11 | Corticosteriods | prednisone |
| 42254036136 | MA11 | Corticosteriods | prednisone |
| 61919040421 | MA11 | Corticosteriods | prednisone |
| 54868090801 | MA11 | Corticosteriods | prednisone |
| 58016021612 | MA11 | Corticosteriods | prednisone |
| 35356081920 | MA11 | Corticosteriods | prednisone |
| 143142501 | MA11 | Corticosteriods | prednisone |
| 43063064421 | MA11 | Corticosteriods | prednisone |
| 51138015615 | MA11 | Corticosteriods | prednisone |
| 58016021632 | MA11 | Corticosteriods | prednisone |
| 43063059014 | MA11 | Corticosteriods | prednisone |
| 58864036220 | MA11 | Corticosteriods | prednisone |
| 71335065101 | MA11 | Corticosteriods | prednisone |
| 51138015610 | MA11 | Corticosteriods | prednisone |
| 43063070315 | MA11 | Corticosteriods | prednisone |
| 49999000820 | MA11 | Corticosteriods | prednisone |
| 57866432406 | MA11 | Corticosteriods | prednisone |
| 52959012630 | MA11 | Corticosteriods | prednisone |
| 57866432605 | MA11 | Corticosteriods | prednisone |
| 53489013905 | MA11 | Corticosteriods | prednisone |
| 23490615902 | MA11 | Corticosteriods | prednisone |
| 52959022040 | MA11 | Corticosteriods | prednisone |
| 21695030707 | MA11 | Corticosteriods | prednisone |
| 23490615803 | MA11 | Corticosteriods | prednisone |
| 55289037321 | MA11 | Corticosteriods | prednisone |
| 143974010 | MA11 | Corticosteriods | prednisone |
| 43063070321 | MA11 | Corticosteriods | prednisone |
| 54868083607 | MA11 | Corticosteriods | prednisone |
| 43063042650 | MA11 | Corticosteriods | prednisone |
| 378064110 | MA11 | Corticosteriods | prednisone |
| 53489013810 | MA11 | Corticosteriods | prednisone |
| 35356081814 | MA11 | Corticosteriods | prednisone |
| 527293537 | MA11 | Corticosteriods | prednisone |
| 55700007230 | MA11 | Corticosteriods | prednisone |
| 63874032742 | MA11 | Corticosteriods | prednisone |
| 440816530 | MA11 | Corticosteriods | prednisone |
| 591544243 | MA11 | Corticosteriods | prednisone |
| 247010018 | MA11 | Corticosteriods | prednisone |
| 55045148005 | MA11 | Corticosteriods | prednisone |
| 50090199007 | MA11 | Corticosteriods | prednisone |
| 45802073367 | MA11 | Corticosteriods | prednisone |
| 60429013001 | MA11 | Corticosteriods | prednisone |

| | | | |
|---|---|---|---|
| 23490615702 | MA11 | Corticosteriods | prednisone |
| 143147310 | MA11 | Corticosteriods | prednisone |
| 60429013010 | MA11 | Corticosteriods | prednisone |
| 58016021690 | MA11 | Corticosteriods | prednisone |
| 58016021800 | MA11 | Corticosteriods | prednisone |
| 51138014520 | MA11 | Corticosteriods | prednisone |
| 23490615901 | MA11 | Corticosteriods | prednisone |
| 440816430 | MA11 | Corticosteriods | prednisone |
| 54569330202 | MA11 | Corticosteriods | prednisone |
| 55289035215 | MA11 | Corticosteriods | prednisone |
| 54868083600 | MA11 | Corticosteriods | prednisone |
| 42254024021 | MA11 | Corticosteriods | prednisone |
| 55045180902 | MA11 | Corticosteriods | prednisone |
| 55289043801 | MA11 | Corticosteriods | prednisone |
| 58016021716 | MA11 | Corticosteriods | prednisone |
| 58864042414 | MA11 | Corticosteriods | prednisone |
| 63874037340 | MA11 | Corticosteriods | prednisone |
| 54569033003 | MA11 | Corticosteriods | prednisone |
| 21695030710 | MA11 | Corticosteriods | prednisone |
| 54569033201 | MA11 | Corticosteriods | prednisone |
| 66336005821 | MA11 | Corticosteriods | prednisone |
| 55289085902 | MA11 | Corticosteriods | prednisone |
| 70934009620 | MA11 | Corticosteriods | prednisone |
| 247007150 | MA11 | Corticosteriods | prednisone |
| 60904028620 | MA11 | Corticosteriods | prednisone |
| 55289035207 | MA11 | Corticosteriods | prednisone |
| 68030432602 | MA11 | Corticosteriods | prednisone |
| 63739058810 | MA11 | Corticosteriods | prednisone |
| 50090199000 | MA11 | Corticosteriods | prednisone |
| 677069810 | MA11 | Corticosteriods | prednisone |
| 70954005830 | MA11 | Corticosteriods | prednisone |
| 58864042430 | MA11 | Corticosteriods | prednisone |
| 67544039960 | MA11 | Corticosteriods | prednisone |
| 23490615800 | MA11 | Corticosteriods | prednisone |
| 54569033007 | MA11 | Corticosteriods | prednisone |
| 440216512 | MA11 | Corticosteriods | prednisone |
| 60429013201 | MA11 | Corticosteriods | prednisone |
| 54001720 | MA11 | Corticosteriods | prednisone |
| 52959012721 | MA11 | Corticosteriods | prednisone |
| 55887064330 | MA11 | Corticosteriods | prednisone |
| 55700020915 | MA11 | Corticosteriods | prednisone |
| 33358029330 | MA11 | Corticosteriods | prednisone |
| 143973910 | MA11 | Corticosteriods | prednisone |
| 23490615801 | MA11 | Corticosteriods | prednisone |
| 49999000840 | MA11 | Corticosteriods | prednisone |
| 54868090805 | MA11 | Corticosteriods | prednisone |
| 55045296301 | MA11 | Corticosteriods | prednisone |
| 54868409600 | MA11 | Corticosteriods | prednisone |
| 259039021 | MA11 | Corticosteriods | prednisone |
| 54868118306 | MA11 | Corticosteriods | prednisone |
| 51138014715 | MA11 | Corticosteriods | prednisone |
| 63739020915 | MA11 | Corticosteriods | prednisone |
| 63874032715 | MA11 | Corticosteriods | prednisone |
| 364046101 | MA11 | Corticosteriods | prednisone |
| 403077030 | MA11 | Corticosteriods | prednisone |
| 55289043815 | MA11 | Corticosteriods | prednisone |
| 143147301 | MA11 | Corticosteriods | prednisone |
| 43063043221 | MA11 | Corticosteriods | prednisone |
| 62584083201 | MA11 | Corticosteriods | prednisone |
| 50090199100 | MA11 | Corticosteriods | prednisone |
| 51079003319 | MA11 | Corticosteriods | prednisone |
| 615053629 | MA11 | Corticosteriods | prednisone |
| 35356081940 | MA11 | Corticosteriods | prednisone |
| 50090335401 | MA11 | Corticosteriods | prednisone |
| 51138015515 | MA11 | Corticosteriods | prednisone |
| 17317066300 | MA11 | Corticosteriods | prednisone |
| 55289037346 | MA11 | Corticosteriods | prednisone |
| 55289043820 | MA11 | Corticosteriods | prednisone |
| 54868083609 | MA11 | Corticosteriods | prednisone |
| 247010027 | MA11 | Corticosteriods | prednisone |
| 615359343 | MA11 | Corticosteriods | prednisone |

| | | | |
|---|---|---|---|
| 51138015520 | MA11 | Corticosteriods | prednisone |
| 43063042620 | MA11 | Corticosteriods | prednisone |
| 63874037320 | MA11 | Corticosteriods | prednisone |
| 49999002815 | MA11 | Corticosteriods | prednisone |
| 51138015620 | MA11 | Corticosteriods | prednisone |
| 66267017360 | MA11 | Corticosteriods | prednisone |
| 55175392001 | MA11 | Corticosteriods | prednisone |
| 615154243 | MA11 | Corticosteriods | prednisone |
| 904214180 | MA11 | Corticosteriods | prednisone |
| 403077050 | MA11 | Corticosteriods | prednisone |
| 55700020820 | MA11 | Corticosteriods | prednisone |
| 33358029430 | MA11 | Corticosteriods | prednisone |
| 52959012600 | MA11 | Corticosteriods | prednisone |
| 70954006020 | MA11 | Corticosteriods | prednisone |
| 378064101 | MA11 | Corticosteriods | prednisone |
| 63187099721 | MA11 | Corticosteriods | prednisone |
| 42254036145 | MA11 | Corticosteriods | prednisone |
| 54569304302 | MA11 | Corticosteriods | prednisone |
| 66267017215 | MA11 | Corticosteriods | prednisone |
| 364021802 | MA11 | Corticosteriods | prednisone |
| 71335065105 | MA11 | Corticosteriods | prednisone |
| 364046102 | MA11 | Corticosteriods | prednisone |
| 66336051540 | MA11 | Corticosteriods | prednisone |
| 247007224 | MA11 | Corticosteriods | prednisone |
| 55175399204 | MA11 | Corticosteriods | prednisone |
| 63187003720 | MA11 | Corticosteriods | prednisone |
| 17317066301 | MA11 | Corticosteriods | prednisone |
| 63739020715 | MA11 | Corticosteriods | prednisone |
| 364044290 | MA11 | Corticosteriods | prednisone |
| 59746017310 | MA11 | Corticosteriods | prednisone |
| 527293137 | MA11 | Corticosteriods | prednisone |
| 63629160503 | MA11 | Corticosteriods | prednisone |
| 42254036115 | MA11 | Corticosteriods | prednisone |
| 69668012020 | MA11 | Corticosteriods | prednisone |
| 35356067420 | MA11 | Corticosteriods | prednisone |
| 51138015430 | MA11 | Corticosteriods | prednisone |
| 54868083601 | MA11 | Corticosteriods | prednisone |
| 591544205 | MA11 | Corticosteriods | prednisone |
| 23490615905 | MA11 | Corticosteriods | prednisone |
| 603533215 | MA11 | Corticosteriods | prednisone |
| 18837026748 | MA11 | Corticosteriods | prednisone |
| 23490615805 | MA11 | Corticosteriods | prednisone |
| 51655002024 | MA11 | Corticosteriods | prednisone |
| 55289035210 | MA11 | Corticosteriods | prednisone |
| 603533231 | MA11 | Corticosteriods | prednisone |
| 33358029460 | MA11 | Corticosteriods | prednisone |
| 18837035330 | MA11 | Corticosteriods | prednisone |
| 52959012725 | MA11 | Corticosteriods | prednisone |
| 50090240304 | MA11 | Corticosteriods | prednisone |
| 66336009421 | MA11 | Corticosteriods | prednisone |
| 43063041501 | MA11 | Corticosteriods | prednisone |
| 23490615707 | MA11 | Corticosteriods | prednisone |
| 440816521 | MA11 | Corticosteriods | prednisone |
| 54569402601 | MA11 | Corticosteriods | prednisone |
| 57866432608 | MA11 | Corticosteriods | prednisone |
| 55887064342 | MA11 | Corticosteriods | prednisone |
| 21695030705 | MA11 | Corticosteriods | prednisone |
| 54569304304 | MA11 | Corticosteriods | prednisone |
| 440816715 | MA11 | Corticosteriods | prednisone |
| 58016021720 | MA11 | Corticosteriods | prednisone |
| 60760013830 | MA11 | Corticosteriods | prednisone |
| 63874037360 | MA11 | Corticosteriods | prednisone |
| 66336009420 | MA11 | Corticosteriods | prednisone |
| 35356067721 | MA11 | Corticosteriods | prednisone |
| 43063059030 | MA11 | Corticosteriods | prednisone |
| 63187080721 | MA11 | Corticosteriods | prednisone |
| 603533421 | MA11 | Corticosteriods | prednisone |
| 49999002848 | MA11 | Corticosteriods | prednisone |
| 54868118300 | MA11 | Corticosteriods | prednisone |
| 66267086004 | MA11 | Corticosteriods | prednisone |
| 43063086660 | MA11 | Corticosteriods | prednisone |

| | | | |
|---|---|---|---|
| 247010014 | MA11 | Corticosteriods | prednisone |
| 70934009615 | MA11 | Corticosteriods | prednisone |
| 63739051910 | MA11 | Corticosteriods | prednisone |
| 63874037310 | MA11 | Corticosteriods | prednisone |
| 259038948 | MA11 | Corticosteriods | prednisone |
| 63739020701 | MA11 | Corticosteriods | prednisone |
| 16590037328 | MA11 | Corticosteriods | prednisone |
| 55175415808 | MA11 | Corticosteriods | prednisone |
| 50090199001 | MA11 | Corticosteriods | prednisone |
| 55289037336 | MA11 | Corticosteriods | prednisone |
| 9004501 | MA11 | Corticosteriods | prednisone |
| 66267017121 | MA11 | Corticosteriods | prednisone |
| 63874032720 | MA11 | Corticosteriods | prednisone |
| 58016021624 | MA11 | Corticosteriods | prednisone |
| 51079002220 | MA11 | Corticosteriods | prednisone |
| 54569033106 | MA11 | Corticosteriods | prednisone |
| 66267017212 | MA11 | Corticosteriods | prednisone |
| 63874032714 | MA11 | Corticosteriods | prednisone |
| 63874039214 | MA11 | Corticosteriods | prednisone |
| 71205008900 | MA11 | Corticosteriods | prednisone |
| 61919034210 | MA11 | Corticosteriods | prednisone |
| 55175276601 | MA11 | Corticosteriods | prednisone |
| 58016021722 | MA11 | Corticosteriods | prednisone |
| 50090010200 | MA11 | Corticosteriods | prednisone |
| 55700020930 | MA11 | Corticosteriods | prednisone |
| 70954006010 | MA11 | Corticosteriods | prednisone |
| 603533521 | MA11 | Corticosteriods | prednisone |
| 35356067740 | MA11 | Corticosteriods | prednisone |
| 63187030042 | MA11 | Corticosteriods | prednisone |
| 63739020803 | MA11 | Corticosteriods | prednisone |
| 45802073321 | MA11 | Corticosteriods | prednisone |
| 49999011018 | MA11 | Corticosteriods | prednisone |
| 43063047225 | MA11 | Corticosteriods | prednisone |
| 60346051536 | MA11 | Corticosteriods | prednisone |
| 63874037315 | MA11 | Corticosteriods | prednisone |
| 55045144404 | MA11 | Corticosteriods | prednisone |
| 63739020702 | MA11 | Corticosteriods | prednisone |
| 75987002001 | MA11 | Corticosteriods | prednisone |
| 54001725 | MA11 | Corticosteriods | prednisone |
| 53489014001 | MA11 | Corticosteriods | prednisone |
| 603533321 | MA11 | Corticosteriods | prednisone |
| 54868111902 | MA11 | Corticosteriods | prednisone |
| 42254036140 | MA11 | Corticosteriods | prednisone |
| 51138014621 | MA11 | Corticosteriods | prednisone |
| 60346051560 | MA11 | Corticosteriods | prednisone |
| 51552002804 | MA11 | Corticosteriods | prednisone |
| 247007143 | MA11 | Corticosteriods | prednisone |
| 60346005820 | MA11 | Corticosteriods | prednisone |
| 58016021628 | MA11 | Corticosteriods | prednisone |
| 51138015614 | MA11 | Corticosteriods | prednisone |
| 55289033010 | MA11 | Corticosteriods | prednisone |
| 54868118307 | MA11 | Corticosteriods | prednisone |
| 904215780 | MA11 | Corticosteriods | prednisone |
| 9016501 | MA11 | Corticosteriods | prednisone |
| 247010015 | MA11 | Corticosteriods | prednisone |
| 49999011010 | MA11 | Corticosteriods | prednisone |
| 55045144403 | MA11 | Corticosteriods | prednisone |
| 35356067320 | MA11 | Corticosteriods | prednisone |
| 57866432501 | MA11 | Corticosteriods | prednisone |
| 63187030025 | MA11 | Corticosteriods | prednisone |
| 440816620 | MA11 | Corticosteriods | prednisone |
| 66336009412 | MA11 | Corticosteriods | prednisone |
| 21695030706 | MA11 | Corticosteriods | prednisone |
| 58016021620 | MA11 | Corticosteriods | prednisone |
| 60346005827 | MA11 | Corticosteriods | prednisone |
| 60687014511 | MA11 | Corticosteriods | prednisone |
| 52959012730 | MA11 | Corticosteriods | prednisone |
| 43063043215 | MA11 | Corticosteriods | prednisone |
| 55289043860 | MA11 | Corticosteriods | prednisone |
| 21695030521 | MA11 | Corticosteriods | prednisone |
| 63187024305 | MA11 | Corticosteriods | prednisone |

| | | | |
|---|---|---|---|
| 247007254 | MA11 | Corticosteriods | prednisone |
| 55175399201 | MA11 | Corticosteriods | prednisone |
| 55289043842 | MA11 | Corticosteriods | prednisone |
| 68115029020 | MA11 | Corticosteriods | prednisone |
| 18837035310 | MA11 | Corticosteriods | prednisone |
| 904214060 | MA11 | Corticosteriods | prednisone |
| 54868323400 | MA11 | Corticosteriods | prednisone |
| 63739020703 | MA11 | Corticosteriods | prednisone |
| 52959012742 | MA11 | Corticosteriods | prednisone |
| 52959012650 | MA11 | Corticosteriods | prednisone |
| 63629157900 | MA11 | Corticosteriods | prednisone |
| 54569033204 | MA11 | Corticosteriods | prednisone |
| 57866432602 | MA11 | Corticosteriods | prednisone |
| 60760017935 | MA11 | Corticosteriods | prednisone |
| 35356067415 | MA11 | Corticosteriods | prednisone |
| 10768728304 | MA11 | Corticosteriods | prednisone |
| 51991046205 | MA11 | Corticosteriods | prednisone |
| 33358029230 | MA11 | Corticosteriods | prednisone |
| 63187030021 | MA11 | Corticosteriods | prednisone |
| 63874037321 | MA11 | Corticosteriods | prednisone |
| 63739020710 | MA11 | Corticosteriods | prednisone |
| 66336005820 | MA11 | Corticosteriods | prednisone |
| 55045153303 | MA11 | Corticosteriods | prednisone |
| 70954005910 | MA11 | Corticosteriods | prednisone |
| 50090336106 | MA11 | Corticosteriods | prednisone |
| 52959012605 | MA11 | Corticosteriods | prednisone |
| 52959012621 | MA11 | Corticosteriods | prednisone |
| 66116042015 | MA11 | Corticosteriods | prednisone |
| 55700004921 | MA11 | Corticosteriods | prednisone |
| 51991045801 | MA11 | Corticosteriods | prednisone |
| 63874032702 | MA11 | Corticosteriods | prednisone |
| 43063086621 | MA11 | Corticosteriods | prednisone |
| 70954005810 | MA11 | Corticosteriods | prednisone |
| 55289043821 | MA11 | Corticosteriods | prednisone |
| 45802030367 | MA11 | Corticosteriods | prednisone |
| 54868409500 | MA11 | Corticosteriods | prednisone |
| 54868118309 | MA11 | Corticosteriods | prednisone |
| 66267017320 | MA11 | Corticosteriods | prednisone |
| 23490615808 | MA11 | Corticosteriods | prednisone |
| 61919032630 | MA11 | Corticosteriods | prednisone |
| 55289037355 | MA11 | Corticosteriods | prednisone |
| 55175391804 | MA11 | Corticosteriods | prednisone |
| 43063009706 | MA11 | Corticosteriods | prednisone |
| 55289035209 | MA11 | Corticosteriods | prednisone |
| 57866432405 | MA11 | Corticosteriods | prednisone |
| 21695030530 | MA11 | Corticosteriods | prednisone |
| 54868025805 | MA11 | Corticosteriods | prednisone |
| 51079002201 | MA11 | Corticosteriods | prednisone |
| 63874037350 | MA11 | Corticosteriods | prednisone |
| 615154263 | MA11 | Corticosteriods | prednisone |
| 49999002828 | MA11 | Corticosteriods | prednisone |
| 403077401 | MA11 | Corticosteriods | prednisone |
| 54569379800 | MA11 | Corticosteriods | prednisone |
| 71335065104 | MA11 | Corticosteriods | prednisone |
| 55700020921 | MA11 | Corticosteriods | prednisone |
| 403196718 | MA11 | Corticosteriods | prednisone |
| 16590032630 | MA11 | Corticosteriods | prednisone |
| 58016021650 | MA11 | Corticosteriods | prednisone |
| 440816612 | MA11 | Corticosteriods | prednisone |
| 50090199005 | MA11 | Corticosteriods | prednisone |
| 54868025803 | MA11 | Corticosteriods | prednisone |
| 60760025540 | MA11 | Corticosteriods | prednisone |
| 63874032750 | MA11 | Corticosteriods | prednisone |
| 247010013 | MA11 | Corticosteriods | prednisone |
| 54868118303 | MA11 | Corticosteriods | prednisone |
| 54474131 | MA11 | Corticosteriods | prednisone |
| 42254036121 | MA11 | Corticosteriods | prednisone |
| 440816590 | MA11 | Corticosteriods | prednisone |
| 54569422700 | MA11 | Corticosteriods | prednisone |
| 70934009520 | MA11 | Corticosteriods | prednisone |
| 603533715 | MA11 | Corticosteriods | prednisone |

| | | | |
|---|---|---|---|
| 63629160502 | MA11 | Corticosteriods | prednisone |
| 55045144401 | MA11 | Corticosteriods | prednisone |
| 677011701 | MA11 | Corticosteriods | prednisone |
| 55700020942 | MA11 | Corticosteriods | prednisone |
| 75987002101 | MA11 | Corticosteriods | prednisone |
| 591505243 | MA11 | Corticosteriods | prednisone |
| 16590040448 | MA11 | Corticosteriods | prednisone |
| 66336009430 | MA11 | Corticosteriods | prednisone |
| 403077042 | MA11 | Corticosteriods | prednisone |
| 63874037333 | MA11 | Corticosteriods | prednisone |
| 49452600003 | MA11 | Corticosteriods | prednisone |
| 66336009418 | MA11 | Corticosteriods | prednisone |
| 615359353 | MA11 | Corticosteriods | prednisone |
| 403077214 | MA11 | Corticosteriods | prednisone |
| 54569033209 | MA11 | Corticosteriods | prednisone |
| 68115028921 | MA11 | Corticosteriods | prednisone |
| 50090198909 | MA11 | Corticosteriods | prednisone |
| 440816560 | MA11 | Corticosteriods | prednisone |
| 10768728301 | MA11 | Corticosteriods | prednisone |
| 43063070330 | MA11 | Corticosteriods | prednisone |
| 55175415800 | MA11 | Corticosteriods | prednisone |
| 21695030714 | MA11 | Corticosteriods | prednisone |
| 54868083604 | MA11 | Corticosteriods | prednisone |
| 55045148002 | MA11 | Corticosteriods | prednisone |
| 58016021700 | MA11 | Corticosteriods | prednisone |
| 50090043900 | MA11 | Corticosteriods | prednisone |
| 16590062421 | MA11 | Corticosteriods | prednisone |
| 60346005830 | MA11 | Corticosteriods | prednisone |
| 54569304307 | MA11 | Corticosteriods | prednisone |
| 51552002825 | MA11 | Corticosteriods | prednisone |
| 51552002805 | MA11 | Corticosteriods | prednisone |
| 58016067220 | MA11 | Corticosteriods | prednisone |
| 55154495000 | MA11 | Corticosteriods | prednisone |
| 43063064440 | MA11 | Corticosteriods | prednisone |
| 54569330200 | MA11 | Corticosteriods | prednisone |
| 43063086612 | MA11 | Corticosteriods | prednisone |
| 247141730 | MA11 | Corticosteriods | prednisone |
| 52959012620 | MA11 | Corticosteriods | prednisone |
| 54473325 | MA11 | Corticosteriods | prednisone |
| 49999002820 | MA11 | Corticosteriods | prednisone |
| 254509423 | MA11 | Corticosteriods | prednisone |
| 10544091510 | MA11 | Corticosteriods | prednisone |
| 23490615701 | MA11 | Corticosteriods | prednisone |
| 247010009 | MA11 | Corticosteriods | prednisone |
| 259036421 | MA11 | Corticosteriods | prednisone |
| 68387024025 | MA11 | Corticosteriods | prednisone |
| 603533721 | MA11 | Corticosteriods | prednisone |
| 58864036256 | MA11 | Corticosteriods | prednisone |
| 603533732 | MA11 | Corticosteriods | prednisone |
| 60760062921 | MA11 | Corticosteriods | prednisone |
| 55289043850 | MA11 | Corticosteriods | prednisone |
| 70934009630 | MA11 | Corticosteriods | prednisone |
| 35356081818 | MA11 | Corticosteriods | prednisone |
| 61919032142 | MA11 | Corticosteriods | prednisone |
| 43063070306 | MA11 | Corticosteriods | prednisone |
| 68387024010 | MA11 | Corticosteriods | prednisone |
| 182108689 | MA11 | Corticosteriods | prednisone |
| 61919032621 | MA11 | Corticosteriods | prednisone |
| 68258301301 | MA11 | Corticosteriods | prednisone |
| 440816730 | MA11 | Corticosteriods | prednisone |
| 55700020815 | MA11 | Corticosteriods | prednisone |
| 591544305 | MA11 | Corticosteriods | prednisone |
| 440216720 | MA11 | Corticosteriods | prednisone |
| 527293037 | MA11 | Corticosteriods | prednisone |
| 16590037330 | MA11 | Corticosteriods | prednisone |
| 54569304300 | MA11 | Corticosteriods | prednisone |
| 615053643 | MA11 | Corticosteriods | prednisone |
| 58016021860 | MA11 | Corticosteriods | prednisone |
| 55700007220 | MA11 | Corticosteriods | prednisone |
| 60346005890 | MA11 | Corticosteriods | prednisone |
| 52959012642 | MA11 | Corticosteriods | prednisone |

| | | | |
|---|---|---|---|
| 55289035205 | MA11 | Corticosteriods | prednisone |
| 60760063060 | MA11 | Corticosteriods | prednisone |
| 70934009530 | MA11 | Corticosteriods | prednisone |
| 247007242 | MA11 | Corticosteriods | prednisone |
| 54868083602 | MA11 | Corticosteriods | prednisone |
| 591505201 | MA11 | Corticosteriods | prednisone |
| 60346051550 | MA11 | Corticosteriods | prednisone |
| 55175415805 | MA11 | Corticosteriods | prednisone |
| 51138015415 | MA11 | Corticosteriods | prednisone |
| 55887069648 | MA11 | Corticosteriods | prednisone |
| 364046190 | MA11 | Corticosteriods | prednisone |
| 50090198901 | MA11 | Corticosteriods | prednisone |
| 43063042628 | MA11 | Corticosteriods | prednisone |
| 54569033208 | MA11 | Corticosteriods | prednisone |
| 43063059025 | MA11 | Corticosteriods | prednisone |
| 55289037330 | MA11 | Corticosteriods | prednisone |
| 247010012 | MA11 | Corticosteriods | prednisone |
| 43063086642 | MA11 | Corticosteriods | prednisone |
| 58016021821 | MA11 | Corticosteriods | prednisone |
| 58016021660 | MA11 | Corticosteriods | prednisone |
| 403077001 | MA11 | Corticosteriods | prednisone |
| 54872925 | MA11 | Corticosteriods | prednisone |
| 254509413 | MA11 | Corticosteriods | prednisone |
| 10768708501 | MA11 | Corticosteriods | prednisone |
| 51079003301 | MA11 | Corticosteriods | prednisone |
| 21695030636 | MA11 | Corticosteriods | prednisone |
| 21695030709 | MA11 | Corticosteriods | prednisone |
| 60346005850 | MA11 | Corticosteriods | prednisone |
| 85084303 | MA11 | Corticosteriods | prednisone |
| 247007245 | MA11 | Corticosteriods | prednisone |
| 57866432606 | MA11 | Corticosteriods | prednisone |
| 440816720 | MA11 | Corticosteriods | prednisone |
| 247007280 | MA11 | Corticosteriods | prednisone |
| 247007222 | MA11 | Corticosteriods | prednisone |
| 21695076421 | MA11 | Corticosteriods | prednisone |
| 60346009424 | MA11 | Corticosteriods | prednisone |
| 35356067430 | MA11 | Corticosteriods | prednisone |
| 55045144402 | MA11 | Corticosteriods | prednisone |
| 63629158705 | MA11 | Corticosteriods | prednisone |
| 16590040420 | MA11 | Corticosteriods | prednisone |
| 54868025802 | MA11 | Corticosteriods | prednisone |
| 55175276608 | MA11 | Corticosteriods | prednisone |
| 70954006030 | MA11 | Corticosteriods | prednisone |
| 52959012625 | MA11 | Corticosteriods | prednisone |
| 33358029340 | MA11 | Corticosteriods | prednisone |
| 59746017306 | MA11 | Corticosteriods | prednisone |
| 45802030321 | MA11 | Corticosteriods | prednisone |
| 63874032712 | MA11 | Corticosteriods | prednisone |
| 43063086620 | MA11 | Corticosteriods | prednisone |
| 603533731 | MA11 | Corticosteriods | prednisone |
| 364021890 | MA11 | Corticosteriods | prednisone |
| 35356081821 | MA11 | Corticosteriods | prednisone |
| 43063059010 | MA11 | Corticosteriods | prednisone |
| 53489013901 | MA11 | Corticosteriods | prednisone |
| 63629157902 | MA11 | Corticosteriods | prednisone |
| 63874032760 | MA11 | Corticosteriods | prednisone |
| 58016021621 | MA11 | Corticosteriods | prednisone |
| 35356081810 | MA11 | Corticosteriods | prednisone |
| 60346051590 | MA11 | Corticosteriods | prednisone |
| 9004516 | MA11 | Corticosteriods | prednisone |
| 247117621 | MA11 | Corticosteriods | prednisone |
| 67296119403 | MA11 | Corticosteriods | prednisone |
| 71335065106 | MA11 | Corticosteriods | prednisone |
| 21695030648 | MA11 | Corticosteriods | prednisone |
| 49999002814 | MA11 | Corticosteriods | prednisone |
| 527293341 | MA11 | Corticosteriods | prednisone |
| 54569033102 | MA11 | Corticosteriods | prednisone |
| 364046105 | MA11 | Corticosteriods | prednisone |
| 51991045810 | MA11 | Corticosteriods | prednisone |
| 16590032645 | MA11 | Corticosteriods | prednisone |
| 33358029215 | MA11 | Corticosteriods | prednisone |

| | | | |
|---|---|---|---|
| 52544083051 | MA11 | Corticosteriods | prednisone |
| 63629158703 | MA11 | Corticosteriods | prednisone |
| 42254036128 | MA11 | Corticosteriods | prednisone |
| 49452600002 | MA11 | Corticosteriods | prednisone |
| 70954005940 | MA11 | Corticosteriods | prednisone |
| 55887079630 | MA11 | Corticosteriods | prednisone |
| 51138015536 | MA11 | Corticosteriods | prednisone |
| 615053663 | MA11 | Corticosteriods | prednisone |
| 43063042660 | MA11 | Corticosteriods | prednisone |
| 603533621 | MA11 | Corticosteriods | prednisone |
| 10544091530 | MA11 | Corticosteriods | prednisone |
| 49999039021 | MA11 | Corticosteriods | prednisone |
| 247010042 | MA11 | Corticosteriods | prednisone |
| 49999002830 | MA11 | Corticosteriods | prednisone |
| 49999011021 | MA11 | Corticosteriods | prednisone |
| 23490785400 | MA11 | Corticosteriods | prednisone |
| 50090240200 | MA11 | Corticosteriods | prednisone |
| 51138014721 | MA11 | Corticosteriods | prednisone |
| 61919023510 | MA11 | Corticosteriods | prednisone |
| 52959012707 | MA11 | Corticosteriods | prednisone |
| 55045144408 | MA11 | Corticosteriods | prednisone |
| 55175399206 | MA11 | Corticosteriods | prednisone |
| 51079003317 | MA11 | Corticosteriods | prednisone |
| 54868523000 | MA11 | Corticosteriods | prednisone |
| 63739051810 | MA11 | Corticosteriods | prednisone |
| 63629160508 | MA11 | Corticosteriods | prednisone |
| 63187080710 | MA11 | Corticosteriods | prednisone |
| 61919037321 | MA11 | Corticosteriods | prednisone |
| 55175399205 | MA11 | Corticosteriods | prednisone |
| 247007240 | MA11 | Corticosteriods | prednisone |
| 42254036142 | MA11 | Corticosteriods | prednisone |
| 21695030712 | MA11 | Corticosteriods | prednisone |
| 52959012718 | MA11 | Corticosteriods | prednisone |
| 58864042420 | MA11 | Corticosteriods | prednisone |
| 66116041005 | MA11 | Corticosteriods | prednisone |
| 378064001 | MA11 | Corticosteriods | prednisone |
| 23490615807 | MA11 | Corticosteriods | prednisone |
| 43063070318 | MA11 | Corticosteriods | prednisone |
| 35356081921 | MA11 | Corticosteriods | prednisone |
| 63187002036 | MA11 | Corticosteriods | prednisone |
| 10768773301 | MA11 | Corticosteriods | prednisone |
| 55175391808 | MA11 | Corticosteriods | prednisone |
| 66267017115 | MA11 | Corticosteriods | prednisone |
| 63739020910 | MA11 | Corticosteriods | prednisone |
| 68084003585 | MA11 | Corticosteriods | prednisone |
| 591544310 | MA11 | Corticosteriods | prednisone |
| 16590032610 | MA11 | Corticosteriods | prednisone |
| 68030432501 | MA11 | Corticosteriods | prednisone |
| 403077471 | MA11 | Corticosteriods | prednisone |
| 53489013910 | MA11 | Corticosteriods | prednisone |
| 54569033009 | MA11 | Corticosteriods | prednisone |
| 615359363 | MA11 | Corticosteriods | prednisone |
| 51138015550 | MA11 | Corticosteriods | prednisone |
| 55045153306 | MA11 | Corticosteriods | prednisone |
| 21695030690 | MA11 | Corticosteriods | prednisone |
| 54569033300 | MA11 | Corticosteriods | prednisone |
| 23490615705 | MA11 | Corticosteriods | prednisone |
| 603533532 | MA11 | Corticosteriods | prednisone |
| 54868025808 | MA11 | Corticosteriods | prednisone |
| 21695058014 | MA11 | Corticosteriods | prednisone |
| 63629157905 | MA11 | Corticosteriods | prednisone |
| 57866432801 | MA11 | Corticosteriods | prednisone |
| 60346009415 | MA11 | Corticosteriods | prednisone |
| 54873925 | MA11 | Corticosteriods | prednisone |
| 54868118301 | MA11 | Corticosteriods | prednisone |
| 63187080706 | MA11 | Corticosteriods | prednisone |
| 54569330201 | MA11 | Corticosteriods | prednisone |
| 55289035221 | MA11 | Corticosteriods | prednisone |
| 63874032790 | MA11 | Corticosteriods | prednisone |
| 68084003580 | MA11 | Corticosteriods | prednisone |
| 55175391809 | MA11 | Corticosteriods | prednisone |

| | | | |
|---|---|---|---|
| 55175399209 | MA11 | Corticosteriods | prednisone |
| 51138014730 | MA11 | Corticosteriods | prednisone |
| 63874039215 | MA11 | Corticosteriods | prednisone |
| 21695030640 | MA11 | Corticosteriods | prednisone |
| 43063010910 | MA11 | Corticosteriods | prednisone |
| 247007236 | MA11 | Corticosteriods | prednisone |
| 49452600001 | MA11 | Corticosteriods | prednisone |
| 55045148008 | MA11 | Corticosteriods | prednisone |
| 10544091421 | MA11 | Corticosteriods | prednisone |
| 55289035214 | MA11 | Corticosteriods | prednisone |
| 54868083605 | MA11 | Corticosteriods | prednisone |
| 54868111901 | MA11 | Corticosteriods | prednisone |
| 63874039202 | MA11 | Corticosteriods | prednisone |
| 70934010121 | MA11 | Corticosteriods | prednisone |
| 247007230 | MA11 | Corticosteriods | prednisone |
| 54569304301 | MA11 | Corticosteriods | prednisone |
| 43063042630 | MA11 | Corticosteriods | prednisone |
| 677042701 | MA11 | Corticosteriods | prednisone |
| 60346051512 | MA11 | Corticosteriods | prednisone |
| 43063061021 | MA11 | Corticosteriods | prednisone |
| 61919040430 | MA11 | Corticosteriods | prednisone |
| 52959012644 | MA11 | Corticosteriods | prednisone |
| 63874032724 | MA11 | Corticosteriods | prednisone |
| 55700020320 | MA11 | Corticosteriods | prednisone |
| 247007252 | MA11 | Corticosteriods | prednisone |
| 49999011007 | MA11 | Corticosteriods | prednisone |
| 52959022010 | MA11 | Corticosteriods | prednisone |
| 63629158706 | MA11 | Corticosteriods | prednisone |
| 42254036148 | MA11 | Corticosteriods | prednisone |
| 54001829 | MA11 | Corticosteriods | prednisone |
| 52544083205 | MA11 | Corticosteriods | prednisone |
| 63187080705 | MA11 | Corticosteriods | prednisone |
| 63629157909 | MA11 | Corticosteriods | prednisone |
| 10544053830 | MA11 | Corticosteriods | prednisone |
| 63874032740 | MA11 | Corticosteriods | prednisone |
| 21695030639 | MA11 | Corticosteriods | prednisone |
| 55175391801 | MA11 | Corticosteriods | prednisone |
| 21695030790 | MA11 | Corticosteriods | prednisone |
| 58016021715 | MA11 | Corticosteriods | prednisone |
| 63874039220 | MA11 | Corticosteriods | prednisone |
| 677042705 | MA11 | Corticosteriods | prednisone |
| 51655002052 | MA11 | Corticosteriods | prednisone |
| 70954006110 | MA11 | Corticosteriods | prednisone |
| 63187024307 | MA11 | Corticosteriods | prednisone |
| 54874025 | MA11 | Corticosteriods | prednisone |
| 364044201 | MA11 | Corticosteriods | prednisone |
| 68115029130 | MA11 | Corticosteriods | prednisone |
| 52959012612 | MA11 | Corticosteriods | prednisone |
| 55700020314 | MA11 | Corticosteriods | prednisone |
| 55175399202 | MA11 | Corticosteriods | prednisone |
| 10768728303 | MA11 | Corticosteriods | prednisone |
| 50090336104 | MA11 | Corticosteriods | prednisone |
| 440816810 | MA11 | Corticosteriods | prednisone |
| 52544083101 | MA11 | Corticosteriods | prednisone |
| 55289037342 | MA11 | Corticosteriods | prednisone |
| 55700007218 | MA11 | Corticosteriods | prednisone |
| 18837026721 | MA11 | Corticosteriods | prednisone |
| 70934009610 | MA11 | Corticosteriods | prednisone |
| 54569330206 | MA11 | Corticosteriods | prednisone |
| 55289033005 | MA11 | Corticosteriods | prednisone |
| 68115028910 | MA11 | Corticosteriods | prednisone |
| 247010008 | MA11 | Corticosteriods | prednisone |
| 51138014636 | MA11 | Corticosteriods | prednisone |
| 68030978601 | MA11 | Corticosteriods | prednisone |
| 10544047320 | MA11 | Corticosteriods | prednisone |
| 52959022036 | MA11 | Corticosteriods | prednisone |
| 440816805 | MA11 | Corticosteriods | prednisone |
| 43353081960 | MA11 | Corticosteriods | prednisone |
| 66116046530 | MA11 | Corticosteriods | prednisone |
| 55700007221 | MA11 | Corticosteriods | prednisone |
| 68030432608 | MA11 | Corticosteriods | prednisone |

| | | | |
|---|---|---|---|
| 603533432 | MA11 | Corticosteriods | prednisone |
| 58016021760 | MA11 | Corticosteriods | prednisone |
| 60346051530 | MA11 | Corticosteriods | prednisone |
| 51655008627 | MA11 | Corticosteriods | prednisone |
| 55045148009 | MA11 | Corticosteriods | prednisone |
| 54569033105 | MA11 | Corticosteriods | prednisone |
| 58016021850 | MA11 | Corticosteriods | prednisone |
| 63187080730 | MA11 | Corticosteriods | prednisone |
| 63629158701 | MA11 | Corticosteriods | prednisone |
| 60346005821 | MA11 | Corticosteriods | prednisone |
| 54868118304 | MA11 | Corticosteriods | prednisone |
| 66336005860 | MA11 | Corticosteriods | prednisone |
| 63629462401 | MA11 | Corticosteriods | prednisone |
| 55700020330 | MA11 | Corticosteriods | prednisone |
| 66116042005 | MA11 | Corticosteriods | prednisone |
| 33358029440 | MA11 | Corticosteriods | prednisone |
| 35356081820 | MA11 | Corticosteriods | prednisone |
| 55045148000 | MA11 | Corticosteriods | prednisone |
| 54569330208 | MA11 | Corticosteriods | prednisone |
| 60346005854 | MA11 | Corticosteriods | prednisone |
| 70954005840 | MA11 | Corticosteriods | prednisone |
| 403077415 | MA11 | Corticosteriods | prednisone |
| 52544083010 | MA11 | Corticosteriods | prednisone |
| 52959022030 | MA11 | Corticosteriods | prednisone |
| 58864042409 | MA11 | Corticosteriods | prednisone |
| 43063043220 | MA11 | Corticosteriods | prednisone |
| 54569402605 | MA11 | Corticosteriods | prednisone |
| 58016021855 | MA11 | Corticosteriods | prednisone |
| 63187002021 | MA11 | Corticosteriods | prednisone |
| 63629158702 | MA11 | Corticosteriods | prednisone |
| 66267017120 | MA11 | Corticosteriods | prednisone |
| 440816604 | MA11 | Corticosteriods | prednisone |
| 57866432502 | MA11 | Corticosteriods | prednisone |
| 63874039230 | MA11 | Corticosteriods | prednisone |
| 10544091420 | MA11 | Corticosteriods | prednisone |
| 58016021830 | MA11 | Corticosteriods | prednisone |
| 49999000842 | MA11 | Corticosteriods | prednisone |
| 143973901 | MA11 | Corticosteriods | prednisone |
| 42254036120 | MA11 | Corticosteriods | prednisone |
| 615359329 | MA11 | Corticosteriods | prednisone |
| 54472831 | MA11 | Corticosteriods | prednisone |
| 43063091121 | MA11 | Corticosteriods | prednisone |
| 55175415804 | MA11 | Corticosteriods | prednisone |
| 55700020310 | MA11 | Corticosteriods | prednisone |
| 247007140 | MA11 | Corticosteriods | prednisone |
| 50090198905 | MA11 | Corticosteriods | prednisone |
| 54569033101 | MA11 | Corticosteriods | prednisone |
| 603533831 | MA11 | Corticosteriods | prednisone |
| 143973810 | MA11 | Corticosteriods | prednisone |
| 403196518 | MA11 | Corticosteriods | prednisone |
| 247007215 | MA11 | Corticosteriods | prednisone |
| 55289043830 | MA11 | Corticosteriods | prednisone |
| 49999000821 | MA11 | Corticosteriods | prednisone |
| 52959012712 | MA11 | Corticosteriods | prednisone |
| 63629160504 | MA11 | Corticosteriods | prednisone |
| 75987002201 | MA11 | Corticosteriods | prednisone |
| 70882011721 | MA11 | Corticosteriods | prednisone |
| 440816803 | MA11 | Corticosteriods | prednisone |
| 10768773303 | MA11 | Corticosteriods | prednisone |
| 43063042610 | MA11 | Corticosteriods | prednisone |
| 52959012615 | MA11 | Corticosteriods | prednisone |
| 55175415803 | MA11 | Corticosteriods | prednisone |
| 54569384700 | MA11 | Corticosteriods | prednisone |
| 54372250 | MA11 | Corticosteriods | prednisone |
| 49999002821 | MA11 | Corticosteriods | prednisone |
| 53489013801 | MA11 | Corticosteriods | prednisone |
| 43063059009 | MA11 | Corticosteriods | prednisone |
| 70882011730 | MA11 | Corticosteriods | prednisone |
| 58016021614 | MA11 | Corticosteriods | prednisone |
| 51138014710 | MA11 | Corticosteriods | prednisone |
| 57866432507 | MA11 | Corticosteriods | prednisone |

| | | | |
|---|---|---|---|
| 60429013101 | MA11 | Corticosteriods | prednisone |
| 55175415802 | MA11 | Corticosteriods | prednisone |
| 63739020810 | MA11 | Corticosteriods | prednisone |
| 57866432503 | MA11 | Corticosteriods | prednisone |
| 440816512 | MA11 | Corticosteriods | prednisone |
| 55289037301 | MA11 | Corticosteriods | prednisone |
| 43063059021 | MA11 | Corticosteriods | prednisone |
| 54569330209 | MA11 | Corticosteriods | prednisone |
| 23490615804 | MA11 | Corticosteriods | prednisone |
| 55887064390 | MA11 | Corticosteriods | prednisone |
| 403077021 | MA11 | Corticosteriods | prednisone |
| 54001925 | MA11 | Corticosteriods | prednisone |
| 54868521300 | MA11 | Corticosteriods | prednisone |
| 63874032718 | MA11 | Corticosteriods | prednisone |
| 21695030718 | MA11 | Corticosteriods | prednisone |
| 43063042621 | MA11 | Corticosteriods | prednisone |
| 57866432601 | MA11 | Corticosteriods | prednisone |
| 70954005820 | MA11 | Corticosteriods | prednisone |
| 54868025806 | MA11 | Corticosteriods | prednisone |
| 66336005810 | MA11 | Corticosteriods | prednisone |
| 16590040445 | MA11 | Corticosteriods | prednisone |
| 60346005818 | MA11 | Corticosteriods | prednisone |
| 55887079620 | MA11 | Corticosteriods | prednisone |
| 60429013110 | MA11 | Corticosteriods | prednisone |
| 70954005710 | MA11 | Corticosteriods | prednisone |
| 43063059020 | MA11 | Corticosteriods | prednisone |
| 52959012720 | MA11 | Corticosteriods | prednisone |
| 55175415801 | MA11 | Corticosteriods | prednisone |
| 60760025560 | MA11 | Corticosteriods | prednisone |
| 43063096855 | MA11 | Corticosteriods | prednisone |
| 49999000855 | MA11 | Corticosteriods | prednisone |
| 63874037330 | MA11 | Corticosteriods | prednisone |
| 43063064450 | MA11 | Corticosteriods | prednisone |
| 55700020318 | MA11 | Corticosteriods | prednisone |
| 63629157904 | MA11 | Corticosteriods | prednisone |
| 54569033109 | MA11 | Corticosteriods | prednisone |
| 58016021630 | MA11 | Corticosteriods | prednisone |
| 55289035220 | MA11 | Corticosteriods | prednisone |
| 182133489 | MA11 | Corticosteriods | prednisone |
| 61919034221 | MA11 | Corticosteriods | prednisone |
| 63874037301 | MA11 | Corticosteriods | prednisone |
| 35356067790 | MA11 | Corticosteriods | prednisone |
| 43063064355 | MA11 | Corticosteriods | prednisone |
| 58016021840 | MA11 | Corticosteriods | prednisone |
| 63629160505 | MA11 | Corticosteriods | prednisone |
| 68115028930 | MA11 | Corticosteriods | prednisone |
| 247097340 | MA11 | Corticosteriods | prednisone |
| 71335065103 | MA11 | Corticosteriods | prednisone |
| 60687013411 | MA11 | Corticosteriods | prednisone |
| 43063086650 | MA11 | Corticosteriods | prednisone |
| 54569033000 | MA11 | Corticosteriods | prednisone |
| 57866432604 | MA11 | Corticosteriods | prednisone |
| 615053653 | MA11 | Corticosteriods | prednisone |
| 63874037336 | MA11 | Corticosteriods | prednisone |
| 58864042340 | MA11 | Corticosteriods | prednisone |
| 23490615802 | MA11 | Corticosteriods | prednisone |
| 35356067321 | MA11 | Corticosteriods | prednisone |
| 43063064420 | MA11 | Corticosteriods | prednisone |
| 63629158708 | MA11 | Corticosteriods | prednisone |
| 591544210 | MA11 | Corticosteriods | prednisone |
| 603533315 | MA11 | Corticosteriods | prednisone |
| 52959012715 | MA11 | Corticosteriods | prednisone |
| 66267017342 | MA11 | Corticosteriods | prednisone |
| 55045153307 | MA11 | Corticosteriods | prednisone |
| 63874032732 | MA11 | Corticosteriods | prednisone |
| 182133410 | MA11 | Corticosteriods | prednisone |
| 591505221 | MA11 | Corticosteriods | prednisone |
| 16590040430 | MA11 | Corticosteriods | prednisone |
| 59746017210 | MA11 | Corticosteriods | prednisone |
| 23490615904 | MA11 | Corticosteriods | prednisone |
| 52959012710 | MA11 | Corticosteriods | prednisone |

| | | | |
|---|---|---|---|
| 603533928 | MA11 | Corticosteriods | prednisone |
| 52959012670 | MA11 | Corticosteriods | prednisone |
| 62584083233 | MA11 | Corticosteriods | prednisone |
| 259040049 | MA11 | Corticosteriods | prednisone |
| 55289043836 | MA11 | Corticosteriods | prednisone |
| 55289033007 | MA11 | Corticosteriods | prednisone |
| 527293243 | MA11 | Corticosteriods | prednisone |
| 62584083401 | MA11 | Corticosteriods | prednisone |
| 591544221 | MA11 | Corticosteriods | prednisone |
| 63629158704 | MA11 | Corticosteriods | prednisone |
| 58016021730 | MA11 | Corticosteriods | prednisone |
| 247007204 | MA11 | Corticosteriods | prednisone |
| 57480047201 | MA11 | Corticosteriods | prednisone |
| 57866432803 | MA11 | Corticosteriods | prednisone |
| 247132321 | MA11 | Corticosteriods | prednisone |
| 63187080718 | MA11 | Corticosteriods | prednisone |
| 43063042640 | MA11 | Corticosteriods | prednisone |
| 54868025809 | MA11 | Corticosteriods | prednisone |
| 54569330205 | MA11 | Corticosteriods | prednisone |
| 35356067418 | MA11 | Corticosteriods | prednisone |
| 63739052010 | MA11 | Corticosteriods | prednisone |
| 440816506 | MA11 | Corticosteriods | prednisone |
| 904215760 | MA11 | Corticosteriods | prednisone |
| 21695030715 | MA11 | Corticosteriods | prednisone |
| 54473029 | MA11 | Corticosteriods | prednisone |
| 21695030620 | MA11 | Corticosteriods | prednisone |
| 23490615708 | MA11 | Corticosteriods | prednisone |
| 18837026730 | MA11 | Corticosteriods | prednisone |
| 603533221 | MA11 | Corticosteriods | prednisone |
| 677011710 | MA11 | Corticosteriods | prednisone |
| 50090199002 | MA11 | Corticosteriods | prednisone |
| 50436173005 | MA11 | Corticosteriods | prednisone |
| 54472925 | MA11 | Corticosteriods | prednisone |
| 143147501 | MA11 | Corticosteriods | prednisone |
| 49999011012 | MA11 | Corticosteriods | prednisone |
| 254509323 | MA11 | Corticosteriods | prednisone |
| 35356081815 | MA11 | Corticosteriods | prednisone |
| 58016021724 | MA11 | Corticosteriods | prednisone |
| 43063096830 | MA11 | Corticosteriods | prednisone |
| 247010007 | MA11 | Corticosteriods | prednisone |
| 403077220 | MA11 | Corticosteriods | prednisone |
| 43063064410 | MA11 | Corticosteriods | prednisone |
| 58016483201 | MA11 | Corticosteriods | prednisone |
| 66267017140 | MA11 | Corticosteriods | prednisone |
| 68115094721 | MA11 | Corticosteriods | prednisone |
| 247010003 | MA11 | Corticosteriods | prednisone |
| 49999002865 | MA11 | Corticosteriods | prednisone |
| 21695058007 | MA11 | Corticosteriods | prednisone |
| 247007244 | MA11 | Corticosteriods | prednisone |
| 51552002806 | MA11 | Corticosteriods | prednisone |
| 33358029320 | MA11 | Corticosteriods | prednisone |
| 53489014005 | MA11 | Corticosteriods | prednisone |
| 60760000221 | MA11 | Corticosteriods | prednisone |
| 63874032710 | MA11 | Corticosteriods | prednisone |
| 55700020315 | MA11 | Corticosteriods | prednisone |
| 21695030645 | MA11 | Corticosteriods | prednisone |
| 43353008060 | MA11 | Corticosteriods | prednisone |
| 49999002805 | MA11 | Corticosteriods | prednisone |
| 440816760 | MA11 | Corticosteriods | prednisone |
| 54868111905 | MA11 | Corticosteriods | prednisone |
| 42254027621 | MA11 | Corticosteriods | prednisone |
| 66336009410 | MA11 | Corticosteriods | prednisone |
| 55175391800 | MA11 | Corticosteriods | prednisone |
| 67296024702 | MA11 | Corticosteriods | prednisone |
| 591544201 | MA11 | Corticosteriods | prednisone |
| 49999011000 | MA11 | Corticosteriods | prednisone |
| 51655002053 | MA11 | Corticosteriods | prednisone |
| 54868111904 | MA11 | Corticosteriods | prednisone |
| 49999002850 | MA11 | Corticosteriods | prednisone |
| 54872425 | MA11 | Corticosteriods | prednisone |
| 54569033205 | MA11 | Corticosteriods | prednisone |

| | | | |
|---|---|---|---|
| 63739020815 | MA11 | Corticosteriods | prednisone |
| 60346009430 | MA11 | Corticosteriods | prednisone |
| 247007203 | MA11 | Corticosteriods | prednisone |
| 60346051540 | MA11 | Corticosteriods | prednisone |
| 52959012610 | MA11 | Corticosteriods | prednisone |
| 16590036548 | MA11 | Corticosteriods | prednisone |
| 51138014620 | MA11 | Corticosteriods | prednisone |
| 54569402600 | MA11 | Corticosteriods | prednisone |
| 43063064430 | MA11 | Corticosteriods | prednisone |
| 63874032730 | MA11 | Corticosteriods | prednisone |
| 43063043212 | MA11 | Corticosteriods | prednisone |
| 10768728302 | MA11 | Corticosteriods | prednisone |
| 71335065107 | MA11 | Corticosteriods | prednisone |
| 603533921 | MA11 | Corticosteriods | prednisone |
| 54868083603 | MA11 | Corticosteriods | prednisone |
| 43063086628 | MA11 | Corticosteriods | prednisone |
| 66267017230 | MA11 | Corticosteriods | prednisone |
| 440816630 | MA11 | Corticosteriods | prednisone |
| 55175415807 | MA11 | Corticosteriods | prednisone |
| 57866432508 | MA11 | Corticosteriods | prednisone |
| 51991046201 | MA11 | Corticosteriods | prednisone |
| 440816703 | MA11 | Corticosteriods | prednisone |
| 54372263 | MA11 | Corticosteriods | prednisone |
| 49999000805 | MA11 | Corticosteriods | prednisone |
| 66116046550 | MA11 | Corticosteriods | prednisone |
| 23490615704 | MA11 | Corticosteriods | prednisone |
| 55700007215 | MA11 | Corticosteriods | prednisone |
| 50436432401 | MA11 | Corticosteriods | prednisone |
| 403077421 | MA11 | Corticosteriods | prednisone |
| 527293437 | MA11 | Corticosteriods | prednisone |
| 52959022021 | MA11 | Corticosteriods | prednisone |
| 440816721 | MA11 | Corticosteriods | prednisone |
| 35356067720 | MA11 | Corticosteriods | prednisone |
| 10768773302 | MA11 | Corticosteriods | prednisone |
| 9016502 | MA11 | Corticosteriods | prednisone |
| 33358029212 | MA11 | Corticosteriods | prednisone |
| 54569033104 | MA11 | Corticosteriods | prednisone |
| 50090327304 | MA11 | Corticosteriods | prednisone |
| 54569033100 | MA11 | Corticosteriods | prednisone |
| 63629157908 | MA11 | Corticosteriods | prednisone |
| 43063059018 | MA11 | Corticosteriods | prednisone |
| 62584083333 | MA11 | Corticosteriods | prednisone |
| 247007221 | MA11 | Corticosteriods | prednisone |
| 43063043230 | MA11 | Corticosteriods | prednisone |
| 66336005812 | MA11 | Corticosteriods | prednisone |
| 58016021640 | MA11 | Corticosteriods | prednisone |
| 66267017130 | MA11 | Corticosteriods | prednisone |
| 68115028942 | MA11 | Corticosteriods | prednisone |
| 35356081930 | MA11 | Corticosteriods | prednisone |
| 51138014714 | MA11 | Corticosteriods | prednisone |
| 54872625 | MA11 | Corticosteriods | prednisone |
| 63187030028 | MA11 | Corticosteriods | prednisone |
| 60346051575 | MA11 | Corticosteriods | prednisone |
| 58016021824 | MA11 | Corticosteriods | prednisone |
| 364021801 | MA11 | Corticosteriods | prednisone |
| 603533832 | MA11 | Corticosteriods | prednisone |
| 43063043210 | MA11 | Corticosteriods | prednisone |
| 54868090802 | MA11 | Corticosteriods | prednisone |
| 55289043838 | MA11 | Corticosteriods | prednisone |
| 51138015539 | MA11 | Corticosteriods | prednisone |
| 63739020801 | MA11 | Corticosteriods | prednisone |
| 143974001 | MA11 | Corticosteriods | prednisone |
| 54868090803 | MA11 | Corticosteriods | prednisone |
| 70954005930 | MA11 | Corticosteriods | prednisone |
| 143973805 | MA11 | Corticosteriods | prednisone |
| 603533331 | MA11 | Corticosteriods | prednisone |
| 43063059006 | MA11 | Corticosteriods | prednisone |
| 55175392108 | MA11 | Corticosteriods | prednisone |
| 50090327300 | MA11 | Corticosteriods | prednisone |
| 21695030621 | MA11 | Corticosteriods | prednisone |
| 51138014515 | MA11 | Corticosteriods | prednisone |

| | | | |
|---|---|---|---|
| 35356081942 | MA11 | Corticosteriods | prednisone |
| 440216715 | MA11 | Corticosteriods | prednisone |
| 59746017510 | MA11 | Corticosteriods | prednisone |
| 66267017330 | MA11 | Corticosteriods | prednisone |
| 63187003730 | MA11 | Corticosteriods | prednisone |
| 55045153308 | MA11 | Corticosteriods | prednisone |
| 63187080720 | MA11 | Corticosteriods | prednisone |
| 21695030590 | MA11 | Corticosteriods | prednisone |
| 58016021600 | MA11 | Corticosteriods | prednisone |
| 43063042642 | MA11 | Corticosteriods | prednisone |
| 58016021740 | MA11 | Corticosteriods | prednisone |
| 49999002890 | MA11 | Corticosteriods | prednisone |
| 51138014615 | MA11 | Corticosteriods | prednisone |
| 66336051530 | MA11 | Corticosteriods | prednisone |
| 247007261 | MA11 | Corticosteriods | prednisone |
| 60346005860 | MA11 | Corticosteriods | prednisone |
| 61919032620 | MA11 | Corticosteriods | prednisone |
| 247010021 | MA11 | Corticosteriods | prednisone |
| 16590032621 | MA11 | Corticosteriods | prednisone |
| 21695076521 | MA11 | Corticosteriods | prednisone |
| 35356081830 | MA11 | Corticosteriods | prednisone |
| 43063070309 | MA11 | Corticosteriods | prednisone |
| 51138014712 | MA11 | Corticosteriods | prednisone |
| 143147510 | MA11 | Corticosteriods | prednisone |
| 591544301 | MA11 | Corticosteriods | prednisone |
| 10544091330 | MA11 | Corticosteriods | prednisone |
| 55887069621 | MA11 | Corticosteriods | prednisone |
| 63874032721 | MA11 | Corticosteriods | prednisone |
| 55700020830 | MA11 | Corticosteriods | prednisone |
| 43063070320 | MA11 | Corticosteriods | prednisone |
| 9019302 | MA11 | Corticosteriods | prednisone |
| 55175399207 | MA11 | Corticosteriods | prednisone |
| 54868118302 | MA11 | Corticosteriods | prednisone |
| 677042710 | MA11 | Corticosteriods | prednisone |
| 54569304305 | MA11 | Corticosteriods | prednisone |
| 403077436 | MA11 | Corticosteriods | prednisone |
| 440816520 | MA11 | Corticosteriods | prednisone |
| 63874077230 | MA11 | Corticosteriods | prednisone |
| 182108610 | MA11 | Corticosteriods | prednisone |
| 58016021615 | MA11 | Corticosteriods | prednisone |
| 63629157901 | MA11 | Corticosteriods | prednisone |
| 71205023910 | MA11 | Corticosteriods | prednisone |
| 615154229 | MA11 | Corticosteriods | prednisone |
| 43353065760 | MA11 | Corticosteriods | prednisone |
| 70954005920 | MA11 | Corticosteriods | prednisone |
| 50090333602 | MA11 | Corticosteriods | prednisone |
| 43063064442 | MA11 | Corticosteriods | prednisone |
| 55175391901 | MA11 | Corticosteriods | prednisone |
| 71205023905 | MA11 | Corticosteriods | prednisone |
| 9019301 | MA11 | Corticosteriods | prednisone |
| 54569304303 | MA11 | Corticosteriods | prednisone |
| 58016021836 | MA11 | Corticosteriods | prednisone |
| 440816510 | MA11 | Corticosteriods | prednisone |
| 254509313 | MA11 | Corticosteriods | prednisone |
| 58016021710 | MA11 | Corticosteriods | prednisone |
| 61919010812 | MA11 | Corticosteriods | prednisone |
| 66267017242 | MA11 | Corticosteriods | prednisone |
| 58864042315 | MA11 | Corticosteriods | prednisone |
| 54868092301 | MA11 | Corticosteriods | prednisone |
| 43063059015 | MA11 | Corticosteriods | prednisone |
| 49999011015 | MA11 | Corticosteriods | prednisone |
| 247007227 | MA11 | Corticosteriods | prednisone |
| 23490615906 | MA11 | Corticosteriods | prednisone |
| 61919023540 | MA11 | Corticosteriods | prednisone |
| 21695076548 | MA11 | Corticosteriods | prednisone |
| 143973801 | MA11 | Corticosteriods | prednisone |
| 54569033200 | MA11 | Corticosteriods | prednisone |
| 182020189 | MA11 | Corticosteriods | prednisone |
| 54472929 | MA11 | Corticosteriods | prednisone |
| 364044205 | MA11 | Corticosteriods | prednisone |
| 43063042612 | MA11 | Corticosteriods | prednisone |

| | | | |
|---|---|---|---|
| 51138014642 | MA11 | Corticosteriods | prednisone |
| 71335065102 | MA11 | Corticosteriods | prednisone |
| 43063086640 | MA11 | Corticosteriods | prednisone |
| 247007210 | MA11 | Corticosteriods | prednisone |
| 54473025 | MA11 | Corticosteriods | prednisone |
| 66267017340 | MA11 | Corticosteriods | prednisone |
| 50090333500 | MA11 | Corticosteriods | prednisone |
| 55045126009 | MA11 | Corticosteriods | prednisone |
| 16590037321 | MA11 | Corticosteriods | prednisone |
| 55175415809 | MA11 | Corticosteriods | prednisone |
| 50090260800 | MA11 | Corticosteriods | prednisone |
| 677069805 | MA11 | Corticosteriods | prednisone |
| 51552002801 | MA11 | Corticosteriods | prednisone |
| 63187030015 | MA11 | Corticosteriods | prednisone |
| 33358029221 | MA11 | Corticosteriods | prednisone |
| 50090100100 | MA11 | Corticosteriods | prednisone |
| 904214160 | MA11 | Corticosteriods | prednisone |
| 49999000800 | MA11 | Corticosteriods | prednisone |
| 54569278500 | MA11 | Corticosteriods | prednisone |
| 21695030730 | MA11 | Corticosteriods | prednisone |
| 66336005840 | MA11 | Corticosteriods | prednisone |
| 55289035230 | MA11 | Corticosteriods | prednisone |
| 51138015420 | MA11 | Corticosteriods | prednisone |
| 52959012607 | MA11 | Corticosteriods | prednisone |
| 60346009418 | MA11 | Corticosteriods | prednisone |
| 52959012737 | MA11 | Corticosteriods | prednisone |
| 63874039210 | MA11 | Corticosteriods | prednisone |
| 55887077021 | MA11 | Corticosteriods | prednisone |
| 247010010 | MA11 | Corticosteriods | prednisone |
| 182020100 | MA11 | Corticosteriods | prednisone |
| 54569033008 | MA11 | Corticosteriods | prednisone |
| 51138015542 | MA11 | Corticosteriods | prednisone |
| 43063096821 | MA11 | Corticosteriods | prednisone |
| 54868025804 | MA11 | Corticosteriods | prednisone |
| 364044202 | MA11 | Corticosteriods | prednisone |
| 63874039224 | MA11 | Corticosteriods | prednisone |
| 60687013401 | MA11 | Corticosteriods | prednisone |
| 55175399203 | MA11 | Corticosteriods | prednisone |
| 615154253 | MA11 | Corticosteriods | prednisone |
| 54569033103 | MA11 | Corticosteriods | prednisone |
| 55175391803 | MA11 | Corticosteriods | prednisone |
| 49999011020 | MA11 | Corticosteriods | prednisone |
| 51138015621 | MA11 | Corticosteriods | prednisone |
| 51138015612 | MA11 | Corticosteriods | prednisone |
| 52959012618 | MA11 | Corticosteriods | prednisone |
| 63874032725 | MA11 | Corticosteriods | prednisone |
| 63629160506 | MA11 | Corticosteriods | prednisone |
| 68115029030 | MA11 | Corticosteriods | prednisone |
| 9038801 | MA11 | Corticosteriods | prednisone |
| 35356067715 | MA11 | Corticosteriods | prednisone |
| 23629016710 | MA11 | Corticosteriods | prednisone |
| 52959022020 | MA11 | Corticosteriods | prednisone |
| 247007200 | MA11 | Corticosteriods | prednisone |
| 54868025800 | MA11 | Corticosteriods | prednisone |
| 259039148 | MA11 | Corticosteriods | prednisone |
| 54569401700 | MA11 | Corticosteriods | prednisone |
| 59746017110 | MA11 | Corticosteriods | prednisone |
| 527293337 | MA11 | Corticosteriods | prednisone |
| 42549064714 | MA11 | Corticosteriods | prednisone |
| 55175391806 | MA11 | Corticosteriods | prednisone |
| 63874032701 | MA11 | Corticosteriods | prednisone |
| 9003201 | MA11 | Corticosteriods | prednisone |
| 66116062530 | MA11 | Corticosteriods | prednisone |
| 55700020840 | MA11 | Corticosteriods | prednisone |
| 60687012201 | MA11 | Corticosteriods | prednisone |
| 57866432607 | MA11 | Corticosteriods | prednisone |
| 16590040421 | MA11 | Corticosteriods | prednisone |
| 55175391802 | MA11 | Corticosteriods | prednisone |
| 43063070325 | MA11 | Corticosteriods | prednisone |
| 247007142 | MA11 | Corticosteriods | prednisone |
| 182108600 | MA11 | Corticosteriods | prednisone |

| | | | |
|---|---|---|---|
| 51079002219 | MA11 | Corticosteriods | prednisone |
| 63629158707 | MA11 | Corticosteriods | prednisone |
| 16590062448 | MA11 | Corticosteriods | prednisone |
| 51138014639 | MA11 | Corticosteriods | prednisone |
| 403077230 | MA11 | Corticosteriods | prednisone |
| 63187002015 | MA11 | Corticosteriods | prednisone |
| 43063041530 | MA11 | Corticosteriods | prednisone |
| 66336051510 | MA11 | Corticosteriods | prednisone |
| 35356067728 | MA11 | Corticosteriods | prednisone |
| 70934009521 | MA11 | Corticosteriods | prednisone |
| 52959022060 | MA11 | Corticosteriods | prednisone |
| 247132360 | MA11 | Corticosteriods | prednisone |
| 54372144 | MA11 | Corticosteriods | prednisone |
| 54569341300 | MA11 | Corticosteriods | prednisone |
| 228233710 | MA11 | Corticosteriods | prednisone |
| 49999011014 | MA11 | Corticosteriods | prednisone |
| 54569422600 | MA11 | Corticosteriods | prednisone |
| 70934013705 | MA11 | Corticosteriods | prednisone |
| 35356067648 | MA11 | Corticosteriods | prednisone |
| 58016021833 | MA11 | Corticosteriods | prednisone |
| 63874039228 | MA11 | Corticosteriods | prednisone |
| 60429013210 | MA11 | Corticosteriods | prednisone |
| 54569033002 | MA11 | Corticosteriods | prednisone |
| 57866432404 | MA11 | Corticosteriods | prednisone |
| 35356067560 | MA11 | Corticosteriods | prednisone |
| 66336005830 | MA11 | Corticosteriods | prednisone |
| 677069801 | MA11 | Corticosteriods | prednisone |
| 54569033005 | MA11 | Corticosteriods | prednisone |
| 51138015528 | MA11 | Corticosteriods | prednisone |
| 63187080715 | MA11 | Corticosteriods | prednisone |
| 43063059005 | MA11 | Corticosteriods | prednisone |
| 66336051521 | MA11 | Corticosteriods | prednisone |
| 603533332 | MA11 | Corticosteriods | prednisone |
| 57480035101 | MA11 | Corticosteriods | prednisone |
| 55887077048 | MA11 | Corticosteriods | prednisone |
| 60346005812 | MA11 | Corticosteriods | prednisone |
| 440816610 | MA11 | Corticosteriods | prednisone |
| 55045148007 | MA11 | Corticosteriods | prednisone |
| 60346072140 | MA11 | Corticosteriods | prednisone |
| 49999011006 | MA11 | Corticosteriods | prednisone |
| 61919032615 | MA11 | Corticosteriods | prednisone |
| 51079003217 | MA11 | Corticosteriods | prednisone |
| 63187030036 | MA11 | Corticosteriods | prednisone |
| 21695030642 | MA11 | Corticosteriods | prednisone |
| 42254036130 | MA11 | Corticosteriods | prednisone |
| 54569033203 | MA11 | Corticosteriods | prednisone |
| 247010030 | MA11 | Corticosteriods | prednisone |
| 49999002860 | MA11 | Corticosteriods | prednisone |
| 16590032660 | MA11 | Corticosteriods | prednisone |
| 55289037360 | MA11 | Corticosteriods | prednisone |
| 63187030027 | MA11 | Corticosteriods | prednisone |
| 66267017142 | MA11 | Corticosteriods | prednisone |
| 21695030713 | MA11 | Corticosteriods | prednisone |
| 66267094821 | MA11 | Corticosteriods | prednisone |
| 61919032623 | MA11 | Corticosteriods | prednisone |
| 143147701 | MA11 | Corticosteriods | prednisone |
| 55700020821 | MA11 | Corticosteriods | prednisone |
| 54474125 | MA11 | Corticosteriods | prednisone |
| 23490615903 | MA11 | Corticosteriods | prednisone |
| 54569033108 | MA11 | Corticosteriods | prednisone |
| 60687012211 | MA11 | Corticosteriods | prednisone |
| 71335065108 | MA11 | Corticosteriods | prednisone |
| 55175415806 | MA11 | Corticosteriods | prednisone |
| 63187030040 | MA11 | Corticosteriods | prednisone |
| 63874039206 | MA11 | Corticosteriods | prednisone |
| 16590032615 | MA11 | Corticosteriods | prednisone |
| 54569584100 | MA11 | Corticosteriods | prednisone |
| 63874037302 | MA11 | Corticosteriods | prednisone |
| 51655008624 | MA11 | Corticosteriods | prednisone |
| 55289035212 | MA11 | Corticosteriods | prednisone |
| 527293237 | MA11 | Corticosteriods | prednisone |

| | | | |
|---|---|---|---|
| 52959012660 | MA11 | Corticosteriods | prednisone |
| 63629160507 | MA11 | Corticosteriods | prednisone |
| 50436432502 | MA11 | Corticosteriods | prednisone |
| 35356067760 | MA11 | Corticosteriods | prednisone |
| 55887064321 | MA11 | Corticosteriods | prednisone |
| 143147710 | MA11 | Corticosteriods | prednisone |
| 228233810 | MA11 | Corticosteriods | prednisone |
| 63187003721 | MA11 | Corticosteriods | prednisone |
| 54474225 | MA11 | Corticosteriods | prednisone |
| 55045144407 | MA11 | Corticosteriods | prednisone |
| 58016021718 | MA11 | Corticosteriods | prednisone |
| 21695058005 | MA11 | Corticosteriods | prednisone |
| 440816704 | MA11 | Corticosteriods | prednisone |
| 51138014628 | MA11 | Corticosteriods | prednisone |
| 61919036521 | MA11 | Corticosteriods | prednisone |
| 54868111903 | MA11 | Corticosteriods | prednisone |
| 55175276501 | MA11 | Corticosteriods | prednisone |
| 603533828 | MA11 | Corticosteriods | prednisone |
| 63629157903 | MA11 | Corticosteriods | prednisone |
| 63187080712 | MA11 | Corticosteriods | prednisone |
| 35356067330 | MA11 | Corticosteriods | prednisone |
| 60346009420 | MA11 | Corticosteriods | prednisone |
| 54001920 | MA11 | Corticosteriods | prednisone |
| 63874039201 | MA11 | Corticosteriods | prednisone |
| 51138015521 | MA11 | Corticosteriods | prednisone |
| 33358029420 | MA11 | Corticosteriods | prednisone |
| 53489014010 | MA11 | Corticosteriods | prednisone |
| 49999011030 | MA11 | Corticosteriods | prednisone |
| 35356067730 | MA11 | Corticosteriods | prednisone |
| 21695030628 | MA11 | Corticosteriods | prednisone |
| 182133400 | MA11 | Corticosteriods | prednisone |
| 61919099507 | MA11 | Corticosteriods | prednisone |
| 42254036108 | MA11 | Corticosteriods | prednisone |
| 63629157906 | MA11 | Corticosteriods | prednisone |
| 63739020802 | MA11 | Corticosteriods | prednisone |
| 49999000830 | MA11 | Corticosteriods | prednisone |
| 66267017210 | MA11 | Corticosteriods | prednisone |
| 51138014530 | MA11 | Corticosteriods | prednisone |
| 54569033001 | MA11 | Corticosteriods | prednisone |
| 63874039240 | MA11 | Corticosteriods | prednisone |
| 60346009490 | MA11 | Corticosteriods | prednisone |
| 57866432802 | MA11 | Corticosteriods | prednisone |
| 54872216 | MA11 | Corticosteriods | prednisone |
| 143147705 | MA11 | Corticosteriods | prednisone |
| 60346005815 | MA11 | Corticosteriods | prednisone |
| 52959012640 | MA11 | Corticosteriods | prednisone |
| 247007135 | MA11 | Corticosteriods | prednisone |
| 55700020920 | MA11 | Corticosteriods | prednisone |
| 55700020321 | MA11 | Corticosteriods | prednisone |
| 59746017309 | MA11 | Corticosteriods | prednisone |
| 440816718 | MA11 | Corticosteriods | prednisone |
| 54569330207 | MA11 | Corticosteriods | prednisone |
| 63187080707 | MA11 | Corticosteriods | prednisone |
| 60346005840 | MA11 | Corticosteriods | prednisone |
| 527293441 | MA11 | Corticosteriods | prednisone |
| 43063070310 | MA11 | Corticosteriods | prednisone |
| 59746017106 | MA11 | Corticosteriods | prednisone |
| 21695030630 | MA11 | Corticosteriods | prednisone |
| 54569402604 | MA11 | Corticosteriods | prednisone |
| 49999043703 | MA11 | Corticosteriods | prednisone |
| 55045148001 | MA11 | Corticosteriods | prednisone |
| 10768773304 | MA11 | Corticosteriods | prednisone |
| 51138015630 | MA11 | Corticosteriods | prednisone |
| 21695030721 | MA11 | Corticosteriods | prednisone |
| 603533815 | MA11 | Corticosteriods | prednisone |
| 60687014501 | MA11 | Corticosteriods | prednisone |
| 71335065100 | MA11 | Corticosteriods | prednisone |
| 51079003219 | MA11 | Corticosteriods | prednisone |
| 42254036114 | MA11 | Corticosteriods | prednisone |
| 55175399200 | MA11 | Corticosteriods | prednisone |
| 71205036206 | MA11 | Corticosteriods | prednisone |

| | | | |
|---|---|---|---|
| 61919037330 | MA11 | Corticosteriods | prednisone |
| 55289043840 | MA11 | Corticosteriods | prednisone |
| 54569033004 | MA11 | Corticosteriods | prednisone |
| 247010000 | MA11 | Corticosteriods | prednisone |
| 55175391805 | MA11 | Corticosteriods | prednisone |
| 35356058130 | MA11 | Corticosteriods | prednisone |
| 33358029278 | MA11 | Corticosteriods | prednisone |
| 60760063028 | MA11 | Corticosteriods | prednisone |
| 50090198902 | MA11 | Corticosteriods | prednisone |
| 60346009410 | MA11 | Corticosteriods | prednisone |
| 17317066308 | MA11 | Corticosteriods | prednisone |
| 57866432401 | MA11 | Corticosteriods | prednisone |
| 33358029415 | MA11 | Corticosteriods | prednisone |
| 52959012700 | MA11 | Corticosteriods | prednisone |
| 51079002217 | MA11 | Corticosteriods | prednisone |
| 247007260 | MA11 | Corticosteriods | prednisone |
| 35356081915 | MA11 | Corticosteriods | prednisone |
| 54001825 | MA11 | Corticosteriods | prednisone |
| 527293043 | MA11 | Corticosteriods | prednisone |
| 58016021890 | MA11 | Corticosteriods | prednisone |
| 51079003220 | MA11 | Corticosteriods | prednisone |
| 60346009440 | MA11 | Corticosteriods | prednisone |
| 43063064415 | MA11 | Corticosteriods | prednisone |
| 43063086630 | MA11 | Corticosteriods | prednisone |
| 378064010 | MA11 | Corticosteriods | prednisone |
| 54569033107 | MA11 | Corticosteriods | prednisone |
| 63187030006 | MA11 | Corticosteriods | prednisone |
| 54472825 | MA11 | Corticosteriods | prednisone |
| 52959022000 | MA11 | Corticosteriods | prednisone |
| 58016021728 | MA11 | Corticosteriods | prednisone |
| 49999002840 | MA11 | Corticosteriods | prednisone |
| 51138014430 | MA11 | Corticosteriods | prednisone |
| 63187002010 | MA11 | Corticosteriods | prednisone |
| 54868090800 | MA11 | Corticosteriods | prednisone |
| 16590036521 | MA11 | Corticosteriods | prednisone |
| 23490615706 | MA11 | Corticosteriods | prednisone |
| 43063009703 | MA11 | Corticosteriods | prednisone |
| 50090327307 | MA11 | Corticosteriods | prednisone |
| 54868118308 | MA11 | Corticosteriods | prednisone |
| 16590040440 | MA11 | Corticosteriods | prednisone |
| 51079003201 | MA11 | Corticosteriods | prednisone |
| 63874032719 | MA11 | Corticosteriods | prednisone |
| 55887064340 | MA11 | Corticosteriods | prednisone |
| 60346009409 | MA11 | Corticosteriods | prednisone |
| 51552002802 | MA11 | Corticosteriods | prednisone |
| 59746017206 | MA11 | Corticosteriods | prednisone |
| 51552002899 | MA11 | Corticosteriods | prednisone |
| 55175391807 | MA11 | Corticosteriods | prednisone |
| 43063059012 | MA11 | Corticosteriods | prednisone |
| 58864042330 | MA11 | Corticosteriods | prednisone |
| 603533821 | MA11 | Corticosteriods | prednisone |
| 63874032728 | MA11 | Corticosteriods | prednisone |
| 23490615809 | MA11 | Corticosteriods | prednisone |
| 247010020 | MA11 | Corticosteriods | prednisone |
| 55045148006 | MA11 | Corticosteriods | prednisone |
| 60346051520 | MA11 | Corticosteriods | prednisone |
| 66116041030 | MA11 | Corticosteriods | prednisone |
| 52959095405 | MA11 | Corticosteriods | prednisone |
| 440816706 | MA11 | Corticosteriods | prednisone |
| 51138014720 | MA11 | Corticosteriods | prednisone |
| 591505210 | MA11 | Corticosteriods | prednisone |
| 16590032620 | MA11 | Corticosteriods | prednisone |
| 21695030650 | MA11 | Corticosteriods | prednisone |
| 54001729 | MA11 | Corticosteriods | prednisone |
| 55289037372 | MA11 | Corticosteriods | prednisone |
| 71205023920 | MA11 | Corticosteriods | prednisone |
| 58016021820 | MA11 | Corticosteriods | prednisone |
| 68387024115 | MA11 | Corticosteriods | prednisone |
| 52544083110 | MA11 | Corticosteriods | prednisone |
| 403077450 | MA11 | Corticosteriods | prednisone |
| 55175184301 | MA11 | Corticosteriods | prednisone |

| | | | |
|---|---|---|---|
| 54868025807 | MA11 | Corticosteriods | prednisone |
| 403077040 | MA11 | Corticosteriods | prednisone |
| 17856372201 | MA11 | Corticosteriods | prednisone |
| 66336021930 | MA11 | Corticosteriods | prednisone |
| 58864042320 | MA11 | Corticosteriods | prednisone |
| 54569304306 | MA11 | Corticosteriods | prednisone |
| 55289037312 | MA11 | Corticosteriods | prednisone |
| 54868083608 | MA11 | Corticosteriods | prednisone |
| 43063064321 | MA11 | Corticosteriods | prednisone |
| 63629160501 | MA11 | Corticosteriods | prednisone |
| 54569033202 | MA11 | Corticosteriods | prednisone |
| 61919010814 | MA11 | Corticosteriods | prednisone |
| 23490615703 | MA11 | Corticosteriods | prednisone |
| 603533932 | MA11 | Corticosteriods | prednisone |
| 54868090804 | MA11 | Corticosteriods | prednisone |
| 58016021721 | MA11 | Corticosteriods | prednisone |
| 54001820 | MA11 | Corticosteriods | prednisone |
| 63874039221 | MA11 | Corticosteriods | prednisone |
| 247007250 | MA11 | Corticosteriods | prednisone |
| 43063086610 | MA11 | Corticosteriods | prednisone |
| 247007299 | MA11 | Corticosteriods | prednisone |
| 247010004 | MA11 | Corticosteriods | prednisone |
| 54569033304 | MA11 | Corticosteriods | prednisone |
| 51079003320 | MA11 | Corticosteriods | prednisone |
| 59746017509 | MA11 | Corticosteriods | prednisone |
| 49999002812 | MA11 | Corticosteriods | prednisone |
| 54868025801 | MA11 | Corticosteriods | prednisone |
| 54872525 | MA11 | Corticosteriods | prednisone |
| 62584083301 | MA11 | Corticosteriods | prednisone |
| 51138014650 | MA11 | Corticosteriods | prednisone |
| 59746017506 | MA11 | Corticosteriods | prednisone |
| 66267017220 | MA11 | Corticosteriods | prednisone |
| 63874077201 | MA11 | Corticosteriods | prednisone |
| 54569584000 | MA11 | Corticosteriods | prednisone |
| 55045153309 | MA11 | Corticosteriods | prednisone |
| 68115029121 | MA11 | Corticosteriods | prednisone |
| 70934009542 | MA11 | Corticosteriods | prednisone |
| 247007248 | MA11 | Corticosteriods | prednisone |
| 440816710 | MA11 | Corticosteriods | prednisone |
| 9004502 | MA11 | Corticosteriods | prednisone |
| 52544079701 | MA11 | Corticosteriods | prednisone |
| 61919032610 | MA11 | Corticosteriods | prednisone |
| 52544083201 | MA11 | Corticosteriods | prednisone |
| 603533232 | MA11 | Corticosteriods | prednisone |
| 21695030720 | MA11 | Corticosteriods | prednisone |
| 52959022075 | MA11 | Corticosteriods | prednisone |
| 182133401 | MA11 | Corticosteriods | prednisone |
| 50090198903 | MA11 | Corticosteriods | prednisone |
| 63629157907 | MA11 | Corticosteriods | prednisone |
| 51927143500 | MA11 | Corticosteriods | prednisone micronized |
| 38779015403 | MA11 | Corticosteriods | prednisone micronized |
| 62991120604 | MA11 | Corticosteriods | prednisone micronized |
| 38779015410 | MA11 | Corticosteriods | prednisone micronized |
| 38779015401 | MA11 | Corticosteriods | prednisone micronized |
| 38779015409 | MA11 | Corticosteriods | prednisone micronized |
| 62991120603 | MA11 | Corticosteriods | prednisone micronized |
| 38779015400 | MA11 | Corticosteriods | prednisone micronized |
| 38779015408 | MA11 | Corticosteriods | prednisone micronized |
| 395816562 | MA11 | Corticosteriods | prednisone micronized |
| 62991120602 | MA11 | Corticosteriods | prednisone micronized |
| 62991120601 | MA11 | Corticosteriods | prednisone micronized |
| 38779015404 | MA11 | Corticosteriods | prednisone micronized |
| 395816519 | MA11 | Corticosteriods | prednisone micronized |
| 38779015450 | MA11 | Corticosteriods | prednisone micronized |
| 38779015425 | MA11 | Corticosteriods | prednisone micronized |
| 38779015405 | MA11 | Corticosteriods | prednisone micronized |

| | | | |
|---|---|---|---|
| M0221 | PXCH | Tixagevimab/Cilgavimab | Injection, tixagevimab and cilgavimab, for the pre-exposure prophylaxis only, for certain adults and pediatric individuals (12 years of age and older weighing at least 40kg) with no known SARS-CoV-2 exposure, who either have moderate to severely compromised immune systems or for whom vaccination with any available COVID-19 vaccine is not recommended due to a history of severe adverse reaction to a COVID-19 vaccine(s) and/or COVID-19 vaccine component(s), includes injection and post administration monitoring in the home or residence; this includes a beneficiary's home that has been made provider-based to the hospital during the COVID-19 public health emergency |
| M0220 | PXCH | Tixagevimab/Cilgavimab | Injection, tixagevimab and cilgavimab, for the pre-exposure prophylaxis only, for certain adults and pediatric individuals (12 years of age and older weighing at least 40kg) with no known SARS-CoV-2 exposure, who either have moderate to severely compromised immune systems or for whom vaccination with any available COVID-19 vaccine is not recommended due to a history of severe adverse reaction to a COVID-19 vaccine(s) and/or COVID-19 vaccine component(s), includes injection and post administration monitoring |
| Q0221 | PXCH | Tixagevimab/Cilgavimab | Injection, tixagevimab and cilgavimab, for the pre-exposure prophylaxis only, for certain adults and pediatric individuals (12 years of age and older weighing at least 40kg) with no known SARS-CoV-2 exposure, who either have moderate to severely compromised immune systems or for whom vaccination with any available COVID-19 vaccine is not recommended due to a history of severe adverse reaction to a COVID-19 vaccine(s) and/or COVID-19 vaccine component(s), 600 mg |
| Q0220 | PXCH | Tixagevimab/Cilgavimab | Injection, tixagevimab and cilgavimab, for the pre-exposure prophylaxis only, for certain adults and pediatric individuals (12 years of age and older weighing at least 40kg) with no known SARS-CoV-2 exposure, who either have moderate to severely compromised immune systems or for whom vaccination with any available COVID-19 vaccine is not recommended due to a history of severe adverse reaction to a COVID-19 vaccine(s) and/or COVID-19 vaccine component(s), 300 mg |
| 2587304 | MA00 | Tixagevimab/Cilgavimab | 1.5 ML tixagevimab 100 MG/ML Injection |
| 2587313 | MA00 | Tixagevimab/Cilgavimab | EVUSHELD 2 vial carton (cilgavimab 150 MG per 1.5 ML, tixagevimab 150 MG per 1.5 ML) (EUA) |
| 2587301 | MA00 | Tixagevimab/Cilgavimab | tixagevimab 100 MG/ML |
| 2587302 | MA00 | Tixagevimab/Cilgavimab | tixagevimab Injectable Product |
| 2587303 | MA00 | Tixagevimab/Cilgavimab | tixagevimab Injection |
| 2587300 | MA00 | Tixagevimab/Cilgavimab | tixagevimab |
| 2587312 | MA00 | Tixagevimab/Cilgavimab | {1 (1.5 ML cilgavimab 100 MG/ML Injection) / 1 (1.5 ML tixagevimab 100 MG/ML Injection) } Pack |
| 310744202 | MA11 | Tixagevimab/Cilgavimab | EVUSHELD co-packaged |
| 310889501 | MA11 | Tixagevimab/Cilgavimab | 1 vial of Tixagevimab 150 mg/1.5 mL (100 mg/mL) |
| 310106101 | MA11 | Tixagevimab/Cilgavimab | 1 vial of Cilgavimab 150 mg/1.5 mL (100 mg/mL) |
| 2587304 | PR00 | Tixagevimab/Cilgavimab | 1.5 ML tixagevimab 100 MG/ML Injection |
| 2587313 | PR00 | Tixagevimab/Cilgavimab | EVUSHELD 2 vial carton (cilgavimab 150 MG per 1.5 ML, tixagevimab 150 MG per 1.5 ML) (EUA) |
| 2587301 | PR00 | Tixagevimab/Cilgavimab | tixagevimab 100 MG/ML |
| 2587302 | PR00 | Tixagevimab/Cilgavimab | tixagevimab Injectable Product |
| 2587303 | PR00 | Tixagevimab/Cilgavimab | tixagevimab Injection |
| 2587300 | PR00 | Tixagevimab/Cilgavimab | tixagevimab |
| 2587312 | PR00 | Tixagevimab/Cilgavimab | {1 (1.5 ML cilgavimab 100 MG/ML Injection) / 1 (1.5 ML tixagevimab 100 MG/ML Injection) } Pack |
